# High-risk human papillomavirus cervical infection prevalence in France, 2020-2023: a nationwide, large-scale, and spatially resolved retrospective study comparing opportunistic and organised screening

**DOI:** 10.1101/2024.10.20.24315479

**Authors:** Olivier Supplisson, Nicolas Tessandier, Mathilde Roussel, Stéphanie Haim-Boukobza, Sonia Burrel, Mircea T. Sofonea, Samuel Alizon

**Affiliations:** Center for Interdisciplinary Research in Biology (CIRB), Collège de France, CNRS, INSERM, Université PSL, Paris, France; Sorbonne University, Paris, France; ExposUM Institute, Université de Montpellier, MIVEGEC, Montpellier, France; Laboratoire Cerba, pole infectiologie, Frépillon, France; Cerba HealthCare, Issy-les-Moulineaux, France; CHU de Bordeaux, Service de virologie, Bordeaux, France; CNRS UMR 5234, Fundamental Microbiology and Pathogenicity, Université de Bordeaux, Bordeaux, France; PCCEI, Univ Montpellier, INSERM, EFS, Montpellier, France; Department of Anesthesiology, Critical Care, Intensive Care, Pain and Emergency Medicine, CHU Nîmes, Nîmes, France, Nîmes, France

**Keywords:** High-risk HPV, Cervical infection prevalence, Organised screening, Opportunistic screening, Gaussian random fields, Gaussian Markov random fields

## Abstract

**Background:** In France, cervical cancer screening for females aged 30 to 65 primarily targets high-risk human papillomavirus (HPV) infections using DNA tests (‘HR HPV test’).

**Aim:** The primary goal was to map the prevalence of cervical infections caused by HPV16 and/or 18, or by any of 12 other carcinogenic genotypes (31, 33, 35, 39, 45, 51, 52, 56, 58, 59, 66, and 68). The secondary goal was to compare prevalence estimates obtained from tests conducted after spontaneous medical visits (‘opportunistic screening’) or as part of the national screening programme (‘organised screening’).

**Methods:** The analytic sample contained 362,963 results of HR HPV tests collected, between 2020 and 2023, in metropolitan France. A full hierarchical Bayesian model was used to compute spatially resolved prevalence maps at the postcode level.

**Results:** Among samples from organised and opportunistic screening, 2.9% and 3.8% were positive for HPV16 and/or 18, respectively. For other genotypes, these percentages were 6.9% and 9.4%, respectively.

During the last week of the study period, among females aged 30, opportunistic screening was associated with a greater HPV infection prevalence for HPV16 and/or 18 (other genotypes) in 97.2 [72.9,100.0]% (99.9 [99.3,100.0]%) of postcodes. The probability of this percentage being lower among females aged 66 was below 95% for both genotype groups.

After correcting for this effect, a pronounced northwest/southeast gradient in HR HPV infection prevalence was found across France, for both genotype groups, with some hotspots located at the border with Spain and Italy and Switzerland.

**Conclusion:** Opportunistic screening is associated with systematic inflation in HR HPV infection prevalence.

## 1. Introduction

Human papillomaviruses (HPV) cause a massive public health burden worldwide. In addition to anogenital warts and papillomatosis, persistent HPV infections caused by oncogenic (high-risk, HR) genotypes are responsible for nearly all cervical cancers and a substantial proportion of penile, vulval, vaginal, anal, and oropharyngeal cancers [1]. In 2020, an estimated 600,000 new cases of cervical cancer were reported globally, making it the fourth most common cancer among females [2, 3]. In the European Union, it ranks as the second most prevalent cancer among females aged 15–44 years, after breast cancer [4]. The World Health Organisation (WHO)’s global strategy to accelerate the elimination of cervical cancer as a public health problem promotes HR HPV screening as one of the key interventions to prevent and control the burden associated with this disease [5].

In 2019, France updated its cervical cancer screening guidelines, shifting from cytology-based screening to a combination of cytological analysis and HPV DNA detection test (further simplified as ‘HR HPV test’). These updated guidelines keep cytological analysis as the primary screening tool for females aged less than 30, but now recommend HR HPV test as the primary screening method for females aged 30 to 65. The first HR HPV test should be performed as early as age 30 or 3 years after the last cytological analysis. If this test is negative for all HR HPV, the next test should be performed 5 years later. In the case of a positive result for one of the targeted genotypes, a cytological analysis should be performed within the year. If the cytological analysis is normal, a new HR HPV test should be performed one year later [6, 7].

The national cervical cancer screening programme was updated in July 2020 with these new guidelines. This programme, which was launched in 2018 following encouraging results from modelling and pilot studies [8, 9], aimed at reducing the incidence of cervical cancer and associated mortality by 30% over 10 years through an increase in screening coverage rate of up to 80% [6, 7, 10]. It was partly motivated by the concerns about the observed decreasing trend in ‘opportunistic screening’ uptake, i.e., prescription-based screening following spontaneous medical consultations [11]. The new version of the French organised screening programme targets females aged 30 to 65 who do not follow French screening guidelines by inviting them to consult a primary care provider and undergo a free HR HPV test (see Supplementary Files S1 for additional details).

As of 2025, many countries have implemented similar organised cervical cancer screening programmes [12, 13]. As in France, these programmes typically rely on a combination of cytological analyses and HR HPV tests. Beyond their contribution to reducing the incidence and mortality of cervical cancer [14–16], screening programmes relying on HR HPV tests offer a unique window on HR HPV cervical infection prevalence. Furthermore, due to different selection mechanisms in screening uptake, prevalence estimates obtained through organised screening programmes are expected to differ from those derived from opportunistic screening [17, 18].

This work aimed to provide the first spatially-resolved picture of cervical infections caused by HPV16 and/or HPV18, or caused by at least one of 12 other carcinogenic genotypes (31, 33, 35, 39, 45, 51, 52, 56, 58, 59, 66, and 68), according to the WHO classification [19], in metropolitan France. It also aimed to study whether opportunistic and organised screening are associated with systematic differences in prevalence estimates.

## 2. Methods

### 2.1. Data

The initial dataset contained all results from HR HPV tests performed on cervical samples, in France, between January 2020 and November 2023, by Cerba, one of the largest networks of medical biology laboratories in the country.

Regardless of the screening pathway, two types of biologically and clinically validated assays were used to analyse collected samples: eiter the Alinity m high risk® assay or the Roche Cobas® HR HPV test [20]. Both assays target the viral DNA of 14 HPV genotypes: 16, 18, 31, 33, 35, 39, 45, 51, 52, 56, 58, 59, 66, and 68. Alinity m high risk® can distinguish between HPV16, HPV18, and HPV45, but not between other genotypes. Roche’s Cobas® provides distinct results only for HPV16 and 18 and cannot distinguish between other HR genotypes. Therefore, the data extracted by Cerba provided binary results (i.e., positive or negative) for HPV16 and HPV18 for both assays, for HPV45 only for Alinity m high risk®, and for all other HR genotypes for both assays.

In addition to test results, the dataset included the screening pathway, the week and year of the sample analysis, patients’ age and postcode of the city of residence. Details about these postcodes are provided in Supplementary Files S2. No additional variables were available.

### 2.2. Analytic sample selection

The detailed description of the sample selection steps and the corresponding flowchart are provided in Supplementary Files S3. In brief, we only analysed test results from females living in France, aged between 15 and 79 years, for whom information about their anonymous ID, age, spatial location, and type of screening was available.

We considered all patients aged 15 to 79, living in metropolitan France and in Overseas Territories for descriptive statistics purposes only (reported in Supplementary Files S4). The statistical analysis, aimed at fulfilling the study objectives, was conducted solely on patients from metropolitan France within the age range of 30 to 66 years, which we refer to as ‘analytic sample’.

The upper age limit was set at 66 years to account for the delay between the receipt of the invitation from the organised screening programme and test uptake; which may lead females of such an age to benefit from the free HR HPV test provided by the organised screening programme. The lower age limit aligns with the recommended age for HR HPV testing.

### 2.3. Statistical inference

We performed a disease mapping statistical analysis [21], which aims at providing disease-related metrics at small-area levels, such as postcodes. It seeks to do so despite scarce sampling and data dependency across space and time. Scarce sampling may prevent the computation of the quantities of interest or may be the source of unstable estimates with large, implausible, uncertainty intervals [22]. Data dependency manifests itself through spatial and temporal autocorrelation and must be accounted for when computing uncertainty intervals [23]. State-of-the-art disease mapping analyses use hierarchical models estimated within a full Bayesian framework, which allow to capture both the relation between data points and to leverage information from densely sampled areas to enhance estimates in sparsely sampled regions, to deal with both issues [21, 24, 25].

The number of tests and positive tests were aggregated at the stratum level. Strata were based on postcode, age, week-year of screening, genotype groups, and screening pathway. Two groups of genotypes were considered: HPV16 and/or HPV18 (‘HPV16/18’) and genotypes other than HPV16/18 (‘other genotypes’).

The number of positive tests within each stratum was assigned a binomial distribution, the size parameter of which was the total number of tests performed within each stratum. The expected proportion of positive tests within a stratum was linked, through the standard logistic function, to a set of predictors using stratum’s characteristics as input.

Each stratum’s predictor included a global intercept and additional parameters associated with the full two-way interaction between dummy variables associated with the group of other genotypes and opportunistic screening. Other parameters were divided into two groups. The first group was common to data collected through opportunistic and organised screening, while the second was specific to data collected through opportunistic screening. Each group of parameters contained spatially varying, age-varying, and time-varying parameters to which we assigned multivariate hierarchical priors. Six different models were considered, each with a different prior spatial structure.

Competing models were discriminated through the log-score [26], which was computed using the leave-one-group-out cross-validation approach described in [27, 28]. All models were estimated using the inlabru package [29, 30], a wrapper around the R-INLA package [31, 32], in R version 4.4.0 [33]. Further details are provided in Supplementary Files S5.

## 3. Results

### Descriptive reporting for the analytic sample

The analytic sample contained the results of 362,963 HR HPV tests. Of these, 3.7% and 9.2% were positive for HPV16/18 and for other genotypes, respectively. For opportunistic screening, these percentages were 3.8% and 9.4%, for HPV16/18 and other genotypes, respectively. For organised screening, they were 2.9% and 6.9% for both groups, respectively. Stratified observed prevalence are reported in Supplementary Files S7.

### Posterior predictive check for the selected model

The selected model’s spatial components were specified through the BYM2 prior, with the first-order Queen contiguity structure as the neighbourhood matrix. It was able to reproduce the observed prevalence and found a posterior average expected [equal-tailed interval at 95%, ETI95%] cervical infection prevalence in the analytic sample of 3.7 [3.7,3.8]% for HPV16/18 and 9.3 [9.2,9.4]% for other genotypes. Keeping the same order for the genotypes, the posterior average [ETI95] predictive cervical infection prevalence among opportunistic tests was 3.8 [3.7,3.8]% and 9.4 [9.3,9.5]%, respectively. For organised screening, they were 2.9 [2.7,3.1]% and 6.9 [6.6,7.3]%, respectively. Additional stratification can be found in Supplementary Files S7.

### Effect of screening pathway on the expected prevalence among females aged 30

The posterior distribution of the spatial-specific difference between the expected HR HPV cervical infection prevalence generated using the model specific to each type of screening made it possible to quantify the difference in expected prevalence associated with opportunistic screening relative to organised screening. For comparison purposes and given the non-linear relation between the expected prevalence and strata characteristics, we focused here on females aged 30 (the age at which the recommendation to use HR HPV tests begins), considering the last observed week, at the end of November 2023.

At the end of the study period, opportunistic screening was associated with a greater HPV infection prevalence among females aged 30, compared with organised screening, in more than the majority of postcodes with high certainty. Such an inflation was identified on average [ETI95%] in 97.2 [72.9,100.0]% of postcodes for HPV16/18 and 99.9 [99.3,100.0]% of postcodes for other genotypes.

The difference in HR HPV infection prevalence, between opportunistic and organized screening, among females aged 30 in the postcode with the greatest ETI95% lower bound was on average [ETI95%] 3.3 [1.1,5.6] percentage points (pp) for HPV16/18 and 12.4 [7.3,18.3]pp for other genotypes (Figure 1).

**Figure 1:**
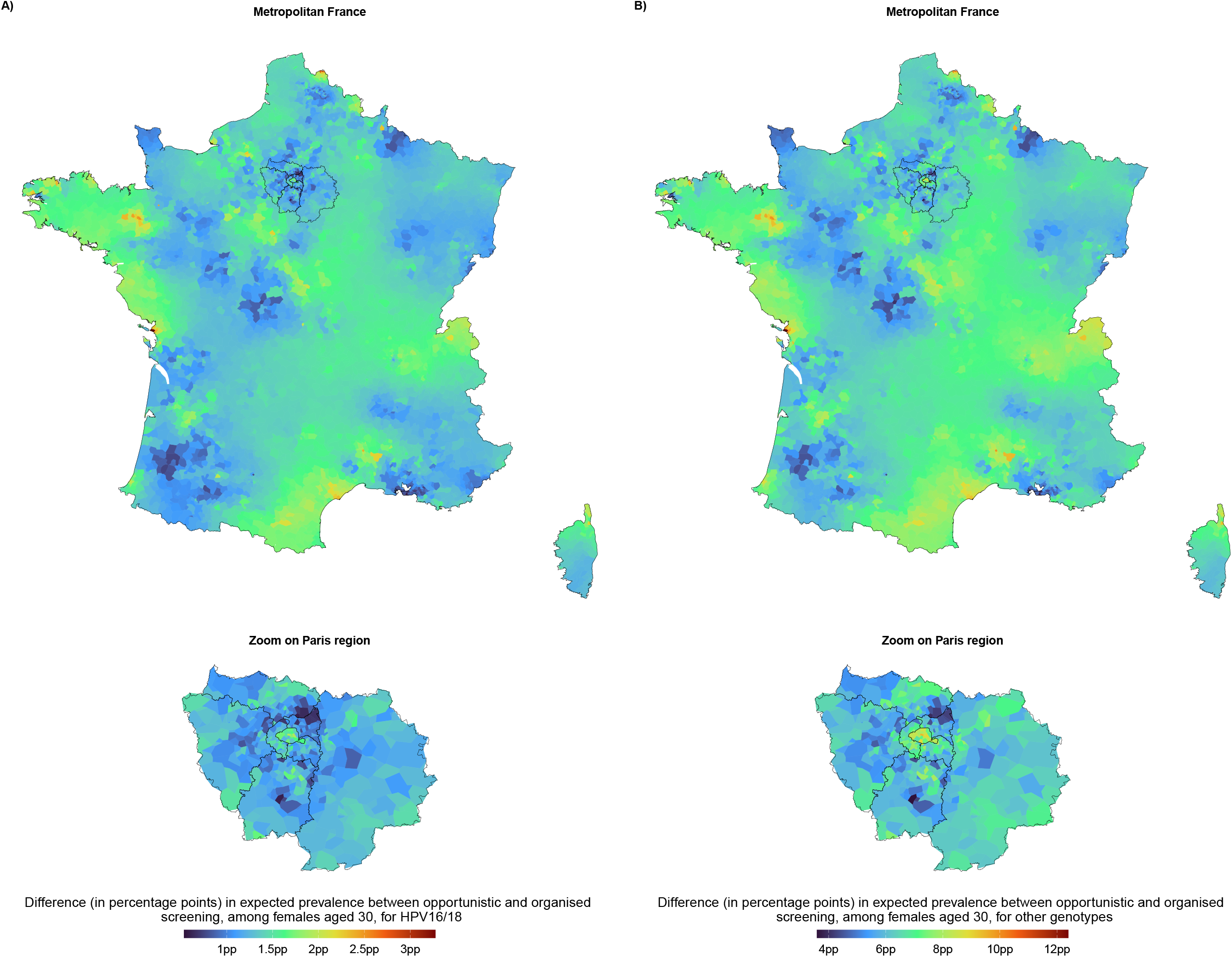
Posterior average difference between opportunistic and organised screening in terms of expected HR HPV cervical infection prevalence in France (upper row) and Paris region (bottom row) for HPV16/18 (A) and other genotypes (B). Values are in percentage points (pp), among females aged 30, at the end of November 2023. Maps with ETI95% are provided in Supplementary Files S8.

### Maps of the expected HR HPV cervical infection prevalence among females aged 30

The joint posterior distribution of the model specific to organised screening allowed us to generate the spatial-specific expected HR HPV cervical infection prevalence under organised screening, among females aged 30, at the end of November 2023. This made it possible to produce what, to our knowledge, is the first spatially resolved nationwide mapping of this quantity for France.

Once removed the systematic inflation associated with opportunistic screening, a pronounced northwest/southeast gradient in HR HPV infection prevalence was found across France, for both genotype groups. Hotspots were identified at the borders with Spain and Italy and Switzerland.

The areas in which we detected the lowest ETI95% lower bound for the expected prevalence had an posterior average [ETI95%] expected prevalence of 2.4 [0.8,5.6]% for HPV16/18 and 8.1 [3.0,16.8]% for other genotypes. By contrast, the area in which the maximum lower bound was detected had a posterior average [ETI95%] expected prevalence of 8.6 [6.3,11.2]% for HPV16/18 and 29.5 [23.7,35.8]% for other genotypes (Figure 2A and B). Maps with ETI95% upper and lower bounds are provided in Supplementary Files S9.

**Figure 2:**
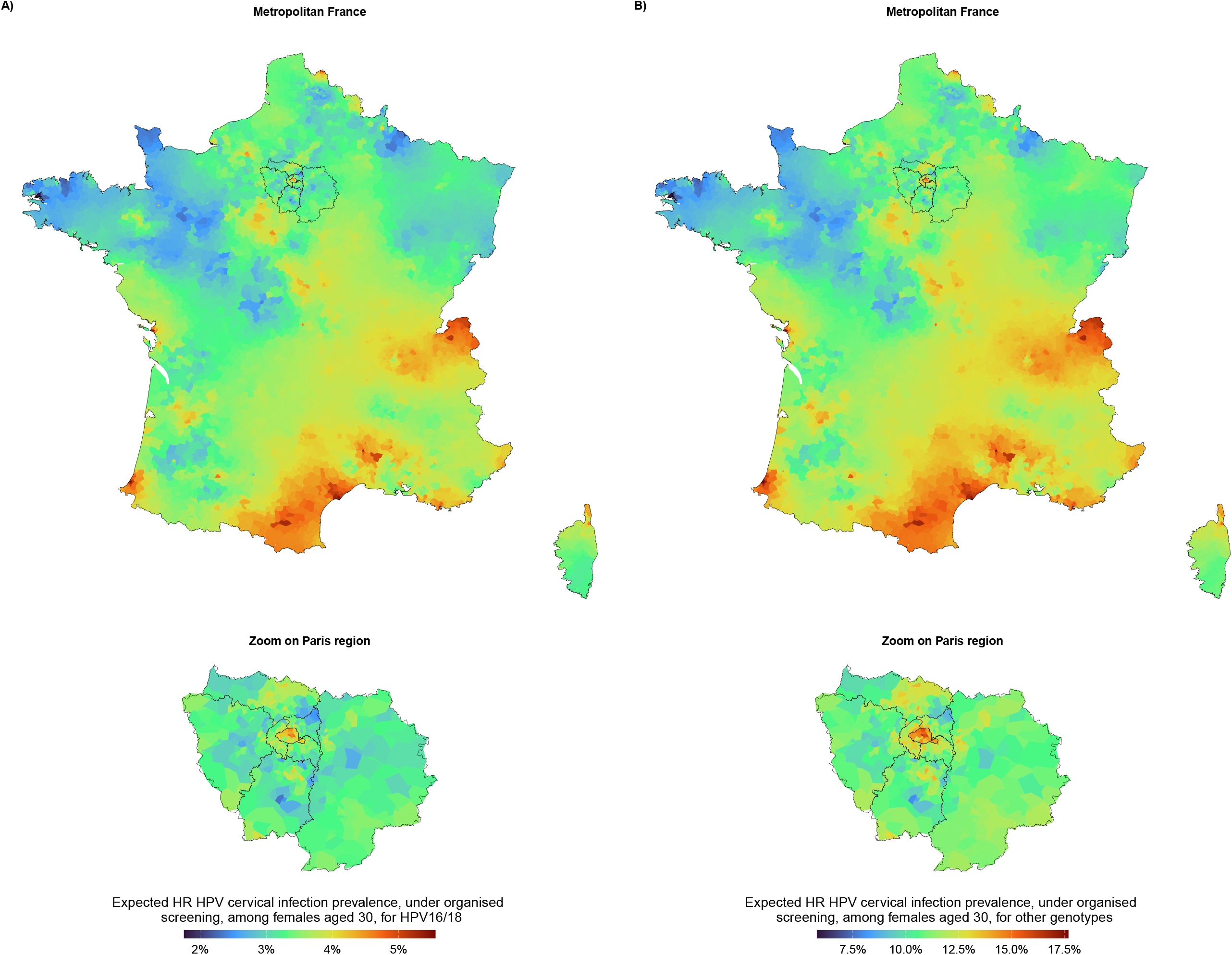
Posterior average expected HR HPV cervical infection prevalence in France (top rows) and Paris region (bottom row), among females aged 30, at the end of November 2023, under organised screening, for HPV16/18 (A) and other genotypes (B). Maps with ETI95% upper and lower bounds are provided in Supplementary Files S9.

### Age-specific expected HR HPV cervical infection prevalence in major French cities

To study how the expected HR HPV cervical infection prevalence changes with age, we restricted the evaluation of the posterior expected prevalence to eleven major French cities, spread around mainland France, at the end of November 2023 (see Supplementary Files S2 for their locations).

The likely systematic upward inflation identified among females aged 30 extended to almost all ages in major French cities. In Paris, the posterior ETI95%lower bound for this upward inflation decreased from 2.0 [0.2,3.7]pp for HPV16/18 and 9.4 [5.1,13.8]pp for other genotypes among females aged 30 to 1.0 [-0.3,2.3]pp and 2.7 [0.1,5.4]pp, respectively, among females aged 66.

When removing the systematic inflation associated with opportunistic screening, the posterior average [ETI95%] expected HPV16/18 cervical infection prevalence in Paris decreased from 4.7 [3.3,6.6]% among females aged 30 to 3.4 [2.3,4.9]% among females aged 66. For other genotypes, it decreased from 16.4 [12.6,20.7]% to 9.7 [7.1,12.8]% (Figure 3A and B). Maps for the entire country are shown in Supplementary Files S10.

**Figure 3:**
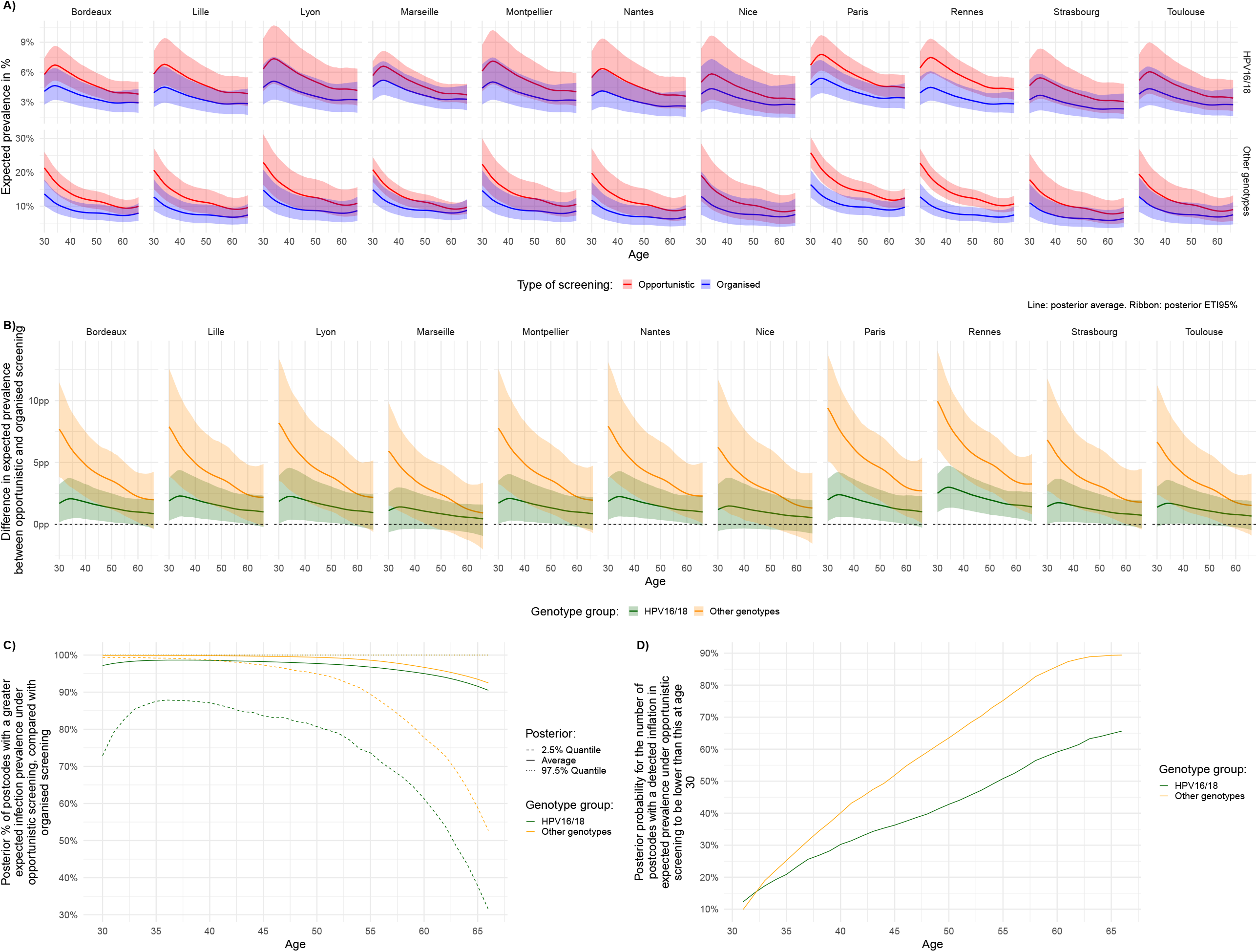
A) Summary of the posterior distribution of the expected HR HPV cervical infection prevalence, in major French cities, stratified by age and genotype group. B) Summary of the posterior distribution of the difference (in percentage points), between the two groups of genotypes (other genotypes and HPV16/18), in the expected prevalence of cervical infection, in major French cities, stratified by age. C) Posterior proportion (in %) of postcodes for which a systematic increase in expected infection prevalence was detected with opportunistic screening, compared with organised screening, stratified by age and genotype group. D) Posterior probability that the number of postcodes with a higher expected prevalence under opportunistic screening, compared to organised screening, is lower than this at age 30. Table counterparts for these plots are provided in Supplementary Files S12. All variables were computed at the end of November 2023.

Among females aged 66, opportunistic screening was associated with a systematic inflation in the expected infection prevalence in 90.5 [31.4,100.0]% of postcodes for HPV16/18 and 92.5 [52.7,100.0]% for other genotypes, see Figure 3C. There was a 65.7% and 89.4% chance for these percentages to be lower than these computed among females aged 30, respectively for HPV16/18 and other genotypes, see 3D.

### Difference in expected infection prevalence between the two groups of genotypes within each screening pathway

For each screening pathway, the posterior mean difference in infection prevalence between the two groups of oncogenic genotypes followed a northeast-to-southwest gradient. In all major French cities, the prevalence of HPV16/18 was almost certainly lower than that of the other genotypes. Moreover, both screening pathways tended to yield smaller and closer differences in expected prevalence between the two genotype groups among older females compared to younger ones (see Supplementary Files S11).

### Temporal changes in the expected HR HPV cervical infection prevalence

The posterior distribution for the expected HR HPV cervical infection prevalence under opportunistic screening favoured an increase in the expected infection prevalence between the last and first week of the study period, for both groups of genotypes. However, when considering organised screening, such a positive temporal trend was not found with high certainty (Figure 4A and Supplementary Files S13).

**Figure 4:**
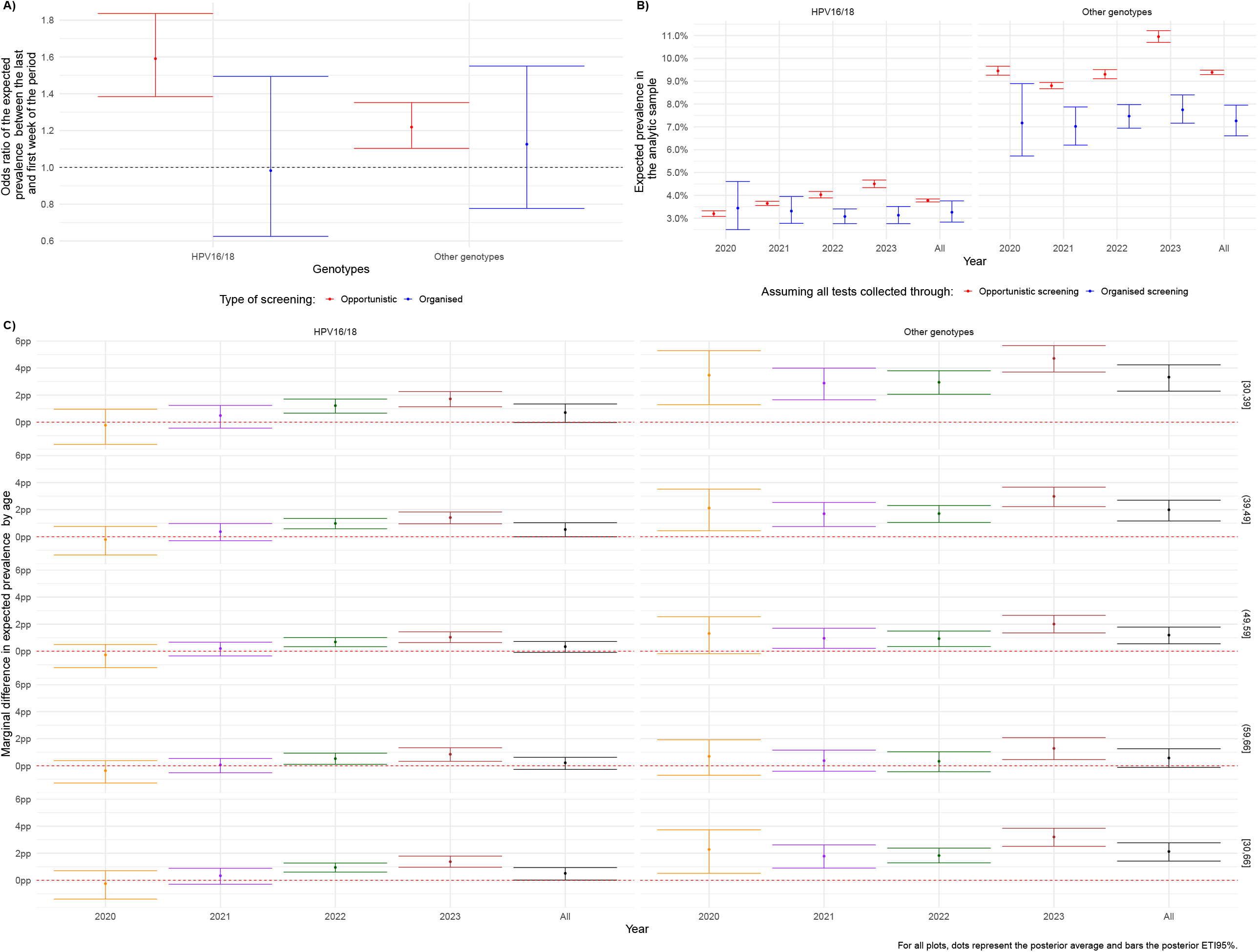
A) Posterior odds ratio between the expected HR HPV cervical infection prevalence during the last and first week of the study period, stratified by type of screening and genotype groups. B) Posterior expected HR HPV cervical infection prevalence simulated for the analytic sample assuming that all data would have been collected only through opportunistic or organised screening, stratified by year of screening. C) Posterior marginal difference (in the analytic sample) in the expected prevalence between opportunistic and organised screening, stratified by year of screening and age. Panels B and C are available in table format in Supplementary Files S14.

### Marginal difference in expected HR HPV cervical infection prevalence associated with screening pathway

To go beyond the conditional picture explored so far, we computed the Marginal Difference in Expected Prevalence (MDEP), which measures the marginal difference in expected prevalence had all data been collected via opportunistic rather than organised screening. It accounts for all interactions and reflects how the within-sample expected prevalence would change depending on screening pathway.

Assuming exclusive collection of data via organised screening, the posterior average [ETI95%] expected prevalence would be 3.3 [2.8,3.8]% for HPV16/18 and 7.3 [6.6,7.9]% for other genotypes. The alternative situation, in which all strata would have been collected through opportunistic screening would be associated with an average [ETI95%] change in the expected prevalence of 0.5 [0.0,0.9]pp and 2.1 [1.4,2.8]pp, for HPV16/18 and other genotypes, respectively (Figure 4A and B). The MDEP stratified by age and year is provided in Figure 4C.

### Sensitivity analyses

The results of the sensitivity analyses can be found in Supplementary Files S15. Only slight changes in the posterior average and ETI95% for the MDEP were observed.

## 4. Discussion

Screening and vaccination are two of the interventions recommended by the WHO strategy to eliminate cervical cancer [2, 3, 34]. The optimisation of their spatial deployment could contribute to making them more effective. To this end, spatially resolved maps of HR HPV infection prevalence can help identify areas where they can be the most effective.

Our study provides the first spatially resolved picture of HR HPV cervical infection prevalence in metropolitan France. Our modelling approach was designed to handle non-linear age-specific prevalence patterns, the spatial structure of the dataset, and multiple genotype groups and screening types within a unified statistical modelling approach.

The analysis highlighted the systematic inflation of HR HPV cervical infection prevalence when using test results collected through opportunistic screening compared to those collected through organised screening. It also identified a northeast/southwest gradient in the HR HPV cervical infection prevalence with prevalence hotspots identified near the border with Spain and Italy and Switzerland. This identified spatial gradient correlates with the spatial gradient in HR HPV vaccines uptake found by Ribassin-Majed *et al*. [35], providing a possible explanation for this pattern.

### HR HPV16/18 cervical infection prevalence

The 2023 Summary Report on HPV and Related Diseases in the World published by the Catalan Institute of Oncology (ICO)/the International Agency for Research on Cancer (IARC) Information Centre on HPV and Cancer, found a pooled 95% confidence interval (CI95%) cervical infection prevalence for HPV16/18 among females with normal cytology of [2.5,2.7]% in Western Europe (see [36], page 103).

For France, the same report identified 12 studies estimating HR HPV cervical infection prevalence in females with normal cervical cytology, all published in 2014 or before (see [36], page 97). Among these, Heard *et al*. [37] stands out with a dataset collected from July 2009 to November 2012, in 16 sampling sites, participating in an experiment for the initial version of the organised screening programme. For normal cytology, they implemented a stratified sampling scheme, according to the age class (25–29, 30–39, 40–49, 50–65 years), resulting in a total of 3,037 cervical samples. HPV16/18 was found in 3.9 [CI95%: 2.8,5.1]% of these samples.

We observed a prevalence for HPV16/18 of 2.9% in the samples collected through organised screening and our model produced a posterior average expected [ETI95%] infection prevalence of 2.9 [2.7,3.1]%. The latter value overlaps with Heard *et al*. [37]’s lower range uncertainty interval while pointing towards lower values that are closer to the ICO/IARC’s estimates for Western Europe, the upper bounds of the uncertainty intervals hint at potentially large difference between our results and Heard et al. [37].

The differences between our results and those from Heard *et al*. [37] could primarily be due to differences in sample composition. The first difference lies in age composition. Heard *et al*. [37] selected participants based on a stratified sampling scheme. Conversely, we used all the tests performed by one of the leading networks of biology laboratories in France but provided model-based age-specific infection prevalence estimates. A second source of heterogeneity is related to vaccination, which increased between previous studies and ours. In 2022, 33.8% of girls aged 16 had a full vaccination scheme, against 15.5% in 2017 [38]. Last but not least, a third source of heterogeneity originates from differences in spatial sampling. We used test results from all over France whereas Heard *et al*. [37] had samples coming from only 5 of the 22 French regions, none of which were located in East of France where we found a lower posterior average expected infection prevalence.

### Public health implications

Many countries, especially in Europe, have implemented nationwide organised HR HPV testing programmes, with the intent to increase screening uptake [13, 39]. In France, according to the latest assessment carried out by the national public health institute (Santé publique France, SpF), the coverage of cervical cancer screening uptake among females aged [25-65] between 2020 and 2022 was 59.5%, below the 80% targeted by the programme [10]. In most of these organised screening programmes, including France, invitations are dispatched based on the time since the previous screening, regardless of residence [13].

The heavy burden of cervical cancer and the benefits associated with HR HPV screening call for HR HPV testing uptake to be encouraged nationwide [14–16]. However, the prevalence hotspots identified in our analysis could help pinpoint areas where increasing screening effort might enhance the public health impact of the organised screening programme while limiting the additional costs.

Our results also shed light on the spatially-differentiated impact of screening pathways on HR HPV infection prevalence, which may contribute to bias prospective modelling studies using routinely collected data and not accounting for such an effect. We provided plausible ranges for this effect, allowing modellers to explore its impact on their results as part of their sensitivity analyses.

### Limits

Our work has three main limitations. First, the large grid used to define strata caused high computational and storage costs, preventing to account for interactions between dimensions. As a result, all models, despite their flexibility, accounted for each dimension independently on the logit-scale.

Second, the dataset did not allow for the study of organised screening uptake among eligible females. A report by SpF found that 11.6% of screening tests for females aged 30-65 in 2020-2022 were performed through the organised screening programme, i.e., following the receipt of an invitation letter [10]. The launch of the updated version of the organised screening programme amidst the COVID-19 pandemic, in July 2020, may have slowed down the roll-out of these invitations and their uptake. However, reported screening coverage for females aged 25-65 was higher in 2020-2022 than in 2017-2019, indicating a plausible limited impact or a catch-up dynamic during time periods with limited disruption of the healthcare sector [10].

Lastly, we shown that data from opportunistic screening were likely associated with a systematic inflation in HR HPV infection prevalence across mainland France compared to organised screening. However, because the screening uptake following receipt of an invitation is not mandatory, data collected through organised screening might not give a more accurate reflection of the true prevalence than opportunistic screening. Therefore, residual selection biases affect both estimated prevalence. Modelling the selection process could address this concern but this is typically impossible using data routinely collected by medical biology laboratories.

### Implication for future research

This study provides an original, nationwide, and spatially resolved picture of HR HPV cervical infection prevalence in France. It also highlights the likely systematic upward inflation in expected prevalence associated with opportunistic screening compared with organised screening.

Similar quantification of the differences between screening pathways for other infections, whose prevalence studies heavily rely on opportunistic sampling, should be performed to improve the reliability of future prospective modelling work assessing the effect of public health interventions.

## Data Availability

Data cannot be shared due to legal constraints associated with the use of the French reference methodology MR-004.
Codes are available on the first author's GitHub page:https://github.com/osupplisson/hpv_prevalence.

## Statements

### Ethical considerations

Study declared to the French Data Protection Authority (the *Commission nationale de l’informatique et des libertés*, CNIL) by the French National Centre for Scientific Research (CNRS) Data Protection Department (DPD) on the 05/07/2024 (number 2228349/3, UMR7241 record). All data processing was compliant with the French reference methodology MR-004 (research not involving the human person, studies and evaluations in the field of health).

### Authors contributions

- Study conceptualisation: O.S., S.H.B., S.A., M.T.S.
- Administrative work: O.S., S.A.
- Funding acquisition: O.S., S.H.B., M.T.S, S.A., S.B.
- Initial data extraction and cleaning: M.R.
- Secondary data cleaning: O.S.
- Initial statistical analysis: O.S.
- Initial draft: O.S.
- Specific domain knowledge: M.R., S.H.B., S.B., S.A., N.T.
- Review of first draft: M.T.S, S.A.
- Review of subsequent drafts: All authors
- Supervision of the statistical analysis: S.A., M.T.S.
- Supervision: S.H.B., M.T.S, S.A., S.B.

### Data and codes

Data cannot be shared due to legal constraints associated with the use of the French reference methodology MR-004.

Codes are available on the first author’s GitHub page: https://github.com/osupplisson/hpv_prevalence.

## Acknowledgements

We are grateful to the genotoul bioinformatics platform Toulouse Occitanie (Bioinfo Genotoul, https://doi.org/10.15454/1.5572369328961167E12) for providing help and computing and storage resources”.

The authors acknowledge the ISO 9001 certified IRD i-Trop HPC (South Green Platform) at IRD Montpellier for providing HPC resources that have contributed to the research results reported within this paper. https://bioinfo.ird.fr/-http://www.southgreen.fr

We are also thankful to Bioclust, the computing cluster of PB-IBENS (LABEX MEMOLIFE members) for providing storage resources. https://www.ibens.bio.ens.psl.eu/spip.php?rubrique55#bioclust

