## Supplementary Files for "High-risk human papillomavirus cervical infection prevalence in France, 2020-2023: a nationwide, large-scale, and spatially resolved retrospective study comparing opportunistic and organised screening"

S7 Number of tests performed in each time step (year-week), stratified by type of screening. 17
S8 Spatial location of all data points (green dots) and mesh (grey edges) used for the finite-elements decomposition approximation of the Gaussian random fields. The blue line delineaetes the areas in which the density of the mesh could change. It was defined to be finner within metropolitan France than outside, where no data could be observed. . 44
S9 Prior density for a Gaussian prior distribution with PC(1,0.5) hyperprior assigned to the precision parameter (blue curve) compared with a standard Gaussian distribution (red curve) . . . . . . 45
S10 PC-prior for the standard deviation of GMRFs. . . . . . 46
S11 Precision for the prior distribution of the latent correlation parameters. . . . . . 47
S12 Spatial neighbourhood matrices considered for BYM2 priors. Black squares indicate that the two areas are neighbours. . . . . . 48
S13 Postcodes with at least one test result observed in the analytic sample. . . . . . 50
S14 Postcodes stratified by the number of HR HPV tests in the analytic sample. . . . . . 51
S15 Postcodes stratified by the number of HR HPV tests in the analytic sample, zoom on Paris region. . . . . . 52
S16 Postcodes only observed through opportunistic screening, organised screening, both opportunistic and organised screening, and not observed in the analytic sample. . . . . 53
S17 Postcodes only observed through opportunistic screening, organised screening, both opportunistic and organised screening, and not observed in the analytic sample, zoom on Paris region. . . . . . 54
S18 Empirical cumulative distribution function of the number of tests within postcodes in the analytic sample. . . . . . 55
S19 Posterior and prior distribution for the correlations between the two groups of genotypes for components specific to organised screening. . . . . . 59
S20 Posterior and prior distribution for the correlations between the two groups of genotypes for components specific to opportunistic screening. . . . . . 60
S21 Posterior difference between the expected HR HPV cervical infection prevalence from opportunistic and organised screening, among females aged 30, at the end of November 2023. . . . . . 115

S22 Posterior difference between the expected HR HPV using opportunistic and organised data, among females aged 30, at the end of November 2023, in Paris region. . . . . . . . 116
S23 Posterior probability of a positive difference in expected HR HPV between opportunistic and organised screening, stratified by genotype group, among females aged 30, at the end of November 2023. . . . . . . . . . . . . . . . . . . . . . . . . . . . . . . . . . . . . . 117
S24 Posterior systematic inflation in HR HPV cervical infection prevalence under opportunistic screening, compared with organised screening, in each of Paris arrondissement, among females aged 30, at the end of November 2023. . . . . . . . . . . 118
S25 Posterior systematic inflation in HR HPV cervical infection prevalence under opportunistic screening, compared with organised screening, in each of Lyon arrondissement, among females aged 30, at the end of November 2023. . . . . . . . . . . 119
S26 Posterior systematic inflation in HR HPV cervical infection prevalence under opportunistic screening, compared with organised screening, in each of Marseille arrondissement, among females aged 30, at the end of November 2023. . . . . . . . . . . 120
S27 Posterior HR HPV cervical infection prevalence in metropolitan France, among females aged 30, at the end of November 2023, correcting for the systematic inflation associated with opportunistic screening, stratified by genotype group. . . . . . . . . . . . . . . . . . 122
S28 Posterior HR HPV cervical infection prevalence in Paris region, among females aged 30, at the end of November 2023, correcting for the systematic inflation associated with opportunistic screening, stratified by genotype group. . . . . . . . . . . . . . . . . . . . . . 123
S29 Posterior HR HPV cervical infection prevalence in each of the Paris arrondissement, among females aged 30, at the end of November 2023, correcting for the systematic inflation associated with opportunistic screening, stratified by genotype group. . . . . . 124
S30 Posterior HR HPV cervical infection prevalence in each of the Lyon arrondissement, among females aged 30, at the end of November 2023, correcting for the systematic inflation associated with opportunistic screening, stratified by genotype group. . . . . . 125
S31 Posterior HR HPV cervical infection prevalence in each of the Marseille arrondissement, among females aged 30, at the end of November 2023, correcting for the systematic inflation associated with opportunistic screening, stratified by genotype group. . . . . . 126
S32 Posterior average expected infection prevalence for HPV16/18 under organised screening, across metropolitan France, stratified by age. . . . . . . . . . . . . . . . . . . . . . 128
S33 Posterior average expected infection prevalence for Other genotypes, under organised screening, across metropolitan France, stratified by age. . . . . . . . . . . . . . . . . . . . . . 129
S34 Summary of the posterior distribution of the difference (in percentage points) in infection prevalence cause by HPB16/18 and other genotypes, under organised screening, among females aged 30, at the end of November 2023. . . . . . . . . . . . . . . . . . . . . . . . . . 130
S35 Summary of the posterior distribution of the difference (in percentage points) in infection prevalence cause by HPB16/18 and other genotypes, under opportunistic screening, among females aged 30, at the end of November 2023. . . . . . . . . . . . . . . . . . . . . . 131
S36 Posterior average expected difference in infection prevalence between other genotypes and HPV16/18, across metropolitan France, stratified by age, under organised screening. 132
S37 Posterior average expected difference in infection prevalence between other genotypes and HPV16/18, across metropolitan France, stratified by age, under opportunistic screening. . . . . . . . . . . . . . . . . . . . . . . . . . . . . . . . . . . . . . . . . . . . . . 133
S38 Summary of the posterior distribution of the difference (in percentage points), between opportunistic and organised screening, in the expected HR HPV cervical infection prevalence, in major French cities, at the end of November 2023, stratified by age and genotype group. . . . . . . . . . . . . . . . . . . . . . . . . . . . . . . . . . . . . . . . . . 134
S39 Difference in expected HR HPV cervical infection prevalence, between the last and first week of the study period in major French cities, stratified by type of screening and age, for HPV16/18. . . . . . . . . . . . . . . . . . . . . . . . . . . . . . . . . . . . . . . . . . . 227

S40 Posterior average (ETI95%) for the expected HR HPV cervical infection prevalence in 11 major cities in France. The precision is defined as 1/variance: the greater the precision, the lower the variance. The baseline case, precision = 0.2, has a U-shape. A precision of 0.4 looks like a uniform distribution on [-1,1]. At a precision of 0.8, extreme values for the correlation are very unlikely. . . . . 234
S41 Posterior average (ETI95%) for the Marginal Difference in Expected Prevalence, stratified by precision for the priors of the latent correlation parameters, age, and year. The precision is defined as 1/variance: the greater the precision, the lower the variance. The baseline case, precision = 0.2, has a U-shape. A precision of 0.4 looks like a uniform distribution on [-1,1]. At a precision of 0.8, extreme values for the correlation are very unlikely. . . . . 235
S42 Posterior average (ETI95%) for the conditional prevalence in major French cities, at the end of November 2023, stratified by genotype group and type of Matérn covariance function (stationary or non-stationary model). . . . . 236
S43 Posterior average (ETI95%) for the Marginal Difference in Expected Prevalence, stratified by the type of spatial component, age, and year. . . . . 237

S5 Logarithmic-score based on LGO-CV. The model with the minimum score was selected. 56
S6 Raw summary provided by R-INLA of the posterior distribution for the latent parameters. 56
S7 Raw summary provided by R-INLA of the posterior distribution for the hyperparameters. 57
S8 Observed HR HPV cervical infection prevalence, posterior predictive HR HPV cervical infection prevalence, and posterior expected HR HPV cervical infection prevalence, stratified by various dimensions. . . . . . . . . . . . . . . . . . . . . . . . . . . . . . . . 61
S9 Posterior expected HR HPV cervical infection prevalence (in %) in major French cities, stratified by type of test, city, and age. Table counterpart to Figure 3A . . . . . . . . . . . 135
S10 Posterior difference in expected HR HPV cervical infection prevalence (in percentage points) in major French cities, between opportunistic and organised screening, stratified by type of test, city, and age. Table counterpart to Figure 3B. . . . . . . . . . . . . . . . . 191
S11 Posterior percentage of postcodes with a greater expected infection prevalence under opportunistic screening, than under organised screening, stratified by age and genotype group, as of November. Table counterpart to Figure 3C. . . . . . . . . . . . . . . . . . . . . 219
S12 Posterior distribution of the difference in the number of postcodes with a higher expected prevalence under opportunistic screening versus organised screening at each age, relative to females aged 30, as of end of November 2023. Table counterpart to Figure 3D. . . . . . . . . . . . . . . . . . . . . . . . . . . . . . . . . . . . . . . . . . . 222
S13 Posterior expected HR HPV infection prevalence (in %) assuming all data would have been collected with opportunistic or organised screening, not stratified or stratified by age, year, or age and year. Provide values from Figure 4B in table format. . . . . . . . . . 228
S14 Marginal Difference in Expected Prevalence (in percentage points), not stratified or stratified by age, year, or age and year. Provide values from Figure 4C in table format. . 232

### S1 Additional details about the French cervical cancer screening guidelines

The latest French guidelines for cervical cancer screening were updated in 2019. These guidelines recommend, for females aged 25 to 29, 2 cytological analyses at a one-year interval if the first result is normal, and, in the absence of abnormality on both cytological analyses, a new cytological analysis 3 years later. In the case of an abnormal cytology result, an HR HPV test should be performed.

For females aged 30 to 65, an HR HPV test should be performed as the first-line test. The first HR HPV test should be performed 3 years after the last normal cytology result, or as early as 30 in the absence of a prior cytology result. If the HR HPV test is negative, the next HR HPV test should be performed 5 years later. In case of a positive HR HPV test for HR HPV, a cytological analysis should be performed within the year. If the cytology result is normal, an HR HPV test should be performed a year later [1, 2]. A detailed note about the management of females with abnormal cervical cytology and positive HR HPV is available in [3].

In France, females can undergo cervical cancer screening through 2 pathways. First, upon reception of an invitation from the French National Social Security System, patients can be screened as part of the French National Cervical Cancer Screening programme.

The current version of this national organised screening programme was set up the 30th of July 2020. The legal document creating this new national screening programme can be accessed on the following link: https://www.legifrance.gouv.fr/loda/id/JORFTEXT000042238343/ (accessed 15th January 2025).

The national organised screening programme proactively invites females who do not follow the screening guidelines by postal mail to visit a doctor, a midwife, or a gynaecologist for a medical examination. As part of this medical examination, a cervical smear can be collected, which is then sent to medical biology or anatomical pathology laboratories for analysis. Alternatively, the biological sampling and analysis can be performed at a medical biology laboratory, following prescription from the primary care provider. Under this screening pathway, referred to as 'organised screening', the biological analysis is free for the patient and the primary care consultation is reimbursed according to standard nationwide rules.

Starting in mid-2024, invitations are also sent online, and the visit to a primary care provider is no longer required: participants can directly visit a medical biology laboratory for biological sampling and analysis.

As an alternative to the organised screening pathway, patients may undergo 'opportunistic screening', meaning that patients spontaneously consult a primary care provider, without any invitation from the French National Social Security System. Any subsequent HR HPV test after this initial consultation is then subject to an out-of-pocket cost.

#### S2 Details about the French administrative organisation

One key issue we had to deal with was the misalignment between two available spatial IDs:

- The France administrative dataset identifies city using a single unique ID, known as the code commune;
- The dataset extracted from Cerba provided the postcode of each patient. Postcodes are defined based on postman's rounds;

Three cases could happen:

- the postcode corresponds to single unique ID in its entirety (one-one relationship);
- the postcode corresponds to several unique IDs in their entirety (one-to-many relationship);
- the postcode corresponds to a single unique ID but not in its entirety (many-to-one relationship);

To deal with this issue, we relied on the only available open source dataset linking both spatial dimension, as of 2014, available on https://www.data.gouv.fr/fr/datasets/fond-de-carte-des-codes-postaux/. As a result of this discrepancy between these two spatial ID, one postcode may include several cities.

The dataset contained only postcodes for metropolitan France. We are not aware of similar dataset for Overseas territories.

The initial data contained 6,048 non-overlapping spatial areas, each associated with a unique postcode in metropolitan France. Among these, we kept only cities not located in small islands around metropolitan France. We ended up with 6,033 unique postcodes, which were used to build the spatial neighbourhood matrix.

Supplementary figure S1 shows the spatial delimitation of all cities considered in the study.

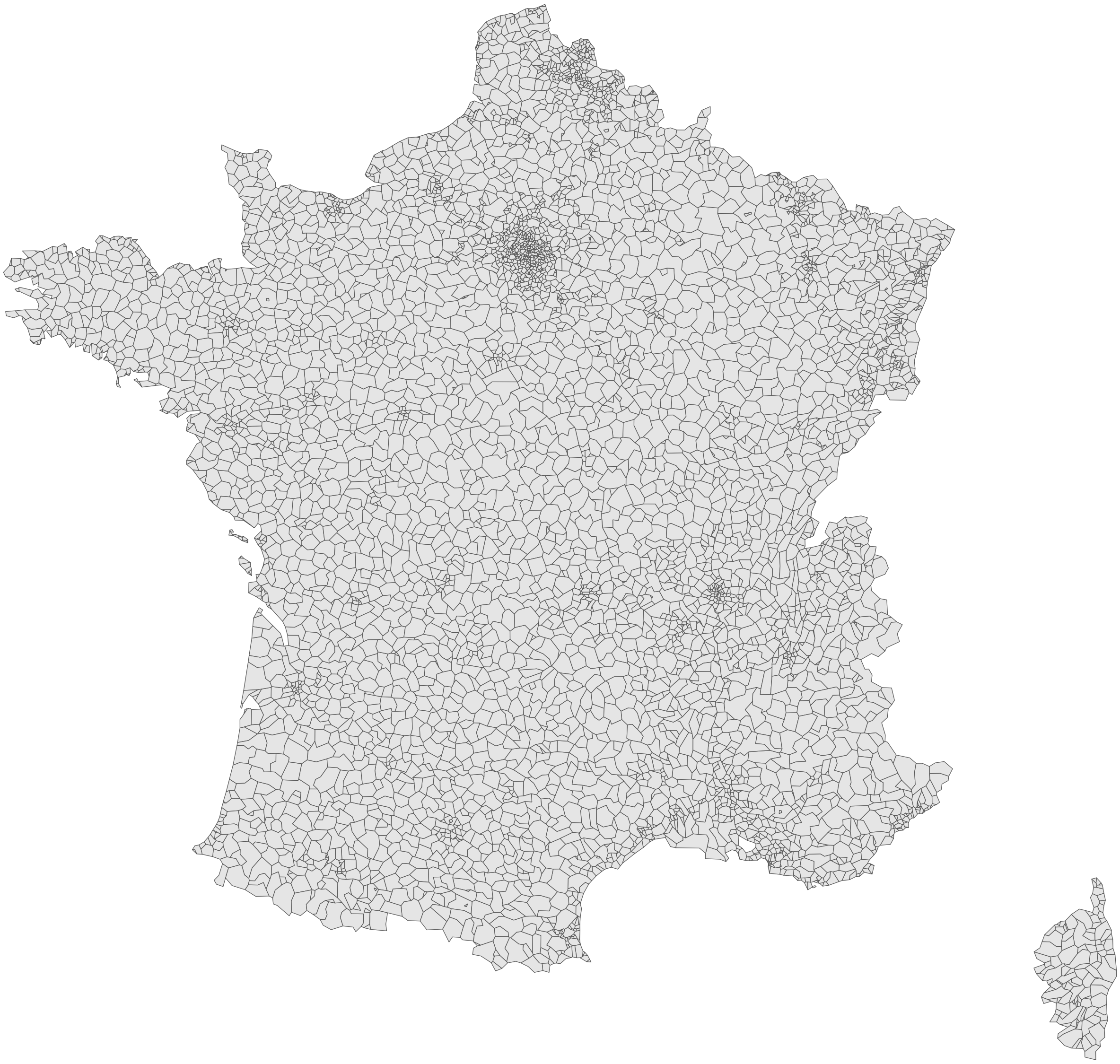

**Figure S1 Layout of postcodes in metropolitan France.**

Among all these cities, we considered specifically 11 of them, for computing the conditional prevalence (Supplementary Figues S2.

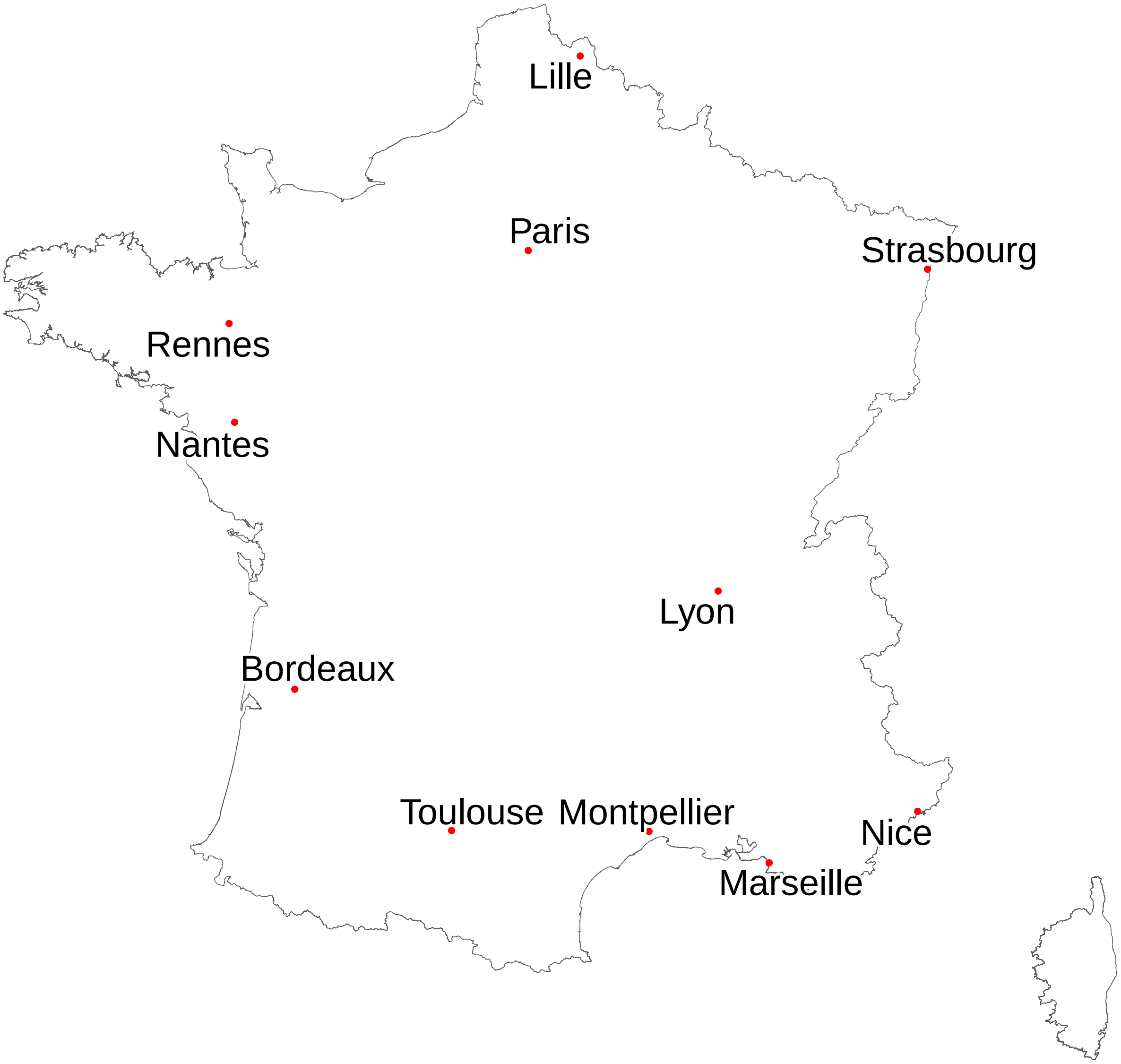

**Figure S2 Centroid of the main French cities in metropolitan France. For Paris, Lyon, and Marseille, the centroid of the 1st arrondissement is displayed.**

In France, cities are organised within administrative divisions known as 'départements' ('districts', in the main text). Metropolitan France comprises 96 districts, grouped into 13 regions. Two of these districts are situated on Corsica, an island located between France and Italy. France's administrative structure is illustrated in Supplementary Figure S3. Paris, France's capital city, lies at the center of the Île-de-France region, depicted in Supplementary Figure S4. In addition to districts located in metropolitan France, 5 districts are located in Overseas territories: Martinique, Guadeloupe, Guyane, Réunion and Mayotte. Of these districts, Guyane is the only one that is continental in nature. Each of this district are also region.

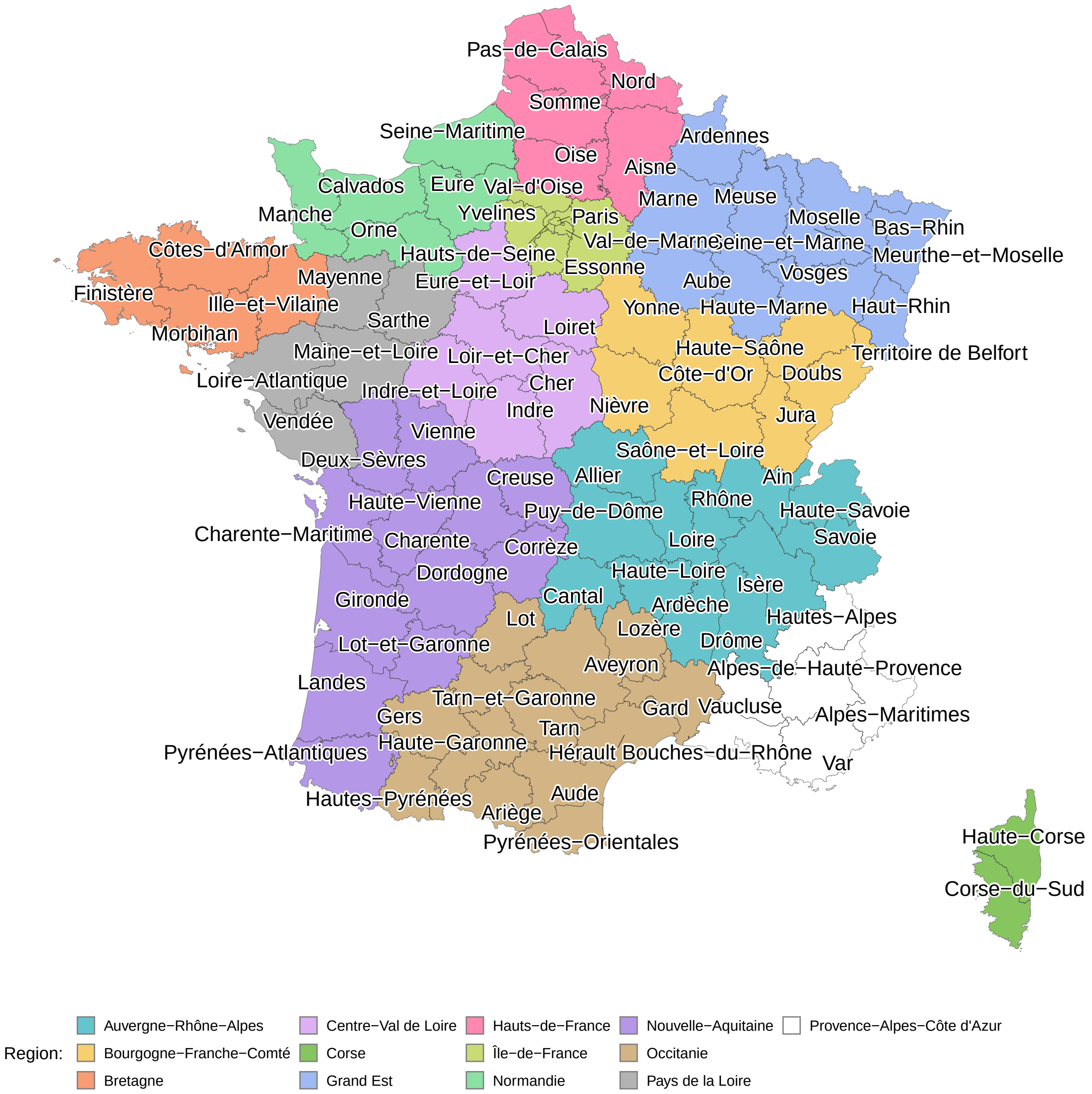

**Figure S3** **Administrative subdivision of metropolitan France.**

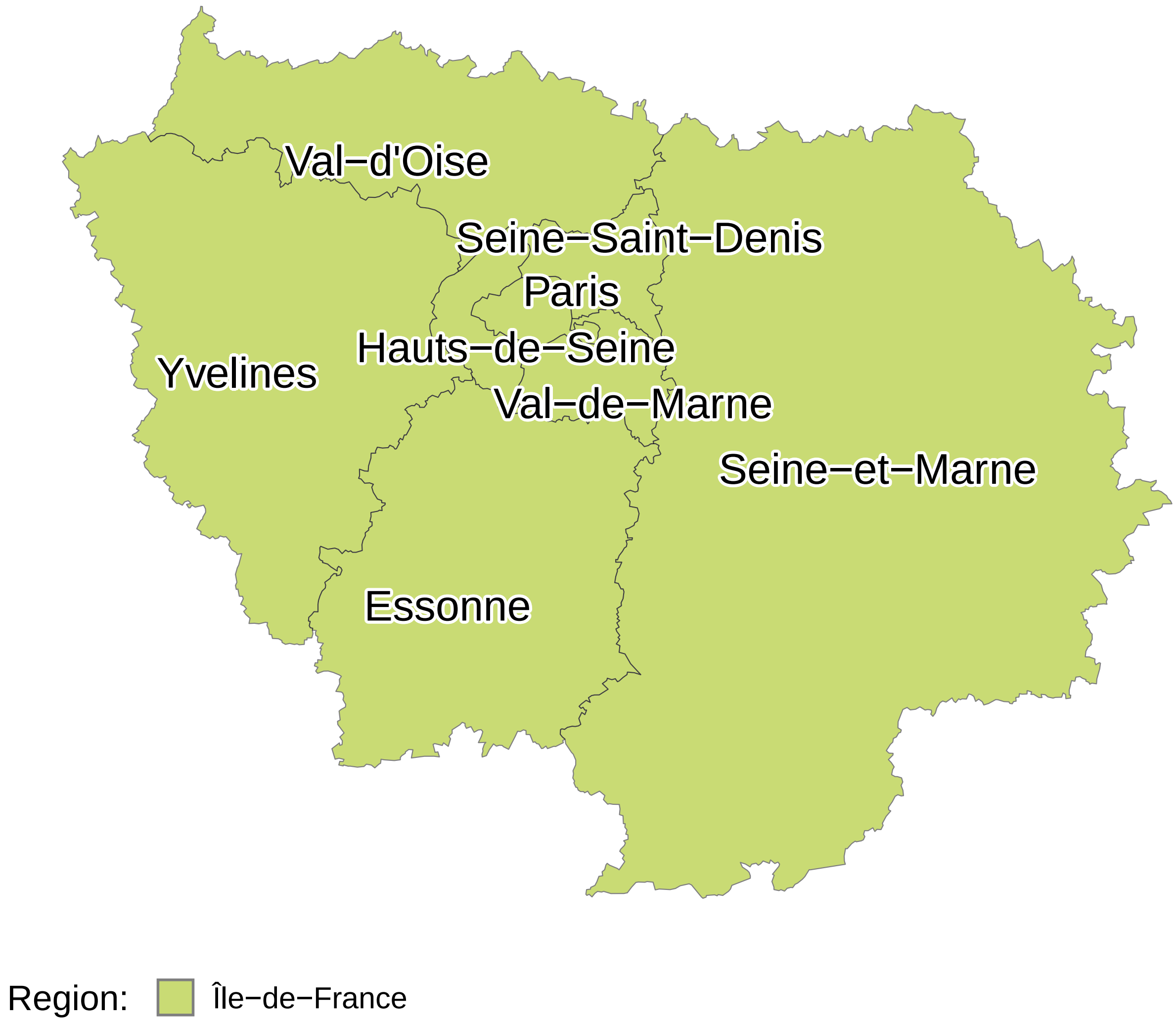

**Figure S4 Administrative subdivision of Paris region (Île-de-France).**

#### S3 Sample selection flowchart

All missing and erroneous items were due to issues at the time of patients' registration, before test uptake, and therefore could be considered as Missing Completely at Random with respect to test results.

Starting from the initial dataset, we excluded all test results associated with patients who had missing age or anonymous ID, because age was required for modelling, and an anonymous ID was necessary to ensure we wouldn't consider the same individuals multiple times in the analysis.

We further restricted the dataset to include only patients aged 15 to 79. We did so because the number of tests outside this age range was small and therefore the descriptive reporting of the results made little sense due to a likely extreme selection process. We also suspected that patients recorded as aged 0 likely had erroneous birthdate entries in Cerba's database.

We then filtered out all tests with missing postcodes or associated with persons living outside France. We did this because our research focused on France.

As a next step, we filtered out tests recorded as both organised and opportunistic screening, which indicated a potential issue at the data reporting step.

Because invitations to take part in the organised screening programme are theoretically restricted to females aged 25 to 65, we removed all tests labeled as organised screening for patients younger than 25 or older than 66. We considered a one-year slack (66 instead of 65-year-old) to allow for a delay between screening invitation and screening uptake.

We then filtered out tests associated with postcodes unmatched in the French administrative dataset.

In the case of individuals testing multiple times, we only kept the most recent test. We did so for three reasons. First, considering population-level modelling helped to slightly alleviate the computational burden associated with model fitting. Second, there was almost no individual testing multiple times within the organised screening pathway, preventing us from considering individual-specific modelling that accounts for screening pathways.

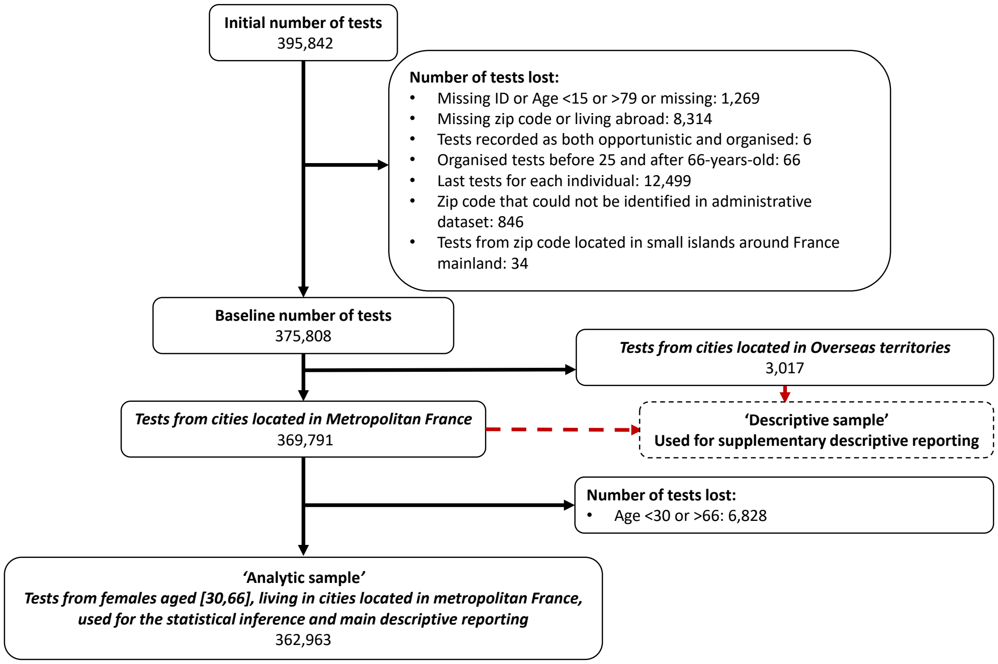

**Figure S5** Analytic sample selection flowchart

### S4 Descriptive reporting for the 'Descriptive samples'

#### S4.1 Descriptive reporting among females aged [15-79] in metropolitan France

The descriptive sample contained the results of 369,791 tests, most of which (around 95%) were collected during opportunistic screening. Test dates ranged from the week of 2020-08-17 to this of 2023-11-27 (i.e., a total of 174 weeks). See Supplementary Figure S7 for the number of tests each week.

The breakdown of these tests by age was highly unbalanced for opportunistic screening but less so for organised screening (Supplementary Figure S6A and B). The number of tests was highly variable between years (Supplementary Figure S6A). This should not be interpreted as a variation in screening effort. First, the lower testing levels in 2020 are attributable to the COVID-19 pandemic context (social distancing measures such as lockdowns, disrupted medical follow-up, and avoidance of medical facilities for fear of higher transmission risk). Second, Cerba and other medical biology laboratories started to implement the 2019 guidelines during 2020. Third, the sharp drop for 2022 and 2023, compared with 2021, was due to a change in Cerba's internal organisation, with an important proportion of collected samples being sent for analysis to another entity of the Cerba group, for which we could not export the data.

During the entire study period, 88.2% of the tests were negative, 2.8% were positive for HPV16, 1.0% were positive for HPV18, and 9.3% were positive for other genotypes. The observed global raw prevalence was greater for opportunistic screening (12.0%) than for the organised one (9.1%). A similar result was found when stratifying by age and year (Supplementary Figure S6D). Details about coinfections are reported in Supplementary Table S2.

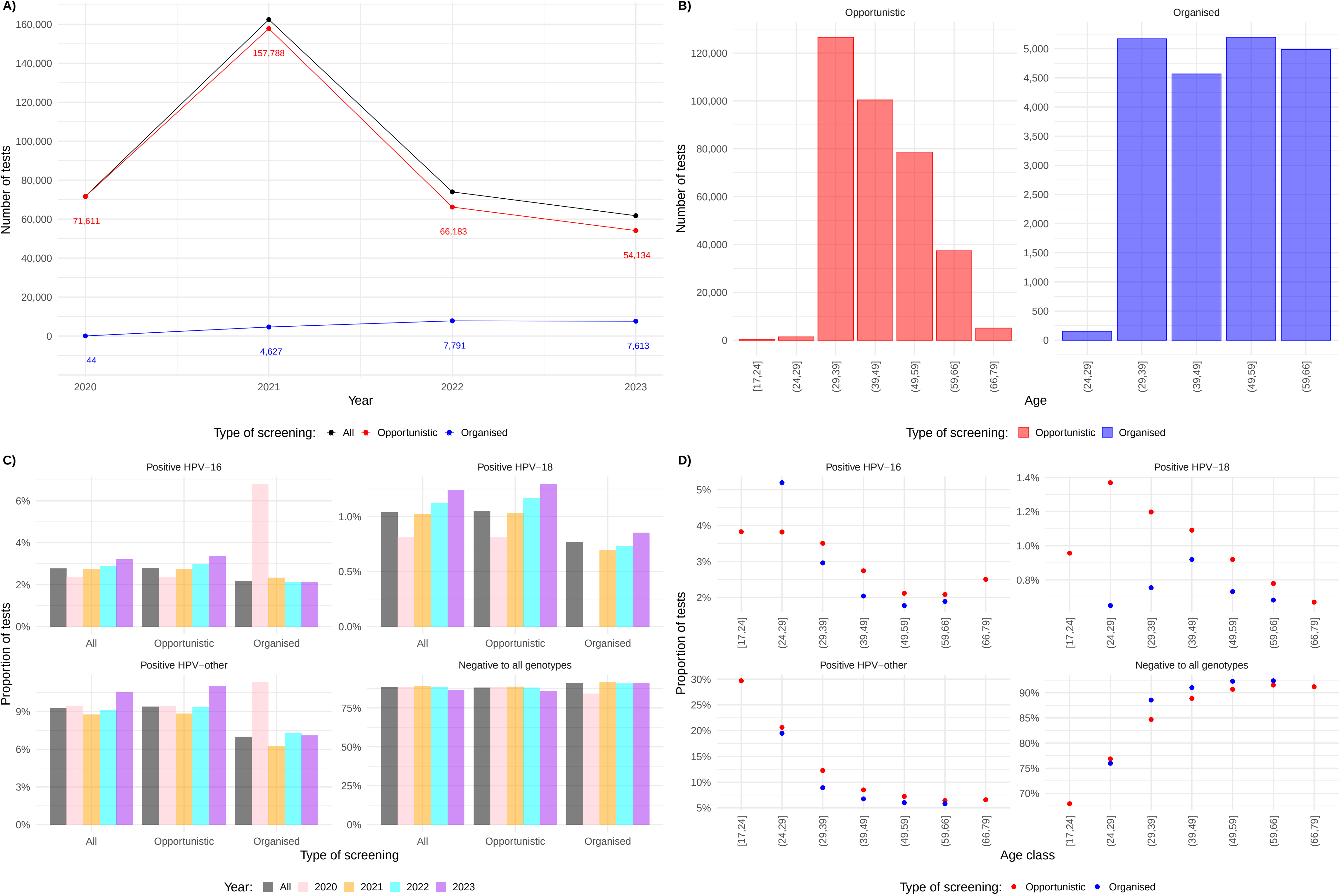

**Figure S6** Descriptive reporting for females aged [15-79] in metropolitan France. A) Number of HR HPV tests, stratified by year and type of screening. B) Number of tests, stratified by age, and type of screening. C) Proportion of tests by result (positive for HPV16, positive for HPV18, positive for other genotypes, and negative), stratified by year and type of screening. D) Proportion of tests by result, stratified by age and type of screening.

**Table S1 Number of tests and observed cervical infection prevalence, stratified by year and type of screening. Table counterpart to Supplementary Figure S6A and C.**

| | | | | Results (% of tests) | | | |
|---|---|---|---|---|---|---|---|
| | | | | Positive | | | |
| Type of screening | Year | Number of tests | % of total test | HPV16 | HPV18 | Other genotypes | Negative |
| All | All | 369,791 | | 2.77 | 1.04 | 9.25 | 88.17 |
| All | 2020 | 71,655 | 19.38 | 2.38 | 0.81 | 9.40 | 88.24 |
| All | 2021 | 162,415 | 43.92 | 2.73 | 1.02 | 8.75 | 88.74 |
| All | 2022 | 73,974 | 20.00 | 2.89 | 1.12 | 9.12 | 88.30 |
| All | 2023 | 61,747 | 16.70 | 3.21 | 1.24 | 10.55 | 86.47 |
| Opportunistic | All | 349,716 | | 2.81 | 1.05 | 9.38 | 88.02 |
| Opportunistic | 2020 | 71,611 | 20.48 | 2.37 | 0.81 | 9.40 | 88.24 |
| Opportunistic | 2021 | 157,788 | 45.12 | 2.74 | 1.03 | 8.82 | 88.65 |
| Opportunistic | 2022 | 66,183 | 18.92 | 2.98 | 1.17 | 9.33 | 88.02 |
| Opportunistic | 2023 | 54,134 | 15.48 | 3.36 | 1.30 | 11.03 | 85.86 |
| Organised | All | 20,075 | | 2.19 | 0.77 | 6.98 | 90.93 |
| Organised | 2020 | 44 | 0.22 | 6.82 | 0.00 | 11.36 | 84.09 |
| Organised | 2021 | 4,627 | 23.05 | 2.33 | 0.69 | 6.27 | 91.74 |
| Organised | 2022 | 7,791 | 38.81 | 2.14 | 0.73 | 7.28 | 90.66 |
| Organised | 2023 | 7,613 | 37.92 | 2.13 | 0.85 | 7.09 | 90.77 |

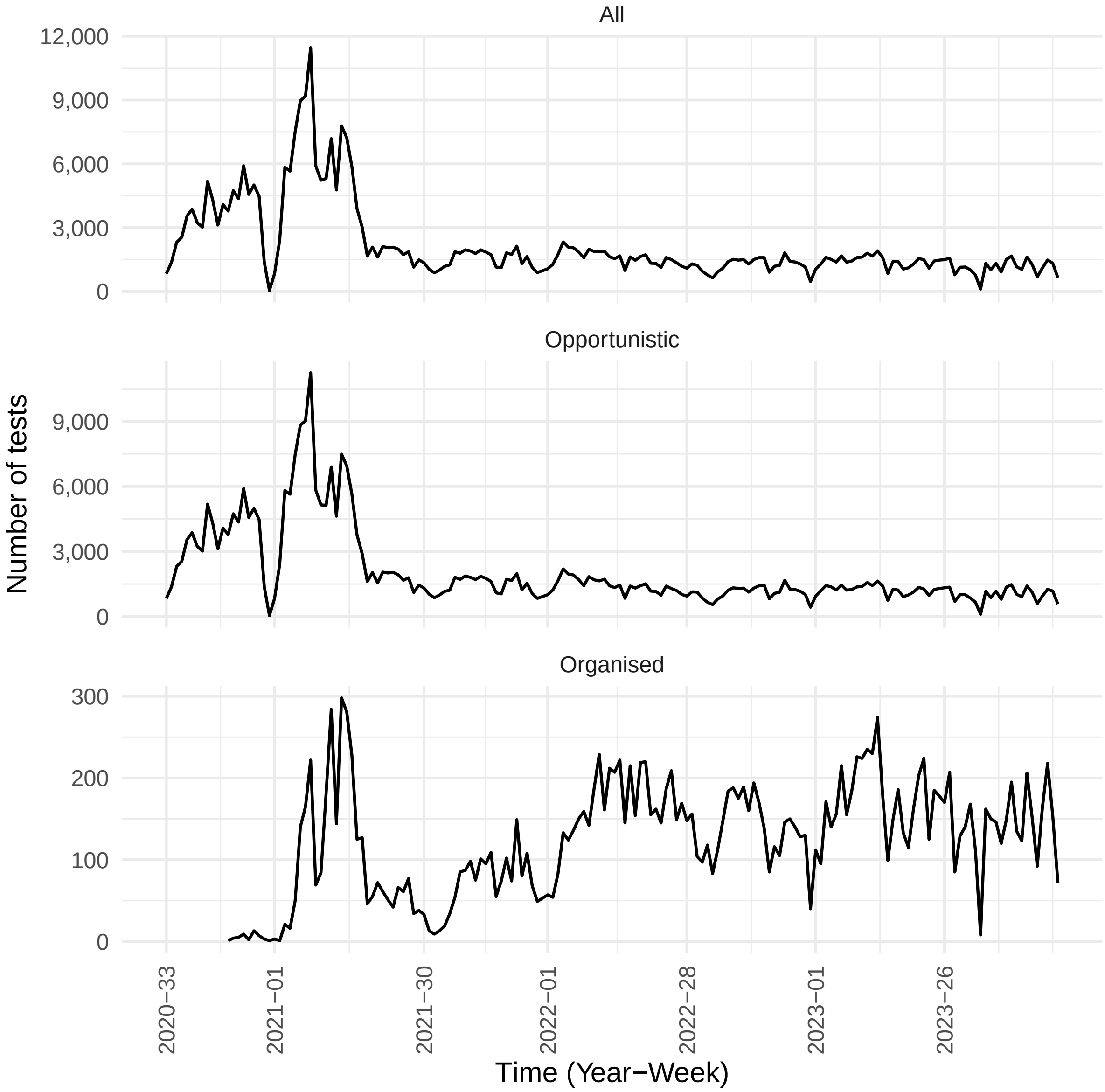

**Figure S7** **Number of tests performed in each time step (year-week), stratified by type of screening.**

**Table S2 Proportion of tests with coinfection, stratified by type of screening and age.**

| Age | Type of screening | Proportion in % (Order: HPV16, HPV18, Other genotypes)* | | | | | | | | Number of tests |
|---|---|---|---|---|---|---|---|---|---|---|
| | | 000 | 001 | 010 | 100 | 011 | 101 | 110 | 111 | |
| All | All | 88.17 | 8.12 | 0.62 | 1.89 | 0.30 | 0.77 | 0.06 | 0.05 | 369,791 |
| All | Organised | 90.93 | 6.16 | 0.48 | 1.58 | 0.24 | 0.56 | 0.02 | 0.02 | 20,075 |
| All | Opportunistic | 88.02 | 8.24 | 0.63 | 1.91 | 0.31 | 0.78 | 0.06 | 0.05 | 349,716 |
| (66,79] | All | 91.20 | 5.65 | 0.41 | 1.79 | 0.24 | 0.69 | 0.02 | 0.00 | 5,078 |
| (66,79] | Opportunistic | 91.20 | 5.65 | 0.41 | 1.79 | 0.24 | 0.69 | 0.02 | 0.00 | 5,078 |
| (59,66] | All | 91.63 | 5.60 | 0.49 | 1.49 | 0.22 | 0.51 | 0.03 | 0.03 | 42,356 |
| (59,66] | Organised | 92.38 | 5.09 | 0.42 | 1.36 | 0.22 | 0.48 | 0.02 | 0.02 | 4,987 |
| (59,66] | Opportunistic | 91.53 | 5.67 | 0.50 | 1.51 | 0.22 | 0.51 | 0.03 | 0.03 | 37,369 |
| (49,59] | All | 90.79 | 6.30 | 0.57 | 1.45 | 0.25 | 0.55 | 0.04 | 0.04 | 83,824 |
| (49,59] | Organised | 92.28 | 5.29 | 0.44 | 1.21 | 0.21 | 0.48 | 0.04 | 0.04 | 5,197 |
| (49,59] | Opportunistic | 90.69 | 6.37 | 0.58 | 1.47 | 0.25 | 0.56 | 0.04 | 0.04 | 78,627 |
| (39,49] | All | 88.96 | 7.37 | 0.66 | 1.90 | 0.31 | 0.69 | 0.06 | 0.05 | 104,989 |
| (39,49] | Organised | 91.02 | 6.04 | 0.61 | 1.60 | 0.28 | 0.42 | 0.02 | 0.00 | 4,567 |
| (39,49] | Opportunistic | 88.86 | 7.43 | 0.66 | 1.92 | 0.31 | 0.70 | 0.07 | 0.05 | 100,422 |
| (29,39] | All | 84.83 | 10.65 | 0.68 | 2.30 | 0.36 | 1.05 | 0.07 | 0.07 | 131,794 |
| (29,39] | Organised | 88.55 | 7.79 | 0.46 | 2.05 | 0.23 | 0.85 | 0.02 | 0.04 | 5,170 |
| (29,39] | Opportunistic | 84.68 | 10.76 | 0.69 | 2.31 | 0.36 | 1.05 | 0.07 | 0.07 | 126,624 |
| (24,29] | All | 76.77 | 18.17 | 0.45 | 2.08 | 0.65 | 1.69 | 0.19 | 0.00 | 1,541 |
| (24,29] | Organised | 75.97 | 18.18 | 0.00 | 4.55 | 0.65 | 0.65 | 0.00 | 0.00 | 154 |
| (24,29] | Opportunistic | 76.86 | 18.17 | 0.50 | 1.80 | 0.65 | 1.80 | 0.22 | 0.00 | 1,387 |
| [17,24] | All | 67.94 | 27.75 | 0.00 | 2.39 | 0.48 | 0.96 | 0.00 | 0.48 | 209 |
| [17,24] | Opportunistic | 67.94 | 27.75 | 0.00 | 2.39 | 0.48 | 0.96 | 0.00 | 0.48 | 209 |

* *: Positive tests are flagged with 1 and negative tests by 0.

**Table S3 Number of tests and observed infection prevalence, stratified by age and type of screening. Table counterpart to Figure S6B and D.**

| Type of screening | Age | Number of tests | % of total test | Results (% of tests) | | | |
|---|---|---|---|---|---|---|---|
| | | | | Positive | | | |
| | | | | HPV16 | HPV18 | Other genotypes | Negative |
| All | [17,24] | 209 | 0.06 | 3.83 | 0.96 | 29.67 | 67.94 |
| All | (24,29] | 1,541 | 0.42 | 3.96 | 1.30 | 20.51 | 76.77 |
| All | (29,39] | 131,794 | 35.64 | 3.48 | 1.18 | 12.12 | 84.83 |
| All | (39,49] | 104,989 | 28.39 | 2.71 | 1.08 | 8.42 | 88.96 |
| All | (49,59] | 83,824 | 22.67 | 2.09 | 0.91 | 7.14 | 90.79 |
| All | (59,66] | 42,356 | 11.45 | 2.06 | 0.77 | 6.36 | 91.63 |
| All | (66,79] | 5,078 | 1.37 | 2.50 | 0.67 | 6.58 | 91.20 |
| Opportunistic | [17,24] | 209 | 0.06 | 3.83 | 0.96 | 29.67 | 67.94 |
| Opportunistic | (24,29] | 1,387 | 0.40 | 3.82 | 1.37 | 20.62 | 76.86 |
| Opportunistic | (29,39] | 126,624 | 36.21 | 3.51 | 1.20 | 12.25 | 84.68 |
| Opportunistic | (39,49] | 100,422 | 28.72 | 2.74 | 1.09 | 8.49 | 88.86 |
| Opportunistic | (49,59] | 78,627 | 22.48 | 2.11 | 0.92 | 7.22 | 90.69 |
| Opportunistic | (59,66] | 37,369 | 10.69 | 2.08 | 0.78 | 6.43 | 91.53 |
| Opportunistic | (66,79] | 5,078 | 1.45 | 2.50 | 0.67 | 6.58 | 91.20 |
| Organised | (24,29] | 154 | 0.77 | 5.19 | 0.65 | 19.48 | 75.97 |
| Organised | (29,39] | 5,170 | 25.75 | 2.96 | 0.75 | 8.92 | 88.55 |
| Organised | (39,49] | 4,567 | 22.75 | 2.04 | 0.92 | 6.74 | 91.02 |
| Organised | (49,59] | 5,197 | 25.89 | 1.77 | 0.73 | 6.02 | 92.28 |
| Organised | (59,66] | 4,987 | 24.84 | 1.88 | 0.68 | 5.82 | 92.38 |

#### S4.2 Descriptive reporting among females aged [15-79] in Overseas Territories

The number of tests for people living in Overseas territories was `formating_text(sum(df_dom_tom$N),0)`, among which the share of tests collected through opportunistic screening was 99.0%. A complete reporting of the descriptive statistics are provided in the Supplementary Table S4.

**Table S4 Number of tests and observed prevalence, stratified by genotype group, type of screening, year, age, and district.**

| | | | | | Positive for... | | | | | | | | | |
|---|---|---|---|---|---|---|---|---|---|---|---|---|---|---|
| Variables | | | | | HPV16 | | HPV18 | | HPV16/18 | | HPV-other | | At least 1 HR HPV | |
| Screening | Year | Age | District | Number of tests (N) | N | % | N | % | N | % | N | % | N | % |
| Opportunistic | | | | 2,988 | 67 | 2.2 | 50 | 1.7 | 116 | 3.9 | 411 | 13.8 | 500 | 16.7 |
| Organised | | | | 29 | 1 | 3.4 | 1 | 3.4 | 2 | 6.9 | 2 | 6.9 | 4 | 13.8 |
| | 2020 | | | 1,296 | 26 | 2.0 | 25 | 1.9 | 51 | 3.9 | 182 | 14.0 | 222 | 17.1 |
| | 2021 | | | 1,654 | 37 | 2.2 | 25 | 1.5 | 61 | 3.7 | 218 | 13.2 | 265 | 16.0 |
| | 2022 | | | 35 | 3 | 8.6 | 0 | 0.0 | 3 | 8.6 | 9 | 25.7 | 10 | 28.6 |
| | 2023 | | | 32 | 2 | 6.2 | 1 | 3.1 | 3 | 9.4 | 4 | 12.5 | 7 | 21.9 |
| | | [24,25] | | 1 | 0 | 0.0 | 0 | 0.0 | 0 | 0.0 | 0 | 0.0 | 0 | 0.0 |
| | | (25,29] | | 3 | 1 | 33.3 | 0 | 0.0 | 1 | 33.3 | 2 | 66.7 | 2 | 66.7 |
| | | (29,39] | | 775 | 29 | 3.7 | 13 | 1.7 | 41 | 5.3 | 152 | 19.6 | 182 | 23.5 |
| | | (39,49] | | 1,043 | 17 | 1.6 | 18 | 1.7 | 35 | 3.4 | 123 | 11.8 | 154 | 14.8 |
| | | (49,59] | | 818 | 15 | 1.8 | 14 | 1.7 | 29 | 3.5 | 94 | 11.5 | 116 | 14.2 |
| | | (59,66] | | 339 | 6 | 1.8 | 3 | 0.9 | 9 | 2.7 | 36 | 10.6 | 43 | 12.7 |
| | | (66,76] | | 38 | 0 | 0.0 | 3 | 7.9 | 3 | 7.9 | 6 | 15.8 | 7 | 18.4 |
| | | | Guadeloupe | 1,775 | 41 | 2.3 | 31 | 1.7 | 71 | 4.0 | 241 | 13.6 | 298 | 16.8 |
| | | | Guyane | 26 | 0 | 0.0 | 0 | 0.0 | 0 | 0.0 | 2 | 7.7 | 2 | 7.7 |
| | | | La Réunion | 32 | 1 | 3.1 | 0 | 0.0 | 1 | 3.1 | 7 | 21.9 | 8 | 25.0 |
| | | | Martinique | 1,162 | 26 | 2.2 | 19 | 1.6 | 45 | 3.9 | 156 | 13.4 | 188 | 16.2 |
| | | | Mayotte | 21 | 0 | 0.0 | 1 | 4.8 | 1 | 4.8 | 6 | 28.6 | 7 | 33.3 |
| | | | | 1 | 0 | 0.0 | 0 | 0.0 | 0 | 0.0 | 1 | 100.0 | 1 | 100.0 |
| Opportunistic | 2020 | | | 1,295 | 26 | 2.0 | 25 | 1.9 | 51 | 3.9 | 182 | 14.1 | 222 | 17.1 |
| Opportunistic | 2021 | | | 1,630 | 37 | 2.3 | 25 | 1.5 | 61 | 3.7 | 216 | 13.3 | 263 | 16.1 |
| Opportunistic | 2022 | | | 34 | 3 | 8.8 | 0 | 0.0 | 3 | 8.8 | 9 | 26.5 | 10 | 29.4 |
| Opportunistic | 2023 | | | 29 | 1 | 3.4 | 0 | 0.0 | 1 | 3.4 | 4 | 13.8 | 5 | 17.2 |

**Table S4 Number of tests and observed prevalence, stratified by genotype group, type of screening, year, age, and district.** ***(continued)***

| Screening | Year | Age | District | Number of tests (N) | N | % | N | % | N | % | N | % | N | % |
|---|---|---|---|---|---|---|---|---|---|---|---|---|---|---|
| Organised | 2020 | | | 1 | 0 | 0.0 | 0 | 0.0 | 0 | 0.0 | 0 | 0.0 | 0 | 0.0 |
| Organised | 2021 | | | 24 | 0 | 0.0 | 0 | 0.0 | 0 | 0.0 | 2 | 8.3 | 2 | 8.3 |
| Organised | 2022 | | | 1 | 0 | 0.0 | 0 | 0.0 | 0 | 0.0 | 0 | 0.0 | 0 | 0.0 |
| Organised | 2023 | | | 3 | 1 | 33.3 | 1 | 33.3 | 2 | 66.7 | 0 | 0.0 | 2 | 66.7 |
| Opportunistic | | (25,29] | | 2 | 1 | 50.0 | 0 | 0.0 | 1 | 50.0 | 2 | 100.0 | 2 | 100.0 |
| Opportunistic | | (29,39] | | 773 | 29 | 3.8 | 13 | 1.7 | 41 | 5.3 | 152 | 19.7 | 182 | 23.5 |
| Opportunistic | | (39,49] | | 1,041 | 17 | 1.6 | 17 | 1.6 | 34 | 3.3 | 122 | 11.7 | 152 | 14.6 |
| Opportunistic | | (49,59] | | 807 | 15 | 1.9 | 14 | 1.7 | 29 | 3.6 | 94 | 11.6 | 116 | 14.4 |
| Opportunistic | | (59,66] | | 327 | 5 | 1.5 | 3 | 0.9 | 8 | 2.4 | 35 | 10.7 | 41 | 12.5 |
| Opportunistic | | (66,76] | | 38 | 0 | 0.0 | 3 | 7.9 | 3 | 7.9 | 6 | 15.8 | 7 | 18.4 |
| Organised | | [24,25] | | 1 | 0 | 0.0 | 0 | 0.0 | 0 | 0.0 | 0 | 0.0 | 0 | 0.0 |
| Organised | | (25,29] | | 1 | 0 | 0.0 | 0 | 0.0 | 0 | 0.0 | 0 | 0.0 | 0 | 0.0 |
| Organised | | (29,39] | | 2 | 0 | 0.0 | 0 | 0.0 | 0 | 0.0 | 0 | 0.0 | 0 | 0.0 |
| Organised | | (39,49] | | 2 | 0 | 0.0 | 1 | 50.0 | 1 | 50.0 | 1 | 50.0 | 2 | 100.0 |
| Organised | | (49,59] | | 11 | 0 | 0.0 | 0 | 0.0 | 0 | 0.0 | 0 | 0.0 | 0 | 0.0 |
| Organised | | (59,66] | | 12 | 1 | 8.3 | 0 | 0.0 | 1 | 8.3 | 1 | 8.3 | 2 | 16.7 |
| Opportunistic | | | Guadeloupe | 1,773 | 40 | 2.3 | 31 | 1.7 | 70 | 3.9 | 241 | 13.6 | 297 | 16.8 |
| Opportunistic | | | Guyane | 25 | 0 | 0.0 | 0 | 0.0 | 0 | 0.0 | 2 | 8.0 | 2 | 8.0 |
| Opportunistic | | | La Réunion | 31 | 1 | 3.2 | 0 | 0.0 | 1 | 3.2 | 7 | 22.6 | 8 | 25.8 |
| Opportunistic | | | Martinique | 1,139 | 26 | 2.3 | 19 | 1.7 | 45 | 4.0 | 155 | 13.6 | 187 | 16.4 |
| Opportunistic | | | Mayotte | 19 | 0 | 0.0 | 0 | 0.0 | 0 | 0.0 | 5 | 26.3 | 5 | 26.3 |
| Opportunistic | | | | 1 | 0 | 0.0 | 0 | 0.0 | 0 | 0.0 | 1 | 100.0 | 1 | 100.0 |
| Organised | | | Guadeloupe | 2 | 1 | 50.0 | 0 | 0.0 | 1 | 50.0 | 0 | 0.0 | 1 | 50.0 |
| Organised | | | Guyane | 1 | 0 | 0.0 | 0 | 0.0 | 0 | 0.0 | 0 | 0.0 | 0 | 0.0 |
| Organised | | | La Réunion | 1 | 0 | 0.0 | 0 | 0.0 | 0 | 0.0 | 0 | 0.0 | 0 | 0.0 |
| Organised | | | Martinique | 23 | 0 | 0.0 | 0 | 0.0 | 0 | 0.0 | 1 | 4.3 | 1 | 4.3 |
| Organised | | | Mayotte | 2 | 0 | 0.0 | 1 | 50.0 | 1 | 50.0 | 1 | 50.0 | 2 | 100.0 |
| | 2020 | (25,29] | | 2 | 1 | 50.0 | 0 | 0.0 | 1 | 50.0 | 2 | 100.0 | 2 | 100.0 |
| | 2020 | (29,39] | | 346 | 10 | 2.9 | 6 | 1.7 | 16 | 4.6 | 63 | 18.2 | 75 | 21.7 |
| | 2020 | (39,49] | | 447 | 5 | 1.1 | 9 | 2.0 | 14 | 3.1 | 58 | 13.0 | 71 | 15.9 |
| | 2020 | (49,59] | | 385 | 10 | 2.6 | 8 | 2.1 | 18 | 4.7 | 51 | 13.2 | 64 | 16.6 |

**Table S4** **Number of tests and observed prevalence, stratified by genotype group, type of screening, year, age, and district.** ***(continued)***

| Screening | Year | Age | District | Number of tests (N) | N | % | N | % | N | % | N | % | N | % |
|---|---|---|---|---|---|---|---|---|---|---|---|---|---|---|
| | 2020 | (59,66] | | 112 | 0 | 0.0 | 2 | 1.8 | 2 | 1.8 | 7 | 6.2 | 9 | 8.0 |
| | 2020 | (66,76] | | 4 | 0 | 0.0 | 0 | 0.0 | 0 | 0.0 | 1 | 25.0 | 1 | 25.0 |
| | 2021 | [24,25] | | 1 | 0 | 0.0 | 0 | 0.0 | 0 | 0.0 | 0 | 0.0 | 0 | 0.0 |
| | 2021 | (25,29] | | 1 | 0 | 0.0 | 0 | 0.0 | 0 | 0.0 | 0 | 0.0 | 0 | 0.0 |
| | 2021 | (29,39] | | 408 | 17 | 4.2 | 7 | 1.7 | 23 | 5.6 | 83 | 20.3 | 100 | 24.5 |
| | 2021 | (39,49] | | 582 | 11 | 1.9 | 8 | 1.4 | 19 | 3.3 | 63 | 10.8 | 79 | 13.6 |
| | 2021 | (49,59] | | 418 | 4 | 1.0 | 6 | 1.4 | 10 | 2.4 | 39 | 9.3 | 48 | 11.5 |
| | 2021 | (59,66] | | 212 | 5 | 2.4 | 1 | 0.5 | 6 | 2.8 | 28 | 13.2 | 32 | 15.1 |
| | 2021 | (66,76] | | 32 | 0 | 0.0 | 3 | 9.4 | 3 | 9.4 | 5 | 15.6 | 6 | 18.8 |
| | 2022 | (29,39] | | 15 | 2 | 13.3 | 0 | 0.0 | 2 | 13.3 | 5 | 33.3 | 6 | 40.0 |
| | 2022 | (39,49] | | 6 | 0 | 0.0 | 0 | 0.0 | 0 | 0.0 | 2 | 33.3 | 2 | 33.3 |
| | 2022 | (49,59] | | 6 | 1 | 16.7 | 0 | 0.0 | 1 | 16.7 | 2 | 33.3 | 2 | 33.3 |
| | 2022 | (59,66] | | 6 | 0 | 0.0 | 0 | 0.0 | 0 | 0.0 | 0 | 0.0 | 0 | 0.0 |
| | 2022 | (66,76] | | 2 | 0 | 0.0 | 0 | 0.0 | 0 | 0.0 | 0 | 0.0 | 0 | 0.0 |
| | 2023 | (29,39] | | 6 | 0 | 0.0 | 0 | 0.0 | 0 | 0.0 | 1 | 16.7 | 1 | 16.7 |
| | 2023 | (39,49] | | 8 | 1 | 12.5 | 1 | 12.5 | 2 | 25.0 | 0 | 0.0 | 2 | 25.0 |
| | 2023 | (49,59] | | 9 | 0 | 0.0 | 0 | 0.0 | 0 | 0.0 | 2 | 22.2 | 2 | 22.2 |
| | 2023 | (59,66] | | 9 | 1 | 11.1 | 0 | 0.0 | 1 | 11.1 | 1 | 11.1 | 2 | 22.2 |
| | 2020 | | Guadeloupe | 759 | 15 | 2.0 | 15 | 2.0 | 30 | 4.0 | 108 | 14.2 | 133 | 17.5 |
| | 2020 | | Guyane | 7 | 0 | 0.0 | 0 | 0.0 | 0 | 0.0 | 1 | 14.3 | 1 | 14.3 |
| | 2020 | | La Réunion | 11 | 0 | 0.0 | 0 | 0.0 | 0 | 0.0 | 4 | 36.4 | 4 | 36.4 |
| | 2020 | | Martinique | 515 | 11 | 2.1 | 10 | 1.9 | 21 | 4.1 | 69 | 13.4 | 84 | 16.3 |
| | 2020 | | Mayotte | 4 | 0 | 0.0 | 0 | 0.0 | 0 | 0.0 | 0 | 0.0 | 0 | 0.0 |
| | 2021 | | Guadeloupe | 988 | 23 | 2.3 | 16 | 1.6 | 38 | 3.8 | 127 | 12.9 | 157 | 15.9 |
| | 2021 | | Guyane | 12 | 0 | 0.0 | 0 | 0.0 | 0 | 0.0 | 0 | 0.0 | 0 | 0.0 |
| | 2021 | | La Réunion | 13 | 1 | 7.7 | 0 | 0.0 | 1 | 7.7 | 3 | 23.1 | 4 | 30.8 |
| | 2021 | | Martinique | 627 | 13 | 2.1 | 9 | 1.4 | 22 | 3.5 | 83 | 13.2 | 99 | 15.8 |
| | 2021 | | Mayotte | 13 | 0 | 0.0 | 0 | 0.0 | 0 | 0.0 | 4 | 30.8 | 4 | 30.8 |
| | 2021 | | | 1 | 0 | 0.0 | 0 | 0.0 | 0 | 0.0 | 1 | 100.0 | 1 | 100.0 |
| | 2022 | | Guadeloupe | 17 | 2 | 11.8 | 0 | 0.0 | 2 | 11.8 | 4 | 23.5 | 5 | 29.4 |
| | 2022 | | Guyane | 3 | 0 | 0.0 | 0 | 0.0 | 0 | 0.0 | 1 | 33.3 | 1 | 33.3 |

**Table S4 Number of tests and observed prevalence, stratified by genotype group, type of screening, year, age, and district.** *(continued)*

| Screening | Year | Age | District | Number of tests (N) | N | % | N | % | N | % | N | % | N | % |
|---|---|---|---|---|---|---|---|---|---|---|---|---|---|---|
| | 2022 | | La Réunion | 5 | 0 | 0.0 | 0 | 0.0 | 0 | 0.0 | 0 | 0.0 | 0 | 0.0 |
| | 2022 | | Martinique | 8 | 1 | 12.5 | 0 | 0.0 | 1 | 12.5 | 2 | 25.0 | 2 | 25.0 |
| | 2022 | | Mayotte | 2 | 0 | 0.0 | 0 | 0.0 | 0 | 0.0 | 2 | 100.0 | 2 | 100.0 |
| | 2023 | | Guadeloupe | 11 | 1 | 9.1 | 0 | 0.0 | 1 | 9.1 | 2 | 18.2 | 3 | 27.3 |
| | 2023 | | Guyane | 4 | 0 | 0.0 | 0 | 0.0 | 0 | 0.0 | 0 | 0.0 | 0 | 0.0 |
| | 2023 | | La Réunion | 3 | 0 | 0.0 | 0 | 0.0 | 0 | 0.0 | 0 | 0.0 | 0 | 0.0 |
| | 2023 | | Martinique | 12 | 1 | 8.3 | 0 | 0.0 | 1 | 8.3 | 2 | 16.7 | 3 | 25.0 |
| | 2023 | | Mayotte | 2 | 0 | 0.0 | 1 | 50.0 | 1 | 50.0 | 0 | 0.0 | 1 | 50.0 |
| | | [24,25] | Martinique | 1 | 0 | 0.0 | 0 | 0.0 | 0 | 0.0 | 0 | 0.0 | 0 | 0.0 |
| | | (25,29] | Guadeloupe | 1 | 1 | 100.0 | 0 | 0.0 | 1 | 100.0 | 1 | 100.0 | 1 | 100.0 |
| | | (25,29] | Martinique | 2 | 0 | 0.0 | 0 | 0.0 | 0 | 0.0 | 1 | 50.0 | 1 | 50.0 |
| | | (29,39] | Guadeloupe | 448 | 18 | 4.0 | 6 | 1.3 | 23 | 5.1 | 88 | 19.6 | 106 | 23.7 |
| | | (29,39] | Guyane | 8 | 0 | 0.0 | 0 | 0.0 | 0 | 0.0 | 0 | 0.0 | 0 | 0.0 |
| | | (29,39] | La Réunion | 13 | 0 | 0.0 | 0 | 0.0 | 0 | 0.0 | 4 | 30.8 | 4 | 30.8 |
| | | (29,39] | Martinique | 299 | 11 | 3.7 | 7 | 2.3 | 18 | 6.0 | 57 | 19.1 | 69 | 23.1 |
| | | (29,39] | Mayotte | 7 | 0 | 0.0 | 0 | 0.0 | 0 | 0.0 | 3 | 42.9 | 3 | 42.9 |
| | | (39,49] | Guadeloupe | 658 | 10 | 1.5 | 13 | 2.0 | 23 | 3.5 | 72 | 10.9 | 93 | 14.1 |
| | | (39,49] | Guyane | 8 | 0 | 0.0 | 0 | 0.0 | 0 | 0.0 | 0 | 0.0 | 0 | 0.0 |
| | | (39,49] | La Réunion | 12 | 1 | 8.3 | 0 | 0.0 | 1 | 8.3 | 3 | 25.0 | 4 | 33.3 |
| | | (39,49] | Martinique | 353 | 6 | 1.7 | 4 | 1.1 | 10 | 2.8 | 44 | 12.5 | 52 | 14.7 |
| | | (39,49] | Mayotte | 11 | 0 | 0.0 | 1 | 9.1 | 1 | 9.1 | 3 | 27.3 | 4 | 36.4 |
| | | (39,49] | | 1 | 0 | 0.0 | 0 | 0.0 | 0 | 0.0 | 1 | 100.0 | 1 | 100.0 |
| | | (49,59] | Guadeloupe | 457 | 8 | 1.8 | 8 | 1.8 | 16 | 3.5 | 60 | 13.1 | 72 | 15.8 |
| | | (49,59] | Guyane | 6 | 0 | 0.0 | 0 | 0.0 | 0 | 0.0 | 2 | 33.3 | 2 | 33.3 |
| | | (49,59] | La Réunion | 4 | 0 | 0.0 | 0 | 0.0 | 0 | 0.0 | 0 | 0.0 | 0 | 0.0 |
| | | (49,59] | Martinique | 349 | 7 | 2.0 | 6 | 1.7 | 13 | 3.7 | 32 | 9.2 | 42 | 12.0 |
| | | (49,59] | Mayotte | 2 | 0 | 0.0 | 0 | 0.0 | 0 | 0.0 | 0 | 0.0 | 0 | 0.0 |
| | | (59,66] | Guadeloupe | 190 | 4 | 2.1 | 2 | 1.1 | 6 | 3.2 | 18 | 9.5 | 23 | 12.1 |
| | | (59,66] | Guyane | 4 | 0 | 0.0 | 0 | 0.0 | 0 | 0.0 | 0 | 0.0 | 0 | 0.0 |
| | | (59,66] | La Réunion | 3 | 0 | 0.0 | 0 | 0.0 | 0 | 0.0 | 0 | 0.0 | 0 | 0.0 |
| | | (59,66] | Martinique | 141 | 2 | 1.4 | 1 | 0.7 | 3 | 2.1 | 18 | 12.8 | 20 | 14.2 |

**Table S4** **Number of tests and observed prevalence, stratified by genotype group, type of screening, year, age, and district.** ***(continued)***

| Screening | Year | Age | District | Number of tests (N) | N | % | N | % | N | % | N | % | N | % |
|---|---|---|---|---|---|---|---|---|---|---|---|---|---|---|
| | | (59,66] | Mayotte | 1 | 0 | 0.0 | 0 | 0.0 | 0 | 0.0 | 0 | 0.0 | 0 | 0.0 |
| | | (66,76] | Guadeloupe | 21 | 0 | 0.0 | 2 | 9.5 | 2 | 9.5 | 2 | 9.5 | 3 | 14.3 |
| | | (66,76] | Martinique | 17 | 0 | 0.0 | 1 | 5.9 | 1 | 5.9 | 4 | 23.5 | 4 | 23.5 |
| Opportunistic | 2020 | (25,29] | | 2 | 1 | 50.0 | 0 | 0.0 | 1 | 50.0 | 2 | 100.0 | 2 | 100.0 |
| Opportunistic | 2020 | (29,39] | | 346 | 10 | 2.9 | 6 | 1.7 | 16 | 4.6 | 63 | 18.2 | 75 | 21.7 |
| Opportunistic | 2020 | (39,49] | | 447 | 5 | 1.1 | 9 | 2.0 | 14 | 3.1 | 58 | 13.0 | 71 | 15.9 |
| Opportunistic | 2020 | (49,59] | | 385 | 10 | 2.6 | 8 | 2.1 | 18 | 4.7 | 51 | 13.2 | 64 | 16.6 |
| Opportunistic | 2020 | (59,66] | | 111 | 0 | 0.0 | 2 | 1.8 | 2 | 1.8 | 7 | 6.3 | 9 | 8.1 |
| Opportunistic | 2020 | (66,76] | | 4 | 0 | 0.0 | 0 | 0.0 | 0 | 0.0 | 1 | 25.0 | 1 | 25.0 |
| Opportunistic | 2021 | (29,39] | | 407 | 17 | 4.2 | 7 | 1.7 | 23 | 5.7 | 83 | 20.4 | 100 | 24.6 |
| Opportunistic | 2021 | (39,49] | | 581 | 11 | 1.9 | 8 | 1.4 | 19 | 3.3 | 62 | 10.7 | 78 | 13.4 |
| Opportunistic | 2021 | (49,59] | | 408 | 4 | 1.0 | 6 | 1.5 | 10 | 2.5 | 39 | 9.6 | 48 | 11.8 |
| Opportunistic | 2021 | (59,66] | | 202 | 5 | 2.5 | 1 | 0.5 | 6 | 3.0 | 27 | 13.4 | 31 | 15.3 |
| Opportunistic | 2021 | (66,76] | | 32 | 0 | 0.0 | 3 | 9.4 | 3 | 9.4 | 5 | 15.6 | 6 | 18.8 |
| Opportunistic | 2022 | (29,39] | | 14 | 2 | 14.3 | 0 | 0.0 | 2 | 14.3 | 5 | 35.7 | 6 | 42.9 |
| Opportunistic | 2022 | (39,49] | | 6 | 0 | 0.0 | 0 | 0.0 | 0 | 0.0 | 2 | 33.3 | 2 | 33.3 |
| Opportunistic | 2022 | (49,59] | | 6 | 1 | 16.7 | 0 | 0.0 | 1 | 16.7 | 2 | 33.3 | 2 | 33.3 |
| Opportunistic | 2022 | (59,66] | | 6 | 0 | 0.0 | 0 | 0.0 | 0 | 0.0 | 0 | 0.0 | 0 | 0.0 |
| Opportunistic | 2022 | (66,76] | | 2 | 0 | 0.0 | 0 | 0.0 | 0 | 0.0 | 0 | 0.0 | 0 | 0.0 |
| Opportunistic | 2023 | (29,39] | | 6 | 0 | 0.0 | 0 | 0.0 | 0 | 0.0 | 1 | 16.7 | 1 | 16.7 |
| Opportunistic | 2023 | (39,49] | | 7 | 1 | 14.3 | 0 | 0.0 | 1 | 14.3 | 0 | 0.0 | 1 | 14.3 |
| Opportunistic | 2023 | (49,59] | | 8 | 0 | 0.0 | 0 | 0.0 | 0 | 0.0 | 2 | 25.0 | 2 | 25.0 |
| Opportunistic | 2023 | (59,66] | | 8 | 0 | 0.0 | 0 | 0.0 | 0 | 0.0 | 1 | 12.5 | 1 | 12.5 |
| Organised | 2020 | (59,66] | | 1 | 0 | 0.0 | 0 | 0.0 | 0 | 0.0 | 0 | 0.0 | 0 | 0.0 |
| Organised | 2021 | [24,25] | | 1 | 0 | 0.0 | 0 | 0.0 | 0 | 0.0 | 0 | 0.0 | 0 | 0.0 |
| Organised | 2021 | (25,29] | | 1 | 0 | 0.0 | 0 | 0.0 | 0 | 0.0 | 0 | 0.0 | 0 | 0.0 |
| Organised | 2021 | (29,39] | | 1 | 0 | 0.0 | 0 | 0.0 | 0 | 0.0 | 0 | 0.0 | 0 | 0.0 |
| Organised | 2021 | (39,49] | | 1 | 0 | 0.0 | 0 | 0.0 | 0 | 0.0 | 1 | 100.0 | 1 | 100.0 |
| Organised | 2021 | (49,59] | | 10 | 0 | 0.0 | 0 | 0.0 | 0 | 0.0 | 0 | 0.0 | 0 | 0.0 |
| Organised | 2021 | (59,66] | | 10 | 0 | 0.0 | 0 | 0.0 | 0 | 0.0 | 1 | 10.0 | 1 | 10.0 |
| Organised | 2022 | (29,39] | | 1 | 0 | 0.0 | 0 | 0.0 | 0 | 0.0 | 0 | 0.0 | 0 | 0.0 |

**Table S4** **Number of tests and observed prevalence, stratified by genotype group, type of screening, year, age, and district.** ***(continued)***

| Screening | Year | Age | District | Number of tests (N) | N | % | N | % | N | % | N | % | N | % |
|---|---|---|---|---|---|---|---|---|---|---|---|---|---|---|
| Organised | 2023 | (39,49] | | 1 | 0 | 0.0 | 1 | 100.0 | 1 | 100.0 | 0 | 0.0 | 1 | 100.0 |
| Organised | 2023 | (49,59] | | 1 | 0 | 0.0 | 0 | 0.0 | 0 | 0.0 | 0 | 0.0 | 0 | 0.0 |
| Organised | 2023 | (59,66] | | 1 | 1 | 100.0 | 0 | 0.0 | 1 | 100.0 | 0 | 0.0 | 1 | 100.0 |
| Opportunistic | 2020 | | Guadeloupe | 759 | 15 | 2.0 | 15 | 2.0 | 30 | 4.0 | 108 | 14.2 | 133 | 17.5 |
| Opportunistic | 2020 | | Guyane | 7 | 0 | 0.0 | 0 | 0.0 | 0 | 0.0 | 1 | 14.3 | 1 | 14.3 |
| Opportunistic | 2020 | | La Réunion | 11 | 0 | 0.0 | 0 | 0.0 | 0 | 0.0 | 4 | 36.4 | 4 | 36.4 |
| Opportunistic | 2020 | | Martinique | 514 | 11 | 2.1 | 10 | 1.9 | 21 | 4.1 | 69 | 13.4 | 84 | 16.3 |
| Opportunistic | 2020 | | Mayotte | 4 | 0 | 0.0 | 0 | 0.0 | 0 | 0.0 | 0 | 0.0 | 0 | 0.0 |
| Opportunistic | 2021 | | Guadeloupe | 987 | 23 | 2.3 | 16 | 1.6 | 38 | 3.9 | 127 | 12.9 | 157 | 15.9 |
| Opportunistic | 2021 | | Guyane | 12 | 0 | 0.0 | 0 | 0.0 | 0 | 0.0 | 0 | 0.0 | 0 | 0.0 |
| Opportunistic | 2021 | | La Réunion | 13 | 1 | 7.7 | 0 | 0.0 | 1 | 7.7 | 3 | 23.1 | 4 | 30.8 |
| Opportunistic | 2021 | | Martinique | 605 | 13 | 2.1 | 9 | 1.5 | 22 | 3.6 | 82 | 13.6 | 98 | 16.2 |
| Opportunistic | 2021 | | Mayotte | 12 | 0 | 0.0 | 0 | 0.0 | 0 | 0.0 | 3 | 25.0 | 3 | 25.0 |
| Opportunistic | 2021 | | | 1 | 0 | 0.0 | 0 | 0.0 | 0 | 0.0 | 1 | 100.0 | 1 | 100.0 |
| Opportunistic | 2022 | | Guadeloupe | 17 | 2 | 11.8 | 0 | 0.0 | 2 | 11.8 | 4 | 23.5 | 5 | 29.4 |
| Opportunistic | 2022 | | Guyane | 3 | 0 | 0.0 | 0 | 0.0 | 0 | 0.0 | 1 | 33.3 | 1 | 33.3 |
| Opportunistic | 2022 | | La Réunion | 4 | 0 | 0.0 | 0 | 0.0 | 0 | 0.0 | 0 | 0.0 | 0 | 0.0 |
| Opportunistic | 2022 | | Martinique | 8 | 1 | 12.5 | 0 | 0.0 | 1 | 12.5 | 2 | 25.0 | 2 | 25.0 |
| Opportunistic | 2022 | | Mayotte | 2 | 0 | 0.0 | 0 | 0.0 | 0 | 0.0 | 2 | 100.0 | 2 | 100.0 |
| Opportunistic | 2023 | | Guadeloupe | 10 | 0 | 0.0 | 0 | 0.0 | 0 | 0.0 | 2 | 20.0 | 2 | 20.0 |
| Opportunistic | 2023 | | Guyane | 3 | 0 | 0.0 | 0 | 0.0 | 0 | 0.0 | 0 | 0.0 | 0 | 0.0 |
| Opportunistic | 2023 | | La Réunion | 3 | 0 | 0.0 | 0 | 0.0 | 0 | 0.0 | 0 | 0.0 | 0 | 0.0 |
| Opportunistic | 2023 | | Martinique | 12 | 1 | 8.3 | 0 | 0.0 | 1 | 8.3 | 2 | 16.7 | 3 | 25.0 |
| Opportunistic | 2023 | | Mayotte | 1 | 0 | 0.0 | 0 | 0.0 | 0 | 0.0 | 0 | 0.0 | 0 | 0.0 |
| Organised | 2020 | | Martinique | 1 | 0 | 0.0 | 0 | 0.0 | 0 | 0.0 | 0 | 0.0 | 0 | 0.0 |
| Organised | 2021 | | Guadeloupe | 1 | 0 | 0.0 | 0 | 0.0 | 0 | 0.0 | 0 | 0.0 | 0 | 0.0 |
| Organised | 2021 | | Martinique | 22 | 0 | 0.0 | 0 | 0.0 | 0 | 0.0 | 1 | 4.5 | 1 | 4.5 |
| Organised | 2021 | | Mayotte | 1 | 0 | 0.0 | 0 | 0.0 | 0 | 0.0 | 1 | 100.0 | 1 | 100.0 |
| Organised | 2022 | | La Réunion | 1 | 0 | 0.0 | 0 | 0.0 | 0 | 0.0 | 0 | 0.0 | 0 | 0.0 |
| Organised | 2023 | | Guadeloupe | 1 | 1 | 100.0 | 0 | 0.0 | 1 | 100.0 | 0 | 0.0 | 1 | 100.0 |
| Organised | 2023 | | Guyane | 1 | 0 | 0.0 | 0 | 0.0 | 0 | 0.0 | 0 | 0.0 | 0 | 0.0 |

**Table S4 Number of tests and observed prevalence, stratified by genotype group, type of screening, year, age, and district.** ***(continued)***

| Screening | Year | Age | District | Number of tests (N) | N | % | N | % | N | % | N | % | N | % |
|---|---|---|---|---|---|---|---|---|---|---|---|---|---|---|
| Organised | 2023 | | Mayotte | 1 | 0 | 0.0 | 1 | 100.0 | 1 | 100.0 | 0 | 0.0 | 1 | 100.0 |
| Opportunistic | | (25,29] | Guadeloupe | 1 | 1 | 100.0 | 0 | 0.0 | 1 | 100.0 | 1 | 100.0 | 1 | 100.0 |
| Opportunistic | | (25,29] | Martinique | 1 | 0 | 0.0 | 0 | 0.0 | 0 | 0.0 | 1 | 100.0 | 1 | 100.0 |
| Opportunistic | | (29,39] | Guadeloupe | 448 | 18 | 4.0 | 6 | 1.3 | 23 | 5.1 | 88 | 19.6 | 106 | 23.7 |
| Opportunistic | | (29,39] | Guyane | 8 | 0 | 0.0 | 0 | 0.0 | 0 | 0.0 | 0 | 0.0 | 0 | 0.0 |
| Opportunistic | | (29,39] | La Réunion | 12 | 0 | 0.0 | 0 | 0.0 | 0 | 0.0 | 4 | 33.3 | 4 | 33.3 |
| Opportunistic | | (29,39] | Martinique | 298 | 11 | 3.7 | 7 | 2.3 | 18 | 6.0 | 57 | 19.1 | 69 | 23.2 |
| Opportunistic | | (29,39] | Mayotte | 7 | 0 | 0.0 | 0 | 0.0 | 0 | 0.0 | 3 | 42.9 | 3 | 42.9 |
| Opportunistic | | (39,49] | Guadeloupe | 658 | 10 | 1.5 | 13 | 2.0 | 23 | 3.5 | 72 | 10.9 | 93 | 14.1 |
| Opportunistic | | (39,49] | Guyane | 8 | 0 | 0.0 | 0 | 0.0 | 0 | 0.0 | 0 | 0.0 | 0 | 0.0 |
| Opportunistic | | (39,49] | La Réunion | 12 | 1 | 8.3 | 0 | 0.0 | 1 | 8.3 | 3 | 25.0 | 4 | 33.3 |
| Opportunistic | | (39,49] | Martinique | 353 | 6 | 1.7 | 4 | 1.1 | 10 | 2.8 | 44 | 12.5 | 52 | 14.7 |
| Opportunistic | | (39,49] | Mayotte | 9 | 0 | 0.0 | 0 | 0.0 | 0 | 0.0 | 2 | 22.2 | 2 | 22.2 |
| Opportunistic | | (39,49] | | 1 | 0 | 0.0 | 0 | 0.0 | 0 | 0.0 | 1 | 100.0 | 1 | 100.0 |
| Opportunistic | | (49,59] | Guadeloupe | 457 | 8 | 1.8 | 8 | 1.8 | 16 | 3.5 | 60 | 13.1 | 72 | 15.8 |
| Opportunistic | | (49,59] | Guyane | 5 | 0 | 0.0 | 0 | 0.0 | 0 | 0.0 | 2 | 40.0 | 2 | 40.0 |
| Opportunistic | | (49,59] | La Réunion | 4 | 0 | 0.0 | 0 | 0.0 | 0 | 0.0 | 0 | 0.0 | 0 | 0.0 |
| Opportunistic | | (49,59] | Martinique | 339 | 7 | 2.1 | 6 | 1.8 | 13 | 3.8 | 32 | 9.4 | 42 | 12.4 |
| Opportunistic | | (49,59] | Mayotte | 2 | 0 | 0.0 | 0 | 0.0 | 0 | 0.0 | 0 | 0.0 | 0 | 0.0 |
| Opportunistic | | (59,66] | Guadeloupe | 188 | 3 | 1.6 | 2 | 1.1 | 5 | 2.7 | 18 | 9.6 | 22 | 11.7 |
| Opportunistic | | (59,66] | Guyane | 4 | 0 | 0.0 | 0 | 0.0 | 0 | 0.0 | 0 | 0.0 | 0 | 0.0 |
| Opportunistic | | (59,66] | La Réunion | 3 | 0 | 0.0 | 0 | 0.0 | 0 | 0.0 | 0 | 0.0 | 0 | 0.0 |
| Opportunistic | | (59,66] | Martinique | 131 | 2 | 1.5 | 1 | 0.8 | 3 | 2.3 | 17 | 13.0 | 19 | 14.5 |
| Opportunistic | | (59,66] | Mayotte | 1 | 0 | 0.0 | 0 | 0.0 | 0 | 0.0 | 0 | 0.0 | 0 | 0.0 |
| Opportunistic | | (66,76] | Guadeloupe | 21 | 0 | 0.0 | 2 | 9.5 | 2 | 9.5 | 2 | 9.5 | 3 | 14.3 |
| Opportunistic | | (66,76] | Martinique | 17 | 0 | 0.0 | 1 | 5.9 | 1 | 5.9 | 4 | 23.5 | 4 | 23.5 |
| Organised | | [24,25] | Martinique | 1 | 0 | 0.0 | 0 | 0.0 | 0 | 0.0 | 0 | 0.0 | 0 | 0.0 |
| Organised | | (25,29] | Martinique | 1 | 0 | 0.0 | 0 | 0.0 | 0 | 0.0 | 0 | 0.0 | 0 | 0.0 |
| Organised | | (29,39] | La Réunion | 1 | 0 | 0.0 | 0 | 0.0 | 0 | 0.0 | 0 | 0.0 | 0 | 0.0 |
| Organised | | (29,39] | Martinique | 1 | 0 | 0.0 | 0 | 0.0 | 0 | 0.0 | 0 | 0.0 | 0 | 0.0 |
| Organised | | (39,49] | Mayotte | 2 | 0 | 0.0 | 1 | 50.0 | 1 | 50.0 | 1 | 50.0 | 2 | 100.0 |

**Table S4 Number of tests and observed prevalence, stratified by genotype group, type of screening, year, age, and district.** *(continued)*

| Screening | Year | Age | District | Number of tests (N) | N | % | N | % | N | % | N | % | N | % |
|---|---|---|---|---|---|---|---|---|---|---|---|---|---|---|
| Organised | | (49,59] | Guyane | 1 | 0 | 0.0 | 0 | 0.0 | 0 | 0.0 | 0 | 0.0 | 0 | 0.0 |
| Organised | | (49,59] | Martinique | 10 | 0 | 0.0 | 0 | 0.0 | 0 | 0.0 | 0 | 0.0 | 0 | 0.0 |
| Organised | | (59,66] | Guadeloupe | 2 | 1 | 50.0 | 0 | 0.0 | 1 | 50.0 | 0 | 0.0 | 1 | 50.0 |
| Organised | | (59,66] | Martinique | 10 | 0 | 0.0 | 0 | 0.0 | 0 | 0.0 | 1 | 10.0 | 1 | 10.0 |
| | 2020 | (25,29] | Guadeloupe | 1 | 1 | 100.0 | 0 | 0.0 | 1 | 100.0 | 1 | 100.0 | 1 | 100.0 |
| | 2020 | (25,29] | Martinique | 1 | 0 | 0.0 | 0 | 0.0 | 0 | 0.0 | 1 | 100.0 | 1 | 100.0 |
| | 2020 | (29,39] | Guadeloupe | 210 | 6 | 2.9 | 3 | 1.4 | 9 | 4.3 | 36 | 17.1 | 43 | 20.5 |
| | 2020 | (29,39] | Guyane | 4 | 0 | 0.0 | 0 | 0.0 | 0 | 0.0 | 0 | 0.0 | 0 | 0.0 |
| | 2020 | (29,39] | La Réunion | 4 | 0 | 0.0 | 0 | 0.0 | 0 | 0.0 | 2 | 50.0 | 2 | 50.0 |
| | 2020 | (29,39] | Martinique | 127 | 4 | 3.1 | 3 | 2.4 | 7 | 5.5 | 25 | 19.7 | 30 | 23.6 |
| | 2020 | (29,39] | Mayotte | 1 | 0 | 0.0 | 0 | 0.0 | 0 | 0.0 | 0 | 0.0 | 0 | 0.0 |
| | 2020 | (39,49] | Guadeloupe | 288 | 4 | 1.4 | 7 | 2.4 | 11 | 3.8 | 39 | 13.5 | 50 | 17.4 |
| | 2020 | (39,49] | La Réunion | 5 | 0 | 0.0 | 0 | 0.0 | 0 | 0.0 | 2 | 40.0 | 2 | 40.0 |
| | 2020 | (39,49] | Martinique | 152 | 1 | 0.7 | 2 | 1.3 | 3 | 2.0 | 17 | 11.2 | 19 | 12.5 |
| | 2020 | (39,49] | Mayotte | 2 | 0 | 0.0 | 0 | 0.0 | 0 | 0.0 | 0 | 0.0 | 0 | 0.0 |
| | 2020 | (49,59] | Guadeloupe | 207 | 4 | 1.9 | 4 | 1.9 | 8 | 3.9 | 29 | 14.0 | 35 | 16.9 |
| | 2020 | (49,59] | Guyane | 2 | 0 | 0.0 | 0 | 0.0 | 0 | 0.0 | 1 | 50.0 | 1 | 50.0 |
| | 2020 | (49,59] | La Réunion | 1 | 0 | 0.0 | 0 | 0.0 | 0 | 0.0 | 0 | 0.0 | 0 | 0.0 |
| | 2020 | (49,59] | Martinique | 175 | 6 | 3.4 | 4 | 2.3 | 10 | 5.7 | 21 | 12.0 | 28 | 16.0 |
| | 2020 | (59,66] | Guadeloupe | 52 | 0 | 0.0 | 1 | 1.9 | 1 | 1.9 | 3 | 5.8 | 4 | 7.7 |
| | 2020 | (59,66] | Guyane | 1 | 0 | 0.0 | 0 | 0.0 | 0 | 0.0 | 0 | 0.0 | 0 | 0.0 |
| | 2020 | (59,66] | La Réunion | 1 | 0 | 0.0 | 0 | 0.0 | 0 | 0.0 | 0 | 0.0 | 0 | 0.0 |
| | 2020 | (59,66] | Martinique | 57 | 0 | 0.0 | 1 | 1.8 | 1 | 1.8 | 4 | 7.0 | 5 | 8.8 |
| | 2020 | (59,66] | Mayotte | 1 | 0 | 0.0 | 0 | 0.0 | 0 | 0.0 | 0 | 0.0 | 0 | 0.0 |
| | 2020 | (66,76] | Guadeloupe | 1 | 0 | 0.0 | 0 | 0.0 | 0 | 0.0 | 0 | 0.0 | 0 | 0.0 |
| | 2020 | (66,76] | Martinique | 3 | 0 | 0.0 | 0 | 0.0 | 0 | 0.0 | 1 | 33.3 | 1 | 33.3 |
| | 2021 | [24,25] | Martinique | 1 | 0 | 0.0 | 0 | 0.0 | 0 | 0.0 | 0 | 0.0 | 0 | 0.0 |
| | 2021 | (25,29] | Martinique | 1 | 0 | 0.0 | 0 | 0.0 | 0 | 0.0 | 0 | 0.0 | 0 | 0.0 |
| | 2021 | (29,39] | Guadeloupe | 231 | 11 | 4.8 | 3 | 1.3 | 13 | 5.6 | 49 | 21.2 | 59 | 25.5 |
| | 2021 | (29,39] | Guyane | 2 | 0 | 0.0 | 0 | 0.0 | 0 | 0.0 | 0 | 0.0 | 0 | 0.0 |
| | 2021 | (29,39] | La Réunion | 4 | 0 | 0.0 | 0 | 0.0 | 0 | 0.0 | 2 | 50.0 | 2 | 50.0 |

**Table S4 Number of tests and observed prevalence, stratified by genotype group, type of screening, year, age, and district.** ***(continued)***

| Screening | Year | Age | District | Number of tests (N) | N | % | N | % | N | % | N | % | N | % |
|---|---|---|---|---|---|---|---|---|---|---|---|---|---|---|
| | 2021 | (29,39] | Martinique | 166 | 6 | 3.6 | 4 | 2.4 | 10 | 6.0 | 30 | 18.1 | 37 | 22.3 |
| | 2021 | (29,39] | Mayotte | 5 | 0 | 0.0 | 0 | 0.0 | 0 | 0.0 | 2 | 40.0 | 2 | 40.0 |
| | 2021 | (39,49] | Guadeloupe | 367 | 6 | 1.6 | 6 | 1.6 | 12 | 3.3 | 33 | 9.0 | 43 | 11.7 |
| | 2021 | (39,49] | Guyane | 6 | 0 | 0.0 | 0 | 0.0 | 0 | 0.0 | 0 | 0.0 | 0 | 0.0 |
| | 2021 | (39,49] | La Réunion | 5 | 1 | 20.0 | 0 | 0.0 | 1 | 20.0 | 1 | 20.0 | 2 | 40.0 |
| | 2021 | (39,49] | Martinique | 196 | 4 | 2.0 | 2 | 1.0 | 6 | 3.1 | 26 | 13.3 | 31 | 15.8 |
| | 2021 | (39,49] | Mayotte | 7 | 0 | 0.0 | 0 | 0.0 | 0 | 0.0 | 2 | 28.6 | 2 | 28.6 |
| | 2021 | (39,49] | | 1 | 0 | 0.0 | 0 | 0.0 | 0 | 0.0 | 1 | 100.0 | 1 | 100.0 |
| | 2021 | (49,59] | Guadeloupe | 242 | 3 | 1.2 | 4 | 1.7 | 7 | 2.9 | 29 | 12.0 | 35 | 14.5 |
| | 2021 | (49,59] | Guyane | 2 | 0 | 0.0 | 0 | 0.0 | 0 | 0.0 | 0 | 0.0 | 0 | 0.0 |
| | 2021 | (49,59] | La Réunion | 3 | 0 | 0.0 | 0 | 0.0 | 0 | 0.0 | 0 | 0.0 | 0 | 0.0 |
| | 2021 | (49,59] | Martinique | 170 | 1 | 0.6 | 2 | 1.2 | 3 | 1.8 | 10 | 5.9 | 13 | 7.6 |
| | 2021 | (49,59] | Mayotte | 1 | 0 | 0.0 | 0 | 0.0 | 0 | 0.0 | 0 | 0.0 | 0 | 0.0 |
| | 2021 | (59,66] | Guadeloupe | 129 | 3 | 2.3 | 1 | 0.8 | 4 | 3.1 | 14 | 10.9 | 17 | 13.2 |
| | 2021 | (59,66] | Guyane | 2 | 0 | 0.0 | 0 | 0.0 | 0 | 0.0 | 0 | 0.0 | 0 | 0.0 |
| | 2021 | (59,66] | La Réunion | 1 | 0 | 0.0 | 0 | 0.0 | 0 | 0.0 | 0 | 0.0 | 0 | 0.0 |
| | 2021 | (59,66] | Martinique | 80 | 2 | 2.5 | 0 | 0.0 | 2 | 2.5 | 14 | 17.5 | 15 | 18.8 |
| | 2021 | (66,76] | Guadeloupe | 19 | 0 | 0.0 | 2 | 10.5 | 2 | 10.5 | 2 | 10.5 | 3 | 15.8 |
| | 2021 | (66,76] | Martinique | 13 | 0 | 0.0 | 1 | 7.7 | 1 | 7.7 | 3 | 23.1 | 3 | 23.1 |
| | 2022 | (29,39] | Guadeloupe | 6 | 1 | 16.7 | 0 | 0.0 | 1 | 16.7 | 3 | 50.0 | 4 | 66.7 |
| | 2022 | (29,39] | Guyane | 1 | 0 | 0.0 | 0 | 0.0 | 0 | 0.0 | 0 | 0.0 | 0 | 0.0 |
| | 2022 | (29,39] | La Réunion | 3 | 0 | 0.0 | 0 | 0.0 | 0 | 0.0 | 0 | 0.0 | 0 | 0.0 |
| | 2022 | (29,39] | Martinique | 4 | 1 | 25.0 | 0 | 0.0 | 1 | 25.0 | 1 | 25.0 | 1 | 25.0 |
| | 2022 | (29,39] | Mayotte | 1 | 0 | 0.0 | 0 | 0.0 | 0 | 0.0 | 1 | 100.0 | 1 | 100.0 |
| | 2022 | (39,49] | Guadeloupe | 1 | 0 | 0.0 | 0 | 0.0 | 0 | 0.0 | 0 | 0.0 | 0 | 0.0 |
| | 2022 | (39,49] | Guyane | 1 | 0 | 0.0 | 0 | 0.0 | 0 | 0.0 | 0 | 0.0 | 0 | 0.0 |
| | 2022 | (39,49] | La Réunion | 2 | 0 | 0.0 | 0 | 0.0 | 0 | 0.0 | 0 | 0.0 | 0 | 0.0 |
| | 2022 | (39,49] | Martinique | 1 | 0 | 0.0 | 0 | 0.0 | 0 | 0.0 | 1 | 100.0 | 1 | 100.0 |
| | 2022 | (39,49] | Mayotte | 1 | 0 | 0.0 | 0 | 0.0 | 0 | 0.0 | 1 | 100.0 | 1 | 100.0 |
| | 2022 | (49,59] | Guadeloupe | 5 | 1 | 20.0 | 0 | 0.0 | 1 | 20.0 | 1 | 20.0 | 1 | 20.0 |
| | 2022 | (49,59] | Guyane | 1 | 0 | 0.0 | 0 | 0.0 | 0 | 0.0 | 1 | 100.0 | 1 | 100.0 |

**Table S4 Number of tests and observed prevalence, stratified by genotype group, type of screening, year, age, and district.** ***(continued)***

| Screening | Year | Age | District | Number of tests (N) | N | % | N | % | N | % | N | % | N | % |
|---|---|---|---|---|---|---|---|---|---|---|---|---|---|---|
| | 2022 | (59,66] | Guadeloupe | 4 | 0 | 0.0 | 0 | 0.0 | 0 | 0.0 | 0 | 0.0 | 0 | 0.0 |
| | 2022 | (59,66] | Martinique | 2 | 0 | 0.0 | 0 | 0.0 | 0 | 0.0 | 0 | 0.0 | 0 | 0.0 |
| | 2022 | (66,76] | Guadeloupe | 1 | 0 | 0.0 | 0 | 0.0 | 0 | 0.0 | 0 | 0.0 | 0 | 0.0 |
| | 2022 | (66,76] | Martinique | 1 | 0 | 0.0 | 0 | 0.0 | 0 | 0.0 | 0 | 0.0 | 0 | 0.0 |
| | 2023 | (29,39] | Guadeloupe | 1 | 0 | 0.0 | 0 | 0.0 | 0 | 0.0 | 0 | 0.0 | 0 | 0.0 |
| | 2023 | (29,39] | Guyane | 1 | 0 | 0.0 | 0 | 0.0 | 0 | 0.0 | 0 | 0.0 | 0 | 0.0 |
| | 2023 | (29,39] | La Réunion | 2 | 0 | 0.0 | 0 | 0.0 | 0 | 0.0 | 0 | 0.0 | 0 | 0.0 |
| | 2023 | (29,39] | Martinique | 2 | 0 | 0.0 | 0 | 0.0 | 0 | 0.0 | 1 | 50.0 | 1 | 50.0 |
| | 2023 | (39,49] | Guadeloupe | 2 | 0 | 0.0 | 0 | 0.0 | 0 | 0.0 | 0 | 0.0 | 0 | 0.0 |
| | 2023 | (39,49] | Guyane | 1 | 0 | 0.0 | 0 | 0.0 | 0 | 0.0 | 0 | 0.0 | 0 | 0.0 |
| | 2023 | (39,49] | Martinique | 4 | 1 | 25.0 | 0 | 0.0 | 1 | 25.0 | 0 | 0.0 | 1 | 25.0 |
| | 2023 | (39,49] | Mayotte | 1 | 0 | 0.0 | 1 | 100.0 | 1 | 100.0 | 0 | 0.0 | 1 | 100.0 |
| | 2023 | (49,59] | Guadeloupe | 3 | 0 | 0.0 | 0 | 0.0 | 0 | 0.0 | 1 | 33.3 | 1 | 33.3 |
| | 2023 | (49,59] | Guyane | 1 | 0 | 0.0 | 0 | 0.0 | 0 | 0.0 | 0 | 0.0 | 0 | 0.0 |
| | 2023 | (49,59] | Martinique | 4 | 0 | 0.0 | 0 | 0.0 | 0 | 0.0 | 1 | 25.0 | 1 | 25.0 |
| | 2023 | (49,59] | Mayotte | 1 | 0 | 0.0 | 0 | 0.0 | 0 | 0.0 | 0 | 0.0 | 0 | 0.0 |
| | 2023 | (59,66] | Guadeloupe | 5 | 1 | 20.0 | 0 | 0.0 | 1 | 20.0 | 1 | 20.0 | 2 | 40.0 |
| | 2023 | (59,66] | Guyane | 1 | 0 | 0.0 | 0 | 0.0 | 0 | 0.0 | 0 | 0.0 | 0 | 0.0 |
| | 2023 | (59,66] | La Réunion | 1 | 0 | 0.0 | 0 | 0.0 | 0 | 0.0 | 0 | 0.0 | 0 | 0.0 |
| | 2023 | (59,66] | Martinique | 2 | 0 | 0.0 | 0 | 0.0 | 0 | 0.0 | 0 | 0.0 | 0 | 0.0 |
| Opportunistic | 2020 | (25,29] | Guadeloupe | 1 | 1 | 100.0 | 0 | 0.0 | 1 | 100.0 | 1 | 100.0 | 1 | 100.0 |
| Opportunistic | 2020 | (25,29] | Martinique | 1 | 0 | 0.0 | 0 | 0.0 | 0 | 0.0 | 1 | 100.0 | 1 | 100.0 |
| Opportunistic | 2020 | (29,39] | Guadeloupe | 210 | 6 | 2.9 | 3 | 1.4 | 9 | 4.3 | 36 | 17.1 | 43 | 20.5 |
| Opportunistic | 2020 | (29,39] | Guyane | 4 | 0 | 0.0 | 0 | 0.0 | 0 | 0.0 | 0 | 0.0 | 0 | 0.0 |
| Opportunistic | 2020 | (29,39] | La Réunion | 4 | 0 | 0.0 | 0 | 0.0 | 0 | 0.0 | 2 | 50.0 | 2 | 50.0 |
| Opportunistic | 2020 | (29,39] | Martinique | 127 | 4 | 3.1 | 3 | 2.4 | 7 | 5.5 | 25 | 19.7 | 30 | 23.6 |
| Opportunistic | 2020 | (29,39] | Mayotte | 1 | 0 | 0.0 | 0 | 0.0 | 0 | 0.0 | 0 | 0.0 | 0 | 0.0 |
| Opportunistic | 2020 | (39,49] | Guadeloupe | 288 | 4 | 1.4 | 7 | 2.4 | 11 | 3.8 | 39 | 13.5 | 50 | 17.4 |
| Opportunistic | 2020 | (39,49] | La Réunion | 5 | 0 | 0.0 | 0 | 0.0 | 0 | 0.0 | 2 | 40.0 | 2 | 40.0 |
| Opportunistic | 2020 | (39,49] | Martinique | 152 | 1 | 0.7 | 2 | 1.3 | 3 | 2.0 | 17 | 11.2 | 19 | 12.5 |
| Opportunistic | 2020 | (39,49] | Mayotte | 2 | 0 | 0.0 | 0 | 0.0 | 0 | 0.0 | 0 | 0.0 | 0 | 0.0 |

**Table S4** **Number of tests and observed prevalence, stratified by genotype group, type of screening, year, age, and district.** ***(continued)***

| Screening | Year | Age | District | Number of tests (N) | N | % | N | % | N | % | N | % | N | % |
|---|---|---|---|---|---|---|---|---|---|---|---|---|---|---|
| Opportunistic | 2020 | (49,59] | Guadeloupe | 207 | 4 | 1.9 | 4 | 1.9 | 8 | 3.9 | 29 | 14.0 | 35 | 16.9 |
| Opportunistic | 2020 | (49,59] | Guyane | 2 | 0 | 0.0 | 0 | 0.0 | 0 | 0.0 | 1 | 50.0 | 1 | 50.0 |
| Opportunistic | 2020 | (49,59] | La Réunion | 1 | 0 | 0.0 | 0 | 0.0 | 0 | 0.0 | 0 | 0.0 | 0 | 0.0 |
| Opportunistic | 2020 | (49,59] | Martinique | 175 | 6 | 3.4 | 4 | 2.3 | 10 | 5.7 | 21 | 12.0 | 28 | 16.0 |
| Opportunistic | 2020 | (59,66] | Guadeloupe | 52 | 0 | 0.0 | 1 | 1.9 | 1 | 1.9 | 3 | 5.8 | 4 | 7.7 |
| Opportunistic | 2020 | (59,66] | Guyane | 1 | 0 | 0.0 | 0 | 0.0 | 0 | 0.0 | 0 | 0.0 | 0 | 0.0 |
| Opportunistic | 2020 | (59,66] | La Réunion | 1 | 0 | 0.0 | 0 | 0.0 | 0 | 0.0 | 0 | 0.0 | 0 | 0.0 |
| Opportunistic | 2020 | (59,66] | Martinique | 56 | 0 | 0.0 | 1 | 1.8 | 1 | 1.8 | 4 | 7.1 | 5 | 8.9 |
| Opportunistic | 2020 | (59,66] | Mayotte | 1 | 0 | 0.0 | 0 | 0.0 | 0 | 0.0 | 0 | 0.0 | 0 | 0.0 |
| Opportunistic | 2020 | (66,76] | Guadeloupe | 1 | 0 | 0.0 | 0 | 0.0 | 0 | 0.0 | 0 | 0.0 | 0 | 0.0 |
| Opportunistic | 2020 | (66,76] | Martinique | 3 | 0 | 0.0 | 0 | 0.0 | 0 | 0.0 | 1 | 33.3 | 1 | 33.3 |
| Opportunistic | 2021 | (29,39] | Guadeloupe | 231 | 11 | 4.8 | 3 | 1.3 | 13 | 5.6 | 49 | 21.2 | 59 | 25.5 |
| Opportunistic | 2021 | (29,39] | Guyane | 2 | 0 | 0.0 | 0 | 0.0 | 0 | 0.0 | 0 | 0.0 | 0 | 0.0 |
| Opportunistic | 2021 | (29,39] | La Réunion | 4 | 0 | 0.0 | 0 | 0.0 | 0 | 0.0 | 2 | 50.0 | 2 | 50.0 |
| Opportunistic | 2021 | (29,39] | Martinique | 165 | 6 | 3.6 | 4 | 2.4 | 10 | 6.1 | 30 | 18.2 | 37 | 22.4 |
| Opportunistic | 2021 | (29,39] | Mayotte | 5 | 0 | 0.0 | 0 | 0.0 | 0 | 0.0 | 2 | 40.0 | 2 | 40.0 |
| Opportunistic | 2021 | (39,49] | Guadeloupe | 367 | 6 | 1.6 | 6 | 1.6 | 12 | 3.3 | 33 | 9.0 | 43 | 11.7 |
| Opportunistic | 2021 | (39,49] | Guyane | 6 | 0 | 0.0 | 0 | 0.0 | 0 | 0.0 | 0 | 0.0 | 0 | 0.0 |
| Opportunistic | 2021 | (39,49] | La Réunion | 5 | 1 | 20.0 | 0 | 0.0 | 1 | 20.0 | 1 | 20.0 | 2 | 40.0 |
| Opportunistic | 2021 | (39,49] | Martinique | 196 | 4 | 2.0 | 2 | 1.0 | 6 | 3.1 | 26 | 13.3 | 31 | 15.8 |
| Opportunistic | 2021 | (39,49] | Mayotte | 6 | 0 | 0.0 | 0 | 0.0 | 0 | 0.0 | 1 | 16.7 | 1 | 16.7 |
| Opportunistic | 2021 | (39,49] | | 1 | 0 | 0.0 | 0 | 0.0 | 0 | 0.0 | 1 | 100.0 | 1 | 100.0 |
| Opportunistic | 2021 | (49,59] | Guadeloupe | 242 | 3 | 1.2 | 4 | 1.7 | 7 | 2.9 | 29 | 12.0 | 35 | 14.5 |
| Opportunistic | 2021 | (49,59] | Guyane | 2 | 0 | 0.0 | 0 | 0.0 | 0 | 0.0 | 0 | 0.0 | 0 | 0.0 |
| Opportunistic | 2021 | (49,59] | La Réunion | 3 | 0 | 0.0 | 0 | 0.0 | 0 | 0.0 | 0 | 0.0 | 0 | 0.0 |
| Opportunistic | 2021 | (49,59] | Martinique | 160 | 1 | 0.6 | 2 | 1.2 | 3 | 1.9 | 10 | 6.2 | 13 | 8.1 |
| Opportunistic | 2021 | (49,59] | Mayotte | 1 | 0 | 0.0 | 0 | 0.0 | 0 | 0.0 | 0 | 0.0 | 0 | 0.0 |
| Opportunistic | 2021 | (59,66] | Guadeloupe | 128 | 3 | 2.3 | 1 | 0.8 | 4 | 3.1 | 14 | 10.9 | 17 | 13.3 |
| Opportunistic | 2021 | (59,66] | Guyane | 2 | 0 | 0.0 | 0 | 0.0 | 0 | 0.0 | 0 | 0.0 | 0 | 0.0 |
| Opportunistic | 2021 | (59,66] | La Réunion | 1 | 0 | 0.0 | 0 | 0.0 | 0 | 0.0 | 0 | 0.0 | 0 | 0.0 |
| Opportunistic | 2021 | (59,66] | Martinique | 71 | 2 | 2.8 | 0 | 0.0 | 2 | 2.8 | 13 | 18.3 | 14 | 19.7 |

**Table S4** **Number of tests and observed prevalence, stratified by genotype group, type of screening, year, age, and district.** ***(continued)***

| Screening | Year | Age | District | Number of tests (N) | N | % | N | % | N | % | N | % | N | % |
|---|---|---|---|---|---|---|---|---|---|---|---|---|---|---|
| Opportunistic | 2021 | (66,76] | Guadeloupe | 19 | 0 | 0.0 | 2 | 10.5 | 2 | 10.5 | 2 | 10.5 | 3 | 15.8 |
| Opportunistic | 2021 | (66,76] | Martinique | 13 | 0 | 0.0 | 1 | 7.7 | 1 | 7.7 | 3 | 23.1 | 3 | 23.1 |
| Opportunistic | 2022 | (29,39] | Guadeloupe | 6 | 1 | 16.7 | 0 | 0.0 | 1 | 16.7 | 3 | 50.0 | 4 | 66.7 |
| Opportunistic | 2022 | (29,39] | Guyane | 1 | 0 | 0.0 | 0 | 0.0 | 0 | 0.0 | 0 | 0.0 | 0 | 0.0 |
| Opportunistic | 2022 | (29,39] | La Réunion | 2 | 0 | 0.0 | 0 | 0.0 | 0 | 0.0 | 0 | 0.0 | 0 | 0.0 |
| Opportunistic | 2022 | (29,39] | Martinique | 4 | 1 | 25.0 | 0 | 0.0 | 1 | 25.0 | 1 | 25.0 | 1 | 25.0 |
| Opportunistic | 2022 | (29,39] | Mayotte | 1 | 0 | 0.0 | 0 | 0.0 | 0 | 0.0 | 1 | 100.0 | 1 | 100.0 |
| Opportunistic | 2022 | (39,49] | Guadeloupe | 1 | 0 | 0.0 | 0 | 0.0 | 0 | 0.0 | 0 | 0.0 | 0 | 0.0 |
| Opportunistic | 2022 | (39,49] | Guyane | 1 | 0 | 0.0 | 0 | 0.0 | 0 | 0.0 | 0 | 0.0 | 0 | 0.0 |
| Opportunistic | 2022 | (39,49] | La Réunion | 2 | 0 | 0.0 | 0 | 0.0 | 0 | 0.0 | 0 | 0.0 | 0 | 0.0 |
| Opportunistic | 2022 | (39,49] | Martinique | 1 | 0 | 0.0 | 0 | 0.0 | 0 | 0.0 | 1 | 100.0 | 1 | 100.0 |
| Opportunistic | 2022 | (39,49] | Mayotte | 1 | 0 | 0.0 | 0 | 0.0 | 0 | 0.0 | 1 | 100.0 | 1 | 100.0 |
| Opportunistic | 2022 | (49,59] | Guadeloupe | 5 | 1 | 20.0 | 0 | 0.0 | 1 | 20.0 | 1 | 20.0 | 1 | 20.0 |
| Opportunistic | 2022 | (49,59] | Guyane | 1 | 0 | 0.0 | 0 | 0.0 | 0 | 0.0 | 1 | 100.0 | 1 | 100.0 |
| Opportunistic | 2022 | (59,66] | Guadeloupe | 4 | 0 | 0.0 | 0 | 0.0 | 0 | 0.0 | 0 | 0.0 | 0 | 0.0 |
| Opportunistic | 2022 | (59,66] | Martinique | 2 | 0 | 0.0 | 0 | 0.0 | 0 | 0.0 | 0 | 0.0 | 0 | 0.0 |
| Opportunistic | 2022 | (66,76] | Guadeloupe | 1 | 0 | 0.0 | 0 | 0.0 | 0 | 0.0 | 0 | 0.0 | 0 | 0.0 |
| Opportunistic | 2022 | (66,76] | Martinique | 1 | 0 | 0.0 | 0 | 0.0 | 0 | 0.0 | 0 | 0.0 | 0 | 0.0 |
| Opportunistic | 2023 | (29,39] | Guadeloupe | 1 | 0 | 0.0 | 0 | 0.0 | 0 | 0.0 | 0 | 0.0 | 0 | 0.0 |
| Opportunistic | 2023 | (29,39] | Guyane | 1 | 0 | 0.0 | 0 | 0.0 | 0 | 0.0 | 0 | 0.0 | 0 | 0.0 |
| Opportunistic | 2023 | (29,39] | La Réunion | 2 | 0 | 0.0 | 0 | 0.0 | 0 | 0.0 | 0 | 0.0 | 0 | 0.0 |
| Opportunistic | 2023 | (29,39] | Martinique | 2 | 0 | 0.0 | 0 | 0.0 | 0 | 0.0 | 1 | 50.0 | 1 | 50.0 |
| Opportunistic | 2023 | (39,49] | Guadeloupe | 2 | 0 | 0.0 | 0 | 0.0 | 0 | 0.0 | 0 | 0.0 | 0 | 0.0 |
| Opportunistic | 2023 | (39,49] | Guyane | 1 | 0 | 0.0 | 0 | 0.0 | 0 | 0.0 | 0 | 0.0 | 0 | 0.0 |
| Opportunistic | 2023 | (39,49] | Martinique | 4 | 1 | 25.0 | 0 | 0.0 | 1 | 25.0 | 0 | 0.0 | 1 | 25.0 |
| Opportunistic | 2023 | (49,59] | Guadeloupe | 3 | 0 | 0.0 | 0 | 0.0 | 0 | 0.0 | 1 | 33.3 | 1 | 33.3 |
| Opportunistic | 2023 | (49,59] | Martinique | 4 | 0 | 0.0 | 0 | 0.0 | 0 | 0.0 | 1 | 25.0 | 1 | 25.0 |
| Opportunistic | 2023 | (49,59] | Mayotte | 1 | 0 | 0.0 | 0 | 0.0 | 0 | 0.0 | 0 | 0.0 | 0 | 0.0 |
| Opportunistic | 2023 | (59,66] | Guadeloupe | 4 | 0 | 0.0 | 0 | 0.0 | 0 | 0.0 | 1 | 25.0 | 1 | 25.0 |
| Opportunistic | 2023 | (59,66] | Guyane | 1 | 0 | 0.0 | 0 | 0.0 | 0 | 0.0 | 0 | 0.0 | 0 | 0.0 |
| Opportunistic | 2023 | (59,66] | La Réunion | 1 | 0 | 0.0 | 0 | 0.0 | 0 | 0.0 | 0 | 0.0 | 0 | 0.0 |

**Table S4 Number of tests and observed prevalence, stratified by genotype group, type of screening, year, age, and district.** ***(continued)***

| Screening | Year | Age | District | Number of tests (N) | N | % | N | % | N | % | N | % | N | % |
|---|---|---|---|---|---|---|---|---|---|---|---|---|---|---|
| Opportunistic | 2023 | (59,66] | Martinique | 2 | 0 | 0.0 | 0 | 0.0 | 0 | 0.0 | 0 | 0.0 | 0 | 0.0 |
| Organised | 2020 | (59,66] | Martinique | 1 | 0 | 0.0 | 0 | 0.0 | 0 | 0.0 | 0 | 0.0 | 0 | 0.0 |
| Organised | 2021 | [24,25] | Martinique | 1 | 0 | 0.0 | 0 | 0.0 | 0 | 0.0 | 0 | 0.0 | 0 | 0.0 |
| Organised | 2021 | (25,29] | Martinique | 1 | 0 | 0.0 | 0 | 0.0 | 0 | 0.0 | 0 | 0.0 | 0 | 0.0 |
| Organised | 2021 | (29,39] | Martinique | 1 | 0 | 0.0 | 0 | 0.0 | 0 | 0.0 | 0 | 0.0 | 0 | 0.0 |
| Organised | 2021 | (39,49] | Mayotte | 1 | 0 | 0.0 | 0 | 0.0 | 0 | 0.0 | 1 | 100.0 | 1 | 100.0 |
| Organised | 2021 | (49,59] | Martinique | 10 | 0 | 0.0 | 0 | 0.0 | 0 | 0.0 | 0 | 0.0 | 0 | 0.0 |
| Organised | 2021 | (59,66] | Guadeloupe | 1 | 0 | 0.0 | 0 | 0.0 | 0 | 0.0 | 0 | 0.0 | 0 | 0.0 |
| Organised | 2021 | (59,66] | Martinique | 9 | 0 | 0.0 | 0 | 0.0 | 0 | 0.0 | 1 | 11.1 | 1 | 11.1 |
| Organised | 2022 | (29,39] | La Réunion | 1 | 0 | 0.0 | 0 | 0.0 | 0 | 0.0 | 0 | 0.0 | 0 | 0.0 |
| Organised | 2023 | (39,49] | Mayotte | 1 | 0 | 0.0 | 1 | 100.0 | 1 | 100.0 | 0 | 0.0 | 1 | 100.0 |
| Organised | 2023 | (49,59] | Guyane | 1 | 0 | 0.0 | 0 | 0.0 | 0 | 0.0 | 0 | 0.0 | 0 | 0.0 |
| Organised | 2023 | (59,66] | Guadeloupe | 1 | 1 | 100.0 | 0 | 0.0 | 1 | 100.0 | 0 | 0.0 | 1 | 100.0 |

### S5 Additional statistical details

#### S5.1 Complete textual description of the statistical model

The statistical inference relied on a full Bayesian approach. We aggregated the number of tests and positive test results at the stratum level. Strata were defined by a grid based on the screening pathway (opportunistic or organised), age (30 to 66 years), week-year of screening, postcode, and genotype groups. Two groups of genotypes were considered: HPV16 and/or HPV18 (further denoted as 'HPV16/18') and genotypes other than HPV16/18 (further denoted 'Other genotypes'). The aggregation of the number of tests positive for HPV16/18 was motivated by the fact that HPV16 and HPV18 stand out in terms of oncogenicity compared to other genotypes [4], such that public health policies often do not distinguish between them. For example, these two genotypes are the only ones included in all the licensed vaccines available in France [5, 6].

The number of positive tests within each stratum was assigned a binomial distribution, the size parameter of which was the total number of tests performed within each stratum. The expected proportion of positive tests within a stratum was linked, through the standard logistic function (i.e., the inverse logit function), to a set of predictor using each stratum's characteristics as input.

Each stratum's linear predictor included a common intercept and additional parameters associated to the full two-ways interaction between dummy variables associated with strata related to other genotypes and opportunistic screening (minus one of those interactions, for identifiability of the intercept). Other parameters were divided into two groups. Other components were split into 2 groups. Each group of parameters contained spatially-varying, age-varying, and time-varying parameters to which we assigned multivariate hierarchical priors. The first group was common to both data collected through opportunistic and organised screening. The second group was specific to data collected through opportunistic screening and allowed to capture the potential systematic difference in expected prevalence associated with this screening pathway.

Spatially-varying parameters were specified through two-dimensional continuous functions, to which we assigned a Gaussian random field (GRF) prior, i.e., a bivariate Gaussian process (GP) [7–9] (Models 1 and 2). Alternatively, for models 3 to 6, the spatially-varying parameters were specified through discrete spatially-indexed parameters to which we assigned a scaled reparametrised Besag-York-Mollié (BYM2) prior, a Gaussian Markov random field prior (GMRF) which is the current workhorse in spatial statistics for areal data [10–13]. Age-specific and time-specific components were each specified through ordered parameters to which we assigned a Random Walk of order 2 (RW2), a GMRF also known as Bayesian P-splines, as priors [10, 14].

For models involving spatial GRFs, we assigned to each stratum the centroid of the corresponding postcode. We used the stochastic partial differential equation approach (SPDE) to build computationally efficient representations of all the GPs [8, 9]. In two dimensions, the approach relies on the use of a triangular mesh, which was built using the French coordinate reference system, Lambert-93 (EPSG:2154), with kilometer as the distance unit. We tested two different GRF: one GRF with a stationary Matérn covariance function [8] (model 1) and the non-stationary Barrier model [15] (model 2). The latter model accounted for the Mediterranean Sea and neighboring countries as a natural barrier, making Corsica a separate area from France Mainland. For this model, the fraction of the range parameter for the barrier domain was set to 0.1 of that of the land domain [15]. The final mesh is available in Supplementary Files S5.

Conversely, for the BYM2 priors, the spatial index was built from the list of French postcodes (see Supplementary Files S2). Four different neighbourhood matrices were considered: the Delaunay Triangulation structure (model 3, [16]), the Sphere of Influence (SOI) subgraph of the Delaunay Triangulation structure (model 4, [17]), and the first-order or second-order Queen contiguity structure (model 5 and 6, [18]).

All models accounted for the bivariate nature of the outcome through an exchangeable separable correlation structure between the two groups of genotypes superimposed on each age, space, and time-specific components following [19]. Riebler *et al.* [19] approach relies on the use of Kronecker product between two precision matrices, hereby assuming that the covariance structure between two dimensions are separable [20, 21]. This is a classical approach in the analysis of multidimensional data [21]. This bivariate structure allowed us to test for between-genotype differences while accounting for the fact that infection prevalences were computed from tests performed on the same participants and originated from similar contact and transmission patterns.

All hyperparameters of G(M)RF components, except those for the correlation structure, were assigned penalising-complexity (PC) priors, to mitigate the risk of overfitting inherent to such highly flexible model components [22–24] Similarly, each parameter specific to other genotypes, opportunistic screening, and the interaction between these two dimensions received a hierarchical prior with their standard deviation being assigned a PC-prior parametrised. These PC-prior allowed the posterior of these components to shrink towards 0 if not supported by the data. In addition, they allowed to consider larger tail, while ensuring regularisation, compared with a standard Gaussian distribution (see Supplementary Figure S9). PC-prior are intuitively parametrised through two parameters, specifying the probability for the parameter of interest to be greater or lower than a threshold, depending on the type of PC-prior. These parameters enable the control of the strength of the penalty applied to the deviation from the base model and hence the amount of support that should be provided by the data to support this additional component.

All assigned priors were very weak priors, meaning that high variance and correlation were not excluded *a priori*. PC priors for the marginal variances of the G(M)RF components were parametrised such that the probability for these parameters to be greater than 10 was 0.01. PC priors for the range parameters were parametrised such that the probability for these parameters to be lower than half the average distance between the two furthest points of metropolitan France on the latitude and longitude axes (i.e., 545 km), was 0.99. The relative weights between the structured and unstructured heterogeneity of BYM2 components received a PC prior parametrised such that the probability to be lower than 0.5 was 0.5. The global intercept, common to all data, received an univariate Gaussian prior with a variance equal to 10 (i.e., a precision equal to 0.1). The density for other additional intercepts was such that half the density was greater than 1. Finally, all latent parameters of the exchangeable correlation structure were parametrised using the general Fisher's z-transformation. The corresponding latent parameters were assigned univariate Gaussian distribution as priors with variance set to 5, resulting in a U-shaped prior allowing us to put sufficient probability mass to extreme values and hence not excluding *a priori* extreme correlation [19].

The six competing spatial structures were discriminated through the use of leave-one-group-out cross-validation approach, following [25, 26]. We first computed the posterior predictive density of each observed data point, conditioning on the non-observation of a group of data, further named leave-one-group-out cross validation (LGO-CV). The group of data used to compute this quantity, comprised 32 observations which were automatically selected using the posterior correlation matrix of the model [25, 26]. We further computed the logarithmic score associated with these LGO-CV, defined as minus the average logarithmic LGO-CV, and selected the model with the lowest value [26, 27].

The model was estimated using the `inlabru` package [28, 29] (development version as of 29/01/2025, test version 2.12.0.9002), a wrapper around the `R-INLA` package (development version as of 29/01/2025, test version 25.01.23, Rocky Linux-8.10 (Green Obsidian) build) [30, 31], in R version 4.4.0 [32]. All quantities were computed using 3,000 draws from the full joint posterior distribution, except for Marginal Difference in Expected Prevalence (see Supplementary subsection S5.4) for which we used only 1,550 draws due to computing requirement.

The sensitivity analyses explored the effect on the selected model of assigning to correlation parameters a prior with a variance of 1.25 and 2.5, instead of 5. Such a change had the effect of decreasing the weights assigned to extreme values of correlation [19]. We also reported the results of the non-selected models.

#### S5.2 Mathematical details of the statistical model

##### Notations and references

- Let $B^{-}$ denote the generalised inverse of the matrix $B$.
- Let $\mathcal{N}\left(\mu, \sigma^2\right)$ be the Gaussian distribution with expectation $\mu$ and variance $\sigma^2$.
- Let $\tau = \sigma^{-2}$ be the precision parameter.
- Let $\mathrm{Cov}(X, Y)$ be the covariance and $\mathrm{Corr}\,(X, Y)$ be the correlation between the two random variables $X$ and $Y$.
- Let $\mathcal{PC}_X$ denote the appropriate penalising-complexity (PC) prior for the random variable X (see [22–24]).
- The PC-prior for the precision parameter $\tau = \sigma^{-2}$ of a GMRF corresponds to a type-2 Gumbel distribution. It corresponds to an Exponential distribution for the standard deviation. Considering the Wasserstein distance within the PC framework introduced in [22, 23], instead of the Kullback-Leibler Divergence (KLD), leads to the same prior [33]. The related R-INLA webpage is: https://inla.r-inla-download.org/r-inla.org/doc/prior/pc.prec.pdf.
- Let $d(z, z')$ be the Euclidean distance for $(z, z') \in \mathbb{R}^d \times \mathbb{R}^d,\ d \geq 1$.
- A Gaussian random field $u : \mathbb{R}^d \times \mathbb{R}^d \to \mathbb{R}$ is defined via the following system:

$$\begin{cases} \mathbb{E}\left(u(z)\right) = m(z) \\ \mathrm{Cov}\left(u(z), u(z')\right) = k\left(z, z'\right) \\ \left(u(z_i)\right)_{i\in\{1,\cdots,N\}} \sim \mathcal{N}\left(\left(m(z_i)\right)_{i\in\{1,\cdots,N\}}, \left(k(z_i, z_j)\right)^2_{(i,j)\in\{1,\cdots,N\}}\right) \end{cases}$$

  for all finite location sets $(z_1, \cdots, z_n)$. With $k\left(.,.\right)$ the covariance function, which is a symmetric positive definite function and $m\left(.\right)$, the mean function (see Chapter 4, [7]).

  If $u$ is a GRF, we will be using the following notation: $u(z) \sim \mathcal{GP}\left(m\left(z\right), k\left(z, z'\right)\right)$.

- Various choice are available for the covariance kernel $k(.,.)$. In spatial statistics, the most widely used is the Matérn covariance function, which has been dominating the field for more than half a century [34, 35].
- Let $C_{\sigma,\nu,\kappa}(z, z')$ be the Matérn covariance function between any two locations $(z, z') \in \mathbb{R}^d \times \mathbb{R}^d$. In our use case, $d = 2$. The Matérn covariance function is parametrised through three parameters: the marginal variance $\sigma^2$, the scaling parameter $\kappa > 0$, and the smoothness parameter $\nu > 0$.

$$C_{\sigma,\nu,\kappa}(z, z') := \sigma^2 \frac{1}{2^{\nu-1}\Gamma(\nu)} \left(\kappa d(z, z')\right)^{\nu} K_{\nu}\left(\kappa d(z, z')\right)$$

  where $\Gamma$ is the gamma function, $K_\nu$ is the modified Bessel function of the second kind.

  Following Lindgren *et al.* [8], one may use the empirically derived definition $\rho := \frac{\sqrt{8\nu}}{\kappa}$, conferring to the range parameter $\rho$ the following natural interpretation: it is the distance at which the covariance function becomes about 0.1 [8].

  The smoothing parameter $\nu$ controls the mean-square differentiability of the underlying process. This parameter is usually poorly identifiable and Lindgren *et al.* [8] define it as $\nu := \alpha - \frac{d}{2} > 0$, with $\alpha \in ]0, 2]$ a fixed parameter. We used $\alpha = 2$, which is the usual default value.

  With such a parametrisation, the covariance function is stationary, meaning that it only depends on the relative position of the two locations, and isotropic, meaning that the covariance between two points depends only on the Euclidean distance between the locations.

Additional details are available in [8, 9].

- Lindgren *et al.* [8] presents a computationally efficient way to compute Matérn models (i.e., GRF with Matérn covariance function). The method allows going from a computational cost scaling as $\mathcal{O}\left(N^3\right)$, with $N$ the sample size, to $\mathcal{O}\left(n^{3/2}\right)$, with $n$ the number of basis functions used to perform the approximation. This approach is routinely available in R-INLA and was used to handle GRF model components. The mesh we used for carrying out this approximation had 18,648 nodes. This mesh is represented in the Supplementary Figure S8.

- The Matérn covariance function presented above is stationary and isotropic. In real application, physical barriers or holes can lead to a violation of this assumption [15]. To handle such cases, Bakka *et al.* [15] introduced a non-stationary GRF accounting for physical barriers using the efficient computation approach introduced by Lindgren *et al.* [8].

  To handle physical barriers or holes from the spatial domain of interest, the Barrier model introduces a partition of the domain into two disjoint subspaces. The first of these subspaces corresponds to the normal area (denoted $\Omega_n$) while the second part corresponds to physical barriers (denoted $\Omega_b$). Each partition receives its own Matérn covariance function. They both share the same marginal standard deviation but they each have their own range parameter.

$$\begin{cases} \forall (z,z') \in \Omega_n : C_{\sigma,\nu,\rho}(z,z') := \sigma^2 \frac{1}{2^{\nu-1}\Gamma(\nu)} \left(\frac{\sqrt{8\nu}}{\rho} d(z,z')\right)^{\nu} K_\nu\left(\frac{\sqrt{8\nu}}{\rho} d(z,z')\right) \\ \forall (z,z') \in \Omega_b : C_{\sigma,\nu,\rho_b}(z,z') := \sigma^2 \frac{1}{2^{\nu-1}\Gamma(\nu)} \left(\frac{\sqrt{8\nu}}{\rho_b} d(z,z')\right)^{\nu} K_\nu\left(\frac{\sqrt{8\nu}}{\rho_b} d(z,z')\right) \end{cases}$$

  To keep the computational cost close to that of the stationary case, the range applied to the barrier domain, $\rho_b$, is specified as a fraction of this of the normal domain, $\rho$: $\rho_b = k \times \rho$. We used the default value for $k$: $k = 0.1$.

  We considered the ocean, seas, and neighbouring countries as natural barriers.

- The PC-prior for the parameters of the Matérn covariance function is derived in [24]. Additional details can be found in the documentation of the `inla.spde2.pcmatern()` function in R-INLA [30, 31], available in https://inla.r-inla-download.org/r-inla.org/doc/prior/pc.matern.pdf. The PC-prior for the barrier model is available in [15].

- The R-INLA webpage, which describes the SPDE representation of the GP process with a stationary Matérn covariance function, is available here: https://inla.r-inla-download.org/r-inla.org/doc/prior/pc.matern.pdf

- The Barrier SPDE model [15] is implemented in the INLAspacetime package, see https://github.com/eliaskrainski/INLAspacetime

- Let $I_S$ be the identity matrix with $S$ rows and columns.

- Let $R$ be a spatially-structured matrix such that $R_{ij}$, the element of row $i$ and column $j$, is defined as $R_{ij} := \begin{cases} n_i \text{ if } i = j \\ -1 \text{ if } i \sim j \\ 0 \textit{ else} \end{cases}$, with $n_i$ the number of neighbouring areas for the spatial unit $i$ and $\sim$ denoting the adjacency (neighbouring) predicate.

- The Matérn and Barrier model considers space in its native continuous form. Because strata are associated with postcodes, we had to assign arbitrary specific coordinates (i.e. those of the centroid) for using these approaches. A more suitable approach not assigning these arbitrary coordinates is to consider that the layout of cities in space form an irregular lattice and then rely on approaches developed specifically to deal with such data, such as the BYM2 model, introduced in [11]. The BYM2 model is the current workhorse in areal spatial statistics, notably because it answers to critics directed towards previously developed priors [10–13].

Let $\alpha = (\alpha_1, \cdots, \alpha_S)$ denotes a vector of parameters such that $\forall i \in \{1, \cdots S\}$, $\alpha_i = \frac{1}{\sqrt{\tau}}\left(\sqrt{1-\phi}v_i + \phi u_i\right)$ with $\tau = \sigma^{-2}$ the marginal precision parameter, $\phi \in [0,1]$ the relative weight of the structured heterogeneity, $v \sim \mathcal{N}(0, I_S)$, and $u \sim \mathcal{N}(0, R_*^-)$. The spatial structure matrix $R_*$ denotes the scaled counterpart of the matrix $R$, meaning that the average variance (the diagonal of the generalised inverse) is equal to 1, see section 3.2. of [11].

The variance-covariance matrix of $\alpha$ is given by $\text{Var}(\alpha|\tau,\phi) = \tau^{-1}((1-\phi)I + \phi R_*^-)$.

The classical sum-to-zero identifiability constraint was used.

- The BYM2 model, like other spatially-structure GMRF, relies on a neighbourhood matrix encoding the conditional dependencies between the data.

  Two distance-based neighbourhood matrices were considered, using the centroid of each postcode. The first one of these distances was the Delaunay Triangulation approach, which is a geometric-based approach connecting areas into triangles such that the minimum angle of all triangles is maximised [16]. The second one consisted in the thinning of the Delaunay Triangulation structure through the sphere of influence approach. Starting from the Delaunay Triangulation neighbourhood structure, two areas area were linked to each other if their circles from the nearest neighbour, defined as the largest circle centred at the centroid of postcode $p$ that contains no other points than this centroid, overlap.

  A third and fourth matrices were defined based on the first-order and second-order Queen contiguity approach, respectively.

  We used the `spdep` package to define spatial neighbourhood [36–38].

- The PC-prior for the BYM2 model are provided in [11] and is implemented in the R-INLA package, see https://inla.r-inla-download.org/r-inla.org/doc/latent/bym2.pdf

- Let $\beta = (\beta_1, \cdots, \beta_n)$. Let $\beta'$ be the transpose of $\beta$. The RW2 prior has an improper (rank deficiency equal to n-2) prior density given by:

$$\pi(\beta|\tau) \propto \tau^{\frac{n-2}{2}} \exp\left(-\frac{1}{2}\beta' Q \beta\right), \; Q = \tau R_{\text{RW2}}$$

  With $R_{\text{RW2}}$ the structure matrix defined as:

$$R_{\text{RW2}} := \begin{pmatrix} 1 & -2 & 1 & 0 & 0 & \cdots & 0 & 0 \\ -2 & 5 & -4 & 1 & 0 & \cdots & 0 & 0 \\ 1 & -4 & 6 & -4 & 1 & \cdots & 0 & 0 \\ 0 & 1 & -4 & 6 & -4 & \ddots & 0 & 0 \\ \vdots & \vdots & \ddots & \ddots & \ddots & \ddots & \vdots & \vdots \\ 0 & 0 & \cdots & 1 & -4 & 6 & -4 & 1 \\ 0 & 0 & \cdots & 0 & 1 & -4 & 5 & -2 \\ 0 & 0 & \cdots & 0 & 0 & 1 & -2 & 1 \end{pmatrix}$$

  The classical sum-to-zero identifiability constraint was used to avoid confounding with the intercept.

  More details are provided in [10, 14] for further details.

- The R-INLA webpage, which describes the RW2 prior, is available here: https://inla.r-inla-download.org/r-inla.org/doc/latent/rw2.pdf
- The uniform exchangeable correlation structure through Gaussian Kronecker product Markov random field is introduced in [19].

**Fit on HPC**

Model fit, computation of GCPO, and draws from the joint posterior were performed within the same R-session for models 1 and 2 and models 3 to 6. The Barrier model (Model 2) used results from the Stationary Matérn model (model 1) as starting values for the optimisation process. Similarly, models 3, 4, and 6 used the results from model 5 (BYM2 with Queen-contiguity structure) as starting values for the optimisation process. Once all fits, GCPOs, and draws were obtained, two new R sessions were launched. The first sessions performed the post-fit analyses for non-selected model, fitted the two additional sensitivity analyses, and run the post-analysis R-function on the sensitivity analyses. The second session run the post-analysis R-function on the selected model, which took almost one week to complete because of all the quantities to compute and the large number of draws considered.

HPC logs are available on the Github webpage: https://github.com/osupplisson/hpv_prevalence/tree/main/log_hpc.

**Full joint model**

Let $s$ denote a stratum. Let $\mathcal{S}$ be the set of all strata.

A stratum $s$ is defined by a 5-tuple $s = (a_s, z_s, t_s, g_s, c_s)$ where $a_s$ denotes the age, $z_s$ the postcode, $t_s$ denotes the week-year of screening, $g_s$ the genotype, and $c_s$ denotes the type of screening.

The postcode is a five-digit number, age $a_s$ is an integer belonging to the set $\{30, \cdots, 66\}$. The week-year goes from 2020-08-17 to that of 2023-11-27, $g_s$ is either 'HPV16/18' or 'Other genotypes', and $c_s$ is either 'Organised screening' or 'Opportunistic screening'.

Depending on the model, this 5-tuple was mapped to a numeric 5-tuple used as inputs of the model components.

- For space: the postcode was mapped to the centroid (longitude,latitude) of the corresponding spatial area for GRF components and to a unique spatial index for GMRF components;
- For age: the age was mapped to an age-specific index starting at 1 for females aged 30 and ending at 36 for females aged 66;
- For time: the week-year was mapped to a time-specific index starting at 1 for the first week-year and ending at 174;
- For genotypes: 'HPV16/18' was assigned index 1 and 'Other genotypes' was assigned index 2;
- For screening pathways: 'Organised screening' was assigned index 1 and 'Opportunistic screening index 2;

Using these notations, the full hierarchical model is given by the set of specifications in the system below. For clarity, the symbol indicating conditional dependencies has been omitted from the system. In short: the observation level is conditional to the latent field and the latent field is itself conditional to the hyperparameter level. Within the observation level, $y_s$ is conditional to $(N_s, p_s)$, and $p_s$ is conditional to $\mu_s$, which is conditional to the latent field.

$$
\begin{cases}
\text{Observation level} \begin{cases}
y_s \sim \mathcal{B}\left(N_s, p_s\right) \\
\begin{cases}
p_s = \frac{1}{1+\exp(-\mu_s)} \\
\\
\mu_s = \psi_0 + \mathbb{1}_{g_s=2}\psi_1 + v_{\text{SPACE}}(z_s, g_s) + \gamma_{\text{AGE}}(a_s, g_s) + \beta_{\text{TIME}}(t_s, g_s) + \\
\mathbb{1}_{c_s=2}\left[\psi_2 + \mathbb{1}_{g_s=2}\psi_3 + v'_{\text{SPACE}}(z_s, g_s) + \gamma'_{\text{AGE}}(a_s, g_s) + \beta'_{\text{TIME}}(t_s, g_s)\right] \text{ Model 1 and 2} \\
\\
\mu_s = \psi_0 + \mathbb{1}_{g_s=2}\psi_1 + \alpha_{\text{SPACE}}(z_s, g_s) + \gamma_{\text{AGE}}(a_s, g_s) + \beta_{\text{TIME}}(t_s, g_s) + \\
\mathbb{1}_{c_s=2}\left[\psi_2 + \mathbb{1}_{g_s=2}\psi_3 + \alpha'_{\text{SPACE}}(z_s, g_s) + \gamma'_{\text{AGE}}(a_s, g_s) + \beta'_{\text{TIME}}(t_s, g_s)\right] \text{ Model 3 to 6}
\end{cases}
\end{cases} \\
\text{Parameters} \begin{cases}
\psi_0 \sim \mathcal{N}\left(0, 10\right) \\
\psi_{j\in\{1,2,3\}} \sim \mathcal{N}\left(0, \sigma^2_{\psi_{j\in\{1,2,3\}}}\right) \\
\text{Hyperparameters for } \psi_{j\in\{1,2,3\}} \begin{cases} \sigma_{\psi_{j\in\{1,2,3\}}} \sim \mathcal{PC}_\sigma\left(1, 0.5\right) \end{cases} \\
\textbf{RW2 for age and exchangeable correlation structure between genotypes} \\
\text{for } f \in \{\gamma, \gamma'\},\ (g_s, g'_s) \in \{1,2\}^2,\ (a_s, a'_s) \in \{1, \cdots, 36\}^2: \\
\text{Latent field} \begin{cases}
f_{\text{AGE}}(a_s, g_s) - 2f_{\text{AGE}}(a_s+1, g_s) + f_{\text{AGE}}(a_s+2, g_s) \sim \mathcal{N}\left(0, \sigma^2\right) \\
\text{Corr}(f_{\text{AGE}}(a_s, 1), f_{\text{AGE}}(a'_s, 2)) = \eta_{f_{\text{AGE}}}
\end{cases} \\
\text{Hyperparameters} \begin{cases}
\eta_{f_{\text{AGE}}} = \frac{\exp(\eta^*_{f_{\text{AGE}}})-1}{\exp(\eta^*_{f_{\text{AGE}}})+2-1} \\
\eta^*_{f_{\text{AGE}}} \sim \mathcal{N}\left(0, 5\right) \\
\sigma_{f_{\text{AGE}}} \sim \mathcal{PC}_\sigma\left(\lambda_{\sigma_{f_{\text{AGE}}}}\right) \\
\lambda_{\sigma_{f_{\text{AGE}}}} = -\frac{-\log\left(p_{\sigma_{f_{\text{AGE}}}}\right)}{\sigma_0},\ \Pr\left(\sigma > \sigma_0\right) = p_{\sigma_{f_{\text{AGE}}}} \\
\left(\sigma_0, p_{\sigma_{f_{\text{AGE}}}}\right) = (10, 0.01)
\end{cases} \\
\textbf{RW2 for time and exchangeable correlation structure between genotypes} \\
\text{for } f \in \{\beta, \beta'\},\ (g_s, g'_s) \in \{1,2\}^2,\ (t_s, t'_s) \in \{1, \cdots, 174\}^2: \\
\text{Latent field} \begin{cases}
f_{\text{TIME}}(t_s, g_s) - 2f_{\text{TIME}}(t_s+1, g_s) + f_{\text{TIME}}(t_s+2, g_s) \sim \mathcal{N}\left(0, \sigma^2\right) \\
\text{Corr}(f_{\text{TIME}}(t_s, 1), f_{\text{TIME}}(t'_s, 2)) = \eta_{f_{\text{TIME}}}
\end{cases} \\
\text{Hyperparameters} \begin{cases}
\eta_{f_{\text{TIME}}} = \frac{\exp(\eta^*_{f_{\text{TIME}}})-1}{\exp(\eta^*_{f_{\text{TIME}}})+2-1} \\
\eta^*_{f_{\text{TIME}}} \sim \mathcal{N}\left(0, 5\right) \\
\sigma_{f_{\text{TIME}}} \sim \mathcal{PC}_\sigma\left(\lambda_{\sigma_{f_{\text{TIME}}}}\right) \\
\lambda_{\sigma_{f_{\text{TIME}}}} = -\frac{-\log\left(p_{\sigma_{f_{\text{TIME}}}}\right)}{\sigma_0},\ \Pr\left(\sigma > \sigma_0\right) = p_{\sigma_{f_{\text{TIME}}}} \\
\left(\sigma_0, p_{\sigma_{f_{\text{TIME}}}}\right) = (10, 0.01)
\end{cases} \\
\text{...see next page}
\end{cases}
\end{cases}
$$

Parameters

*...next*

**Model 1, spatial GRF: GRF with stationary Matérn covariance for space and exchangeable correlation structure between genotypes**

for $f \in \{v, v'\}$, $(g_s, g'_s) \in \{1,2\}^2$, $\forall (z_s, z'_s) \in \Omega^2$ :

Latent field
$$\begin{cases} f_{\text{SPACE}}(z_s, g_s) \sim \mathcal{GP}\left(0, C_{\sigma_{f_{\text{SPACE}}}, \nu_{f_{\text{SPACE}}}, \rho_{f_{\text{SPACE}}}}(z_s, z'_s)\right) \\ \text{Corr}(f_{\text{SPACE}}(z_s, 1), f_{\text{SPACE}}(z_s, 2)) = \eta_{f_{\text{SPACE}}} \end{cases}$$

Hyperparameters
$$\begin{cases} \eta_{f_{\text{SPACE}}} = \frac{\exp(\eta^*_{f_{\text{SPACE}}})-1}{\exp(\eta^*_{f_{\text{SPACE}}})+2-1} \\ \eta^*_{f_{\text{SPACE}}} \sim \mathcal{N}(0,5) \\ \left(\rho_{f_{\text{SPACE}}}, \sigma_{f_{\text{SPACE}}}\right) \sim \mathcal{PC}_{(\rho,\sigma)}\left(\lambda_{\rho_{f_{\text{SPACE}}}}, \lambda_{\sigma_{f_{\text{SPACE}}}}\right) \\ \lambda_{\sigma_{f_{\text{SPACE}}}} = \arg_{\in \mathbb{R}^+} : \Pr_{\lambda_{\sigma_{f_{\text{SPACE}}}}}\left(\sigma_{f_{\text{SPACE}}} \geq \sigma_0\right) = p_{\sigma_{f_{\text{SPACE}}}} \\ \lambda_{\rho_{f_{\text{SPACE}}}} = \arg_{\in \mathbb{R}^+} : \Pr_{\lambda_{\sigma_{f_{\text{SPACE}}}}}\left(\rho_{f_{\text{SPACE}}} \leq \rho_0\right) = p_{\rho_{f_{\text{SPACE}}}} \\ \left(\sigma_0, p_{\sigma_{f_{\text{SPACE}}}}\right) = (10, 0.01) \\ \left(\rho_0, p_{\rho_{f_{\text{SPACE}}}}\right) = (545, 0.99) \\ \alpha_{f_{\text{SPACE}}} = 2 \end{cases}$$

**Model 2, spatial GRF: GRF with non-stationary Matérn covariance (Barrier model) for space and exchangeable correlation structure between genotypes**

$\Omega$ = Spatial domain, definition domain for the GRF as given by the mesh

$\Omega_b$ = Northern and Mediterranean seas, the Atlantic Ocean, the English Channel...

..., and foreign neighbouring countries (Spain, Italy...)

$\Omega_n$ = All France mainland and Corsica

$\Omega = \Omega_n \cup \Omega_b$, $\Omega_n \cap \Omega_b = \varnothing$

for $f \in \{v, v'\}$, $(g_s, g'_s) \in \{1,2\}^2$ :

Latent field
$$\begin{cases} (z_s, z'_s) \in \Omega_n^2 : f_{\text{SPACE}}(z_s, g_s) \sim \mathcal{GP}\left(0, C_{\sigma_{f_{\text{SPACE}}}, \nu_{f_{\text{SPACE}}}, \rho_{f_{\text{SPACE}}}}(z_s, z'_s)\right) \\ (z_s, z'_s) \in \Omega_b^2 : f_{\text{SPACE}}(z_s, g_s) \sim \mathcal{GP}\left(0, C_{\sigma_{f_{\text{SPACE}}}, \nu_{f_{\text{SPACE}}}, \rho_{b, f_{\text{SPACE}}}}(z_s, z'_s)\right) \\ \text{Corr}(f_{\text{SPACE}}(z_s, 1), f_{\text{SPACE}}(z_s, 2)) = \eta_{f_{\text{SPACE}}} \end{cases}$$

Hyperparameters
$$\begin{cases} \eta_{f_{\text{SPACE}}} = \frac{\exp(\eta^*_{f_{\text{SPACE}}})-1}{\exp(\eta^*_{f_{\text{SPACE}}})+2-1} \\ \eta^*_{f_{\text{SPACE}}} \sim \mathcal{N}(0,5) \\ \left(\rho_{f_{\text{SPACE}}}, \sigma_{f_{\text{SPACE}}}\right) \sim \mathcal{PC}_{(\rho,\sigma)}\left(\lambda_{\rho_{f_{\text{SPACE}}}}, \lambda_{\sigma_{f_{\text{SPACE}}}}\right) \\ \lambda_{\sigma_{f_{\text{SPACE}}}} = \arg_{\in \mathbb{R}^+} : \Pr\left(\sigma_{f_{\text{SPACE}}} \geq \sigma_0\right) = p_{\sigma_{f_{\text{SPACE}}}} \\ \lambda_{\rho_{f_{\text{SPACE}}}} = \arg_{\in \mathbb{R}^+} : \Pr\left(\rho_{f_{\text{SPACE}}} \leq \rho_0\right) = p_{\rho_{f_{\text{SPACE}}}} \\ \left(\sigma_0, p_{\sigma_{f_{\text{SPACE}}}}\right) = (10, 0.01) \\ \left(\rho_0, p_{\rho_{f_{\text{SPACE}}}}\right) = (545, 0.99) \\ \alpha_{f_{\text{SPACE}}} = 2 \end{cases}$$

...see next page

$$
\left\{ \text{Parameters} \left\{
\begin{array}{l}
\textit{...next} \\
\textbf{Models 3 to 6, spatial GMRF: GMRF with BYM2 structure for space} \\
\textbf{and exchangeable correlation structure between genotypes} \\
\text{for } f \in \{\alpha, \alpha'\},\ (g_s, g_s') \in \{1,2\}^2,\ \forall (z_s, z_s') \in \text{Set of postcodes}^2 : \\
\text{Latent field} \begin{cases} f_{SPACE}(z_s, g_s) \sim BYM2\,(\sigma, \phi, R) \\ \text{Corr}(f_{\text{SPACE}}(z_s, 1), f_{\text{SPACE}}(z_s, 2)) = \eta_{f_{\text{SPACE}}} \end{cases} \\
\text{Hyperparameters} \begin{cases}
\eta_{f_{\text{SPACE}}} = \frac{\exp(\eta^*_{f_{\text{SPACE}}})-1}{\exp(\eta^*_{f_{\text{SPACE}}})+2-1} \\
\eta^*_{f_{\text{SPACE}}} \sim \mathcal{N}\,(0, 5) \\
\phi_{f_{SPACE}} \sim \mathcal{PC}_{\phi}\left(\lambda_{f_{SPACE}}\right) \\
\lambda_{f_{SPACE}} = \arg_{\in \mathbb{R}^+} : \Pr_{\lambda_{\sigma_{f_{\phi_{\text{SPACE}}}}}}\left(\phi_{f_{\text{SPACE}}} \leq \phi_0\right) = p_{\phi_{f_{\text{SPACE}}}} \\
\sigma_{f_{SPACE}} \sim \mathcal{PC}_{\sigma}\left(\lambda_{\sigma_{f_{SPACE}}}\right) \\
\lambda_{\sigma_{f_{SPACE}}} = -\frac{-\log\left(p_{\sigma_{f_{SPACE}}}\right)}{\sigma_0},\ \Pr\left(\sigma > \sigma_0\right) = p_{\sigma_{f_{SPACE}}} \\
\left(\sigma_0, p_{\sigma_{f_{SPACE}}}\right) = (10, 0.01) \\
\left(\phi_0, p_{\phi_{f_{SPACE}}}\right) = (0.5, 0.5) \\
R : \begin{cases} \text{Delaunay Triangulation (model 3)} \\ \text{SOI subgraph of the Delaunay Triangulation (model 4)} \\ \text{1st-order Queen contiguity (model 5)} \\ \text{2nd-order Queen contiguity (model 6)} \end{cases}
\end{cases}
\end{array}
\right.\right.
$$

- We discriminated between competing models using the logarithmic score, computed using the leave-one-group-out conditional predictive ordinate (LOO-CPO), also known as leave-one-group-out cross validation (LGO-CV). Left-out groups comprised 32 observations which were automatically selected using the posterior correlation matrix of the model [26].
- Denoting $\pi$ the probability density function and $y_{-\mathcal{G}_s}$ all observed data leaving out the group of data $\mathcal{G}_s$, defined using the posterior correlation matrix between data points [26]:

$$\begin{cases} \text{LGO-CV}_s = \pi(y_s|y_{-\mathcal{G}_s}) \\ \text{log-score} = \frac{1}{\text{card}(\mathcal{S})}\left(\sum_{s\in\mathcal{S}} \text{LGO-CV}_s\right) \end{cases}$$

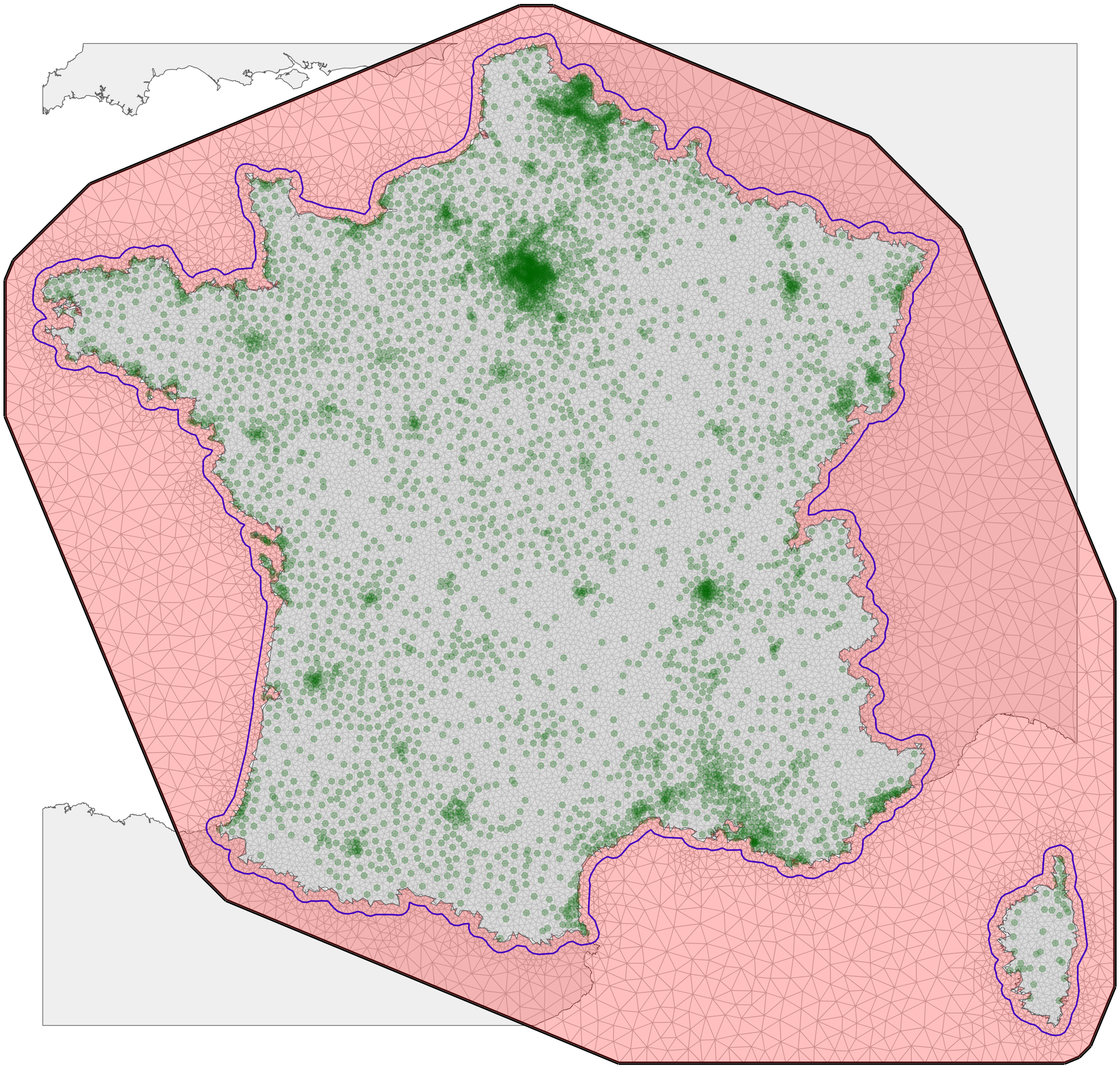

Red shaded area indicates the barrier accounted for by the Barrier model.

**Figure S8 Spatial location of all data points (green dots) and mesh (grey edges) used for the finite-elements decomposition approximation of the Gaussian random fields. The blue line delineaetes the areas in which the density of the mesh could change. It was defined to be finner within metropolitan France than outside, where no data could be observed.**

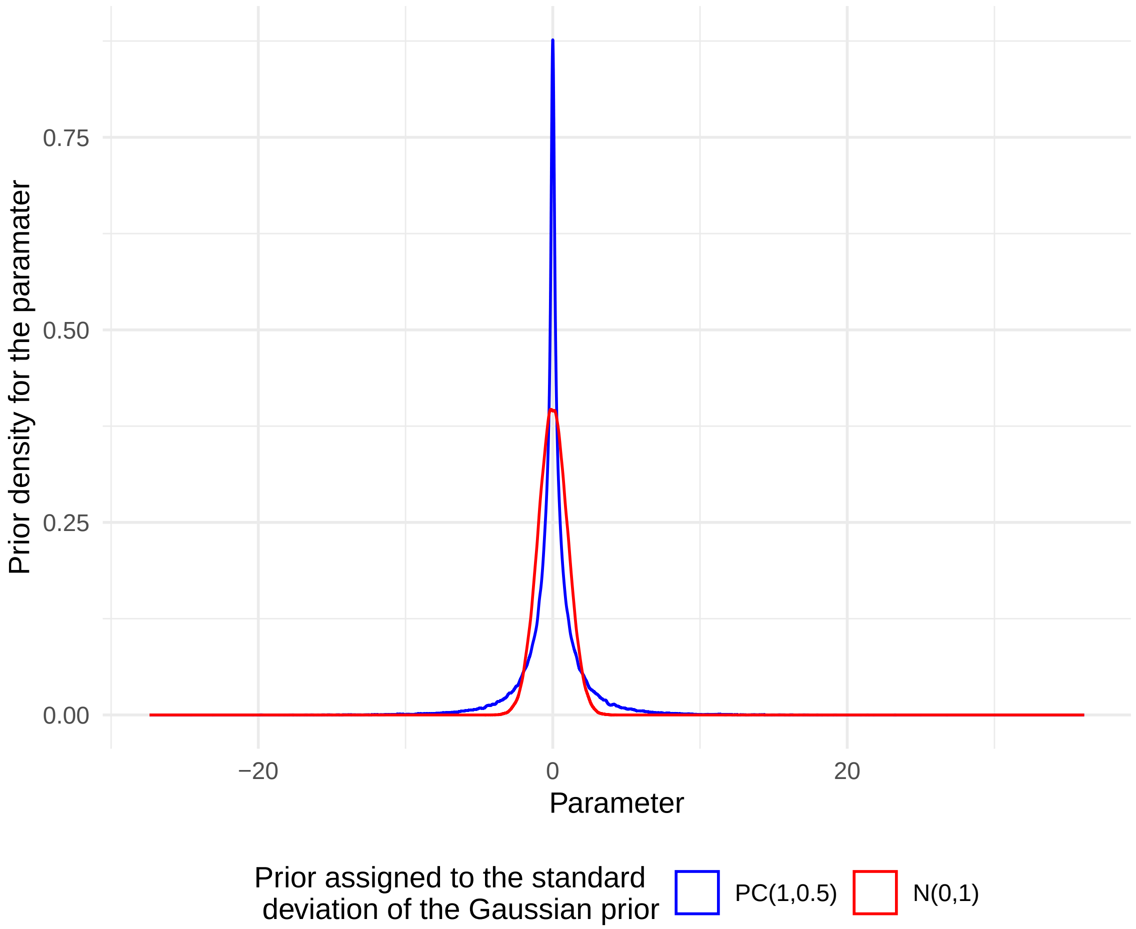

**Figure S9 Prior density for a Gaussian prior distribution with PC(1,0.5) hyperprior assigned to the precision parameter (blue curve) compared with a standard Gaussian distribution (red curve) .**

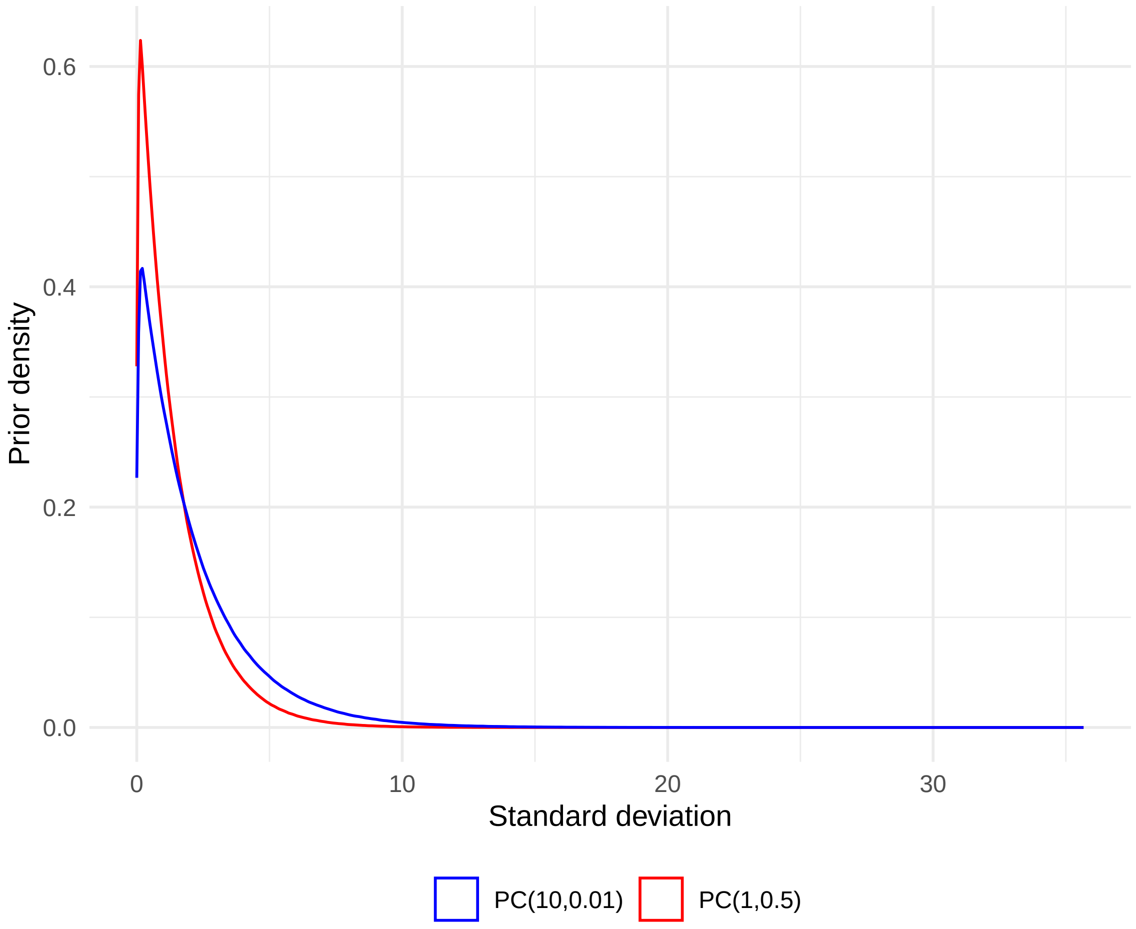

**Figure S10 PC-prior for the standard deviation of GMRFs.**

#### S5.3 Details about the sensitivity analyses

Two sensitivity analyses were explored:

1. Reporting of the non-selected competing models;
2. Change in the prior for the correlation parameters: variance of 1.25 and 2.5, instead of 5. These changes are illustrated in the Supplementary Figures S11.

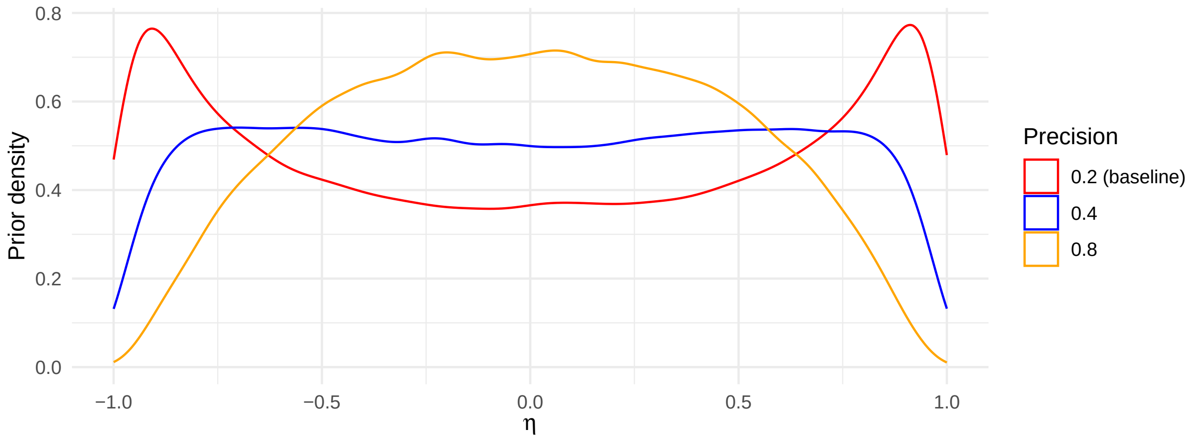

**Figure S11** **Precision for the prior distribution of the latent correlation parameters.**

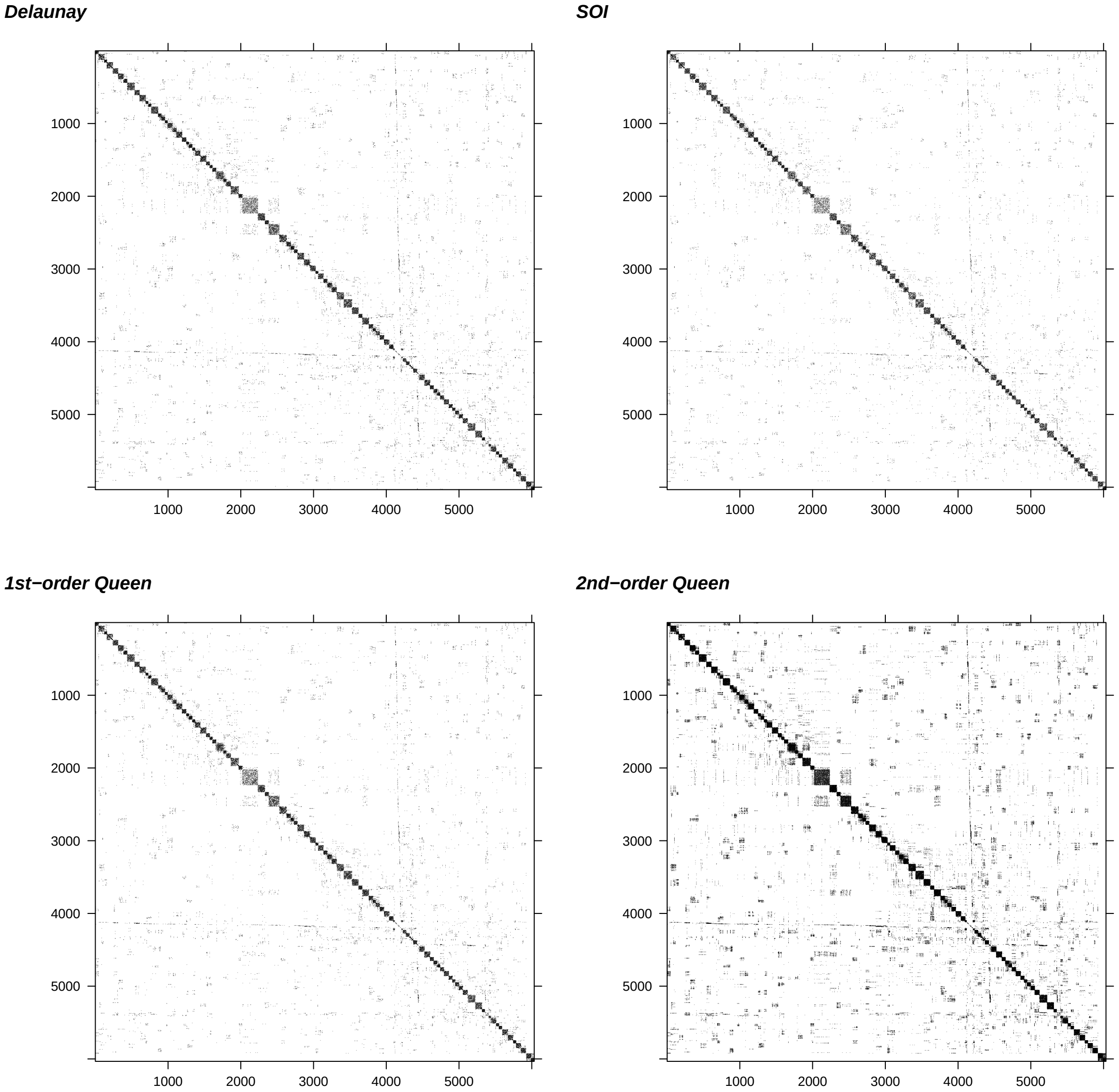

**Figure S12 Spatial neighbourhood matrices considered for BYM2 priors. Black squares indicate that the two areas are neighbours.**

#### S5.4 Details about the Marginal Difference in Expected Prevalence (MDEP)

To compute the MDEP, we considered each unique 4-tuple $(a_s, z_s, t_s, g_s)$ and computed the corresponding number of observed tests (i.e., we summed the number of tests across the two screening pathways).

For each of these 4-tuples we then made 1,550 draws from the joint posterior distribution to obtain the expected proportion of positive tests. We did so considering, first, the model specific to the organised screening pathway and, second, the model specific to the opportunistic screening pathway. We then computed the expected number of positive tests under each screening pathway. We finally took the difference between the expected number of positive tests under opportunistic and organised screening and then divided this difference by the total number of tests, yielding the Marginal Difference in Expected Prevalence.

In mathematical form:

- Step 1. Compute the observed number of tests, integrating out the screening pathway: $N_{a_s,z_s,t_s,g_s} = N_{a_s,z_s,t_s,g_s,1} + N_{a_s,z_s,t_s,g_s,2}$
- Step 2. Draws from the full joint posterior distribution $\hat{p}_{a_s,z_s,t_s,g_s,1}$ and $\hat{p}_{a_s,z_s,t_s,g_s,2}$;
- Step 3. For each draw, compute the expected number of positive test under each screening pathway: $\hat{p}_{a_s,z_s,t_s,g_s,1} \times N_{a_s,z_s,t_s,g_s}$ and $\hat{p}_{a_s,z_s,t_s,g_s,2} \times N_{a_s,z_s,t_s,g_s}$
- Step 4A. For each draw, compute the marginal prevalence integrating out various dimension. To do so, sum over all focal dimension the expected number of positive tests. Then, divide this sum by the observed number of tests. For example, for a given screening pathway $c_s$, integrating out all dimensions but the genotype one: $\frac{\sum_{a_s,z_s,t_s} \hat{p}_{a_s,z_s,t_s,g_s,c_s} \times N_{a_s,z_s,t_s,g_s}}{\sum_{a_s,z_s,t_s} N_{a_s,z_s,t_s,g_s}}$
- Step 4B. For each draw, compute the difference between the marginal expected prevalence under opportunistic and organised screening.

#### S6 Descriptive reporting for the analytic sample

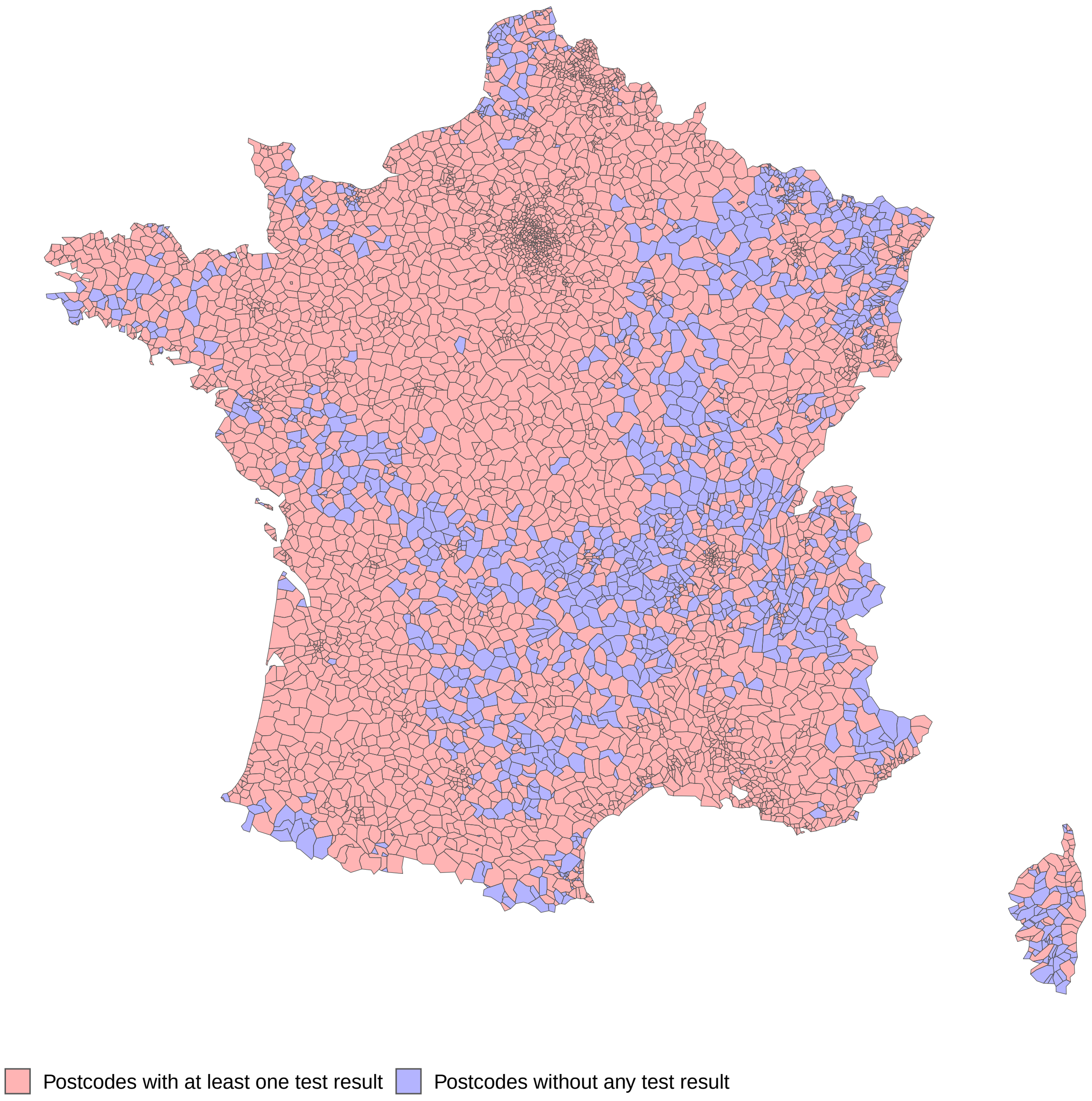

**Figure S13 Postcodes with at least one test result observed in the analytic sample.**

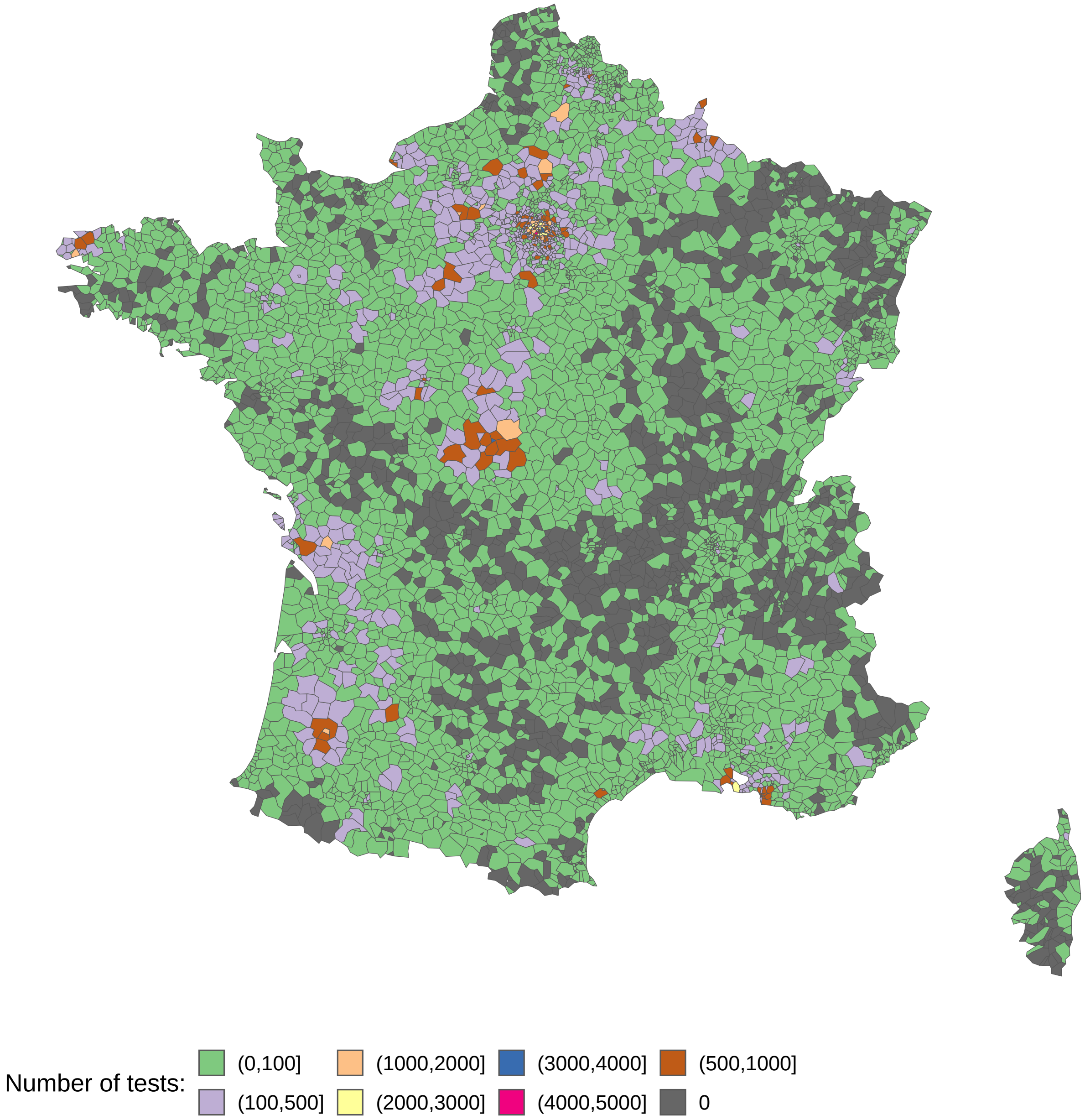

**Figure S14 Postcodes stratified by the number of HR HPV tests in the analytic sample.**

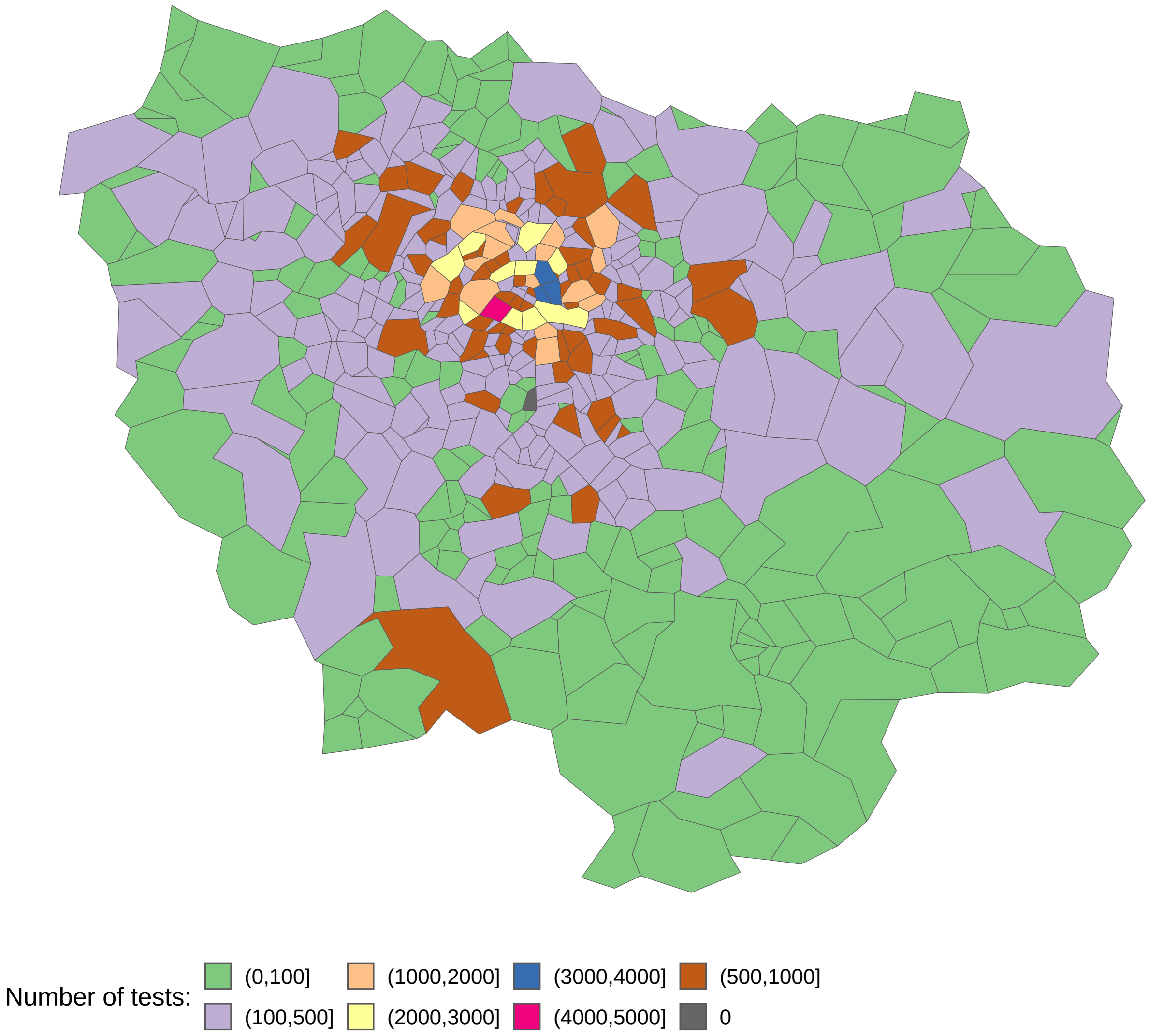

**Figure S15 Postcodes stratified by the number of HR HPV tests in the analytic sample, zoom on Paris region.**

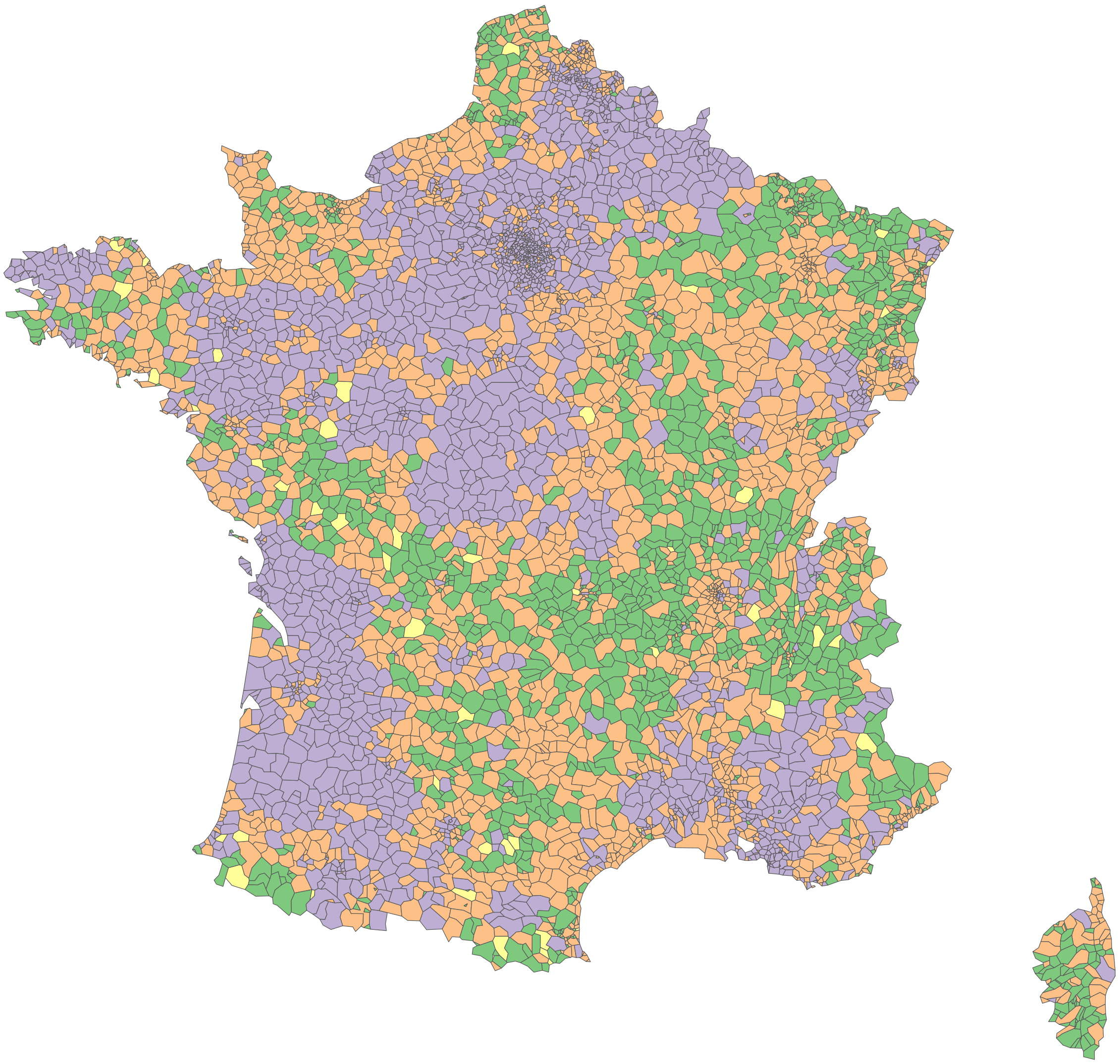

**Figure S16** **Postcodes only observed through opportunistic screening, organised screening, both opportunistic and organised screening, and not observed in the analytic sample.**

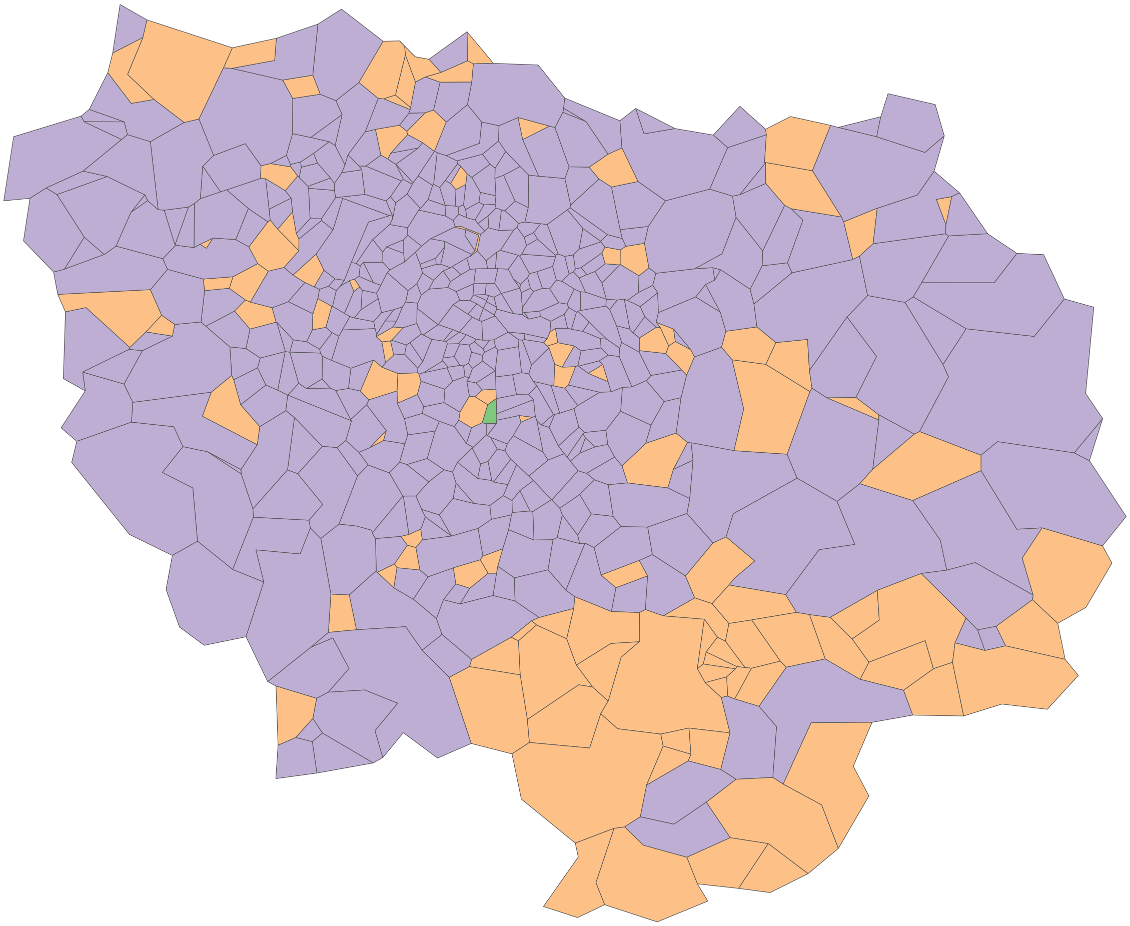

**Figure S17 Postcodes only observed through opportunistic screening, organised screening, both opportunistic and organised screening, and not observed in the analytic sample, zoom on Paris region.**

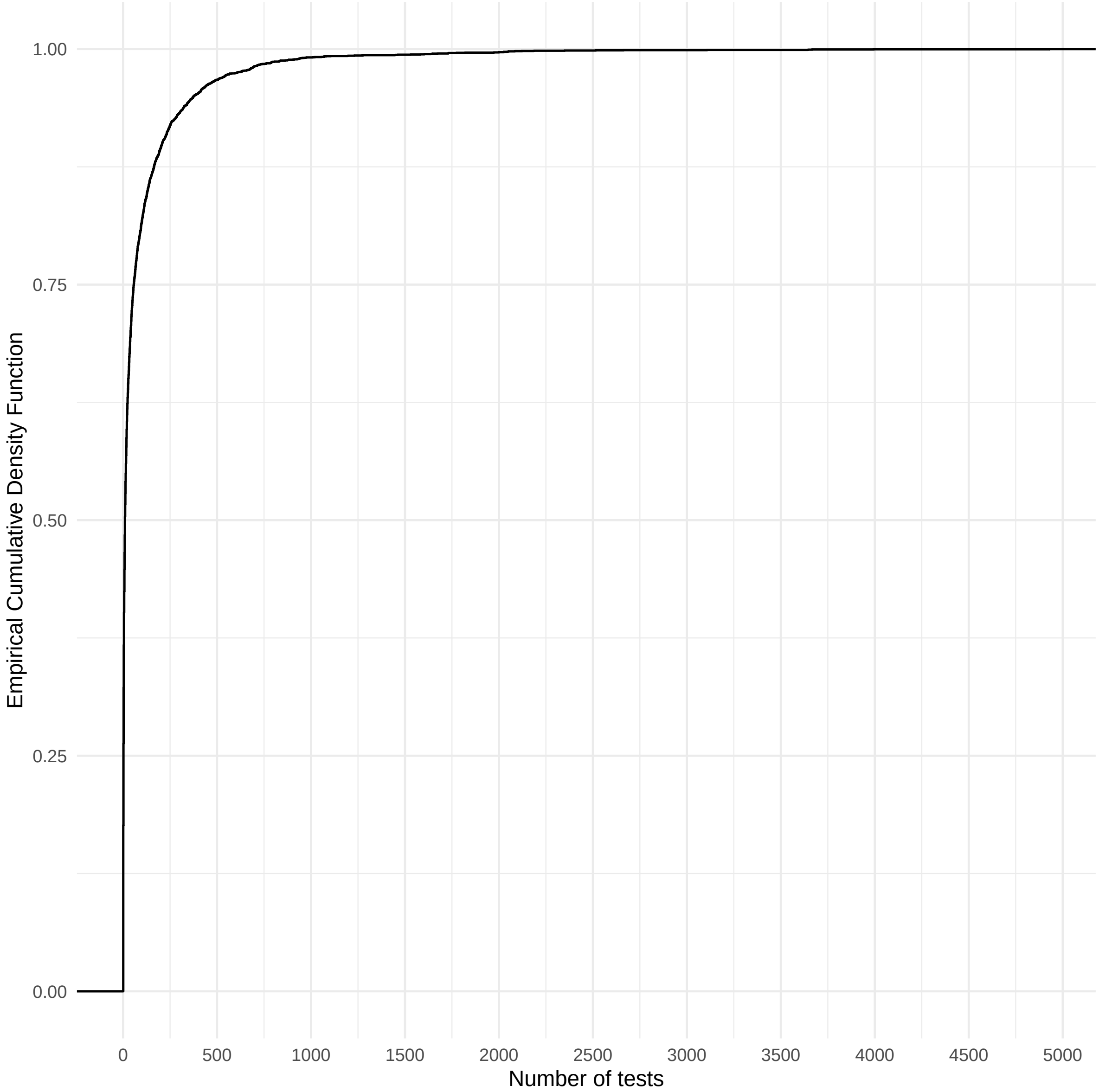

**Figure S18 Empirical cumulative distribution function of the number of tests within postcodes in the analytic sample.**

### S7 Performance of the selected model

#### S7.1 Model selection and raw reporting of the selected model

**Table S5 Logarithmic-score based on LGO-CV. The model with the minimum score was selected.**

| Model | Type | Structure | Log-score |
|---|---|---|---|
| Model 5 | BYM2 | 1st-order Queen | 0.2351 |
| Model 4 | BYM2 | SOI | 0.2351 |
| Model 3 | BYM2 | Delaunay | 0.2351 |
| Model 6 | BYM2 | 2nd-order Queen | 0.2351 |
| Model 1 | Matérn | Stationary | 0.2352 |
| Model 2 | Matérn | Non-stationary (Barrier model) | 0.2352 |

**Table S6 Raw summary provided by R-INLA of the posterior distribution for the latent parameters.**

| | Description of the posterior distribution | | | | | |
|---|---|---|---|---|---|---|
| | | | Quantile | | | |
| Parameters | Average | SD | 0.025 | 0.5 | 0.975 | Mode |
| $\psi_0$ | -3.48 | 0.05 | -3.58 | -3.48 | -3.38 | -3.48 |
| $\psi_1$ | 0.88 | 0.05 | 0.76 | 0.88 | 0.98 | 0.90 |
| $\psi_2$ | 0.23 | 0.05 | 0.13 | 0.23 | 0.32 | 0.23 |
| $\psi_3$ | 0.04 | 0.06 | -0.07 | 0.03 | 0.15 | 0.02 |

**Table S7 Raw summary provided by R-INLA of the posterior distribution for the hyperparameters.**

| Parameters | Description of the posterior distribution | | | | | |
|---|---|---|---|---|---|---|
| | | | Quantile | | | |
| | Average | SD | 0.025 | 0.5 | 0.975 | Mode |
| $\sigma^{-2}$ for $\psi_1$ | 1.63 | 3.08 | 0.04 | 0.71 | 8.82 | 0.08 |
| $\sigma^{-2}$ for $\psi_2$ | 20.50 | 104.01 | 0.03 | 3.06 | 146.30 | 0.01 |
| $\sigma^{-2}$ for $\psi_3$ | 3,782.89 | 192,792.03 | 0.04 | 23.92 | 13,055.10 | 0.00 |
| $\sigma^{-2}$ for $\gamma_{AGE}(age_s, g_s)$ | 75.32 | 49.27 | 19.78 | 62.96 | 204.37 | 44.24 |
| $\eta$ for $\gamma_{AGE}(age_s, g_s)$ | -0.21 | 0.41 | -0.86 | -0.26 | 0.63 | -0.47 |
| $\sigma^{-2}$ for $\gamma'_{AGE}(age_s, g_s)$ | 4,445.98 | 21,985.49 | 33.97 | 874.46 | 30,422.39 | 59.18 |
| $\eta$ for $\gamma'_{AGE}(age_s, g_s)$ | 0.12 | 0.67 | -0.97 | 0.22 | 0.99 | 0.99 |
| $\sigma^{-2}$ for $\alpha_{SPACE}(z_s, g_s)$ | 15.44 | 2.44 | 11.07 | 15.29 | 20.67 | 15.06 |
| $\phi$ for $\alpha_{SPACE}(z_s, g_s)$ | 0.95 | 0.05 | 0.82 | 0.96 | 0.99 | 0.98 |
| $\eta$ for $\alpha_{SPACE}(z_s, g_s)$ | 0.95 | 0.03 | 0.89 | 0.96 | 0.99 | 0.97 |
| $\sigma^{-2}$ for $\alpha'_{SPACE}(z_s, g_s)$ | 82.27 | 46.95 | 27.33 | 70.98 | 205.00 | 53.72 |
| $\phi$ for $\alpha'_{SPACE}(z_s, g_s)$ | 0.80 | 0.16 | 0.38 | 0.84 | 0.98 | 0.95 |
| $\eta$ for $\alpha'_{SPACE}(z_s, g_s)$ | 0.91 | 0.10 | 0.62 | 0.94 | 0.99 | 0.98 |
| $\sigma^{-2}$ for $\beta_{TIME}(t_s, g_s)$ | 1,018.77 | 4,944.59 | 5.73 | 197.67 | 6,991.05 | 7.90 |
| $\eta$ for $\beta_{TIME}(t_s, g_s)$ | 0.13 | 0.62 | -0.94 | 0.21 | 0.98 | 0.97 |
| $\sigma^{-2}$ for $\beta'_{TIME}(t_s, g_s)$ | 323.38 | 814.88 | 7.28 | 118.44 | 1,929.37 | 15.94 |
| $\eta$ for $\beta'_{TIME}(t_s, g_s)$ | -0.04 | 0.63 | -0.96 | -0.06 | 0.96 | -0.95 |

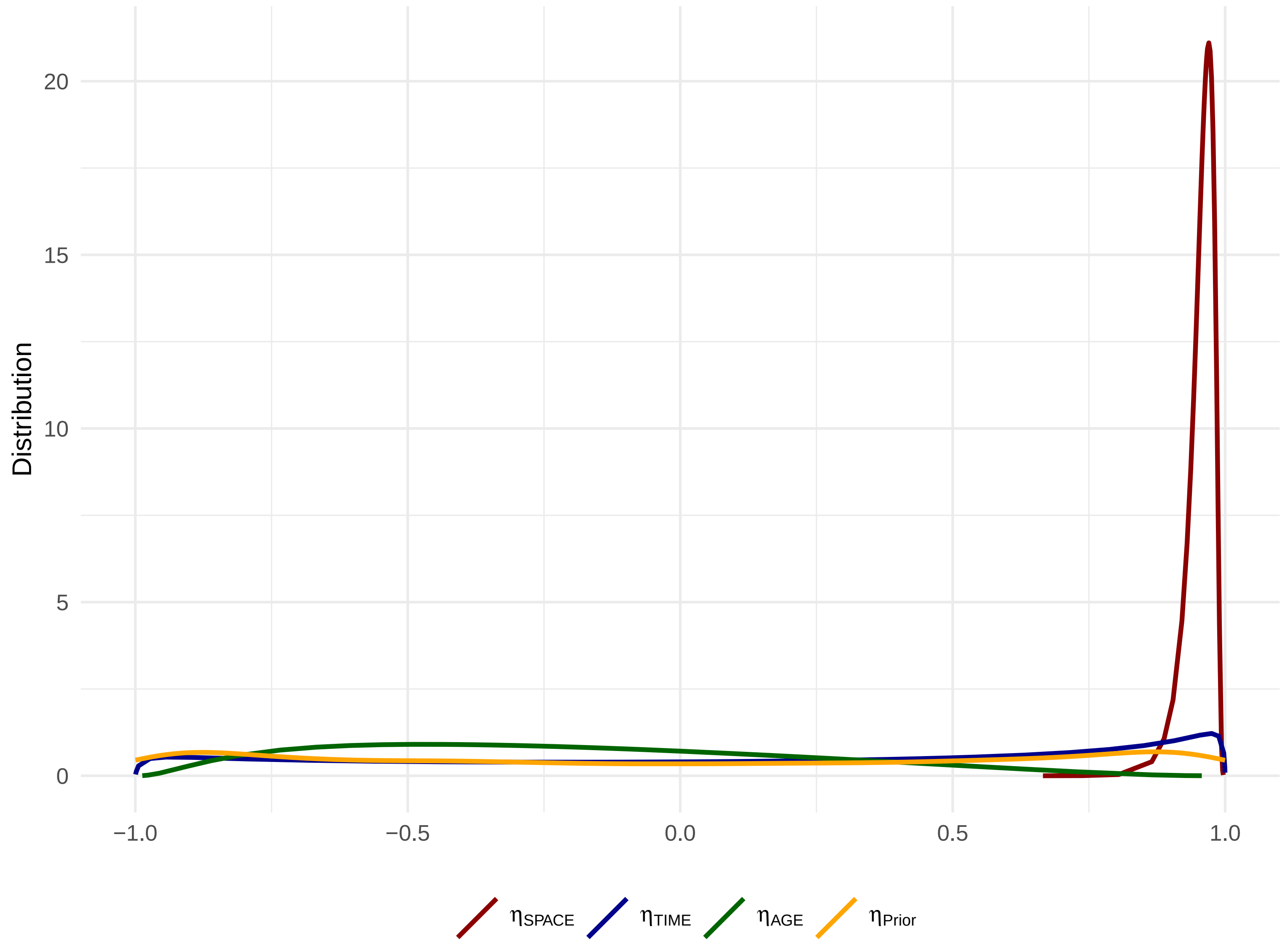

**Figure S19 Posterior and prior distribution for the correlations between the two groups of genotypes for components specific to organised screening.**

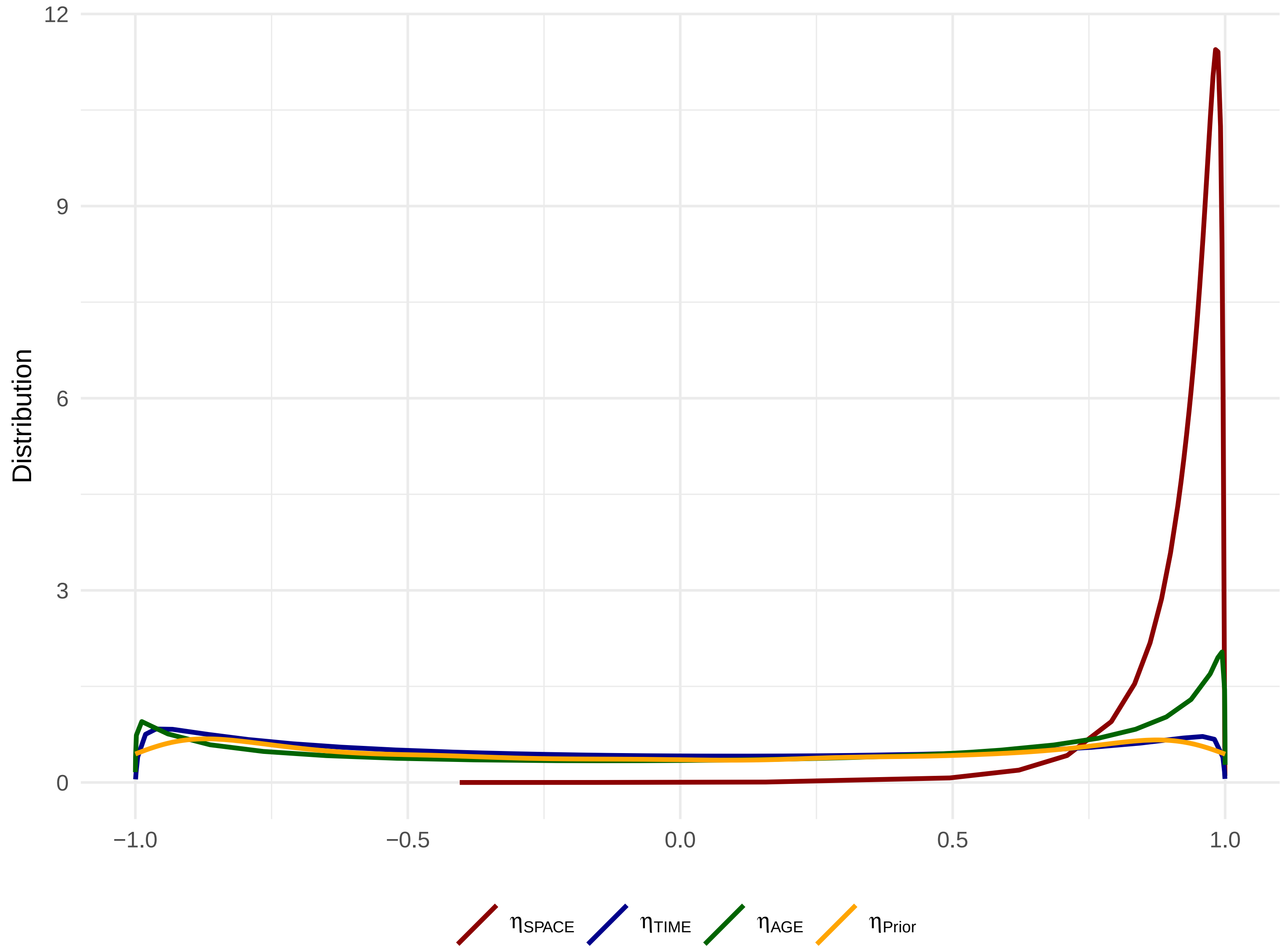

**Figure S20 Posterior and prior distribution for the correlations between the two groups of genotypes for components specific to opportunistic screening.**

#### S7.2 Posterior predictive check from the selected model

**Table S8 Observed HR HPV cervical infection prevalence, posterior predictive HR HPV cervical infection prevalence, and posterior expected HR HPV cervical infection prevalence, stratified by various dimensions.**

| | | | | | | | Posterior distribution | | | | | | | |
|---|---|---|---|---|---|---|---|---|---|---|---|---|---|---|
| | | | | | | | Predictive prevalence | | | | Expected prevalence | | | |
| | | | | | | | | | ETI95% | | | | ETI95% | |
| Region | Genotypes | Type of screening | Year | Age | Number of tests | Observed | Average | Median | LB | UB | Average | Median | LB | UB |
| | HPV16/18 | | | | 362,963 | 3.70 | 3.72 | 3.73 | 3.64 | 3.81 | 3.72 | 3.72 | 3.66 | 3.78 |
| | Other genotypes | | | | 362,963 | 9.23 | 9.27 | 9.27 | 9.14 | 9.41 | 9.27 | 9.27 | 9.18 | 9.37 |
| | HPV16/18 | | 2020 | | 71,081 | 3.12 | 3.19 | 3.19 | 3.02 | 3.38 | 3.19 | 3.19 | 3.07 | 3.32 |
| | HPV16/18 | | 2021 | | 159,182 | 3.65 | 3.64 | 3.64 | 3.51 | 3.77 | 3.64 | 3.64 | 3.55 | 3.73 |
| | HPV16/18 | | 2022 | | 72,265 | 3.89 | 3.93 | 3.93 | 3.75 | 4.12 | 3.93 | 3.93 | 3.80 | 4.06 |
| | HPV16/18 | | 2023 | | 60,435 | 4.32 | 4.33 | 4.33 | 4.11 | 4.55 | 4.33 | 4.33 | 4.18 | 4.49 |
| | Other genotypes | | 2020 | | 71,081 | 9.36 | 9.44 | 9.44 | 9.15 | 9.74 | 9.44 | 9.44 | 9.24 | 9.65 |
| | Other genotypes | | 2021 | | 159,182 | 8.75 | 8.77 | 8.77 | 8.58 | 8.95 | 8.77 | 8.77 | 8.64 | 8.91 |
| | Other genotypes | | 2022 | | 72,265 | 9.08 | 9.12 | 9.12 | 8.84 | 9.41 | 9.12 | 9.12 | 8.93 | 9.32 |
| | Other genotypes | | 2023 | | 60,435 | 10.49 | 10.57 | 10.57 | 10.24 | 10.92 | 10.58 | 10.58 | 10.35 | 10.82 |
| | HPV16/18 | | | [30,39] | | | 4.54 | 4.55 | 4.39 | 4.70 | 4.54 | 4.55 | 4.44 | 4.65 |
| | HPV16/18 | | | (39,49] | 104,989 | 3.68 | 3.71 | 3.70 | 3.55 | 3.86 | 3.70 | 3.70 | 3.60 | 3.82 |
| | HPV16/18 | | | (49,59] | 83,824 | 2.91 | 2.95 | 2.95 | 2.80 | 3.12 | 2.95 | 2.95 | 2.85 | 3.06 |
| | HPV16/18 | | | (59,66] | 42,356 | 2.77 | 2.75 | 2.74 | 2.54 | 2.96 | 2.75 | 2.75 | 2.60 | 2.89 |
| | Other genotypes | | | [30,39] | | | 12.16 | 12.15 | 11.91 | 12.40 | 12.16 | 12.16 | 11.98 | 12.33 |
| | Other genotypes | | | (39,49] | 104,989 | 8.42 | 8.49 | 8.49 | 8.26 | 8.72 | 8.49 | 8.49 | 8.32 | 8.65 |
| | Other genotypes | | | (49,59] | 83,824 | 7.14 | 7.18 | 7.17 | 6.94 | 7.42 | 7.18 | 7.18 | 7.02 | 7.35 |
| | Other genotypes | | | (59,66] | 42,356 | 6.36 | 6.38 | 6.38 | 6.05 | 6.71 | 6.38 | 6.38 | 6.16 | 6.61 |
| Auvergne-Rhône-Alpes | HPV16/18 | | | | 4,671 | 3.81 | 4.01 | 4.00 | 3.38 | 4.71 | 4.02 | 4.01 | 3.69 | 4.35 |
| Bourgogne-Franche-Comté | HPV16/18 | | | | 4,035 | 3.64 | 3.67 | 3.67 | 3.02 | 4.36 | 3.66 | 3.66 | 3.33 | 4.03 |
| Bretagne | HPV16/18 | | | | 11,062 | 3.88 | 3.87 | 3.87 | 3.42 | 4.36 | 3.87 | 3.87 | 3.58 | 4.19 |
| Centre-Val de Loire | HPV16/18 | | | | 32,749 | 3.70 | 3.59 | 3.59 | 3.34 | 3.86 | 3.59 | 3.59 | 3.44 | 3.76 |
| Corse | HPV16/18 | | | | 430 | 3.49 | 3.43 | 3.49 | 1.63 | 5.35 | 3.40 | 3.38 | 2.71 | 4.25 |
| Grand Est | HPV16/18 | | | | 11,886 | 3.58 | 3.60 | 3.59 | 3.18 | 4.05 | 3.59 | 3.59 | 3.35 | 3.85 |
| Hauts-de-France | HPV16/18 | | | | 31,938 | 3.72 | 3.74 | 3.73 | 3.48 | 4.02 | 3.74 | 3.74 | 3.57 | 3.91 |
| Normandie | HPV16/18 | | | | 19,963 | 4.32 | 4.15 | 4.15 | 3.79 | 4.52 | 4.15 | 4.15 | 3.92 | 4.38 |
| Nouvelle-Aquitaine | HPV16/18 | | | | 30,721 | 3.76 | 3.81 | 3.81 | 3.54 | 4.10 | 3.82 | 3.82 | 3.64 | 4.00 |
| Occitanie | HPV16/18 | | | | 12,524 | 4.07 | 4.09 | 4.08 | 3.66 | 4.51 | 4.09 | 4.09 | 3.84 | 4.34 |
| Pays de la Loire | HPV16/18 | | | | 6,705 | 2.98 | 3.14 | 3.13 | 2.67 | 3.64 | 3.14 | 3.14 | 2.89 | 3.40 |
| Provence-Alpes-Côte d'Azur | HPV16/18 | | | | 27,889 | 4.11 | 4.12 | 4.12 | 3.81 | 4.44 | 4.12 | 4.12 | 3.92 | 4.34 |
| Île-de-France | HPV16/18 | | | | 168,390 | 3.55 | 3.61 | 3.61 | 3.48 | 3.73 | 3.60 | 3.60 | 3.52 | 3.69 |
| Auvergne-Rhône-Alpes | Other genotypes | | | | 4,671 | 9.98 | 9.90 | 9.89 | 8.82 | 10.98 | 9.90 | 9.89 | 9.24 | 10.59 |
| Bourgogne-Franche-Comté | Other genotypes | | | | 4,035 | 9.59 | 9.00 | 9.00 | 7.88 | 10.14 | 9.00 | 9.00 | 8.35 | 9.73 |
| Bretagne | Other genotypes | | | | 11,062 | 8.82 | 8.83 | 8.81 | 8.14 | 9.56 | 8.82 | 8.82 | 8.35 | 9.31 |
| Centre-Val de Loire | Other genotypes | | | | 32,749 | 8.18 | 8.34 | 8.34 | 7.94 | 8.73 | 8.34 | 8.34 | 8.07 | 8.61 |
| Corse | Other genotypes | | | | 430 | 10.93 | 10.13 | 10.00 | 6.98 | 13.72 | 10.11 | 10.07 | 8.35 | 12.22 |

**Table S8** **Observed HR HPV cervical infection prevalence, posterior predictive HR HPV cervical infection prevalence, and posterior expected HR HPV cervical infection prevalence, stratified by various dimensions.** ***(continued)***

| | | | | | | | Posterior distribution | | | | | | | |
|---|---|---|---|---|---|---|---|---|---|---|---|---|---|---|
| | | | | | | | Predictive prevalence | | | | Expected prevalence | | | |
| | | | | | | | | | ETI95% | | | | ETI95% | |
| Region | Genotypes | Type of screening | Year | Age | Number of tests | Observed | Average | Median | LB | UB | Average | Median | LB | UB |
| Grand Est | Other genotypes | | | | 11,886 | 8.51 | 8.61 | 8.61 | 7.93 | 9.30 | 8.61 | 8.60 | 8.18 | 9.08 |
| Hauts-de-France | Other genotypes | | | | 31,938 | 8.80 | 8.91 | 8.91 | 8.49 | 9.33 | 8.91 | 8.92 | 8.63 | 9.20 |
| Normandie | Other genotypes | | | | 19,963 | 8.97 | 9.22 | 9.22 | 8.66 | 9.76 | 9.22 | 9.22 | 8.86 | 9.59 |
| Nouvelle-Aquitaine | Other genotypes | | | | 30,721 | 9.22 | 9.20 | 9.20 | 8.77 | 9.65 | 9.20 | 9.20 | 8.92 | 9.51 |
| Occitanie | Other genotypes | | | | 12,524 | 9.85 | 9.86 | 9.86 | 9.17 | 10.54 | 9.86 | 9.86 | 9.42 | 10.33 |
| Pays de la Loire | Other genotypes | | | | 6,705 | 7.05 | 7.36 | 7.35 | 6.58 | 8.16 | 7.36 | 7.35 | 6.90 | 7.86 |
| Provence-Alpes-Côte d'Azur | Other genotypes | | | | 27,889 | 9.82 | 9.88 | 9.87 | 9.41 | 10.36 | 9.88 | 9.88 | 9.55 | 10.21 |
| Île-de-France | Other genotypes | | | | 168,390 | 9.53 | 9.53 | 9.53 | 9.34 | 9.73 | 9.53 | 9.53 | 9.40 | 9.68 |
| | HPV16/18 | Opportunistic | | | 343,042 | 3.75 | 3.77 | 3.77 | 3.68 | 3.86 | 3.77 | 3.77 | 3.71 | 3.83 |
| | HPV16/18 | Organised | | | 19,921 | 2.89 | 2.90 | 2.91 | 2.58 | 3.22 | 2.91 | 2.91 | 2.69 | 3.14 |
| | Other genotypes | Opportunistic | | | 343,042 | 9.36 | 9.41 | 9.41 | 9.27 | 9.55 | 9.41 | 9.41 | 9.31 | 9.50 |
| | Other genotypes | Organised | | | 19,921 | 6.89 | 6.93 | 6.92 | 6.46 | 7.39 | 6.94 | 6.94 | 6.59 | 7.28 |
| | HPV16/18 | | 2020 | [30,39] | | | 3.86 | 3.86 | 3.59 | 4.15 | 3.86 | 3.86 | 3.70 | 4.03 |
| | HPV16/18 | | 2020 | (39,49] | 21,620 | 2.86 | 3.17 | 3.17 | 2.90 | 3.46 | 3.17 | 3.17 | 3.03 | 3.32 |
| | HPV16/18 | | 2020 | (49,59] | 16,498 | 2.36 | 2.53 | 2.53 | 2.27 | 2.81 | 2.53 | 2.53 | 2.41 | 2.66 |
| | HPV16/18 | | 2020 | (59,66] | 7,086 | 2.27 | 2.35 | 2.34 | 1.98 | 2.75 | 2.35 | 2.35 | 2.20 | 2.50 |
| | HPV16/18 | | 2021 | [30,39] | | | 4.45 | 4.45 | 4.23 | 4.67 | 4.45 | 4.45 | 4.32 | 4.57 |
| | HPV16/18 | | 2021 | (39,49] | 46,281 | 3.54 | 3.64 | 3.64 | 3.44 | 3.85 | 3.64 | 3.64 | 3.52 | 3.77 |
| | HPV16/18 | | 2021 | (49,59] | 36,867 | 2.82 | 2.90 | 2.90 | 2.70 | 3.12 | 2.91 | 2.90 | 2.79 | 3.02 |
| | HPV16/18 | | 2021 | (59,66] | 19,728 | 2.69 | 2.70 | 2.70 | 2.44 | 2.99 | 2.70 | 2.70 | 2.55 | 2.86 |
| | HPV16/18 | | 2022 | [30,39] | | | 4.79 | 4.79 | 4.48 | 5.09 | 4.79 | 4.79 | 4.61 | 4.97 |
| | HPV16/18 | | 2022 | (39,49] | 20,354 | 4.08 | 3.91 | 3.91 | 3.59 | 4.24 | 3.91 | 3.91 | 3.75 | 4.07 |
| | HPV16/18 | | 2022 | (49,59] | 16,678 | 2.99 | 3.10 | 3.10 | 2.80 | 3.39 | 3.10 | 3.09 | 2.96 | 3.24 |
| | HPV16/18 | | 2022 | (59,66] | 8,295 | 2.83 | 2.86 | 2.86 | 2.47 | 3.25 | 2.86 | 2.85 | 2.68 | 3.03 |
| | HPV16/18 | | 2023 | [30,39] | | | 5.27 | 5.27 | 4.93 | 5.64 | 5.28 | 5.28 | 5.08 | 5.48 |
| | HPV16/18 | | 2023 | (39,49] | 16,734 | 4.63 | 4.32 | 4.32 | 3.98 | 4.68 | 4.32 | 4.32 | 4.14 | 4.52 |
| | HPV16/18 | | 2023 | (49,59] | 13,781 | 3.74 | 3.43 | 3.43 | 3.09 | 3.77 | 3.43 | 3.43 | 3.26 | 3.60 |
| | HPV16/18 | | 2023 | (59,66] | 7,247 | 3.38 | 3.13 | 3.12 | 2.69 | 3.59 | 3.13 | 3.13 | 2.93 | 3.34 |
| | Other genotypes | | 2020 | [30,39] | | | 12.29 | 12.29 | 11.81 | 12.81 | 12.30 | 12.30 | 12.01 | 12.59 |
| | Other genotypes | | 2020 | (39,49] | 21,620 | 8.41 | 8.64 | 8.64 | 8.19 | 9.08 | 8.64 | 8.64 | 8.41 | 8.87 |
| | Other genotypes | | 2020 | (49,59] | 16,498 | 7.18 | 7.30 | 7.29 | 6.86 | 7.77 | 7.30 | 7.29 | 7.08 | 7.53 |
| | Other genotypes | | 2020 | (59,66] | 7,086 | 5.91 | 6.48 | 6.48 | 5.86 | 7.11 | 6.47 | 6.47 | 6.21 | 6.74 |
| | Other genotypes | | 2021 | [30,39] | | | 11.53 | 11.53 | 11.19 | 11.88 | 11.53 | 11.53 | 11.32 | 11.75 |
| | Other genotypes | | 2021 | (39,49] | 46,281 | 8.01 | 8.09 | 8.09 | 7.80 | 8.40 | 8.09 | 8.09 | 7.91 | 8.27 |
| | Other genotypes | | 2021 | (49,59] | 36,867 | 6.69 | 6.83 | 6.83 | 6.52 | 7.14 | 6.83 | 6.83 | 6.66 | 7.01 |
| | Other genotypes | | 2021 | (59,66] | 19,728 | 6.03 | 6.08 | 6.08 | 5.68 | 6.48 | 6.08 | 6.08 | 5.86 | 6.31 |
| | Other genotypes | | 2022 | [30,39] | | | 11.92 | 11.92 | 11.44 | 12.39 | 11.91 | 11.91 | 11.64 | 12.19 |
| | Other genotypes | | 2022 | (39,49] | 20,354 | 8.24 | 8.30 | 8.29 | 7.86 | 8.74 | 8.30 | 8.30 | 8.08 | 8.53 |
| | Other genotypes | | 2022 | (49,59] | 16,678 | 7.25 | 7.02 | 7.02 | 6.60 | 7.49 | 7.03 | 7.03 | 6.82 | 7.24 |

**Table S8 Observed HR HPV cervical infection prevalence, posterior predictive HR HPV cervical infection prevalence, and posterior expected HR HPV cervical infection prevalence, stratified by various dimensions.** ***(continued)***

| | | | | | | | Posterior distribution | | | | | | | |
|---|---|---|---|---|---|---|---|---|---|---|---|---|---|---|
| | | | | | | | Predictive prevalence | | | | Expected prevalence | | | |
| | | | | | | | | | ETI95% | | | | ETI95% | |
| Region | Genotypes | Type of screening | Year | Age | Number of tests | Observed | Average | Median | LB | UB | Average | Median | LB | UB |
| | Other genotypes | | 2022 | (59,66] | 8,295 | 6.04 | 6.29 | 6.29 | 5.74 | 6.91 | 6.29 | 6.29 | 6.03 | 6.55 |
| | Other genotypes | | 2023 | [30,39] | | | 13.84 | 13.84 | 13.30 | 14.39 | 13.84 | 13.84 | 13.51 | 14.17 |
| | Other genotypes | | 2023 | (39,49] | 16,734 | 9.75 | 9.62 | 9.62 | 9.11 | 10.14 | 9.62 | 9.62 | 9.35 | 9.89 |
| | Other genotypes | | 2023 | (49,59] | 13,781 | 8.20 | 8.15 | 8.14 | 7.64 | 8.69 | 8.15 | 8.15 | 7.91 | 8.42 |
| | Other genotypes | | 2023 | (59,66] | 7,247 | 8.06 | 7.20 | 7.19 | 6.55 | 7.85 | 7.20 | 7.20 | 6.91 | 7.50 |
| Auvergne-Rhône-Alpes | HPV16/18 | | 2020 | | 988 | 3.44 | 3.46 | 3.44 | 2.33 | 4.76 | 3.48 | 3.47 | 3.14 | 3.83 |
| Bourgogne-Franche-Comté | HPV16/18 | | 2020 | | 537 | 2.98 | 3.08 | 2.98 | 1.68 | 4.66 | 3.05 | 3.04 | 2.73 | 3.39 |
| Bretagne | HPV16/18 | | 2020 | | 559 | 3.76 | 3.24 | 3.22 | 1.79 | 4.83 | 3.23 | 3.23 | 2.88 | 3.62 |
| Centre-Val de Loire | HPV16/18 | | 2020 | | 3,670 | 2.64 | 3.07 | 3.08 | 2.51 | 3.68 | 3.08 | 3.07 | 2.89 | 3.27 |
| Corse | HPV16/18 | | 2020 | | 377 | 3.45 | 3.36 | 3.18 | 1.59 | 5.57 | 3.32 | 3.30 | 2.55 | 4.28 |
| Grand Est | HPV16/18 | | 2020 | | 882 | 3.17 | 2.77 | 2.72 | 1.70 | 3.97 | 2.77 | 2.77 | 2.48 | 3.08 |
| Hauts-de-France | HPV16/18 | | 2020 | | 2,840 | 3.49 | 3.01 | 2.99 | 2.36 | 3.70 | 3.02 | 3.01 | 2.83 | 3.21 |
| Normandie | HPV16/18 | | 2020 | | 2,313 | 3.67 | 3.30 | 3.29 | 2.55 | 4.11 | 3.30 | 3.30 | 3.06 | 3.54 |
| Nouvelle-Aquitaine | HPV16/18 | | 2020 | | 1,689 | 3.37 | 3.16 | 3.14 | 2.31 | 4.09 | 3.16 | 3.16 | 2.95 | 3.38 |
| Occitanie | HPV16/18 | | 2020 | | 2,179 | 4.22 | 3.91 | 3.90 | 3.03 | 4.82 | 3.91 | 3.91 | 3.55 | 4.31 |
| Pays de la Loire | HPV16/18 | | 2020 | | 1,161 | 1.72 | 2.65 | 2.67 | 1.72 | 3.62 | 2.67 | 2.66 | 2.40 | 2.96 |
| Provence-Alpes-Côte d'Azur | HPV16/18 | | 2020 | | 2,131 | 3.10 | 3.24 | 3.24 | 2.49 | 4.08 | 3.24 | 3.23 | 3.00 | 3.49 |
| Île-de-France | HPV16/18 | | 2020 | | 51,755 | 3.07 | 3.19 | 3.18 | 3.00 | 3.39 | 3.19 | 3.19 | 3.06 | 3.32 |
| Auvergne-Rhône-Alpes | HPV16/18 | | 2021 | | 3,147 | 3.37 | 4.03 | 4.04 | 3.30 | 4.80 | 4.04 | 4.03 | 3.70 | 4.38 |
| Bourgogne-Franche-Comté | HPV16/18 | | 2021 | | 1,537 | 3.97 | 3.58 | 3.58 | 2.67 | 4.62 | 3.58 | 3.58 | 3.24 | 3.95 |
| Bretagne | HPV16/18 | | 2021 | | 2,374 | 3.45 | 3.60 | 3.62 | 2.82 | 4.42 | 3.60 | 3.59 | 3.27 | 3.95 |
| Centre-Val de Loire | HPV16/18 | | 2021 | | 15,029 | 3.35 | 3.47 | 3.47 | 3.14 | 3.81 | 3.47 | 3.47 | 3.31 | 3.66 |
| Corse | HPV16/18 | | 2021 | | 35 | 2.86 | 3.73 | 2.86 | 0.00 | 11.43 | 3.77 | 3.76 | 3.27 | 4.35 |
| Grand Est | HPV16/18 | | 2021 | | 3,923 | 3.14 | 3.35 | 3.34 | 2.75 | 3.98 | 3.34 | 3.33 | 3.10 | 3.58 |
| Hauts-de-France | HPV16/18 | | 2021 | | 10,748 | 3.67 | 3.55 | 3.54 | 3.16 | 3.94 | 3.55 | 3.55 | 3.37 | 3.73 |
| Normandie | HPV16/18 | | 2021 | | 8,079 | 4.30 | 3.93 | 3.94 | 3.44 | 4.43 | 3.93 | 3.93 | 3.69 | 4.17 |
| Nouvelle-Aquitaine | HPV16/18 | | 2021 | | 8,917 | 3.51 | 3.54 | 3.54 | 3.12 | 3.99 | 3.54 | 3.54 | 3.36 | 3.74 |
| Occitanie | HPV16/18 | | 2021 | | 5,839 | 4.01 | 4.06 | 4.06 | 3.49 | 4.66 | 4.07 | 4.07 | 3.81 | 4.34 |
| Pays de la Loire | HPV16/18 | | 2021 | | 3,081 | 3.21 | 3.06 | 3.05 | 2.43 | 3.77 | 3.06 | 3.06 | 2.78 | 3.34 |
| Provence-Alpes-Côte d'Azur | HPV16/18 | | 2021 | | 11,313 | 3.81 | 3.88 | 3.88 | 3.47 | 4.31 | 3.88 | 3.88 | 3.67 | 4.11 |
| Île-de-France | HPV16/18 | | 2021 | | 85,160 | 3.65 | 3.62 | 3.62 | 3.46 | 3.79 | 3.62 | 3.62 | 3.52 | 3.73 |
| Auvergne-Rhône-Alpes | HPV16/18 | | 2022 | | 295 | 8.47 | 4.63 | 4.41 | 2.37 | 7.12 | 4.62 | 4.61 | 4.13 | 5.18 |
| Bourgogne-Franche-Comté | HPV16/18 | | 2022 | | 1,220 | 3.36 | 3.90 | 3.85 | 2.79 | 5.16 | 3.89 | 3.89 | 3.51 | 4.33 |
| Bretagne | HPV16/18 | | 2022 | | 3,082 | 3.50 | 3.89 | 3.89 | 3.18 | 4.64 | 3.89 | 3.89 | 3.58 | 4.23 |
| Centre-Val de Loire | HPV16/18 | | 2022 | | 7,977 | 4.25 | 3.63 | 3.64 | 3.18 | 4.10 | 3.64 | 3.64 | 3.45 | 3.85 |
| Corse | HPV16/18 | | 2022 | | 13 | 0.00 | 4.44 | 0.00 | 0.00 | 15.38 | 4.33 | 4.31 | 3.69 | 5.09 |
| Grand Est | HPV16/18 | | 2022 | | 3,704 | 3.75 | 3.66 | 3.64 | 3.00 | 4.37 | 3.66 | 3.66 | 3.37 | 3.95 |
| Hauts-de-France | HPV16/18 | | 2022 | | 9,943 | 3.53 | 3.77 | 3.76 | 3.34 | 4.20 | 3.77 | 3.77 | 3.57 | 3.98 |
| Normandie | HPV16/18 | | 2022 | | 5,912 | 4.35 | 4.39 | 4.38 | 3.82 | 4.99 | 4.38 | 4.38 | 4.11 | 4.66 |

**Table S8 Observed HR HPV cervical infection prevalence, posterior predictive HR HPV cervical infection prevalence, and posterior expected HR HPV cervical infection prevalence, stratified by various dimensions.** ***(continued)***

| | | | | | | | Posterior distribution | | | | | | | |
|---|---|---|---|---|---|---|---|---|---|---|---|---|---|---|
| | | | | | | | Predictive prevalence | | | | Expected prevalence | | | |
| | | | | | | | | | ETI95% | | | | ETI95% | |
| Region | Genotypes | Type of screening | Year | Age | Number of tests | Observed | Average | Median | LB | UB | Average | Median | LB | UB |
| Nouvelle-Aquitaine | HPV16/18 | | 2022 | | 10,965 | 3.80 | 3.79 | 3.78 | 3.39 | 4.22 | 3.79 | 3.79 | 3.58 | 3.99 |
| Occitanie | HPV16/18 | | 2022 | | 3,131 | 3.93 | 4.08 | 4.09 | 3.35 | 4.85 | 4.08 | 4.08 | 3.79 | 4.37 |
| Pays de la Loire | HPV16/18 | | 2022 | | 962 | 2.49 | 3.40 | 3.33 | 2.29 | 4.57 | 3.37 | 3.36 | 3.08 | 3.71 |
| Provence-Alpes-Côte d'Azur | HPV16/18 | | 2022 | | 8,238 | 4.44 | 4.24 | 4.24 | 3.74 | 4.76 | 4.24 | 4.24 | 4.00 | 4.49 |
| Île-de-France | HPV16/18 | | 2022 | | 16,823 | 3.67 | 3.99 | 3.99 | 3.64 | 4.32 | 3.99 | 3.99 | 3.81 | 4.17 |
| Auvergne-Rhône-Alpes | HPV16/18 | | 2023 | | 241 | 5.39 | 5.21 | 4.98 | 2.49 | 8.30 | 5.23 | 5.21 | 4.50 | 6.07 |
| Bourgogne-Franche-Comté | HPV16/18 | | 2023 | | 741 | 3.91 | 3.92 | 3.91 | 2.43 | 5.40 | 3.91 | 3.90 | 3.45 | 4.42 |
| Bretagne | HPV16/18 | | 2023 | | 5,047 | 4.32 | 4.06 | 4.06 | 3.45 | 4.72 | 4.06 | 4.05 | 3.73 | 4.41 |
| Centre-Val de Loire | HPV16/18 | | 2023 | | 6,073 | 4.46 | 4.13 | 4.12 | 3.59 | 4.69 | 4.13 | 4.13 | 3.90 | 4.37 |
| Corse | HPV16/18 | | 2023 | | 5 | 20.00 | 3.77 | 0.00 | 0.00 | 20.00 | 4.08 | 4.02 | 3.08 | 5.35 |
| Grand Est | HPV16/18 | | 2023 | | 3,377 | 4.03 | 4.03 | 4.03 | 3.32 | 4.77 | 4.03 | 4.03 | 3.72 | 4.35 |
| Hauts-de-France | HPV16/18 | | 2023 | | 8,407 | 4.09 | 4.19 | 4.19 | 3.71 | 4.67 | 4.19 | 4.19 | 3.96 | 4.43 |
| Normandie | HPV16/18 | | 2023 | | 3,659 | 4.73 | 4.80 | 4.78 | 4.04 | 5.60 | 4.81 | 4.81 | 4.49 | 5.13 |
| Nouvelle-Aquitaine | HPV16/18 | | 2023 | | 9,150 | 4.02 | 4.23 | 4.23 | 3.77 | 4.71 | 4.23 | 4.23 | 4.00 | 4.47 |
| Occitanie | HPV16/18 | | 2023 | | 1,375 | 4.44 | 4.48 | 4.51 | 3.27 | 5.75 | 4.49 | 4.49 | 4.12 | 4.88 |
| Pays de la Loire | HPV16/18 | | 2023 | | 1,501 | 3.80 | 3.51 | 3.46 | 2.60 | 4.53 | 3.52 | 3.52 | 3.19 | 3.89 |
| Provence-Alpes-Côte d'Azur | HPV16/18 | | 2023 | | 6,207 | 4.58 | 4.70 | 4.70 | 4.14 | 5.28 | 4.70 | 4.70 | 4.42 | 4.99 |
| Île-de-France | HPV16/18 | | 2023 | | 14,652 | 4.49 | 4.53 | 4.52 | 4.13 | 4.93 | 4.52 | 4.53 | 4.32 | 4.74 |
| Auvergne-Rhône-Alpes | Other genotypes | | 2020 | | 988 | 9.11 | 10.01 | 10.02 | 8.00 | 12.15 | 10.01 | 10.00 | 9.25 | 10.80 |
| Bourgogne-Franche-Comté | Other genotypes | | 2020 | | 537 | 9.50 | 8.73 | 8.75 | 6.33 | 11.17 | 8.74 | 8.73 | 8.04 | 9.50 |
| Bretagne | Other genotypes | | 2020 | | 559 | 8.77 | 8.59 | 8.59 | 6.26 | 11.09 | 8.61 | 8.61 | 7.87 | 9.40 |
| Centre-Val de Loire | Other genotypes | | 2020 | | 3,670 | 7.08 | 8.40 | 8.39 | 7.41 | 9.37 | 8.39 | 8.39 | 8.03 | 8.76 |
| Corse | Other genotypes | | 2020 | | 377 | 10.61 | 10.26 | 10.08 | 6.90 | 14.06 | 10.24 | 10.19 | 8.28 | 12.60 |
| Grand Est | Other genotypes | | 2020 | | 882 | 7.82 | 8.00 | 7.94 | 6.24 | 9.98 | 8.01 | 8.00 | 7.32 | 8.75 |
| Hauts-de-France | Other genotypes | | 2020 | | 2,840 | 8.06 | 8.56 | 8.56 | 7.46 | 9.65 | 8.55 | 8.55 | 8.17 | 8.94 |
| Normandie | Other genotypes | | 2020 | | 2,313 | 9.21 | 8.80 | 8.78 | 7.57 | 10.07 | 8.80 | 8.80 | 8.35 | 9.27 |
| Nouvelle-Aquitaine | Other genotypes | | 2020 | | 1,689 | 8.94 | 9.15 | 9.12 | 7.76 | 10.60 | 9.16 | 9.16 | 8.77 | 9.57 |
| Occitanie | Other genotypes | | 2020 | | 2,179 | 9.91 | 10.92 | 10.92 | 9.45 | 12.44 | 10.92 | 10.91 | 10.13 | 11.72 |
| Pays de la Loire | Other genotypes | | 2020 | | 1,161 | 6.63 | 7.25 | 7.24 | 5.68 | 8.96 | 7.26 | 7.25 | 6.65 | 7.92 |
| Provence-Alpes-Côte d'Azur | Other genotypes | | 2020 | | 2,131 | 9.62 | 9.41 | 9.39 | 8.12 | 10.75 | 9.41 | 9.41 | 8.91 | 9.92 |
| Île-de-France | Other genotypes | | 2020 | | 51,755 | 9.67 | 9.62 | 9.61 | 9.29 | 9.96 | 9.62 | 9.61 | 9.41 | 9.84 |
| Auvergne-Rhône-Alpes | Other genotypes | | 2021 | | 3,147 | 9.12 | 9.56 | 9.53 | 8.33 | 10.80 | 9.56 | 9.55 | 8.91 | 10.26 |
| Bourgogne-Franche-Comté | Other genotypes | | 2021 | | 1,537 | 9.24 | 8.56 | 8.52 | 6.96 | 10.21 | 8.55 | 8.55 | 7.91 | 9.25 |
| Bretagne | Other genotypes | | 2021 | | 2,374 | 7.03 | 7.97 | 7.96 | 6.74 | 9.22 | 7.94 | 7.94 | 7.40 | 8.49 |
| Centre-Val de Loire | Other genotypes | | 2021 | | 15,029 | 7.58 | 7.85 | 7.84 | 7.36 | 8.35 | 7.85 | 7.84 | 7.57 | 8.13 |
| Corse | Other genotypes | | 2021 | | 35 | 8.57 | 9.00 | 8.57 | 0.00 | 20.00 | 9.07 | 9.05 | 7.95 | 10.37 |
| Grand Est | Other genotypes | | 2021 | | 3,923 | 6.96 | 7.79 | 7.77 | 6.91 | 8.74 | 7.79 | 7.78 | 7.37 | 8.24 |
| Hauts-de-France | Other genotypes | | 2021 | | 10,748 | 7.75 | 8.27 | 8.27 | 7.68 | 8.87 | 8.27 | 8.27 | 7.99 | 8.58 |
| Normandie | Other genotypes | | 2021 | | 8,079 | 8.39 | 8.58 | 8.59 | 7.88 | 9.28 | 8.58 | 8.58 | 8.22 | 8.95 |

**Table S8 Observed HR HPV cervical infection prevalence, posterior predictive HR HPV cervical infection prevalence, and posterior expected HR HPV cervical infection prevalence, stratified by various dimensions.** ***(continued)***

| | | | | | | | Posterior distribution | | | | | | | |
|---|---|---|---|---|---|---|---|---|---|---|---|---|---|---|
| | | | | | | | Predictive prevalence | | | | Expected prevalence | | | |
| | | | | | | | | | ETI95% | | | | ETI95% | |
| Region | Genotypes | Type of screening | Year | Age | Number of tests | Observed | Average | Median | LB | UB | Average | Median | LB | UB |
| Nouvelle-Aquitaine | Other genotypes | | 2021 | | 8,917 | 8.32 | 8.39 | 8.39 | 7.74 | 9.05 | 8.39 | 8.39 | 8.09 | 8.72 |
| Occitanie | Other genotypes | | 2021 | | 5,839 | 9.71 | 9.46 | 9.45 | 8.58 | 10.34 | 9.47 | 9.46 | 9.00 | 9.95 |
| Pays de la Loire | Other genotypes | | 2021 | | 3,081 | 6.04 | 6.87 | 6.88 | 5.87 | 7.89 | 6.87 | 6.87 | 6.39 | 7.40 |
| Provence-Alpes-Côte d'Azur | Other genotypes | | 2021 | | 11,313 | 9.15 | 9.14 | 9.13 | 8.53 | 9.79 | 9.13 | 9.13 | 8.79 | 9.49 |
| Île-de-France | Other genotypes | | 2021 | | 85,160 | 9.25 | 9.06 | 9.06 | 8.82 | 9.31 | 9.06 | 9.06 | 8.90 | 9.23 |
| Auvergne-Rhône-Alpes | Other genotypes | | 2022 | | 295 | 18.31 | 10.72 | 10.51 | 7.12 | 14.58 | 10.76 | 10.75 | 9.75 | 11.78 |
| Bourgogne-Franche-Comté | Other genotypes | | 2022 | | 1,220 | 9.18 | 9.19 | 9.18 | 7.46 | 11.07 | 9.20 | 9.19 | 8.48 | 9.98 |
| Bretagne | Other genotypes | | 2022 | | 3,082 | 8.70 | 8.58 | 8.60 | 7.50 | 9.73 | 8.58 | 8.58 | 8.06 | 9.09 |
| Centre-Val de Loire | Other genotypes | | 2022 | | 7,977 | 8.65 | 8.16 | 8.16 | 7.47 | 8.83 | 8.15 | 8.15 | 7.83 | 8.47 |
| Corse | Other genotypes | | 2022 | | 13 | 23.08 | 9.62 | 7.69 | 0.00 | 30.77 | 9.43 | 9.40 | 8.13 | 10.83 |
| Grand Est | Other genotypes | | 2022 | | 3,704 | 9.10 | 8.53 | 8.50 | 7.51 | 9.58 | 8.53 | 8.52 | 8.06 | 9.04 |
| Hauts-de-France | Other genotypes | | 2022 | | 9,943 | 8.90 | 8.71 | 8.72 | 8.09 | 9.34 | 8.71 | 8.71 | 8.40 | 9.03 |
| Normandie | Other genotypes | | 2022 | | 5,912 | 9.08 | 9.38 | 9.37 | 8.51 | 10.28 | 9.37 | 9.37 | 8.95 | 9.78 |
| Nouvelle-Aquitaine | Other genotypes | | 2022 | | 10,965 | 8.75 | 8.88 | 8.88 | 8.27 | 9.51 | 8.89 | 8.88 | 8.58 | 9.21 |
| Occitanie | Other genotypes | | 2022 | | 3,131 | 9.55 | 9.40 | 9.39 | 8.27 | 10.57 | 9.39 | 9.39 | 8.89 | 9.94 |
| Pays de la Loire | Other genotypes | | 2022 | | 962 | 8.42 | 7.60 | 7.59 | 6.03 | 9.36 | 7.60 | 7.59 | 7.02 | 8.22 |
| Provence-Alpes-Côte d'Azur | Other genotypes | | 2022 | | 8,238 | 9.48 | 9.86 | 9.86 | 9.12 | 10.56 | 9.87 | 9.86 | 9.49 | 10.25 |
| Île-de-France | Other genotypes | | 2022 | | 16,823 | 9.27 | 9.76 | 9.76 | 9.23 | 10.29 | 9.76 | 9.76 | 9.49 | 10.03 |
| Auvergne-Rhône-Alpes | Other genotypes | | 2023 | | 241 | 14.52 | 12.80 | 12.86 | 8.30 | 17.43 | 12.77 | 12.77 | 11.18 | 14.45 |
| Bourgogne-Franche-Comté | Other genotypes | | 2023 | | 741 | 11.07 | 9.81 | 9.85 | 7.56 | 12.15 | 9.81 | 9.80 | 8.91 | 10.79 |
| Bretagne | Other genotypes | | 2023 | | 5,047 | 9.75 | 9.41 | 9.41 | 8.46 | 10.44 | 9.41 | 9.40 | 8.89 | 9.97 |
| Centre-Val de Loire | Other genotypes | | 2023 | | 6,073 | 9.70 | 9.76 | 9.76 | 8.94 | 10.60 | 9.78 | 9.78 | 9.41 | 10.18 |
| Corse | Other genotypes | | 2023 | | 5 | 20.00 | 8.98 | 0.00 | 0.00 | 40.00 | 9.04 | 8.94 | 6.97 | 11.58 |
| Grand Est | Other genotypes | | 2023 | | 3,377 | 9.86 | 9.81 | 9.83 | 8.68 | 10.96 | 9.80 | 9.80 | 9.25 | 10.40 |
| Hauts-de-France | Other genotypes | | 2023 | | 8,407 | 10.29 | 10.09 | 10.09 | 9.36 | 10.87 | 10.09 | 10.09 | 9.71 | 10.49 |
| Normandie | Other genotypes | | 2023 | | 3,659 | 9.92 | 10.64 | 10.63 | 9.54 | 11.78 | 10.65 | 10.65 | 10.16 | 11.15 |
| Nouvelle-Aquitaine | Other genotypes | | 2023 | | 9,150 | 10.73 | 10.39 | 10.37 | 9.68 | 11.13 | 10.38 | 10.38 | 10.02 | 10.77 |
| Occitanie | Other genotypes | | 2023 | | 1,375 | 11.05 | 10.92 | 10.91 | 9.16 | 12.73 | 10.92 | 10.91 | 10.24 | 11.67 |
| Pays de la Loire | Other genotypes | | 2023 | | 1,501 | 8.59 | 8.28 | 8.26 | 6.80 | 9.93 | 8.29 | 8.28 | 7.65 | 8.99 |
| Provence-Alpes-Côte d'Azur | Other genotypes | | 2023 | | 6,207 | 11.58 | 11.40 | 11.41 | 10.47 | 12.32 | 11.41 | 11.41 | 10.96 | 11.87 |
| Île-de-France | Other genotypes | | 2023 | | 14,652 | 10.92 | 11.72 | 11.72 | 11.12 | 12.37 | 11.73 | 11.72 | 11.40 | 12.07 |
| | HPV16/18 | Opportunistic | 2020 | | 71,043 | 3.11 | 3.19 | 3.19 | 3.02 | 3.38 | 3.19 | 3.19 | 3.07 | 3.32 |
| | HPV16/18 | Organised | 2020 | | 38 | 7.89 | 3.08 | 2.63 | 0.00 | 10.53 | 3.12 | 3.09 | 2.42 | 3.98 |
| | HPV16/18 | Opportunistic | 2021 | | 154,586 | 3.67 | 3.66 | 3.66 | 3.53 | 3.79 | 3.66 | 3.66 | 3.57 | 3.75 |
| | HPV16/18 | Organised | 2021 | | 4,596 | 2.87 | 2.91 | 2.92 | 2.31 | 3.59 | 2.91 | 2.90 | 2.51 | 3.36 |
| | HPV16/18 | Opportunistic | 2022 | | 64,519 | 4.02 | 4.05 | 4.05 | 3.85 | 4.26 | 4.05 | 4.05 | 3.91 | 4.19 |
| | HPV16/18 | Organised | 2022 | | 7,746 | 2.80 | 2.89 | 2.88 | 2.41 | 3.37 | 2.90 | 2.89 | 2.62 | 3.18 |
| | HPV16/18 | Opportunistic | 2023 | | 52,894 | 4.52 | 4.53 | 4.53 | 4.29 | 4.78 | 4.53 | 4.54 | 4.37 | 4.71 |
| | HPV16/18 | Organised | 2023 | | 7,541 | 2.96 | 2.92 | 2.90 | 2.43 | 3.47 | 2.92 | 2.92 | 2.58 | 3.27 |

**Table S8** **Observed HR HPV cervical infection prevalence, posterior predictive HR HPV cervical infection prevalence, and posterior expected HR HPV cervical infection prevalence, stratified by various dimensions.** ***(continued)***

| | | | | | | | Posterior distribution | | | | | | | |
|---|---|---|---|---|---|---|---|---|---|---|---|---|---|---|
| | | | | | | | Predictive prevalence | | | | Expected prevalence | | | |
| | | | | | | | | | ETI95% | | | | ETI95% | |
| Region | Genotypes | Type of screening | Year | Age | Number of tests | Observed | Average | Median | LB | UB | Average | Median | LB | UB |
| | Other genotypes | Opportunistic | 2020 | | 71,043 | 9.37 | 9.44 | 9.44 | 9.15 | 9.74 | 9.44 | 9.44 | 9.24 | 9.66 |
| | Other genotypes | Organised | 2020 | | 38 | 5.26 | 6.87 | 5.26 | 0.00 | 15.79 | 6.96 | 6.93 | 5.75 | 8.33 |
| | Other genotypes | Opportunistic | 2021 | | 154,586 | 8.83 | 8.84 | 8.84 | 8.65 | 9.03 | 8.84 | 8.84 | 8.70 | 8.98 |
| | Other genotypes | Organised | 2021 | | 4,596 | 6.24 | 6.42 | 6.40 | 5.53 | 7.38 | 6.42 | 6.42 | 5.82 | 7.07 |
| | Other genotypes | Opportunistic | 2022 | | 64,519 | 9.31 | 9.38 | 9.38 | 9.07 | 9.69 | 9.38 | 9.37 | 9.17 | 9.58 |
| | Other genotypes | Organised | 2022 | | 7,746 | 7.18 | 7.01 | 7.00 | 6.29 | 7.73 | 7.01 | 7.00 | 6.55 | 7.48 |
| | Other genotypes | Opportunistic | 2023 | | 52,894 | 10.99 | 11.06 | 11.06 | 10.69 | 11.43 | 11.06 | 11.07 | 10.81 | 11.33 |
| | Other genotypes | Organised | 2023 | | 7,541 | 6.99 | 7.15 | 7.15 | 6.42 | 7.93 | 7.17 | 7.17 | 6.65 | 7.72 |
| Auvergne-Rhône-Alpes | HPV16/18 | | | [30,39] | | | 4.82 | 4.80 | 3.78 | 5.93 | 4.82 | 4.82 | 4.41 | 5.24 |
| Bourgogne-Franche-Comté | HPV16/18 | | | [30,39] | | | 4.42 | 4.40 | 3.38 | 5.55 | 4.42 | 4.41 | 3.98 | 4.88 |
| Bretagne | HPV16/18 | | | [30,39] | | | 4.79 | 4.80 | 4.07 | 5.58 | 4.80 | 4.79 | 4.42 | 5.20 |
| Centre-Val de Loire | HPV16/18 | | | [30,39] | | | 4.48 | 4.48 | 4.02 | 4.95 | 4.48 | 4.48 | 4.28 | 4.71 |
| Corse | HPV16/18 | | | [30,39] | | | 4.32 | 4.55 | 0.91 | 9.09 | 4.32 | 4.30 | 3.48 | 5.38 |
| Grand Est | HPV16/18 | | | [30,39] | | | 4.43 | 4.44 | 3.72 | 5.14 | 4.42 | 4.42 | 4.12 | 4.76 |
| Hauts-de-France | HPV16/18 | | | [30,39] | | | 4.40 | 4.40 | 4.01 | 4.82 | 4.40 | 4.40 | 4.18 | 4.62 |
| Normandie | HPV16/18 | | | [30,39] | | | 5.08 | 5.07 | 4.50 | 5.67 | 5.08 | 5.08 | 4.78 | 5.38 |
| Nouvelle-Aquitaine | HPV16/18 | | | [30,39] | | | 4.72 | 4.72 | 4.25 | 5.20 | 4.72 | 4.72 | 4.48 | 4.96 |
| Occitanie | HPV16/18 | | | [30,39] | | | 5.04 | 5.04 | 4.30 | 5.81 | 5.04 | 5.04 | 4.72 | 5.36 |
| Pays de la Loire | HPV16/18 | | | [30,39] | | | 3.82 | 3.82 | 3.00 | 4.68 | 3.81 | 3.81 | 3.50 | 4.14 |
| Provence-Alpes-Côte d'Azur | HPV16/18 | | | [30,39] | | | 5.02 | 5.02 | 4.53 | 5.51 | 5.02 | 5.02 | 4.76 | 5.30 |
| Île-de-France | HPV16/18 | | | [30,39] | | | 4.39 | 4.39 | 4.18 | 4.60 | 4.39 | 4.39 | 4.26 | 4.53 |
| Auvergne-Rhône-Alpes | HPV16/18 | | | (39,49] | 1,258 | 3.66 | 3.95 | 3.97 | 2.86 | 5.09 | 3.96 | 3.96 | 3.63 | 4.31 |
| Bourgogne-Franche-Comté | HPV16/18 | | | (39,49] | 1,096 | 3.19 | 3.64 | 3.65 | 2.55 | 4.93 | 3.62 | 3.62 | 3.27 | 4.02 |
| Bretagne | HPV16/18 | | | (39,49] | 2,970 | 4.18 | 3.86 | 3.84 | 3.13 | 4.68 | 3.85 | 3.85 | 3.55 | 4.19 |
| Centre-Val de Loire | HPV16/18 | | | (39,49] | 9,293 | 3.94 | 3.66 | 3.66 | 3.24 | 4.10 | 3.66 | 3.66 | 3.48 | 3.86 |
| Corse | HPV16/18 | | | (39,49] | 129 | 4.65 | 3.61 | 3.10 | 0.78 | 6.98 | 3.55 | 3.53 | 2.76 | 4.53 |
| Grand Est | HPV16/18 | | | (39,49] | 3,360 | 3.96 | 3.66 | 3.66 | 2.98 | 4.38 | 3.66 | 3.66 | 3.39 | 3.95 |
| Hauts-de-France | HPV16/18 | | | (39,49] | 8,705 | 4.24 | 3.66 | 3.66 | 3.24 | 4.10 | 3.66 | 3.66 | 3.47 | 3.86 |
| Normandie | HPV16/18 | | | (39,49] | 5,956 | 4.30 | 4.17 | 4.16 | 3.61 | 4.79 | 4.16 | 4.16 | 3.91 | 4.42 |
| Nouvelle-Aquitaine | HPV16/18 | | | (39,49] | 8,585 | 3.84 | 3.88 | 3.88 | 3.41 | 4.36 | 3.88 | 3.88 | 3.67 | 4.09 |
| Occitanie | HPV16/18 | | | (39,49] | 3,661 | 3.91 | 4.16 | 4.15 | 3.47 | 4.89 | 4.18 | 4.18 | 3.90 | 4.45 |
| Pays de la Loire | HPV16/18 | | | (39,49] | 1,993 | 2.96 | 3.16 | 3.16 | 2.36 | 4.01 | 3.17 | 3.16 | 2.91 | 3.45 |
| Provence-Alpes-Côte d'Azur | HPV16/18 | | | (39,49] | 7,655 | 4.22 | 4.08 | 4.08 | 3.59 | 4.56 | 4.08 | 4.07 | 3.85 | 4.31 |
| Île-de-France | HPV16/18 | | | (39,49] | 50,328 | 3.32 | 3.56 | 3.56 | 3.35 | 3.77 | 3.56 | 3.55 | 3.44 | 3.68 |
| Auvergne-Rhône-Alpes | HPV16/18 | | | (49,59] | 1,035 | 3.09 | 3.16 | 3.19 | 2.13 | 4.35 | 3.19 | 3.19 | 2.91 | 3.47 |
| Bourgogne-Franche-Comté | HPV16/18 | | | (49,59] | 826 | 2.42 | 2.95 | 2.91 | 1.82 | 4.24 | 2.94 | 2.93 | 2.67 | 3.25 |
| Bretagne | HPV16/18 | | | (49,59] | 2,517 | 2.86 | 3.04 | 3.02 | 2.34 | 3.77 | 3.04 | 3.04 | 2.78 | 3.31 |
| Centre-Val de Loire | HPV16/18 | | | (49,59] | 8,682 | 2.59 | 2.92 | 2.91 | 2.56 | 3.32 | 2.92 | 2.92 | 2.77 | 3.09 |
| Corse | HPV16/18 | | | (49,59] | 123 | 2.44 | 2.83 | 2.44 | 0.00 | 6.50 | 2.79 | 2.77 | 2.18 | 3.53 |

**Table S8 Observed HR HPV cervical infection prevalence, posterior predictive HR HPV cervical infection prevalence, and posterior expected HR HPV cervical infection prevalence, stratified by various dimensions.** ***(continued)***

| | | | | | | | Posterior distribution | | | | | | | |
|---|---|---|---|---|---|---|---|---|---|---|---|---|---|---|
| | | | | | | | Predictive prevalence | | | | Expected prevalence | | | |
| | | | | | | | | | ETI95% | | | | ETI95% | |
| Region | Genotypes | Type of screening | Year | Age | Number of tests | Observed | Average | Median | LB | UB | Average | Median | LB | UB |
| Grand Est | HPV16/18 | | | (49,59] | 3,050 | 2.82 | 2.94 | 2.92 | 2.33 | 3.57 | 2.93 | 2.93 | 2.70 | 3.16 |
| Hauts-de-France | HPV16/18 | | | (49,59] | 6,400 | 3.23 | 2.91 | 2.91 | 2.48 | 3.36 | 2.91 | 2.91 | 2.75 | 3.08 |
| Normandie | HPV16/18 | | | (49,59] | 4,427 | 3.50 | 3.27 | 3.25 | 2.71 | 3.86 | 3.27 | 3.26 | 3.06 | 3.48 |
| Nouvelle-Aquitaine | HPV16/18 | | | (49,59] | 7,434 | 3.12 | 3.05 | 3.04 | 2.65 | 3.47 | 3.05 | 3.05 | 2.89 | 3.24 |
| Occitanie | HPV16/18 | | | (49,59] | 3,221 | 3.51 | 3.35 | 3.35 | 2.73 | 4.04 | 3.36 | 3.36 | 3.13 | 3.60 |
| Pays de la Loire | HPV16/18 | | | (49,59] | 1,688 | 2.31 | 2.58 | 2.55 | 1.84 | 3.44 | 2.58 | 2.58 | 2.36 | 2.82 |
| Provence-Alpes-Côte d'Azur | HPV16/18 | | | (49,59] | 6,437 | 3.08 | 3.24 | 3.23 | 2.77 | 3.76 | 3.25 | 3.24 | 3.05 | 3.45 |
| Île-de-France | HPV16/18 | | | (49,59] | 37,984 | 2.79 | 2.84 | 2.83 | 2.64 | 3.05 | 2.84 | 2.84 | 2.72 | 2.95 |
| Auvergne-Rhône-Alpes | HPV16/18 | | | (59,66] | 524 | 1.34 | 2.96 | 2.86 | 1.53 | 4.58 | 2.96 | 2.95 | 2.68 | 3.25 |
| Bourgogne-Franche-Comté | HPV16/18 | | | (59,66] | 546 | 3.85 | 2.67 | 2.56 | 1.47 | 4.03 | 2.69 | 2.68 | 2.41 | 2.98 |
| Bretagne | HPV16/18 | | | (59,66] | 1,451 | 2.83 | 2.72 | 2.69 | 1.93 | 3.59 | 2.72 | 2.72 | 2.46 | 3.01 |
| Centre-Val de Loire | HPV16/18 | | | (59,66] | 4,471 | 2.75 | 2.68 | 2.68 | 2.17 | 3.20 | 2.68 | 2.68 | 2.51 | 2.88 |
| Corse | HPV16/18 | | | (59,66] | 68 | 1.47 | 2.72 | 2.94 | 0.00 | 7.35 | 2.72 | 2.69 | 2.20 | 3.37 |
| Grand Est | HPV16/18 | | | (59,66] | 1,602 | 2.31 | 2.71 | 2.68 | 1.87 | 3.56 | 2.71 | 2.71 | 2.49 | 2.95 |
| Hauts-de-France | HPV16/18 | | | (59,66] | 3,095 | 2.42 | 2.71 | 2.71 | 2.13 | 3.33 | 2.72 | 2.72 | 2.54 | 2.91 |
| Normandie | HPV16/18 | | | (59,66] | 2,387 | 3.02 | 2.97 | 2.97 | 2.30 | 3.69 | 2.96 | 2.96 | 2.74 | 3.20 |
| Nouvelle-Aquitaine | HPV16/18 | | | (59,66] | 4,321 | 3.03 | 2.83 | 2.82 | 2.31 | 3.40 | 2.83 | 2.83 | 2.64 | 3.04 |
| Occitanie | HPV16/18 | | | (59,66] | 1,714 | 3.33 | 3.11 | 3.09 | 2.28 | 4.03 | 3.11 | 3.11 | 2.88 | 3.36 |
| Pays de la Loire | HPV16/18 | | | (59,66] | 888 | 2.48 | 2.52 | 2.48 | 1.58 | 3.60 | 2.51 | 2.50 | 2.27 | 2.77 |
| Provence-Alpes-Côte d'Azur | HPV16/18 | | | (59,66] | 3,201 | 2.94 | 3.02 | 3.00 | 2.41 | 3.66 | 3.02 | 3.01 | 2.80 | 3.24 |
| Île-de-France | HPV16/18 | | | (59,66] | 18,088 | 2.71 | 2.65 | 2.65 | 2.39 | 2.92 | 2.64 | 2.64 | 2.50 | 2.80 |
| Auvergne-Rhône-Alpes | Other genotypes | | | [30,39] | | | 12.73 | 12.73 | 11.11 | 14.46 | 12.73 | 12.72 | 11.88 | 13.60 |
| Bourgogne-Franche-Comté | Other genotypes | | | [30,39] | | | 11.66 | 11.61 | 9.83 | 13.53 | 11.67 | 11.67 | 10.82 | 12.65 |
| Bretagne | Other genotypes | | | [30,39] | | | 11.65 | 11.64 | 10.50 | 12.88 | 11.63 | 11.63 | 10.99 | 12.31 |
| Centre-Val de Loire | Other genotypes | | | [30,39] | | | 11.26 | 11.26 | 10.54 | 11.95 | 11.26 | 11.26 | 10.88 | 11.64 |
| Corse | Other genotypes | | | [30,39] | | | 13.54 | 13.64 | 7.27 | 20.00 | 13.59 | 13.53 | 11.37 | 16.20 |
| Grand Est | Other genotypes | | | [30,39] | | | 11.38 | 11.38 | 10.22 | 12.52 | 11.38 | 11.37 | 10.80 | 12.01 |
| Hauts-de-France | Other genotypes | | | [30,39] | | | 11.20 | 11.20 | 10.56 | 11.84 | 11.20 | 11.20 | 10.84 | 11.58 |
| Normandie | Other genotypes | | | [30,39] | | | 12.09 | 12.08 | 11.23 | 13.00 | 12.08 | 12.08 | 11.59 | 12.58 |
| Nouvelle-Aquitaine | Other genotypes | | | [30,39] | | | 12.23 | 12.22 | 11.47 | 12.99 | 12.23 | 12.22 | 11.83 | 12.64 |
| Occitanie | Other genotypes | | | [30,39] | | | 13.04 | 13.03 | 11.84 | 14.23 | 13.04 | 13.03 | 12.44 | 13.68 |
| Pays de la Loire | Other genotypes | | | [30,39] | | | 9.59 | 9.60 | 8.19 | 11.05 | 9.61 | 9.60 | 8.98 | 10.26 |
| Provence-Alpes-Côte d'Azur | Other genotypes | | | [30,39] | | | 12.87 | 12.86 | 12.11 | 13.66 | 12.87 | 12.88 | 12.44 | 13.31 |
| Île-de-France | Other genotypes | | | [30,39] | | | 12.50 | 12.50 | 12.16 | 12.85 | 12.50 | 12.50 | 12.29 | 12.73 |
| Auvergne-Rhône-Alpes | Other genotypes | | | (39,49] | 1,258 | 8.43 | 8.89 | 8.90 | 7.23 | 10.65 | 8.89 | 8.88 | 8.28 | 9.56 |
| Bourgogne-Franche-Comté | Other genotypes | | | (39,49] | 1,096 | 8.67 | 8.14 | 8.12 | 6.39 | 9.95 | 8.12 | 8.11 | 7.49 | 8.79 |
| Bretagne | Other genotypes | | | (39,49] | 2,970 | 8.96 | 8.05 | 8.01 | 7.00 | 9.19 | 8.05 | 8.05 | 7.60 | 8.53 |
| Centre-Val de Loire | Other genotypes | | | (39,49] | 9,293 | 7.95 | 7.85 | 7.86 | 7.23 | 8.47 | 7.85 | 7.85 | 7.57 | 8.14 |
| Corse | Other genotypes | | | (39,49] | 129 | 14.73 | 10.15 | 10.08 | 5.43 | 16.28 | 10.10 | 10.05 | 8.19 | 12.40 |

**Table S8 Observed HR HPV cervical infection prevalence, posterior predictive HR HPV cervical infection prevalence, and posterior expected HR HPV cervical infection prevalence, stratified by various dimensions.** ***(continued)***

| | | | | | | | Posterior distribution | | | | | | | |
|---|---|---|---|---|---|---|---|---|---|---|---|---|---|---|
| | | | | | | | Predictive prevalence | | | | Expected prevalence | | | |
| | | | | | | | | | ETI95% | | | | ETI95% | |
| Region | Genotypes | Type of screening | Year | Age | Number of tests | Observed | Average | Median | LB | UB | Average | Median | LB | UB |
| Grand Est | Other genotypes | | | (39,49] | 3,360 | 8.96 | 8.10 | 8.10 | 7.14 | 9.17 | 8.11 | 8.11 | 7.69 | 8.60 |
| Hauts-de-France | Other genotypes | | | (39,49] | 8,705 | 8.65 | 7.92 | 7.93 | 7.27 | 8.57 | 7.92 | 7.92 | 7.63 | 8.21 |
| Normandie | Other genotypes | | | (39,49] | 5,956 | 8.28 | 8.52 | 8.51 | 7.72 | 9.35 | 8.52 | 8.52 | 8.15 | 8.90 |
| Nouvelle-Aquitaine | Other genotypes | | | (39,49] | 8,585 | 8.69 | 8.60 | 8.60 | 7.97 | 9.27 | 8.61 | 8.60 | 8.31 | 8.92 |
| Occitanie | Other genotypes | | | (39,49] | 3,661 | 9.01 | 9.35 | 9.34 | 8.36 | 10.41 | 9.35 | 9.35 | 8.90 | 9.84 |
| Pays de la Loire | Other genotypes | | | (39,49] | 1,993 | 6.67 | 6.92 | 6.92 | 5.72 | 8.13 | 6.91 | 6.91 | 6.45 | 7.41 |
| Provence-Alpes-Côte d'Azur | Other genotypes | | | (39,49] | 7,655 | 8.83 | 8.96 | 8.95 | 8.27 | 9.68 | 8.96 | 8.96 | 8.62 | 9.31 |
| Île-de-France | Other genotypes | | | (39,49] | 50,328 | 8.31 | 8.65 | 8.65 | 8.35 | 8.96 | 8.65 | 8.65 | 8.47 | 8.84 |
| Auvergne-Rhône-Alpes | Other genotypes | | | (49,59] | 1,035 | 7.63 | 7.62 | 7.63 | 5.89 | 9.37 | 7.63 | 7.61 | 7.09 | 8.20 |
| Bourgogne-Franche-Comté | Other genotypes | | | (49,59] | 826 | 7.38 | 6.96 | 6.90 | 5.21 | 8.72 | 6.96 | 6.96 | 6.43 | 7.52 |
| Bretagne | Other genotypes | | | (49,59] | 2,517 | 6.87 | 6.79 | 6.79 | 5.72 | 7.87 | 6.78 | 6.78 | 6.39 | 7.19 |
| Centre-Val de Loire | Other genotypes | | | (49,59] | 8,682 | 6.15 | 6.66 | 6.66 | 6.07 | 7.24 | 6.66 | 6.66 | 6.40 | 6.92 |
| Corse | Other genotypes | | | (49,59] | 123 | 5.69 | 8.46 | 8.13 | 3.25 | 13.82 | 8.40 | 8.35 | 6.81 | 10.29 |
| Grand Est | Other genotypes | | | (49,59] | 3,050 | 6.00 | 6.90 | 6.89 | 5.93 | 7.87 | 6.90 | 6.90 | 6.53 | 7.32 |
| Hauts-de-France | Other genotypes | | | (49,59] | 6,400 | 7.58 | 6.71 | 6.70 | 6.05 | 7.39 | 6.72 | 6.72 | 6.47 | 6.99 |
| Normandie | Other genotypes | | | (49,59] | 4,427 | 6.44 | 7.11 | 7.12 | 6.28 | 7.95 | 7.11 | 7.11 | 6.80 | 7.44 |
| Nouvelle-Aquitaine | Other genotypes | | | (49,59] | 7,434 | 7.37 | 7.24 | 7.24 | 6.63 | 7.90 | 7.24 | 7.24 | 6.97 | 7.52 |
| Occitanie | Other genotypes | | | (49,59] | 3,221 | 8.20 | 7.99 | 7.98 | 7.02 | 9.03 | 7.99 | 7.99 | 7.60 | 8.41 |
| Pays de la Loire | Other genotypes | | | (49,59] | 1,688 | 6.10 | 5.97 | 5.98 | 4.80 | 7.23 | 5.98 | 5.97 | 5.58 | 6.43 |
| Provence-Alpes-Côte d'Azur | Other genotypes | | | (49,59] | 6,437 | 8.16 | 7.57 | 7.57 | 6.87 | 8.28 | 7.58 | 7.58 | 7.27 | 7.89 |
| Île-de-France | Other genotypes | | | (49,59] | 37,984 | 7.22 | 7.32 | 7.32 | 7.01 | 7.65 | 7.32 | 7.32 | 7.14 | 7.52 |
| Auvergne-Rhône-Alpes | Other genotypes | | | (59,66] | 524 | 6.11 | 6.77 | 6.68 | 4.58 | 8.97 | 6.80 | 6.79 | 6.29 | 7.34 |
| Bourgogne-Franche-Comté | Other genotypes | | | (59,66] | 546 | 8.06 | 6.19 | 6.23 | 4.21 | 8.42 | 6.21 | 6.20 | 5.71 | 6.76 |
| Bretagne | Other genotypes | | | (59,66] | 1,451 | 5.86 | 5.93 | 5.93 | 4.69 | 7.31 | 5.94 | 5.94 | 5.53 | 6.36 |
| Centre-Val de Loire | Other genotypes | | | (59,66] | 4,471 | 6.08 | 5.90 | 5.90 | 5.14 | 6.64 | 5.90 | 5.90 | 5.61 | 6.17 |
| Corse | Other genotypes | | | (59,66] | 68 | 2.94 | 7.56 | 7.35 | 1.47 | 14.71 | 7.59 | 7.55 | 6.32 | 9.10 |
| Grand Est | Other genotypes | | | (59,66] | 1,602 | 4.62 | 6.21 | 6.18 | 4.99 | 7.55 | 6.19 | 6.18 | 5.81 | 6.57 |
| Hauts-de-France | Other genotypes | | | (59,66] | 3,095 | 6.20 | 6.07 | 6.04 | 5.23 | 6.98 | 6.07 | 6.07 | 5.78 | 6.36 |
| Normandie | Other genotypes | | | (59,66] | 2,387 | 5.70 | 6.24 | 6.24 | 5.24 | 7.25 | 6.25 | 6.24 | 5.91 | 6.58 |
| Nouvelle-Aquitaine | Other genotypes | | | (59,66] | 4,321 | 6.23 | 6.51 | 6.50 | 5.72 | 7.31 | 6.51 | 6.51 | 6.21 | 6.84 |
| Occitanie | Other genotypes | | | (59,66] | 1,714 | 7.29 | 7.16 | 7.18 | 5.89 | 8.40 | 7.18 | 7.17 | 6.78 | 7.60 |
| Pays de la Loire | Other genotypes | | | (59,66] | 888 | 4.73 | 5.58 | 5.52 | 4.05 | 7.21 | 5.60 | 5.59 | 5.17 | 6.09 |
| Provence-Alpes-Côte d'Azur | Other genotypes | | | (59,66] | 3,201 | 7.19 | 6.78 | 6.78 | 5.87 | 7.75 | 6.78 | 6.78 | 6.44 | 7.12 |
| Île-de-France | Other genotypes | | | (59,66] | 18,088 | 6.58 | 6.47 | 6.47 | 6.04 | 6.91 | 6.47 | 6.47 | 6.23 | 6.71 |
| | HPV16/18 | Opportunistic | | [30,39] | | | 4.58 | 4.58 | 4.42 | 4.74 | 4.58 | 4.58 | 4.47 | 4.69 |
| | HPV16/18 | Organised | | [30,39] | | | 3.65 | 3.66 | 3.02 | 4.33 | 3.66 | 3.65 | 3.22 | 4.12 |
| | HPV16/18 | Opportunistic | | (39,49] | 100,422 | 3.71 | 3.74 | 3.74 | 3.58 | 3.90 | 3.74 | 3.74 | 3.63 | 3.85 |
| | HPV16/18 | Organised | | (39,49] | 4,567 | 2.93 | 3.01 | 3.00 | 2.45 | 3.59 | 3.01 | 3.00 | 2.71 | 3.31 |
| | HPV16/18 | Opportunistic | | (49,59] | 78,627 | 2.95 | 2.98 | 2.98 | 2.82 | 3.16 | 2.98 | 2.98 | 2.88 | 3.09 |

**Table S8 Observed HR HPV cervical infection prevalence, posterior predictive HR HPV cervical infection prevalence, and posterior expected HR HPV cervical infection prevalence, stratified by various dimensions. *(continued)***

| | | | | | | | Posterior distribution | | | | | | | |
|---|---|---|---|---|---|---|---|---|---|---|---|---|---|---|
| | | | | | | | Predictive prevalence | | | | Expected prevalence | | | |
| | | | | | | | | | ETI95% | | | | ETI95% | |
| Region | Genotypes | Type of screening | Year | Age | Number of tests | Observed | Average | Median | LB | UB | Average | Median | LB | UB |
| | HPV16/18 | Organised | | (49,59] | 5,197 | 2.42 | 2.49 | 2.48 | 2.02 | 3.00 | 2.50 | 2.50 | 2.25 | 2.77 |
| | HPV16/18 | Opportunistic | | (59,66] | 37,369 | 2.80 | 2.78 | 2.78 | 2.57 | 3.02 | 2.78 | 2.78 | 2.63 | 2.94 |
| | HPV16/18 | Organised | | (59,66] | 4,987 | 2.53 | 2.46 | 2.47 | 1.93 | 3.03 | 2.47 | 2.46 | 2.14 | 2.84 |
| | Other genotypes | Opportunistic | | [30,39] | | | 12.28 | 12.28 | 12.03 | 12.54 | 12.28 | 12.29 | 12.10 | 12.46 |
| | Other genotypes | Organised | | [30,39] | | | 9.02 | 8.99 | 7.99 | 10.14 | 9.04 | 9.03 | 8.35 | 9.76 |
| | Other genotypes | Opportunistic | | (39,49] | 100,422 | 8.49 | 8.57 | 8.57 | 8.34 | 8.81 | 8.57 | 8.57 | 8.40 | 8.73 |
| | Other genotypes | Organised | | (39,49] | 4,567 | 6.74 | 6.66 | 6.66 | 5.82 | 7.55 | 6.68 | 6.68 | 6.21 | 7.17 |
| | Other genotypes | Opportunistic | | (49,59] | 78,627 | 7.22 | 7.25 | 7.24 | 7.01 | 7.50 | 7.25 | 7.25 | 7.08 | 7.42 |
| | Other genotypes | Organised | | (49,59] | 5,197 | 6.02 | 6.13 | 6.12 | 5.39 | 6.89 | 6.13 | 6.13 | 5.71 | 6.58 |
| | Other genotypes | Opportunistic | | (59,66] | 37,369 | 6.43 | 6.45 | 6.45 | 6.10 | 6.80 | 6.45 | 6.45 | 6.22 | 6.70 |
| | Other genotypes | Organised | | (59,66] | 4,987 | 5.82 | 5.83 | 5.82 | 5.01 | 6.68 | 5.83 | 5.82 | 5.30 | 6.40 |
| Auvergne-Rhône-Alpes | HPV16/18 | Opportunistic | | | 4,422 | 3.78 | 4.03 | 4.03 | 3.37 | 4.75 | 4.04 | 4.04 | 3.71 | 4.38 |
| Auvergne-Rhône-Alpes | HPV16/18 | Organised | | | 249 | 4.42 | 3.60 | 3.61 | 1.61 | 6.02 | 3.61 | 3.59 | 3.10 | 4.19 |
| Bourgogne-Franche-Comté | HPV16/18 | Opportunistic | | | 3,773 | 3.74 | 3.73 | 3.74 | 3.05 | 4.45 | 3.73 | 3.72 | 3.38 | 4.11 |
| Bourgogne-Franche-Comté | HPV16/18 | Organised | | | 262 | 2.29 | 2.77 | 2.67 | 1.15 | 4.96 | 2.77 | 2.76 | 2.30 | 3.30 |
| Bretagne | HPV16/18 | Opportunistic | | | 8,765 | 4.26 | 4.22 | 4.21 | 3.70 | 4.76 | 4.22 | 4.21 | 3.89 | 4.57 |
| Bretagne | HPV16/18 | Organised | | | 2,297 | 2.44 | 2.55 | 2.53 | 1.83 | 3.40 | 2.56 | 2.55 | 2.16 | 3.04 |
| Centre-Val de Loire | HPV16/18 | Opportunistic | | | 30,545 | 3.73 | 3.65 | 3.65 | 3.39 | 3.92 | 3.65 | 3.64 | 3.49 | 3.83 |
| Centre-Val de Loire | HPV16/18 | Organised | | | 2,204 | 3.27 | 2.79 | 2.77 | 2.09 | 3.58 | 2.80 | 2.80 | 2.53 | 3.11 |
| Corse | HPV16/18 | Opportunistic | | | 427 | 3.51 | 3.43 | 3.28 | 1.64 | 5.39 | 3.40 | 3.38 | 2.70 | 4.26 |
| Corse | HPV16/18 | Organised | | | 3 | 0.00 | 2.90 | 0.00 | 0.00 | 33.33 | 3.04 | 3.01 | 2.38 | 3.84 |
| Grand Est | HPV16/18 | Opportunistic | | | 10,708 | 3.70 | 3.70 | 3.69 | 3.26 | 4.15 | 3.69 | 3.68 | 3.44 | 3.96 |
| Grand Est | HPV16/18 | Organised | | | 1,178 | 2.55 | 2.71 | 2.72 | 1.70 | 3.74 | 2.71 | 2.70 | 2.34 | 3.11 |
| Hauts-de-France | HPV16/18 | Opportunistic | | | 29,988 | 3.80 | 3.80 | 3.79 | 3.52 | 4.09 | 3.80 | 3.80 | 3.62 | 3.98 |
| Hauts-de-France | HPV16/18 | Organised | | | 1,950 | 2.46 | 2.82 | 2.82 | 2.05 | 3.64 | 2.83 | 2.82 | 2.51 | 3.17 |
| Normandie | HPV16/18 | Opportunistic | | | 19,589 | 4.36 | 4.18 | 4.18 | 3.81 | 4.55 | 4.18 | 4.17 | 3.94 | 4.41 |
| Normandie | HPV16/18 | Organised | | | 374 | 2.14 | 2.83 | 2.67 | 1.34 | 4.81 | 2.82 | 2.82 | 2.46 | 3.20 |
| Nouvelle-Aquitaine | HPV16/18 | Opportunistic | | | 26,985 | 3.90 | 3.94 | 3.94 | 3.64 | 4.25 | 3.94 | 3.94 | 3.75 | 4.13 |
| Nouvelle-Aquitaine | HPV16/18 | Organised | | | 3,736 | 2.76 | 2.88 | 2.86 | 2.27 | 3.51 | 2.89 | 2.88 | 2.59 | 3.22 |
| Occitanie | HPV16/18 | Opportunistic | | | 11,511 | 4.20 | 4.15 | 4.14 | 3.71 | 4.60 | 4.15 | 4.15 | 3.90 | 4.41 |
| Occitanie | HPV16/18 | Organised | | | 1,013 | 2.57 | 3.38 | 3.36 | 2.27 | 4.64 | 3.39 | 3.38 | 3.00 | 3.82 |
| Pays de la Loire | HPV16/18 | Opportunistic | | | 5,691 | 3.09 | 3.25 | 3.25 | 2.72 | 3.80 | 3.24 | 3.24 | 2.98 | 3.52 |
| Pays de la Loire | HPV16/18 | Organised | | | 1,014 | 2.37 | 2.55 | 2.56 | 1.58 | 3.65 | 2.55 | 2.54 | 2.22 | 2.89 |
| Provence-Alpes-Côte d'Azur | HPV16/18 | Opportunistic | | | 26,765 | 4.09 | 4.14 | 4.14 | 3.82 | 4.46 | 4.14 | 4.13 | 3.93 | 4.36 |
| Provence-Alpes-Côte d'Azur | HPV16/18 | Organised | | | 1,124 | 4.63 | 3.76 | 3.74 | 2.58 | 5.07 | 3.75 | 3.74 | 3.22 | 4.37 |
| Île-de-France | HPV16/18 | Opportunistic | | | 163,873 | 3.56 | 3.62 | 3.62 | 3.50 | 3.75 | 3.62 | 3.62 | 3.53 | 3.71 |
| Île-de-France | HPV16/18 | Organised | | | 4,517 | 3.08 | 2.98 | 2.99 | 2.44 | 3.59 | 2.98 | 2.97 | 2.68 | 3.32 |
| Auvergne-Rhône-Alpes | Other genotypes | Opportunistic | | | 4,422 | 9.93 | 10.00 | 10.00 | 8.89 | 11.13 | 10.00 | 9.98 | 9.33 | 10.71 |
| Auvergne-Rhône-Alpes | Other genotypes | Organised | | | 249 | 10.84 | 8.07 | 8.03 | 4.82 | 11.65 | 8.14 | 8.13 | 7.13 | 9.15 |

**Table S8** **Observed HR HPV cervical infection prevalence, posterior predictive HR HPV cervical infection prevalence, and posterior expected HR HPV cervical infection prevalence, stratified by various dimensions.** ***(continued)***

| | | | | | | | Posterior distribution | | | | | | | |
|---|---|---|---|---|---|---|---|---|---|---|---|---|---|---|
| | | | | | | | Predictive prevalence | | | | Expected prevalence | | | |
| | | | | | | | | | ETI95% | | | | ETI95% | |
| Region | Genotypes | Type of screening | Year | Age | Number of tests | Observed | Average | Median | LB | UB | Average | Median | LB | UB |
| Bourgogne-Franche-Comté | Other genotypes | Opportunistic | | | 3,773 | 9.78 | 9.16 | 9.14 | 8.03 | 10.36 | 9.16 | 9.15 | 8.48 | 9.90 |
| Bourgogne-Franche-Comté | Other genotypes | Organised | | | 262 | 6.87 | 6.76 | 6.87 | 3.82 | 9.92 | 6.81 | 6.79 | 5.84 | 7.92 |
| Bretagne | Other genotypes | Opportunistic | | | 8,765 | 9.82 | 9.57 | 9.57 | 8.73 | 10.41 | 9.57 | 9.57 | 9.00 | 10.16 |
| Bretagne | Other genotypes | Organised | | | 2,297 | 5.01 | 5.98 | 5.96 | 4.75 | 7.27 | 5.98 | 5.96 | 5.21 | 6.86 |
| Centre-Val de Loire | Other genotypes | Opportunistic | | | 30,545 | 8.30 | 8.48 | 8.47 | 8.08 | 8.89 | 8.48 | 8.48 | 8.20 | 8.76 |
| Centre-Val de Loire | Other genotypes | Organised | | | 2,204 | 6.44 | 6.39 | 6.35 | 5.31 | 7.53 | 6.39 | 6.38 | 5.90 | 6.92 |
| Corse | Other genotypes | Opportunistic | | | 427 | 11.01 | 10.15 | 10.07 | 7.03 | 13.82 | 10.13 | 10.09 | 8.37 | 12.27 |
| Corse | Other genotypes | Organised | | | 3 | 0.00 | 6.46 | 0.00 | 0.00 | 33.33 | 6.67 | 6.63 | 5.35 | 8.25 |
| Grand Est | Other genotypes | Opportunistic | | | 10,708 | 8.78 | 8.84 | 8.84 | 8.13 | 9.55 | 8.84 | 8.84 | 8.39 | 9.34 |
| Grand Est | Other genotypes | Organised | | | 1,178 | 6.11 | 6.47 | 6.45 | 5.01 | 8.06 | 6.48 | 6.47 | 5.76 | 7.24 |
| Hauts-de-France | Other genotypes | Opportunistic | | | 29,988 | 8.97 | 9.06 | 9.06 | 8.62 | 9.50 | 9.06 | 9.07 | 8.77 | 9.36 |
| Hauts-de-France | Other genotypes | Organised | | | 1,950 | 6.26 | 6.61 | 6.56 | 5.38 | 7.90 | 6.61 | 6.60 | 6.03 | 7.23 |
| Normandie | Other genotypes | Opportunistic | | | 19,589 | 9.03 | 9.27 | 9.27 | 8.71 | 9.83 | 9.27 | 9.27 | 8.91 | 9.64 |
| Normandie | Other genotypes | Organised | | | 374 | 5.88 | 6.51 | 6.42 | 4.01 | 9.09 | 6.49 | 6.49 | 5.83 | 7.20 |
| Nouvelle-Aquitaine | Other genotypes | Opportunistic | | | 26,985 | 9.48 | 9.49 | 9.49 | 9.02 | 9.98 | 9.49 | 9.49 | 9.20 | 9.82 |
| Nouvelle-Aquitaine | Other genotypes | Organised | | | 3,736 | 7.39 | 7.10 | 7.09 | 6.18 | 8.11 | 7.11 | 7.11 | 6.60 | 7.69 |
| Occitanie | Other genotypes | Opportunistic | | | 11,511 | 9.99 | 10.03 | 10.03 | 9.32 | 10.74 | 10.03 | 10.03 | 9.58 | 10.52 |
| Occitanie | Other genotypes | Organised | | | 1,013 | 8.29 | 7.88 | 7.90 | 6.12 | 9.77 | 7.90 | 7.88 | 7.16 | 8.70 |
| Pays de la Loire | Other genotypes | Opportunistic | | | 5,691 | 7.43 | 7.63 | 7.63 | 6.78 | 8.52 | 7.63 | 7.62 | 7.14 | 8.18 |
| Pays de la Loire | Other genotypes | Organised | | | 1,014 | 4.93 | 5.84 | 5.82 | 4.34 | 7.50 | 5.84 | 5.83 | 5.24 | 6.49 |
| Provence-Alpes-Côte d'Azur | Other genotypes | Opportunistic | | | 26,765 | 9.84 | 9.90 | 9.90 | 9.42 | 10.40 | 9.90 | 9.91 | 9.57 | 10.24 |
| Provence-Alpes-Côte d'Azur | Other genotypes | Organised | | | 1,124 | 9.34 | 9.19 | 9.16 | 7.21 | 11.30 | 9.23 | 9.20 | 8.18 | 10.50 |
| Île-de-France | Other genotypes | Opportunistic | | | 163,873 | 9.58 | 9.60 | 9.60 | 9.40 | 9.80 | 9.60 | 9.60 | 9.46 | 9.75 |
| Île-de-France | Other genotypes | Organised | | | 4,517 | 7.50 | 7.23 | 7.22 | 6.31 | 8.19 | 7.24 | 7.23 | 6.72 | 7.82 |
| Auvergne-Rhône-Alpes | HPV16/18 | | 2020 | [30,39] | | | 4.14 | 4.10 | 2.41 | 6.27 | 4.14 | 4.13 | 3.73 | 4.58 |
| Bourgogne-Franche-Comté | HPV16/18 | | 2020 | [30,39] | | | 3.69 | 3.86 | 1.45 | 6.76 | 3.66 | 3.66 | 3.26 | 4.11 |
| Bretagne | HPV16/18 | | 2020 | [30,39] | | | 3.89 | 3.93 | 1.75 | 6.55 | 3.88 | 3.87 | 3.46 | 4.33 |
| Centre-Val de Loire | HPV16/18 | | 2020 | [30,39] | | | 3.79 | 3.77 | 2.68 | 4.94 | 3.79 | 3.79 | 3.55 | 4.05 |
| Corse | HPV16/18 | | 2020 | [30,39] | | | 4.22 | 4.35 | 1.09 | 8.70 | 4.22 | 4.20 | 3.24 | 5.42 |
| Grand Est | HPV16/18 | | 2020 | [30,39] | | | 3.62 | 3.40 | 1.28 | 6.38 | 3.58 | 3.57 | 3.21 | 3.96 |
| Hauts-de-France | HPV16/18 | | 2020 | [30,39] | | | 3.58 | 3.56 | 2.54 | 4.75 | 3.59 | 3.58 | 3.34 | 3.83 |
| Normandie | HPV16/18 | | 2020 | [30,39] | | | 4.05 | 4.09 | 2.81 | 5.50 | 4.06 | 4.06 | 3.75 | 4.37 |
| Nouvelle-Aquitaine | HPV16/18 | | 2020 | [30,39] | | | 3.82 | 3.80 | 2.38 | 5.39 | 3.84 | 3.84 | 3.57 | 4.10 |
| Occitanie | HPV16/18 | | 2020 | [30,39] | | | 4.76 | 4.68 | 3.16 | 6.46 | 4.77 | 4.76 | 4.31 | 5.26 |
| Pays de la Loire | HPV16/18 | | 2020 | [30,39] | | | 3.19 | 3.24 | 1.62 | 5.14 | 3.23 | 3.22 | 2.89 | 3.61 |
| Provence-Alpes-Côte d'Azur | HPV16/18 | | 2020 | [30,39] | | | 3.96 | 3.91 | 2.65 | 5.42 | 3.96 | 3.96 | 3.67 | 4.28 |
| Île-de-France | HPV16/18 | | 2020 | [30,39] | | | 3.84 | 3.84 | 3.55 | 4.17 | 3.85 | 3.85 | 3.68 | 4.02 |
| Auvergne-Rhône-Alpes | HPV16/18 | | 2020 | (39,49] | 262 | 0.76 | 3.37 | 3.44 | 1.52 | 5.73 | 3.38 | 3.37 | 3.04 | 3.74 |
| Bourgogne-Franche-Comté | HPV16/18 | | 2020 | (39,49] | 144 | 2.08 | 3.02 | 2.78 | 0.69 | 6.25 | 2.99 | 2.99 | 2.64 | 3.36 |

**Table S8** **Observed HR HPV cervical infection prevalence, posterior predictive HR HPV cervical infection prevalence, and posterior expected HR HPV cervical infection prevalence, stratified by various dimensions.** ***(continued)***

| | | | | | | | Posterior distribution | | | | | | | |
|---|---|---|---|---|---|---|---|---|---|---|---|---|---|---|
| | | | | | | | Predictive prevalence | | | | Expected prevalence | | | |
| | | | | | | | | | ETI95% | | | | ETI95% | |
| Region | Genotypes | Type of screening | Year | Age | Number of tests | Observed | Average | Median | LB | UB | Average | Median | LB | UB |
| Bretagne | HPV16/18 | | 2020 | (39,49] | 158 | 5.06 | 3.20 | 3.16 | 0.63 | 6.33 | 3.20 | 3.19 | 2.82 | 3.62 |
| Centre-Val de Loire | HPV16/18 | | 2020 | (39,49] | 1,001 | 2.50 | 3.12 | 3.10 | 2.10 | 4.30 | 3.13 | 3.14 | 2.92 | 3.36 |
| Corse | HPV16/18 | | 2020 | (39,49] | 118 | 3.39 | 3.56 | 3.39 | 0.85 | 7.63 | 3.50 | 3.47 | 2.66 | 4.54 |
| Grand Est | HPV16/18 | | 2020 | (39,49] | 241 | 2.90 | 2.90 | 2.90 | 0.83 | 5.39 | 2.89 | 2.88 | 2.57 | 3.23 |
| Hauts-de-France | HPV16/18 | | 2020 | (39,49] | 759 | 3.82 | 2.97 | 2.90 | 1.84 | 4.22 | 2.99 | 2.99 | 2.78 | 3.19 |
| Normandie | HPV16/18 | | 2020 | (39,49] | 675 | 3.56 | 3.26 | 3.26 | 1.93 | 4.74 | 3.25 | 3.24 | 2.99 | 3.50 |
| Nouvelle-Aquitaine | HPV16/18 | | 2020 | (39,49] | 424 | 2.83 | 3.20 | 3.07 | 1.65 | 4.95 | 3.20 | 3.20 | 2.96 | 3.43 |
| Occitanie | HPV16/18 | | 2020 | (39,49] | 628 | 3.66 | 4.05 | 3.98 | 2.55 | 5.73 | 4.05 | 4.05 | 3.64 | 4.49 |
| Pays de la Loire | HPV16/18 | | 2020 | (39,49] | 370 | 1.89 | 2.67 | 2.70 | 1.08 | 4.59 | 2.66 | 2.66 | 2.38 | 2.98 |
| Provence-Alpes-Côte d'Azur | HPV16/18 | | 2020 | (39,49] | 620 | 3.71 | 3.19 | 3.23 | 1.94 | 4.68 | 3.18 | 3.18 | 2.92 | 3.45 |
| Île-de-France | HPV16/18 | | 2020 | (39,49] | 16,220 | 2.78 | 3.15 | 3.15 | 2.85 | 3.48 | 3.15 | 3.16 | 3.01 | 3.30 |
| Auvergne-Rhône-Alpes | HPV16/18 | | 2020 | (49,59] | 215 | 4.65 | 2.68 | 2.79 | 0.93 | 5.12 | 2.74 | 2.73 | 2.45 | 3.04 |
| Bourgogne-Franche-Comté | HPV16/18 | | 2020 | (49,59] | 98 | 1.02 | 2.55 | 2.04 | 0.00 | 6.12 | 2.48 | 2.48 | 2.22 | 2.78 |
| Bretagne | HPV16/18 | | 2020 | (49,59] | 107 | 1.87 | 2.51 | 2.80 | 0.00 | 5.61 | 2.51 | 2.50 | 2.20 | 2.86 |
| Centre-Val de Loire | HPV16/18 | | 2020 | (49,59] | 973 | 1.64 | 2.52 | 2.47 | 1.64 | 3.60 | 2.52 | 2.52 | 2.35 | 2.70 |
| Corse | HPV16/18 | | 2020 | (49,59] | 111 | 2.70 | 2.81 | 2.70 | 0.00 | 6.31 | 2.76 | 2.74 | 2.12 | 3.56 |
| Grand Est | HPV16/18 | | 2020 | (49,59] | 279 | 3.23 | 2.25 | 2.15 | 0.72 | 4.30 | 2.29 | 2.29 | 2.02 | 2.58 |
| Hauts-de-France | HPV16/18 | | 2020 | (49,59] | 626 | 2.72 | 2.34 | 2.24 | 1.28 | 3.67 | 2.35 | 2.35 | 2.18 | 2.52 |
| Normandie | HPV16/18 | | 2020 | (49,59] | 521 | 3.07 | 2.58 | 2.50 | 1.34 | 4.03 | 2.58 | 2.58 | 2.37 | 2.80 |
| Nouvelle-Aquitaine | HPV16/18 | | 2020 | (49,59] | 367 | 3.00 | 2.52 | 2.45 | 1.09 | 4.09 | 2.53 | 2.53 | 2.34 | 2.73 |
| Occitanie | HPV16/18 | | 2020 | (49,59] | 536 | 2.99 | 3.11 | 2.99 | 1.68 | 4.66 | 3.13 | 3.12 | 2.83 | 3.44 |
| Pays de la Loire | HPV16/18 | | 2020 | (49,59] | 279 | 0.72 | 2.17 | 2.15 | 0.72 | 3.94 | 2.19 | 2.19 | 1.96 | 2.45 |
| Provence-Alpes-Côte d'Azur | HPV16/18 | | 2020 | (49,59] | 509 | 1.96 | 2.51 | 2.55 | 1.18 | 3.93 | 2.52 | 2.52 | 2.32 | 2.74 |
| Île-de-France | HPV16/18 | | 2020 | (49,59] | 11,877 | 2.32 | 2.52 | 2.52 | 2.21 | 2.84 | 2.52 | 2.51 | 2.40 | 2.64 |
| Auvergne-Rhône-Alpes | HPV16/18 | | 2020 | (59,66] | 96 | 3.12 | 2.46 | 2.08 | 0.00 | 6.25 | 2.52 | 2.52 | 2.24 | 2.84 |
| Bourgogne-Franche-Comté | HPV16/18 | | 2020 | (59,66] | 88 | 4.55 | 2.34 | 2.27 | 0.00 | 5.68 | 2.31 | 2.30 | 2.04 | 2.60 |
| Bretagne | HPV16/18 | | 2020 | (59,66] | 65 | 1.54 | 2.24 | 1.54 | 0.00 | 6.15 | 2.24 | 2.23 | 1.92 | 2.59 |
| Centre-Val de Loire | HPV16/18 | | 2020 | (59,66] | 501 | 1.60 | 2.34 | 2.20 | 1.20 | 3.79 | 2.33 | 2.33 | 2.15 | 2.53 |
| Corse | HPV16/18 | | 2020 | (59,66] | 56 | 1.79 | 2.63 | 1.79 | 0.00 | 7.14 | 2.60 | 2.57 | 2.03 | 3.31 |
| Grand Est | HPV16/18 | | 2020 | (59,66] | 127 | 3.15 | 2.11 | 1.97 | 0.00 | 4.72 | 2.10 | 2.10 | 1.84 | 2.40 |
| Hauts-de-France | HPV16/18 | | 2020 | (59,66] | 276 | 2.54 | 2.15 | 2.17 | 0.72 | 3.99 | 2.17 | 2.17 | 1.99 | 2.36 |
| Normandie | HPV16/18 | | 2020 | (59,66] | 262 | 3.82 | 2.34 | 2.29 | 0.76 | 4.20 | 2.35 | 2.34 | 2.13 | 2.58 |
| Nouvelle-Aquitaine | HPV16/18 | | 2020 | (59,66] | 267 | 2.25 | 2.41 | 2.25 | 0.75 | 4.49 | 2.40 | 2.39 | 2.18 | 2.62 |
| Occitanie | HPV16/18 | | 2020 | (59,66] | 288 | 4.51 | 2.92 | 2.78 | 1.04 | 5.21 | 2.93 | 2.92 | 2.63 | 3.25 |
| Pays de la Loire | HPV16/18 | | 2020 | (59,66] | 142 | 0.70 | 2.15 | 2.11 | 0.00 | 4.93 | 2.15 | 2.15 | 1.91 | 2.43 |
| Provence-Alpes-Côte d'Azur | HPV16/18 | | 2020 | (59,66] | 209 | 1.44 | 2.39 | 2.39 | 0.48 | 4.78 | 2.40 | 2.40 | 2.18 | 2.62 |
| Île-de-France | HPV16/18 | | 2020 | (59,66] | 4,709 | 2.12 | 2.33 | 2.34 | 1.89 | 2.80 | 2.33 | 2.33 | 2.18 | 2.48 |
| Auvergne-Rhône-Alpes | HPV16/18 | | 2021 | [30,39] | | | 4.89 | 4.88 | 3.66 | 6.18 | 4.87 | 4.87 | 4.46 | 5.31 |
| Bourgogne-Franche-Comté | HPV16/18 | | 2021 | [30,39] | | | 4.31 | 4.27 | 2.73 | 5.97 | 4.32 | 4.31 | 3.89 | 4.79 |

**Table S8 Observed HR HPV cervical infection prevalence, posterior predictive HR HPV cervical infection prevalence, and posterior expected HR HPV cervical infection prevalence, stratified by various dimensions.** ***(continued)***

| | | | | | | | Posterior distribution | | | | | | | |
|---|---|---|---|---|---|---|---|---|---|---|---|---|---|---|
| | | | | | | | Predictive prevalence | | | | Expected prevalence | | | |
| | | | | | | | | | ETI95% | | | | ETI95% | |
| Region | Genotypes | Type of screening | Year | Age | Number of tests | Observed | Average | Median | LB | UB | Average | Median | LB | UB |
| Bretagne | HPV16/18 | | 2021 | [30,39] | | | 4.43 | 4.36 | 3.05 | 5.88 | 4.41 | 4.40 | 4.00 | 4.85 |
| Centre-Val de Loire | HPV16/18 | | 2021 | [30,39] | | | 4.35 | 4.35 | 3.72 | 4.98 | 4.36 | 4.36 | 4.15 | 4.60 |
| Corse | HPV16/18 | | 2021 | [30,39] | | | 4.70 | 0.00 | 0.00 | 15.38 | 4.73 | 4.70 | 3.85 | 5.80 |
| Grand Est | HPV16/18 | | 2021 | [30,39] | | | 4.20 | 4.23 | 3.00 | 5.38 | 4.18 | 4.18 | 3.87 | 4.51 |
| Hauts-de-France | HPV16/18 | | 2021 | [30,39] | | | 4.17 | 4.17 | 3.56 | 4.82 | 4.17 | 4.17 | 3.95 | 4.39 |
| Normandie | HPV16/18 | | 2021 | [30,39] | | | 4.82 | 4.79 | 3.98 | 5.67 | 4.81 | 4.80 | 4.50 | 5.11 |
| Nouvelle-Aquitaine | HPV16/18 | | 2021 | [30,39] | | | 4.41 | 4.43 | 3.65 | 5.20 | 4.40 | 4.40 | 4.16 | 4.66 |
| Occitanie | HPV16/18 | | 2021 | [30,39] | | | 5.01 | 5.01 | 3.99 | 6.09 | 5.01 | 5.01 | 4.66 | 5.36 |
| Pays de la Loire | HPV16/18 | | 2021 | [30,39] | | | 3.72 | 3.69 | 2.49 | 4.99 | 3.70 | 3.69 | 3.35 | 4.06 |
| Provence-Alpes-Côte d'Azur | HPV16/18 | | 2021 | [30,39] | | | 4.74 | 4.74 | 4.08 | 5.48 | 4.74 | 4.74 | 4.48 | 5.02 |
| Île-de-France | HPV16/18 | | 2021 | [30,39] | | | 4.42 | 4.41 | 4.14 | 4.70 | 4.42 | 4.42 | 4.27 | 4.57 |
| Auvergne-Rhône-Alpes | HPV16/18 | | 2021 | (39,49] | 852 | 3.76 | 3.98 | 3.99 | 2.70 | 5.40 | 3.98 | 3.98 | 3.64 | 4.35 |
| Bourgogne-Franche-Comté | HPV16/18 | | 2021 | (39,49] | 392 | 3.06 | 3.58 | 3.57 | 1.79 | 5.61 | 3.58 | 3.57 | 3.21 | 3.96 |
| Bretagne | HPV16/18 | | 2021 | (39,49] | 604 | 3.31 | 3.58 | 3.48 | 2.15 | 5.14 | 3.59 | 3.58 | 3.26 | 3.98 |
| Centre-Val de Loire | HPV16/18 | | 2021 | (39,49] | 4,339 | 3.13 | 3.58 | 3.57 | 3.02 | 4.17 | 3.57 | 3.57 | 3.38 | 3.78 |
| Corse | HPV16/18 | | 2021 | (39,49] | 5 | 20.00 | 3.87 | 0.00 | 0.00 | 20.00 | 3.96 | 3.95 | 3.14 | 4.93 |
| Grand Est | HPV16/18 | | 2021 | (39,49] | 1,119 | 3.31 | 3.44 | 3.40 | 2.32 | 4.56 | 3.44 | 3.44 | 3.17 | 3.72 |
| Hauts-de-France | HPV16/18 | | 2021 | (39,49] | 3,026 | 3.80 | 3.47 | 3.47 | 2.84 | 4.13 | 3.46 | 3.46 | 3.27 | 3.65 |
| Normandie | HPV16/18 | | 2021 | (39,49] | 2,315 | 4.92 | 3.92 | 3.89 | 3.11 | 4.84 | 3.92 | 3.92 | 3.66 | 4.18 |
| Nouvelle-Aquitaine | HPV16/18 | | 2021 | (39,49] | 2,362 | 3.64 | 3.59 | 3.60 | 2.88 | 4.36 | 3.60 | 3.60 | 3.38 | 3.82 |
| Occitanie | HPV16/18 | | 2021 | (39,49] | 1,748 | 4.12 | 4.09 | 4.06 | 3.15 | 5.09 | 4.10 | 4.11 | 3.82 | 4.39 |
| Pays de la Loire | HPV16/18 | | 2021 | (39,49] | 919 | 2.61 | 3.08 | 3.05 | 1.96 | 4.35 | 3.09 | 3.09 | 2.81 | 3.39 |
| Provence-Alpes-Côte d'Azur | HPV16/18 | | 2021 | (39,49] | 3,177 | 3.71 | 3.86 | 3.84 | 3.12 | 4.56 | 3.85 | 3.85 | 3.62 | 4.10 |
| Île-de-France | HPV16/18 | | 2021 | (39,49] | 25,423 | 3.43 | 3.62 | 3.62 | 3.36 | 3.87 | 3.62 | 3.62 | 3.48 | 3.75 |
| Auvergne-Rhône-Alpes | HPV16/18 | | 2021 | (49,59] | 704 | 2.13 | 3.17 | 3.12 | 1.99 | 4.55 | 3.19 | 3.19 | 2.91 | 3.50 |
| Bourgogne-Franche-Comté | HPV16/18 | | 2021 | (49,59] | 330 | 3.33 | 2.90 | 2.73 | 1.21 | 4.85 | 2.90 | 2.90 | 2.62 | 3.23 |
| Bretagne | HPV16/18 | | 2021 | (49,59] | 592 | 2.03 | 2.79 | 2.70 | 1.52 | 4.22 | 2.78 | 2.78 | 2.51 | 3.09 |
| Centre-Val de Loire | HPV16/18 | | 2021 | (49,59] | 4,068 | 2.21 | 2.84 | 2.83 | 2.31 | 3.39 | 2.84 | 2.84 | 2.68 | 3.01 |
| Corse | HPV16/18 | | 2021 | (49,59] | 10 | 0.00 | 2.90 | 0.00 | 0.00 | 20.00 | 2.95 | 2.94 | 2.54 | 3.41 |
| Grand Est | HPV16/18 | | 2021 | (49,59] | 1,062 | 2.64 | 2.79 | 2.82 | 1.79 | 3.86 | 2.77 | 2.76 | 2.55 | 3.00 |
| Hauts-de-France | HPV16/18 | | 2021 | (49,59] | 2,111 | 3.17 | 2.76 | 2.75 | 2.08 | 3.51 | 2.77 | 2.76 | 2.60 | 2.93 |
| Normandie | HPV16/18 | | 2021 | (49,59] | 1,834 | 2.89 | 3.09 | 3.05 | 2.29 | 3.98 | 3.09 | 3.09 | 2.87 | 3.31 |
| Nouvelle-Aquitaine | HPV16/18 | | 2021 | (49,59] | 2,165 | 2.86 | 2.86 | 2.86 | 2.17 | 3.60 | 2.86 | 2.86 | 2.69 | 3.03 |
| Occitanie | HPV16/18 | | 2021 | (49,59] | 1,467 | 3.89 | 3.34 | 3.34 | 2.45 | 4.36 | 3.35 | 3.35 | 3.11 | 3.60 |
| Pays de la Loire | HPV16/18 | | 2021 | (49,59] | 781 | 3.07 | 2.50 | 2.43 | 1.41 | 3.71 | 2.49 | 2.49 | 2.26 | 2.74 |
| Provence-Alpes-Côte d'Azur | HPV16/18 | | 2021 | (49,59] | 2,584 | 2.90 | 3.05 | 3.06 | 2.36 | 3.79 | 3.06 | 3.05 | 2.86 | 3.26 |
| Île-de-France | HPV16/18 | | 2021 | (49,59] | 19,159 | 2.84 | 2.88 | 2.89 | 2.62 | 3.16 | 2.88 | 2.89 | 2.75 | 3.01 |
| Auvergne-Rhône-Alpes | HPV16/18 | | 2021 | (59,66] | 361 | 0.83 | 2.95 | 2.77 | 1.39 | 4.99 | 2.94 | 2.93 | 2.66 | 3.24 |
| Bourgogne-Franche-Comté | HPV16/18 | | 2021 | (59,66] | 229 | 4.80 | 2.66 | 2.62 | 0.87 | 4.80 | 2.68 | 2.68 | 2.41 | 2.99 |

**Table S8** **Observed HR HPV cervical infection prevalence, posterior predictive HR HPV cervical infection prevalence, and posterior expected HR HPV cervical infection prevalence, stratified by various dimensions.** ***(continued)***

| | | | | | | | Posterior distribution | | | | | | | |
|---|---|---|---|---|---|---|---|---|---|---|---|---|---|---|
| | | | | | | | Predictive prevalence | | | | Expected prevalence | | | |
| | | | | | | | | | ETI95% | | | | ETI95% | |
| Region | Genotypes | Type of screening | Year | Age | Number of tests | Observed | Average | Median | LB | UB | Average | Median | LB | UB |
| Bretagne | HPV16/18 | | 2021 | (59,66] | 260 | 2.31 | 2.60 | 2.69 | 0.77 | 4.62 | 2.61 | 2.60 | 2.34 | 2.91 |
| Centre-Val de Loire | HPV16/18 | | 2021 | (59,66] | 2,161 | 2.59 | 2.63 | 2.64 | 1.94 | 3.33 | 2.64 | 2.64 | 2.46 | 2.84 |
| Corse | HPV16/18 | | 2021 | (59,66] | 7 | 0.00 | 3.00 | 0.00 | 0.00 | 14.29 | 3.03 | 3.01 | 2.44 | 3.74 |
| Grand Est | HPV16/18 | | 2021 | (59,66] | 608 | 2.14 | 2.57 | 2.47 | 1.32 | 3.95 | 2.57 | 2.57 | 2.35 | 2.80 |
| Hauts-de-France | HPV16/18 | | 2021 | (59,66] | 980 | 2.24 | 2.56 | 2.55 | 1.63 | 3.57 | 2.56 | 2.55 | 2.37 | 2.75 |
| Normandie | HPV16/18 | | 2021 | (59,66] | 968 | 2.79 | 2.86 | 2.79 | 1.86 | 4.03 | 2.85 | 2.85 | 2.62 | 3.10 |
| Nouvelle-Aquitaine | HPV16/18 | | 2021 | (59,66] | 1,407 | 2.91 | 2.69 | 2.70 | 1.85 | 3.62 | 2.69 | 2.69 | 2.50 | 2.89 |
| Occitanie | HPV16/18 | | 2021 | (59,66] | 768 | 2.86 | 3.10 | 3.12 | 1.95 | 4.30 | 3.11 | 3.10 | 2.86 | 3.37 |
| Pays de la Loire | HPV16/18 | | 2021 | (59,66] | 378 | 2.65 | 2.44 | 2.38 | 1.06 | 4.23 | 2.44 | 2.43 | 2.19 | 2.71 |
| Provence-Alpes-Côte d'Azur | HPV16/18 | | 2021 | (59,66] | 1,336 | 2.84 | 2.83 | 2.84 | 2.02 | 3.74 | 2.83 | 2.83 | 2.62 | 3.06 |
| Île-de-France | HPV16/18 | | 2021 | (59,66] | 10,265 | 2.75 | 2.68 | 2.67 | 2.33 | 3.05 | 2.68 | 2.67 | 2.52 | 2.84 |
| Auvergne-Rhône-Alpes | HPV16/18 | | 2022 | [30,39] | | | 5.44 | 5.04 | 1.68 | 10.08 | 5.48 | 5.47 | 4.93 | 6.09 |
| Bourgogne-Franche-Comté | HPV16/18 | | 2022 | [30,39] | | | 4.63 | 4.52 | 2.95 | 6.48 | 4.62 | 4.61 | 4.15 | 5.14 |
| Bretagne | HPV16/18 | | 2022 | [30,39] | | | 4.81 | 4.80 | 3.60 | 6.17 | 4.82 | 4.82 | 4.41 | 5.27 |
| Centre-Val de Loire | HPV16/18 | | 2022 | [30,39] | | | 4.54 | 4.53 | 3.72 | 5.42 | 4.54 | 4.54 | 4.29 | 4.82 |
| Corse | HPV16/18 | | 2022 | [30,39] | | | 5.11 | 0.00 | 0.00 | 25.00 | 5.19 | 5.15 | 4.27 | 6.30 |
| Grand Est | HPV16/18 | | 2022 | [30,39] | | | 4.36 | 4.32 | 3.22 | 5.56 | 4.36 | 4.35 | 4.01 | 4.72 |
| Hauts-de-France | HPV16/18 | | 2022 | [30,39] | | | 4.43 | 4.41 | 3.77 | 5.09 | 4.43 | 4.43 | 4.19 | 4.68 |
| Normandie | HPV16/18 | | 2022 | [30,39] | | | 5.36 | 5.33 | 4.40 | 6.39 | 5.35 | 5.35 | 5.02 | 5.72 |
| Nouvelle-Aquitaine | HPV16/18 | | 2022 | [30,39] | | | 4.72 | 4.72 | 4.00 | 5.47 | 4.72 | 4.72 | 4.44 | 4.98 |
| Occitanie | HPV16/18 | | 2022 | [30,39] | | | 5.08 | 5.10 | 3.64 | 6.56 | 5.07 | 5.07 | 4.69 | 5.46 |
| Pays de la Loire | HPV16/18 | | 2022 | [30,39] | | | 4.15 | 4.09 | 2.20 | 6.60 | 4.11 | 4.11 | 3.75 | 4.52 |
| Provence-Alpes-Côte d'Azur | HPV16/18 | | 2022 | [30,39] | | | 5.13 | 5.13 | 4.31 | 5.95 | 5.12 | 5.12 | 4.82 | 5.43 |
| Île-de-France | HPV16/18 | | 2022 | [30,39] | | | 4.88 | 4.89 | 4.31 | 5.47 | 4.88 | 4.88 | 4.65 | 5.11 |
| Auvergne-Rhône-Alpes | HPV16/18 | | 2022 | (39,49] | 79 | 11.39 | 4.54 | 3.80 | 1.27 | 10.13 | 4.55 | 4.54 | 3.97 | 5.21 |
| Bourgogne-Franche-Comté | HPV16/18 | | 2022 | (39,49] | 334 | 3.59 | 3.83 | 3.89 | 1.80 | 5.99 | 3.82 | 3.81 | 3.40 | 4.26 |
| Bretagne | HPV16/18 | | 2022 | (39,49] | 836 | 4.43 | 3.83 | 3.83 | 2.51 | 5.26 | 3.83 | 3.82 | 3.50 | 4.18 |
| Centre-Val de Loire | HPV16/18 | | 2022 | (39,49] | 2,327 | 4.86 | 3.70 | 3.70 | 2.92 | 4.51 | 3.71 | 3.71 | 3.50 | 3.94 |
| Corse | HPV16/18 | | 2022 | (39,49] | 5 | 0.00 | 4.42 | 0.00 | 0.00 | 20.00 | 4.12 | 4.08 | 3.27 | 5.10 |
| Grand Est | HPV16/18 | | 2022 | (39,49] | 1,056 | 4.36 | 3.64 | 3.60 | 2.56 | 4.83 | 3.63 | 3.63 | 3.33 | 3.95 |
| Hauts-de-France | HPV16/18 | | 2022 | (39,49] | 2,668 | 4.20 | 3.66 | 3.67 | 2.92 | 4.42 | 3.67 | 3.67 | 3.45 | 3.88 |
| Normandie | HPV16/18 | | 2022 | (39,49] | 1,836 | 3.81 | 4.39 | 4.36 | 3.43 | 5.39 | 4.38 | 4.38 | 4.08 | 4.67 |
| Nouvelle-Aquitaine | HPV16/18 | | 2022 | (39,49] | 3,149 | 4.16 | 3.87 | 3.87 | 3.18 | 4.57 | 3.86 | 3.86 | 3.63 | 4.10 |
| Occitanie | HPV16/18 | | 2022 | (39,49] | 881 | 3.41 | 4.15 | 4.09 | 2.84 | 5.56 | 4.16 | 4.16 | 3.84 | 4.49 |
| Pays de la Loire | HPV16/18 | | 2022 | (39,49] | 274 | 2.19 | 3.41 | 3.28 | 1.46 | 5.84 | 3.37 | 3.37 | 3.06 | 3.72 |
| Provence-Alpes-Côte d'Azur | HPV16/18 | | 2022 | (39,49] | 2,168 | 4.20 | 4.19 | 4.20 | 3.32 | 5.07 | 4.19 | 4.20 | 3.93 | 4.47 |
| Île-de-France | HPV16/18 | | 2022 | (39,49] | 4,741 | 3.67 | 3.91 | 3.92 | 3.35 | 4.51 | 3.91 | 3.90 | 3.71 | 4.11 |
| Auvergne-Rhône-Alpes | HPV16/18 | | 2022 | (49,59] | 62 | 6.45 | 3.89 | 3.23 | 0.00 | 9.68 | 3.78 | 3.77 | 3.27 | 4.37 |
| Bourgogne-Franche-Comté | HPV16/18 | | 2022 | (49,59] | 240 | 2.50 | 3.11 | 2.92 | 1.25 | 5.42 | 3.09 | 3.09 | 2.78 | 3.46 |

**Table S8** **Observed HR HPV cervical infection prevalence, posterior predictive HR HPV cervical infection prevalence, and posterior expected HR HPV cervical infection prevalence, stratified by various dimensions.** ***(continued)***

| | | | | | | | Posterior distribution | | | | | | | |
|---|---|---|---|---|---|---|---|---|---|---|---|---|---|---|
| | | | | | | | Predictive prevalence | | | | Expected prevalence | | | |
| | | | | | | | | | ETI95% | | | | ETI95% | |
| Region | Genotypes | Type of screening | Year | Age | Number of tests | Observed | Average | Median | LB | UB | Average | Median | LB | UB |
| Bretagne | HPV16/18 | | 2022 | (49,59] | 679 | 2.36 | 3.06 | 3.09 | 1.77 | 4.42 | 3.06 | 3.06 | 2.78 | 3.35 |
| Centre-Val de Loire | HPV16/18 | | 2022 | (49,59] | 2,116 | 3.54 | 2.96 | 2.93 | 2.22 | 3.73 | 2.97 | 2.97 | 2.79 | 3.17 |
| Corse | HPV16/18 | | 2022 | (49,59] | 2 | 0.00 | 3.60 | 0.00 | 0.00 | 50.00 | 3.54 | 3.49 | 2.51 | 4.83 |
| Grand Est | HPV16/18 | | 2022 | (49,59] | 887 | 2.25 | 3.02 | 3.04 | 1.92 | 4.17 | 3.02 | 3.01 | 2.75 | 3.28 |
| Hauts-de-France | HPV16/18 | | 2022 | (49,59] | 1,992 | 3.01 | 2.93 | 2.91 | 2.21 | 3.71 | 2.93 | 2.93 | 2.75 | 3.12 |
| Normandie | HPV16/18 | | 2022 | (49,59] | 1,212 | 4.21 | 3.41 | 3.38 | 2.39 | 4.54 | 3.42 | 3.42 | 3.19 | 3.67 |
| Nouvelle-Aquitaine | HPV16/18 | | 2022 | (49,59] | 2,734 | 2.89 | 3.02 | 3.00 | 2.38 | 3.69 | 3.02 | 3.02 | 2.84 | 3.21 |
| Occitanie | HPV16/18 | | 2022 | (49,59] | 851 | 2.94 | 3.37 | 3.41 | 2.23 | 4.70 | 3.37 | 3.37 | 3.11 | 3.63 |
| Pays de la Loire | HPV16/18 | | 2022 | (49,59] | 225 | 1.33 | 2.76 | 2.67 | 0.89 | 4.89 | 2.75 | 2.75 | 2.49 | 3.05 |
| Provence-Alpes-Côte d'Azur | HPV16/18 | | 2022 | (49,59] | 1,876 | 3.14 | 3.32 | 3.30 | 2.51 | 4.21 | 3.32 | 3.32 | 3.10 | 3.54 |
| Île-de-France | HPV16/18 | | 2022 | (49,59] | 3,802 | 2.66 | 3.07 | 3.05 | 2.52 | 3.63 | 3.06 | 3.06 | 2.89 | 3.23 |
| Auvergne-Rhône-Alpes | HPV16/18 | | 2022 | (59,66] | 35 | 2.86 | 3.44 | 2.86 | 0.00 | 11.43 | 3.36 | 3.35 | 2.93 | 3.86 |
| Bourgogne-Franche-Comté | HPV16/18 | | 2022 | (59,66] | 137 | 2.19 | 2.77 | 2.92 | 0.73 | 5.84 | 2.79 | 2.78 | 2.47 | 3.13 |
| Bretagne | HPV16/18 | | 2022 | (59,66] | 401 | 2.99 | 2.76 | 2.74 | 1.25 | 4.49 | 2.75 | 2.75 | 2.49 | 3.04 |
| Centre-Val de Loire | HPV16/18 | | 2022 | (59,66] | 1,060 | 3.21 | 2.71 | 2.74 | 1.70 | 3.77 | 2.71 | 2.71 | 2.51 | 2.93 |
| Corse | HPV16/18 | | 2022 | (59,66] | 2 | 0.00 | 3.98 | 0.00 | 0.00 | 50.00 | 3.96 | 3.89 | 2.74 | 5.63 |
| Grand Est | HPV16/18 | | 2022 | (59,66] | 394 | 2.54 | 2.77 | 2.79 | 1.27 | 4.57 | 2.76 | 2.75 | 2.51 | 3.02 |
| Hauts-de-France | HPV16/18 | | 2022 | (59,66] | 883 | 2.49 | 2.71 | 2.72 | 1.70 | 3.85 | 2.72 | 2.72 | 2.53 | 2.93 |
| Normandie | HPV16/18 | | 2022 | (59,66] | 705 | 2.84 | 3.09 | 3.12 | 1.84 | 4.40 | 3.07 | 3.07 | 2.83 | 3.33 |
| Nouvelle-Aquitaine | HPV16/18 | | 2022 | (59,66] | 1,480 | 2.91 | 2.79 | 2.77 | 1.96 | 3.72 | 2.80 | 2.79 | 2.58 | 3.03 |
| Occitanie | HPV16/18 | | 2022 | (59,66] | 438 | 3.42 | 3.11 | 2.97 | 1.60 | 4.79 | 3.11 | 3.11 | 2.84 | 3.39 |
| Pays de la Loire | HPV16/18 | | 2022 | (59,66] | 145 | 2.07 | 2.74 | 2.76 | 0.69 | 5.52 | 2.68 | 2.68 | 2.39 | 3.01 |
| Provence-Alpes-Côte d'Azur | HPV16/18 | | 2022 | (59,66] | 902 | 3.10 | 3.06 | 2.99 | 2.00 | 4.21 | 3.06 | 3.06 | 2.82 | 3.31 |
| Île-de-France | HPV16/18 | | 2022 | (59,66] | 1,713 | 2.57 | 2.86 | 2.86 | 2.10 | 3.68 | 2.85 | 2.85 | 2.66 | 3.05 |
| Auvergne-Rhône-Alpes | HPV16/18 | | 2023 | [30,39] | | | 6.30 | 6.67 | 2.22 | 12.22 | 6.29 | 6.27 | 5.41 | 7.25 |
| Bourgogne-Franche-Comté | HPV16/18 | | 2023 | [30,39] | | | 4.84 | 4.91 | 2.64 | 7.55 | 4.83 | 4.82 | 4.25 | 5.46 |
| Bretagne | HPV16/18 | | 2023 | [30,39] | | | 5.09 | 5.08 | 4.03 | 6.24 | 5.09 | 5.09 | 4.67 | 5.54 |
| Centre-Val de Loire | HPV16/18 | | 2023 | [30,39] | | | 5.05 | 5.02 | 4.10 | 6.03 | 5.04 | 5.04 | 4.74 | 5.35 |
| Corse | HPV16/18 | | 2023 | [30,39] | | | 5.13 | 0.00 | 0.00 | 100.00 | 5.09 | 4.93 | 2.81 | 8.48 |
| Grand Est | HPV16/18 | | 2023 | [30,39] | | | 4.91 | 4.92 | 3.69 | 6.24 | 4.91 | 4.91 | 4.52 | 5.33 |
| Hauts-de-France | HPV16/18 | | 2023 | [30,39] | | | 4.95 | 4.93 | 4.20 | 5.70 | 4.95 | 4.94 | 4.66 | 5.24 |
| Normandie | HPV16/18 | | 2023 | [30,39] | | | 5.94 | 5.92 | 4.60 | 7.40 | 5.96 | 5.96 | 5.54 | 6.38 |
| Nouvelle-Aquitaine | HPV16/18 | | 2023 | [30,39] | | | 5.20 | 5.18 | 4.39 | 6.03 | 5.20 | 5.20 | 4.90 | 5.51 |
| Occitanie | HPV16/18 | | 2023 | [30,39] | | | 5.61 | 5.47 | 3.39 | 8.07 | 5.62 | 5.62 | 5.15 | 6.11 |
| Pays de la Loire | HPV16/18 | | 2023 | [30,39] | | | 4.34 | 4.27 | 2.47 | 6.29 | 4.34 | 4.33 | 3.92 | 4.80 |
| Provence-Alpes-Côte d'Azur | HPV16/18 | | 2023 | [30,39] | | | 5.74 | 5.73 | 4.75 | 6.80 | 5.75 | 5.75 | 5.41 | 6.12 |
| Île-de-France | HPV16/18 | | 2023 | [30,39] | | | 5.45 | 5.44 | 4.84 | 6.09 | 5.45 | 5.45 | 5.18 | 5.72 |
| Auvergne-Rhône-Alpes | HPV16/18 | | 2023 | (39,49] | 65 | 4.62 | 5.15 | 4.62 | 0.00 | 10.77 | 5.19 | 5.18 | 4.40 | 6.11 |
| Bourgogne-Franche-Comté | HPV16/18 | | 2023 | (39,49] | 226 | 3.54 | 3.88 | 3.98 | 1.33 | 6.64 | 3.82 | 3.81 | 3.36 | 4.36 |

**Table S8 Observed HR HPV cervical infection prevalence, posterior predictive HR HPV cervical infection prevalence, and posterior expected HR HPV cervical infection prevalence, stratified by various dimensions.** ***(continued)***

| | | | | | | | Posterior distribution | | | | | | | |
|---|---|---|---|---|---|---|---|---|---|---|---|---|---|---|
| | | | | | | | Predictive prevalence | | | | Expected prevalence | | | |
| | | | | | | | | | ETI95% | | | | ETI95% | |
| Region | Genotypes | Type of screening | Year | Age | Number of tests | Observed | Average | Median | LB | UB | Average | Median | LB | UB |
| Bretagne | HPV16/18 | | 2023 | (39,49] | 1,372 | 4.30 | 4.07 | 4.08 | 3.06 | 5.25 | 4.06 | 4.05 | 3.72 | 4.44 |
| Centre-Val de Loire | HPV16/18 | | 2023 | (39,49] | 1,626 | 5.66 | 4.16 | 4.12 | 3.20 | 5.23 | 4.17 | 4.17 | 3.92 | 4.44 |
| Corse | HPV16/18 | | 2023 | (39,49] | 1 | 100.00 | 4.50 | 0.00 | 0.00 | 100.00 | 5.06 | 4.84 | 2.69 | 8.51 |
| Grand Est | HPV16/18 | | 2023 | (39,49] | 944 | 4.56 | 4.15 | 4.13 | 2.86 | 5.51 | 4.15 | 4.14 | 3.81 | 4.50 |
| Hauts-de-France | HPV16/18 | | 2023 | (39,49] | 2,252 | 5.02 | 4.17 | 4.17 | 3.33 | 5.02 | 4.16 | 4.16 | 3.91 | 4.42 |
| Normandie | HPV16/18 | | 2023 | (39,49] | 1,130 | 4.25 | 4.86 | 4.87 | 3.63 | 6.19 | 4.86 | 4.86 | 4.52 | 5.21 |
| Nouvelle-Aquitaine | HPV16/18 | | 2023 | (39,49] | 2,650 | 3.81 | 4.25 | 4.23 | 3.47 | 5.06 | 4.25 | 4.25 | 4.01 | 4.53 |
| Occitanie | HPV16/18 | | 2023 | (39,49] | 404 | 4.46 | 4.70 | 4.70 | 2.72 | 6.93 | 4.71 | 4.71 | 4.28 | 5.16 |
| Pays de la Loire | HPV16/18 | | 2023 | (39,49] | 430 | 5.12 | 3.62 | 3.49 | 2.09 | 5.58 | 3.62 | 3.62 | 3.27 | 4.01 |
| Provence-Alpes-Côte d'Azur | HPV16/18 | | 2023 | (39,49] | 1,690 | 5.38 | 4.68 | 4.67 | 3.73 | 5.80 | 4.67 | 4.66 | 4.37 | 4.99 |
| Île-de-France | HPV16/18 | | 2023 | (39,49] | 3,944 | 4.46 | 4.40 | 4.41 | 3.73 | 5.07 | 4.39 | 4.39 | 4.16 | 4.62 |
| Auvergne-Rhône-Alpes | HPV16/18 | | 2023 | (49,59] | 54 | 5.56 | 4.17 | 3.70 | 0.00 | 11.11 | 4.23 | 4.22 | 3.56 | 5.04 |
| Bourgogne-Franche-Comté | HPV16/18 | | 2023 | (49,59] | 158 | 1.27 | 3.07 | 3.16 | 0.63 | 5.70 | 3.06 | 3.05 | 2.69 | 3.47 |
| Bretagne | HPV16/18 | | 2023 | (49,59] | 1,139 | 3.69 | 3.21 | 3.16 | 2.19 | 4.30 | 3.21 | 3.20 | 2.92 | 3.52 |
| Centre-Val de Loire | HPV16/18 | | 2023 | (49,59] | 1,525 | 2.89 | 3.34 | 3.34 | 2.43 | 4.26 | 3.34 | 3.34 | 3.12 | 3.56 |
| Grand Est | HPV16/18 | | 2023 | (49,59] | 822 | 3.53 | 3.27 | 3.28 | 2.07 | 4.62 | 3.26 | 3.25 | 2.98 | 3.55 |
| Hauts-de-France | HPV16/18 | | 2023 | (49,59] | 1,671 | 3.77 | 3.28 | 3.23 | 2.45 | 4.19 | 3.29 | 3.28 | 3.09 | 3.51 |
| Normandie | HPV16/18 | | 2023 | (49,59] | 860 | 4.07 | 3.87 | 3.84 | 2.67 | 5.23 | 3.85 | 3.84 | 3.56 | 4.13 |
| Nouvelle-Aquitaine | HPV16/18 | | 2023 | (49,59] | 2,168 | 3.69 | 3.38 | 3.37 | 2.63 | 4.15 | 3.38 | 3.38 | 3.18 | 3.60 |
| Occitanie | HPV16/18 | | 2023 | (49,59] | 367 | 4.09 | 3.71 | 3.81 | 1.91 | 5.72 | 3.72 | 3.72 | 3.38 | 4.09 |
| Pays de la Loire | HPV16/18 | | 2023 | (49,59] | 403 | 2.48 | 2.91 | 2.73 | 1.49 | 4.71 | 2.93 | 2.93 | 2.64 | 3.26 |
| Provence-Alpes-Côte d'Azur | HPV16/18 | | 2023 | (49,59] | 1,468 | 3.68 | 3.74 | 3.75 | 2.79 | 4.77 | 3.73 | 3.73 | 3.48 | 3.99 |
| Île-de-France | HPV16/18 | | 2023 | (49,59] | 3,146 | 4.39 | 3.47 | 3.46 | 2.83 | 4.16 | 3.47 | 3.47 | 3.28 | 3.68 |
| Auvergne-Rhône-Alpes | HPV16/18 | | 2023 | (59,66] | 32 | 0.00 | 4.07 | 3.12 | 0.00 | 12.50 | 3.99 | 3.98 | 3.36 | 4.72 |
| Bourgogne-Franche-Comté | HPV16/18 | | 2023 | (59,66] | 92 | 3.26 | 2.85 | 2.17 | 0.00 | 6.52 | 2.89 | 2.89 | 2.51 | 3.32 |
| Bretagne | HPV16/18 | | 2023 | (59,66] | 725 | 3.03 | 2.79 | 2.76 | 1.66 | 4.14 | 2.79 | 2.78 | 2.48 | 3.12 |
| Centre-Val de Loire | HPV16/18 | | 2023 | (59,66] | 749 | 3.34 | 3.02 | 2.94 | 1.87 | 4.41 | 3.01 | 3.00 | 2.79 | 3.25 |
| Corse | HPV16/18 | | 2023 | (59,66] | 3 | 0.00 | 3.07 | 0.00 | 0.00 | 33.33 | 3.41 | 3.35 | 2.42 | 4.69 |
| Grand Est | HPV16/18 | | 2023 | (59,66] | 473 | 2.11 | 3.00 | 2.96 | 1.48 | 4.65 | 3.01 | 3.01 | 2.74 | 3.31 |
| Hauts-de-France | HPV16/18 | | 2023 | (59,66] | 956 | 2.51 | 3.04 | 3.03 | 1.99 | 4.18 | 3.04 | 3.03 | 2.81 | 3.28 |
| Normandie | HPV16/18 | | 2023 | (59,66] | 452 | 3.32 | 3.38 | 3.32 | 1.77 | 5.31 | 3.39 | 3.38 | 3.11 | 3.69 |
| Nouvelle-Aquitaine | HPV16/18 | | 2023 | (59,66] | 1,167 | 3.51 | 3.13 | 3.08 | 2.14 | 4.20 | 3.13 | 3.13 | 2.90 | 3.41 |
| Occitanie | HPV16/18 | | 2023 | (59,66] | 220 | 3.18 | 3.40 | 3.18 | 1.36 | 5.91 | 3.40 | 3.40 | 3.06 | 3.77 |
| Pays de la Loire | HPV16/18 | | 2023 | (59,66] | 223 | 3.59 | 2.74 | 2.69 | 0.90 | 4.93 | 2.74 | 2.73 | 2.43 | 3.08 |
| Provence-Alpes-Côte d'Azur | HPV16/18 | | 2023 | (59,66] | 754 | 3.32 | 3.47 | 3.45 | 2.25 | 4.78 | 3.47 | 3.47 | 3.20 | 3.76 |
| Île-de-France | HPV16/18 | | 2023 | (59,66] | 1,401 | 4.64 | 3.21 | 3.21 | 2.28 | 4.21 | 3.21 | 3.21 | 2.99 | 3.43 |
| Auvergne-Rhône-Alpes | Other genotypes | | 2020 | [30,39] | | | 12.65 | 12.53 | 9.40 | 15.90 | 12.64 | 12.63 | 11.67 | 13.64 |
| Bourgogne-Franche-Comté | Other genotypes | | 2020 | [30,39] | | | 11.21 | 11.11 | 6.76 | 15.94 | 11.23 | 11.22 | 10.27 | 12.29 |
| Bretagne | Other genotypes | | 2020 | [30,39] | | | 10.92 | 10.92 | 6.99 | 14.86 | 10.91 | 10.91 | 10.02 | 11.85 |

**Table S8** **Observed HR HPV cervical infection prevalence, posterior predictive HR HPV cervical infection prevalence, and posterior expected HR HPV cervical infection prevalence, stratified by various dimensions.** ***(continued)***

| | | | | | | | Posterior distribution | | | | | | | |
|---|---|---|---|---|---|---|---|---|---|---|---|---|---|---|
| | | | | | | | Predictive prevalence | | | | Expected prevalence | | | |
| | | | | | | | | | ETI95% | | | | ETI95% | |
| Region | Genotypes | Type of screening | Year | Age | Number of tests | Observed | Average | Median | LB | UB | Average | Median | LB | UB |
| Centre-Val de Loire | Other genotypes | | 2020 | [30,39] | | | 11.25 | 11.21 | 9.37 | 13.14 | 11.25 | 11.25 | 10.77 | 11.77 |
| Corse | Other genotypes | | 2020 | [30,39] | | | 13.82 | 14.13 | 6.52 | 21.74 | 13.89 | 13.83 | 11.30 | 16.88 |
| Grand Est | Other genotypes | | 2020 | [30,39] | | | 11.13 | 11.06 | 7.23 | 15.32 | 11.17 | 11.16 | 10.28 | 12.16 |
| Hauts-de-France | Other genotypes | | 2020 | [30,39] | | | 10.94 | 10.94 | 8.99 | 12.81 | 10.91 | 10.91 | 10.43 | 11.42 |
| Normandie | Other genotypes | | 2020 | [30,39] | | | 11.63 | 11.58 | 9.47 | 13.80 | 11.62 | 11.62 | 11.01 | 12.24 |
| Nouvelle-Aquitaine | Other genotypes | | 2020 | [30,39] | | | 12.00 | 11.89 | 9.51 | 14.74 | 12.01 | 12.01 | 11.47 | 12.56 |
| Occitanie | Other genotypes | | 2020 | [30,39] | | | 14.12 | 14.03 | 11.42 | 16.78 | 14.16 | 14.15 | 13.15 | 15.22 |
| Pays de la Loire | Other genotypes | | 2020 | [30,39] | | | 9.43 | 9.46 | 6.49 | 12.43 | 9.46 | 9.44 | 8.62 | 10.35 |
| Provence-Alpes-Côte d'Azur | Other genotypes | | 2020 | [30,39] | | | 12.42 | 12.36 | 10.21 | 14.88 | 12.43 | 12.44 | 11.78 | 13.08 |
| Île-de-France | Other genotypes | | 2020 | [30,39] | | | 12.49 | 12.49 | 11.95 | 13.06 | 12.50 | 12.49 | 12.20 | 12.80 |
| Auvergne-Rhône-Alpes | Other genotypes | | 2020 | (39,49] | 262 | 7.25 | 8.93 | 8.78 | 5.34 | 12.60 | 8.91 | 8.91 | 8.20 | 9.66 |
| Bourgogne-Franche-Comté | Other genotypes | | 2020 | (39,49] | 144 | 6.94 | 7.95 | 7.64 | 3.47 | 12.50 | 7.93 | 7.92 | 7.25 | 8.68 |
| Bretagne | Other genotypes | | 2020 | (39,49] | 158 | 10.76 | 7.80 | 7.59 | 3.80 | 12.03 | 7.86 | 7.86 | 7.10 | 8.71 |
| Centre-Val de Loire | Other genotypes | | 2020 | (39,49] | 1,001 | 7.59 | 7.91 | 7.89 | 6.29 | 9.69 | 7.89 | 7.89 | 7.54 | 8.27 |
| Corse | Other genotypes | | 2020 | (39,49] | 118 | 15.25 | 10.29 | 10.17 | 5.08 | 16.10 | 10.22 | 10.17 | 8.16 | 12.70 |
| Grand Est | Other genotypes | | 2020 | (39,49] | 241 | 8.71 | 7.85 | 7.88 | 4.56 | 11.21 | 7.84 | 7.83 | 7.16 | 8.61 |
| Hauts-de-France | Other genotypes | | 2020 | (39,49] | 759 | 7.25 | 7.66 | 7.64 | 5.67 | 9.62 | 7.66 | 7.65 | 7.29 | 8.05 |
| Normandie | Other genotypes | | 2020 | (39,49] | 675 | 8.44 | 8.00 | 8.00 | 5.93 | 10.22 | 7.99 | 7.99 | 7.55 | 8.46 |
| Nouvelle-Aquitaine | Other genotypes | | 2020 | (39,49] | 424 | 7.78 | 8.33 | 8.25 | 5.90 | 11.08 | 8.37 | 8.36 | 7.97 | 8.78 |
| Occitanie | Other genotypes | | 2020 | (39,49] | 628 | 8.60 | 10.56 | 10.51 | 8.12 | 13.22 | 10.54 | 10.54 | 9.69 | 11.39 |
| Pays de la Loire | Other genotypes | | 2020 | (39,49] | 370 | 7.03 | 6.86 | 6.76 | 4.32 | 9.46 | 6.84 | 6.84 | 6.22 | 7.50 |
| Provence-Alpes-Côte d'Azur | Other genotypes | | 2020 | (39,49] | 620 | 9.52 | 8.43 | 8.39 | 6.29 | 10.81 | 8.44 | 8.43 | 7.94 | 8.96 |
| Île-de-France | Other genotypes | | 2020 | (39,49] | 16,220 | 8.47 | 8.75 | 8.75 | 8.26 | 9.24 | 8.74 | 8.74 | 8.51 | 8.99 |
| Auvergne-Rhône-Alpes | Other genotypes | | 2020 | (49,59] | 215 | 6.51 | 7.63 | 7.44 | 4.19 | 11.16 | 7.68 | 7.68 | 7.05 | 8.36 |
| Bourgogne-Franche-Comté | Other genotypes | | 2020 | (49,59] | 98 | 6.12 | 6.87 | 7.14 | 2.04 | 12.24 | 6.92 | 6.90 | 6.33 | 7.57 |
| Bretagne | Other genotypes | | 2020 | (49,59] | 107 | 6.54 | 6.61 | 6.54 | 2.80 | 12.15 | 6.58 | 6.56 | 5.91 | 7.32 |
| Centre-Val de Loire | Other genotypes | | 2020 | (49,59] | 973 | 5.76 | 6.63 | 6.63 | 5.14 | 8.32 | 6.64 | 6.64 | 6.33 | 6.98 |
| Corse | Other genotypes | | 2020 | (49,59] | 111 | 6.31 | 8.62 | 8.11 | 3.60 | 14.41 | 8.55 | 8.51 | 6.84 | 10.62 |
| Grand Est | Other genotypes | | 2020 | (49,59] | 279 | 5.73 | 6.52 | 6.45 | 3.58 | 9.68 | 6.51 | 6.50 | 5.86 | 7.24 |
| Hauts-de-France | Other genotypes | | 2020 | (49,59] | 626 | 7.03 | 6.44 | 6.39 | 4.63 | 8.47 | 6.45 | 6.45 | 6.11 | 6.80 |
| Normandie | Other genotypes | | 2020 | (49,59] | 521 | 6.91 | 6.68 | 6.72 | 4.61 | 9.02 | 6.69 | 6.69 | 6.32 | 7.09 |
| Nouvelle-Aquitaine | Other genotypes | | 2020 | (49,59] | 367 | 8.45 | 7.10 | 7.08 | 4.63 | 9.82 | 7.12 | 7.12 | 6.75 | 7.52 |
| Occitanie | Other genotypes | | 2020 | (49,59] | 536 | 9.89 | 8.67 | 8.58 | 6.34 | 11.19 | 8.65 | 8.65 | 8.01 | 9.33 |
| Pays de la Loire | Other genotypes | | 2020 | (49,59] | 279 | 5.73 | 5.80 | 5.73 | 3.23 | 8.96 | 5.82 | 5.81 | 5.33 | 6.37 |
| Provence-Alpes-Côte d'Azur | Other genotypes | | 2020 | (49,59] | 509 | 8.64 | 7.13 | 7.07 | 5.11 | 9.43 | 7.12 | 7.12 | 6.69 | 7.55 |
| Île-de-France | Other genotypes | | 2020 | (49,59] | 11,877 | 7.20 | 7.42 | 7.42 | 6.92 | 7.96 | 7.42 | 7.42 | 7.19 | 7.66 |
| Auvergne-Rhône-Alpes | Other genotypes | | 2020 | (59,66] | 96 | 10.42 | 6.84 | 6.25 | 2.08 | 12.50 | 6.87 | 6.86 | 6.26 | 7.53 |
| Bourgogne-Franche-Comté | Other genotypes | | 2020 | (59,66] | 88 | 7.95 | 6.23 | 5.68 | 2.27 | 11.36 | 6.23 | 6.22 | 5.65 | 6.84 |
| Bretagne | Other genotypes | | 2020 | (59,66] | 65 | 4.62 | 5.57 | 6.15 | 0.00 | 12.31 | 5.67 | 5.64 | 5.01 | 6.42 |

**Table S8** **Observed HR HPV cervical infection prevalence, posterior predictive HR HPV cervical infection prevalence, and posterior expected HR HPV cervical infection prevalence, stratified by various dimensions.** ***(continued)***

| | | | | | | | Posterior distribution | | | | | | | |
|---|---|---|---|---|---|---|---|---|---|---|---|---|---|---|
| | | | | | | | Predictive prevalence | | | | Expected prevalence | | | |
| | | | | | | | | | ETI95% | | | | ETI95% | |
| Region | Genotypes | Type of screening | Year | Age | Number of tests | Observed | Average | Median | LB | UB | Average | Median | LB | UB |
| Centre-Val de Loire | Other genotypes | | 2020 | (59,66] | 501 | 3.99 | 5.98 | 5.99 | 3.99 | 8.18 | 5.94 | 5.94 | 5.60 | 6.29 |
| Corse | Other genotypes | | 2020 | (59,66] | 56 | 1.79 | 7.61 | 7.14 | 1.79 | 16.07 | 7.65 | 7.59 | 6.23 | 9.39 |
| Grand Est | Other genotypes | | 2020 | (59,66] | 127 | 3.94 | 5.74 | 5.51 | 1.57 | 10.24 | 5.76 | 5.75 | 5.14 | 6.46 |
| Hauts-de-France | Other genotypes | | 2020 | (59,66] | 276 | 6.16 | 5.72 | 5.80 | 3.26 | 8.70 | 5.69 | 5.69 | 5.34 | 6.04 |
| Normandie | Other genotypes | | 2020 | (59,66] | 262 | 4.58 | 5.81 | 5.73 | 3.05 | 8.78 | 5.88 | 5.87 | 5.48 | 6.29 |
| Nouvelle-Aquitaine | Other genotypes | | 2020 | (59,66] | 267 | 7.12 | 6.49 | 6.37 | 3.75 | 9.74 | 6.48 | 6.48 | 6.09 | 6.90 |
| Occitanie | Other genotypes | | 2020 | (59,66] | 288 | 6.25 | 7.80 | 7.64 | 4.86 | 10.77 | 7.78 | 7.78 | 7.17 | 8.43 |
| Pays de la Loire | Other genotypes | | 2020 | (59,66] | 142 | 3.52 | 5.43 | 5.63 | 2.11 | 9.86 | 5.44 | 5.43 | 4.93 | 6.03 |
| Provence-Alpes-Côte d'Azur | Other genotypes | | 2020 | (59,66] | 209 | 5.26 | 6.39 | 6.22 | 3.35 | 9.58 | 6.38 | 6.38 | 5.98 | 6.79 |
| Île-de-France | Other genotypes | | 2020 | (59,66] | 4,709 | 6.18 | 6.58 | 6.58 | 5.82 | 7.35 | 6.57 | 6.57 | 6.30 | 6.86 |
| Auvergne-Rhône-Alpes | Other genotypes | | 2021 | [30,39] | | | 12.41 | 12.36 | 10.41 | 14.47 | 12.42 | 12.40 | 11.57 | 13.30 |
| Bourgogne-Franche-Comté | Other genotypes | | 2021 | [30,39] | | | 11.13 | 11.09 | 8.53 | 13.99 | 11.14 | 11.13 | 10.28 | 12.07 |
| Bretagne | Other genotypes | | 2021 | [30,39] | | | 10.45 | 10.46 | 8.28 | 12.64 | 10.42 | 10.41 | 9.72 | 11.15 |
| Centre-Val de Loire | Other genotypes | | 2021 | [30,39] | | | 10.64 | 10.63 | 9.68 | 11.63 | 10.63 | 10.63 | 10.24 | 11.03 |
| Corse | Other genotypes | | 2021 | [30,39] | | | 12.13 | 7.69 | 0.00 | 30.77 | 12.30 | 12.23 | 10.20 | 14.78 |
| Grand Est | Other genotypes | | 2021 | [30,39] | | | 10.56 | 10.58 | 8.73 | 12.43 | 10.57 | 10.57 | 9.99 | 11.21 |
| Hauts-de-France | Other genotypes | | 2021 | [30,39] | | | 10.38 | 10.39 | 9.48 | 11.36 | 10.39 | 10.39 | 10.02 | 10.79 |
| Normandie | Other genotypes | | 2021 | [30,39] | | | 11.24 | 11.21 | 10.03 | 12.42 | 11.23 | 11.23 | 10.73 | 11.74 |
| Nouvelle-Aquitaine | Other genotypes | | 2021 | [30,39] | | | 11.28 | 11.26 | 10.06 | 12.54 | 11.27 | 11.26 | 10.84 | 11.73 |
| Occitanie | Other genotypes | | 2021 | [30,39] | | | 12.50 | 12.50 | 10.88 | 14.12 | 12.51 | 12.50 | 11.89 | 13.15 |
| Pays de la Loire | Other genotypes | | 2021 | [30,39] | | | 8.94 | 8.97 | 7.08 | 10.87 | 8.97 | 8.96 | 8.30 | 9.70 |
| Provence-Alpes-Côte d'Azur | Other genotypes | | 2021 | [30,39] | | | 11.96 | 11.95 | 10.91 | 13.05 | 11.95 | 11.96 | 11.51 | 12.43 |
| Île-de-France | Other genotypes | | 2021 | [30,39] | | | 11.90 | 11.89 | 11.44 | 12.35 | 11.90 | 11.90 | 11.66 | 12.15 |
| Auvergne-Rhône-Alpes | Other genotypes | | 2021 | (39,49] | 852 | 8.10 | 8.59 | 8.57 | 6.69 | 10.56 | 8.60 | 8.59 | 7.95 | 9.27 |
| Bourgogne-Franche-Comté | Other genotypes | | 2021 | (39,49] | 392 | 9.44 | 7.81 | 7.91 | 5.10 | 10.71 | 7.78 | 7.78 | 7.14 | 8.46 |
| Bretagne | Other genotypes | | 2021 | (39,49] | 604 | 8.61 | 7.26 | 7.28 | 5.13 | 9.44 | 7.24 | 7.24 | 6.70 | 7.79 |
| Centre-Val de Loire | Other genotypes | | 2021 | (39,49] | 4,339 | 7.12 | 7.50 | 7.49 | 6.64 | 8.37 | 7.50 | 7.50 | 7.20 | 7.80 |
| Corse | Other genotypes | | 2021 | (39,49] | 5 | 0.00 | 8.41 | 0.00 | 0.00 | 40.00 | 8.56 | 8.53 | 6.83 | 10.46 |
| Grand Est | Other genotypes | | 2021 | (39,49] | 1,119 | 7.06 | 7.48 | 7.42 | 5.81 | 9.12 | 7.48 | 7.48 | 7.04 | 7.96 |
| Hauts-de-France | Other genotypes | | 2021 | (39,49] | 3,026 | 7.60 | 7.34 | 7.34 | 6.38 | 8.36 | 7.34 | 7.34 | 7.05 | 7.63 |
| Normandie | Other genotypes | | 2021 | (39,49] | 2,315 | 7.95 | 7.89 | 7.90 | 6.78 | 9.07 | 7.90 | 7.89 | 7.54 | 8.27 |
| Nouvelle-Aquitaine | Other genotypes | | 2021 | (39,49] | 2,362 | 7.96 | 7.85 | 7.83 | 6.73 | 9.06 | 7.86 | 7.86 | 7.54 | 8.19 |
| Occitanie | Other genotypes | | 2021 | (39,49] | 1,748 | 10.07 | 8.88 | 8.87 | 7.49 | 10.35 | 8.87 | 8.87 | 8.40 | 9.36 |
| Pays de la Loire | Other genotypes | | 2021 | (39,49] | 919 | 6.42 | 6.45 | 6.42 | 4.79 | 8.05 | 6.43 | 6.43 | 5.96 | 6.95 |
| Provence-Alpes-Côte d'Azur | Other genotypes | | 2021 | (39,49] | 3,177 | 8.31 | 8.34 | 8.34 | 7.40 | 9.38 | 8.33 | 8.33 | 7.97 | 8.71 |
| Île-de-France | Other genotypes | | 2021 | (39,49] | 25,423 | 8.11 | 8.34 | 8.33 | 7.96 | 8.72 | 8.33 | 8.34 | 8.13 | 8.54 |
| Auvergne-Rhône-Alpes | Other genotypes | | 2021 | (49,59] | 704 | 6.96 | 7.34 | 7.24 | 5.39 | 9.38 | 7.33 | 7.32 | 6.78 | 7.90 |
| Bourgogne-Franche-Comté | Other genotypes | | 2021 | (49,59] | 330 | 6.67 | 6.69 | 6.67 | 3.94 | 9.39 | 6.68 | 6.67 | 6.13 | 7.25 |
| Bretagne | Other genotypes | | 2021 | (49,59] | 592 | 3.89 | 5.96 | 5.91 | 4.05 | 8.11 | 5.94 | 5.94 | 5.50 | 6.40 |

**Table S8 Observed HR HPV cervical infection prevalence, posterior predictive HR HPV cervical infection prevalence, and posterior expected HR HPV cervical infection prevalence, stratified by various dimensions.** ***(continued)***

| | | | | | | | Posterior distribution | | | | | | | |
|---|---|---|---|---|---|---|---|---|---|---|---|---|---|---|
| | | | | | | | Predictive prevalence | | | | Expected prevalence | | | |
| | | | | | | | | | ETI95% | | | | ETI95% | |
| Region | Genotypes | Type of screening | Year | Age | Number of tests | Observed | Average | Median | LB | UB | Average | Median | LB | UB |
| Centre-Val de Loire | Other genotypes | | 2021 | (49,59] | 4,068 | 5.92 | 6.33 | 6.34 | 5.53 | 7.15 | 6.33 | 6.33 | 6.07 | 6.59 |
| Corse | Other genotypes | | 2021 | (49,59] | 10 | 0.00 | 6.75 | 10.00 | 0.00 | 30.00 | 6.83 | 6.81 | 5.96 | 7.77 |
| Grand Est | Other genotypes | | 2021 | (49,59] | 1,062 | 4.90 | 6.35 | 6.31 | 4.90 | 7.91 | 6.35 | 6.34 | 5.97 | 6.75 |
| Hauts-de-France | Other genotypes | | 2021 | (49,59] | 2,111 | 6.44 | 6.22 | 6.21 | 5.21 | 7.30 | 6.23 | 6.23 | 5.98 | 6.51 |
| Normandie | Other genotypes | | 2021 | (49,59] | 1,834 | 5.34 | 6.60 | 6.60 | 5.45 | 7.74 | 6.60 | 6.60 | 6.28 | 6.92 |
| Nouvelle-Aquitaine | Other genotypes | | 2021 | (49,59] | 2,165 | 6.47 | 6.59 | 6.61 | 5.54 | 7.71 | 6.59 | 6.59 | 6.32 | 6.88 |
| Occitanie | Other genotypes | | 2021 | (49,59] | 1,467 | 7.70 | 7.68 | 7.63 | 6.34 | 9.20 | 7.69 | 7.68 | 7.26 | 8.12 |
| Pays de la Loire | Other genotypes | | 2021 | (49,59] | 781 | 5.51 | 5.52 | 5.51 | 3.97 | 7.30 | 5.52 | 5.51 | 5.12 | 5.98 |
| Provence-Alpes-Côte d'Azur | Other genotypes | | 2021 | (49,59] | 2,584 | 7.39 | 7.00 | 7.00 | 6.00 | 8.09 | 7.01 | 7.00 | 6.70 | 7.34 |
| Île-de-France | Other genotypes | | 2021 | (49,59] | 19,159 | 7.08 | 7.05 | 7.05 | 6.64 | 7.47 | 7.05 | 7.05 | 6.86 | 7.26 |
| Auvergne-Rhône-Alpes | Other genotypes | | 2021 | (59,66] | 361 | 5.54 | 6.46 | 6.37 | 4.16 | 9.14 | 6.49 | 6.48 | 5.98 | 7.03 |
| Bourgogne-Franche-Comté | Other genotypes | | 2021 | (59,66] | 229 | 7.86 | 5.95 | 5.68 | 3.06 | 9.17 | 5.97 | 5.97 | 5.46 | 6.51 |
| Bretagne | Other genotypes | | 2021 | (59,66] | 260 | 4.23 | 5.37 | 5.38 | 2.69 | 8.46 | 5.37 | 5.36 | 4.92 | 5.82 |
| Centre-Val de Loire | Other genotypes | | 2021 | (59,66] | 2,161 | 6.06 | 5.64 | 5.65 | 4.63 | 6.71 | 5.65 | 5.65 | 5.36 | 5.94 |
| Corse | Other genotypes | | 2021 | (59,66] | 7 | 0.00 | 6.80 | 0.00 | 0.00 | 28.57 | 6.66 | 6.61 | 5.48 | 8.06 |
| Grand Est | Other genotypes | | 2021 | (59,66] | 608 | 3.29 | 5.69 | 5.76 | 3.78 | 7.73 | 5.69 | 5.69 | 5.32 | 6.08 |
| Hauts-de-France | Other genotypes | | 2021 | (59,66] | 980 | 4.18 | 5.56 | 5.51 | 4.18 | 7.04 | 5.55 | 5.55 | 5.27 | 5.84 |
| Normandie | Other genotypes | | 2021 | (59,66] | 968 | 5.79 | 5.87 | 5.89 | 4.34 | 7.44 | 5.88 | 5.88 | 5.54 | 6.22 |
| Nouvelle-Aquitaine | Other genotypes | | 2021 | (59,66] | 1,407 | 4.83 | 5.95 | 5.90 | 4.76 | 7.25 | 5.97 | 5.97 | 5.67 | 6.29 |
| Occitanie | Other genotypes | | 2021 | (59,66] | 768 | 7.94 | 6.84 | 6.90 | 4.95 | 8.72 | 6.88 | 6.88 | 6.45 | 7.32 |
| Pays de la Loire | Other genotypes | | 2021 | (59,66] | 378 | 3.17 | 5.19 | 5.03 | 2.91 | 7.67 | 5.19 | 5.18 | 4.77 | 5.67 |
| Provence-Alpes-Côte d'Azur | Other genotypes | | 2021 | (59,66] | 1,336 | 7.11 | 6.26 | 6.29 | 4.94 | 7.56 | 6.24 | 6.24 | 5.91 | 6.58 |
| Île-de-France | Other genotypes | | 2021 | (59,66] | 10,265 | 6.39 | 6.24 | 6.23 | 5.74 | 6.78 | 6.24 | 6.24 | 5.99 | 6.49 |
| Auvergne-Rhône-Alpes | Other genotypes | | 2022 | [30,39] | | | 13.45 | 13.45 | 7.56 | 20.17 | 13.49 | 13.47 | 12.36 | 14.62 |
| Bourgogne-Franche-Comté | Other genotypes | | 2022 | [30,39] | | | 11.72 | 11.79 | 8.84 | 14.73 | 11.74 | 11.73 | 10.82 | 12.77 |
| Bretagne | Other genotypes | | 2022 | [30,39] | | | 11.32 | 11.32 | 9.43 | 13.29 | 11.29 | 11.29 | 10.60 | 11.99 |
| Centre-Val de Loire | Other genotypes | | 2022 | [30,39] | | | 11.00 | 10.99 | 9.74 | 12.29 | 10.97 | 10.97 | 10.53 | 11.43 |
| Corse | Other genotypes | | 2022 | [30,39] | | | 12.17 | 0.00 | 0.00 | 50.00 | 11.67 | 11.61 | 9.76 | 13.89 |
| Grand Est | Other genotypes | | 2022 | [30,39] | | | 10.88 | 10.90 | 9.22 | 12.66 | 10.85 | 10.85 | 10.26 | 11.50 |
| Hauts-de-France | Other genotypes | | 2022 | [30,39] | | | 10.90 | 10.91 | 9.91 | 11.86 | 10.89 | 10.89 | 10.48 | 11.30 |
| Normandie | Other genotypes | | 2022 | [30,39] | | | 12.29 | 12.27 | 10.84 | 13.80 | 12.28 | 12.27 | 11.72 | 12.83 |
| Nouvelle-Aquitaine | Other genotypes | | 2022 | [30,39] | | | 11.80 | 11.80 | 10.72 | 12.97 | 11.81 | 11.80 | 11.38 | 12.26 |
| Occitanie | Other genotypes | | 2022 | [30,39] | | | 12.51 | 12.49 | 10.41 | 14.78 | 12.50 | 12.49 | 11.81 | 13.21 |
| Pays de la Loire | Other genotypes | | 2022 | [30,39] | | | 9.92 | 9.75 | 6.60 | 13.21 | 9.91 | 9.90 | 9.16 | 10.72 |
| Provence-Alpes-Côte d'Azur | Other genotypes | | 2022 | [30,39] | | | 12.70 | 12.73 | 11.42 | 13.88 | 12.71 | 12.70 | 12.21 | 13.22 |
| Île-de-France | Other genotypes | | 2022 | [30,39] | | | 12.81 | 12.80 | 11.94 | 13.69 | 12.81 | 12.81 | 12.44 | 13.18 |
| Auvergne-Rhône-Alpes | Other genotypes | | 2022 | (39,49] | 79 | 16.46 | 9.63 | 8.86 | 3.80 | 16.46 | 9.71 | 9.69 | 8.61 | 10.85 |
| Bourgogne-Franche-Comté | Other genotypes | | 2022 | (39,49] | 334 | 8.68 | 8.17 | 8.08 | 5.09 | 11.38 | 8.17 | 8.16 | 7.48 | 8.90 |
| Bretagne | Other genotypes | | 2022 | (39,49] | 836 | 7.30 | 7.73 | 7.66 | 5.86 | 9.69 | 7.73 | 7.73 | 7.23 | 8.22 |

**Table S8** **Observed HR HPV cervical infection prevalence, posterior predictive HR HPV cervical infection prevalence, and posterior expected HR HPV cervical infection prevalence, stratified by various dimensions.** ***(continued)***

| | | | | | | | Posterior distribution | | | | | | | |
|---|---|---|---|---|---|---|---|---|---|---|---|---|---|---|
| | | | | | | | Predictive prevalence | | | | Expected prevalence | | | |
| | | | | | | | | | ETI95% | | | | ETI95% | |
| Region | Genotypes | Type of screening | Year | Age | Number of tests | Observed | Average | Median | LB | UB | Average | Median | LB | UB |
| Centre-Val de Loire | Other genotypes | | 2022 | (39,49] | 2,327 | 8.12 | 7.67 | 7.69 | 6.53 | 8.85 | 7.68 | 7.67 | 7.34 | 8.02 |
| Corse | Other genotypes | | 2022 | (39,49] | 5 | 20.00 | 8.65 | 0.00 | 0.00 | 40.00 | 8.54 | 8.48 | 6.93 | 10.44 |
| Grand Est | Other genotypes | | 2022 | (39,49] | 1,056 | 9.85 | 7.76 | 7.77 | 6.16 | 9.47 | 7.78 | 7.77 | 7.32 | 8.29 |
| Hauts-de-France | Other genotypes | | 2022 | (39,49] | 2,668 | 8.43 | 7.67 | 7.68 | 6.60 | 8.73 | 7.66 | 7.66 | 7.35 | 7.99 |
| Normandie | Other genotypes | | 2022 | (39,49] | 1,836 | 8.33 | 8.62 | 8.61 | 7.30 | 10.02 | 8.61 | 8.61 | 8.19 | 9.03 |
| Nouvelle-Aquitaine | Other genotypes | | 2022 | (39,49] | 3,149 | 7.94 | 8.35 | 8.35 | 7.34 | 9.37 | 8.36 | 8.36 | 8.04 | 8.71 |
| Occitanie | Other genotypes | | 2022 | (39,49] | 881 | 8.17 | 8.88 | 8.85 | 6.92 | 10.78 | 8.90 | 8.90 | 8.38 | 9.46 |
| Pays de la Loire | Other genotypes | | 2022 | (39,49] | 274 | 6.20 | 7.01 | 6.93 | 4.01 | 10.22 | 7.04 | 7.03 | 6.47 | 7.65 |
| Provence-Alpes-Côte d'Azur | Other genotypes | | 2022 | (39,49] | 2,168 | 8.30 | 8.91 | 8.90 | 7.70 | 10.15 | 8.92 | 8.92 | 8.55 | 9.30 |
| Île-de-France | Other genotypes | | 2022 | (39,49] | 4,741 | 8.08 | 8.68 | 8.67 | 7.85 | 9.53 | 8.69 | 8.68 | 8.41 | 8.98 |
| Auvergne-Rhône-Alpes | Other genotypes | | 2022 | (49,59] | 62 | 17.74 | 8.71 | 8.06 | 1.61 | 16.13 | 8.68 | 8.66 | 7.62 | 9.81 |
| Bourgogne-Franche-Comté | Other genotypes | | 2022 | (49,59] | 240 | 7.92 | 6.98 | 7.08 | 3.75 | 10.42 | 6.99 | 6.98 | 6.40 | 7.62 |
| Bretagne | Other genotypes | | 2022 | (49,59] | 679 | 6.77 | 6.61 | 6.63 | 4.71 | 8.69 | 6.63 | 6.62 | 6.18 | 7.06 |
| Centre-Val de Loire | Other genotypes | | 2022 | (49,59] | 2,116 | 6.66 | 6.54 | 6.52 | 5.53 | 7.66 | 6.54 | 6.54 | 6.23 | 6.84 |
| Corse | Other genotypes | | 2022 | (49,59] | 2 | 0.00 | 8.30 | 0.00 | 0.00 | 50.00 | 7.65 | 7.56 | 5.63 | 10.09 |
| Grand Est | Other genotypes | | 2022 | (49,59] | 887 | 5.86 | 6.86 | 6.88 | 5.30 | 8.57 | 6.89 | 6.88 | 6.46 | 7.35 |
| Hauts-de-France | Other genotypes | | 2022 | (49,59] | 1,992 | 8.08 | 6.53 | 6.53 | 5.47 | 7.68 | 6.54 | 6.54 | 6.26 | 6.85 |
| Normandie | Other genotypes | | 2022 | (49,59] | 1,212 | 6.77 | 7.15 | 7.10 | 5.69 | 8.66 | 7.16 | 7.16 | 6.80 | 7.53 |
| Nouvelle-Aquitaine | Other genotypes | | 2022 | (49,59] | 2,734 | 7.06 | 7.03 | 7.02 | 6.07 | 8.05 | 7.03 | 7.02 | 6.73 | 7.33 |
| Occitanie | Other genotypes | | 2022 | (49,59] | 851 | 7.76 | 7.68 | 7.64 | 5.88 | 9.52 | 7.69 | 7.68 | 7.24 | 8.18 |
| Pays de la Loire | Other genotypes | | 2022 | (49,59] | 225 | 6.67 | 6.19 | 6.22 | 3.11 | 9.33 | 6.16 | 6.16 | 5.66 | 6.73 |
| Provence-Alpes-Côte d'Azur | Other genotypes | | 2022 | (49,59] | 1,876 | 7.36 | 7.49 | 7.52 | 6.34 | 8.69 | 7.51 | 7.51 | 7.16 | 7.85 |
| Île-de-France | Other genotypes | | 2022 | (49,59] | 3,802 | 7.50 | 7.27 | 7.29 | 6.44 | 8.15 | 7.27 | 7.27 | 7.01 | 7.53 |
| Auvergne-Rhône-Alpes | Other genotypes | | 2022 | (59,66] | 35 | 2.86 | 7.44 | 5.71 | 0.00 | 17.14 | 7.53 | 7.52 | 6.66 | 8.47 |
| Bourgogne-Franche-Comté | Other genotypes | | 2022 | (59,66] | 137 | 5.84 | 6.09 | 5.84 | 2.19 | 10.22 | 6.14 | 6.14 | 5.59 | 6.74 |
| Bretagne | Other genotypes | | 2022 | (59,66] | 401 | 6.23 | 5.76 | 5.74 | 3.49 | 8.23 | 5.78 | 5.78 | 5.34 | 6.23 |
| Centre-Val de Loire | Other genotypes | | 2022 | (59,66] | 1,060 | 6.98 | 5.80 | 5.75 | 4.43 | 7.26 | 5.81 | 5.81 | 5.49 | 6.13 |
| Corse | Other genotypes | | 2022 | (59,66] | 2 | 50.00 | 8.23 | 0.00 | 0.00 | 50.00 | 8.93 | 8.79 | 6.33 | 12.22 |
| Grand Est | Other genotypes | | 2022 | (59,66] | 394 | 5.58 | 6.19 | 6.09 | 3.81 | 8.63 | 6.15 | 6.15 | 5.75 | 6.60 |
| Hauts-de-France | Other genotypes | | 2022 | (59,66] | 883 | 6.00 | 5.91 | 5.89 | 4.42 | 7.59 | 5.91 | 5.91 | 5.60 | 6.23 |
| Normandie | Other genotypes | | 2022 | (59,66] | 705 | 4.68 | 6.29 | 6.24 | 4.54 | 8.09 | 6.26 | 6.26 | 5.90 | 6.63 |
| Nouvelle-Aquitaine | Other genotypes | | 2022 | (59,66] | 1,480 | 6.42 | 6.34 | 6.35 | 5.07 | 7.64 | 6.34 | 6.34 | 6.00 | 6.70 |
| Occitanie | Other genotypes | | 2022 | (59,66] | 438 | 5.94 | 6.94 | 6.85 | 4.57 | 9.59 | 6.90 | 6.90 | 6.46 | 7.40 |
| Pays de la Loire | Other genotypes | | 2022 | (59,66] | 145 | 5.52 | 5.79 | 5.52 | 2.07 | 9.67 | 5.80 | 5.79 | 5.25 | 6.41 |
| Provence-Alpes-Côte d'Azur | Other genotypes | | 2022 | (59,66] | 902 | 6.21 | 6.70 | 6.65 | 4.99 | 8.43 | 6.69 | 6.69 | 6.34 | 7.06 |
| Île-de-France | Other genotypes | | 2022 | (59,66] | 1,713 | 5.78 | 6.56 | 6.54 | 5.43 | 7.82 | 6.55 | 6.55 | 6.25 | 6.85 |
| Auvergne-Rhône-Alpes | Other genotypes | | 2023 | [30,39] | | | 16.36 | 16.67 | 8.89 | 24.44 | 16.36 | 16.36 | 14.50 | 18.35 |
| Bourgogne-Franche-Comté | Other genotypes | | 2023 | [30,39] | | | 13.04 | 12.83 | 8.68 | 17.74 | 13.08 | 13.07 | 11.89 | 14.33 |
| Bretagne | Other genotypes | | 2023 | [30,39] | | | 12.56 | 12.53 | 10.88 | 14.30 | 12.56 | 12.55 | 11.85 | 13.34 |

**Table S8** **Observed HR HPV cervical infection prevalence, posterior predictive HR HPV cervical infection prevalence, and posterior expected HR HPV cervical infection prevalence, stratified by various dimensions.** ***(continued)***

| | | | | | | | Posterior distribution | | | | | | | |
|---|---|---|---|---|---|---|---|---|---|---|---|---|---|---|
| | | | | | | | Predictive prevalence | | | | Expected prevalence | | | |
| | | | | | | | | | ETI95% | | | | ETI95% | |
| Region | Genotypes | Type of screening | Year | Age | Number of tests | Observed | Average | Median | LB | UB | Average | Median | LB | UB |
| Centre-Val de Loire | Other genotypes | | 2023 | [30,39] | | | 12.84 | 12.84 | 11.41 | 14.36 | 12.87 | 12.86 | 12.34 | 13.43 |
| Corse | Other genotypes | | 2023 | [30,39] | | | 11.07 | 0.00 | 0.00 | 100.00 | 10.92 | 10.55 | 6.24 | 17.33 |
| Grand Est | Other genotypes | | 2023 | [30,39] | | | 12.87 | 12.83 | 10.81 | 15.03 | 12.87 | 12.86 | 12.13 | 13.65 |
| Hauts-de-France | Other genotypes | | 2023 | [30,39] | | | 12.75 | 12.73 | 11.59 | 13.97 | 12.76 | 12.76 | 12.27 | 13.25 |
| Normandie | Other genotypes | | 2023 | [30,39] | | | 14.13 | 14.13 | 12.08 | 16.19 | 14.14 | 14.14 | 13.49 | 14.82 |
| Nouvelle-Aquitaine | Other genotypes | | 2023 | [30,39] | | | 13.65 | 13.65 | 12.35 | 14.98 | 13.64 | 13.64 | 13.15 | 14.17 |
| Occitanie | Other genotypes | | 2023 | [30,39] | | | 14.86 | 14.84 | 11.45 | 18.75 | 14.83 | 14.82 | 13.94 | 15.85 |
| Pays de la Loire | Other genotypes | | 2023 | [30,39] | | | 10.94 | 11.01 | 8.09 | 14.16 | 10.97 | 10.95 | 10.10 | 11.89 |
| Provence-Alpes-Côte d'Azur | Other genotypes | | 2023 | [30,39] | | | 14.95 | 14.95 | 13.42 | 16.51 | 14.95 | 14.94 | 14.35 | 15.56 |
| Île-de-France | Other genotypes | | 2023 | [30,39] | | | 15.13 | 15.11 | 14.15 | 16.15 | 15.13 | 15.13 | 14.69 | 15.58 |
| Auvergne-Rhône-Alpes | Other genotypes | | 2023 | (39,49] | 65 | 7.69 | 11.80 | 12.31 | 4.62 | 20.00 | 11.71 | 11.69 | 10.09 | 13.39 |
| Bourgogne-Franche-Comté | Other genotypes | | 2023 | (39,49] | 226 | 8.41 | 8.79 | 8.85 | 5.31 | 12.83 | 8.74 | 8.74 | 7.86 | 9.71 |
| Bretagne | Other genotypes | | 2023 | (39,49] | 1,372 | 9.91 | 8.61 | 8.60 | 7.07 | 10.20 | 8.63 | 8.63 | 8.12 | 9.18 |
| Centre-Val de Loire | Other genotypes | | 2023 | (39,49] | 1,626 | 10.15 | 9.01 | 8.98 | 7.63 | 10.52 | 9.01 | 9.02 | 8.63 | 9.42 |
| Corse | Other genotypes | | 2023 | (39,49] | 1 | 0.00 | 9.77 | 0.00 | 0.00 | 100.00 | 11.01 | 10.74 | 6.05 | 17.73 |
| Grand Est | Other genotypes | | 2023 | (39,49] | 944 | 10.28 | 9.30 | 9.32 | 7.42 | 11.33 | 9.31 | 9.31 | 8.75 | 9.90 |
| Hauts-de-France | Other genotypes | | 2023 | (39,49] | 2,252 | 10.79 | 9.10 | 9.10 | 7.86 | 10.30 | 9.10 | 9.10 | 8.72 | 9.49 |
| Normandie | Other genotypes | | 2023 | (39,49] | 1,130 | 8.76 | 9.96 | 10.00 | 8.14 | 11.77 | 9.99 | 9.98 | 9.48 | 10.50 |
| Nouvelle-Aquitaine | Other genotypes | | 2023 | (39,49] | 2,650 | 10.38 | 9.61 | 9.62 | 8.45 | 10.83 | 9.61 | 9.60 | 9.23 | 10.03 |
| Occitanie | Other genotypes | | 2023 | (39,49] | 404 | 6.93 | 10.58 | 10.64 | 7.67 | 13.61 | 10.59 | 10.58 | 9.85 | 11.40 |
| Pays de la Loire | Other genotypes | | 2023 | (39,49] | 430 | 7.21 | 7.93 | 7.91 | 5.35 | 10.70 | 7.91 | 7.90 | 7.27 | 8.62 |
| Provence-Alpes-Côte d'Azur | Other genotypes | | 2023 | (39,49] | 1,690 | 10.24 | 10.37 | 10.36 | 8.87 | 11.89 | 10.37 | 10.37 | 9.91 | 10.84 |
| Île-de-France | Other genotypes | | 2023 | (39,49] | 3,944 | 9.15 | 10.28 | 10.27 | 9.28 | 11.28 | 10.28 | 10.28 | 9.94 | 10.64 |
| Auvergne-Rhône-Alpes | Other genotypes | | 2023 | (49,59] | 54 | 9.26 | 10.06 | 9.26 | 3.66 | 18.52 | 10.13 | 10.11 | 8.68 | 11.73 |
| Bourgogne-Franche-Comté | Other genotypes | | 2023 | (49,59] | 158 | 8.86 | 7.56 | 7.59 | 3.80 | 12.03 | 7.53 | 7.52 | 6.78 | 8.36 |
| Bretagne | Other genotypes | | 2023 | (49,59] | 1,139 | 8.52 | 7.34 | 7.33 | 5.79 | 8.96 | 7.33 | 7.33 | 6.88 | 7.83 |
| Centre-Val de Loire | Other genotypes | | 2023 | (49,59] | 1,525 | 6.30 | 7.69 | 7.67 | 6.30 | 9.11 | 7.71 | 7.71 | 7.37 | 8.07 |
| Grand Est | Other genotypes | | 2023 | (49,59] | 822 | 7.66 | 7.78 | 7.79 | 5.96 | 9.61 | 7.77 | 7.76 | 7.29 | 8.29 |
| Hauts-de-France | Other genotypes | | 2023 | (49,59] | 1,671 | 8.62 | 7.66 | 7.66 | 6.34 | 9.04 | 7.66 | 7.66 | 7.33 | 8.01 |
| Normandie | Other genotypes | | 2023 | (49,59] | 860 | 8.02 | 8.39 | 8.37 | 6.51 | 10.35 | 8.40 | 8.40 | 7.96 | 8.86 |
| Nouvelle-Aquitaine | Other genotypes | | 2023 | (49,59] | 2,168 | 8.49 | 8.19 | 8.16 | 7.06 | 9.46 | 8.18 | 8.18 | 7.84 | 8.54 |
| Occitanie | Other genotypes | | 2023 | (49,59] | 367 | 8.72 | 8.97 | 8.99 | 6.27 | 11.99 | 8.95 | 8.94 | 8.32 | 9.65 |
| Pays de la Loire | Other genotypes | | 2023 | (49,59] | 403 | 7.20 | 6.84 | 6.70 | 4.47 | 9.68 | 6.87 | 6.86 | 6.29 | 7.51 |
| Provence-Alpes-Côte d'Azur | Other genotypes | | 2023 | (49,59] | 1,468 | 10.35 | 8.84 | 8.86 | 7.43 | 10.35 | 8.86 | 8.85 | 8.44 | 9.29 |
| Île-de-France | Other genotypes | | 2023 | (49,59] | 3,146 | 7.79 | 8.66 | 8.65 | 7.63 | 9.73 | 8.66 | 8.65 | 8.35 | 8.99 |
| Auvergne-Rhône-Alpes | Other genotypes | | 2023 | (59,66] | 32 | 3.12 | 9.42 | 9.38 | 0.00 | 21.88 | 9.31 | 9.30 | 8.03 | 10.71 |
| Bourgogne-Franche-Comté | Other genotypes | | 2023 | (59,66] | 92 | 11.96 | 6.91 | 6.52 | 2.17 | 11.96 | 6.90 | 6.89 | 6.16 | 7.71 |
| Bretagne | Other genotypes | | 2023 | (59,66] | 725 | 6.34 | 6.26 | 6.21 | 4.41 | 8.14 | 6.26 | 6.26 | 5.76 | 6.79 |
| Centre-Val de Loire | Other genotypes | | 2023 | (59,66] | 749 | 6.28 | 6.72 | 6.68 | 4.94 | 8.68 | 6.71 | 6.71 | 6.34 | 7.08 |

**Table S8 Observed HR HPV cervical infection prevalence, posterior predictive HR HPV cervical infection prevalence, and posterior expected HR HPV cervical infection prevalence, stratified by various dimensions.** ***(continued)***

| | | | | | | | Posterior distribution | | | | | | | |
|---|---|---|---|---|---|---|---|---|---|---|---|---|---|---|
| | | | | | | | Predictive prevalence | | | | Expected prevalence | | | |
| | | | | | | | | | ETI95% | | | | ETI95% | |
| Region | Genotypes | Type of screening | Year | Age | Number of tests | Observed | Average | Median | LB | UB | Average | Median | LB | UB |
| Corse | Other genotypes | | 2023 | (59,66] | 3 | 0.00 | 8.02 | 0.00 | 0.00 | 33.33 | 7.75 | 7.67 | 5.63 | 10.49 |
| Grand Est | Other genotypes | | 2023 | (59,66] | 473 | 5.71 | 7.01 | 6.98 | 4.65 | 9.51 | 6.96 | 6.96 | 6.48 | 7.47 |
| Hauts-de-France | Other genotypes | | 2023 | (59,66] | 956 | 8.47 | 6.84 | 6.80 | 5.23 | 8.58 | 6.86 | 6.85 | 6.49 | 7.21 |
| Normandie | Other genotypes | | 2023 | (59,66] | 452 | 7.74 | 7.23 | 7.30 | 4.87 | 9.73 | 7.23 | 7.22 | 6.81 | 7.68 |
| Nouvelle-Aquitaine | Other genotypes | | 2023 | (59,66] | 1,167 | 7.46 | 7.39 | 7.37 | 5.91 | 9.00 | 7.39 | 7.39 | 7.01 | 7.80 |
| Occitanie | Other genotypes | | 2023 | (59,66] | 220 | 9.09 | 7.91 | 7.73 | 4.55 | 11.82 | 7.97 | 7.97 | 7.38 | 8.63 |
| Pays de la Loire | Other genotypes | | 2023 | (59,66] | 223 | 7.62 | 6.21 | 6.28 | 3.14 | 9.42 | 6.25 | 6.24 | 5.67 | 6.86 |
| Provence-Alpes-Côte d'Azur | Other genotypes | | 2023 | (59,66] | 754 | 9.02 | 7.90 | 7.82 | 5.97 | 9.95 | 7.93 | 7.93 | 7.51 | 8.36 |
| Île-de-France | Other genotypes | | 2023 | (59,66] | 1,401 | 10.28 | 7.71 | 7.71 | 6.35 | 9.14 | 7.71 | 7.71 | 7.37 | 8.08 |
| | HPV16/18 | Opportunistic | 2020 | [30,39] | | | 3.86 | 3.86 | 3.59 | 4.15 | 3.86 | 3.86 | 3.70 | 4.03 |
| | HPV16/18 | Organised | 2020 | [30,39] | | | 3.49 | 0.00 | 0.00 | 16.67 | 3.73 | 3.69 | 2.76 | 4.92 |
| | HPV16/18 | Opportunistic | 2020 | (39,49] | 21,616 | 2.86 | 3.17 | 3.17 | 2.91 | 3.46 | 3.17 | 3.17 | 3.03 | 3.32 |
| | HPV16/18 | Organised | 2020 | (39,49] | 4 | 0.00 | 2.73 | 0.00 | 0.00 | 25.00 | 2.84 | 2.82 | 2.07 | 3.77 |
| | HPV16/18 | Opportunistic | 2020 | (49,59] | 16,489 | 2.36 | 2.53 | 2.53 | 2.27 | 2.81 | 2.53 | 2.53 | 2.41 | 2.66 |
| | HPV16/18 | Organised | 2020 | (49,59] | 9 | 0.00 | 3.21 | 0.00 | 0.00 | 22.22 | 3.07 | 3.03 | 2.32 | 4.00 |
| | HPV16/18 | Opportunistic | 2020 | (59,66] | 7,073 | 2.23 | 2.35 | 2.35 | 1.98 | 2.74 | 2.35 | 2.35 | 2.20 | 2.50 |
| | HPV16/18 | Organised | 2020 | (59,66] | 13 | 23.08 | 2.71 | 0.00 | 0.00 | 15.38 | 2.67 | 2.65 | 2.03 | 3.49 |
| | HPV16/18 | Opportunistic | 2021 | [30,39] | | | 4.46 | 4.46 | 4.24 | 4.68 | 4.46 | 4.46 | 4.33 | 4.59 |
| | HPV16/18 | Organised | 2021 | [30,39] | | | 3.80 | 3.81 | 2.47 | 5.25 | 3.78 | 3.77 | 3.15 | 4.52 |
| | HPV16/18 | Opportunistic | 2021 | (39,49] | 45,649 | 3.56 | 3.65 | 3.65 | 3.44 | 3.86 | 3.65 | 3.65 | 3.53 | 3.78 |
| | HPV16/18 | Organised | 2021 | (39,49] | 632 | 1.90 | 3.13 | 3.16 | 1.74 | 4.75 | 3.15 | 3.13 | 2.67 | 3.67 |
| | HPV16/18 | Opportunistic | 2021 | (49,59] | 35,896 | 2.83 | 2.91 | 2.91 | 2.71 | 3.13 | 2.91 | 2.91 | 2.80 | 3.03 |
| | HPV16/18 | Organised | 2021 | (49,59] | 971 | 2.47 | 2.55 | 2.57 | 1.54 | 3.60 | 2.57 | 2.56 | 2.21 | 2.97 |
| | HPV16/18 | Opportunistic | 2021 | (59,66] | 17,707 | 2.71 | 2.71 | 2.71 | 2.43 | 3.00 | 2.71 | 2.71 | 2.56 | 2.87 |
| | HPV16/18 | Organised | 2021 | (59,66] | 2,021 | 2.57 | 2.59 | 2.57 | 1.78 | 3.46 | 2.58 | 2.57 | 2.14 | 3.11 |
| | HPV16/18 | Opportunistic | 2022 | [30,39] | | | 4.89 | 4.89 | 4.56 | 5.21 | 4.88 | 4.88 | 4.70 | 5.07 |
| | HPV16/18 | Organised | 2022 | [30,39] | | | 3.62 | 3.60 | 2.74 | 4.61 | 3.63 | 3.62 | 3.18 | 4.14 |
| | HPV16/18 | Opportunistic | 2022 | (39,49] | 18,412 | 4.16 | 4.01 | 4.01 | 3.67 | 4.34 | 4.01 | 4.01 | 3.84 | 4.18 |
| | HPV16/18 | Organised | 2022 | (39,49] | 1,942 | 3.35 | 2.96 | 2.94 | 2.16 | 3.81 | 2.96 | 2.96 | 2.61 | 3.31 |
| | HPV16/18 | Opportunistic | 2022 | (49,59] | 14,453 | 3.11 | 3.19 | 3.19 | 2.86 | 3.51 | 3.19 | 3.19 | 3.04 | 3.34 |
| | HPV16/18 | Organised | 2022 | (49,59] | 2,225 | 2.25 | 2.49 | 2.47 | 1.80 | 3.19 | 2.49 | 2.48 | 2.20 | 2.79 |
| | HPV16/18 | Opportunistic | 2022 | (59,66] | 6,798 | 2.96 | 2.96 | 2.96 | 2.53 | 3.41 | 2.96 | 2.96 | 2.77 | 3.15 |
| | HPV16/18 | Organised | 2022 | (59,66] | 1,497 | 2.27 | 2.38 | 2.34 | 1.54 | 3.27 | 2.39 | 2.39 | 2.04 | 2.79 |
| | HPV16/18 | Opportunistic | 2023 | [30,39] | | | 5.44 | 5.45 | 5.07 | 5.82 | 5.45 | 5.44 | 5.23 | 5.66 |
| | HPV16/18 | Organised | 2023 | [30,39] | | | 3.62 | 3.61 | 2.71 | 4.56 | 3.62 | 3.62 | 3.12 | 4.15 |
| | HPV16/18 | Opportunistic | 2023 | (39,49] | 14,745 | 4.87 | 4.50 | 4.50 | 4.12 | 4.89 | 4.49 | 4.49 | 4.30 | 4.69 |
| | HPV16/18 | Organised | 2023 | (39,49] | 1,989 | 2.87 | 3.01 | 3.02 | 2.26 | 3.92 | 3.01 | 3.01 | 2.64 | 3.42 |
| | HPV16/18 | Opportunistic | 2023 | (49,59] | 11,789 | 3.93 | 3.59 | 3.59 | 3.22 | 3.97 | 3.59 | 3.59 | 3.42 | 3.77 |
| | HPV16/18 | Organised | 2023 | (49,59] | 1,992 | 2.61 | 2.47 | 2.46 | 1.71 | 3.26 | 2.48 | 2.48 | 2.15 | 2.86 |

**Table S8** **Observed HR HPV cervical infection prevalence, posterior predictive HR HPV cervical infection prevalence, and posterior expected HR HPV cervical infection prevalence, stratified by various dimensions.** ***(continued)***

| | | | | | | | Posterior distribution | | | | | | | |
|---|---|---|---|---|---|---|---|---|---|---|---|---|---|---|
| | | | | | | | Predictive prevalence | | | | Expected prevalence | | | |
| | | | | | | | | | ETI95% | | | | ETI95% | |
| Region | Genotypes | Type of screening | Year | Age | Number of tests | Observed | Average | Median | LB | UB | Average | Median | LB | UB |
| | HPV16/18 | Opportunistic | 2023 | (59,66] | 5,791 | 3.59 | 3.32 | 3.32 | 2.81 | 3.85 | 3.32 | 3.32 | 3.11 | 3.54 |
| | HPV16/18 | Organised | 2023 | (59,66] | 1,456 | 2.54 | 2.38 | 2.34 | 1.58 | 3.37 | 2.38 | 2.37 | 1.96 | 2.83 |
| | Other genotypes | Opportunistic | 2020 | [30,39] | | | 12.30 | 12.29 | 11.81 | 12.81 | 12.30 | 12.30 | 12.01 | 12.59 |
| | Other genotypes | Organised | 2020 | [30,39] | | | 8.88 | 8.33 | 0.00 | 25.00 | 9.08 | 9.05 | 7.36 | 11.12 |
| | Other genotypes | Opportunistic | 2020 | (39,49] | 21,616 | 8.42 | 8.64 | 8.64 | 8.19 | 9.07 | 8.64 | 8.64 | 8.41 | 8.87 |
| | Other genotypes | Organised | 2020 | (39,49] | 4 | 0.00 | 5.48 | 0.00 | 0.00 | 25.00 | 5.60 | 5.56 | 4.35 | 7.06 |
| | Other genotypes | Opportunistic | 2020 | (49,59] | 16,489 | 7.18 | 7.30 | 7.29 | 6.86 | 7.77 | 7.30 | 7.29 | 7.08 | 7.53 |
| | Other genotypes | Organised | 2020 | (49,59] | 9 | 11.11 | 6.39 | 0.00 | 0.00 | 22.22 | 6.52 | 6.48 | 5.25 | 8.00 |
| | Other genotypes | Opportunistic | 2020 | (59,66] | 7,073 | 5.91 | 6.48 | 6.48 | 5.85 | 7.11 | 6.47 | 6.47 | 6.21 | 6.74 |
| | Other genotypes | Organised | 2020 | (59,66] | 13 | 7.69 | 5.78 | 7.69 | 0.00 | 23.08 | 5.72 | 5.69 | 4.59 | 7.02 |
| | Other genotypes | Opportunistic | 2021 | [30,39] | | | 11.58 | 11.57 | 11.24 | 11.93 | 11.58 | 11.58 | 11.37 | 11.80 |
| | Other genotypes | Organised | 2021 | [30,39] | | | 8.77 | 8.74 | 6.79 | 10.91 | 8.79 | 8.76 | 7.80 | 9.90 |
| | Other genotypes | Opportunistic | 2021 | (39,49] | 45,649 | 8.07 | 8.12 | 8.12 | 7.83 | 8.43 | 8.12 | 8.12 | 7.94 | 8.30 |
| | Other genotypes | Organised | 2021 | (39,49] | 632 | 3.80 | 6.23 | 6.17 | 4.27 | 8.23 | 6.24 | 6.23 | 5.52 | 6.97 |
| | Other genotypes | Opportunistic | 2021 | (49,59] | 35,896 | 6.71 | 6.86 | 6.86 | 6.54 | 7.18 | 6.86 | 6.86 | 6.69 | 7.05 |
| | Other genotypes | Organised | 2021 | (49,59] | 971 | 5.77 | 5.70 | 5.66 | 4.22 | 7.31 | 5.71 | 5.71 | 5.13 | 6.31 |
| | Other genotypes | Opportunistic | 2021 | (59,66] | 17,707 | 6.05 | 6.12 | 6.12 | 5.72 | 6.55 | 6.13 | 6.13 | 5.89 | 6.37 |
| | Other genotypes | Organised | 2021 | (59,66] | 2,021 | 5.79 | 5.69 | 5.69 | 4.50 | 6.98 | 5.69 | 5.67 | 5.00 | 6.45 |
| | Other genotypes | Opportunistic | 2022 | [30,39] | | | 12.15 | 12.15 | 11.64 | 12.67 | 12.15 | 12.15 | 11.86 | 12.44 |
| | Other genotypes | Organised | 2022 | [30,39] | | | 9.07 | 9.08 | 7.64 | 10.57 | 9.07 | 9.06 | 8.30 | 9.91 |
| | Other genotypes | Opportunistic | 2022 | (39,49] | 18,412 | 8.37 | 8.47 | 8.47 | 8.01 | 8.94 | 8.48 | 8.48 | 8.25 | 8.71 |
| | Other genotypes | Organised | 2022 | (39,49] | 1,942 | 7.00 | 6.61 | 6.59 | 5.41 | 7.88 | 6.63 | 6.62 | 6.09 | 7.18 |
| | Other genotypes | Opportunistic | 2022 | (49,59] | 14,453 | 7.35 | 7.16 | 7.15 | 6.69 | 7.65 | 7.16 | 7.16 | 6.94 | 7.38 |
| | Other genotypes | Organised | 2022 | (49,59] | 2,225 | 6.56 | 6.17 | 6.20 | 5.08 | 7.33 | 6.17 | 6.17 | 5.66 | 6.70 |
| | Other genotypes | Opportunistic | 2022 | (59,66] | 6,798 | 6.18 | 6.38 | 6.37 | 5.77 | 7.05 | 6.38 | 6.38 | 6.11 | 6.65 |
| | Other genotypes | Organised | 2022 | (59,66] | 1,497 | 5.41 | 5.90 | 5.88 | 4.54 | 7.35 | 5.88 | 5.87 | 5.28 | 6.57 |
| | Other genotypes | Opportunistic | 2023 | [30,39] | | | 14.32 | 14.32 | 13.73 | 14.91 | 14.32 | 14.33 | 13.99 | 14.67 |
| | Other genotypes | Organised | 2023 | [30,39] | | | 9.08 | 9.08 | 7.65 | 10.65 | 9.12 | 9.11 | 8.29 | 9.99 |
| | Other genotypes | Opportunistic | 2023 | (39,49] | 14,745 | 10.06 | 9.99 | 9.99 | 9.45 | 10.55 | 9.99 | 9.99 | 9.70 | 10.29 |
| | Other genotypes | Organised | 2023 | (39,49] | 1,989 | 7.44 | 6.86 | 6.84 | 5.63 | 8.14 | 6.88 | 6.87 | 6.28 | 7.53 |
| | Other genotypes | Opportunistic | 2023 | (49,59] | 11,789 | 8.65 | 8.46 | 8.46 | 7.89 | 9.03 | 8.46 | 8.46 | 8.21 | 8.75 |
| | Other genotypes | Organised | 2023 | (49,59] | 1,992 | 5.52 | 6.29 | 6.28 | 5.12 | 7.53 | 6.28 | 6.28 | 5.72 | 6.87 |
| | Other genotypes | Opportunistic | 2023 | (59,66] | 5,791 | 8.51 | 7.51 | 7.51 | 6.77 | 8.24 | 7.51 | 7.51 | 7.20 | 7.83 |
| | Other genotypes | Organised | 2023 | (59,66] | 1,456 | 6.25 | 5.94 | 5.98 | 4.60 | 7.35 | 5.98 | 5.97 | 5.30 | 6.70 |
| Auvergne-Rhône-Alpes | HPV16/18 | Opportunistic | 2020 | | 988 | 3.44 | 3.46 | 3.44 | 2.33 | 4.76 | 3.48 | 3.47 | 3.14 | 3.83 |
| Bourgogne-Franche-Comté | HPV16/18 | Opportunistic | 2020 | | 537 | 2.98 | 3.08 | 2.98 | 1.68 | 4.66 | 3.05 | 3.04 | 2.73 | 3.39 |
| Bretagne | HPV16/18 | Opportunistic | 2020 | | 559 | 3.76 | 3.24 | 3.22 | 1.79 | 4.83 | 3.23 | 3.23 | 2.88 | 3.62 |
| Centre-Val de Loire | HPV16/18 | Opportunistic | 2020 | | 3,667 | 2.62 | 3.07 | 3.08 | 2.51 | 3.68 | 3.08 | 3.07 | 2.89 | 3.27 |
| Centre-Val de Loire | HPV16/18 | Organised | 2020 | | 3 | 33.33 | 3.13 | 0.00 | 0.00 | 33.33 | 2.98 | 2.93 | 2.15 | 4.04 |

**Table S8** **Observed HR HPV cervical infection prevalence, posterior predictive HR HPV cervical infection prevalence, and posterior expected HR HPV cervical infection prevalence, stratified by various dimensions.** ***(continued)***

| | | | | | | | Posterior distribution | | | | | | | |
|---|---|---|---|---|---|---|---|---|---|---|---|---|---|---|
| | | | | | | | Predictive prevalence | | | | Expected prevalence | | | |
| | | | | | | | | | ETI95% | | | | ETI95% | |
| Region | Genotypes | Type of screening | Year | Age | Number of tests | Observed | Average | Median | LB | UB | Average | Median | LB | UB |
| Corse | HPV16/18 | Opportunistic | 2020 | | 377 | 3.45 | 3.36 | 3.18 | 1.59 | 5.57 | 3.32 | 3.30 | 2.55 | 4.28 |
| Grand Est | HPV16/18 | Opportunistic | 2020 | | 882 | 3.17 | 2.77 | 2.72 | 1.70 | 3.97 | 2.77 | 2.77 | 2.48 | 3.08 |
| Hauts-de-France | HPV16/18 | Opportunistic | 2020 | | 2,833 | 3.46 | 3.01 | 3.00 | 2.36 | 3.71 | 3.02 | 3.01 | 2.83 | 3.22 |
| Hauts-de-France | HPV16/18 | Organised | 2020 | | 7 | 14.29 | 2.78 | 0.00 | 0.00 | 14.29 | 2.84 | 2.81 | 2.13 | 3.74 |
| Normandie | HPV16/18 | Opportunistic | 2020 | | 2,313 | 3.67 | 3.30 | 3.29 | 2.55 | 4.11 | 3.30 | 3.30 | 3.06 | 3.54 |
| Nouvelle-Aquitaine | HPV16/18 | Opportunistic | 2020 | | 1,688 | 3.38 | 3.16 | 3.14 | 2.31 | 4.09 | 3.16 | 3.16 | 2.95 | 3.38 |
| Nouvelle-Aquitaine | HPV16/18 | Organised | 2020 | | 1 | 0.00 | 2.33 | 0.00 | 0.00 | 0.00 | 2.52 | 2.43 | 1.46 | 4.06 |
| Occitanie | HPV16/18 | Opportunistic | 2020 | | 2,175 | 4.23 | 3.91 | 3.91 | 3.03 | 4.83 | 3.92 | 3.91 | 3.56 | 4.31 |
| Occitanie | HPV16/18 | Organised | 2020 | | 4 | 0.00 | 3.35 | 0.00 | 0.00 | 25.00 | 3.19 | 3.13 | 2.18 | 4.54 |
| Pays de la Loire | HPV16/18 | Opportunistic | 2020 | | 1,156 | 1.73 | 2.65 | 2.68 | 1.73 | 3.63 | 2.67 | 2.66 | 2.40 | 2.96 |
| Pays de la Loire | HPV16/18 | Organised | 2020 | | 5 | 0.00 | 2.57 | 0.00 | 0.00 | 20.00 | 2.80 | 2.75 | 2.00 | 3.77 |
| Provence-Alpes-Côte d'Azur | HPV16/18 | Opportunistic | 2020 | | 2,129 | 3.05 | 3.24 | 3.24 | 2.49 | 4.04 | 3.24 | 3.23 | 3.00 | 3.49 |
| Provence-Alpes-Côte d'Azur | HPV16/18 | Organised | 2020 | | 2 | 50.00 | 2.80 | 0.00 | 0.00 | 50.00 | 2.71 | 2.66 | 1.88 | 3.84 |
| Île-de-France | HPV16/18 | Opportunistic | 2020 | | 51,739 | 3.07 | 3.19 | 3.18 | 3.00 | 3.39 | 3.19 | 3.19 | 3.06 | 3.32 |
| Île-de-France | HPV16/18 | Organised | 2020 | | 16 | 0.00 | 3.37 | 0.00 | 0.00 | 12.50 | 3.44 | 3.41 | 2.64 | 4.44 |
| Auvergne-Rhône-Alpes | HPV16/18 | Opportunistic | 2021 | | 2,995 | 3.47 | 4.06 | 4.04 | 3.31 | 4.84 | 4.06 | 4.06 | 3.72 | 4.42 |
| Auvergne-Rhône-Alpes | HPV16/18 | Organised | 2021 | | 152 | 1.32 | 3.56 | 3.29 | 0.66 | 6.58 | 3.54 | 3.52 | 2.93 | 4.23 |
| Bourgogne-Franche-Comté | HPV16/18 | Opportunistic | 2021 | | 1,510 | 3.91 | 3.59 | 3.58 | 2.65 | 4.64 | 3.60 | 3.59 | 3.26 | 3.97 |
| Bourgogne-Franche-Comté | HPV16/18 | Organised | 2021 | | 27 | 7.41 | 2.58 | 3.70 | 0.00 | 11.11 | 2.64 | 2.63 | 2.15 | 3.22 |
| Bretagne | HPV16/18 | Opportunistic | 2021 | | 2,260 | 3.41 | 3.66 | 3.67 | 2.83 | 4.51 | 3.66 | 3.65 | 3.32 | 4.02 |
| Bretagne | HPV16/18 | Organised | 2021 | | 114 | 4.39 | 2.46 | 2.63 | 0.00 | 5.26 | 2.47 | 2.45 | 1.94 | 3.11 |
| Centre-Val de Loire | HPV16/18 | Opportunistic | 2021 | | 14,308 | 3.39 | 3.50 | 3.49 | 3.15 | 3.86 | 3.50 | 3.50 | 3.33 | 3.68 |
| Centre-Val de Loire | HPV16/18 | Organised | 2021 | | 721 | 2.64 | 2.91 | 2.91 | 1.66 | 4.30 | 2.92 | 2.91 | 2.51 | 3.39 |
| Corse | HPV16/18 | Opportunistic | 2021 | | 34 | 2.94 | 3.77 | 2.94 | 0.00 | 11.76 | 3.80 | 3.79 | 3.29 | 4.39 |
| Corse | HPV16/18 | Organised | 2021 | | 1 | 0.00 | 2.37 | 0.00 | 0.00 | 0.00 | 2.84 | 2.77 | 1.84 | 4.21 |
| Grand Est | HPV16/18 | Opportunistic | 2021 | | 3,787 | 3.22 | 3.38 | 3.38 | 2.77 | 4.01 | 3.37 | 3.37 | 3.13 | 3.62 |
| Grand Est | HPV16/18 | Organised | 2021 | | 136 | 0.74 | 2.41 | 2.21 | 0.00 | 5.15 | 2.43 | 2.43 | 2.01 | 2.92 |
| Hauts-de-France | HPV16/18 | Opportunistic | 2021 | | 10,358 | 3.72 | 3.57 | 3.57 | 3.18 | 3.97 | 3.57 | 3.57 | 3.39 | 3.75 |
| Hauts-de-France | HPV16/18 | Organised | 2021 | | 390 | 2.31 | 2.98 | 2.82 | 1.28 | 4.87 | 3.01 | 3.00 | 2.56 | 3.51 |
| Normandie | HPV16/18 | Opportunistic | 2021 | | 8,045 | 4.29 | 3.94 | 3.93 | 3.46 | 4.44 | 3.93 | 3.93 | 3.69 | 4.17 |
| Normandie | HPV16/18 | Organised | 2021 | | 34 | 5.88 | 3.03 | 2.94 | 0.00 | 8.82 | 2.90 | 2.89 | 2.40 | 3.48 |
| Nouvelle-Aquitaine | HPV16/18 | Opportunistic | 2021 | | 8,526 | 3.48 | 3.58 | 3.58 | 3.16 | 4.02 | 3.58 | 3.59 | 3.39 | 3.78 |
| Nouvelle-Aquitaine | HPV16/18 | Organised | 2021 | | 391 | 4.09 | 2.66 | 2.56 | 1.28 | 4.35 | 2.65 | 2.64 | 2.27 | 3.09 |
| Occitanie | HPV16/18 | Opportunistic | 2021 | | 5,534 | 4.03 | 4.09 | 4.08 | 3.51 | 4.70 | 4.09 | 4.09 | 3.83 | 4.37 |
| Occitanie | HPV16/18 | Organised | 2021 | | 305 | 3.61 | 3.67 | 3.61 | 1.64 | 6.23 | 3.66 | 3.66 | 3.06 | 4.36 |
| Pays de la Loire | HPV16/18 | Opportunistic | 2021 | | 2,811 | 3.34 | 3.11 | 3.09 | 2.45 | 3.84 | 3.10 | 3.10 | 2.82 | 3.39 |
| Pays de la Loire | HPV16/18 | Organised | 2021 | | 270 | 1.85 | 2.57 | 2.59 | 0.74 | 4.81 | 2.58 | 2.56 | 2.12 | 3.12 |
| Provence-Alpes-Côte d'Azur | HPV16/18 | Opportunistic | 2021 | | 11,257 | 3.82 | 3.89 | 3.88 | 3.46 | 4.32 | 3.88 | 3.88 | 3.67 | 4.11 |
| Provence-Alpes-Côte d'Azur | HPV16/18 | Organised | 2021 | | 56 | 1.79 | 3.49 | 3.57 | 0.00 | 8.93 | 3.50 | 3.49 | 2.91 | 4.23 |

**Table S8** **Observed HR HPV cervical infection prevalence, posterior predictive HR HPV cervical infection prevalence, and posterior expected HR HPV cervical infection prevalence, stratified by various dimensions.** ***(continued)***

| | | | | | | | Posterior distribution | | | | | | | |
|---|---|---|---|---|---|---|---|---|---|---|---|---|---|---|
| | | | | | | | Predictive prevalence | | | | Expected prevalence | | | |
| | | | | | | | | | ETI95% | | | | ETI95% | |
| Region | Genotypes | Type of screening | Year | Age | Number of tests | Observed | Average | Median | LB | UB | Average | Median | LB | UB |
| Île-de-France | HPV16/18 | Opportunistic | 2021 | | 83,161 | 3.67 | 3.64 | 3.64 | 3.48 | 3.80 | 3.64 | 3.64 | 3.53 | 3.75 |
| Île-de-France | HPV16/18 | Organised | 2021 | | 1,999 | 2.95 | 2.87 | 2.85 | 2.05 | 3.80 | 2.87 | 2.85 | 2.41 | 3.38 |
| Auvergne-Rhône-Alpes | HPV16/18 | Opportunistic | 2022 | | 247 | 7.29 | 4.85 | 4.86 | 2.43 | 7.69 | 4.83 | 4.82 | 4.33 | 5.42 |
| Auvergne-Rhône-Alpes | HPV16/18 | Organised | 2022 | | 48 | 14.58 | 3.51 | 4.17 | 0.00 | 8.33 | 3.54 | 3.52 | 2.92 | 4.27 |
| Bourgogne-Franche-Comté | HPV16/18 | Opportunistic | 2022 | | 1,175 | 3.40 | 3.94 | 3.91 | 2.81 | 5.19 | 3.93 | 3.92 | 3.53 | 4.37 |
| Bourgogne-Franche-Comté | HPV16/18 | Organised | 2022 | | 45 | 2.22 | 2.94 | 2.22 | 0.00 | 8.89 | 3.03 | 3.01 | 2.50 | 3.65 |
| Bretagne | HPV16/18 | Opportunistic | 2022 | | 2,496 | 3.85 | 4.22 | 4.21 | 3.41 | 5.09 | 4.22 | 4.21 | 3.87 | 4.61 |
| Bretagne | HPV16/18 | Organised | 2022 | | 586 | 2.05 | 2.49 | 2.39 | 1.19 | 3.92 | 2.50 | 2.49 | 2.06 | 3.00 |
| Centre-Val de Loire | HPV16/18 | Opportunistic | 2022 | | 7,059 | 4.32 | 3.75 | 3.75 | 3.27 | 4.25 | 3.76 | 3.76 | 3.55 | 3.98 |
| Centre-Val de Loire | HPV16/18 | Organised | 2022 | | 918 | 3.70 | 2.70 | 2.72 | 1.63 | 3.92 | 2.72 | 2.71 | 2.41 | 3.04 |
| Corse | HPV16/18 | Opportunistic | 2022 | | 11 | 0.00 | 4.67 | 0.00 | 0.00 | 18.18 | 4.55 | 4.52 | 3.86 | 5.40 |
| Corse | HPV16/18 | Organised | 2022 | | 2 | 0.00 | 3.17 | 0.00 | 0.00 | 50.00 | 3.15 | 3.11 | 2.28 | 4.23 |
| Grand Est | HPV16/18 | Opportunistic | 2022 | | 3,082 | 3.89 | 3.83 | 3.83 | 3.11 | 4.61 | 3.83 | 3.83 | 3.52 | 4.15 |
| Grand Est | HPV16/18 | Organised | 2022 | | 622 | 3.05 | 2.82 | 2.73 | 1.61 | 4.34 | 2.81 | 2.81 | 2.40 | 3.26 |
| Hauts-de-France | HPV16/18 | Opportunistic | 2022 | | 8,863 | 3.69 | 3.89 | 3.88 | 3.43 | 4.34 | 3.89 | 3.89 | 3.68 | 4.10 |
| Hauts-de-France | HPV16/18 | Organised | 2022 | | 1,080 | 2.22 | 2.80 | 2.78 | 1.85 | 3.89 | 2.81 | 2.81 | 2.47 | 3.18 |
| Normandie | HPV16/18 | Opportunistic | 2022 | | 5,759 | 4.43 | 4.43 | 4.43 | 3.85 | 5.05 | 4.43 | 4.43 | 4.15 | 4.71 |
| Normandie | HPV16/18 | Organised | 2022 | | 153 | 1.31 | 2.67 | 2.61 | 0.65 | 5.23 | 2.69 | 2.68 | 2.30 | 3.10 |
| Nouvelle-Aquitaine | HPV16/18 | Opportunistic | 2022 | | 9,232 | 4.09 | 3.98 | 3.98 | 3.54 | 4.46 | 3.98 | 3.98 | 3.75 | 4.20 |
| Nouvelle-Aquitaine | HPV16/18 | Organised | 2022 | | 1,733 | 2.25 | 2.79 | 2.77 | 1.96 | 3.64 | 2.79 | 2.79 | 2.47 | 3.14 |
| Occitanie | HPV16/18 | Opportunistic | 2022 | | 2,624 | 4.27 | 4.23 | 4.23 | 3.43 | 5.11 | 4.23 | 4.23 | 3.92 | 4.54 |
| Occitanie | HPV16/18 | Organised | 2022 | | 507 | 2.17 | 3.28 | 3.16 | 1.78 | 4.93 | 3.30 | 3.29 | 2.87 | 3.80 |
| Pays de la Loire | HPV16/18 | Opportunistic | 2022 | | 721 | 2.91 | 3.70 | 3.61 | 2.36 | 5.27 | 3.66 | 3.66 | 3.33 | 4.03 |
| Pays de la Loire | HPV16/18 | Organised | 2022 | | 241 | 1.24 | 2.51 | 2.49 | 0.83 | 4.56 | 2.49 | 2.49 | 2.13 | 2.88 |
| Provence-Alpes-Côte d'Azur | HPV16/18 | Opportunistic | 2022 | | 7,734 | 4.38 | 4.27 | 4.27 | 3.75 | 4.80 | 4.27 | 4.26 | 4.02 | 4.52 |
| Provence-Alpes-Côte d'Azur | HPV16/18 | Organised | 2022 | | 504 | 5.36 | 3.85 | 3.77 | 2.18 | 5.75 | 3.86 | 3.84 | 3.27 | 4.55 |
| Île-de-France | HPV16/18 | Opportunistic | 2022 | | 15,516 | 3.74 | 4.08 | 4.08 | 3.71 | 4.43 | 4.07 | 4.07 | 3.89 | 4.26 |
| Île-de-France | HPV16/18 | Organised | 2022 | | 1,307 | 2.91 | 2.98 | 2.98 | 1.99 | 3.98 | 2.98 | 2.97 | 2.63 | 3.38 |
| Auvergne-Rhône-Alpes | HPV16/18 | Opportunistic | 2023 | | 192 | 5.73 | 5.57 | 5.21 | 2.60 | 9.38 | 5.56 | 5.54 | 4.78 | 6.47 |
| Auvergne-Rhône-Alpes | HPV16/18 | Organised | 2023 | | 49 | 4.08 | 3.81 | 4.08 | 0.00 | 10.20 | 3.90 | 3.87 | 3.11 | 4.82 |
| Bourgogne-Franche-Comté | HPV16/18 | Opportunistic | 2023 | | 551 | 4.72 | 4.32 | 4.36 | 2.54 | 6.17 | 4.31 | 4.31 | 3.81 | 4.87 |
| Bourgogne-Franche-Comté | HPV16/18 | Organised | 2023 | | 190 | 1.58 | 2.76 | 2.63 | 0.53 | 5.26 | 2.72 | 2.71 | 2.20 | 3.33 |
| Bretagne | HPV16/18 | Opportunistic | 2023 | | 3,450 | 5.19 | 4.74 | 4.72 | 3.97 | 5.57 | 4.74 | 4.73 | 4.35 | 5.16 |
| Bretagne | HPV16/18 | Organised | 2023 | | 1,597 | 2.44 | 2.58 | 2.57 | 1.69 | 3.57 | 2.59 | 2.57 | 2.15 | 3.11 |
| Centre-Val de Loire | HPV16/18 | Opportunistic | 2023 | | 5,511 | 4.59 | 4.27 | 4.26 | 3.70 | 4.86 | 4.27 | 4.27 | 4.03 | 4.52 |
| Centre-Val de Loire | HPV16/18 | Organised | 2023 | | 562 | 3.20 | 2.79 | 2.85 | 1.42 | 4.27 | 2.79 | 2.78 | 2.42 | 3.21 |
| Corse | HPV16/18 | Opportunistic | 2023 | | 5 | 20.00 | 3.77 | 0.00 | 0.00 | 20.00 | 4.08 | 4.02 | 3.08 | 5.35 |
| Grand Est | HPV16/18 | Opportunistic | 2023 | | 2,957 | 4.26 | 4.23 | 4.23 | 3.45 | 5.07 | 4.23 | 4.22 | 3.90 | 4.58 |
| Grand Est | HPV16/18 | Organised | 2023 | | 420 | 2.38 | 2.63 | 2.62 | 1.19 | 4.29 | 2.64 | 2.63 | 2.22 | 3.13 |

**Table S8 Observed HR HPV cervical infection prevalence, posterior predictive HR HPV cervical infection prevalence, and posterior expected HR HPV cervical infection prevalence, stratified by various dimensions.** ***(continued)***

| | | | | | | | Posterior distribution | | | | | | | |
|---|---|---|---|---|---|---|---|---|---|---|---|---|---|---|
| | | | | | | | Predictive prevalence | | | | Expected prevalence | | | |
| | | | | | | | | | ETI95% | | | | ETI95% | |
| Region | Genotypes | Type of screening | Year | Age | Number of tests | Observed | Average | Median | LB | UB | Average | Median | LB | UB |
| Hauts-de-France | HPV16/18 | Opportunistic | 2023 | | 7,934 | 4.16 | 4.28 | 4.27 | 3.77 | 4.79 | 4.28 | 4.27 | 4.04 | 4.52 |
| Hauts-de-France | HPV16/18 | Organised | 2023 | | 473 | 2.96 | 2.73 | 2.75 | 1.27 | 4.23 | 2.72 | 2.71 | 2.34 | 3.16 |
| Normandie | HPV16/18 | Opportunistic | 2023 | | 3,472 | 4.87 | 4.90 | 4.90 | 4.12 | 5.73 | 4.91 | 4.91 | 4.58 | 5.25 |
| Normandie | HPV16/18 | Organised | 2023 | | 187 | 2.14 | 2.91 | 2.67 | 0.53 | 5.88 | 2.91 | 2.91 | 2.49 | 3.39 |
| Nouvelle-Aquitaine | HPV16/18 | Opportunistic | 2023 | | 7,539 | 4.24 | 4.48 | 4.48 | 3.97 | 5.03 | 4.49 | 4.49 | 4.23 | 4.75 |
| Nouvelle-Aquitaine | HPV16/18 | Organised | 2023 | | 1,611 | 2.98 | 3.03 | 3.04 | 2.17 | 3.97 | 3.05 | 3.04 | 2.65 | 3.48 |
| Occitanie | HPV16/18 | Opportunistic | 2023 | | 1,178 | 4.84 | 4.70 | 4.67 | 3.40 | 6.03 | 4.71 | 4.70 | 4.31 | 5.12 |
| Occitanie | HPV16/18 | Organised | 2023 | | 197 | 2.03 | 3.20 | 3.05 | 1.02 | 6.09 | 3.21 | 3.20 | 2.71 | 3.76 |
| Pays de la Loire | HPV16/18 | Opportunistic | 2023 | | 1,003 | 4.09 | 3.98 | 3.99 | 2.79 | 5.28 | 4.00 | 3.99 | 3.63 | 4.42 |
| Pays de la Loire | HPV16/18 | Organised | 2023 | | 498 | 3.21 | 2.55 | 2.41 | 1.20 | 4.22 | 2.55 | 2.55 | 2.16 | 3.00 |
| Provence-Alpes-Côte d'Azur | HPV16/18 | Opportunistic | 2023 | | 5,645 | 4.62 | 4.80 | 4.80 | 4.20 | 5.46 | 4.80 | 4.80 | 4.52 | 5.11 |
| Provence-Alpes-Côte d'Azur | HPV16/18 | Organised | 2023 | | 562 | 4.09 | 3.70 | 3.74 | 2.14 | 5.52 | 3.69 | 3.67 | 3.09 | 4.38 |
| Île-de-France | HPV16/18 | Opportunistic | 2023 | | 13,457 | 4.58 | 4.65 | 4.64 | 4.23 | 5.08 | 4.64 | 4.64 | 4.43 | 4.87 |
| Île-de-France | HPV16/18 | Organised | 2023 | | 1,195 | 3.51 | 3.17 | 3.18 | 2.09 | 4.27 | 3.17 | 3.15 | 2.74 | 3.67 |
| Auvergne-Rhône-Alpes | Other genotypes | Opportunistic | 2020 | | 988 | 9.11 | 10.01 | 10.02 | 8.00 | 12.15 | 10.01 | 10.00 | 9.25 | 10.80 |
| Bourgogne-Franche-Comté | Other genotypes | Opportunistic | 2020 | | 537 | 9.50 | 8.73 | 8.75 | 6.33 | 11.17 | 8.74 | 8.73 | 8.04 | 9.50 |
| Bretagne | Other genotypes | Opportunistic | 2020 | | 559 | 8.77 | 8.59 | 8.59 | 6.26 | 11.09 | 8.61 | 8.61 | 7.87 | 9.40 |
| Centre-Val de Loire | Other genotypes | Opportunistic | 2020 | | 3,667 | 7.06 | 8.40 | 8.40 | 7.42 | 9.38 | 8.39 | 8.39 | 8.03 | 8.76 |
| Centre-Val de Loire | Other genotypes | Organised | 2020 | | 3 | 33.33 | 6.02 | 0.00 | 0.00 | 33.33 | 6.03 | 5.96 | 4.51 | 7.84 |
| Corse | Other genotypes | Opportunistic | 2020 | | 377 | 10.61 | 10.26 | 10.08 | 6.90 | 14.06 | 10.24 | 10.19 | 8.28 | 12.60 |
| Grand Est | Other genotypes | Opportunistic | 2020 | | 882 | 7.82 | 8.00 | 7.94 | 6.24 | 9.98 | 8.01 | 8.00 | 7.32 | 8.75 |
| Hauts-de-France | Other genotypes | Opportunistic | 2020 | | 2,833 | 8.08 | 8.57 | 8.58 | 7.48 | 9.67 | 8.56 | 8.55 | 8.18 | 8.95 |
| Hauts-de-France | Other genotypes | Organised | 2020 | | 7 | 0.00 | 6.22 | 0.00 | 0.00 | 28.57 | 6.12 | 6.09 | 4.88 | 7.58 |
| Normandie | Other genotypes | Opportunistic | 2020 | | 2,313 | 9.21 | 8.80 | 8.78 | 7.57 | 10.07 | 8.80 | 8.80 | 8.35 | 9.27 |
| Nouvelle-Aquitaine | Other genotypes | Opportunistic | 2020 | | 1,688 | 8.95 | 9.15 | 9.12 | 7.76 | 10.60 | 9.16 | 9.16 | 8.77 | 9.57 |
| Nouvelle-Aquitaine | Other genotypes | Organised | 2020 | | 1 | 0.00 | 6.10 | 0.00 | 0.00 | 100.00 | 5.80 | 5.65 | 3.58 | 8.90 |
| Occitanie | Other genotypes | Opportunistic | 2020 | | 2,175 | 9.89 | 10.93 | 10.94 | 9.47 | 12.46 | 10.92 | 10.92 | 10.13 | 11.73 |
| Occitanie | Other genotypes | Organised | 2020 | | 4 | 25.00 | 6.53 | 0.00 | 0.00 | 25.00 | 6.83 | 6.76 | 4.88 | 9.26 |
| Pays de la Loire | Other genotypes | Opportunistic | 2020 | | 1,156 | 6.66 | 7.26 | 7.27 | 5.62 | 8.91 | 7.27 | 7.25 | 6.65 | 7.92 |
| Pays de la Loire | Other genotypes | Organised | 2020 | | 5 | 0.00 | 5.35 | 0.00 | 0.00 | 40.00 | 5.50 | 5.44 | 4.24 | 7.05 |
| Provence-Alpes-Côte d'Azur | Other genotypes | Opportunistic | 2020 | | 2,129 | 9.63 | 9.41 | 9.39 | 8.13 | 10.76 | 9.41 | 9.41 | 8.92 | 9.92 |
| Provence-Alpes-Côte d'Azur | Other genotypes | Organised | 2020 | | 2 | 0.00 | 5.45 | 0.00 | 0.00 | 50.00 | 5.75 | 5.66 | 4.15 | 7.76 |
| Île-de-France | Other genotypes | Opportunistic | 2020 | | 51,739 | 9.68 | 9.62 | 9.61 | 9.29 | 9.96 | 9.62 | 9.61 | 9.41 | 9.84 |
| Île-de-France | Other genotypes | Organised | 2020 | | 16 | 0.00 | 8.10 | 6.25 | 0.00 | 25.00 | 8.22 | 8.18 | 6.74 | 9.90 |
| Auvergne-Rhône-Alpes | Other genotypes | Opportunistic | 2021 | | 2,995 | 9.12 | 9.67 | 9.65 | 8.41 | 10.98 | 9.67 | 9.66 | 9.00 | 10.38 |
| Auvergne-Rhône-Alpes | Other genotypes | Organised | 2021 | | 152 | 9.21 | 7.43 | 7.24 | 3.29 | 11.84 | 7.52 | 7.51 | 6.44 | 8.59 |
| Bourgogne-Franche-Comté | Other genotypes | Opportunistic | 2021 | | 1,510 | 9.21 | 8.61 | 8.61 | 7.02 | 10.26 | 8.60 | 8.60 | 7.95 | 9.31 |
| Bourgogne-Franche-Comté | Other genotypes | Organised | 2021 | | 27 | 11.11 | 5.68 | 3.70 | 0.00 | 14.81 | 5.89 | 5.87 | 4.97 | 6.92 |
| Bretagne | Other genotypes | Opportunistic | 2021 | | 2,260 | 7.26 | 8.10 | 8.10 | 6.81 | 9.42 | 8.08 | 8.08 | 7.51 | 8.66 |

**Table S8 Observed HR HPV cervical infection prevalence, posterior predictive HR HPV cervical infection prevalence, and posterior expected HR HPV cervical infection prevalence, stratified by various dimensions. *(continued)***

| | | | | | | | Posterior distribution | | | | | | | |
|---|---|---|---|---|---|---|---|---|---|---|---|---|---|---|
| | | | | | | | Predictive prevalence | | | | Expected prevalence | | | |
| | | | | | | | | | ETI95% | | | | ETI95% | |
| Region | Genotypes | Type of screening | Year | Age | Number of tests | Observed | Average | Median | LB | UB | Average | Median | LB | UB |
| Bretagne | Other genotypes | Organised | 2021 | | 114 | 2.63 | 5.24 | 5.26 | 1.75 | 9.65 | 5.23 | 5.21 | 4.33 | 6.25 |
| Centre-Val de Loire | Other genotypes | Opportunistic | 2021 | | 14,308 | 7.68 | 7.93 | 7.92 | 7.43 | 8.46 | 7.93 | 7.93 | 7.64 | 8.22 |
| Centre-Val de Loire | Other genotypes | Organised | 2021 | | 721 | 5.55 | 6.24 | 6.24 | 4.58 | 8.18 | 6.22 | 6.22 | 5.59 | 6.91 |
| Corse | Other genotypes | Opportunistic | 2021 | | 34 | 8.82 | 9.07 | 8.82 | 0.00 | 20.59 | 9.16 | 9.13 | 8.01 | 10.50 |
| Corse | Other genotypes | Organised | 2021 | | 1 | 0.00 | 6.40 | 0.00 | 0.00 | 100.00 | 6.17 | 6.05 | 4.07 | 8.97 |
| Grand Est | Other genotypes | Opportunistic | 2021 | | 3,787 | 7.10 | 7.87 | 7.84 | 6.97 | 8.82 | 7.88 | 7.87 | 7.45 | 8.34 |
| Grand Est | Other genotypes | Organised | 2021 | | 136 | 2.94 | 5.35 | 5.15 | 2.21 | 9.56 | 5.36 | 5.35 | 4.58 | 6.16 |
| Hauts-de-France | Other genotypes | Opportunistic | 2021 | | 10,358 | 7.81 | 8.34 | 8.33 | 7.74 | 8.96 | 8.34 | 8.34 | 8.05 | 8.65 |
| Hauts-de-France | Other genotypes | Organised | 2021 | | 390 | 6.15 | 6.45 | 6.41 | 3.85 | 8.97 | 6.43 | 6.42 | 5.71 | 7.19 |
| Normandie | Other genotypes | Opportunistic | 2021 | | 8,045 | 8.39 | 8.60 | 8.60 | 7.89 | 9.30 | 8.59 | 8.59 | 8.23 | 8.96 |
| Normandie | Other genotypes | Organised | 2021 | | 34 | 8.82 | 5.93 | 5.88 | 0.00 | 14.71 | 6.00 | 5.99 | 5.18 | 6.85 |
| Nouvelle-Aquitaine | Other genotypes | Opportunistic | 2021 | | 8,526 | 8.40 | 8.50 | 8.49 | 7.83 | 9.17 | 8.50 | 8.50 | 8.18 | 8.83 |
| Nouvelle-Aquitaine | Other genotypes | Organised | 2021 | | 391 | 6.65 | 6.11 | 6.14 | 3.83 | 8.70 | 6.14 | 6.13 | 5.45 | 6.84 |
| Occitanie | Other genotypes | Opportunistic | 2021 | | 5,534 | 9.78 | 9.56 | 9.56 | 8.64 | 10.46 | 9.56 | 9.56 | 9.09 | 10.05 |
| Occitanie | Other genotypes | Organised | 2021 | | 305 | 8.52 | 7.70 | 7.54 | 4.92 | 10.82 | 7.75 | 7.73 | 6.75 | 8.84 |
| Pays de la Loire | Other genotypes | Opportunistic | 2021 | | 2,811 | 6.19 | 7.03 | 7.04 | 5.98 | 8.11 | 7.03 | 7.02 | 6.53 | 7.58 |
| Pays de la Loire | Other genotypes | Organised | 2021 | | 270 | 4.44 | 5.24 | 5.19 | 2.59 | 8.15 | 5.25 | 5.24 | 4.49 | 6.07 |
| Provence-Alpes-Côte d'Azur | Other genotypes | Opportunistic | 2021 | | 11,257 | 9.16 | 9.14 | 9.14 | 8.55 | 9.79 | 9.14 | 9.14 | 8.80 | 9.50 |
| Provence-Alpes-Côte d'Azur | Other genotypes | Organised | 2021 | | 56 | 7.14 | 7.97 | 7.14 | 1.79 | 16.07 | 7.94 | 7.90 | 6.85 | 9.19 |
| Île-de-France | Other genotypes | Opportunistic | 2021 | | 83,161 | 9.32 | 9.12 | 9.12 | 8.87 | 9.38 | 9.12 | 9.12 | 8.96 | 9.30 |
| Île-de-France | Other genotypes | Organised | 2021 | | 1,999 | 6.40 | 6.53 | 6.50 | 5.25 | 7.90 | 6.54 | 6.52 | 5.81 | 7.35 |
| Auvergne-Rhône-Alpes | Other genotypes | Opportunistic | 2022 | | 247 | 19.43 | 11.17 | 10.93 | 7.29 | 15.38 | 11.22 | 11.21 | 10.20 | 12.28 |
| Auvergne-Rhône-Alpes | Other genotypes | Organised | 2022 | | 48 | 12.50 | 8.40 | 8.33 | 2.08 | 16.67 | 8.38 | 8.36 | 7.12 | 9.76 |
| Bourgogne-Franche-Comté | Other genotypes | Opportunistic | 2022 | | 1,175 | 9.36 | 9.24 | 9.19 | 7.49 | 11.15 | 9.25 | 9.24 | 8.51 | 10.03 |
| Bourgogne-Franche-Comté | Other genotypes | Organised | 2022 | | 45 | 4.44 | 7.83 | 6.67 | 2.17 | 15.56 | 7.89 | 7.88 | 6.67 | 9.21 |
| Bretagne | Other genotypes | Opportunistic | 2022 | | 2,496 | 9.70 | 9.24 | 9.25 | 7.93 | 10.58 | 9.23 | 9.23 | 8.62 | 9.85 |
| Bretagne | Other genotypes | Organised | 2022 | | 586 | 4.44 | 5.79 | 5.80 | 3.92 | 8.02 | 5.81 | 5.77 | 4.95 | 6.78 |
| Centre-Val de Loire | Other genotypes | Opportunistic | 2022 | | 7,059 | 8.85 | 8.39 | 8.40 | 7.65 | 9.12 | 8.39 | 8.39 | 8.05 | 8.72 |
| Centre-Val de Loire | Other genotypes | Organised | 2022 | | 918 | 7.08 | 6.33 | 6.32 | 4.68 | 8.06 | 6.32 | 6.32 | 5.77 | 6.91 |
| Corse | Other genotypes | Opportunistic | 2022 | | 11 | 27.27 | 10.19 | 9.09 | 0.00 | 27.27 | 9.88 | 9.85 | 8.51 | 11.43 |
| Corse | Other genotypes | Organised | 2022 | | 2 | 0.00 | 6.48 | 0.00 | 0.00 | 50.00 | 6.92 | 6.82 | 5.15 | 9.08 |
| Grand Est | Other genotypes | Opportunistic | 2022 | | 3,082 | 9.34 | 8.90 | 8.89 | 7.75 | 10.06 | 8.90 | 8.89 | 8.39 | 9.45 |
| Grand Est | Other genotypes | Organised | 2022 | | 622 | 7.88 | 6.69 | 6.59 | 4.82 | 8.84 | 6.70 | 6.69 | 5.89 | 7.54 |
| Hauts-de-France | Other genotypes | Opportunistic | 2022 | | 8,863 | 9.20 | 8.97 | 8.97 | 8.29 | 9.65 | 8.97 | 8.96 | 8.64 | 9.30 |
| Hauts-de-France | Other genotypes | Organised | 2022 | | 1,080 | 6.48 | 6.63 | 6.57 | 5.09 | 8.24 | 6.62 | 6.61 | 6.00 | 7.30 |
| Normandie | Other genotypes | Opportunistic | 2022 | | 5,759 | 9.20 | 9.46 | 9.45 | 8.56 | 10.38 | 9.45 | 9.45 | 9.03 | 9.87 |
| Normandie | Other genotypes | Organised | 2022 | | 153 | 4.58 | 6.32 | 5.88 | 2.61 | 10.46 | 6.26 | 6.25 | 5.55 | 7.03 |
| Nouvelle-Aquitaine | Other genotypes | Opportunistic | 2022 | | 9,232 | 8.99 | 9.25 | 9.25 | 8.57 | 9.92 | 9.26 | 9.26 | 8.92 | 9.62 |
| Nouvelle-Aquitaine | Other genotypes | Organised | 2022 | | 1,733 | 7.44 | 6.93 | 6.92 | 5.71 | 8.31 | 6.92 | 6.91 | 6.35 | 7.54 |

**Table S8 Observed HR HPV cervical infection prevalence, posterior predictive HR HPV cervical infection prevalence, and posterior expected HR HPV cervical infection prevalence, stratified by various dimensions.** ***(continued)***

| | | | | | | | Posterior distribution | | | | | | | |
|---|---|---|---|---|---|---|---|---|---|---|---|---|---|---|
| | | | | | | | Predictive prevalence | | | | Expected prevalence | | | |
| | | | | | | | | | ETI95% | | | | ETI95% | |
| Region | Genotypes | Type of screening | Year | Age | Number of tests | Observed | Average | Median | LB | UB | Average | Median | LB | UB |
| Occitanie | Other genotypes | Opportunistic | 2022 | | 2,624 | 9.83 | 9.67 | 9.64 | 8.38 | 10.94 | 9.67 | 9.66 | 9.12 | 10.24 |
| Occitanie | Other genotypes | Organised | 2022 | | 507 | 8.09 | 8.00 | 7.89 | 5.52 | 10.45 | 7.99 | 7.97 | 7.17 | 8.91 |
| Pays de la Loire | Other genotypes | Opportunistic | 2022 | | 721 | 9.71 | 8.18 | 8.18 | 6.24 | 10.26 | 8.18 | 8.18 | 7.56 | 8.87 |
| Pays de la Loire | Other genotypes | Organised | 2022 | | 241 | 4.56 | 5.87 | 5.81 | 2.90 | 9.13 | 5.85 | 5.83 | 5.15 | 6.60 |
| Provence-Alpes-Côte d'Azur | Other genotypes | Opportunistic | 2022 | | 7,734 | 9.53 | 9.89 | 9.88 | 9.13 | 10.62 | 9.89 | 9.89 | 9.51 | 10.30 |
| Provence-Alpes-Côte d'Azur | Other genotypes | Organised | 2022 | | 504 | 8.73 | 9.45 | 9.33 | 6.75 | 12.50 | 9.51 | 9.45 | 8.36 | 10.90 |
| Île-de-France | Other genotypes | Opportunistic | 2022 | | 15,516 | 9.36 | 9.95 | 9.95 | 9.40 | 10.49 | 9.95 | 9.95 | 9.67 | 10.24 |
| Île-de-France | Other genotypes | Organised | 2022 | | 1,307 | 8.11 | 7.49 | 7.50 | 5.97 | 9.10 | 7.50 | 7.50 | 6.87 | 8.20 |
| Auvergne-Rhône-Alpes | Other genotypes | Opportunistic | 2023 | | 192 | 14.58 | 13.58 | 13.54 | 8.33 | 18.75 | 13.52 | 13.51 | 11.81 | 15.33 |
| Auvergne-Rhône-Alpes | Other genotypes | Organised | 2023 | | 49 | 14.29 | 9.71 | 10.20 | 2.04 | 18.37 | 9.84 | 9.81 | 8.15 | 11.79 |
| Bourgogne-Franche-Comté | Other genotypes | Opportunistic | 2023 | | 551 | 12.52 | 10.90 | 10.89 | 8.34 | 13.79 | 10.88 | 10.87 | 9.86 | 11.95 |
| Bourgogne-Franche-Comté | Other genotypes | Organised | 2023 | | 190 | 6.84 | 6.66 | 6.84 | 3.16 | 10.53 | 6.68 | 6.66 | 5.60 | 7.91 |
| Bretagne | Other genotypes | Opportunistic | 2023 | | 3,450 | 11.77 | 10.93 | 10.93 | 9.71 | 12.23 | 10.94 | 10.93 | 10.25 | 11.65 |
| Bretagne | Other genotypes | Organised | 2023 | | 1,597 | 5.39 | 6.11 | 6.07 | 4.76 | 7.58 | 6.10 | 6.08 | 5.30 | 7.02 |
| Centre-Val de Loire | Other genotypes | Opportunistic | 2023 | | 5,511 | 10.03 | 10.08 | 10.07 | 9.22 | 10.98 | 10.09 | 10.09 | 9.69 | 10.52 |
| Centre-Val de Loire | Other genotypes | Organised | 2023 | | 562 | 6.41 | 6.67 | 6.58 | 4.63 | 8.90 | 6.71 | 6.70 | 6.06 | 7.41 |
| Corse | Other genotypes | Opportunistic | 2023 | | 5 | 20.00 | 8.98 | 0.00 | 0.00 | 40.00 | 9.04 | 8.94 | 6.97 | 11.58 |
| Grand Est | Other genotypes | Opportunistic | 2023 | | 2,957 | 10.62 | 10.28 | 10.28 | 9.03 | 11.53 | 10.27 | 10.27 | 9.67 | 10.90 |
| Grand Est | Other genotypes | Organised | 2023 | | 420 | 4.52 | 6.49 | 6.43 | 4.28 | 9.05 | 6.53 | 6.51 | 5.68 | 7.44 |
| Hauts-de-France | Other genotypes | Opportunistic | 2023 | | 7,934 | 10.55 | 10.29 | 10.28 | 9.53 | 11.10 | 10.29 | 10.30 | 9.89 | 10.70 |
| Hauts-de-France | Other genotypes | Organised | 2023 | | 473 | 5.92 | 6.68 | 6.55 | 4.44 | 9.30 | 6.73 | 6.72 | 5.99 | 7.54 |
| Normandie | Other genotypes | Opportunistic | 2023 | | 3,472 | 10.11 | 10.85 | 10.83 | 9.71 | 12.04 | 10.86 | 10.86 | 10.36 | 11.37 |
| Normandie | Other genotypes | Organised | 2023 | | 187 | 6.42 | 6.77 | 6.42 | 3.21 | 10.70 | 6.77 | 6.77 | 6.02 | 7.58 |
| Nouvelle-Aquitaine | Other genotypes | Opportunistic | 2023 | | 7,539 | 11.42 | 11.00 | 10.98 | 10.20 | 11.83 | 10.99 | 10.98 | 10.58 | 11.43 |
| Nouvelle-Aquitaine | Other genotypes | Organised | 2023 | | 1,611 | 7.51 | 7.53 | 7.51 | 6.08 | 9.00 | 7.55 | 7.54 | 6.88 | 8.28 |
| Occitanie | Other genotypes | Opportunistic | 2023 | | 1,178 | 11.54 | 11.43 | 11.38 | 9.51 | 13.33 | 11.42 | 11.40 | 10.70 | 12.22 |
| Occitanie | Other genotypes | Organised | 2023 | | 197 | 8.12 | 7.87 | 7.61 | 4.57 | 12.18 | 7.92 | 7.91 | 6.98 | 8.96 |
| Pays de la Loire | Other genotypes | Opportunistic | 2023 | | 1,003 | 10.17 | 9.33 | 9.27 | 7.48 | 11.37 | 9.35 | 9.33 | 8.62 | 10.17 |
| Pays de la Loire | Other genotypes | Organised | 2023 | | 498 | 5.42 | 6.15 | 6.02 | 4.02 | 8.43 | 6.16 | 6.14 | 5.43 | 6.98 |
| Provence-Alpes-Côte d'Azur | Other genotypes | Opportunistic | 2023 | | 5,645 | 11.73 | 11.63 | 11.64 | 10.68 | 12.60 | 11.64 | 11.64 | 11.16 | 12.12 |
| Provence-Alpes-Côte d'Azur | Other genotypes | Organised | 2023 | | 562 | 10.14 | 9.08 | 9.07 | 6.41 | 11.92 | 9.13 | 9.10 | 7.97 | 10.52 |
| Île-de-France | Other genotypes | Opportunistic | 2023 | | 13,457 | 11.11 | 12.05 | 12.05 | 11.41 | 12.72 | 12.05 | 12.04 | 11.70 | 12.41 |
| Île-de-France | Other genotypes | Organised | 2023 | | 1,195 | 8.79 | 8.11 | 8.12 | 6.44 | 9.87 | 8.12 | 8.11 | 7.36 | 9.01 |
| Auvergne-Rhône-Alpes | HPV16/18 | Opportunistic | | [30,39] | | | 4.84 | 4.87 | 3.80 | 5.93 | 4.83 | 4.83 | 4.42 | 5.26 |
| Auvergne-Rhône-Alpes | HPV16/18 | Organised | | [30,39] | | | 4.42 | 4.55 | 0.00 | 10.61 | 4.40 | 4.37 | 3.67 | 5.25 |
| Bourgogne-Franche-Comté | HPV16/18 | Opportunistic | | [30,39] | | | 4.47 | 4.47 | 3.40 | 5.61 | 4.46 | 4.45 | 4.03 | 4.93 |
| Bourgogne-Franche-Comté | HPV16/18 | Organised | | [30,39] | | | 3.44 | 2.90 | 0.00 | 8.70 | 3.45 | 3.43 | 2.82 | 4.17 |
| Bretagne | HPV16/18 | Opportunistic | | [30,39] | | | 5.10 | 5.09 | 4.28 | 5.95 | 5.11 | 5.10 | 4.69 | 5.55 |
| Bretagne | HPV16/18 | Organised | | [30,39] | | | 3.20 | 3.17 | 1.81 | 4.68 | 3.20 | 3.17 | 2.61 | 3.89 |

**Table S8** **Observed HR HPV cervical infection prevalence, posterior predictive HR HPV cervical infection prevalence, and posterior expected HR HPV cervical infection prevalence, stratified by various dimensions.** ***(continued)***

| | | | | | | | Posterior distribution | | | | | | | |
|---|---|---|---|---|---|---|---|---|---|---|---|---|---|---|
| | | | | | | | Predictive prevalence | | | | Expected prevalence | | | |
| | | | | | | | | | ETI95% | | | | ETI95% | |
| Region | Genotypes | Type of screening | Year | Age | Number of tests | Observed | Average | Median | LB | UB | Average | Median | LB | UB |
| Centre-Val de Loire | HPV16/18 | Opportunistic | | [30,39] | | | 4.54 | 4.54 | 4.06 | 5.02 | 4.54 | 4.54 | 4.32 | 4.77 |
| Centre-Val de Loire | HPV16/18 | Organised | | [30,39] | | | 3.52 | 3.39 | 1.96 | 5.18 | 3.52 | 3.51 | 3.03 | 4.05 |
| Corse | HPV16/18 | Opportunistic | | [30,39] | | | 4.32 | 4.55 | 0.91 | 9.09 | 4.32 | 4.30 | 3.48 | 5.38 |
| Grand Est | HPV16/18 | Opportunistic | | [30,39] | | | 4.53 | 4.54 | 3.78 | 5.30 | 4.53 | 4.52 | 4.21 | 4.88 |
| Grand Est | HPV16/18 | Organised | | [30,39] | | | 3.29 | 3.35 | 1.52 | 5.49 | 3.29 | 3.28 | 2.79 | 3.86 |
| Hauts-de-France | HPV16/18 | Opportunistic | | [30,39] | | | 4.45 | 4.45 | 4.04 | 4.88 | 4.45 | 4.45 | 4.22 | 4.67 |
| Hauts-de-France | HPV16/18 | Organised | | [30,39] | | | 3.42 | 3.48 | 2.05 | 4.90 | 3.44 | 3.43 | 2.94 | 3.96 |
| Normandie | HPV16/18 | Opportunistic | | [30,39] | | | 5.09 | 5.08 | 4.52 | 5.70 | 5.09 | 5.09 | 4.79 | 5.39 |
| Normandie | HPV16/18 | Organised | | [30,39] | | | 3.61 | 2.99 | 0.00 | 8.96 | 3.67 | 3.67 | 3.13 | 4.25 |
| Nouvelle-Aquitaine | HPV16/18 | Opportunistic | | [30,39] | | | 4.80 | 4.81 | 4.31 | 5.31 | 4.80 | 4.80 | 4.54 | 5.05 |
| Nouvelle-Aquitaine | HPV16/18 | Organised | | [30,39] | | | 3.68 | 3.60 | 2.26 | 5.19 | 3.71 | 3.70 | 3.19 | 4.29 |
| Occitanie | HPV16/18 | Opportunistic | | [30,39] | | | 5.09 | 5.08 | 4.30 | 5.89 | 5.09 | 5.09 | 4.76 | 5.42 |
| Occitanie | HPV16/18 | Organised | | [30,39] | | | 4.19 | 3.95 | 1.75 | 7.02 | 4.19 | 4.18 | 3.57 | 4.87 |
| Pays de la Loire | HPV16/18 | Opportunistic | | [30,39] | | | 3.92 | 3.90 | 3.04 | 4.86 | 3.91 | 3.91 | 3.58 | 4.25 |
| Pays de la Loire | HPV16/18 | Organised | | [30,39] | | | 3.10 | 3.05 | 1.15 | 5.34 | 3.11 | 3.10 | 2.62 | 3.65 |
| Provence-Alpes-Côte d'Azur | HPV16/18 | Opportunistic | | [30,39] | | | 5.05 | 5.04 | 4.55 | 5.54 | 5.05 | 5.04 | 4.78 | 5.33 |
| Provence-Alpes-Côte d'Azur | HPV16/18 | Organised | | [30,39] | | | 4.47 | 4.51 | 2.58 | 6.65 | 4.45 | 4.44 | 3.72 | 5.33 |
| Île-de-France | HPV16/18 | Opportunistic | | [30,39] | | | 4.40 | 4.40 | 4.19 | 4.61 | 4.40 | 4.40 | 4.27 | 4.54 |
| Île-de-France | HPV16/18 | Organised | | [30,39] | | | 3.87 | 3.81 | 2.69 | 5.20 | 3.86 | 3.85 | 3.32 | 4.46 |
| Auvergne-Rhône-Alpes | HPV16/18 | Opportunistic | | (39,49] | 1,178 | 3.57 | 3.97 | 3.99 | 2.89 | 5.18 | 3.97 | 3.97 | 3.64 | 4.33 |
| Auvergne-Rhône-Alpes | HPV16/18 | Organised | | (39,49] | 80 | 5.00 | 3.68 | 3.75 | 0.00 | 8.75 | 3.73 | 3.71 | 3.15 | 4.39 |
| Bourgogne-Franche-Comté | HPV16/18 | Opportunistic | | (39,49] | 1,030 | 3.30 | 3.69 | 3.69 | 2.52 | 4.95 | 3.67 | 3.67 | 3.31 | 4.07 |
| Bourgogne-Franche-Comté | HPV16/18 | Organised | | (39,49] | 66 | 1.52 | 2.86 | 3.03 | 0.00 | 7.58 | 2.84 | 2.83 | 2.30 | 3.48 |
| Bretagne | HPV16/18 | Opportunistic | | (39,49] | 2,375 | 4.38 | 4.17 | 4.17 | 3.37 | 5.05 | 4.17 | 4.16 | 3.84 | 4.54 |
| Bretagne | HPV16/18 | Organised | | (39,49] | 595 | 3.36 | 2.60 | 2.52 | 1.34 | 4.04 | 2.61 | 2.59 | 2.16 | 3.14 |
| Centre-Val de Loire | HPV16/18 | Opportunistic | | (39,49] | 8,774 | 3.99 | 3.70 | 3.70 | 3.28 | 4.16 | 3.71 | 3.70 | 3.52 | 3.91 |
| Centre-Val de Loire | HPV16/18 | Organised | | (39,49] | 519 | 3.08 | 2.92 | 2.89 | 1.54 | 4.43 | 2.93 | 2.92 | 2.57 | 3.30 |
| Corse | HPV16/18 | Opportunistic | | (39,49] | 127 | 4.72 | 3.62 | 3.15 | 0.79 | 7.09 | 3.56 | 3.53 | 2.76 | 4.55 |
| Corse | HPV16/18 | Organised | | (39,49] | 2 | 0.00 | 3.17 | 0.00 | 0.00 | 50.00 | 3.15 | 3.11 | 2.28 | 4.23 |
| Grand Est | HPV16/18 | Opportunistic | | (39,49] | 3,035 | 4.09 | 3.76 | 3.76 | 3.03 | 4.51 | 3.76 | 3.75 | 3.48 | 4.05 |
| Grand Est | HPV16/18 | Organised | | (39,49] | 325 | 2.77 | 2.78 | 2.77 | 1.23 | 4.92 | 2.76 | 2.75 | 2.35 | 3.20 |
| Hauts-de-France | HPV16/18 | Opportunistic | | (39,49] | 8,207 | 4.37 | 3.71 | 3.72 | 3.29 | 4.17 | 3.71 | 3.71 | 3.52 | 3.91 |
| Hauts-de-France | HPV16/18 | Organised | | (39,49] | 498 | 2.01 | 2.86 | 2.81 | 1.41 | 4.42 | 2.85 | 2.85 | 2.48 | 3.25 |
| Normandie | HPV16/18 | Opportunistic | | (39,49] | 5,887 | 4.33 | 4.18 | 4.16 | 3.60 | 4.79 | 4.18 | 4.18 | 3.92 | 4.43 |
| Normandie | HPV16/18 | Organised | | (39,49] | 69 | 1.45 | 3.16 | 2.90 | 0.00 | 7.25 | 3.13 | 3.12 | 2.70 | 3.60 |
| Nouvelle-Aquitaine | HPV16/18 | Opportunistic | | (39,49] | 7,804 | 3.86 | 3.96 | 3.96 | 3.47 | 4.45 | 3.96 | 3.95 | 3.74 | 4.18 |
| Nouvelle-Aquitaine | HPV16/18 | Organised | | (39,49] | 781 | 3.71 | 3.09 | 3.07 | 1.92 | 4.35 | 3.09 | 3.08 | 2.71 | 3.51 |
| Occitanie | HPV16/18 | Opportunistic | | (39,49] | 3,365 | 4.04 | 4.22 | 4.22 | 3.51 | 4.99 | 4.24 | 4.23 | 3.96 | 4.52 |
| Occitanie | HPV16/18 | Organised | | (39,49] | 296 | 2.36 | 3.48 | 3.38 | 1.35 | 5.74 | 3.50 | 3.49 | 3.01 | 4.01 |

**Table S8 Observed HR HPV cervical infection prevalence, posterior predictive HR HPV cervical infection prevalence, and posterior expected HR HPV cervical infection prevalence, stratified by various dimensions.** ***(continued)***

| | | | | | | | Posterior distribution | | | | | | | |
|---|---|---|---|---|---|---|---|---|---|---|---|---|---|---|
| | | | | | | | Predictive prevalence | | | | Expected prevalence | | | |
| | | | | | | | | | ETI95% | | | | ETI95% | |
| Region | Genotypes | Type of screening | Year | Age | Number of tests | Observed | Average | Median | LB | UB | Average | Median | LB | UB |
| Pays de la Loire | HPV16/18 | Opportunistic | | (39,49] | 1,705 | 2.93 | 3.26 | 3.23 | 2.40 | 4.16 | 3.26 | 3.26 | 2.99 | 3.56 |
| Pays de la Loire | HPV16/18 | Organised | | (39,49] | 288 | 3.12 | 2.58 | 2.43 | 1.04 | 4.51 | 2.59 | 2.59 | 2.23 | 3.00 |
| Provence-Alpes-Côte d'Azur | HPV16/18 | Opportunistic | | (39,49] | 7,397 | 4.24 | 4.09 | 4.10 | 3.58 | 4.58 | 4.09 | 4.08 | 3.85 | 4.32 |
| Provence-Alpes-Côte d'Azur | HPV16/18 | Organised | | (39,49] | 258 | 3.49 | 3.70 | 3.49 | 1.55 | 6.20 | 3.70 | 3.69 | 3.14 | 4.37 |
| Île-de-France | HPV16/18 | Opportunistic | | (39,49] | 49,538 | 3.34 | 3.56 | 3.56 | 3.36 | 3.77 | 3.56 | 3.56 | 3.44 | 3.69 |
| Île-de-France | HPV16/18 | Organised | | (39,49] | 790 | 2.41 | 3.16 | 3.16 | 1.90 | 4.43 | 3.15 | 3.15 | 2.76 | 3.59 |
| Auvergne-Rhône-Alpes | HPV16/18 | Opportunistic | | (49,59] | 975 | 3.18 | 3.17 | 3.18 | 2.05 | 4.41 | 3.20 | 3.19 | 2.91 | 3.50 |
| Auvergne-Rhône-Alpes | HPV16/18 | Organised | | (49,59] | 60 | 1.67 | 3.01 | 3.33 | 0.00 | 8.33 | 3.06 | 3.05 | 2.57 | 3.62 |
| Bourgogne-Franche-Comté | HPV16/18 | Opportunistic | | (49,59] | 770 | 2.60 | 2.99 | 2.99 | 1.82 | 4.29 | 2.98 | 2.98 | 2.70 | 3.30 |
| Bourgogne-Franche-Comté | HPV16/18 | Organised | | (49,59] | 56 | 0.00 | 2.41 | 1.79 | 0.00 | 7.14 | 2.35 | 2.34 | 1.86 | 2.93 |
| Bretagne | HPV16/18 | Opportunistic | | (49,59] | 1,948 | 3.08 | 3.30 | 3.29 | 2.52 | 4.16 | 3.30 | 3.29 | 3.01 | 3.60 |
| Bretagne | HPV16/18 | Organised | | (49,59] | 569 | 2.11 | 2.16 | 2.11 | 1.05 | 3.51 | 2.16 | 2.15 | 1.78 | 2.61 |
| Centre-Val de Loire | HPV16/18 | Opportunistic | | (49,59] | 8,009 | 2.58 | 2.96 | 2.96 | 2.56 | 3.38 | 2.96 | 2.96 | 2.80 | 3.14 |
| Centre-Val de Loire | HPV16/18 | Organised | | (49,59] | 673 | 2.67 | 2.44 | 2.38 | 1.34 | 3.71 | 2.44 | 2.43 | 2.14 | 2.77 |
| Corse | HPV16/18 | Opportunistic | | (49,59] | 122 | 2.46 | 2.84 | 2.46 | 0.00 | 6.56 | 2.79 | 2.77 | 2.18 | 3.54 |
| Corse | HPV16/18 | Organised | | (49,59] | 1 | 0.00 | 2.37 | 0.00 | 0.00 | 0.00 | 2.84 | 2.77 | 1.84 | 4.21 |
| Grand Est | HPV16/18 | Opportunistic | | (49,59] | 2,742 | 2.88 | 3.01 | 3.03 | 2.33 | 3.72 | 3.00 | 2.99 | 2.76 | 3.23 |
| Grand Est | HPV16/18 | Organised | | (49,59] | 308 | 2.27 | 2.30 | 2.27 | 0.65 | 4.22 | 2.33 | 2.32 | 1.98 | 2.71 |
| Hauts-de-France | HPV16/18 | Opportunistic | | (49,59] | 5,919 | 3.26 | 2.95 | 2.94 | 2.52 | 3.43 | 2.96 | 2.96 | 2.80 | 3.13 |
| Hauts-de-France | HPV16/18 | Organised | | (49,59] | 481 | 2.91 | 2.35 | 2.29 | 1.04 | 3.74 | 2.36 | 2.35 | 2.06 | 2.70 |
| Normandie | HPV16/18 | Opportunistic | | (49,59] | 4,371 | 3.52 | 3.28 | 3.27 | 2.72 | 3.87 | 3.27 | 3.27 | 3.07 | 3.49 |
| Normandie | HPV16/18 | Organised | | (49,59] | 56 | 1.79 | 2.64 | 1.79 | 0.00 | 7.14 | 2.56 | 2.55 | 2.20 | 2.97 |
| Nouvelle-Aquitaine | HPV16/18 | Opportunistic | | (49,59] | 6,164 | 3.28 | 3.16 | 3.15 | 2.71 | 3.62 | 3.16 | 3.16 | 2.98 | 3.34 |
| Nouvelle-Aquitaine | HPV16/18 | Organised | | (49,59] | 1,270 | 2.36 | 2.55 | 2.52 | 1.73 | 3.46 | 2.56 | 2.55 | 2.25 | 2.90 |
| Occitanie | HPV16/18 | Opportunistic | | (49,59] | 2,922 | 3.70 | 3.39 | 3.39 | 2.74 | 4.11 | 3.40 | 3.39 | 3.16 | 3.64 |
| Occitanie | HPV16/18 | Organised | | (49,59] | 299 | 1.67 | 3.01 | 3.01 | 1.34 | 5.02 | 3.01 | 3.01 | 2.62 | 3.46 |
| Pays de la Loire | HPV16/18 | Opportunistic | | (49,59] | 1,413 | 2.34 | 2.65 | 2.62 | 1.84 | 3.54 | 2.65 | 2.65 | 2.42 | 2.90 |
| Pays de la Loire | HPV16/18 | Organised | | (49,59] | 275 | 2.18 | 2.23 | 2.18 | 0.73 | 4.00 | 2.23 | 2.22 | 1.91 | 2.57 |
| Provence-Alpes-Côte d'Azur | HPV16/18 | Opportunistic | | (49,59] | 6,214 | 3.07 | 3.25 | 3.25 | 2.77 | 3.77 | 3.25 | 3.25 | 3.06 | 3.46 |
| Provence-Alpes-Côte d'Azur | HPV16/18 | Organised | | (49,59] | 223 | 3.14 | 3.04 | 3.14 | 0.90 | 5.38 | 3.05 | 3.03 | 2.56 | 3.61 |
| Île-de-France | HPV16/18 | Opportunistic | | (49,59] | 37,058 | 2.79 | 2.85 | 2.84 | 2.64 | 3.07 | 2.84 | 2.84 | 2.73 | 2.96 |
| Île-de-France | HPV16/18 | Organised | | (49,59] | 926 | 2.70 | 2.55 | 2.48 | 1.51 | 3.67 | 2.56 | 2.55 | 2.24 | 2.92 |
| Auvergne-Rhône-Alpes | HPV16/18 | Opportunistic | | (59,66] | 481 | 1.46 | 2.96 | 2.91 | 1.46 | 4.57 | 2.96 | 2.95 | 2.67 | 3.26 |
| Auvergne-Rhône-Alpes | HPV16/18 | Organised | | (59,66] | 43 | 0.00 | 3.02 | 2.33 | 0.00 | 9.30 | 2.96 | 2.94 | 2.42 | 3.55 |
| Bourgogne-Franche-Comté | HPV16/18 | Opportunistic | | (59,66] | 475 | 3.79 | 2.72 | 2.74 | 1.47 | 4.21 | 2.73 | 2.73 | 2.45 | 3.04 |
| Bourgogne-Franche-Comté | HPV16/18 | Organised | | (59,66] | 71 | 4.23 | 2.32 | 1.41 | 0.00 | 7.04 | 2.37 | 2.36 | 1.94 | 2.89 |
| Bretagne | HPV16/18 | Opportunistic | | (59,66] | 981 | 3.47 | 3.03 | 3.06 | 2.04 | 4.18 | 3.03 | 3.02 | 2.73 | 3.33 |
| Bretagne | HPV16/18 | Organised | | (59,66] | 470 | 1.49 | 2.08 | 2.13 | 0.85 | 3.62 | 2.09 | 2.08 | 1.66 | 2.60 |
| Centre-Val de Loire | HPV16/18 | Opportunistic | | (59,66] | 4,019 | 2.74 | 2.73 | 2.71 | 2.21 | 3.26 | 2.73 | 2.72 | 2.54 | 2.93 |

**Table S8 Observed HR HPV cervical infection prevalence, posterior predictive HR HPV cervical infection prevalence, and posterior expected HR HPV cervical infection prevalence, stratified by various dimensions.** ***(continued)***

| | | | | | | | Posterior distribution | | | | | | | |
|---|---|---|---|---|---|---|---|---|---|---|---|---|---|---|
| | | | | | | | Predictive prevalence | | | | Expected prevalence | | | |
| | | | | | | | | | ETI95% | | | | ETI95% | |
| Region | Genotypes | Type of screening | Year | Age | Number of tests | Observed | Average | Median | LB | UB | Average | Median | LB | UB |
| Centre-Val de Loire | HPV16/18 | Organised | | (59,66] | 452 | 2.88 | 2.29 | 2.21 | 1.11 | 3.76 | 2.31 | 2.30 | 1.96 | 2.71 |
| Corse | HPV16/18 | Opportunistic | | (59,66] | 68 | 1.47 | 2.72 | 2.94 | 0.00 | 7.35 | 2.72 | 2.69 | 2.20 | 3.37 |
| Grand Est | HPV16/18 | Opportunistic | | (59,66] | 1,385 | 2.45 | 2.77 | 2.74 | 1.95 | 3.68 | 2.78 | 2.77 | 2.54 | 3.03 |
| Grand Est | HPV16/18 | Organised | | (59,66] | 217 | 1.38 | 2.28 | 2.30 | 0.46 | 4.61 | 2.29 | 2.28 | 1.87 | 2.76 |
| Hauts-de-France | HPV16/18 | Opportunistic | | (59,66] | 2,757 | 2.32 | 2.76 | 2.76 | 2.14 | 3.41 | 2.76 | 2.76 | 2.57 | 2.97 |
| Hauts-de-France | HPV16/18 | Organised | | (59,66] | 338 | 3.25 | 2.31 | 2.37 | 0.89 | 4.14 | 2.33 | 2.32 | 1.96 | 2.75 |
| Normandie | HPV16/18 | Opportunistic | | (59,66] | 2,205 | 2.99 | 3.01 | 2.99 | 2.31 | 3.72 | 3.00 | 3.00 | 2.77 | 3.25 |
| Normandie | HPV16/18 | Organised | | (59,66] | 182 | 3.30 | 2.47 | 2.20 | 0.55 | 4.95 | 2.47 | 2.45 | 2.05 | 2.95 |
| Nouvelle-Aquitaine | HPV16/18 | Opportunistic | | (59,66] | 3,387 | 3.16 | 2.91 | 2.89 | 2.33 | 3.54 | 2.92 | 2.92 | 2.72 | 3.13 |
| Nouvelle-Aquitaine | HPV16/18 | Organised | | (59,66] | 934 | 2.57 | 2.50 | 2.46 | 1.39 | 3.64 | 2.50 | 2.49 | 2.13 | 2.93 |
| Occitanie | HPV16/18 | Opportunistic | | (59,66] | 1,524 | 3.54 | 3.14 | 3.15 | 2.29 | 4.13 | 3.15 | 3.14 | 2.90 | 3.41 |
| Occitanie | HPV16/18 | Organised | | (59,66] | 190 | 1.58 | 2.85 | 2.63 | 0.53 | 5.26 | 2.87 | 2.85 | 2.41 | 3.41 |
| Pays de la Loire | HPV16/18 | Opportunistic | | (59,66] | 699 | 2.72 | 2.61 | 2.58 | 1.43 | 3.86 | 2.60 | 2.60 | 2.34 | 2.88 |
| Pays de la Loire | HPV16/18 | Organised | | (59,66] | 189 | 1.59 | 2.19 | 2.12 | 0.53 | 4.76 | 2.17 | 2.16 | 1.80 | 2.58 |
| Provence-Alpes-Côte d'Azur | HPV16/18 | Opportunistic | | (59,66] | 3,024 | 2.84 | 3.02 | 3.01 | 2.38 | 3.70 | 3.03 | 3.02 | 2.81 | 3.26 |
| Provence-Alpes-Côte d'Azur | HPV16/18 | Organised | | (59,66] | 177 | 4.52 | 2.89 | 2.82 | 0.56 | 5.65 | 2.87 | 2.86 | 2.37 | 3.49 |
| Île-de-France | HPV16/18 | Opportunistic | | (59,66] | 16,364 | 2.73 | 2.65 | 2.65 | 2.37 | 2.94 | 2.65 | 2.65 | 2.50 | 2.81 |
| Île-de-France | HPV16/18 | Organised | | (59,66] | 1,724 | 2.61 | 2.58 | 2.55 | 1.74 | 3.54 | 2.58 | 2.57 | 2.15 | 3.08 |
| Auvergne-Rhône-Alpes | Other genotypes | Opportunistic | | [30,39] | | | 12.81 | 12.81 | 11.13 | 14.60 | 12.81 | 12.80 | 11.95 | 13.71 |
| Auvergne-Rhône-Alpes | Other genotypes | Organised | | [30,39] | | | 10.41 | 10.61 | 3.03 | 18.18 | 10.40 | 10.41 | 9.00 | 11.86 |
| Bourgogne-Franche-Comté | Other genotypes | Opportunistic | | [30,39] | | | 11.79 | 11.75 | 9.88 | 13.75 | 11.81 | 11.80 | 10.93 | 12.80 |
| Bourgogne-Franche-Comté | Other genotypes | Organised | | [30,39] | | | 8.66 | 8.70 | 2.90 | 15.94 | 8.79 | 8.75 | 7.43 | 10.32 |
| Bretagne | Other genotypes | Opportunistic | | [30,39] | | | 12.41 | 12.42 | 11.10 | 13.78 | 12.39 | 12.39 | 11.65 | 13.16 |
| Bretagne | Other genotypes | Organised | | [30,39] | | | 7.68 | 7.54 | 5.58 | 10.11 | 7.67 | 7.63 | 6.59 | 8.93 |
| Centre-Val de Loire | Other genotypes | Opportunistic | | [30,39] | | | 11.44 | 11.43 | 10.71 | 12.14 | 11.43 | 11.43 | 11.05 | 11.83 |
| Centre-Val de Loire | Other genotypes | Organised | | [30,39] | | | 8.16 | 8.21 | 5.89 | 10.54 | 8.16 | 8.15 | 7.40 | 9.02 |
| Corse | Other genotypes | Opportunistic | | [30,39] | | | 13.54 | 13.64 | 7.27 | 20.00 | 13.59 | 13.53 | 11.37 | 16.20 |
| Grand Est | Other genotypes | Opportunistic | | [30,39] | | | 11.69 | 11.68 | 10.43 | 12.92 | 11.69 | 11.68 | 11.08 | 12.33 |
| Grand Est | Other genotypes | Organised | | [30,39] | | | 8.04 | 7.93 | 5.18 | 11.28 | 8.04 | 8.03 | 7.01 | 9.09 |
| Hauts-de-France | Other genotypes | Opportunistic | | [30,39] | | | 11.35 | 11.35 | 10.70 | 12.00 | 11.35 | 11.35 | 10.97 | 11.74 |
| Hauts-de-France | Other genotypes | Organised | | [30,39] | | | 8.15 | 8.06 | 5.85 | 10.74 | 8.15 | 8.14 | 7.29 | 9.05 |
| Normandie | Other genotypes | Opportunistic | | [30,39] | | | 12.12 | 12.12 | 11.25 | 13.04 | 12.11 | 12.11 | 11.62 | 12.61 |
| Normandie | Other genotypes | Organised | | [30,39] | | | 8.52 | 8.96 | 2.99 | 14.93 | 8.55 | 8.54 | 7.57 | 9.59 |
| Nouvelle-Aquitaine | Other genotypes | Opportunistic | | [30,39] | | | 12.45 | 12.44 | 11.67 | 13.26 | 12.44 | 12.43 | 12.02 | 12.87 |
| Nouvelle-Aquitaine | Other genotypes | Organised | | [30,39] | | | 9.45 | 9.45 | 7.19 | 11.72 | 9.48 | 9.46 | 8.56 | 10.44 |
| Occitanie | Other genotypes | Opportunistic | | [30,39] | | | 13.22 | 13.22 | 11.92 | 14.46 | 13.22 | 13.21 | 12.62 | 13.88 |
| Occitanie | Other genotypes | Organised | | [30,39] | | | 10.13 | 10.09 | 6.14 | 14.47 | 10.11 | 10.08 | 9.04 | 11.31 |
| Pays de la Loire | Other genotypes | Opportunistic | | [30,39] | | | 9.91 | 9.93 | 8.43 | 11.47 | 9.93 | 9.91 | 9.27 | 10.63 |
| Pays de la Loire | Other genotypes | Organised | | [30,39] | | | 7.27 | 7.25 | 4.20 | 10.69 | 7.34 | 7.33 | 6.47 | 8.26 |

**Table S8** **Observed HR HPV cervical infection prevalence, posterior predictive HR HPV cervical infection prevalence, and posterior expected HR HPV cervical infection prevalence, stratified by various dimensions.** ***(continued)***

| | | | | | | | Posterior distribution | | | | | | | |
|---|---|---|---|---|---|---|---|---|---|---|---|---|---|---|
| | | | | | | | Predictive prevalence | | | | Expected prevalence | | | |
| | | | | | | | | | ETI95% | | | | ETI95% | |
| Region | Genotypes | Type of screening | Year | Age | Number of tests | Observed | Average | Median | LB | UB | Average | Median | LB | UB |
| Provence-Alpes-Côte d'Azur | Other genotypes | Opportunistic | | [30,39] | | | 12.95 | 12.94 | 12.17 | 13.77 | 12.94 | 12.94 | 12.50 | 13.40 |
| Provence-Alpes-Côte d'Azur | Other genotypes | Organised | | [30,39] | | | 11.25 | 11.16 | 8.15 | 14.59 | 11.31 | 11.26 | 9.90 | 12.99 |
| Île-de-France | Other genotypes | Opportunistic | | [30,39] | | | 12.54 | 12.54 | 12.21 | 12.90 | 12.54 | 12.55 | 12.33 | 12.77 |
| Île-de-France | Other genotypes | Organised | | [30,39] | | | 9.98 | 9.94 | 7.99 | 12.07 | 10.01 | 9.99 | 9.03 | 11.05 |
| Auvergne-Rhône-Alpes | Other genotypes | Opportunistic | | (39,49] | 1,178 | 8.32 | 8.97 | 8.91 | 7.22 | 10.78 | 8.97 | 8.95 | 8.33 | 9.64 |
| Auvergne-Rhône-Alpes | Other genotypes | Organised | | (39,49] | 80 | 10.00 | 7.71 | 7.50 | 2.50 | 13.75 | 7.79 | 7.79 | 6.73 | 8.89 |
| Bourgogne-Franche-Comté | Other genotypes | Opportunistic | | (39,49] | 1,030 | 8.93 | 8.24 | 8.25 | 6.50 | 10.10 | 8.22 | 8.21 | 7.58 | 8.91 |
| Bourgogne-Franche-Comté | Other genotypes | Organised | | (39,49] | 66 | 4.55 | 6.58 | 6.06 | 1.52 | 13.64 | 6.59 | 6.56 | 5.49 | 7.84 |
| Bretagne | Other genotypes | Opportunistic | | (39,49] | 2,375 | 9.81 | 8.66 | 8.67 | 7.45 | 9.98 | 8.66 | 8.65 | 8.12 | 9.22 |
| Bretagne | Other genotypes | Organised | | (39,49] | 595 | 5.55 | 5.60 | 5.55 | 3.70 | 7.56 | 5.64 | 5.62 | 4.87 | 6.47 |
| Centre-Val de Loire | Other genotypes | Opportunistic | | (39,49] | 8,774 | 8.09 | 7.95 | 7.94 | 7.31 | 8.58 | 7.95 | 7.95 | 7.66 | 8.24 |
| Centre-Val de Loire | Other genotypes | Organised | | (39,49] | 519 | 5.59 | 6.20 | 6.17 | 4.05 | 8.48 | 6.21 | 6.21 | 5.64 | 6.80 |
| Corse | Other genotypes | Opportunistic | | (39,49] | 127 | 14.96 | 10.21 | 10.24 | 4.72 | 15.75 | 10.15 | 10.10 | 8.20 | 12.51 |
| Corse | Other genotypes | Organised | | (39,49] | 2 | 0.00 | 6.48 | 0.00 | 0.00 | 50.00 | 6.92 | 6.82 | 5.15 | 9.08 |
| Grand Est | Other genotypes | Opportunistic | | (39,49] | 3,035 | 9.13 | 8.31 | 8.30 | 7.25 | 9.42 | 8.32 | 8.31 | 7.87 | 8.82 |
| Grand Est | Other genotypes | Organised | | (39,49] | 325 | 7.38 | 6.13 | 6.15 | 3.69 | 8.93 | 6.18 | 6.17 | 5.42 | 7.00 |
| Hauts-de-France | Other genotypes | Opportunistic | | (39,49] | 8,207 | 8.87 | 8.03 | 8.03 | 7.35 | 8.69 | 8.03 | 8.03 | 7.73 | 8.32 |
| Hauts-de-France | Other genotypes | Organised | | (39,49] | 498 | 5.02 | 6.19 | 6.22 | 4.02 | 8.43 | 6.17 | 6.16 | 5.54 | 6.80 |
| Normandie | Other genotypes | Opportunistic | | (39,49] | 5,887 | 8.26 | 8.54 | 8.53 | 7.73 | 9.38 | 8.55 | 8.54 | 8.17 | 8.92 |
| Normandie | Other genotypes | Organised | | (39,49] | 69 | 10.14 | 6.64 | 5.80 | 1.45 | 13.04 | 6.72 | 6.72 | 5.96 | 7.52 |
| Nouvelle-Aquitaine | Other genotypes | Opportunistic | | (39,49] | 7,804 | 8.73 | 8.75 | 8.75 | 8.05 | 9.46 | 8.76 | 8.75 | 8.44 | 9.09 |
| Nouvelle-Aquitaine | Other genotypes | Organised | | (39,49] | 781 | 8.32 | 7.14 | 7.17 | 5.25 | 9.09 | 7.14 | 7.12 | 6.48 | 7.85 |
| Occitanie | Other genotypes | Opportunistic | | (39,49] | 3,365 | 9.27 | 9.52 | 9.51 | 8.47 | 10.64 | 9.51 | 9.51 | 9.04 | 10.00 |
| Occitanie | Other genotypes | Organised | | (39,49] | 296 | 6.08 | 7.49 | 7.43 | 4.73 | 10.81 | 7.55 | 7.54 | 6.73 | 8.47 |
| Pays de la Loire | Other genotypes | Opportunistic | | (39,49] | 1,705 | 7.10 | 7.15 | 7.16 | 5.87 | 8.50 | 7.15 | 7.14 | 6.66 | 7.68 |
| Pays de la Loire | Other genotypes | Organised | | (39,49] | 288 | 4.17 | 5.56 | 5.56 | 3.12 | 8.33 | 5.52 | 5.51 | 4.88 | 6.19 |
| Provence-Alpes-Côte d'Azur | Other genotypes | Opportunistic | | (39,49] | 7,397 | 8.85 | 8.98 | 8.98 | 8.27 | 9.71 | 8.98 | 8.98 | 8.64 | 9.33 |
| Provence-Alpes-Côte d'Azur | Other genotypes | Organised | | (39,49] | 258 | 8.14 | 8.30 | 8.14 | 4.65 | 12.02 | 8.34 | 8.30 | 7.29 | 9.62 |
| Île-de-France | Other genotypes | Opportunistic | | (39,49] | 49,538 | 8.31 | 8.68 | 8.68 | 8.37 | 8.99 | 8.67 | 8.67 | 8.49 | 8.86 |
| Île-de-France | Other genotypes | Organised | | (39,49] | 790 | 7.97 | 7.27 | 7.22 | 5.44 | 9.37 | 7.30 | 7.29 | 6.65 | 8.04 |
| Auvergne-Rhône-Alpes | Other genotypes | Opportunistic | | (49,59] | 975 | 7.49 | 7.67 | 7.69 | 5.95 | 9.54 | 7.66 | 7.65 | 7.11 | 8.24 |
| Auvergne-Rhône-Alpes | Other genotypes | Organised | | (49,59] | 60 | 10.00 | 6.94 | 6.67 | 1.67 | 15.00 | 7.10 | 7.09 | 6.13 | 8.16 |
| Bourgogne-Franche-Comté | Other genotypes | Opportunistic | | (49,59] | 770 | 7.40 | 7.03 | 7.01 | 5.19 | 8.96 | 7.03 | 7.02 | 6.49 | 7.60 |
| Bourgogne-Franche-Comté | Other genotypes | Organised | | (49,59] | 56 | 7.14 | 6.05 | 5.36 | 0.00 | 12.50 | 6.02 | 5.99 | 4.98 | 7.23 |
| Bretagne | Other genotypes | Opportunistic | | (49,59] | 1,948 | 7.55 | 7.25 | 7.24 | 6.06 | 8.52 | 7.25 | 7.25 | 6.77 | 7.73 |
| Bretagne | Other genotypes | Organised | | (49,59] | 569 | 4.57 | 5.21 | 5.27 | 3.34 | 7.38 | 5.20 | 5.17 | 4.48 | 6.03 |
| Centre-Val de Loire | Other genotypes | Opportunistic | | (49,59] | 8,009 | 6.21 | 6.74 | 6.74 | 6.12 | 7.34 | 6.74 | 6.74 | 6.48 | 7.00 |
| Centre-Val de Loire | Other genotypes | Organised | | (49,59] | 673 | 5.50 | 5.69 | 5.65 | 3.86 | 7.58 | 5.70 | 5.69 | 5.19 | 6.26 |
| Corse | Other genotypes | Opportunistic | | (49,59] | 122 | 5.74 | 8.48 | 8.20 | 3.28 | 13.93 | 8.42 | 8.37 | 6.82 | 10.33 |

**Table S8 Observed HR HPV cervical infection prevalence, posterior predictive HR HPV cervical infection prevalence, and posterior expected HR HPV cervical infection prevalence, stratified by various dimensions.** ***(continued)***

| | | | | | | | Posterior distribution | | | | | | | |
|---|---|---|---|---|---|---|---|---|---|---|---|---|---|---|
| | | | | | | | Predictive prevalence | | | | Expected prevalence | | | |
| | | | | | | | | | ETI95% | | | | ETI95% | |
| Region | Genotypes | Type of screening | Year | Age | Number of tests | Observed | Average | Median | LB | UB | Average | Median | LB | UB |
| Corse | Other genotypes | Organised | | (49,59] | 1 | 0.00 | 6.40 | 0.00 | 0.00 | 100.00 | 6.17 | 6.05 | 4.07 | 8.97 |
| Grand Est | Other genotypes | Opportunistic | | (49,59] | 2,742 | 6.24 | 7.03 | 7.04 | 6.02 | 8.02 | 7.03 | 7.03 | 6.64 | 7.48 |
| Grand Est | Other genotypes | Organised | | (49,59] | 308 | 3.90 | 5.76 | 5.84 | 3.25 | 8.44 | 5.74 | 5.73 | 5.05 | 6.48 |
| Hauts-de-France | Other genotypes | Opportunistic | | (49,59] | 5,919 | 7.72 | 6.80 | 6.79 | 6.10 | 7.48 | 6.81 | 6.81 | 6.55 | 7.08 |
| Hauts-de-France | Other genotypes | Organised | | (49,59] | 481 | 5.82 | 5.69 | 5.61 | 3.74 | 7.90 | 5.71 | 5.70 | 5.14 | 6.34 |
| Normandie | Other genotypes | Opportunistic | | (49,59] | 4,371 | 6.47 | 7.12 | 7.12 | 6.29 | 7.96 | 7.13 | 7.13 | 6.81 | 7.46 |
| Normandie | Other genotypes | Organised | | (49,59] | 56 | 3.57 | 6.13 | 5.36 | 0.00 | 12.50 | 6.10 | 6.10 | 5.40 | 6.85 |
| Nouvelle-Aquitaine | Other genotypes | Opportunistic | | (49,59] | 6,164 | 7.54 | 7.41 | 7.40 | 6.73 | 8.10 | 7.41 | 7.41 | 7.13 | 7.72 |
| Nouvelle-Aquitaine | Other genotypes | Organised | | (49,59] | 1,270 | 6.54 | 6.42 | 6.38 | 5.04 | 7.87 | 6.40 | 6.39 | 5.85 | 7.02 |
| Occitanie | Other genotypes | Opportunistic | | (49,59] | 2,922 | 8.35 | 8.08 | 8.08 | 7.05 | 9.17 | 8.07 | 8.07 | 7.67 | 8.50 |
| Occitanie | Other genotypes | Organised | | (49,59] | 299 | 6.69 | 7.14 | 7.02 | 4.35 | 10.37 | 7.19 | 7.17 | 6.42 | 8.03 |
| Pays de la Loire | Other genotypes | Opportunistic | | (49,59] | 1,413 | 6.09 | 6.11 | 6.09 | 4.81 | 7.43 | 6.12 | 6.11 | 5.70 | 6.59 |
| Pays de la Loire | Other genotypes | Organised | | (49,59] | 275 | 6.18 | 5.25 | 5.09 | 2.55 | 8.00 | 5.25 | 5.24 | 4.66 | 5.90 |
| Provence-Alpes-Côte d'Azur | Other genotypes | Opportunistic | | (49,59] | 6,214 | 8.18 | 7.57 | 7.56 | 6.86 | 8.29 | 7.58 | 7.58 | 7.27 | 7.90 |
| Provence-Alpes-Côte d'Azur | Other genotypes | Organised | | (49,59] | 223 | 7.62 | 7.61 | 7.62 | 4.04 | 11.21 | 7.63 | 7.59 | 6.61 | 8.87 |
| Île-de-France | Other genotypes | Opportunistic | | (49,59] | 37,058 | 7.23 | 7.34 | 7.34 | 7.03 | 7.67 | 7.34 | 7.34 | 7.16 | 7.54 |
| Île-de-France | Other genotypes | Organised | | (49,59] | 926 | 6.59 | 6.52 | 6.48 | 4.86 | 8.21 | 6.51 | 6.49 | 5.91 | 7.15 |
| Auvergne-Rhône-Alpes | Other genotypes | Opportunistic | | (59,66] | 481 | 6.44 | 6.78 | 6.65 | 4.57 | 9.15 | 6.81 | 6.79 | 6.28 | 7.36 |
| Auvergne-Rhône-Alpes | Other genotypes | Organised | | (59,66] | 43 | 2.33 | 6.72 | 6.98 | 0.00 | 13.95 | 6.76 | 6.75 | 5.77 | 7.84 |
| Bourgogne-Franche-Comté | Other genotypes | Opportunistic | | (59,66] | 475 | 8.00 | 6.27 | 6.32 | 4.00 | 8.63 | 6.28 | 6.28 | 5.76 | 6.85 |
| Bourgogne-Franche-Comté | Other genotypes | Organised | | (59,66] | 71 | 8.45 | 5.64 | 5.63 | 1.41 | 11.27 | 5.72 | 5.71 | 4.83 | 6.68 |
| Bretagne | Other genotypes | Opportunistic | | (59,66] | 981 | 6.83 | 6.39 | 6.32 | 4.79 | 8.05 | 6.40 | 6.40 | 5.93 | 6.87 |
| Bretagne | Other genotypes | Organised | | (59,66] | 470 | 3.83 | 4.99 | 4.89 | 2.98 | 7.23 | 4.99 | 4.98 | 4.21 | 5.88 |
| Centre-Val de Loire | Other genotypes | Opportunistic | | (59,66] | 4,019 | 6.10 | 5.95 | 5.95 | 5.18 | 6.77 | 5.95 | 5.95 | 5.65 | 6.25 |
| Centre-Val de Loire | Other genotypes | Organised | | (59,66] | 452 | 5.97 | 5.43 | 5.31 | 3.32 | 7.74 | 5.41 | 5.39 | 4.82 | 6.10 |
| Corse | Other genotypes | Opportunistic | | (59,66] | 68 | 2.94 | 7.56 | 7.35 | 1.47 | 14.71 | 7.59 | 7.55 | 6.32 | 9.10 |
| Grand Est | Other genotypes | Opportunistic | | (59,66] | 1,385 | 4.69 | 6.30 | 6.28 | 4.98 | 7.65 | 6.28 | 6.27 | 5.88 | 6.69 |
| Grand Est | Other genotypes | Organised | | (59,66] | 217 | 4.15 | 5.60 | 5.53 | 2.76 | 8.76 | 5.62 | 5.61 | 4.83 | 6.45 |
| Hauts-de-France | Other genotypes | Opportunistic | | (59,66] | 2,757 | 6.13 | 6.13 | 6.09 | 5.22 | 7.07 | 6.12 | 6.12 | 5.82 | 6.43 |
| Hauts-de-France | Other genotypes | Organised | | (59,66] | 338 | 6.80 | 5.63 | 5.62 | 3.25 | 8.28 | 5.64 | 5.63 | 4.95 | 6.38 |
| Normandie | Other genotypes | Opportunistic | | (59,66] | 2,205 | 5.80 | 6.28 | 6.26 | 5.26 | 7.35 | 6.28 | 6.28 | 5.95 | 6.63 |
| Normandie | Other genotypes | Organised | | (59,66] | 182 | 4.40 | 5.83 | 5.49 | 2.75 | 9.34 | 5.77 | 5.75 | 5.01 | 6.62 |
| Nouvelle-Aquitaine | Other genotypes | Opportunistic | | (59,66] | 3,387 | 6.17 | 6.61 | 6.61 | 5.73 | 7.56 | 6.61 | 6.61 | 6.29 | 6.95 |
| Nouvelle-Aquitaine | Other genotypes | Organised | | (59,66] | 934 | 6.42 | 6.12 | 6.10 | 4.50 | 7.82 | 6.15 | 6.14 | 5.48 | 6.90 |
| Occitanie | Other genotypes | Opportunistic | | (59,66] | 1,524 | 7.15 | 7.19 | 7.15 | 5.84 | 8.53 | 7.21 | 7.21 | 6.79 | 7.65 |
| Occitanie | Other genotypes | Organised | | (59,66] | 190 | 8.42 | 6.93 | 6.84 | 3.68 | 10.54 | 6.92 | 6.90 | 6.08 | 7.89 |
| Pays de la Loire | Other genotypes | Opportunistic | | (59,66] | 699 | 5.29 | 5.71 | 5.72 | 4.01 | 7.58 | 5.73 | 5.72 | 5.27 | 6.25 |
| Pays de la Loire | Other genotypes | Organised | | (59,66] | 189 | 2.65 | 5.12 | 5.29 | 2.12 | 8.47 | 5.10 | 5.09 | 4.42 | 5.87 |
| Provence-Alpes-Côte d'Azur | Other genotypes | Opportunistic | | (59,66] | 3,024 | 7.21 | 6.76 | 6.75 | 5.82 | 7.77 | 6.76 | 6.76 | 6.42 | 7.10 |

**Table S8 Observed HR HPV cervical infection prevalence, posterior predictive HR HPV cervical infection prevalence, and posterior expected HR HPV cervical infection prevalence, stratified by various dimensions.** ***(continued)***

| | | | | | | | Posterior distribution | | | | | | | |
|---|---|---|---|---|---|---|---|---|---|---|---|---|---|---|
| | | | | | | | Predictive prevalence | | | | Expected prevalence | | | |
| | | | | | | | | | ETI95% | | | | ETI95% | |
| Region | Genotypes | Type of screening | Year | Age | Number of tests | Observed | Average | Median | LB | UB | Average | Median | LB | UB |
| Provence-Alpes-Côte d'Azur | Other genotypes | Organised | | (59,66] | 177 | 6.78 | 7.02 | 6.78 | 3.39 | 11.30 | 7.07 | 7.04 | 6.09 | 8.26 |
| Île-de-France | Other genotypes | Opportunistic | | (59,66] | 16,364 | 6.63 | 6.54 | 6.54 | 6.09 | 6.98 | 6.53 | 6.53 | 6.29 | 6.79 |
| Île-de-France | Other genotypes | Organised | | (59,66] | 1,724 | 6.09 | 5.88 | 5.86 | 4.58 | 7.31 | 5.88 | 5.86 | 5.19 | 6.63 |
| Auvergne-Rhône-Alpes | HPV16/18 | Opportunistic | 2020 | [30,39] | | | 4.14 | 4.10 | 2.41 | 6.27 | 4.14 | 4.13 | 3.73 | 4.58 |
| Bourgogne-Franche-Comté | HPV16/18 | Opportunistic | 2020 | [30,39] | | | 3.69 | 3.86 | 1.45 | 6.76 | 3.66 | 3.66 | 3.26 | 4.11 |
| Bretagne | HPV16/18 | Opportunistic | 2020 | [30,39] | | | 3.89 | 3.93 | 1.75 | 6.55 | 3.88 | 3.87 | 3.46 | 4.33 |
| Centre-Val de Loire | HPV16/18 | Opportunistic | 2020 | [30,39] | | | 3.79 | 3.77 | 2.68 | 4.94 | 3.79 | 3.79 | 3.55 | 4.05 |
| Corse | HPV16/18 | Opportunistic | 2020 | [30,39] | | | 4.22 | 4.35 | 1.09 | 8.70 | 4.22 | 4.20 | 3.24 | 5.42 |
| Grand Est | HPV16/18 | Opportunistic | 2020 | [30,39] | | | 3.62 | 3.40 | 1.28 | 6.38 | 3.58 | 3.57 | 3.21 | 3.96 |
| Hauts-de-France | HPV16/18 | Opportunistic | 2020 | [30,39] | | | 3.58 | 3.57 | 2.55 | 4.75 | 3.59 | 3.58 | 3.34 | 3.83 |
| Hauts-de-France | HPV16/18 | Organised | 2020 | [30,39] | | | 3.37 | 0.00 | 0.00 | 100.00 | 3.78 | 3.70 | 2.44 | 5.48 |
| Normandie | HPV16/18 | Opportunistic | 2020 | [30,39] | | | 4.05 | 4.09 | 2.81 | 5.50 | 4.06 | 4.06 | 3.75 | 4.37 |
| Nouvelle-Aquitaine | HPV16/18 | Opportunistic | 2020 | [30,39] | | | 3.82 | 3.80 | 2.38 | 5.39 | 3.84 | 3.84 | 3.57 | 4.10 |
| Occitanie | HPV16/18 | Opportunistic | 2020 | [30,39] | | | 4.76 | 4.68 | 3.16 | 6.46 | 4.77 | 4.76 | 4.31 | 5.26 |
| Pays de la Loire | HPV16/18 | Opportunistic | 2020 | [30,39] | | | 3.20 | 3.00 | 1.36 | 5.18 | 3.23 | 3.23 | 2.89 | 3.61 |
| Pays de la Loire | HPV16/18 | Organised | 2020 | [30,39] | | | 2.99 | 0.00 | 0.00 | 33.33 | 3.11 | 3.06 | 2.15 | 4.31 |
| Provence-Alpes-Côte d'Azur | HPV16/18 | Opportunistic | 2020 | [30,39] | | | 3.96 | 3.91 | 2.65 | 5.42 | 3.96 | 3.96 | 3.67 | 4.28 |
| Île-de-France | HPV16/18 | Opportunistic | 2020 | [30,39] | | | 3.84 | 3.84 | 3.55 | 4.17 | 3.85 | 3.85 | 3.68 | 4.02 |
| Île-de-France | HPV16/18 | Organised | 2020 | [30,39] | | | 3.70 | 0.00 | 0.00 | 25.00 | 3.96 | 3.91 | 2.90 | 5.25 |
| Auvergne-Rhône-Alpes | HPV16/18 | Opportunistic | 2020 | (39,49] | 262 | 0.76 | 3.37 | 3.44 | 1.52 | 5.73 | 3.38 | 3.37 | 3.04 | 3.74 |
| Bourgogne-Franche-Comté | HPV16/18 | Opportunistic | 2020 | (39,49] | 144 | 2.08 | 3.02 | 2.78 | 0.69 | 6.25 | 2.99 | 2.99 | 2.64 | 3.36 |
| Bretagne | HPV16/18 | Opportunistic | 2020 | (39,49] | 158 | 5.06 | 3.20 | 3.16 | 0.63 | 6.33 | 3.20 | 3.19 | 2.82 | 3.62 |
| Centre-Val de Loire | HPV16/18 | Opportunistic | 2020 | (39,49] | 1,001 | 2.50 | 3.12 | 3.10 | 2.10 | 4.30 | 3.13 | 3.14 | 2.92 | 3.36 |
| Corse | HPV16/18 | Opportunistic | 2020 | (39,49] | 118 | 3.39 | 3.56 | 3.39 | 0.85 | 7.63 | 3.50 | 3.47 | 2.66 | 4.54 |
| Grand Est | HPV16/18 | Opportunistic | 2020 | (39,49] | 241 | 2.90 | 2.90 | 2.90 | 0.83 | 5.39 | 2.89 | 2.88 | 2.57 | 3.23 |
| Hauts-de-France | HPV16/18 | Opportunistic | 2020 | (39,49] | 757 | 3.83 | 2.97 | 2.91 | 1.85 | 4.23 | 2.99 | 2.99 | 2.78 | 3.19 |
| Hauts-de-France | HPV16/18 | Organised | 2020 | (39,49] | 2 | 0.00 | 2.83 | 0.00 | 0.00 | 50.00 | 2.89 | 2.85 | 2.01 | 3.99 |
| Normandie | HPV16/18 | Opportunistic | 2020 | (39,49] | 675 | 3.56 | 3.26 | 3.26 | 1.93 | 4.74 | 3.25 | 3.24 | 2.99 | 3.50 |
| Nouvelle-Aquitaine | HPV16/18 | Opportunistic | 2020 | (39,49] | 424 | 2.83 | 3.20 | 3.07 | 1.65 | 4.95 | 3.20 | 3.20 | 2.96 | 3.43 |
| Occitanie | HPV16/18 | Opportunistic | 2020 | (39,49] | 628 | 3.66 | 4.05 | 3.98 | 2.55 | 5.73 | 4.05 | 4.05 | 3.64 | 4.49 |
| Pays de la Loire | HPV16/18 | Opportunistic | 2020 | (39,49] | 369 | 1.90 | 2.67 | 2.71 | 1.08 | 4.61 | 2.67 | 2.66 | 2.38 | 2.98 |
| Pays de la Loire | HPV16/18 | Organised | 2020 | (39,49] | 1 | 0.00 | 1.70 | 0.00 | 0.00 | 0.00 | 2.49 | 2.43 | 1.62 | 3.67 |
| Provence-Alpes-Côte d'Azur | HPV16/18 | Opportunistic | 2020 | (39,49] | 620 | 3.71 | 3.19 | 3.23 | 1.94 | 4.68 | 3.18 | 3.18 | 2.92 | 3.45 |
| Île-de-France | HPV16/18 | Opportunistic | 2020 | (39,49] | 16,219 | 2.78 | 3.15 | 3.15 | 2.85 | 3.48 | 3.15 | 3.16 | 3.01 | 3.30 |
| Île-de-France | HPV16/18 | Organised | 2020 | (39,49] | 1 | 0.00 | 3.57 | 0.00 | 0.00 | 100.00 | 3.11 | 3.07 | 2.21 | 4.24 |
| Auvergne-Rhône-Alpes | HPV16/18 | Opportunistic | 2020 | (49,59] | 215 | 4.65 | 2.68 | 2.79 | 0.93 | 5.12 | 2.74 | 2.73 | 2.45 | 3.04 |
| Bourgogne-Franche-Comté | HPV16/18 | Opportunistic | 2020 | (49,59] | 98 | 1.02 | 2.55 | 2.04 | 0.00 | 6.12 | 2.48 | 2.48 | 2.22 | 2.78 |
| Bretagne | HPV16/18 | Opportunistic | 2020 | (49,59] | 107 | 1.87 | 2.51 | 2.80 | 0.00 | 5.61 | 2.51 | 2.50 | 2.20 | 2.86 |
| Centre-Val de Loire | HPV16/18 | Opportunistic | 2020 | (49,59] | 971 | 1.65 | 2.52 | 2.47 | 1.54 | 3.60 | 2.52 | 2.52 | 2.35 | 2.70 |

**Table S8** **Observed HR HPV cervical infection prevalence, posterior predictive HR HPV cervical infection prevalence, and posterior expected HR HPV cervical infection prevalence, stratified by various dimensions.** ***(continued)***

| | | | | | | | Posterior distribution | | | | | | | |
|---|---|---|---|---|---|---|---|---|---|---|---|---|---|---|
| | | | | | | | Predictive prevalence | | | | Expected prevalence | | | |
| | | | | | | | | | ETI95% | | | | ETI95% | |
| Region | Genotypes | Type of screening | Year | Age | Number of tests | Observed | Average | Median | LB | UB | Average | Median | LB | UB |
| Centre-Val de Loire | HPV16/18 | Organised | 2020 | (49,59] | 2 | 0.00 | 2.83 | 0.00 | 0.00 | 50.00 | 2.76 | 2.72 | 1.96 | 3.76 |
| Corse | HPV16/18 | Opportunistic | 2020 | (49,59] | 111 | 2.70 | 2.81 | 2.70 | 0.00 | 6.31 | 2.76 | 2.74 | 2.12 | 3.56 |
| Grand Est | HPV16/18 | Opportunistic | 2020 | (49,59] | 279 | 3.23 | 2.25 | 2.15 | 0.72 | 4.30 | 2.29 | 2.29 | 2.02 | 2.58 |
| Hauts-de-France | HPV16/18 | Opportunistic | 2020 | (49,59] | 626 | 2.72 | 2.34 | 2.24 | 1.28 | 3.67 | 2.35 | 2.35 | 2.18 | 2.52 |
| Normandie | HPV16/18 | Opportunistic | 2020 | (49,59] | 521 | 3.07 | 2.58 | 2.50 | 1.34 | 4.03 | 2.58 | 2.58 | 2.37 | 2.80 |
| Nouvelle-Aquitaine | HPV16/18 | Opportunistic | 2020 | (49,59] | 367 | 3.00 | 2.52 | 2.45 | 1.09 | 4.09 | 2.53 | 2.53 | 2.34 | 2.73 |
| Occitanie | HPV16/18 | Opportunistic | 2020 | (49,59] | 533 | 3.00 | 3.11 | 3.00 | 1.69 | 4.69 | 3.13 | 3.12 | 2.83 | 3.44 |
| Occitanie | HPV16/18 | Organised | 2020 | (49,59] | 3 | 0.00 | 3.39 | 0.00 | 0.00 | 33.33 | 3.24 | 3.17 | 2.20 | 4.63 |
| Pays de la Loire | HPV16/18 | Opportunistic | 2020 | (49,59] | 278 | 0.72 | 2.17 | 2.16 | 0.72 | 3.96 | 2.19 | 2.19 | 1.96 | 2.45 |
| Pays de la Loire | HPV16/18 | Organised | 2020 | (49,59] | 1 | 0.00 | 2.20 | 0.00 | 0.00 | 0.00 | 2.15 | 2.10 | 1.43 | 3.15 |
| Provence-Alpes-Côte d'Azur | HPV16/18 | Opportunistic | 2020 | (49,59] | 509 | 1.96 | 2.51 | 2.55 | 1.18 | 3.93 | 2.52 | 2.52 | 2.32 | 2.74 |
| Île-de-France | HPV16/18 | Opportunistic | 2020 | (49,59] | 11,874 | 2.32 | 2.52 | 2.52 | 2.21 | 2.84 | 2.52 | 2.51 | 2.40 | 2.64 |
| Île-de-France | HPV16/18 | Organised | 2020 | (49,59] | 3 | 0.00 | 3.63 | 0.00 | 0.00 | 33.33 | 3.40 | 3.36 | 2.58 | 4.40 |
| Auvergne-Rhône-Alpes | HPV16/18 | Opportunistic | 2020 | (59,66] | 96 | 3.12 | 2.46 | 2.08 | 0.00 | 6.25 | 2.52 | 2.52 | 2.24 | 2.84 |
| Bourgogne-Franche-Comté | HPV16/18 | Opportunistic | 2020 | (59,66] | 88 | 4.55 | 2.34 | 2.27 | 0.00 | 5.68 | 2.31 | 2.30 | 2.04 | 2.60 |
| Bretagne | HPV16/18 | Opportunistic | 2020 | (59,66] | 65 | 1.54 | 2.24 | 1.54 | 0.00 | 6.15 | 2.24 | 2.23 | 1.92 | 2.59 |
| Centre-Val de Loire | HPV16/18 | Opportunistic | 2020 | (59,66] | 500 | 1.40 | 2.33 | 2.20 | 1.20 | 3.80 | 2.33 | 2.33 | 2.15 | 2.52 |
| Centre-Val de Loire | HPV16/18 | Organised | 2020 | (59,66] | 1 | 100.00 | 3.73 | 0.00 | 0.00 | 100.00 | 3.41 | 3.34 | 2.19 | 5.11 |
| Corse | HPV16/18 | Opportunistic | 2020 | (59,66] | 56 | 1.79 | 2.63 | 1.79 | 0.00 | 7.14 | 2.60 | 2.57 | 2.03 | 3.31 |
| Grand Est | HPV16/18 | Opportunistic | 2020 | (59,66] | 127 | 3.15 | 2.11 | 1.97 | 0.00 | 4.72 | 2.10 | 2.10 | 1.84 | 2.40 |
| Hauts-de-France | HPV16/18 | Opportunistic | 2020 | (59,66] | 272 | 2.21 | 2.14 | 2.21 | 0.74 | 4.04 | 2.16 | 2.16 | 1.98 | 2.35 |
| Hauts-de-France | HPV16/18 | Organised | 2020 | (59,66] | 4 | 25.00 | 2.61 | 0.00 | 0.00 | 25.00 | 2.58 | 2.54 | 1.89 | 3.46 |
| Normandie | HPV16/18 | Opportunistic | 2020 | (59,66] | 262 | 3.82 | 2.34 | 2.29 | 0.76 | 4.20 | 2.35 | 2.34 | 2.13 | 2.58 |
| Nouvelle-Aquitaine | HPV16/18 | Opportunistic | 2020 | (59,66] | 266 | 2.26 | 2.41 | 2.26 | 0.75 | 4.51 | 2.40 | 2.39 | 2.18 | 2.62 |
| Nouvelle-Aquitaine | HPV16/18 | Organised | 2020 | (59,66] | 1 | 0.00 | 2.33 | 0.00 | 0.00 | 0.00 | 2.52 | 2.43 | 1.46 | 4.06 |
| Occitanie | HPV16/18 | Opportunistic | 2020 | (59,66] | 287 | 4.53 | 2.92 | 2.79 | 1.05 | 4.89 | 2.93 | 2.92 | 2.63 | 3.25 |
| Occitanie | HPV16/18 | Organised | 2020 | (59,66] | 1 | 0.00 | 3.23 | 0.00 | 0.00 | 100.00 | 3.05 | 2.98 | 2.01 | 4.39 |
| Pays de la Loire | HPV16/18 | Opportunistic | 2020 | (59,66] | 142 | 0.70 | 2.15 | 2.11 | 0.00 | 4.93 | 2.15 | 2.15 | 1.91 | 2.43 |
| Provence-Alpes-Côte d'Azur | HPV16/18 | Opportunistic | 2020 | (59,66] | 207 | 0.97 | 2.38 | 2.42 | 0.48 | 4.83 | 2.40 | 2.39 | 2.18 | 2.62 |
| Provence-Alpes-Côte d'Azur | HPV16/18 | Organised | 2020 | (59,66] | 2 | 50.00 | 2.80 | 0.00 | 0.00 | 50.00 | 2.71 | 2.66 | 1.88 | 3.84 |
| Île-de-France | HPV16/18 | Opportunistic | 2020 | (59,66] | 4,705 | 2.13 | 2.33 | 2.34 | 1.89 | 2.81 | 2.33 | 2.33 | 2.18 | 2.48 |
| Île-de-France | HPV16/18 | Organised | 2020 | (59,66] | 4 | 0.00 | 2.47 | 0.00 | 0.00 | 25.00 | 2.50 | 2.47 | 1.87 | 3.32 |
| Auvergne-Rhône-Alpes | HPV16/18 | Opportunistic | 2021 | [30,39] | | | 4.90 | 4.94 | 3.68 | 6.19 | 4.89 | 4.88 | 4.47 | 5.33 |
| Auvergne-Rhône-Alpes | HPV16/18 | Organised | 2021 | [30,39] | | | 4.58 | 2.86 | 0.00 | 11.43 | 4.46 | 4.43 | 3.59 | 5.52 |
| Bourgogne-Franche-Comté | HPV16/18 | Opportunistic | 2021 | [30,39] | | | 4.32 | 4.29 | 2.74 | 6.00 | 4.32 | 4.31 | 3.89 | 4.79 |
| Bourgogne-Franche-Comté | HPV16/18 | Organised | 2021 | [30,39] | | | 3.82 | 0.00 | 0.00 | 33.33 | 3.94 | 3.89 | 2.95 | 5.21 |
| Bretagne | HPV16/18 | Opportunistic | 2021 | [30,39] | | | 4.49 | 4.45 | 3.08 | 5.93 | 4.48 | 4.47 | 4.05 | 4.93 |
| Bretagne | HPV16/18 | Organised | 2021 | [30,39] | | | 3.00 | 2.44 | 0.00 | 9.76 | 3.01 | 2.98 | 2.29 | 3.86 |
| Centre-Val de Loire | HPV16/18 | Opportunistic | 2021 | [30,39] | | | 4.39 | 4.38 | 3.75 | 5.04 | 4.40 | 4.40 | 4.17 | 4.63 |

**Table S8** **Observed HR HPV cervical infection prevalence, posterior predictive HR HPV cervical infection prevalence, and posterior expected HR HPV cervical infection prevalence, stratified by various dimensions.** ***(continued)***

| | | | | | | | Posterior distribution | | | | | | | |
|---|---|---|---|---|---|---|---|---|---|---|---|---|---|---|
| | | | | | | | Predictive prevalence | | | | Expected prevalence | | | |
| | | | | | | | | | ETI95% | | | | ETI95% | |
| Region | Genotypes | Type of screening | Year | Age | Number of tests | Observed | Average | Median | LB | UB | Average | Median | LB | UB |
| Centre-Val de Loire | HPV16/18 | Organised | 2021 | [30,39] | | | 3.68 | 3.70 | 1.39 | 6.48 | 3.66 | 3.64 | 3.03 | 4.36 |
| Corse | HPV16/18 | Opportunistic | 2021 | [30,39] | | | 4.70 | 0.00 | 0.00 | 15.38 | 4.73 | 4.70 | 3.85 | 5.80 |
| Grand Est | HPV16/18 | Opportunistic | 2021 | [30,39] | | | 4.20 | 4.16 | 3.01 | 5.40 | 4.18 | 4.18 | 3.88 | 4.51 |
| Grand Est | HPV16/18 | Organised | 2021 | [30,39] | | | 3.63 | 0.00 | 0.00 | 20.00 | 3.37 | 3.33 | 2.56 | 4.36 |
| Hauts-de-France | HPV16/18 | Opportunistic | 2021 | [30,39] | | | 4.19 | 4.18 | 3.58 | 4.85 | 4.19 | 4.18 | 3.96 | 4.41 |
| Hauts-de-France | HPV16/18 | Organised | 2021 | [30,39] | | | 3.58 | 3.70 | 0.74 | 7.41 | 3.62 | 3.61 | 3.00 | 4.34 |
| Normandie | HPV16/18 | Opportunistic | 2021 | [30,39] | | | 4.82 | 4.80 | 3.99 | 5.68 | 4.81 | 4.81 | 4.50 | 5.12 |
| Normandie | HPV16/18 | Organised | 2021 | [30,39] | | | 4.03 | 0.00 | 0.00 | 16.67 | 3.84 | 3.81 | 2.96 | 4.88 |
| Nouvelle-Aquitaine | HPV16/18 | Opportunistic | 2021 | [30,39] | | | 4.42 | 4.41 | 3.66 | 5.22 | 4.41 | 4.41 | 4.16 | 4.67 |
| Nouvelle-Aquitaine | HPV16/18 | Organised | 2021 | [30,39] | | | 3.55 | 3.03 | 0.00 | 12.12 | 3.62 | 3.60 | 2.87 | 4.55 |
| Occitanie | HPV16/18 | Opportunistic | 2021 | [30,39] | | | 5.02 | 4.99 | 3.99 | 6.15 | 5.02 | 5.02 | 4.68 | 5.38 |
| Occitanie | HPV16/18 | Organised | 2021 | [30,39] | | | 4.64 | 3.85 | 0.00 | 11.54 | 4.58 | 4.56 | 3.77 | 5.53 |
| Pays de la Loire | HPV16/18 | Opportunistic | 2021 | [30,39] | | | 3.76 | 3.69 | 2.53 | 5.06 | 3.73 | 3.73 | 3.37 | 4.11 |
| Pays de la Loire | HPV16/18 | Organised | 2021 | [30,39] | | | 3.15 | 3.64 | 0.00 | 9.09 | 3.14 | 3.12 | 2.48 | 3.94 |
| Provence-Alpes-Côte d'Azur | HPV16/18 | Opportunistic | 2021 | [30,39] | | | 4.75 | 4.74 | 4.08 | 5.48 | 4.74 | 4.74 | 4.48 | 5.02 |
| Provence-Alpes-Côte d'Azur | HPV16/18 | Organised | 2021 | [30,39] | | | 4.35 | 5.00 | 0.00 | 15.00 | 4.41 | 4.39 | 3.53 | 5.49 |
| Île-de-France | HPV16/18 | Opportunistic | 2021 | [30,39] | | | 4.42 | 4.42 | 4.14 | 4.71 | 4.42 | 4.43 | 4.27 | 4.57 |
| Île-de-France | HPV16/18 | Organised | 2021 | [30,39] | | | 3.93 | 3.77 | 1.89 | 6.20 | 3.90 | 3.88 | 3.16 | 4.76 |
| Auvergne-Rhône-Alpes | HPV16/18 | Opportunistic | 2021 | (39,49] | 804 | 3.98 | 4.00 | 3.98 | 2.61 | 5.35 | 4.00 | 4.00 | 3.65 | 4.38 |
| Auvergne-Rhône-Alpes | HPV16/18 | Organised | 2021 | (39,49] | 48 | 0.00 | 3.69 | 4.17 | 0.00 | 10.42 | 3.66 | 3.64 | 2.96 | 4.45 |
| Bourgogne-Franche-Comté | HPV16/18 | Opportunistic | 2021 | (39,49] | 391 | 3.07 | 3.58 | 3.58 | 1.79 | 5.63 | 3.58 | 3.57 | 3.22 | 3.97 |
| Bourgogne-Franche-Comté | HPV16/18 | Organised | 2021 | (39,49] | 1 | 0.00 | 2.33 | 0.00 | 0.00 | 0.00 | 2.94 | 2.89 | 2.06 | 4.06 |
| Bretagne | HPV16/18 | Opportunistic | 2021 | (39,49] | 588 | 3.06 | 3.61 | 3.57 | 2.21 | 5.27 | 3.62 | 3.62 | 3.29 | 4.02 |
| Bretagne | HPV16/18 | Organised | 2021 | (39,49] | 16 | 12.50 | 2.35 | 0.00 | 0.00 | 12.50 | 2.40 | 2.37 | 1.84 | 3.06 |
| Centre-Val de Loire | HPV16/18 | Opportunistic | 2021 | (39,49] | 4,201 | 3.19 | 3.60 | 3.59 | 3.02 | 4.19 | 3.59 | 3.59 | 3.39 | 3.80 |
| Centre-Val de Loire | HPV16/18 | Organised | 2021 | (39,49] | 138 | 1.45 | 3.01 | 2.90 | 0.72 | 6.52 | 3.05 | 3.03 | 2.56 | 3.61 |
| Corse | HPV16/18 | Opportunistic | 2021 | (39,49] | 5 | 20.00 | 3.87 | 0.00 | 0.00 | 20.00 | 3.96 | 3.95 | 3.14 | 4.93 |
| Grand Est | HPV16/18 | Opportunistic | 2021 | (39,49] | 1,093 | 3.39 | 3.45 | 3.48 | 2.38 | 4.57 | 3.46 | 3.46 | 3.19 | 3.74 |
| Grand Est | HPV16/18 | Organised | 2021 | (39,49] | 26 | 0.00 | 2.79 | 3.85 | 0.00 | 11.54 | 2.70 | 2.69 | 2.21 | 3.24 |
| Hauts-de-France | HPV16/18 | Opportunistic | 2021 | (39,49] | 2,916 | 3.88 | 3.49 | 3.46 | 2.81 | 4.15 | 3.48 | 3.48 | 3.29 | 3.68 |
| Hauts-de-France | HPV16/18 | Organised | 2021 | (39,49] | 110 | 1.82 | 2.99 | 2.73 | 0.00 | 6.36 | 2.99 | 2.98 | 2.48 | 3.55 |
| Normandie | HPV16/18 | Opportunistic | 2021 | (39,49] | 2,310 | 4.94 | 3.92 | 3.90 | 3.12 | 4.81 | 3.92 | 3.92 | 3.66 | 4.18 |
| Normandie | HPV16/18 | Organised | 2021 | (39,49] | 5 | 0.00 | 3.55 | 0.00 | 0.00 | 20.00 | 3.40 | 3.38 | 2.63 | 4.31 |
| Nouvelle-Aquitaine | HPV16/18 | Opportunistic | 2021 | (39,49] | 2,359 | 3.65 | 3.59 | 3.60 | 2.88 | 4.37 | 3.60 | 3.60 | 3.38 | 3.82 |
| Nouvelle-Aquitaine | HPV16/18 | Organised | 2021 | (39,49] | 3 | 0.00 | 3.20 | 0.00 | 0.00 | 33.33 | 3.20 | 3.17 | 2.43 | 4.10 |
| Occitanie | HPV16/18 | Opportunistic | 2021 | (39,49] | 1,638 | 4.15 | 4.12 | 4.09 | 3.11 | 5.19 | 4.13 | 4.13 | 3.85 | 4.43 |
| Occitanie | HPV16/18 | Organised | 2021 | (39,49] | 110 | 3.64 | 3.68 | 3.64 | 0.91 | 7.27 | 3.70 | 3.69 | 3.02 | 4.45 |
| Pays de la Loire | HPV16/18 | Opportunistic | 2021 | (39,49] | 834 | 2.76 | 3.13 | 3.12 | 1.92 | 4.44 | 3.14 | 3.13 | 2.85 | 3.44 |
| Pays de la Loire | HPV16/18 | Organised | 2021 | (39,49] | 85 | 1.18 | 2.59 | 2.35 | 0.00 | 5.88 | 2.68 | 2.67 | 2.15 | 3.28 |

**Table S8** **Observed HR HPV cervical infection prevalence, posterior predictive HR HPV cervical infection prevalence, and posterior expected HR HPV cervical infection prevalence, stratified by various dimensions.** ***(continued)***

| | | | | | | | Posterior distribution | | | | | | | |
|---|---|---|---|---|---|---|---|---|---|---|---|---|---|---|
| | | | | | | | Predictive prevalence | | | | Expected prevalence | | | |
| | | | | | | | | | ETI95% | | | | ETI95% | |
| Region | Genotypes | Type of screening | Year | Age | Number of tests | Observed | Average | Median | LB | UB | Average | Median | LB | UB |
| Provence-Alpes-Côte d'Azur | HPV16/18 | Opportunistic | 2021 | (39,49] | 3,170 | 3.72 | 3.86 | 3.85 | 3.12 | 4.57 | 3.85 | 3.85 | 3.62 | 4.10 |
| Provence-Alpes-Côte d'Azur | HPV16/18 | Organised | 2021 | (39,49] | 7 | 0.00 | 3.79 | 0.00 | 0.00 | 28.57 | 3.73 | 3.70 | 3.00 | 4.56 |
| Île-de-France | HPV16/18 | Opportunistic | 2021 | (39,49] | 25,340 | 3.44 | 3.62 | 3.62 | 3.36 | 3.88 | 3.62 | 3.62 | 3.48 | 3.76 |
| Île-de-France | HPV16/18 | Organised | 2021 | (39,49] | 83 | 1.20 | 3.19 | 2.41 | 0.00 | 7.23 | 3.18 | 3.17 | 2.69 | 3.73 |
| Auvergne-Rhône-Alpes | HPV16/18 | Opportunistic | 2021 | (49,59] | 666 | 2.25 | 3.18 | 3.15 | 1.95 | 4.65 | 3.20 | 3.20 | 2.91 | 3.52 |
| Auvergne-Rhône-Alpes | HPV16/18 | Organised | 2021 | (49,59] | 38 | 0.00 | 2.96 | 2.63 | 0.00 | 7.89 | 3.03 | 3.02 | 2.46 | 3.71 |
| Bourgogne-Franche-Comté | HPV16/18 | Opportunistic | 2021 | (49,59] | 329 | 3.34 | 2.90 | 2.74 | 1.22 | 4.86 | 2.90 | 2.90 | 2.62 | 3.23 |
| Bourgogne-Franche-Comté | HPV16/18 | Organised | 2021 | (49,59] | 1 | 0.00 | 2.77 | 0.00 | 0.00 | 100.00 | 2.46 | 2.42 | 1.73 | 3.35 |
| Bretagne | HPV16/18 | Opportunistic | 2021 | (49,59] | 556 | 2.16 | 2.85 | 2.88 | 1.44 | 4.32 | 2.83 | 2.83 | 2.55 | 3.14 |
| Bretagne | HPV16/18 | Organised | 2021 | (49,59] | 36 | 0.00 | 1.99 | 2.78 | 0.00 | 8.33 | 2.04 | 2.03 | 1.59 | 2.61 |
| Centre-Val de Loire | HPV16/18 | Opportunistic | 2021 | (49,59] | 3,844 | 2.21 | 2.86 | 2.86 | 2.32 | 3.43 | 2.86 | 2.86 | 2.70 | 3.04 |
| Centre-Val de Loire | HPV16/18 | Organised | 2021 | (49,59] | 224 | 2.23 | 2.48 | 2.23 | 0.45 | 4.91 | 2.48 | 2.47 | 2.11 | 2.92 |
| Corse | HPV16/18 | Opportunistic | 2021 | (49,59] | 9 | 0.00 | 2.96 | 0.00 | 0.00 | 11.11 | 2.97 | 2.95 | 2.53 | 3.44 |
| Corse | HPV16/18 | Organised | 2021 | (49,59] | 1 | 0.00 | 2.37 | 0.00 | 0.00 | 0.00 | 2.84 | 2.77 | 1.84 | 4.21 |
| Grand Est | HPV16/18 | Opportunistic | 2021 | (49,59] | 1,003 | 2.79 | 2.82 | 2.79 | 1.79 | 3.89 | 2.79 | 2.79 | 2.57 | 3.02 |
| Grand Est | HPV16/18 | Organised | 2021 | (49,59] | 59 | 0.00 | 2.27 | 1.69 | 0.00 | 6.78 | 2.34 | 2.33 | 1.91 | 2.85 |
| Hauts-de-France | HPV16/18 | Opportunistic | 2021 | (49,59] | 2,015 | 3.18 | 2.77 | 2.78 | 2.08 | 3.52 | 2.78 | 2.78 | 2.61 | 2.95 |
| Hauts-de-France | HPV16/18 | Organised | 2021 | (49,59] | 96 | 3.12 | 2.40 | 2.08 | 0.00 | 6.25 | 2.44 | 2.43 | 2.07 | 2.85 |
| Normandie | HPV16/18 | Opportunistic | 2021 | (49,59] | 1,829 | 2.90 | 3.09 | 3.06 | 2.30 | 3.94 | 3.09 | 3.09 | 2.87 | 3.31 |
| Normandie | HPV16/18 | Organised | 2021 | (49,59] | 5 | 0.00 | 2.74 | 0.00 | 0.00 | 20.00 | 2.63 | 2.62 | 2.03 | 3.34 |
| Nouvelle-Aquitaine | HPV16/18 | Opportunistic | 2021 | (49,59] | 1,995 | 2.86 | 2.89 | 2.86 | 2.16 | 3.66 | 2.88 | 2.88 | 2.71 | 3.07 |
| Nouvelle-Aquitaine | HPV16/18 | Organised | 2021 | (49,59] | 170 | 2.94 | 2.53 | 2.35 | 0.59 | 5.29 | 2.54 | 2.53 | 2.16 | 2.97 |
| Occitanie | HPV16/18 | Opportunistic | 2021 | (49,59] | 1,372 | 3.94 | 3.34 | 3.35 | 2.41 | 4.37 | 3.35 | 3.35 | 3.10 | 3.61 |
| Occitanie | HPV16/18 | Organised | 2021 | (49,59] | 95 | 3.16 | 3.35 | 3.16 | 0.00 | 7.37 | 3.35 | 3.33 | 2.73 | 4.05 |
| Pays de la Loire | HPV16/18 | Opportunistic | 2021 | (49,59] | 703 | 3.13 | 2.52 | 2.56 | 1.42 | 3.84 | 2.52 | 2.51 | 2.27 | 2.77 |
| Pays de la Loire | HPV16/18 | Organised | 2021 | (49,59] | 78 | 2.56 | 2.33 | 2.56 | 0.00 | 6.41 | 2.29 | 2.27 | 1.86 | 2.79 |
| Provence-Alpes-Côte d'Azur | HPV16/18 | Opportunistic | 2021 | (49,59] | 2,577 | 2.91 | 3.05 | 3.03 | 2.37 | 3.76 | 3.06 | 3.05 | 2.86 | 3.27 |
| Provence-Alpes-Côte d'Azur | HPV16/18 | Organised | 2021 | (49,59] | 7 | 0.00 | 3.00 | 0.00 | 0.00 | 14.29 | 2.98 | 2.95 | 2.35 | 3.75 |
| Île-de-France | HPV16/18 | Opportunistic | 2021 | (49,59] | 18,998 | 2.84 | 2.89 | 2.89 | 2.63 | 3.16 | 2.89 | 2.89 | 2.76 | 3.01 |
| Île-de-France | HPV16/18 | Organised | 2021 | (49,59] | 161 | 3.73 | 2.51 | 2.48 | 0.62 | 4.97 | 2.53 | 2.52 | 2.15 | 2.96 |
| Auvergne-Rhône-Alpes | HPV16/18 | Opportunistic | 2021 | (59,66] | 330 | 0.91 | 2.95 | 3.03 | 1.21 | 4.85 | 2.95 | 2.94 | 2.66 | 3.25 |
| Auvergne-Rhône-Alpes | HPV16/18 | Organised | 2021 | (59,66] | 31 | 0.00 | 2.96 | 3.23 | 0.00 | 9.68 | 2.92 | 2.90 | 2.34 | 3.59 |
| Bourgogne-Franche-Comté | HPV16/18 | Opportunistic | 2021 | (59,66] | 207 | 4.83 | 2.69 | 2.42 | 0.97 | 5.31 | 2.71 | 2.70 | 2.42 | 3.01 |
| Bourgogne-Franche-Comté | HPV16/18 | Organised | 2021 | (59,66] | 22 | 4.55 | 2.42 | 0.00 | 0.00 | 9.09 | 2.46 | 2.45 | 1.96 | 3.06 |
| Bretagne | HPV16/18 | Opportunistic | 2021 | (59,66] | 239 | 1.67 | 2.63 | 2.51 | 0.84 | 5.02 | 2.65 | 2.64 | 2.36 | 2.96 |
| Bretagne | HPV16/18 | Organised | 2021 | (59,66] | 21 | 9.52 | 2.29 | 0.00 | 0.00 | 9.52 | 2.17 | 2.14 | 1.64 | 2.85 |
| Centre-Val de Loire | HPV16/18 | Opportunistic | 2021 | (59,66] | 2,018 | 2.63 | 2.65 | 2.63 | 1.93 | 3.42 | 2.66 | 2.65 | 2.47 | 2.86 |
| Centre-Val de Loire | HPV16/18 | Organised | 2021 | (59,66] | 143 | 2.10 | 2.35 | 2.10 | 0.00 | 4.90 | 2.38 | 2.36 | 1.98 | 2.82 |
| Corse | HPV16/18 | Opportunistic | 2021 | (59,66] | 7 | 0.00 | 3.00 | 0.00 | 0.00 | 14.29 | 3.03 | 3.01 | 2.44 | 3.74 |

**Table S8** **Observed HR HPV cervical infection prevalence, posterior predictive HR HPV cervical infection prevalence, and posterior expected HR HPV cervical infection prevalence, stratified by various dimensions.** ***(continued)***

| | | | | | | | Posterior distribution | | | | | | | |
|---|---|---|---|---|---|---|---|---|---|---|---|---|---|---|
| | | | | | | | Predictive prevalence | | | | Expected prevalence | | | |
| | | | | | | | | | ETI95% | | | | ETI95% | |
| Region | Genotypes | Type of screening | Year | Age | Number of tests | Observed | Average | Median | LB | UB | Average | Median | LB | UB |
| Grand Est | HPV16/18 | Opportunistic | 2021 | (59,66] | 562 | 2.14 | 2.60 | 2.49 | 1.25 | 3.91 | 2.59 | 2.59 | 2.37 | 2.83 |
| Grand Est | HPV16/18 | Organised | 2021 | (59,66] | 46 | 2.17 | 2.24 | 2.17 | 0.00 | 6.52 | 2.29 | 2.27 | 1.82 | 2.85 |
| Hauts-de-France | HPV16/18 | Opportunistic | 2021 | (59,66] | 931 | 2.26 | 2.56 | 2.58 | 1.61 | 3.65 | 2.56 | 2.56 | 2.37 | 2.76 |
| Hauts-de-France | HPV16/18 | Organised | 2021 | (59,66] | 49 | 2.04 | 2.43 | 2.04 | 0.00 | 8.16 | 2.49 | 2.47 | 2.03 | 3.03 |
| Normandie | HPV16/18 | Opportunistic | 2021 | (59,66] | 950 | 2.63 | 2.86 | 2.84 | 1.89 | 4.00 | 2.86 | 2.85 | 2.62 | 3.11 |
| Normandie | HPV16/18 | Organised | 2021 | (59,66] | 18 | 11.11 | 2.63 | 0.00 | 0.00 | 11.11 | 2.51 | 2.50 | 2.04 | 3.08 |
| Nouvelle-Aquitaine | HPV16/18 | Opportunistic | 2021 | (59,66] | 1,222 | 2.70 | 2.70 | 2.70 | 1.80 | 3.68 | 2.71 | 2.71 | 2.51 | 2.92 |
| Nouvelle-Aquitaine | HPV16/18 | Organised | 2021 | (59,66] | 185 | 4.32 | 2.61 | 2.70 | 0.54 | 5.41 | 2.58 | 2.57 | 2.14 | 3.06 |
| Occitanie | HPV16/18 | Opportunistic | 2021 | (59,66] | 720 | 3.06 | 3.09 | 3.06 | 1.94 | 4.44 | 3.10 | 3.09 | 2.85 | 3.36 |
| Occitanie | HPV16/18 | Organised | 2021 | (59,66] | 48 | 0.00 | 3.21 | 2.08 | 0.00 | 8.33 | 3.22 | 3.20 | 2.59 | 3.93 |
| Pays de la Loire | HPV16/18 | Opportunistic | 2021 | (59,66] | 326 | 3.07 | 2.47 | 2.45 | 0.92 | 4.29 | 2.46 | 2.46 | 2.20 | 2.74 |
| Pays de la Loire | HPV16/18 | Organised | 2021 | (59,66] | 52 | 0.00 | 2.28 | 1.92 | 0.00 | 7.69 | 2.27 | 2.25 | 1.80 | 2.83 |
| Provence-Alpes-Côte d'Azur | HPV16/18 | Opportunistic | 2021 | (59,66] | 1,314 | 2.82 | 2.83 | 2.82 | 1.98 | 3.73 | 2.83 | 2.83 | 2.62 | 3.06 |
| Provence-Alpes-Côte d'Azur | HPV16/18 | Organised | 2021 | (59,66] | 22 | 4.55 | 2.78 | 0.00 | 0.00 | 9.09 | 2.78 | 2.75 | 2.19 | 3.52 |
| Île-de-France | HPV16/18 | Opportunistic | 2021 | (59,66] | 8,881 | 2.80 | 2.69 | 2.69 | 2.32 | 3.07 | 2.69 | 2.69 | 2.53 | 2.86 |
| Île-de-France | HPV16/18 | Organised | 2021 | (59,66] | 1,384 | 2.38 | 2.61 | 2.60 | 1.66 | 3.61 | 2.61 | 2.59 | 2.12 | 3.17 |
| Auvergne-Rhône-Alpes | HPV16/18 | Opportunistic | 2022 | [30,39] | | | 5.58 | 5.61 | 1.87 | 10.28 | 5.62 | 5.60 | 5.03 | 6.25 |
| Auvergne-Rhône-Alpes | HPV16/18 | Organised | 2022 | [30,39] | | | 4.14 | 0.00 | 0.00 | 16.67 | 4.24 | 4.22 | 3.45 | 5.19 |
| Bourgogne-Franche-Comté | HPV16/18 | Opportunistic | 2022 | [30,39] | | | 4.69 | 4.55 | 2.90 | 6.63 | 4.68 | 4.67 | 4.19 | 5.22 |
| Bourgogne-Franche-Comté | HPV16/18 | Organised | 2022 | [30,39] | | | 3.35 | 3.85 | 0.00 | 11.54 | 3.48 | 3.46 | 2.78 | 4.31 |
| Bretagne | HPV16/18 | Opportunistic | 2022 | [30,39] | | | 5.07 | 5.05 | 3.77 | 6.54 | 5.08 | 5.08 | 4.64 | 5.58 |
| Bretagne | HPV16/18 | Organised | 2022 | [30,39] | | | 3.12 | 3.18 | 0.64 | 6.37 | 3.15 | 3.13 | 2.52 | 3.91 |
| Centre-Val de Loire | HPV16/18 | Opportunistic | 2022 | [30,39] | | | 4.65 | 4.63 | 3.74 | 5.59 | 4.65 | 4.64 | 4.38 | 4.94 |
| Centre-Val de Loire | HPV16/18 | Organised | 2022 | [30,39] | | | 3.38 | 3.43 | 0.98 | 5.88 | 3.41 | 3.40 | 2.91 | 3.95 |
| Corse | HPV16/18 | Opportunistic | 2022 | [30,39] | | | 5.11 | 0.00 | 0.00 | 25.00 | 5.19 | 5.15 | 4.27 | 6.30 |
| Grand Est | HPV16/18 | Opportunistic | 2022 | [30,39] | | | 4.56 | 4.54 | 3.32 | 5.85 | 4.57 | 4.56 | 4.20 | 4.95 |
| Grand Est | HPV16/18 | Organised | 2022 | [30,39] | | | 3.30 | 3.15 | 0.90 | 5.86 | 3.30 | 3.29 | 2.77 | 3.92 |
| Hauts-de-France | HPV16/18 | Opportunistic | 2022 | [30,39] | | | 4.52 | 4.50 | 3.83 | 5.24 | 4.52 | 4.52 | 4.26 | 4.78 |
| Hauts-de-France | HPV16/18 | Organised | 2022 | [30,39] | | | 3.40 | 3.35 | 1.68 | 5.59 | 3.41 | 3.40 | 2.90 | 3.98 |
| Normandie | HPV16/18 | Opportunistic | 2022 | [30,39] | | | 5.38 | 5.38 | 4.40 | 6.41 | 5.37 | 5.37 | 5.03 | 5.74 |
| Normandie | HPV16/18 | Organised | 2022 | [30,39] | | | 3.43 | 4.55 | 0.00 | 13.64 | 3.54 | 3.52 | 2.91 | 4.29 |
| Nouvelle-Aquitaine | HPV16/18 | Opportunistic | 2022 | [30,39] | | | 4.82 | 4.80 | 4.04 | 5.59 | 4.82 | 4.81 | 4.52 | 5.10 |
| Nouvelle-Aquitaine | HPV16/18 | Organised | 2022 | [30,39] | | | 3.65 | 3.53 | 1.60 | 5.77 | 3.66 | 3.65 | 3.12 | 4.26 |
| Occitanie | HPV16/18 | Opportunistic | 2022 | [30,39] | | | 5.26 | 5.25 | 3.79 | 6.96 | 5.24 | 5.24 | 4.86 | 5.66 |
| Occitanie | HPV16/18 | Organised | 2022 | [30,39] | | | 4.04 | 4.23 | 1.41 | 7.75 | 4.07 | 4.06 | 3.42 | 4.78 |
| Pays de la Loire | HPV16/18 | Opportunistic | 2022 | [30,39] | | | 4.43 | 4.31 | 1.96 | 7.06 | 4.39 | 4.38 | 3.98 | 4.83 |
| Pays de la Loire | HPV16/18 | Organised | 2022 | [30,39] | | | 2.98 | 3.17 | 0.00 | 7.94 | 3.00 | 2.99 | 2.50 | 3.59 |
| Provence-Alpes-Côte d'Azur | HPV16/18 | Opportunistic | 2022 | [30,39] | | | 5.18 | 5.18 | 4.36 | 6.06 | 5.17 | 5.17 | 4.86 | 5.49 |
| Provence-Alpes-Côte d'Azur | HPV16/18 | Organised | 2022 | [30,39] | | | 4.44 | 4.15 | 2.07 | 7.47 | 4.44 | 4.41 | 3.69 | 5.33 |

**Table S8 Observed HR HPV cervical infection prevalence, posterior predictive HR HPV cervical infection prevalence, and posterior expected HR HPV cervical infection prevalence, stratified by various dimensions.** ***(continued)***

| | | | | | | | Posterior distribution | | | | | | | |
|---|---|---|---|---|---|---|---|---|---|---|---|---|---|---|
| | | | | | | | Predictive prevalence | | | | Expected prevalence | | | |
| | | | | | | | | | ETI95% | | | | ETI95% | |
| Region | Genotypes | Type of screening | Year | Age | Number of tests | Observed | Average | Median | LB | UB | Average | Median | LB | UB |
| Île-de-France | HPV16/18 | Opportunistic | 2022 | [30,39] | | | 4.94 | 4.93 | 4.37 | 5.54 | 4.94 | 4.94 | 4.70 | 5.18 |
| Île-de-France | HPV16/18 | Organised | 2022 | [30,39] | | | 3.79 | 3.72 | 1.86 | 6.19 | 3.79 | 3.77 | 3.24 | 4.40 |
| Auvergne-Rhône-Alpes | HPV16/18 | Opportunistic | 2022 | (39,49] | 64 | 9.38 | 4.78 | 4.69 | 0.00 | 10.94 | 4.78 | 4.77 | 4.16 | 5.51 |
| Auvergne-Rhône-Alpes | HPV16/18 | Organised | 2022 | (39,49] | 15 | 20.00 | 3.48 | 0.00 | 0.00 | 13.33 | 3.59 | 3.56 | 2.93 | 4.37 |
| Bourgogne-Franche-Comté | HPV16/18 | Opportunistic | 2022 | (39,49] | 333 | 3.60 | 3.83 | 3.90 | 1.80 | 6.01 | 3.82 | 3.81 | 3.40 | 4.26 |
| Bourgogne-Franche-Comté | HPV16/18 | Organised | 2022 | (39,49] | 1 | 0.00 | 3.07 | 0.00 | 0.00 | 100.00 | 2.85 | 2.81 | 2.02 | 3.94 |
| Bretagne | HPV16/18 | Opportunistic | 2022 | (39,49] | 663 | 4.37 | 4.16 | 4.07 | 2.71 | 5.88 | 4.16 | 4.15 | 3.79 | 4.56 |
| Bretagne | HPV16/18 | Organised | 2022 | (39,49] | 173 | 4.62 | 2.56 | 2.31 | 0.58 | 5.20 | 2.55 | 2.54 | 2.09 | 3.11 |
| Centre-Val de Loire | HPV16/18 | Opportunistic | 2022 | (39,49] | 2,079 | 4.95 | 3.80 | 3.80 | 2.98 | 4.67 | 3.82 | 3.81 | 3.58 | 4.05 |
| Centre-Val de Loire | HPV16/18 | Organised | 2022 | (39,49] | 248 | 4.03 | 2.82 | 2.82 | 0.81 | 5.24 | 2.83 | 2.82 | 2.45 | 3.22 |
| Corse | HPV16/18 | Opportunistic | 2022 | (39,49] | 3 | 0.00 | 5.26 | 0.00 | 0.00 | 33.33 | 4.76 | 4.72 | 3.68 | 6.06 |
| Corse | HPV16/18 | Organised | 2022 | (39,49] | 2 | 0.00 | 3.17 | 0.00 | 0.00 | 50.00 | 3.15 | 3.11 | 2.28 | 4.23 |
| Grand Est | HPV16/18 | Opportunistic | 2022 | (39,49] | 858 | 4.55 | 3.84 | 3.85 | 2.56 | 5.24 | 3.84 | 3.83 | 3.52 | 4.19 |
| Grand Est | HPV16/18 | Organised | 2022 | (39,49] | 198 | 3.54 | 2.78 | 2.53 | 0.51 | 5.56 | 2.74 | 2.73 | 2.29 | 3.20 |
| Hauts-de-France | HPV16/18 | Opportunistic | 2022 | (39,49] | 2,382 | 4.49 | 3.76 | 3.78 | 2.98 | 4.58 | 3.77 | 3.77 | 3.54 | 4.00 |
| Hauts-de-France | HPV16/18 | Organised | 2022 | (39,49] | 286 | 1.75 | 2.80 | 2.80 | 1.05 | 4.90 | 2.81 | 2.80 | 2.42 | 3.23 |
| Normandie | HPV16/18 | Opportunistic | 2022 | (39,49] | 1,814 | 3.80 | 4.40 | 4.41 | 3.42 | 5.40 | 4.39 | 4.39 | 4.10 | 4.69 |
| Normandie | HPV16/18 | Organised | 2022 | (39,49] | 22 | 4.55 | 3.04 | 4.55 | 0.00 | 13.64 | 3.05 | 3.05 | 2.59 | 3.56 |
| Nouvelle-Aquitaine | HPV16/18 | Opportunistic | 2022 | (39,49] | 2,812 | 4.30 | 3.97 | 3.98 | 3.24 | 4.77 | 3.97 | 3.97 | 3.72 | 4.22 |
| Nouvelle-Aquitaine | HPV16/18 | Organised | 2022 | (39,49] | 337 | 2.97 | 2.99 | 2.97 | 1.19 | 5.04 | 3.00 | 2.99 | 2.61 | 3.41 |
| Occitanie | HPV16/18 | Opportunistic | 2022 | (39,49] | 746 | 3.62 | 4.30 | 4.29 | 2.82 | 5.76 | 4.31 | 4.30 | 3.98 | 4.66 |
| Occitanie | HPV16/18 | Organised | 2022 | (39,49] | 135 | 2.22 | 3.32 | 2.96 | 0.74 | 6.67 | 3.34 | 3.33 | 2.83 | 3.91 |
| Pays de la Loire | HPV16/18 | Opportunistic | 2022 | (39,49] | 203 | 1.48 | 3.68 | 3.45 | 1.48 | 6.40 | 3.66 | 3.65 | 3.31 | 4.04 |
| Pays de la Loire | HPV16/18 | Organised | 2022 | (39,49] | 71 | 4.23 | 2.62 | 2.82 | 0.00 | 7.04 | 2.55 | 2.55 | 2.15 | 3.00 |
| Provence-Alpes-Côte d'Azur | HPV16/18 | Opportunistic | 2022 | (39,49] | 2,056 | 4.23 | 4.21 | 4.18 | 3.36 | 5.16 | 4.22 | 4.22 | 3.96 | 4.50 |
| Provence-Alpes-Côte d'Azur | HPV16/18 | Organised | 2022 | (39,49] | 112 | 3.57 | 3.72 | 3.57 | 0.89 | 7.14 | 3.74 | 3.72 | 3.13 | 4.44 |
| Île-de-France | HPV16/18 | Opportunistic | 2022 | (39,49] | 4,399 | 3.71 | 3.98 | 3.98 | 3.39 | 4.59 | 3.97 | 3.97 | 3.77 | 4.18 |
| Île-de-France | HPV16/18 | Organised | 2022 | (39,49] | 342 | 3.22 | 3.12 | 2.92 | 1.46 | 5.26 | 3.12 | 3.12 | 2.70 | 3.58 |
| Auvergne-Rhône-Alpes | HPV16/18 | Opportunistic | 2022 | (49,59] | 49 | 6.12 | 4.10 | 4.08 | 0.00 | 10.20 | 3.97 | 3.95 | 3.43 | 4.59 |
| Auvergne-Rhône-Alpes | HPV16/18 | Organised | 2022 | (49,59] | 13 | 7.69 | 3.12 | 0.00 | 0.00 | 15.38 | 3.10 | 3.07 | 2.41 | 3.90 |
| Bourgogne-Franche-Comté | HPV16/18 | Opportunistic | 2022 | (49,59] | 239 | 2.51 | 3.11 | 2.93 | 1.26 | 5.44 | 3.10 | 3.09 | 2.78 | 3.47 |
| Bourgogne-Franche-Comté | HPV16/18 | Organised | 2022 | (49,59] | 1 | 0.00 | 2.57 | 0.00 | 0.00 | 100.00 | 2.47 | 2.43 | 1.71 | 3.46 |
| Bretagne | HPV16/18 | Opportunistic | 2022 | (49,59] | 535 | 2.62 | 3.32 | 3.36 | 1.87 | 4.86 | 3.31 | 3.31 | 3.00 | 3.65 |
| Bretagne | HPV16/18 | Organised | 2022 | (49,59] | 144 | 1.39 | 2.11 | 2.08 | 0.00 | 4.86 | 2.11 | 2.10 | 1.71 | 2.56 |
| Centre-Val de Loire | HPV16/18 | Opportunistic | 2022 | (49,59] | 1,838 | 3.54 | 3.04 | 3.05 | 2.29 | 3.86 | 3.05 | 3.05 | 2.86 | 3.27 |
| Centre-Val de Loire | HPV16/18 | Organised | 2022 | (49,59] | 278 | 3.60 | 2.40 | 2.16 | 0.72 | 4.32 | 2.40 | 2.40 | 2.08 | 2.76 |
| Corse | HPV16/18 | Opportunistic | 2022 | (49,59] | 2 | 0.00 | 3.60 | 0.00 | 0.00 | 50.00 | 3.54 | 3.49 | 2.51 | 4.83 |
| Grand Est | HPV16/18 | Opportunistic | 2022 | (49,59] | 767 | 2.35 | 3.13 | 3.13 | 1.96 | 4.43 | 3.12 | 3.11 | 2.84 | 3.40 |
| Grand Est | HPV16/18 | Organised | 2022 | (49,59] | 120 | 1.67 | 2.36 | 2.50 | 0.00 | 5.00 | 2.38 | 2.37 | 2.00 | 2.79 |

**Table S8 Observed HR HPV cervical infection prevalence, posterior predictive HR HPV cervical infection prevalence, and posterior expected HR HPV cervical infection prevalence, stratified by various dimensions. *(continued)***

| | | | | | | | Posterior distribution | | | | | | | |
|---|---|---|---|---|---|---|---|---|---|---|---|---|---|---|
| | | | | | | | Predictive prevalence | | | | Expected prevalence | | | |
| | | | | | | | | | ETI95% | | | | ETI95% | |
| Region | Genotypes | Type of screening | Year | Age | Number of tests | Observed | Average | Median | LB | UB | Average | Median | LB | UB |
| Hauts-de-France | HPV16/18 | Opportunistic | 2022 | (49,59] | 1,725 | 3.01 | 3.02 | 3.01 | 2.20 | 3.88 | 3.02 | 3.02 | 2.83 | 3.22 |
| Hauts-de-France | HPV16/18 | Organised | 2022 | (49,59] | 267 | 3.00 | 2.35 | 2.25 | 0.75 | 4.12 | 2.34 | 2.34 | 2.02 | 2.70 |
| Normandie | HPV16/18 | Opportunistic | 2022 | (49,59] | 1,196 | 4.26 | 3.42 | 3.43 | 2.42 | 4.52 | 3.43 | 3.43 | 3.20 | 3.68 |
| Normandie | HPV16/18 | Organised | 2022 | (49,59] | 16 | 0.00 | 2.55 | 0.00 | 0.00 | 12.50 | 2.48 | 2.47 | 2.08 | 2.94 |
| Nouvelle-Aquitaine | HPV16/18 | Opportunistic | 2022 | (49,59] | 2,090 | 3.11 | 3.18 | 3.16 | 2.44 | 3.97 | 3.18 | 3.18 | 2.99 | 3.39 |
| Nouvelle-Aquitaine | HPV16/18 | Organised | 2022 | (49,59] | 644 | 2.17 | 2.52 | 2.48 | 1.40 | 3.88 | 2.51 | 2.51 | 2.18 | 2.87 |
| Occitanie | HPV16/18 | Opportunistic | 2022 | (49,59] | 710 | 3.38 | 3.47 | 3.38 | 2.11 | 4.93 | 3.47 | 3.47 | 3.19 | 3.75 |
| Occitanie | HPV16/18 | Organised | 2022 | (49,59] | 141 | 0.71 | 2.86 | 2.84 | 0.71 | 5.67 | 2.86 | 2.85 | 2.43 | 3.35 |
| Pays de la Loire | HPV16/18 | Opportunistic | 2022 | (49,59] | 158 | 1.90 | 3.02 | 3.16 | 0.63 | 5.70 | 3.01 | 3.01 | 2.71 | 3.33 |
| Pays de la Loire | HPV16/18 | Organised | 2022 | (49,59] | 67 | 0.00 | 2.12 | 1.49 | 0.00 | 5.97 | 2.15 | 2.15 | 1.82 | 2.52 |
| Provence-Alpes-Côte d'Azur | HPV16/18 | Opportunistic | 2022 | (49,59] | 1,791 | 3.13 | 3.33 | 3.29 | 2.46 | 4.24 | 3.33 | 3.33 | 3.11 | 3.56 |
| Provence-Alpes-Côte d'Azur | HPV16/18 | Organised | 2022 | (49,59] | 85 | 3.53 | 3.14 | 2.35 | 0.00 | 7.06 | 3.11 | 3.10 | 2.59 | 3.73 |
| Île-de-France | HPV16/18 | Opportunistic | 2022 | (49,59] | 3,353 | 2.74 | 3.14 | 3.13 | 2.56 | 3.76 | 3.13 | 3.13 | 2.96 | 3.31 |
| Île-de-France | HPV16/18 | Organised | 2022 | (49,59] | 449 | 2.00 | 2.53 | 2.45 | 1.11 | 4.23 | 2.53 | 2.53 | 2.20 | 2.93 |
| Auvergne-Rhône-Alpes | HPV16/18 | Opportunistic | 2022 | (59,66] | 27 | 3.70 | 3.50 | 3.70 | 0.00 | 11.11 | 3.43 | 3.42 | 3.00 | 3.91 |
| Auvergne-Rhône-Alpes | HPV16/18 | Organised | 2022 | (59,66] | 8 | 0.00 | 3.25 | 0.00 | 0.00 | 25.00 | 3.13 | 3.11 | 2.37 | 4.08 |
| Bourgogne-Franche-Comté | HPV16/18 | Opportunistic | 2022 | (59,66] | 120 | 1.67 | 2.83 | 2.50 | 0.00 | 5.83 | 2.85 | 2.84 | 2.52 | 3.21 |
| Bourgogne-Franche-Comté | HPV16/18 | Organised | 2022 | (59,66] | 17 | 5.88 | 2.33 | 0.00 | 0.00 | 11.76 | 2.39 | 2.37 | 1.89 | 2.99 |
| Bretagne | HPV16/18 | Opportunistic | 2022 | (59,66] | 289 | 4.15 | 3.05 | 3.11 | 1.38 | 5.19 | 3.04 | 3.04 | 2.73 | 3.38 |
| Bretagne | HPV16/18 | Organised | 2022 | (59,66] | 112 | 0.00 | 2.00 | 1.79 | 0.00 | 5.36 | 2.01 | 2.00 | 1.59 | 2.51 |
| Centre-Val de Loire | HPV16/18 | Opportunistic | 2022 | (59,66] | 872 | 3.21 | 2.81 | 2.75 | 1.72 | 4.01 | 2.81 | 2.81 | 2.59 | 3.04 |
| Centre-Val de Loire | HPV16/18 | Organised | 2022 | (59,66] | 188 | 3.19 | 2.25 | 2.13 | 0.53 | 4.79 | 2.28 | 2.27 | 1.91 | 2.70 |
| Corse | HPV16/18 | Opportunistic | 2022 | (59,66] | 2 | 0.00 | 3.98 | 0.00 | 0.00 | 50.00 | 3.96 | 3.89 | 2.74 | 5.63 |
| Grand Est | HPV16/18 | Opportunistic | 2022 | (59,66] | 312 | 2.56 | 2.89 | 2.88 | 1.28 | 4.81 | 2.87 | 2.87 | 2.61 | 3.16 |
| Grand Est | HPV16/18 | Organised | 2022 | (59,66] | 82 | 2.44 | 2.32 | 2.44 | 0.00 | 6.10 | 2.32 | 2.31 | 1.89 | 2.81 |
| Hauts-de-France | HPV16/18 | Opportunistic | 2022 | (59,66] | 714 | 2.38 | 2.81 | 2.80 | 1.68 | 4.06 | 2.82 | 2.82 | 2.61 | 3.04 |
| Hauts-de-France | HPV16/18 | Organised | 2022 | (59,66] | 169 | 2.96 | 2.27 | 2.37 | 0.59 | 4.73 | 2.29 | 2.28 | 1.90 | 2.72 |
| Normandie | HPV16/18 | Opportunistic | 2022 | (59,66] | 612 | 3.10 | 3.19 | 3.10 | 1.80 | 4.58 | 3.17 | 3.16 | 2.91 | 3.44 |
| Normandie | HPV16/18 | Organised | 2022 | (59,66] | 93 | 1.08 | 2.43 | 2.15 | 0.00 | 6.45 | 2.44 | 2.42 | 2.00 | 2.96 |
| Nouvelle-Aquitaine | HPV16/18 | Opportunistic | 2022 | (59,66] | 1,040 | 3.37 | 2.95 | 2.88 | 2.02 | 4.04 | 2.95 | 2.95 | 2.73 | 3.18 |
| Nouvelle-Aquitaine | HPV16/18 | Organised | 2022 | (59,66] | 440 | 1.82 | 2.42 | 2.27 | 1.14 | 4.09 | 2.43 | 2.43 | 2.04 | 2.88 |
| Occitanie | HPV16/18 | Opportunistic | 2022 | (59,66] | 349 | 3.72 | 3.22 | 3.15 | 1.43 | 5.16 | 3.21 | 3.21 | 2.92 | 3.51 |
| Occitanie | HPV16/18 | Organised | 2022 | (59,66] | 89 | 2.25 | 2.69 | 2.25 | 0.00 | 6.74 | 2.72 | 2.70 | 2.26 | 3.26 |
| Pays de la Loire | HPV16/18 | Opportunistic | 2022 | (59,66] | 105 | 2.86 | 2.94 | 2.86 | 0.00 | 6.67 | 2.89 | 2.88 | 2.55 | 3.26 |
| Pays de la Loire | HPV16/18 | Organised | 2022 | (59,66] | 40 | 0.00 | 2.22 | 2.50 | 0.00 | 7.50 | 2.15 | 2.14 | 1.75 | 2.62 |
| Provence-Alpes-Côte d'Azur | HPV16/18 | Opportunistic | 2022 | (59,66] | 836 | 2.99 | 3.07 | 2.99 | 1.91 | 4.31 | 3.08 | 3.07 | 2.83 | 3.33 |
| Provence-Alpes-Côte d'Azur | HPV16/18 | Organised | 2022 | (59,66] | 66 | 4.55 | 2.88 | 3.03 | 0.00 | 7.58 | 2.88 | 2.87 | 2.35 | 3.55 |
| Île-de-France | HPV16/18 | Opportunistic | 2022 | (59,66] | 1,520 | 2.50 | 2.91 | 2.89 | 2.11 | 3.82 | 2.91 | 2.90 | 2.71 | 3.12 |
| Île-de-France | HPV16/18 | Organised | 2022 | (59,66] | 193 | 3.11 | 2.41 | 2.07 | 0.52 | 4.66 | 2.43 | 2.41 | 2.04 | 2.89 |

**Table S8 Observed HR HPV cervical infection prevalence, posterior predictive HR HPV cervical infection prevalence, and posterior expected HR HPV cervical infection prevalence, stratified by various dimensions.** ***(continued)***

| | | | | | | | Posterior distribution | | | | | | | |
|---|---|---|---|---|---|---|---|---|---|---|---|---|---|---|
| | | | | | | | Predictive prevalence | | | | Expected prevalence | | | |
| | | | | | | | | | ETI95% | | | | ETI95% | |
| Region | Genotypes | Type of screening | Year | Age | Number of tests | Observed | Average | Median | LB | UB | Average | Median | LB | UB |
| Auvergne-Rhône-Alpes | HPV16/18 | Opportunistic | 2023 | [30,39] | | | 6.83 | 7.04 | 1.41 | 14.08 | 6.79 | 6.77 | 5.83 | 7.88 |
| Auvergne-Rhône-Alpes | HPV16/18 | Organised | 2023 | [30,39] | | | 4.31 | 5.26 | 0.00 | 15.79 | 4.38 | 4.34 | 3.45 | 5.49 |
| Bourgogne-Franche-Comté | HPV16/18 | Opportunistic | 2023 | [30,39] | | | 5.08 | 4.89 | 2.22 | 8.00 | 5.09 | 5.08 | 4.48 | 5.76 |
| Bourgogne-Franche-Comté | HPV16/18 | Organised | 2023 | [30,39] | | | 3.47 | 2.50 | 0.00 | 10.00 | 3.39 | 3.37 | 2.70 | 4.21 |
| Bretagne | HPV16/18 | Opportunistic | 2023 | [30,39] | | | 5.72 | 5.72 | 4.38 | 7.06 | 5.74 | 5.74 | 5.26 | 6.25 |
| Bretagne | HPV16/18 | Organised | 2023 | [30,39] | | | 3.24 | 3.23 | 1.72 | 5.16 | 3.23 | 3.21 | 2.63 | 3.96 |
| Centre-Val de Loire | HPV16/18 | Opportunistic | 2023 | [30,39] | | | 5.16 | 5.12 | 4.18 | 6.20 | 5.15 | 5.15 | 4.83 | 5.47 |
| Centre-Val de Loire | HPV16/18 | Organised | 2023 | [30,39] | | | 3.46 | 3.57 | 0.71 | 7.14 | 3.47 | 3.45 | 2.92 | 4.06 |
| Corse | HPV16/18 | Opportunistic | 2023 | [30,39] | | | 5.13 | 0.00 | 0.00 | 100.00 | 5.09 | 4.93 | 2.81 | 8.48 |
| Grand Est | HPV16/18 | Opportunistic | 2023 | [30,39] | | | 5.07 | 5.01 | 3.76 | 6.46 | 5.07 | 5.06 | 4.66 | 5.51 |
| Grand Est | HPV16/18 | Organised | 2023 | [30,39] | | | 3.27 | 2.97 | 0.00 | 6.93 | 3.27 | 3.26 | 2.70 | 3.91 |
| Hauts-de-France | HPV16/18 | Opportunistic | 2023 | [30,39] | | | 5.02 | 5.02 | 4.22 | 5.81 | 5.01 | 5.01 | 4.73 | 5.31 |
| Hauts-de-France | HPV16/18 | Organised | 2023 | [30,39] | | | 3.33 | 2.88 | 0.72 | 6.47 | 3.32 | 3.31 | 2.76 | 3.93 |
| Normandie | HPV16/18 | Opportunistic | 2023 | [30,39] | | | 6.02 | 6.03 | 4.58 | 7.47 | 6.03 | 6.03 | 5.61 | 6.46 |
| Normandie | HPV16/18 | Organised | 2023 | [30,39] | | | 3.64 | 2.56 | 0.00 | 10.26 | 3.73 | 3.71 | 3.15 | 4.39 |
| Nouvelle-Aquitaine | HPV16/18 | Opportunistic | 2023 | [30,39] | | | 5.42 | 5.40 | 4.53 | 6.34 | 5.42 | 5.41 | 5.08 | 5.75 |
| Nouvelle-Aquitaine | HPV16/18 | Organised | 2023 | [30,39] | | | 3.72 | 3.69 | 1.97 | 5.67 | 3.76 | 3.75 | 3.19 | 4.41 |
| Occitanie | HPV16/18 | Opportunistic | 2023 | [30,39] | | | 5.75 | 5.71 | 3.43 | 8.29 | 5.77 | 5.77 | 5.29 | 6.28 |
| Occitanie | HPV16/18 | Organised | 2023 | [30,39] | | | 4.15 | 2.94 | 0.00 | 11.76 | 4.12 | 4.11 | 3.43 | 4.87 |
| Pays de la Loire | HPV16/18 | Opportunistic | 2023 | [30,39] | | | 4.89 | 4.93 | 2.63 | 7.57 | 4.89 | 4.88 | 4.41 | 5.42 |
| Pays de la Loire | HPV16/18 | Organised | 2023 | [30,39] | | | 3.14 | 2.84 | 0.71 | 6.38 | 3.15 | 3.14 | 2.58 | 3.77 |
| Provence-Alpes-Côte d'Azur | HPV16/18 | Opportunistic | 2023 | [30,39] | | | 5.86 | 5.84 | 4.83 | 6.99 | 5.88 | 5.88 | 5.52 | 6.26 |
| Provence-Alpes-Côte d'Azur | HPV16/18 | Organised | 2023 | [30,39] | | | 4.51 | 4.39 | 1.95 | 7.80 | 4.47 | 4.46 | 3.67 | 5.43 |
| Île-de-France | HPV16/18 | Opportunistic | 2023 | [30,39] | | | 5.55 | 5.55 | 4.93 | 6.20 | 5.55 | 5.55 | 5.28 | 5.83 |
| Île-de-France | HPV16/18 | Organised | 2023 | [30,39] | | | 3.87 | 3.73 | 1.87 | 6.13 | 3.88 | 3.87 | 3.25 | 4.60 |
| Auvergne-Rhône-Alpes | HPV16/18 | Opportunistic | 2023 | (39,49] | 48 | 4.17 | 5.62 | 6.25 | 0.00 | 12.50 | 5.61 | 5.59 | 4.77 | 6.61 |
| Auvergne-Rhône-Alpes | HPV16/18 | Organised | 2023 | (39,49] | 17 | 5.88 | 3.84 | 0.00 | 0.00 | 17.65 | 4.02 | 3.99 | 3.05 | 5.16 |
| Bourgogne-Franche-Comté | HPV16/18 | Opportunistic | 2023 | (39,49] | 162 | 4.32 | 4.28 | 4.32 | 1.23 | 7.41 | 4.21 | 4.20 | 3.69 | 4.80 |
| Bourgogne-Franche-Comté | HPV16/18 | Organised | 2023 | (39,49] | 64 | 1.56 | 2.86 | 3.12 | 0.00 | 7.81 | 2.84 | 2.82 | 2.29 | 3.49 |
| Bretagne | HPV16/18 | Opportunistic | 2023 | (39,49] | 966 | 5.07 | 4.68 | 4.66 | 3.42 | 6.11 | 4.66 | 4.65 | 4.26 | 5.11 |
| Bretagne | HPV16/18 | Organised | 2023 | (39,49] | 406 | 2.46 | 2.62 | 2.46 | 1.23 | 4.43 | 2.64 | 2.62 | 2.16 | 3.20 |
| Centre-Val de Loire | HPV16/18 | Opportunistic | 2023 | (39,49] | 1,493 | 5.89 | 4.26 | 4.22 | 3.28 | 5.43 | 4.28 | 4.27 | 4.02 | 4.56 |
| Centre-Val de Loire | HPV16/18 | Organised | 2023 | (39,49] | 133 | 3.01 | 3.00 | 3.01 | 0.75 | 6.02 | 2.98 | 2.98 | 2.57 | 3.47 |
| Corse | HPV16/18 | Opportunistic | 2023 | (39,49] | 1 | 100.00 | 4.50 | 0.00 | 0.00 | 100.00 | 5.06 | 4.84 | 2.69 | 8.51 |
| Grand Est | HPV16/18 | Opportunistic | 2023 | (39,49] | 843 | 4.86 | 4.31 | 4.27 | 2.97 | 5.81 | 4.31 | 4.30 | 3.95 | 4.69 |
| Grand Est | HPV16/18 | Organised | 2023 | (39,49] | 101 | 1.98 | 2.78 | 2.97 | 0.00 | 6.93 | 2.81 | 2.80 | 2.33 | 3.35 |
| Hauts-de-France | HPV16/18 | Opportunistic | 2023 | (39,49] | 2,152 | 5.11 | 4.23 | 4.23 | 3.39 | 5.11 | 4.22 | 4.22 | 3.97 | 4.49 |
| Hauts-de-France | HPV16/18 | Organised | 2023 | (39,49] | 100 | 3.00 | 2.88 | 3.00 | 0.00 | 7.00 | 2.83 | 2.82 | 2.40 | 3.32 |
| Normandie | HPV16/18 | Opportunistic | 2023 | (39,49] | 1,088 | 4.41 | 4.92 | 4.87 | 3.68 | 6.34 | 4.93 | 4.93 | 4.58 | 5.28 |

**Table S8 Observed HR HPV cervical infection prevalence, posterior predictive HR HPV cervical infection prevalence, and posterior expected HR HPV cervical infection prevalence, stratified by various dimensions.** ***(continued)***

| | | | | | | | Posterior distribution | | | | | | | |
|---|---|---|---|---|---|---|---|---|---|---|---|---|---|---|
| | | | | | | | Predictive prevalence | | | | Expected prevalence | | | |
| | | | | | | | | | ETI95% | | | | ETI95% | |
| Region | Genotypes | Type of screening | Year | Age | Number of tests | Observed | Average | Median | LB | UB | Average | Median | LB | UB |
| Normandie | HPV16/18 | Organised | 2023 | (39,49] | 42 | 0.00 | 3.18 | 2.38 | 0.00 | 9.52 | 3.13 | 3.12 | 2.65 | 3.67 |
| Nouvelle-Aquitaine | HPV16/18 | Opportunistic | 2023 | (39,49] | 2,209 | 3.71 | 4.47 | 4.44 | 3.58 | 5.39 | 4.47 | 4.47 | 4.20 | 4.76 |
| Nouvelle-Aquitaine | HPV16/18 | Organised | 2023 | (39,49] | 441 | 4.31 | 3.16 | 3.17 | 1.59 | 4.99 | 3.17 | 3.15 | 2.72 | 3.67 |
| Occitanie | HPV16/18 | Opportunistic | 2023 | (39,49] | 353 | 5.10 | 4.88 | 4.82 | 2.83 | 7.37 | 4.89 | 4.88 | 4.44 | 5.36 |
| Occitanie | HPV16/18 | Organised | 2023 | (39,49] | 51 | 0.00 | 3.48 | 3.92 | 0.00 | 9.80 | 3.49 | 3.48 | 2.92 | 4.10 |
| Pays de la Loire | HPV16/18 | Opportunistic | 2023 | (39,49] | 299 | 5.69 | 4.08 | 4.01 | 2.01 | 6.35 | 4.09 | 4.08 | 3.69 | 4.55 |
| Pays de la Loire | HPV16/18 | Organised | 2023 | (39,49] | 131 | 3.82 | 2.56 | 2.29 | 0.00 | 5.34 | 2.56 | 2.55 | 2.13 | 3.04 |
| Provence-Alpes-Côte d'Azur | HPV16/18 | Opportunistic | 2023 | (39,49] | 1,551 | 5.54 | 4.77 | 4.77 | 3.74 | 5.93 | 4.76 | 4.76 | 4.45 | 5.09 |
| Provence-Alpes-Côte d'Azur | HPV16/18 | Organised | 2023 | (39,49] | 139 | 3.60 | 3.68 | 3.60 | 0.72 | 7.19 | 3.67 | 3.65 | 3.07 | 4.39 |
| Île-de-France | HPV16/18 | Opportunistic | 2023 | (39,49] | 3,580 | 4.72 | 4.52 | 4.53 | 3.83 | 5.22 | 4.51 | 4.51 | 4.28 | 4.75 |
| Île-de-France | HPV16/18 | Organised | 2023 | (39,49] | 364 | 1.92 | 3.18 | 3.02 | 1.37 | 5.22 | 3.18 | 3.16 | 2.72 | 3.71 |
| Auvergne-Rhône-Alpes | HPV16/18 | Opportunistic | 2023 | (49,59] | 45 | 6.67 | 4.39 | 4.44 | 0.00 | 11.11 | 4.46 | 4.44 | 3.73 | 5.32 |
| Auvergne-Rhône-Alpes | HPV16/18 | Organised | 2023 | (49,59] | 9 | 0.00 | 3.05 | 0.00 | 0.00 | 11.11 | 3.10 | 3.06 | 2.37 | 4.03 |
| Bourgogne-Franche-Comté | HPV16/18 | Opportunistic | 2023 | (49,59] | 104 | 1.92 | 3.42 | 2.88 | 0.00 | 7.69 | 3.43 | 3.42 | 3.02 | 3.89 |
| Bourgogne-Franche-Comté | HPV16/18 | Organised | 2023 | (49,59] | 54 | 0.00 | 2.40 | 1.85 | 0.00 | 7.41 | 2.35 | 2.34 | 1.85 | 2.94 |
| Bretagne | HPV16/18 | Opportunistic | 2023 | (49,59] | 750 | 4.27 | 3.74 | 3.73 | 2.40 | 5.20 | 3.74 | 3.73 | 3.40 | 4.11 |
| Bretagne | HPV16/18 | Organised | 2023 | (49,59] | 389 | 2.57 | 2.19 | 2.06 | 0.77 | 3.86 | 2.19 | 2.18 | 1.78 | 2.67 |
| Centre-Val de Loire | HPV16/18 | Opportunistic | 2023 | (49,59] | 1,356 | 3.02 | 3.46 | 3.47 | 2.51 | 4.42 | 3.45 | 3.45 | 3.22 | 3.68 |
| Centre-Val de Loire | HPV16/18 | Organised | 2023 | (49,59] | 169 | 1.78 | 2.43 | 2.37 | 0.59 | 5.33 | 2.45 | 2.44 | 2.07 | 2.88 |
| Grand Est | HPV16/18 | Opportunistic | 2023 | (49,59] | 693 | 3.46 | 3.46 | 3.46 | 2.16 | 4.91 | 3.44 | 3.43 | 3.13 | 3.76 |
| Grand Est | HPV16/18 | Organised | 2023 | (49,59] | 129 | 3.88 | 2.25 | 2.33 | 0.00 | 5.43 | 2.28 | 2.26 | 1.86 | 2.75 |
| Hauts-de-France | HPV16/18 | Opportunistic | 2023 | (49,59] | 1,553 | 3.86 | 3.36 | 3.35 | 2.45 | 4.31 | 3.36 | 3.36 | 3.15 | 3.59 |
| Hauts-de-France | HPV16/18 | Organised | 2023 | (49,59] | 118 | 2.54 | 2.32 | 2.54 | 0.00 | 5.08 | 2.34 | 2.33 | 1.96 | 2.80 |
| Normandie | HPV16/18 | Opportunistic | 2023 | (49,59] | 825 | 4.12 | 3.92 | 3.88 | 2.67 | 5.33 | 3.90 | 3.89 | 3.61 | 4.19 |
| Normandie | HPV16/18 | Organised | 2023 | (49,59] | 35 | 2.86 | 2.67 | 2.86 | 0.00 | 8.57 | 2.59 | 2.58 | 2.18 | 3.05 |
| Nouvelle-Aquitaine | HPV16/18 | Opportunistic | 2023 | (49,59] | 1,712 | 4.03 | 3.58 | 3.56 | 2.69 | 4.50 | 3.58 | 3.58 | 3.36 | 3.82 |
| Nouvelle-Aquitaine | HPV16/18 | Organised | 2023 | (49,59] | 456 | 2.41 | 2.60 | 2.63 | 1.32 | 4.17 | 2.63 | 2.63 | 2.25 | 3.06 |
| Occitanie | HPV16/18 | Opportunistic | 2023 | (49,59] | 307 | 4.56 | 3.90 | 3.91 | 1.95 | 6.19 | 3.90 | 3.89 | 3.53 | 4.29 |
| Occitanie | HPV16/18 | Organised | 2023 | (49,59] | 60 | 1.67 | 2.78 | 3.33 | 0.00 | 6.67 | 2.83 | 2.82 | 2.35 | 3.38 |
| Pays de la Loire | HPV16/18 | Opportunistic | 2023 | (49,59] | 274 | 2.19 | 3.23 | 3.28 | 1.46 | 5.47 | 3.26 | 3.26 | 2.93 | 3.63 |
| Pays de la Loire | HPV16/18 | Organised | 2023 | (49,59] | 129 | 3.10 | 2.22 | 2.33 | 0.00 | 5.43 | 2.23 | 2.22 | 1.85 | 2.65 |
| Provence-Alpes-Côte d'Azur | HPV16/18 | Opportunistic | 2023 | (49,59] | 1,337 | 3.74 | 3.81 | 3.81 | 2.77 | 4.86 | 3.80 | 3.80 | 3.54 | 4.08 |
| Provence-Alpes-Côte d'Azur | HPV16/18 | Organised | 2023 | (49,59] | 131 | 3.05 | 2.97 | 3.05 | 0.76 | 6.11 | 3.01 | 2.99 | 2.46 | 3.65 |
| Île-de-France | HPV16/18 | Opportunistic | 2023 | (49,59] | 2,833 | 4.52 | 3.57 | 3.57 | 2.89 | 4.27 | 3.57 | 3.57 | 3.37 | 3.79 |
| Île-de-France | HPV16/18 | Organised | 2023 | (49,59] | 313 | 3.19 | 2.60 | 2.56 | 0.96 | 4.47 | 2.60 | 2.58 | 2.20 | 3.06 |
| Auvergne-Rhône-Alpes | HPV16/18 | Opportunistic | 2023 | (59,66] | 28 | 0.00 | 4.22 | 3.57 | 0.00 | 14.29 | 4.15 | 4.13 | 3.47 | 4.93 |
| Auvergne-Rhône-Alpes | HPV16/18 | Organised | 2023 | (59,66] | 4 | 0.00 | 3.01 | 0.00 | 0.00 | 25.00 | 2.90 | 2.87 | 2.15 | 3.77 |
| Bourgogne-Franche-Comté | HPV16/18 | Opportunistic | 2023 | (59,66] | 60 | 3.33 | 3.18 | 3.33 | 0.00 | 8.33 | 3.21 | 3.20 | 2.79 | 3.68 |
| Bourgogne-Franche-Comté | HPV16/18 | Organised | 2023 | (59,66] | 32 | 3.12 | 2.25 | 3.12 | 0.00 | 9.38 | 2.30 | 2.28 | 1.77 | 2.95 |

**Table S8** **Observed HR HPV cervical infection prevalence, posterior predictive HR HPV cervical infection prevalence, and posterior expected HR HPV cervical infection prevalence, stratified by various dimensions.** ***(continued)***

| | | | | | | | Posterior distribution | | | | | | | |
|---|---|---|---|---|---|---|---|---|---|---|---|---|---|---|
| | | | | | | | Predictive prevalence | | | | Expected prevalence | | | |
| | | | | | | | | | ETI95% | | | | ETI95% | |
| Region | Genotypes | Type of screening | Year | Age | Number of tests | Observed | Average | Median | LB | UB | Average | Median | LB | UB |
| Bretagne | HPV16/18 | Opportunistic | 2023 | (59,66] | 388 | 4.38 | 3.40 | 3.35 | 1.80 | 5.15 | 3.38 | 3.38 | 3.04 | 3.75 |
| Bretagne | HPV16/18 | Organised | 2023 | (59,66] | 337 | 1.48 | 2.09 | 2.08 | 0.59 | 3.86 | 2.11 | 2.10 | 1.66 | 2.67 |
| Centre-Val de Loire | HPV16/18 | Opportunistic | 2023 | (59,66] | 629 | 3.50 | 3.17 | 3.18 | 1.91 | 4.61 | 3.15 | 3.15 | 2.91 | 3.41 |
| Centre-Val de Loire | HPV16/18 | Organised | 2023 | (59,66] | 120 | 2.50 | 2.28 | 2.50 | 0.00 | 5.00 | 2.27 | 2.26 | 1.85 | 2.76 |
| Corse | HPV16/18 | Opportunistic | 2023 | (59,66] | 3 | 0.00 | 3.07 | 0.00 | 0.00 | 33.33 | 3.41 | 3.35 | 2.42 | 4.69 |
| Grand Est | HPV16/18 | Opportunistic | 2023 | (59,66] | 384 | 2.60 | 3.16 | 3.12 | 1.56 | 4.95 | 3.19 | 3.19 | 2.90 | 3.51 |
| Grand Est | HPV16/18 | Organised | 2023 | (59,66] | 89 | 0.00 | 2.27 | 2.25 | 0.00 | 5.62 | 2.25 | 2.24 | 1.77 | 2.81 |
| Hauts-de-France | HPV16/18 | Opportunistic | 2023 | (59,66] | 840 | 2.38 | 3.15 | 3.10 | 2.02 | 4.40 | 3.14 | 3.14 | 2.89 | 3.39 |
| Hauts-de-France | HPV16/18 | Organised | 2023 | (59,66] | 116 | 3.45 | 2.31 | 1.72 | 0.00 | 5.17 | 2.31 | 2.30 | 1.87 | 2.82 |
| Normandie | HPV16/18 | Opportunistic | 2023 | (59,66] | 381 | 3.15 | 3.55 | 3.41 | 1.84 | 5.51 | 3.55 | 3.55 | 3.26 | 3.87 |
| Normandie | HPV16/18 | Organised | 2023 | (59,66] | 71 | 4.23 | 2.47 | 2.82 | 0.00 | 7.04 | 2.50 | 2.49 | 2.02 | 3.05 |
| Nouvelle-Aquitaine | HPV16/18 | Opportunistic | 2023 | (59,66] | 859 | 3.84 | 3.33 | 3.26 | 2.10 | 4.54 | 3.34 | 3.34 | 3.09 | 3.63 |
| Nouvelle-Aquitaine | HPV16/18 | Organised | 2023 | (59,66] | 308 | 2.60 | 2.57 | 2.60 | 0.97 | 4.55 | 2.55 | 2.53 | 2.08 | 3.08 |
| Occitanie | HPV16/18 | Opportunistic | 2023 | (59,66] | 168 | 3.57 | 3.59 | 3.57 | 1.19 | 6.55 | 3.59 | 3.58 | 3.23 | 3.97 |
| Occitanie | HPV16/18 | Organised | 2023 | (59,66] | 52 | 1.92 | 2.78 | 1.92 | 0.00 | 7.69 | 2.79 | 2.77 | 2.23 | 3.48 |
| Pays de la Loire | HPV16/18 | Opportunistic | 2023 | (59,66] | 126 | 3.97 | 3.21 | 3.17 | 0.79 | 6.35 | 3.22 | 3.21 | 2.85 | 3.61 |
| Pays de la Loire | HPV16/18 | Organised | 2023 | (59,66] | 97 | 3.09 | 2.13 | 2.06 | 0.00 | 5.15 | 2.12 | 2.11 | 1.70 | 2.59 |
| Provence-Alpes-Côte d'Azur | HPV16/18 | Opportunistic | 2023 | (59,66] | 667 | 3.30 | 3.54 | 3.45 | 2.25 | 5.10 | 3.55 | 3.55 | 3.26 | 3.85 |
| Provence-Alpes-Côte d'Azur | HPV16/18 | Organised | 2023 | (59,66] | 87 | 3.45 | 2.93 | 2.30 | 0.00 | 6.90 | 2.89 | 2.88 | 2.31 | 3.60 |
| Île-de-France | HPV16/18 | Opportunistic | 2023 | (59,66] | 1,258 | 4.69 | 3.29 | 3.26 | 2.31 | 4.37 | 3.29 | 3.28 | 3.07 | 3.53 |
| Île-de-France | HPV16/18 | Organised | 2023 | (59,66] | 143 | 4.20 | 2.53 | 2.10 | 0.68 | 5.59 | 2.51 | 2.49 | 2.03 | 3.06 |
| Auvergne-Rhône-Alpes | Other genotypes | Opportunistic | 2020 | [30,39] | | | 12.65 | 12.53 | 9.40 | 15.90 | 12.64 | 12.63 | 11.67 | 13.64 |
| Bourgogne-Franche-Comté | Other genotypes | Opportunistic | 2020 | [30,39] | | | 11.21 | 11.11 | 6.76 | 15.94 | 11.23 | 11.22 | 10.27 | 12.29 |
| Bretagne | Other genotypes | Opportunistic | 2020 | [30,39] | | | 10.92 | 10.92 | 6.99 | 14.86 | 10.91 | 10.91 | 10.02 | 11.85 |
| Centre-Val de Loire | Other genotypes | Opportunistic | 2020 | [30,39] | | | 11.25 | 11.21 | 9.37 | 13.14 | 11.25 | 11.25 | 10.77 | 11.77 |
| Corse | Other genotypes | Opportunistic | 2020 | [30,39] | | | 13.82 | 14.13 | 6.52 | 21.74 | 13.89 | 13.83 | 11.30 | 16.88 |
| Grand Est | Other genotypes | Opportunistic | 2020 | [30,39] | | | 11.13 | 11.06 | 7.23 | 15.32 | 11.17 | 11.16 | 10.28 | 12.16 |
| Hauts-de-France | Other genotypes | Opportunistic | 2020 | [30,39] | | | 10.94 | 10.95 | 9.00 | 12.82 | 10.91 | 10.91 | 10.43 | 11.42 |
| Hauts-de-France | Other genotypes | Organised | 2020 | [30,39] | | | 9.17 | 0.00 | 0.00 | 100.00 | 9.64 | 9.53 | 6.86 | 13.08 |
| Normandie | Other genotypes | Opportunistic | 2020 | [30,39] | | | 11.63 | 11.58 | 9.47 | 13.80 | 11.62 | 11.62 | 11.01 | 12.24 |
| Nouvelle-Aquitaine | Other genotypes | Opportunistic | 2020 | [30,39] | | | 12.00 | 11.89 | 9.51 | 14.74 | 12.01 | 12.01 | 11.47 | 12.56 |
| Occitanie | Other genotypes | Opportunistic | 2020 | [30,39] | | | 14.12 | 14.03 | 11.42 | 16.78 | 14.16 | 14.15 | 13.15 | 15.22 |
| Pays de la Loire | Other genotypes | Opportunistic | 2020 | [30,39] | | | 9.46 | 9.54 | 6.54 | 12.53 | 9.49 | 9.47 | 8.64 | 10.38 |
| Pays de la Loire | Other genotypes | Organised | 2020 | [30,39] | | | 6.02 | 0.00 | 0.00 | 33.33 | 6.13 | 6.07 | 4.61 | 8.08 |
| Provence-Alpes-Côte d'Azur | Other genotypes | Opportunistic | 2020 | [30,39] | | | 12.42 | 12.36 | 10.21 | 14.88 | 12.43 | 12.44 | 11.78 | 13.08 |
| Île-de-France | Other genotypes | Opportunistic | 2020 | [30,39] | | | 12.49 | 12.49 | 11.95 | 13.06 | 12.50 | 12.49 | 12.20 | 12.80 |
| Île-de-France | Other genotypes | Organised | 2020 | [30,39] | | | 9.92 | 12.50 | 0.00 | 37.50 | 10.12 | 10.09 | 8.11 | 12.47 |
| Auvergne-Rhône-Alpes | Other genotypes | Opportunistic | 2020 | (39,49] | 262 | 7.25 | 8.93 | 8.78 | 5.34 | 12.60 | 8.91 | 8.91 | 8.20 | 9.66 |
| Bourgogne-Franche-Comté | Other genotypes | Opportunistic | 2020 | (39,49] | 144 | 6.94 | 7.95 | 7.64 | 3.47 | 12.50 | 7.93 | 7.92 | 7.25 | 8.68 |

**Table S8 Observed HR HPV cervical infection prevalence, posterior predictive HR HPV cervical infection prevalence, and posterior expected HR HPV cervical infection prevalence, stratified by various dimensions.** ***(continued)***

| | | | | | | | Posterior distribution | | | | | | | |
|---|---|---|---|---|---|---|---|---|---|---|---|---|---|---|
| | | | | | | | Predictive prevalence | | | | Expected prevalence | | | |
| | | | | | | | | | ETI95% | | | | ETI95% | |
| Region | Genotypes | Type of screening | Year | Age | Number of tests | Observed | Average | Median | LB | UB | Average | Median | LB | UB |
| Bretagne | Other genotypes | Opportunistic | 2020 | (39,49] | 158 | 10.76 | 7.80 | 7.59 | 3.80 | 12.03 | 7.86 | 7.86 | 7.10 | 8.71 |
| Centre-Val de Loire | Other genotypes | Opportunistic | 2020 | (39,49] | 1,001 | 7.59 | 7.91 | 7.89 | 6.29 | 9.69 | 7.89 | 7.89 | 7.54 | 8.27 |
| Corse | Other genotypes | Opportunistic | 2020 | (39,49] | 118 | 15.25 | 10.29 | 10.17 | 5.08 | 16.10 | 10.22 | 10.17 | 8.16 | 12.70 |
| Grand Est | Other genotypes | Opportunistic | 2020 | (39,49] | 241 | 8.71 | 7.85 | 7.88 | 4.56 | 11.21 | 7.84 | 7.83 | 7.16 | 8.61 |
| Hauts-de-France | Other genotypes | Opportunistic | 2020 | (39,49] | 757 | 7.27 | 7.66 | 7.66 | 5.68 | 9.51 | 7.66 | 7.66 | 7.30 | 8.05 |
| Hauts-de-France | Other genotypes | Organised | 2020 | (39,49] | 2 | 0.00 | 5.80 | 0.00 | 0.00 | 50.00 | 5.73 | 5.67 | 4.20 | 7.58 |
| Normandie | Other genotypes | Opportunistic | 2020 | (39,49] | 675 | 8.44 | 8.00 | 8.00 | 5.93 | 10.22 | 7.99 | 7.99 | 7.55 | 8.46 |
| Nouvelle-Aquitaine | Other genotypes | Opportunistic | 2020 | (39,49] | 424 | 7.78 | 8.33 | 8.25 | 5.90 | 11.08 | 8.37 | 8.36 | 7.97 | 8.78 |
| Occitanie | Other genotypes | Opportunistic | 2020 | (39,49] | 628 | 8.60 | 10.56 | 10.51 | 8.12 | 13.22 | 10.54 | 10.54 | 9.69 | 11.39 |
| Pays de la Loire | Other genotypes | Opportunistic | 2020 | (39,49] | 369 | 7.05 | 6.86 | 6.78 | 4.34 | 9.49 | 6.85 | 6.84 | 6.23 | 7.51 |
| Pays de la Loire | Other genotypes | Organised | 2020 | (39,49] | 1 | 0.00 | 4.23 | 0.00 | 0.00 | 100.00 | 4.61 | 4.54 | 3.14 | 6.47 |
| Provence-Alpes-Côte d'Azur | Other genotypes | Opportunistic | 2020 | (39,49] | 620 | 9.52 | 8.43 | 8.39 | 6.29 | 10.81 | 8.44 | 8.43 | 7.94 | 8.96 |
| Île-de-France | Other genotypes | Opportunistic | 2020 | (39,49] | 16,219 | 8.47 | 8.75 | 8.75 | 8.26 | 9.24 | 8.75 | 8.74 | 8.51 | 8.99 |
| Île-de-France | Other genotypes | Organised | 2020 | (39,49] | 1 | 0.00 | 6.10 | 0.00 | 0.00 | 100.00 | 6.33 | 6.29 | 4.81 | 8.22 |
| Auvergne-Rhône-Alpes | Other genotypes | Opportunistic | 2020 | (49,59] | 215 | 6.51 | 7.63 | 7.44 | 4.19 | 11.16 | 7.68 | 7.68 | 7.05 | 8.36 |
| Bourgogne-Franche-Comté | Other genotypes | Opportunistic | 2020 | (49,59] | 98 | 6.12 | 6.87 | 7.14 | 2.04 | 12.24 | 6.92 | 6.90 | 6.33 | 7.57 |
| Bretagne | Other genotypes | Opportunistic | 2020 | (49,59] | 107 | 6.54 | 6.61 | 6.54 | 2.80 | 12.15 | 6.58 | 6.56 | 5.91 | 7.32 |
| Centre-Val de Loire | Other genotypes | Opportunistic | 2020 | (49,59] | 971 | 5.77 | 6.64 | 6.59 | 5.15 | 8.34 | 6.65 | 6.64 | 6.33 | 6.98 |
| Centre-Val de Loire | Other genotypes | Organised | 2020 | (49,59] | 2 | 0.00 | 5.60 | 0.00 | 0.00 | 50.00 | 5.65 | 5.58 | 4.23 | 7.34 |
| Corse | Other genotypes | Opportunistic | 2020 | (49,59] | 111 | 6.31 | 8.62 | 8.11 | 3.60 | 14.41 | 8.55 | 8.51 | 6.84 | 10.62 |
| Grand Est | Other genotypes | Opportunistic | 2020 | (49,59] | 279 | 5.73 | 6.52 | 6.45 | 3.58 | 9.68 | 6.51 | 6.50 | 5.86 | 7.24 |
| Hauts-de-France | Other genotypes | Opportunistic | 2020 | (49,59] | 626 | 7.03 | 6.44 | 6.39 | 4.63 | 8.47 | 6.45 | 6.45 | 6.11 | 6.80 |
| Normandie | Other genotypes | Opportunistic | 2020 | (49,59] | 521 | 6.91 | 6.68 | 6.72 | 4.61 | 9.02 | 6.69 | 6.69 | 6.32 | 7.09 |
| Nouvelle-Aquitaine | Other genotypes | Opportunistic | 2020 | (49,59] | 367 | 8.45 | 7.10 | 7.08 | 4.63 | 9.82 | 7.12 | 7.12 | 6.75 | 7.52 |
| Occitanie | Other genotypes | Opportunistic | 2020 | (49,59] | 533 | 9.76 | 8.69 | 8.63 | 6.19 | 11.26 | 8.66 | 8.66 | 8.01 | 9.34 |
| Occitanie | Other genotypes | Organised | 2020 | (49,59] | 3 | 33.33 | 6.54 | 0.00 | 0.00 | 33.33 | 6.84 | 6.77 | 4.87 | 9.33 |
| Pays de la Loire | Other genotypes | Opportunistic | 2020 | (49,59] | 278 | 5.76 | 5.80 | 5.76 | 3.24 | 8.63 | 5.83 | 5.82 | 5.33 | 6.37 |
| Pays de la Loire | Other genotypes | Organised | 2020 | (49,59] | 1 | 0.00 | 4.43 | 0.00 | 0.00 | 100.00 | 4.49 | 4.41 | 3.13 | 6.28 |
| Provence-Alpes-Côte d'Azur | Other genotypes | Opportunistic | 2020 | (49,59] | 509 | 8.64 | 7.13 | 7.07 | 5.11 | 9.43 | 7.12 | 7.12 | 6.69 | 7.55 |
| Île-de-France | Other genotypes | Opportunistic | 2020 | (49,59] | 11,874 | 7.20 | 7.42 | 7.42 | 6.91 | 7.96 | 7.42 | 7.42 | 7.19 | 7.66 |
| Île-de-France | Other genotypes | Organised | 2020 | (49,59] | 3 | 0.00 | 7.42 | 0.00 | 0.00 | 33.33 | 7.46 | 7.43 | 5.98 | 9.16 |
| Auvergne-Rhône-Alpes | Other genotypes | Opportunistic | 2020 | (59,66] | 96 | 10.42 | 6.84 | 6.25 | 2.08 | 12.50 | 6.87 | 6.86 | 6.26 | 7.53 |
| Bourgogne-Franche-Comté | Other genotypes | Opportunistic | 2020 | (59,66] | 88 | 7.95 | 6.23 | 5.68 | 2.27 | 11.36 | 6.23 | 6.22 | 5.65 | 6.84 |
| Bretagne | Other genotypes | Opportunistic | 2020 | (59,66] | 65 | 4.62 | 5.57 | 6.15 | 0.00 | 12.31 | 5.67 | 5.64 | 5.01 | 6.42 |
| Centre-Val de Loire | Other genotypes | Opportunistic | 2020 | (59,66] | 500 | 3.80 | 5.98 | 6.00 | 4.00 | 8.20 | 5.94 | 5.94 | 5.60 | 6.29 |
| Centre-Val de Loire | Other genotypes | Organised | 2020 | (59,66] | 1 | 100.00 | 6.87 | 0.00 | 0.00 | 100.00 | 6.79 | 6.68 | 4.51 | 9.64 |
| Corse | Other genotypes | Opportunistic | 2020 | (59,66] | 56 | 1.79 | 7.61 | 7.14 | 1.79 | 16.07 | 7.65 | 7.59 | 6.23 | 9.39 |
| Grand Est | Other genotypes | Opportunistic | 2020 | (59,66] | 127 | 3.94 | 5.74 | 5.51 | 1.57 | 10.24 | 5.76 | 5.75 | 5.14 | 6.46 |
| Hauts-de-France | Other genotypes | Opportunistic | 2020 | (59,66] | 272 | 6.25 | 5.72 | 5.51 | 3.31 | 8.46 | 5.70 | 5.69 | 5.34 | 6.05 |

**Table S8 Observed HR HPV cervical infection prevalence, posterior predictive HR HPV cervical infection prevalence, and posterior expected HR HPV cervical infection prevalence, stratified by various dimensions.** ***(continued)***

| | | | | | | | Posterior distribution | | | | | | | |
|---|---|---|---|---|---|---|---|---|---|---|---|---|---|---|
| | | | | | | | Predictive prevalence | | | | Expected prevalence | | | |
| | | | | | | | | | ETI95% | | | | ETI95% | |
| Region | Genotypes | Type of screening | Year | Age | Number of tests | Observed | Average | Median | LB | UB | Average | Median | LB | UB |
| Hauts-de-France | Other genotypes | Organised | 2020 | (59,66] | 4 | 0.00 | 5.70 | 0.00 | 0.00 | 25.00 | 5.44 | 5.41 | 4.20 | 6.86 |
| Normandie | Other genotypes | Opportunistic | 2020 | (59,66] | 262 | 4.58 | 5.81 | 5.73 | 3.05 | 8.78 | 5.88 | 5.87 | 5.48 | 6.29 |
| Nouvelle-Aquitaine | Other genotypes | Opportunistic | 2020 | (59,66] | 266 | 7.14 | 6.49 | 6.39 | 3.76 | 9.77 | 6.48 | 6.48 | 6.09 | 6.90 |
| Nouvelle-Aquitaine | Other genotypes | Organised | 2020 | (59,66] | 1 | 0.00 | 6.10 | 0.00 | 0.00 | 100.00 | 5.80 | 5.65 | 3.58 | 8.90 |
| Occitanie | Other genotypes | Opportunistic | 2020 | (59,66] | 287 | 6.27 | 7.80 | 7.67 | 4.88 | 10.80 | 7.79 | 7.78 | 7.17 | 8.43 |
| Occitanie | Other genotypes | Organised | 2020 | (59,66] | 1 | 0.00 | 6.50 | 0.00 | 0.00 | 100.00 | 6.79 | 6.69 | 4.78 | 9.28 |
| Pays de la Loire | Other genotypes | Opportunistic | 2020 | (59,66] | 142 | 3.52 | 5.43 | 5.63 | 2.11 | 9.86 | 5.44 | 5.43 | 4.93 | 6.03 |
| Provence-Alpes-Côte d'Azur | Other genotypes | Opportunistic | 2020 | (59,66] | 207 | 5.31 | 6.40 | 6.28 | 3.38 | 9.66 | 6.39 | 6.39 | 5.98 | 6.79 |
| Provence-Alpes-Côte d'Azur | Other genotypes | Organised | 2020 | (59,66] | 2 | 0.00 | 5.45 | 0.00 | 0.00 | 50.00 | 5.75 | 5.66 | 4.15 | 7.76 |
| Île-de-France | Other genotypes | Opportunistic | 2020 | (59,66] | 4,705 | 6.18 | 6.58 | 6.57 | 5.82 | 7.35 | 6.57 | 6.57 | 6.30 | 6.86 |
| Île-de-France | Other genotypes | Organised | 2020 | (59,66] | 4 | 0.00 | 5.49 | 0.00 | 0.00 | 25.00 | 5.44 | 5.41 | 4.28 | 6.81 |
| Auvergne-Rhône-Alpes | Other genotypes | Opportunistic | 2021 | [30,39] | | | 12.49 | 12.47 | 10.46 | 14.56 | 12.49 | 12.48 | 11.62 | 13.40 |
| Auvergne-Rhône-Alpes | Other genotypes | Organised | 2021 | [30,39] | | | 9.79 | 8.57 | 0.00 | 20.00 | 9.79 | 9.76 | 8.28 | 11.37 |
| Bourgogne-Franche-Comté | Other genotypes | Opportunistic | 2021 | [30,39] | | | 11.14 | 11.15 | 8.58 | 13.89 | 11.15 | 11.14 | 10.29 | 12.09 |
| Bourgogne-Franche-Comté | Other genotypes | Organised | 2021 | [30,39] | | | 8.57 | 0.00 | 0.00 | 33.33 | 8.89 | 8.81 | 7.00 | 11.23 |
| Bretagne | Other genotypes | Opportunistic | 2021 | [30,39] | | | 10.63 | 10.60 | 8.44 | 12.88 | 10.60 | 10.59 | 9.88 | 11.36 |
| Bretagne | Other genotypes | Organised | 2021 | [30,39] | | | 6.63 | 7.32 | 0.00 | 14.63 | 6.66 | 6.60 | 5.40 | 8.12 |
| Centre-Val de Loire | Other genotypes | Opportunistic | 2021 | [30,39] | | | 10.78 | 10.77 | 9.78 | 11.85 | 10.77 | 10.77 | 10.38 | 11.18 |
| Centre-Val de Loire | Other genotypes | Organised | 2021 | [30,39] | | | 7.99 | 7.87 | 4.63 | 12.04 | 7.97 | 7.96 | 7.03 | 9.03 |
| Corse | Other genotypes | Opportunistic | 2021 | [30,39] | | | 12.13 | 7.69 | 0.00 | 30.77 | 12.30 | 12.23 | 10.20 | 14.78 |
| Grand Est | Other genotypes | Opportunistic | 2021 | [30,39] | | | 10.57 | 10.54 | 8.77 | 12.40 | 10.58 | 10.58 | 10.00 | 11.22 |
| Grand Est | Other genotypes | Organised | 2021 | [30,39] | | | 8.59 | 0.00 | 0.00 | 40.00 | 8.37 | 8.33 | 6.78 | 10.24 |
| Hauts-de-France | Other genotypes | Opportunistic | 2021 | [30,39] | | | 10.46 | 10.45 | 9.54 | 11.43 | 10.46 | 10.46 | 10.08 | 10.87 |
| Hauts-de-France | Other genotypes | Organised | 2021 | [30,39] | | | 7.92 | 8.15 | 3.70 | 12.59 | 7.92 | 7.91 | 6.95 | 9.00 |
| Normandie | Other genotypes | Opportunistic | 2021 | [30,39] | | | 11.24 | 11.23 | 10.05 | 12.45 | 11.23 | 11.24 | 10.74 | 11.74 |
| Normandie | Other genotypes | Organised | 2021 | [30,39] | | | 7.52 | 0.00 | 0.00 | 33.33 | 7.84 | 7.80 | 6.39 | 9.51 |
| Nouvelle-Aquitaine | Other genotypes | Opportunistic | 2021 | [30,39] | | | 11.30 | 11.29 | 10.10 | 12.54 | 11.28 | 11.28 | 10.85 | 11.74 |
| Nouvelle-Aquitaine | Other genotypes | Organised | 2021 | [30,39] | | | 9.94 | 9.09 | 0.00 | 21.21 | 10.12 | 10.09 | 8.63 | 11.80 |
| Occitanie | Other genotypes | Opportunistic | 2021 | [30,39] | | | 12.57 | 12.58 | 10.92 | 14.25 | 12.58 | 12.57 | 11.95 | 13.23 |
| Occitanie | Other genotypes | Organised | 2021 | [30,39] | | | 10.14 | 9.62 | 1.92 | 19.23 | 10.12 | 10.09 | 8.77 | 11.62 |
| Pays de la Loire | Other genotypes | Opportunistic | 2021 | [30,39] | | | 9.10 | 9.07 | 7.07 | 11.18 | 9.11 | 9.11 | 8.42 | 9.86 |
| Pays de la Loire | Other genotypes | Organised | 2021 | [30,39] | | | 6.29 | 5.45 | 0.00 | 12.73 | 6.47 | 6.46 | 5.43 | 7.61 |
| Provence-Alpes-Côte d'Azur | Other genotypes | Opportunistic | 2021 | [30,39] | | | 11.96 | 11.94 | 10.92 | 13.06 | 11.96 | 11.96 | 11.51 | 12.44 |
| Provence-Alpes-Côte d'Azur | Other genotypes | Organised | 2021 | [30,39] | | | 10.74 | 10.00 | 0.00 | 25.00 | 10.62 | 10.59 | 8.96 | 12.51 |
| Île-de-France | Other genotypes | Opportunistic | 2021 | [30,39] | | | 11.92 | 11.92 | 11.47 | 12.38 | 11.93 | 11.93 | 11.69 | 12.18 |
| Île-de-France | Other genotypes | Organised | 2021 | [30,39] | | | 9.68 | 9.70 | 6.47 | 13.21 | 9.69 | 9.66 | 8.42 | 11.08 |
| Auvergne-Rhône-Alpes | Other genotypes | Opportunistic | 2021 | (39,49] | 804 | 7.96 | 8.68 | 8.71 | 6.59 | 10.82 | 8.68 | 8.67 | 8.03 | 9.36 |
| Auvergne-Rhône-Alpes | Other genotypes | Organised | 2021 | (39,49] | 48 | 10.42 | 7.19 | 6.25 | 0.00 | 14.58 | 7.23 | 7.22 | 6.09 | 8.43 |
| Bourgogne-Franche-Comté | Other genotypes | Opportunistic | 2021 | (39,49] | 391 | 9.46 | 7.82 | 7.67 | 5.12 | 10.74 | 7.79 | 7.78 | 7.15 | 8.47 |

**Table S8 Observed HR HPV cervical infection prevalence, posterior predictive HR HPV cervical infection prevalence, and posterior expected HR HPV cervical infection prevalence, stratified by various dimensions.** ***(continued)***

| | | | | | | | Posterior distribution | | | | | | | |
|---|---|---|---|---|---|---|---|---|---|---|---|---|---|---|
| | | | | | | | Predictive prevalence | | | | Expected prevalence | | | |
| | | | | | | | | | ETI95% | | | | ETI95% | |
| Region | Genotypes | Type of screening | Year | Age | Number of tests | Observed | Average | Median | LB | UB | Average | Median | LB | UB |
| Bourgogne-Franche-Comté | Other genotypes | Organised | 2021 | (39,49] | 1 | 0.00 | 4.93 | 0.00 | 0.00 | 100.00 | 5.90 | 5.83 | 4.33 | 7.84 |
| Bretagne | Other genotypes | Opportunistic | 2021 | (39,49] | 588 | 8.84 | 7.34 | 7.31 | 5.27 | 9.52 | 7.31 | 7.31 | 6.76 | 7.87 |
| Bretagne | Other genotypes | Organised | 2021 | (39,49] | 16 | 0.00 | 4.55 | 6.25 | 0.00 | 18.75 | 4.60 | 4.58 | 3.68 | 5.59 |
| Centre-Val de Loire | Other genotypes | Opportunistic | 2021 | (39,49] | 4,201 | 7.28 | 7.55 | 7.55 | 6.69 | 8.43 | 7.55 | 7.55 | 7.25 | 7.85 |
| Centre-Val de Loire | Other genotypes | Organised | 2021 | (39,49] | 138 | 2.17 | 5.97 | 5.80 | 2.17 | 10.14 | 5.96 | 5.96 | 5.22 | 6.75 |
| Corse | Other genotypes | Opportunistic | 2021 | (39,49] | 5 | 0.00 | 8.41 | 0.00 | 0.00 | 40.00 | 8.56 | 8.53 | 6.83 | 10.46 |
| Grand Est | Other genotypes | Opportunistic | 2021 | (39,49] | 1,093 | 7.14 | 7.52 | 7.50 | 5.86 | 9.24 | 7.52 | 7.52 | 7.08 | 8.00 |
| Grand Est | Other genotypes | Organised | 2021 | (39,49] | 26 | 3.85 | 5.61 | 3.85 | 0.00 | 15.38 | 5.64 | 5.63 | 4.78 | 6.58 |
| Hauts-de-France | Other genotypes | Opportunistic | 2021 | (39,49] | 2,916 | 7.82 | 7.39 | 7.37 | 6.41 | 8.44 | 7.39 | 7.39 | 7.10 | 7.69 |
| Hauts-de-France | Other genotypes | Organised | 2021 | (39,49] | 110 | 1.82 | 5.97 | 5.45 | 1.82 | 10.91 | 5.93 | 5.92 | 5.17 | 6.77 |
| Normandie | Other genotypes | Opportunistic | 2021 | (39,49] | 2,310 | 7.92 | 7.90 | 7.88 | 6.80 | 9.09 | 7.90 | 7.90 | 7.54 | 8.27 |
| Normandie | Other genotypes | Organised | 2021 | (39,49] | 5 | 20.00 | 6.05 | 0.00 | 0.00 | 40.00 | 6.23 | 6.21 | 5.07 | 7.53 |
| Nouvelle-Aquitaine | Other genotypes | Opportunistic | 2021 | (39,49] | 2,359 | 7.97 | 7.85 | 7.84 | 6.74 | 9.03 | 7.86 | 7.86 | 7.55 | 8.19 |
| Nouvelle-Aquitaine | Other genotypes | Organised | 2021 | (39,49] | 3 | 0.00 | 5.99 | 0.00 | 0.00 | 33.33 | 6.19 | 6.14 | 4.93 | 7.68 |
| Occitanie | Other genotypes | Opportunistic | 2021 | (39,49] | 1,638 | 10.32 | 8.98 | 8.97 | 7.51 | 10.50 | 8.97 | 8.97 | 8.49 | 9.47 |
| Occitanie | Other genotypes | Organised | 2021 | (39,49] | 110 | 6.36 | 7.34 | 7.27 | 2.73 | 12.73 | 7.37 | 7.35 | 6.34 | 8.52 |
| Pays de la Loire | Other genotypes | Opportunistic | 2021 | (39,49] | 834 | 6.71 | 6.58 | 6.59 | 4.92 | 8.39 | 6.57 | 6.57 | 6.08 | 7.10 |
| Pays de la Loire | Other genotypes | Organised | 2021 | (39,49] | 85 | 3.53 | 5.15 | 4.71 | 1.18 | 10.59 | 5.09 | 5.07 | 4.30 | 5.96 |
| Provence-Alpes-Côte d'Azur | Other genotypes | Opportunistic | 2021 | (39,49] | 3,170 | 8.30 | 8.34 | 8.36 | 7.38 | 9.37 | 8.34 | 8.33 | 7.97 | 8.71 |
| Provence-Alpes-Côte d'Azur | Other genotypes | Organised | 2021 | (39,49] | 7 | 14.29 | 7.61 | 0.00 | 0.00 | 28.57 | 7.49 | 7.46 | 6.26 | 8.95 |
| Île-de-France | Other genotypes | Opportunistic | 2021 | (39,49] | 25,340 | 8.13 | 8.34 | 8.34 | 7.96 | 8.73 | 8.34 | 8.34 | 8.14 | 8.55 |
| Île-de-France | Other genotypes | Organised | 2021 | (39,49] | 83 | 1.20 | 6.53 | 6.02 | 1.20 | 12.05 | 6.62 | 6.61 | 5.83 | 7.44 |
| Auvergne-Rhône-Alpes | Other genotypes | Opportunistic | 2021 | (49,59] | 666 | 6.91 | 7.39 | 7.36 | 5.26 | 9.46 | 7.36 | 7.35 | 6.81 | 7.94 |
| Auvergne-Rhône-Alpes | Other genotypes | Organised | 2021 | (49,59] | 38 | 7.89 | 6.42 | 5.26 | 0.00 | 15.79 | 6.68 | 6.67 | 5.62 | 7.83 |
| Bourgogne-Franche-Comté | Other genotypes | Opportunistic | 2021 | (49,59] | 329 | 6.69 | 6.70 | 6.69 | 3.95 | 9.42 | 6.68 | 6.67 | 6.14 | 7.25 |
| Bourgogne-Franche-Comté | Other genotypes | Organised | 2021 | (49,59] | 1 | 0.00 | 4.77 | 0.00 | 0.00 | 100.00 | 5.37 | 5.31 | 3.99 | 7.04 |
| Bretagne | Other genotypes | Opportunistic | 2021 | (49,59] | 556 | 3.96 | 6.07 | 6.12 | 3.96 | 8.09 | 6.05 | 6.05 | 5.58 | 6.52 |
| Bretagne | Other genotypes | Organised | 2021 | (49,59] | 36 | 2.78 | 4.37 | 2.78 | 0.00 | 11.11 | 4.34 | 4.32 | 3.54 | 5.23 |
| Centre-Val de Loire | Other genotypes | Opportunistic | 2021 | (49,59] | 3,844 | 5.93 | 6.39 | 6.37 | 5.59 | 7.21 | 6.38 | 6.39 | 6.11 | 6.66 |
| Centre-Val de Loire | Other genotypes | Organised | 2021 | (49,59] | 224 | 5.80 | 5.41 | 5.36 | 2.68 | 8.49 | 5.38 | 5.38 | 4.77 | 6.03 |
| Corse | Other genotypes | Opportunistic | 2021 | (49,59] | 9 | 0.00 | 6.79 | 0.00 | 0.00 | 22.22 | 6.90 | 6.89 | 5.96 | 7.93 |
| Corse | Other genotypes | Organised | 2021 | (49,59] | 1 | 0.00 | 6.40 | 0.00 | 0.00 | 100.00 | 6.17 | 6.05 | 4.07 | 8.97 |
| Grand Est | Other genotypes | Opportunistic | 2021 | (49,59] | 1,003 | 5.08 | 6.42 | 6.38 | 4.99 | 7.98 | 6.42 | 6.41 | 6.03 | 6.82 |
| Grand Est | Other genotypes | Organised | 2021 | (49,59] | 59 | 1.69 | 5.23 | 5.08 | 0.00 | 11.86 | 5.21 | 5.21 | 4.40 | 6.07 |
| Hauts-de-France | Other genotypes | Opportunistic | 2021 | (49,59] | 2,015 | 6.50 | 6.25 | 6.25 | 5.26 | 7.39 | 6.27 | 6.27 | 6.01 | 6.55 |
| Hauts-de-France | Other genotypes | Organised | 2021 | (49,59] | 96 | 5.21 | 5.42 | 5.21 | 1.04 | 10.42 | 5.43 | 5.43 | 4.80 | 6.12 |
| Normandie | Other genotypes | Opportunistic | 2021 | (49,59] | 1,829 | 5.36 | 6.61 | 6.56 | 5.47 | 7.76 | 6.60 | 6.60 | 6.29 | 6.93 |
| Normandie | Other genotypes | Organised | 2021 | (49,59] | 5 | 0.00 | 5.33 | 0.00 | 0.00 | 20.50 | 5.46 | 5.44 | 4.45 | 6.62 |
| Nouvelle-Aquitaine | Other genotypes | Opportunistic | 2021 | (49,59] | 1,995 | 6.47 | 6.65 | 6.62 | 5.56 | 7.82 | 6.65 | 6.65 | 6.38 | 6.95 |

**Table S8** **Observed HR HPV cervical infection prevalence, posterior predictive HR HPV cervical infection prevalence, and posterior expected HR HPV cervical infection prevalence, stratified by various dimensions.** ***(continued)***

| | | | | | | | Posterior distribution | | | | | | | |
|---|---|---|---|---|---|---|---|---|---|---|---|---|---|---|
| | | | | | | | Predictive prevalence | | | | Expected prevalence | | | |
| | | | | | | | | | ETI95% | | | | ETI95% | |
| Region | Genotypes | Type of screening | Year | Age | Number of tests | Observed | Average | Median | LB | UB | Average | Median | LB | UB |
| Nouvelle-Aquitaine | Other genotypes | Organised | 2021 | (49,59] | 170 | 6.47 | 5.88 | 5.88 | 2.35 | 10.00 | 5.84 | 5.82 | 5.16 | 6.56 |
| Occitanie | Other genotypes | Opportunistic | 2021 | (49,59] | 1,372 | 7.94 | 7.72 | 7.65 | 6.34 | 9.26 | 7.72 | 7.71 | 7.29 | 8.16 |
| Occitanie | Other genotypes | Organised | 2021 | (49,59] | 95 | 4.21 | 7.17 | 7.37 | 2.11 | 12.63 | 7.28 | 7.25 | 6.20 | 8.45 |
| Pays de la Loire | Other genotypes | Opportunistic | 2021 | (49,59] | 703 | 5.41 | 5.60 | 5.55 | 3.98 | 7.40 | 5.60 | 5.58 | 5.17 | 6.07 |
| Pays de la Loire | Other genotypes | Organised | 2021 | (49,59] | 78 | 6.41 | 4.85 | 5.13 | 0.00 | 10.26 | 4.84 | 4.83 | 4.11 | 5.63 |
| Provence-Alpes-Côte d'Azur | Other genotypes | Opportunistic | 2021 | (49,59] | 2,577 | 7.37 | 7.00 | 7.02 | 5.98 | 8.07 | 7.01 | 7.00 | 6.70 | 7.34 |
| Provence-Alpes-Côte d'Azur | Other genotypes | Organised | 2021 | (49,59] | 7 | 14.29 | 6.50 | 0.00 | 0.00 | 28.57 | 6.68 | 6.64 | 5.46 | 8.13 |
| Île-de-France | Other genotypes | Opportunistic | 2021 | (49,59] | 18,998 | 7.08 | 7.06 | 7.06 | 6.67 | 7.48 | 7.06 | 7.06 | 6.87 | 7.27 |
| Île-de-France | Other genotypes | Organised | 2021 | (49,59] | 161 | 7.45 | 5.90 | 5.59 | 2.48 | 9.94 | 5.93 | 5.92 | 5.30 | 6.66 |
| Auvergne-Rhône-Alpes | Other genotypes | Opportunistic | 2021 | (59,66] | 330 | 5.76 | 6.47 | 6.36 | 3.94 | 9.09 | 6.50 | 6.49 | 5.98 | 7.04 |
| Auvergne-Rhône-Alpes | Other genotypes | Organised | 2021 | (59,66] | 31 | 3.23 | 6.38 | 6.45 | 0.00 | 16.13 | 6.42 | 6.41 | 5.43 | 7.53 |
| Bourgogne-Franche-Comté | Other genotypes | Opportunistic | 2021 | (59,66] | 207 | 7.25 | 6.01 | 5.80 | 2.90 | 9.66 | 6.02 | 6.02 | 5.49 | 6.57 |
| Bourgogne-Franche-Comté | Other genotypes | Organised | 2021 | (59,66] | 22 | 13.64 | 5.36 | 4.55 | 0.00 | 18.18 | 5.51 | 5.49 | 4.60 | 6.55 |
| Bretagne | Other genotypes | Opportunistic | 2021 | (59,66] | 239 | 4.18 | 5.45 | 5.44 | 2.93 | 8.79 | 5.44 | 5.44 | 4.99 | 5.92 |
| Bretagne | Other genotypes | Organised | 2021 | (59,66] | 21 | 4.76 | 4.51 | 4.76 | 0.00 | 14.29 | 4.48 | 4.44 | 3.59 | 5.56 |
| Centre-Val de Loire | Other genotypes | Opportunistic | 2021 | (59,66] | 2,018 | 6.19 | 5.67 | 5.65 | 4.66 | 6.79 | 5.68 | 5.68 | 5.38 | 5.99 |
| Centre-Val de Loire | Other genotypes | Organised | 2021 | (59,66] | 143 | 4.20 | 5.19 | 4.90 | 2.10 | 9.09 | 5.16 | 5.14 | 4.51 | 5.88 |
| Corse | Other genotypes | Opportunistic | 2021 | (59,66] | 7 | 0.00 | 6.80 | 0.00 | 0.00 | 28.57 | 6.66 | 6.61 | 5.48 | 8.06 |
| Grand Est | Other genotypes | Opportunistic | 2021 | (59,66] | 562 | 3.38 | 5.75 | 5.69 | 3.91 | 7.83 | 5.75 | 5.74 | 5.37 | 6.14 |
| Grand Est | Other genotypes | Organised | 2021 | (59,66] | 46 | 2.17 | 5.02 | 4.35 | 0.00 | 13.04 | 5.05 | 5.05 | 4.21 | 5.93 |
| Hauts-de-France | Other genotypes | Opportunistic | 2021 | (59,66] | 931 | 4.08 | 5.57 | 5.59 | 4.19 | 7.09 | 5.56 | 5.56 | 5.27 | 5.86 |
| Hauts-de-France | Other genotypes | Organised | 2021 | (59,66] | 49 | 6.12 | 5.48 | 6.12 | 0.00 | 12.24 | 5.39 | 5.38 | 4.63 | 6.19 |
| Normandie | Other genotypes | Opportunistic | 2021 | (59,66] | 950 | 5.68 | 5.88 | 5.89 | 4.32 | 7.47 | 5.88 | 5.89 | 5.55 | 6.23 |
| Normandie | Other genotypes | Organised | 2021 | (59,66] | 18 | 11.11 | 5.54 | 5.56 | 0.00 | 16.67 | 5.47 | 5.46 | 4.64 | 6.39 |
| Nouvelle-Aquitaine | Other genotypes | Opportunistic | 2021 | (59,66] | 1,222 | 4.58 | 6.00 | 5.97 | 4.66 | 7.36 | 6.01 | 6.01 | 5.70 | 6.34 |
| Nouvelle-Aquitaine | Other genotypes | Organised | 2021 | (59,66] | 185 | 6.49 | 5.65 | 5.41 | 2.70 | 9.19 | 5.72 | 5.71 | 4.96 | 6.49 |
| Occitanie | Other genotypes | Opportunistic | 2021 | (59,66] | 720 | 7.64 | 6.83 | 6.81 | 5.00 | 8.75 | 6.87 | 6.87 | 6.44 | 7.32 |
| Occitanie | Other genotypes | Organised | 2021 | (59,66] | 48 | 12.50 | 6.93 | 6.25 | 0.00 | 14.58 | 6.99 | 6.95 | 5.97 | 8.17 |
| Pays de la Loire | Other genotypes | Opportunistic | 2021 | (59,66] | 326 | 3.37 | 5.25 | 5.21 | 2.76 | 7.98 | 5.25 | 5.24 | 4.81 | 5.73 |
| Pays de la Loire | Other genotypes | Organised | 2021 | (59,66] | 52 | 1.92 | 4.85 | 3.85 | 0.00 | 11.54 | 4.81 | 4.80 | 4.02 | 5.72 |
| Provence-Alpes-Côte d'Azur | Other genotypes | Opportunistic | 2021 | (59,66] | 1,314 | 7.15 | 6.26 | 6.24 | 4.95 | 7.53 | 6.25 | 6.25 | 5.91 | 6.59 |
| Provence-Alpes-Côte d'Azur | Other genotypes | Organised | 2021 | (59,66] | 22 | 4.55 | 6.03 | 4.55 | 0.00 | 18.18 | 6.03 | 6.00 | 4.99 | 7.20 |
| Île-de-France | Other genotypes | Opportunistic | 2021 | (59,66] | 8,881 | 6.49 | 6.32 | 6.32 | 5.78 | 6.88 | 6.31 | 6.31 | 6.07 | 6.57 |
| Île-de-France | Other genotypes | Organised | 2021 | (59,66] | 1,384 | 5.78 | 5.77 | 5.78 | 4.34 | 7.37 | 5.76 | 5.74 | 5.01 | 6.60 |
| Auvergne-Rhône-Alpes | Other genotypes | Opportunistic | 2022 | [30,39] | | | 13.83 | 14.02 | 7.48 | 20.56 | 13.89 | 13.88 | 12.74 | 15.08 |
| Auvergne-Rhône-Alpes | Other genotypes | Organised | 2022 | [30,39] | | | 10.02 | 8.33 | 0.00 | 25.00 | 9.90 | 9.88 | 8.34 | 11.61 |
| Bourgogne-Franche-Comté | Other genotypes | Opportunistic | 2022 | [30,39] | | | 11.85 | 11.80 | 8.90 | 15.11 | 11.86 | 11.85 | 10.91 | 12.89 |
| Bourgogne-Franche-Comté | Other genotypes | Organised | 2022 | [30,39] | | | 9.33 | 7.69 | 0.00 | 23.08 | 9.41 | 9.36 | 7.81 | 11.20 |
| Bretagne | Other genotypes | Opportunistic | 2022 | [30,39] | | | 11.88 | 11.89 | 9.81 | 14.07 | 11.85 | 11.85 | 11.07 | 12.64 |

**Table S8** **Observed HR HPV cervical infection prevalence, posterior predictive HR HPV cervical infection prevalence, and posterior expected HR HPV cervical infection prevalence, stratified by various dimensions.** ***(continued)***

| | | | | | | | Posterior distribution | | | | | | | |
|---|---|---|---|---|---|---|---|---|---|---|---|---|---|---|
| | | | | | | | Predictive prevalence | | | | Expected prevalence | | | |
| | | | | | | | | | ETI95% | | | | ETI95% | |
| Region | Genotypes | Type of screening | Year | Age | Number of tests | Observed | Average | Median | LB | UB | Average | Median | LB | UB |
| Bretagne | Other genotypes | Organised | 2022 | [30,39] | | | 7.72 | 7.64 | 3.82 | 12.12 | 7.71 | 7.67 | 6.49 | 9.15 |
| Centre-Val de Loire | Other genotypes | Opportunistic | 2022 | [30,39] | | | 11.25 | 11.23 | 9.91 | 12.64 | 11.23 | 11.23 | 10.77 | 11.70 |
| Centre-Val de Loire | Other genotypes | Organised | 2022 | [30,39] | | | 8.16 | 8.33 | 4.41 | 12.25 | 8.12 | 8.10 | 7.29 | 9.04 |
| Corse | Other genotypes | Opportunistic | 2022 | [30,39] | | | 12.17 | 0.00 | 0.00 | 50.00 | 11.67 | 11.61 | 9.76 | 13.89 |
| Grand Est | Other genotypes | Opportunistic | 2022 | [30,39] | | | 11.44 | 11.44 | 9.52 | 13.45 | 11.42 | 11.41 | 10.77 | 12.12 |
| Grand Est | Other genotypes | Organised | 2022 | [30,39] | | | 7.98 | 8.11 | 4.50 | 11.71 | 7.97 | 7.95 | 6.90 | 9.05 |
| Hauts-de-France | Other genotypes | Opportunistic | 2022 | [30,39] | | | 11.14 | 11.16 | 10.07 | 12.20 | 11.13 | 11.13 | 10.71 | 11.56 |
| Hauts-de-France | Other genotypes | Organised | 2022 | [30,39] | | | 8.14 | 8.10 | 5.31 | 11.17 | 8.15 | 8.13 | 7.23 | 9.10 |
| Normandie | Other genotypes | Opportunistic | 2022 | [30,39] | | | 12.33 | 12.31 | 10.86 | 13.85 | 12.31 | 12.31 | 11.75 | 12.87 |
| Normandie | Other genotypes | Organised | 2022 | [30,39] | | | 8.72 | 9.09 | 0.00 | 22.73 | 8.83 | 8.79 | 7.59 | 10.21 |
| Nouvelle-Aquitaine | Other genotypes | Opportunistic | 2022 | [30,39] | | | 12.03 | 12.01 | 10.82 | 13.25 | 12.04 | 12.03 | 11.60 | 12.51 |
| Nouvelle-Aquitaine | Other genotypes | Organised | 2022 | [30,39] | | | 9.45 | 9.29 | 6.09 | 12.82 | 9.44 | 9.42 | 8.48 | 10.48 |
| Occitanie | Other genotypes | Opportunistic | 2022 | [30,39] | | | 12.92 | 12.82 | 10.62 | 15.38 | 12.91 | 12.90 | 12.19 | 13.66 |
| Occitanie | Other genotypes | Organised | 2022 | [30,39] | | | 10.15 | 9.86 | 5.63 | 15.49 | 10.11 | 10.09 | 8.95 | 11.39 |
| Pays de la Loire | Other genotypes | Opportunistic | 2022 | [30,39] | | | 10.59 | 10.59 | 7.06 | 14.51 | 10.60 | 10.58 | 9.77 | 11.47 |
| Pays de la Loire | Other genotypes | Organised | 2022 | [30,39] | | | 7.20 | 6.35 | 1.59 | 14.29 | 7.15 | 7.13 | 6.18 | 8.17 |
| Provence-Alpes-Côte d'Azur | Other genotypes | Opportunistic | 2022 | [30,39] | | | 12.81 | 12.82 | 11.50 | 14.09 | 12.81 | 12.81 | 12.31 | 13.35 |
| Provence-Alpes-Côte d'Azur | Other genotypes | Organised | 2022 | [30,39] | | | 11.27 | 11.20 | 7.47 | 15.77 | 11.33 | 11.27 | 9.88 | 13.08 |
| Île-de-France | Other genotypes | Opportunistic | 2022 | [30,39] | | | 12.96 | 12.96 | 12.08 | 13.85 | 12.96 | 12.96 | 12.57 | 13.34 |
| Île-de-France | Other genotypes | Organised | 2022 | [30,39] | | | 9.93 | 9.91 | 6.50 | 13.62 | 9.95 | 9.94 | 8.92 | 11.04 |
| Auvergne-Rhône-Alpes | Other genotypes | Opportunistic | 2022 | (39,49] | 64 | 17.19 | 10.04 | 9.38 | 3.12 | 18.75 | 10.11 | 10.09 | 8.95 | 11.36 |
| Auvergne-Rhône-Alpes | Other genotypes | Organised | 2022 | (39,49] | 15 | 13.33 | 7.92 | 6.67 | 0.00 | 26.67 | 8.01 | 7.98 | 6.71 | 9.44 |
| Bourgogne-Franche-Comté | Other genotypes | Opportunistic | 2022 | (39,49] | 333 | 8.71 | 8.18 | 8.11 | 5.11 | 11.41 | 8.18 | 8.17 | 7.49 | 8.90 |
| Bourgogne-Franche-Comté | Other genotypes | Organised | 2022 | (39,49] | 1 | 0.00 | 6.63 | 0.00 | 0.00 | 100.00 | 6.45 | 6.38 | 4.66 | 8.65 |
| Bretagne | Other genotypes | Opportunistic | 2022 | (39,49] | 663 | 7.69 | 8.33 | 8.30 | 6.18 | 10.56 | 8.33 | 8.32 | 7.75 | 8.92 |
| Bretagne | Other genotypes | Organised | 2022 | (39,49] | 173 | 5.78 | 5.40 | 5.20 | 2.31 | 9.25 | 5.44 | 5.43 | 4.62 | 6.37 |
| Centre-Val de Loire | Other genotypes | Opportunistic | 2022 | (39,49] | 2,079 | 8.37 | 7.86 | 7.84 | 6.69 | 9.09 | 7.86 | 7.86 | 7.51 | 8.22 |
| Centre-Val de Loire | Other genotypes | Organised | 2022 | (39,49] | 248 | 6.05 | 6.14 | 6.05 | 3.23 | 9.27 | 6.14 | 6.13 | 5.52 | 6.78 |
| Corse | Other genotypes | Opportunistic | 2022 | (39,49] | 3 | 33.33 | 10.10 | 0.00 | 0.00 | 66.67 | 9.62 | 9.54 | 7.61 | 11.95 |
| Corse | Other genotypes | Organised | 2022 | (39,49] | 2 | 0.00 | 6.48 | 0.00 | 0.00 | 50.00 | 6.92 | 6.82 | 5.15 | 9.08 |
| Grand Est | Other genotypes | Opportunistic | 2022 | (39,49] | 858 | 10.14 | 8.15 | 8.16 | 6.29 | 10.14 | 8.17 | 8.16 | 7.67 | 8.72 |
| Grand Est | Other genotypes | Organised | 2022 | (39,49] | 198 | 8.59 | 6.06 | 6.06 | 3.03 | 9.60 | 6.12 | 6.11 | 5.29 | 6.98 |
| Hauts-de-France | Other genotypes | Opportunistic | 2022 | (39,49] | 2,382 | 8.73 | 7.85 | 7.85 | 6.72 | 9.03 | 7.84 | 7.84 | 7.51 | 8.18 |
| Hauts-de-France | Other genotypes | Organised | 2022 | (39,49] | 286 | 5.94 | 6.21 | 6.29 | 3.50 | 9.44 | 6.18 | 6.17 | 5.53 | 6.86 |
| Normandie | Other genotypes | Opportunistic | 2022 | (39,49] | 1,814 | 8.27 | 8.64 | 8.65 | 7.28 | 10.03 | 8.64 | 8.63 | 8.21 | 9.05 |
| Normandie | Other genotypes | Organised | 2022 | (39,49] | 22 | 13.64 | 6.52 | 4.55 | 0.00 | 18.18 | 6.58 | 6.57 | 5.79 | 7.45 |
| Nouvelle-Aquitaine | Other genotypes | Opportunistic | 2022 | (39,49] | 2,812 | 8.04 | 8.52 | 8.50 | 7.43 | 9.64 | 8.54 | 8.53 | 8.19 | 8.90 |
| Nouvelle-Aquitaine | Other genotypes | Organised | 2022 | (39,49] | 337 | 7.12 | 6.90 | 6.82 | 4.15 | 9.79 | 6.89 | 6.88 | 6.24 | 7.63 |
| Occitanie | Other genotypes | Opportunistic | 2022 | (39,49] | 746 | 8.58 | 9.14 | 9.12 | 7.10 | 11.26 | 9.15 | 9.14 | 8.60 | 9.73 |

**Table S8 Observed HR HPV cervical infection prevalence, posterior predictive HR HPV cervical infection prevalence, and posterior expected HR HPV cervical infection prevalence, stratified by various dimensions.** ***(continued)***

| | | | | | | | Posterior distribution | | | | | | | |
|---|---|---|---|---|---|---|---|---|---|---|---|---|---|---|
| | | | | | | | Predictive prevalence | | | | Expected prevalence | | | |
| | | | | | | | | | ETI95% | | | | ETI95% | |
| Region | Genotypes | Type of screening | Year | Age | Number of tests | Observed | Average | Median | LB | UB | Average | Median | LB | UB |
| Occitanie | Other genotypes | Organised | 2022 | (39,49] | 135 | 5.93 | 7.43 | 7.41 | 3.70 | 11.85 | 7.50 | 7.48 | 6.62 | 8.47 |
| Pays de la Loire | Other genotypes | Opportunistic | 2022 | (39,49] | 203 | 7.88 | 7.51 | 7.39 | 3.94 | 11.33 | 7.53 | 7.52 | 6.91 | 8.20 |
| Pays de la Loire | Other genotypes | Organised | 2022 | (39,49] | 71 | 1.41 | 5.59 | 5.63 | 1.41 | 11.27 | 5.63 | 5.61 | 4.89 | 6.40 |
| Provence-Alpes-Côte d'Azur | Other genotypes | Opportunistic | 2022 | (39,49] | 2,056 | 8.46 | 8.94 | 8.95 | 7.68 | 10.21 | 8.95 | 8.95 | 8.58 | 9.35 |
| Provence-Alpes-Côte d'Azur | Other genotypes | Organised | 2022 | (39,49] | 112 | 5.36 | 8.29 | 8.04 | 3.57 | 14.29 | 8.37 | 8.33 | 7.27 | 9.71 |
| Île-de-France | Other genotypes | Opportunistic | 2022 | (39,49] | 4,399 | 7.96 | 8.80 | 8.80 | 7.91 | 9.68 | 8.80 | 8.80 | 8.51 | 9.10 |
| Île-de-France | Other genotypes | Organised | 2022 | (39,49] | 342 | 9.65 | 7.20 | 7.02 | 4.39 | 10.23 | 7.23 | 7.21 | 6.53 | 8.00 |
| Auvergne-Rhône-Alpes | Other genotypes | Opportunistic | 2022 | (49,59] | 49 | 20.41 | 8.94 | 8.16 | 2.04 | 18.37 | 8.92 | 8.90 | 7.86 | 10.07 |
| Auvergne-Rhône-Alpes | Other genotypes | Organised | 2022 | (49,59] | 13 | 7.69 | 7.86 | 7.69 | 0.00 | 23.08 | 7.79 | 7.75 | 6.31 | 9.49 |
| Bourgogne-Franche-Comté | Other genotypes | Opportunistic | 2022 | (49,59] | 239 | 7.53 | 6.98 | 7.11 | 3.77 | 10.46 | 6.99 | 6.98 | 6.41 | 7.63 |
| Bourgogne-Franche-Comté | Other genotypes | Organised | 2022 | (49,59] | 1 | 100.00 | 6.30 | 0.00 | 0.00 | 100.00 | 6.41 | 6.32 | 4.58 | 8.74 |
| Bretagne | Other genotypes | Opportunistic | 2022 | (49,59] | 535 | 7.85 | 7.04 | 7.10 | 4.86 | 9.35 | 7.06 | 7.05 | 6.54 | 7.59 |
| Bretagne | Other genotypes | Organised | 2022 | (49,59] | 144 | 2.78 | 5.02 | 4.86 | 2.08 | 9.03 | 5.04 | 5.01 | 4.26 | 5.95 |
| Centre-Val de Loire | Other genotypes | Opportunistic | 2022 | (49,59] | 1,838 | 6.58 | 6.66 | 6.64 | 5.55 | 7.83 | 6.66 | 6.66 | 6.34 | 6.98 |
| Centre-Val de Loire | Other genotypes | Organised | 2022 | (49,59] | 278 | 7.19 | 5.73 | 5.76 | 3.24 | 8.63 | 5.75 | 5.75 | 5.18 | 6.37 |
| Corse | Other genotypes | Opportunistic | 2022 | (49,59] | 2 | 0.00 | 8.30 | 0.00 | 0.00 | 50.00 | 7.65 | 7.56 | 5.63 | 10.09 |
| Grand Est | Other genotypes | Opportunistic | 2022 | (49,59] | 767 | 6.13 | 7.00 | 7.04 | 5.22 | 8.87 | 7.04 | 7.03 | 6.57 | 7.53 |
| Grand Est | Other genotypes | Organised | 2022 | (49,59] | 120 | 4.17 | 5.94 | 5.83 | 1.67 | 10.02 | 5.92 | 5.91 | 5.18 | 6.72 |
| Hauts-de-France | Other genotypes | Opportunistic | 2022 | (49,59] | 1,725 | 8.17 | 6.66 | 6.67 | 5.51 | 7.94 | 6.67 | 6.67 | 6.38 | 6.98 |
| Hauts-de-France | Other genotypes | Organised | 2022 | (49,59] | 267 | 7.49 | 5.71 | 5.62 | 3.00 | 8.99 | 5.72 | 5.71 | 5.11 | 6.40 |
| Normandie | Other genotypes | Opportunistic | 2022 | (49,59] | 1,196 | 6.77 | 7.16 | 7.11 | 5.69 | 8.70 | 7.18 | 7.18 | 6.82 | 7.55 |
| Normandie | Other genotypes | Organised | 2022 | (49,59] | 16 | 6.25 | 6.12 | 6.25 | 0.00 | 18.75 | 5.98 | 5.98 | 5.18 | 6.82 |
| Nouvelle-Aquitaine | Other genotypes | Opportunistic | 2022 | (49,59] | 2,090 | 7.32 | 7.25 | 7.22 | 6.17 | 8.47 | 7.25 | 7.25 | 6.93 | 7.59 |
| Nouvelle-Aquitaine | Other genotypes | Organised | 2022 | (49,59] | 644 | 6.21 | 6.31 | 6.21 | 4.35 | 8.39 | 6.30 | 6.29 | 5.71 | 6.96 |
| Occitanie | Other genotypes | Opportunistic | 2022 | (49,59] | 710 | 7.75 | 7.80 | 7.75 | 5.77 | 9.86 | 7.81 | 7.80 | 7.34 | 8.33 |
| Occitanie | Other genotypes | Organised | 2022 | (49,59] | 141 | 7.80 | 7.08 | 7.09 | 2.84 | 11.35 | 7.09 | 7.06 | 6.26 | 8.02 |
| Pays de la Loire | Other genotypes | Opportunistic | 2022 | (49,59] | 158 | 7.59 | 6.57 | 6.33 | 2.53 | 10.76 | 6.55 | 6.55 | 6.00 | 7.17 |
| Pays de la Loire | Other genotypes | Organised | 2022 | (49,59] | 67 | 4.48 | 5.29 | 4.48 | 0.00 | 10.49 | 5.24 | 5.24 | 4.58 | 5.99 |
| Provence-Alpes-Côte d'Azur | Other genotypes | Opportunistic | 2022 | (49,59] | 1,791 | 7.31 | 7.48 | 7.48 | 6.25 | 8.77 | 7.50 | 7.50 | 7.15 | 7.85 |
| Provence-Alpes-Côte d'Azur | Other genotypes | Organised | 2022 | (49,59] | 85 | 8.24 | 7.62 | 7.06 | 2.35 | 14.12 | 7.70 | 7.66 | 6.65 | 9.03 |
| Île-de-France | Other genotypes | Opportunistic | 2022 | (49,59] | 3,353 | 7.52 | 7.38 | 7.37 | 6.47 | 8.35 | 7.38 | 7.38 | 7.10 | 7.65 |
| Île-de-France | Other genotypes | Organised | 2022 | (49,59] | 449 | 7.35 | 6.50 | 6.46 | 4.23 | 8.91 | 6.49 | 6.48 | 5.83 | 7.19 |
| Auvergne-Rhône-Alpes | Other genotypes | Opportunistic | 2022 | (59,66] | 27 | 3.70 | 7.35 | 7.41 | 0.00 | 18.52 | 7.46 | 7.46 | 6.65 | 8.34 |
| Auvergne-Rhône-Alpes | Other genotypes | Organised | 2022 | (59,66] | 8 | 0.00 | 7.75 | 0.00 | 0.00 | 25.00 | 7.74 | 7.69 | 6.09 | 9.70 |
| Bourgogne-Franche-Comté | Other genotypes | Opportunistic | 2022 | (59,66] | 120 | 5.83 | 6.15 | 5.83 | 2.50 | 10.83 | 6.19 | 6.19 | 5.62 | 6.83 |
| Bourgogne-Franche-Comté | Other genotypes | Organised | 2022 | (59,66] | 17 | 5.88 | 5.69 | 5.88 | 0.00 | 17.65 | 5.75 | 5.73 | 4.76 | 6.86 |
| Bretagne | Other genotypes | Opportunistic | 2022 | (59,66] | 289 | 7.96 | 6.18 | 6.23 | 3.46 | 9.00 | 6.21 | 6.20 | 5.71 | 6.71 |
| Bretagne | Other genotypes | Organised | 2022 | (59,66] | 112 | 1.79 | 4.67 | 4.46 | 0.89 | 8.93 | 4.69 | 4.67 | 3.89 | 5.64 |
| Centre-Val de Loire | Other genotypes | Opportunistic | 2022 | (59,66] | 872 | 6.88 | 5.88 | 5.85 | 4.36 | 7.46 | 5.88 | 5.88 | 5.56 | 6.22 |

**Table S8** **Observed HR HPV cervical infection prevalence, posterior predictive HR HPV cervical infection prevalence, and posterior expected HR HPV cervical infection prevalence, stratified by various dimensions.** ***(continued)***

| | | | | | | | Posterior distribution | | | | | | | |
|---|---|---|---|---|---|---|---|---|---|---|---|---|---|---|
| | | | | | | | Predictive prevalence | | | | Expected prevalence | | | |
| | | | | | | | | | ETI95% | | | | ETI95% | |
| Region | Genotypes | Type of screening | Year | Age | Number of tests | Observed | Average | Median | LB | UB | Average | Median | LB | UB |
| Centre-Val de Loire | Other genotypes | Organised | 2022 | (59,66] | 188 | 7.45 | 5.46 | 5.32 | 2.66 | 9.04 | 5.45 | 5.43 | 4.80 | 6.20 |
| Corse | Other genotypes | Opportunistic | 2022 | (59,66] | 2 | 50.00 | 8.23 | 0.00 | 0.00 | 50.00 | 8.93 | 8.79 | 6.33 | 12.22 |
| Grand Est | Other genotypes | Opportunistic | 2022 | (59,66] | 312 | 5.77 | 6.29 | 6.09 | 3.85 | 8.97 | 6.25 | 6.25 | 5.81 | 6.71 |
| Grand Est | Other genotypes | Organised | 2022 | (59,66] | 82 | 4.88 | 5.83 | 6.10 | 1.22 | 10.98 | 5.80 | 5.78 | 4.95 | 6.73 |
| Hauts-de-France | Other genotypes | Opportunistic | 2022 | (59,66] | 714 | 6.16 | 5.99 | 6.02 | 4.34 | 7.84 | 5.99 | 5.99 | 5.68 | 6.32 |
| Hauts-de-France | Other genotypes | Organised | 2022 | (59,66] | 169 | 5.33 | 5.59 | 5.33 | 2.37 | 9.47 | 5.58 | 5.56 | 4.85 | 6.38 |
| Normandie | Other genotypes | Opportunistic | 2022 | (59,66] | 612 | 5.07 | 6.37 | 6.37 | 4.41 | 8.33 | 6.35 | 6.35 | 5.98 | 6.74 |
| Normandie | Other genotypes | Organised | 2022 | (59,66] | 93 | 2.15 | 5.73 | 5.38 | 1.08 | 10.75 | 5.62 | 5.60 | 4.83 | 6.52 |
| Nouvelle-Aquitaine | Other genotypes | Opportunistic | 2022 | (59,66] | 1,040 | 6.06 | 6.45 | 6.44 | 5.00 | 7.98 | 6.45 | 6.45 | 6.10 | 6.80 |
| Nouvelle-Aquitaine | Other genotypes | Organised | 2022 | (59,66] | 440 | 7.27 | 6.07 | 6.14 | 3.86 | 8.64 | 6.08 | 6.06 | 5.36 | 6.89 |
| Occitanie | Other genotypes | Opportunistic | 2022 | (59,66] | 349 | 6.02 | 6.96 | 6.88 | 4.58 | 9.74 | 6.94 | 6.93 | 6.47 | 7.46 |
| Occitanie | Other genotypes | Organised | 2022 | (59,66] | 89 | 5.62 | 6.87 | 6.74 | 2.25 | 12.36 | 6.77 | 6.75 | 5.90 | 7.79 |
| Pays de la Loire | Other genotypes | Opportunistic | 2022 | (59,66] | 105 | 7.62 | 6.00 | 5.71 | 1.90 | 10.48 | 6.03 | 6.02 | 5.44 | 6.66 |
| Pays de la Loire | Other genotypes | Organised | 2022 | (59,66] | 40 | 0.00 | 5.22 | 5.00 | 0.00 | 12.50 | 5.21 | 5.20 | 4.42 | 6.07 |
| Provence-Alpes-Côte d'Azur | Other genotypes | Opportunistic | 2022 | (59,66] | 836 | 6.22 | 6.66 | 6.70 | 4.90 | 8.49 | 6.66 | 6.66 | 6.29 | 7.03 |
| Provence-Alpes-Côte d'Azur | Other genotypes | Organised | 2022 | (59,66] | 66 | 6.06 | 7.15 | 7.58 | 1.52 | 13.64 | 7.13 | 7.08 | 6.03 | 8.37 |
| Île-de-France | Other genotypes | Opportunistic | 2022 | (59,66] | 1,520 | 5.99 | 6.60 | 6.58 | 5.39 | 7.89 | 6.59 | 6.58 | 6.28 | 6.90 |
| Île-de-France | Other genotypes | Organised | 2022 | (59,66] | 193 | 4.15 | 6.24 | 6.22 | 3.11 | 9.84 | 6.23 | 6.21 | 5.52 | 7.04 |
| Auvergne-Rhône-Alpes | Other genotypes | Opportunistic | 2023 | [30,39] | | | 17.58 | 16.90 | 8.45 | 26.76 | 17.57 | 17.57 | 15.51 | 19.69 |
| Auvergne-Rhône-Alpes | Other genotypes | Organised | 2023 | [30,39] | | | 11.80 | 10.53 | 0.00 | 26.32 | 11.85 | 11.84 | 9.79 | 14.15 |
| Bourgogne-Franche-Comté | Other genotypes | Opportunistic | 2023 | [30,39] | | | 13.90 | 13.78 | 9.33 | 19.11 | 13.92 | 13.91 | 12.63 | 15.28 |
| Bourgogne-Franche-Comté | Other genotypes | Organised | 2023 | [30,39] | | | 8.24 | 7.50 | 0.00 | 17.50 | 8.37 | 8.34 | 6.99 | 10.03 |
| Bretagne | Other genotypes | Opportunistic | 2023 | [30,39] | | | 14.22 | 14.19 | 12.11 | 16.34 | 14.22 | 14.21 | 13.35 | 15.13 |
| Bretagne | Other genotypes | Organised | 2023 | [30,39] | | | 7.76 | 7.74 | 5.16 | 10.54 | 7.75 | 7.71 | 6.66 | 9.01 |
| Centre-Val de Loire | Other genotypes | Opportunistic | 2023 | [30,39] | | | 13.14 | 13.13 | 11.61 | 14.71 | 13.16 | 13.16 | 12.63 | 13.74 |
| Centre-Val de Loire | Other genotypes | Organised | 2023 | [30,39] | | | 8.43 | 8.57 | 3.57 | 13.57 | 8.53 | 8.50 | 7.59 | 9.57 |
| Corse | Other genotypes | Opportunistic | 2023 | [30,39] | | | 11.07 | 0.00 | 0.00 | 100.00 | 10.92 | 10.55 | 6.24 | 17.33 |
| Grand Est | Other genotypes | Opportunistic | 2023 | [30,39] | | | 13.33 | 13.31 | 11.09 | 15.62 | 13.32 | 13.31 | 12.55 | 14.13 |
| Grand Est | Other genotypes | Organised | 2023 | [30,39] | | | 8.14 | 7.92 | 2.97 | 13.86 | 8.19 | 8.19 | 7.08 | 9.41 |
| Hauts-de-France | Other genotypes | Opportunistic | 2023 | [30,39] | | | 12.93 | 12.92 | 11.71 | 14.19 | 12.94 | 12.94 | 12.44 | 13.44 |
| Hauts-de-France | Other genotypes | Organised | 2023 | [30,39] | | | 8.37 | 7.91 | 4.32 | 13.67 | 8.38 | 8.37 | 7.35 | 9.51 |
| Normandie | Other genotypes | Opportunistic | 2023 | [30,39] | | | 14.31 | 14.26 | 12.22 | 16.47 | 14.32 | 14.33 | 13.66 | 15.01 |
| Normandie | Other genotypes | Organised | 2023 | [30,39] | | | 8.55 | 7.69 | 0.00 | 17.95 | 8.50 | 8.50 | 7.45 | 9.61 |
| Nouvelle-Aquitaine | Other genotypes | Opportunistic | 2023 | [30,39] | | | 14.27 | 14.28 | 12.87 | 15.69 | 14.26 | 14.26 | 13.72 | 14.83 |
| Nouvelle-Aquitaine | Other genotypes | Organised | 2023 | [30,39] | | | 9.40 | 9.36 | 6.65 | 12.56 | 9.46 | 9.43 | 8.47 | 10.50 |
| Occitanie | Other genotypes | Opportunistic | 2023 | [30,39] | | | 15.33 | 15.14 | 11.71 | 19.43 | 15.30 | 15.27 | 14.36 | 16.34 |
| Occitanie | Other genotypes | Organised | 2023 | [30,39] | | | 10.06 | 8.82 | 0.00 | 20.59 | 10.07 | 10.05 | 8.81 | 11.50 |
| Pays de la Loire | Other genotypes | Opportunistic | 2023 | [30,39] | | | 12.44 | 12.50 | 8.88 | 16.45 | 12.44 | 12.42 | 11.44 | 13.52 |
| Pays de la Loire | Other genotypes | Organised | 2023 | [30,39] | | | 7.72 | 7.80 | 3.55 | 12.77 | 7.79 | 7.78 | 6.76 | 8.93 |

**Table S8** **Observed HR HPV cervical infection prevalence, posterior predictive HR HPV cervical infection prevalence, and posterior expected HR HPV cervical infection prevalence, stratified by various dimensions.** ***(continued)***

| | | | | | | | Posterior distribution | | | | | | | |
|---|---|---|---|---|---|---|---|---|---|---|---|---|---|---|
| | | | | | | | Predictive prevalence | | | | Expected prevalence | | | |
| | | | | | | | | | ETI95% | | | | ETI95% | |
| Region | Genotypes | Type of screening | Year | Age | Number of tests | Observed | Average | Median | LB | UB | Average | Median | LB | UB |
| Provence-Alpes-Côte d'Azur | Other genotypes | Opportunistic | 2023 | [30,39] | | | 15.31 | 15.31 | 13.68 | 16.99 | 15.31 | 15.31 | 14.68 | 15.93 |
| Provence-Alpes-Côte d'Azur | Other genotypes | Organised | 2023 | [30,39] | | | 11.28 | 11.22 | 6.83 | 16.10 | 11.36 | 11.31 | 9.81 | 13.12 |
| Île-de-France | Other genotypes | Opportunistic | 2023 | [30,39] | | | 15.44 | 15.43 | 14.43 | 16.45 | 15.44 | 15.44 | 14.99 | 15.90 |
| Île-de-France | Other genotypes | Organised | 2023 | [30,39] | | | 10.34 | 10.40 | 7.20 | 13.87 | 10.37 | 10.34 | 9.27 | 11.64 |
| Auvergne-Rhône-Alpes | Other genotypes | Opportunistic | 2023 | (39,49] | 48 | 8.33 | 12.81 | 12.50 | 4.17 | 22.92 | 12.60 | 12.58 | 10.91 | 14.43 |
| Auvergne-Rhône-Alpes | Other genotypes | Organised | 2023 | (39,49] | 17 | 5.88 | 8.97 | 5.88 | 0.00 | 23.53 | 9.20 | 9.14 | 7.30 | 11.48 |
| Bourgogne-Franche-Comté | Other genotypes | Opportunistic | 2023 | (39,49] | 162 | 9.88 | 9.65 | 9.26 | 5.56 | 14.81 | 9.59 | 9.58 | 8.60 | 10.61 |
| Bourgogne-Franche-Comté | Other genotypes | Organised | 2023 | (39,49] | 64 | 4.69 | 6.61 | 6.25 | 1.56 | 14.06 | 6.60 | 6.57 | 5.50 | 7.87 |
| Bretagne | Other genotypes | Opportunistic | 2023 | (39,49] | 966 | 11.70 | 9.82 | 9.83 | 7.97 | 11.80 | 9.84 | 9.83 | 9.19 | 10.51 |
| Bretagne | Other genotypes | Organised | 2023 | (39,49] | 406 | 5.67 | 5.73 | 5.67 | 3.45 | 8.37 | 5.76 | 5.75 | 4.95 | 6.63 |
| Centre-Val de Loire | Other genotypes | Opportunistic | 2023 | (39,49] | 1,493 | 10.31 | 9.23 | 9.24 | 7.70 | 10.79 | 9.23 | 9.23 | 8.82 | 9.64 |
| Centre-Val de Loire | Other genotypes | Organised | 2023 | (39,49] | 133 | 8.27 | 6.56 | 6.77 | 2.26 | 11.28 | 6.62 | 6.61 | 5.88 | 7.40 |
| Corse | Other genotypes | Opportunistic | 2023 | (39,49] | 1 | 0.00 | 9.77 | 0.00 | 0.00 | 100.00 | 11.01 | 10.74 | 6.05 | 17.73 |
| Grand Est | Other genotypes | Opportunistic | 2023 | (39,49] | 843 | 10.79 | 9.65 | 9.61 | 7.59 | 11.86 | 9.65 | 9.65 | 9.07 | 10.28 |
| Grand Est | Other genotypes | Organised | 2023 | (39,49] | 101 | 5.94 | 6.40 | 5.94 | 1.98 | 11.88 | 6.46 | 6.44 | 5.59 | 7.42 |
| Hauts-de-France | Other genotypes | Opportunistic | 2023 | (39,49] | 2,152 | 11.01 | 9.22 | 9.20 | 7.95 | 10.50 | 9.23 | 9.23 | 8.83 | 9.62 |
| Hauts-de-France | Other genotypes | Organised | 2023 | (39,49] | 100 | 6.00 | 6.39 | 6.00 | 2.00 | 11.03 | 6.40 | 6.39 | 5.62 | 7.27 |
| Normandie | Other genotypes | Opportunistic | 2023 | (39,49] | 1,088 | 8.82 | 10.08 | 10.11 | 8.27 | 11.95 | 10.11 | 10.10 | 9.58 | 10.63 |
| Normandie | Other genotypes | Organised | 2023 | (39,49] | 42 | 7.14 | 6.78 | 7.14 | 0.00 | 14.29 | 6.84 | 6.84 | 5.98 | 7.78 |
| Nouvelle-Aquitaine | Other genotypes | Opportunistic | 2023 | (39,49] | 2,209 | 10.59 | 10.07 | 10.05 | 8.78 | 11.41 | 10.06 | 10.06 | 9.65 | 10.51 |
| Nouvelle-Aquitaine | Other genotypes | Organised | 2023 | (39,49] | 441 | 9.30 | 7.34 | 7.26 | 4.76 | 10.20 | 7.33 | 7.31 | 6.58 | 8.17 |
| Occitanie | Other genotypes | Opportunistic | 2023 | (39,49] | 353 | 7.08 | 10.96 | 11.05 | 7.93 | 14.45 | 10.96 | 10.94 | 10.18 | 11.82 |
| Occitanie | Other genotypes | Organised | 2023 | (39,49] | 51 | 5.88 | 7.97 | 7.84 | 1.96 | 15.69 | 8.05 | 8.05 | 6.98 | 9.20 |
| Pays de la Loire | Other genotypes | Opportunistic | 2023 | (39,49] | 299 | 7.69 | 8.86 | 8.70 | 5.69 | 12.37 | 8.86 | 8.85 | 8.14 | 9.68 |
| Pays de la Loire | Other genotypes | Organised | 2023 | (39,49] | 131 | 6.11 | 5.82 | 5.34 | 2.29 | 9.92 | 5.74 | 5.72 | 4.98 | 6.57 |
| Provence-Alpes-Côte d'Azur | Other genotypes | Opportunistic | 2023 | (39,49] | 1,551 | 10.25 | 10.55 | 10.51 | 8.96 | 12.19 | 10.55 | 10.55 | 10.08 | 11.04 |
| Provence-Alpes-Côte d'Azur | Other genotypes | Organised | 2023 | (39,49] | 139 | 10.07 | 8.34 | 7.91 | 3.60 | 13.67 | 8.37 | 8.33 | 7.22 | 9.76 |
| Île-de-France | Other genotypes | Opportunistic | 2023 | (39,49] | 3,580 | 9.27 | 10.56 | 10.56 | 9.47 | 11.65 | 10.56 | 10.56 | 10.20 | 10.93 |
| Île-de-France | Other genotypes | Organised | 2023 | (39,49] | 364 | 7.97 | 7.51 | 7.42 | 4.67 | 10.44 | 7.53 | 7.50 | 6.73 | 8.45 |
| Auvergne-Rhône-Alpes | Other genotypes | Opportunistic | 2023 | (49,59] | 45 | 6.67 | 10.51 | 11.11 | 2.22 | 20.00 | 10.57 | 10.56 | 8.98 | 12.29 |
| Auvergne-Rhône-Alpes | Other genotypes | Organised | 2023 | (49,59] | 9 | 22.22 | 7.79 | 11.11 | 0.00 | 33.33 | 7.88 | 7.84 | 6.29 | 9.71 |
| Bourgogne-Franche-Comté | Other genotypes | Opportunistic | 2023 | (49,59] | 104 | 10.58 | 8.34 | 7.69 | 3.85 | 13.46 | 8.31 | 8.30 | 7.52 | 9.16 |
| Bourgogne-Franche-Comté | Other genotypes | Organised | 2023 | (49,59] | 54 | 5.56 | 6.07 | 5.56 | 0.00 | 12.96 | 6.03 | 5.99 | 4.96 | 7.27 |
| Bretagne | Other genotypes | Opportunistic | 2023 | (49,59] | 750 | 10.13 | 8.37 | 8.40 | 6.40 | 10.53 | 8.37 | 8.37 | 7.80 | 8.96 |
| Bretagne | Other genotypes | Organised | 2023 | (49,59] | 389 | 5.40 | 5.36 | 5.40 | 3.08 | 7.97 | 5.33 | 5.31 | 4.59 | 6.17 |
| Centre-Val de Loire | Other genotypes | Opportunistic | 2023 | (49,59] | 1,356 | 6.78 | 7.90 | 7.89 | 6.42 | 9.37 | 7.92 | 7.91 | 7.56 | 8.30 |
| Centre-Val de Loire | Other genotypes | Organised | 2023 | (49,59] | 169 | 2.37 | 6.00 | 5.92 | 2.37 | 10.06 | 6.03 | 6.03 | 5.37 | 6.76 |
| Grand Est | Other genotypes | Opportunistic | 2023 | (49,59] | 693 | 8.23 | 8.14 | 8.08 | 6.20 | 10.25 | 8.14 | 8.13 | 7.62 | 8.69 |
| Grand Est | Other genotypes | Organised | 2023 | (49,59] | 129 | 4.65 | 5.84 | 5.43 | 2.33 | 10.08 | 5.81 | 5.80 | 5.01 | 6.69 |

**Table S8 Observed HR HPV cervical infection prevalence, posterior predictive HR HPV cervical infection prevalence, and posterior expected HR HPV cervical infection prevalence, stratified by various dimensions.** ***(continued)***

| | | | | | | | Posterior distribution | | | | | | | |
|---|---|---|---|---|---|---|---|---|---|---|---|---|---|---|
| | | | | | | | Predictive prevalence | | | | Expected prevalence | | | |
| | | | | | | | | | ETI95% | | | | ETI95% | |
| Region | Genotypes | Type of screening | Year | Age | Number of tests | Observed | Average | Median | LB | UB | Average | Median | LB | UB |
| Hauts-de-France | Other genotypes | Opportunistic | 2023 | (49,59] | 1,553 | 9.08 | 7.80 | 7.79 | 6.44 | 9.21 | 7.80 | 7.79 | 7.45 | 8.16 |
| Hauts-de-France | Other genotypes | Organised | 2023 | (49,59] | 118 | 2.54 | 5.86 | 5.93 | 1.69 | 11.02 | 5.92 | 5.91 | 5.19 | 6.71 |
| Normandie | Other genotypes | Opportunistic | 2023 | (49,59] | 825 | 8.24 | 8.48 | 8.48 | 6.55 | 10.43 | 8.50 | 8.49 | 8.05 | 8.95 |
| Normandie | Other genotypes | Organised | 2023 | (49,59] | 35 | 2.86 | 6.25 | 5.71 | 0.00 | 14.29 | 6.24 | 6.24 | 5.45 | 7.07 |
| Nouvelle-Aquitaine | Other genotypes | Opportunistic | 2023 | (49,59] | 1,712 | 8.88 | 8.57 | 8.59 | 7.24 | 9.93 | 8.56 | 8.56 | 8.20 | 8.95 |
| Nouvelle-Aquitaine | Other genotypes | Organised | 2023 | (49,59] | 456 | 7.02 | 6.77 | 6.80 | 4.60 | 9.21 | 6.75 | 6.74 | 6.07 | 7.50 |
| Occitanie | Other genotypes | Opportunistic | 2023 | (49,59] | 307 | 9.12 | 9.31 | 9.12 | 6.19 | 12.70 | 9.28 | 9.26 | 8.61 | 10.02 |
| Occitanie | Other genotypes | Organised | 2023 | (49,59] | 60 | 6.67 | 7.24 | 6.67 | 1.67 | 15.00 | 7.28 | 7.27 | 6.33 | 8.35 |
| Pays de la Loire | Other genotypes | Opportunistic | 2023 | (49,59] | 274 | 7.30 | 7.49 | 7.30 | 4.38 | 10.95 | 7.51 | 7.50 | 6.88 | 8.23 |
| Pays de la Loire | Other genotypes | Organised | 2023 | (49,59] | 129 | 6.98 | 5.47 | 5.43 | 2.33 | 10.08 | 5.50 | 5.49 | 4.78 | 6.31 |
| Provence-Alpes-Côte d'Azur | Other genotypes | Opportunistic | 2023 | (49,59] | 1,337 | 10.70 | 8.96 | 8.98 | 7.40 | 10.55 | 8.98 | 8.98 | 8.55 | 9.42 |
| Provence-Alpes-Côte d'Azur | Other genotypes | Organised | 2023 | (49,59] | 131 | 6.87 | 7.67 | 7.63 | 3.05 | 12.21 | 7.63 | 7.59 | 6.53 | 9.01 |
| Île-de-France | Other genotypes | Opportunistic | 2023 | (49,59] | 2,833 | 8.08 | 8.86 | 8.86 | 7.80 | 9.99 | 8.86 | 8.86 | 8.54 | 9.20 |
| Île-de-France | Other genotypes | Organised | 2023 | (49,59] | 313 | 5.11 | 6.85 | 6.71 | 4.15 | 9.90 | 6.83 | 6.81 | 6.07 | 7.65 |
| Auvergne-Rhône-Alpes | Other genotypes | Opportunistic | 2023 | (59,66] | 28 | 3.57 | 9.72 | 10.71 | 0.00 | 21.43 | 9.58 | 9.57 | 8.18 | 11.10 |
| Auvergne-Rhône-Alpes | Other genotypes | Organised | 2023 | (59,66] | 4 | 0.00 | 7.30 | 0.00 | 0.00 | 50.00 | 7.45 | 7.39 | 5.90 | 9.28 |
| Bourgogne-Franche-Comté | Other genotypes | Opportunistic | 2023 | (59,66] | 60 | 15.00 | 7.50 | 6.67 | 1.67 | 15.00 | 7.46 | 7.45 | 6.68 | 8.31 |
| Bourgogne-Franche-Comté | Other genotypes | Organised | 2023 | (59,66] | 32 | 6.25 | 5.80 | 6.25 | 0.00 | 15.62 | 5.85 | 5.83 | 4.76 | 7.08 |
| Bretagne | Other genotypes | Opportunistic | 2023 | (59,66] | 388 | 7.99 | 7.25 | 7.22 | 4.64 | 9.79 | 7.25 | 7.25 | 6.72 | 7.82 |
| Bretagne | Other genotypes | Organised | 2023 | (59,66] | 337 | 4.45 | 5.13 | 5.04 | 2.67 | 7.72 | 5.12 | 5.10 | 4.30 | 6.07 |
| Centre-Val de Loire | Other genotypes | Opportunistic | 2023 | (59,66] | 629 | 6.52 | 6.91 | 6.84 | 4.93 | 9.06 | 6.91 | 6.91 | 6.52 | 7.29 |
| Centre-Val de Loire | Other genotypes | Organised | 2023 | (59,66] | 120 | 5.00 | 5.67 | 5.83 | 1.67 | 10.00 | 5.64 | 5.63 | 4.88 | 6.48 |
| Corse | Other genotypes | Opportunistic | 2023 | (59,66] | 3 | 0.00 | 8.02 | 0.00 | 0.00 | 33.33 | 7.75 | 7.67 | 5.63 | 10.49 |
| Grand Est | Other genotypes | Opportunistic | 2023 | (59,66] | 384 | 5.99 | 7.31 | 7.29 | 4.69 | 10.16 | 7.24 | 7.24 | 6.73 | 7.77 |
| Grand Est | Other genotypes | Organised | 2023 | (59,66] | 89 | 4.49 | 5.68 | 5.62 | 1.12 | 11.24 | 5.74 | 5.73 | 4.81 | 6.75 |
| Hauts-de-France | Other genotypes | Opportunistic | 2023 | (59,66] | 840 | 8.33 | 6.99 | 6.90 | 5.36 | 8.81 | 6.99 | 6.99 | 6.61 | 7.38 |
| Hauts-de-France | Other genotypes | Organised | 2023 | (59,66] | 116 | 9.48 | 5.74 | 6.03 | 1.72 | 10.34 | 5.85 | 5.83 | 5.04 | 6.78 |
| Normandie | Other genotypes | Opportunistic | 2023 | (59,66] | 381 | 8.14 | 7.45 | 7.35 | 4.99 | 10.24 | 7.45 | 7.44 | 7.01 | 7.92 |
| Normandie | Other genotypes | Organised | 2023 | (59,66] | 71 | 5.63 | 6.04 | 5.63 | 1.41 | 12.68 | 6.04 | 6.02 | 5.17 | 7.00 |
| Nouvelle-Aquitaine | Other genotypes | Opportunistic | 2023 | (59,66] | 859 | 8.27 | 7.72 | 7.68 | 5.94 | 9.55 | 7.70 | 7.70 | 7.29 | 8.13 |
| Nouvelle-Aquitaine | Other genotypes | Organised | 2023 | (59,66] | 308 | 5.19 | 6.48 | 6.49 | 3.90 | 9.74 | 6.53 | 6.52 | 5.73 | 7.43 |
| Occitanie | Other genotypes | Opportunistic | 2023 | (59,66] | 168 | 8.93 | 8.18 | 8.33 | 4.17 | 12.50 | 8.23 | 8.22 | 7.61 | 8.94 |
| Occitanie | Other genotypes | Organised | 2023 | (59,66] | 52 | 9.62 | 7.04 | 5.77 | 1.92 | 15.38 | 7.13 | 7.10 | 6.03 | 8.36 |
| Pays de la Loire | Other genotypes | Opportunistic | 2023 | (59,66] | 126 | 10.32 | 6.98 | 7.14 | 3.17 | 11.13 | 7.05 | 7.04 | 6.37 | 7.79 |
| Pays de la Loire | Other genotypes | Organised | 2023 | (59,66] | 97 | 4.12 | 5.22 | 5.15 | 1.03 | 9.30 | 5.22 | 5.20 | 4.44 | 6.10 |
| Provence-Alpes-Côte d'Azur | Other genotypes | Opportunistic | 2023 | (59,66] | 667 | 9.15 | 8.00 | 7.95 | 6.00 | 10.19 | 8.01 | 8.01 | 7.57 | 8.46 |
| Provence-Alpes-Côte d'Azur | Other genotypes | Organised | 2023 | (59,66] | 87 | 8.05 | 7.21 | 6.90 | 2.30 | 12.64 | 7.32 | 7.28 | 6.21 | 8.65 |
| Île-de-France | Other genotypes | Opportunistic | 2023 | (59,66] | 1,258 | 10.10 | 7.84 | 7.79 | 6.44 | 9.38 | 7.84 | 7.84 | 7.48 | 8.22 |

| | | | | | | | | | | | | | | |
|---|---|---|---|---|---|---|---|---|---|---|---|---|---|---|
| Île-de-France | Other genotypes | Organised | 2023 | (59,66] | 143 | 11.89 | 6.54 | 6.29 | 2.80 | 10.49 | 6.58 | 6.56 | 5.74 | 7.53 |

#### S8 Systematic difference in expected HR HPV cervical infection prevalence between opportunistic and organised screening, among females aged 30, at the end of November 2023

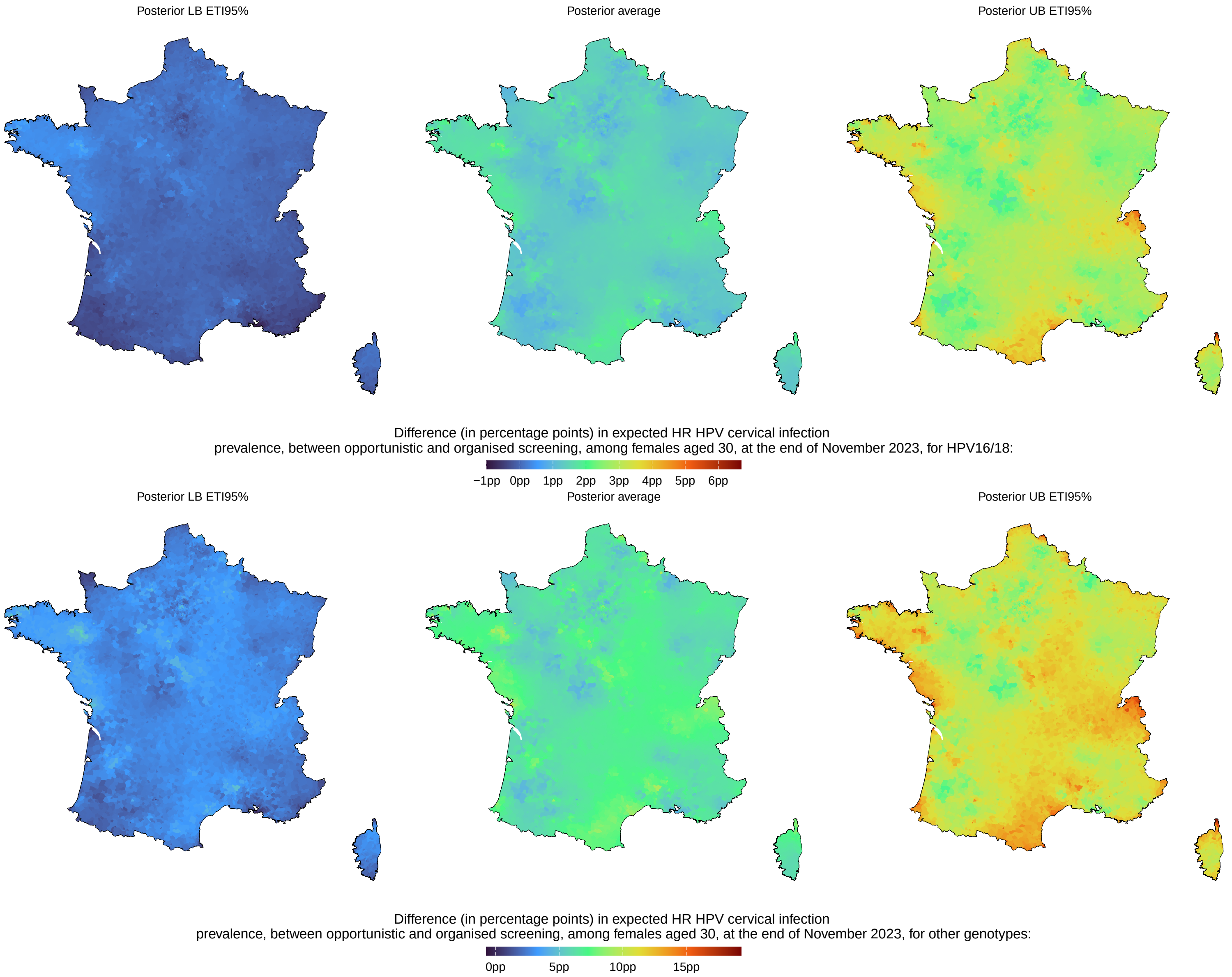

**Figure S21 Posterior difference between the expected HR HPV cervical infection prevalence from opportunistic and organised screening, among females aged 30, at the end of November 2023.**

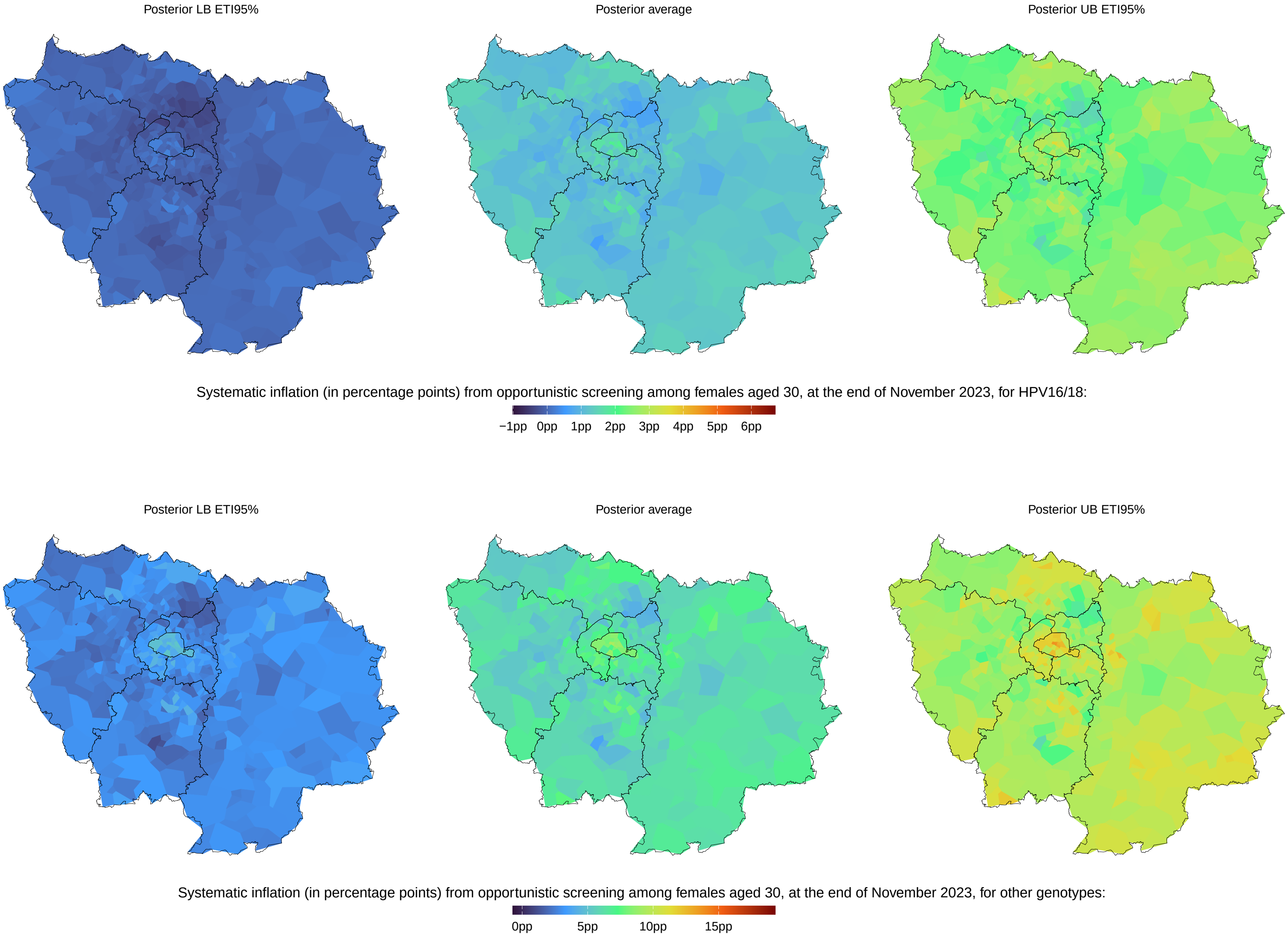

**Figure S22 Posterior difference between the expected HR HPV using opportunistic and organised data, among females aged 30, at the end of November 2023, in Paris region.**

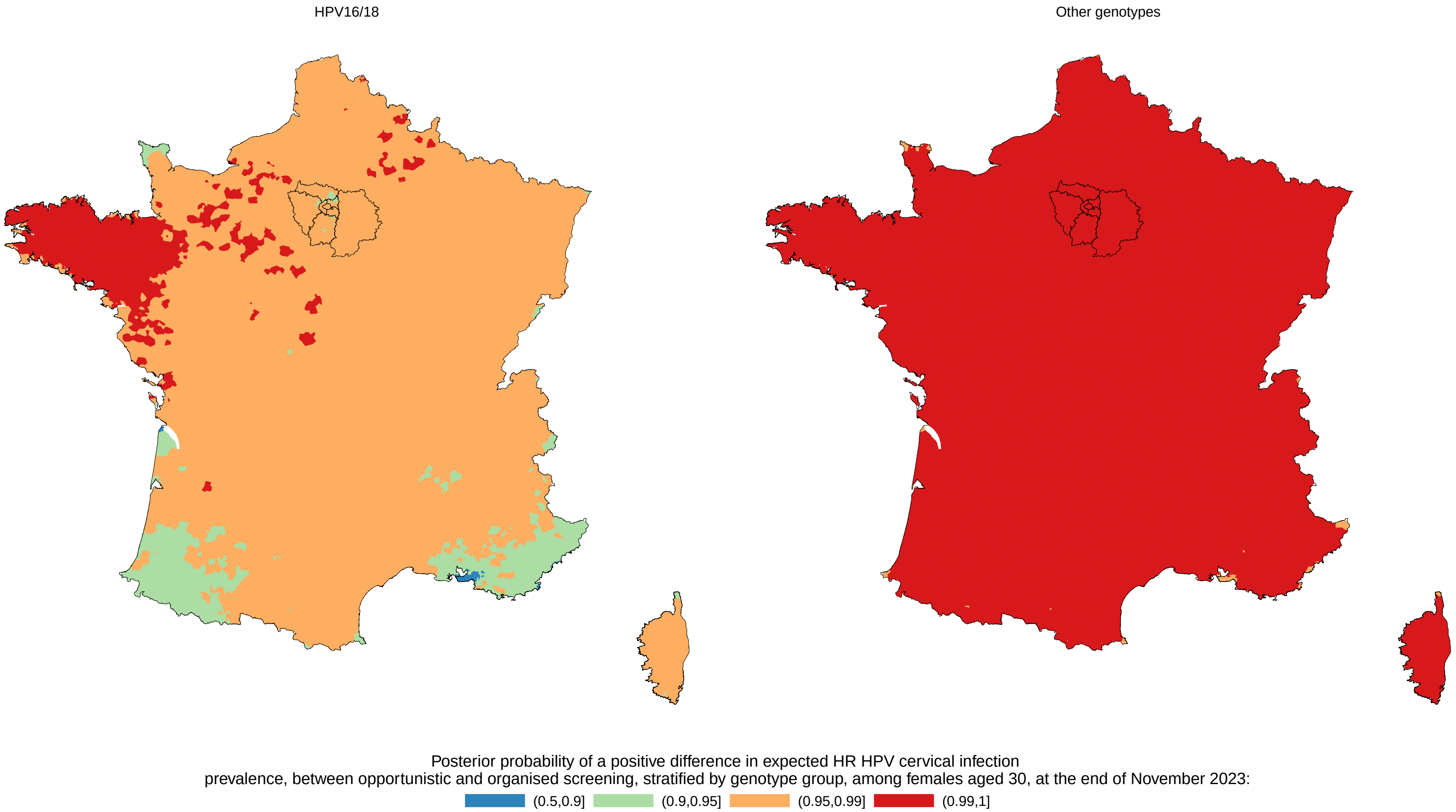

**Figure S23 Posterior probability of a positive difference in expected HR HPV between opportunistic and organised screening, stratified by genotype group, among females aged 30, at the end of November 2023.**

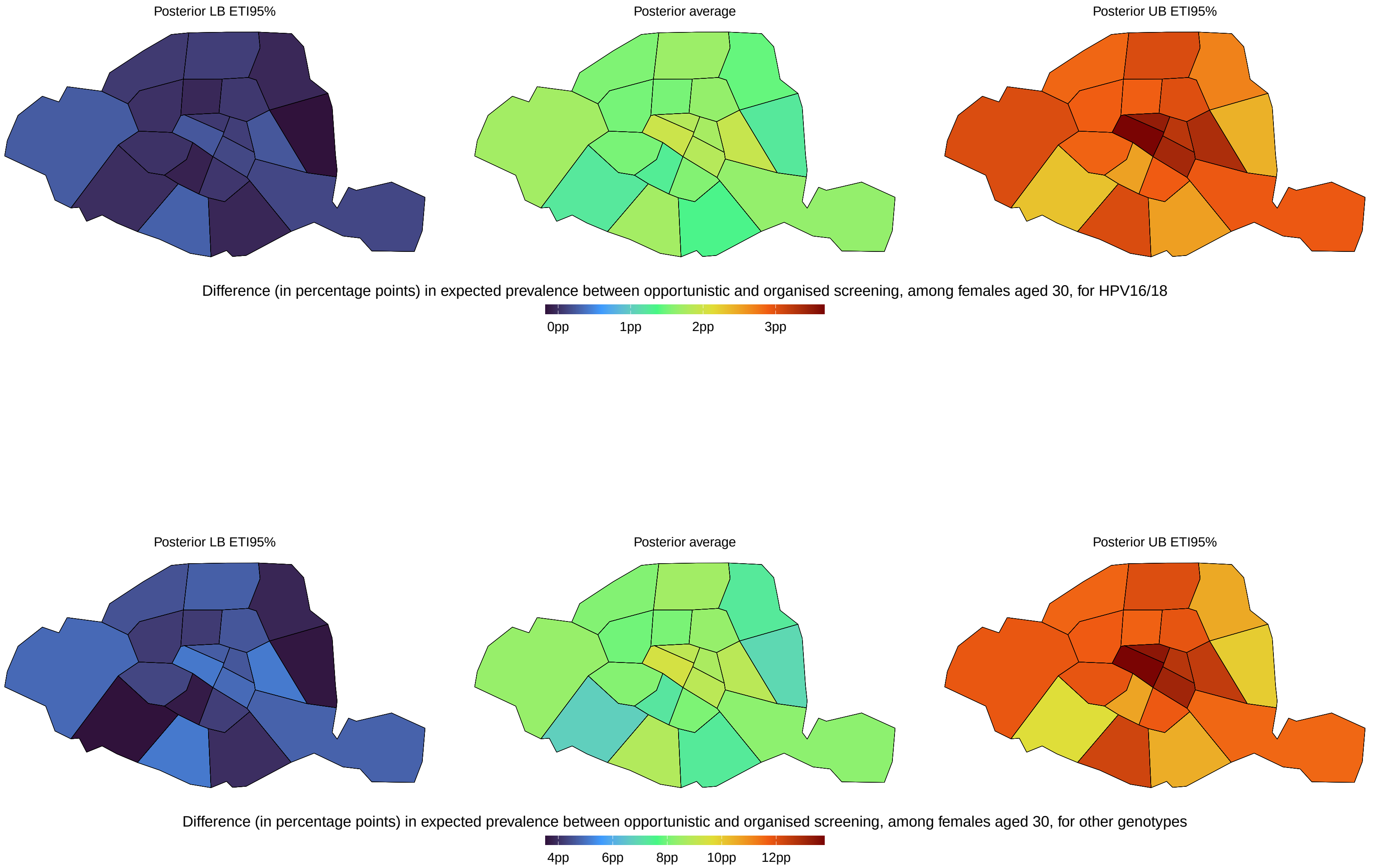

**Figure S24 Posterior systematic inflation in HR HPV cervical infection prevalence under opportunistic screening, compared with organised screening, in each of Paris arrondissement, among females aged 30, at the end of November 2023.**

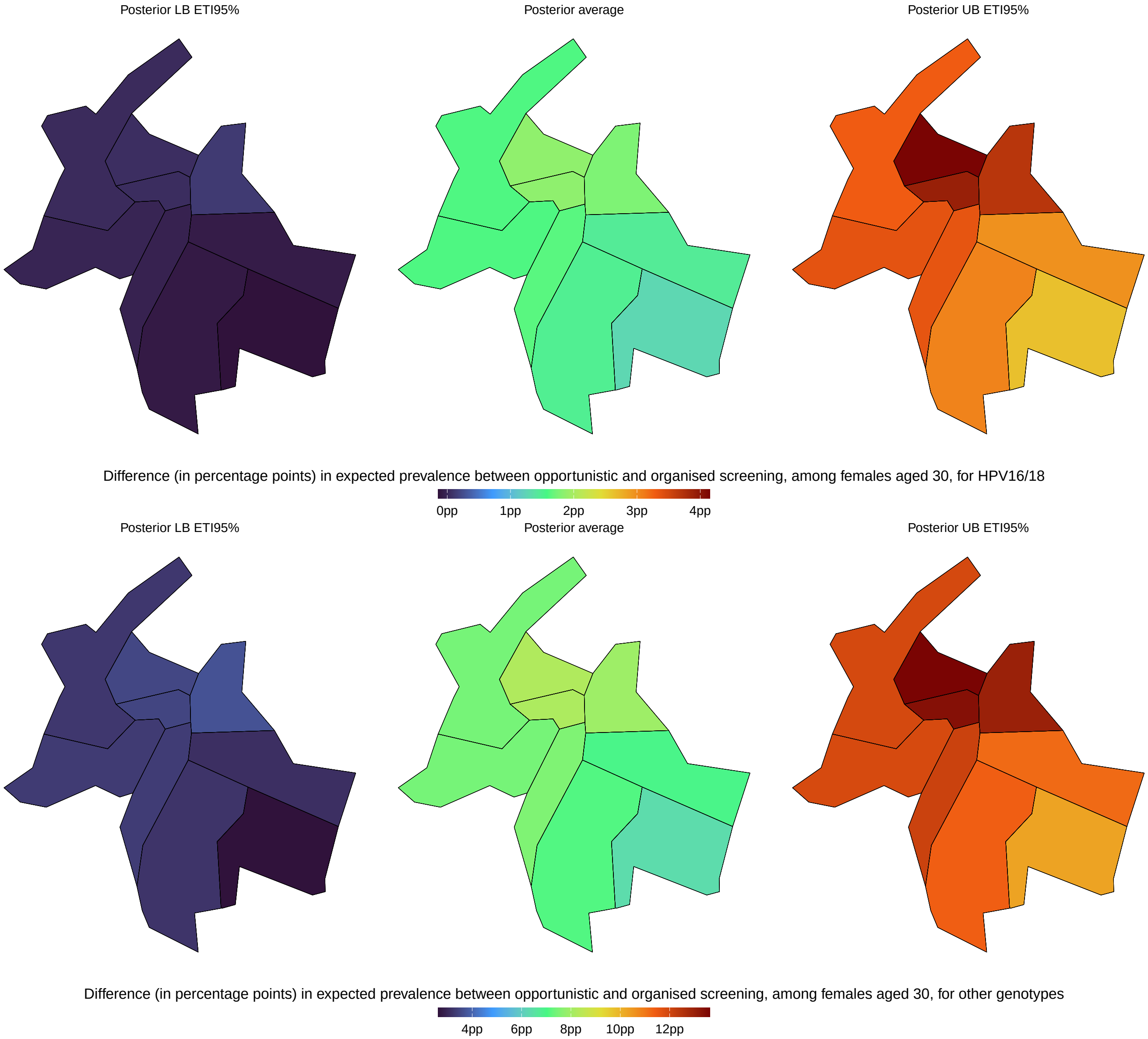

**Figure S25 Posterior systematic inflation in HR HPV cervical infection prevalence under opportunistic screening, compared with organised screening, in each of Lyon arrondissement, among females aged 30, at the end of November 2023.**

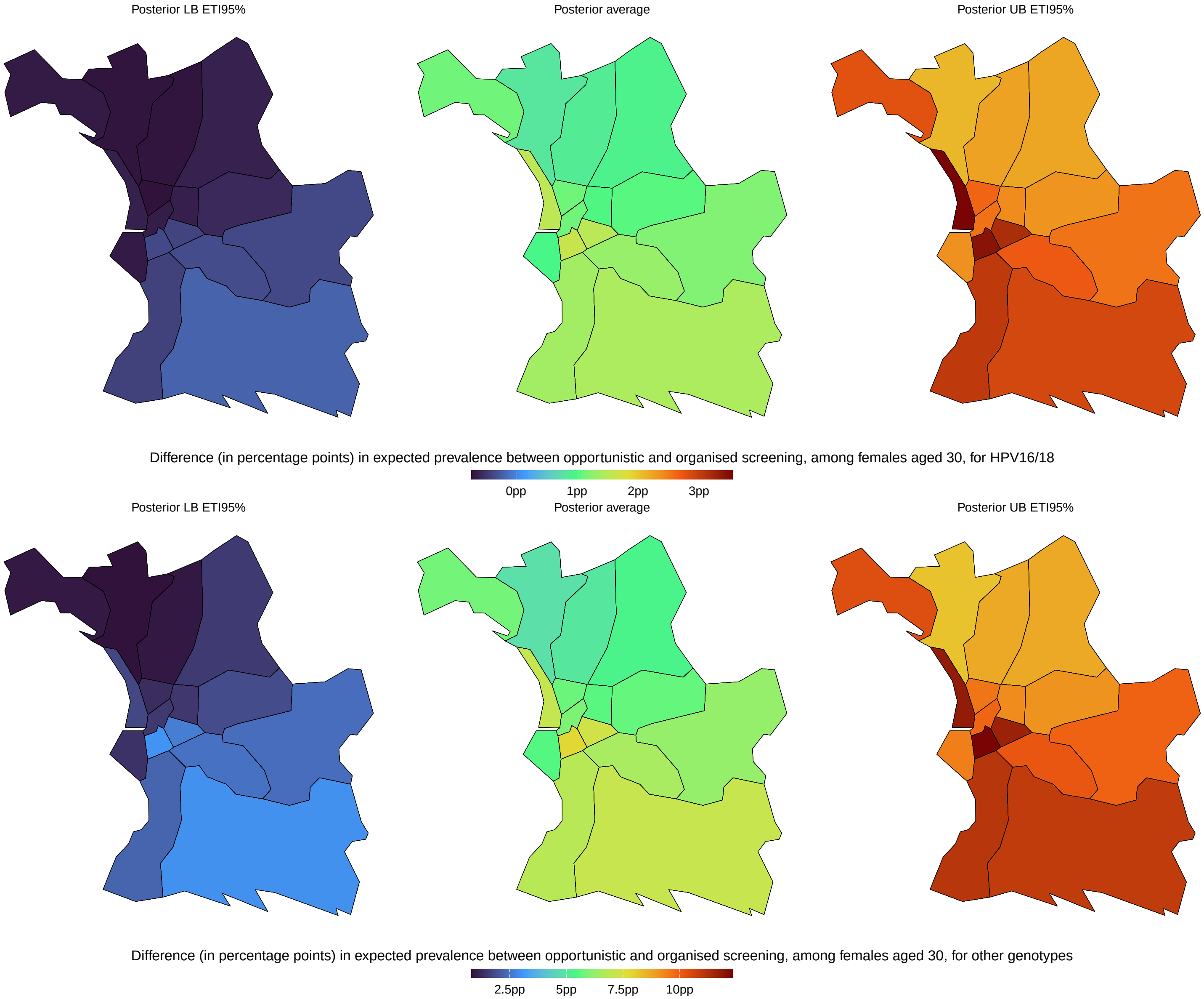

**Figure S26 Posterior systematic inflation in HR HPV cervical infection prevalence under opportunistic screening, compared with organised screening, in each of Marseille arrondissement, among females aged 30, at the end of November 2023.**

### S9 Expected HR HPV cervical infection prevalence, among females aged 30, at the end of November 2023, under organised screening

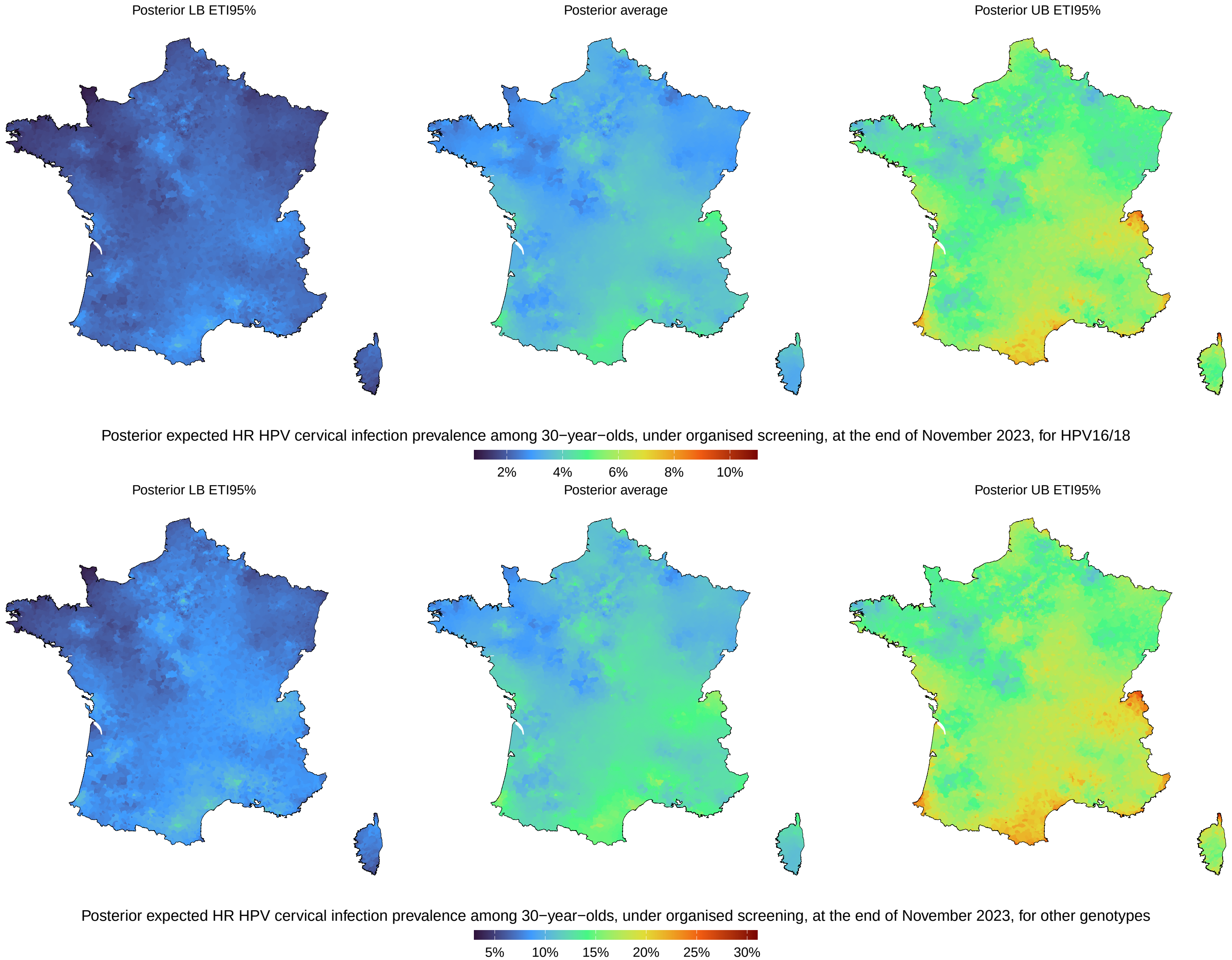

**Figure S27 Posterior HR HPV cervical infection prevalence in metropolitan France, among females aged 30, at the end of November 2023, correcting for the systematic inflation associated with opportunistic screening, stratified by genotype group.**

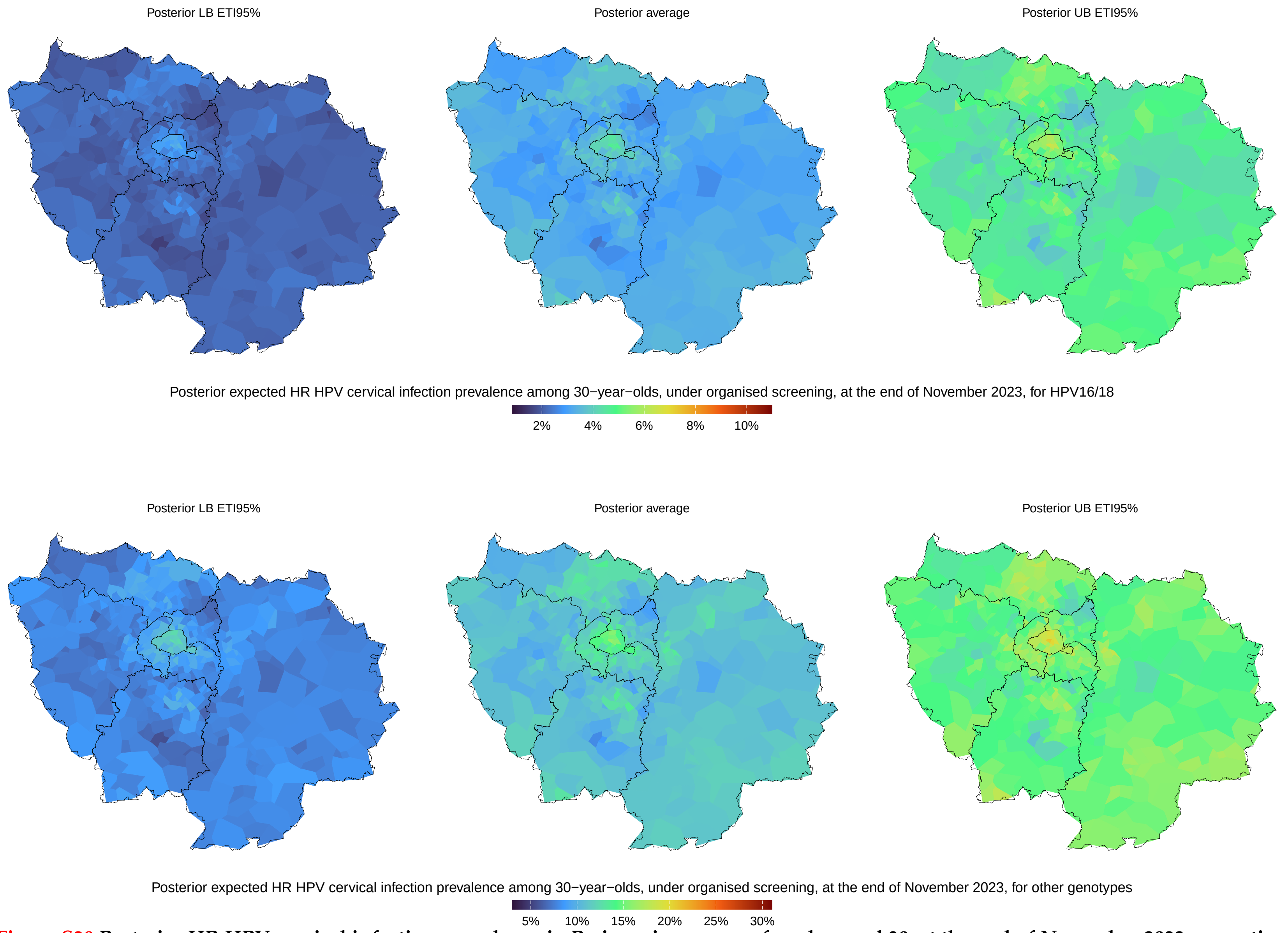

**Figure S28 Posterior HR HPV cervical infection prevalence in Paris region, among females aged 30, at the end of November 2023, correcting for the systematic inflation associated with opportunistic screening, stratified by genotype group.**

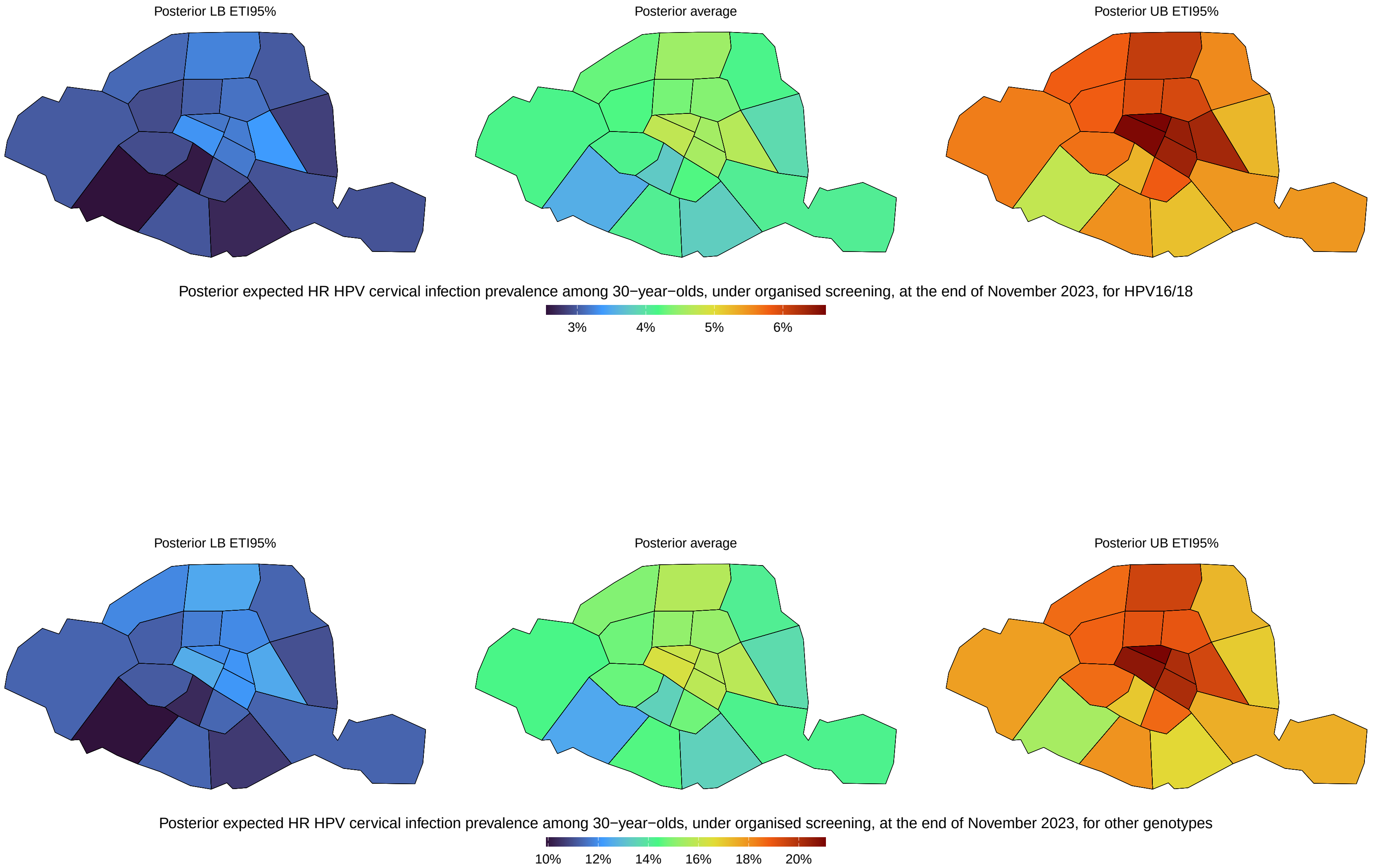

**Figure S29 Posterior HR HPV cervical infection prevalence in each of the Paris arrondissement, among females aged 30, at the end of November 2023, correcting for the systematic inflation associated with opportunistic screening, stratified by genotype group.**

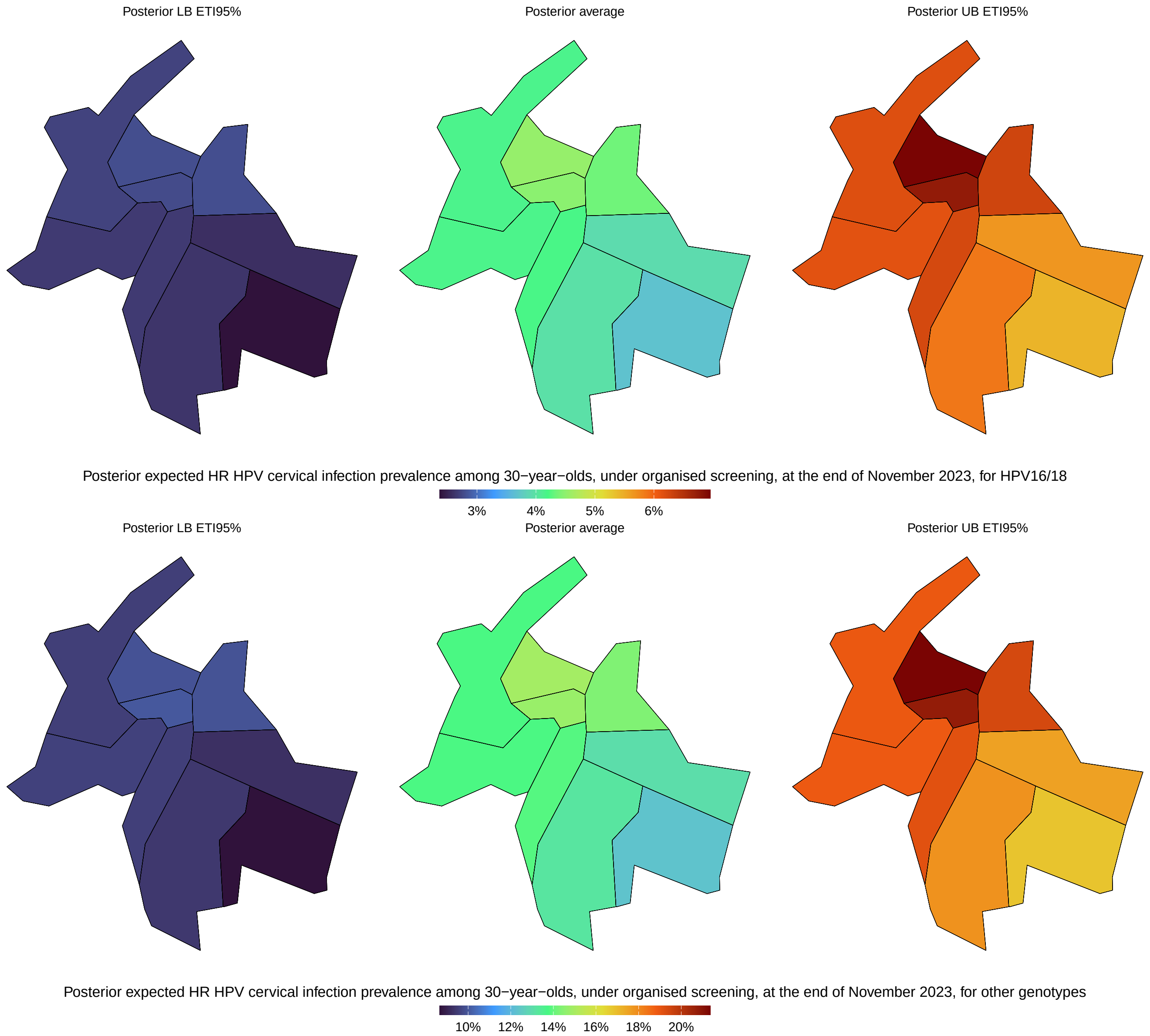

**Figure S30 Posterior HR HPV cervical infection prevalence in each of the Lyon arrondissement, among females aged 30, at the end of November 2023, correcting for the systematic inflation associated with opportunistic screening, stratified by genotype group.**

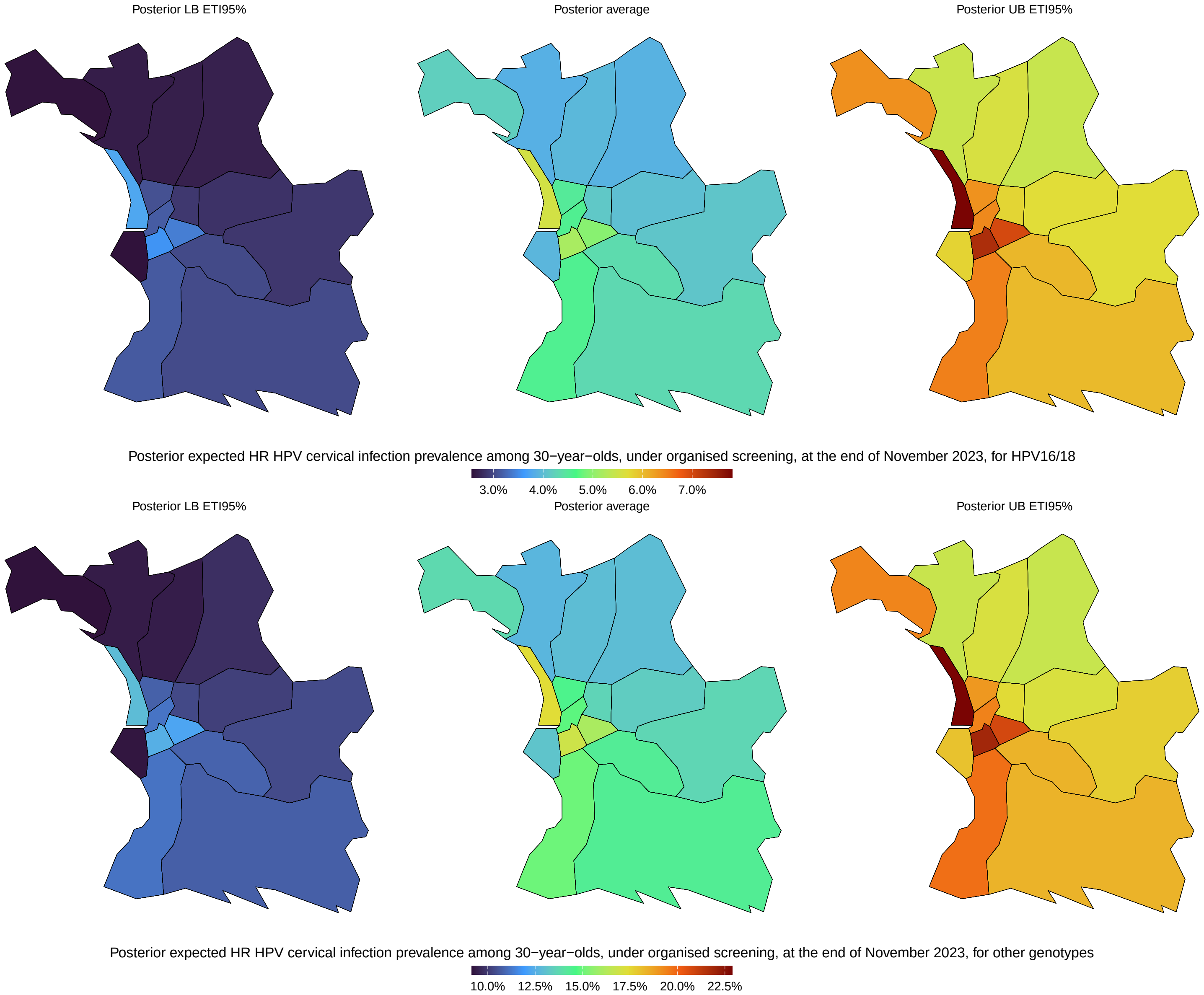

**Figure S31 Posterior HR HPV cervical infection prevalence in each of the Marseille arrondissement, among females aged 30, at the end of November 2023, correcting for the systematic inflation associated with opportunistic screening, stratified by genotype group.**

#### S10 Maps of the posterior average expected infection prevalence stratified by age, at the end of November 2023, under organised screening

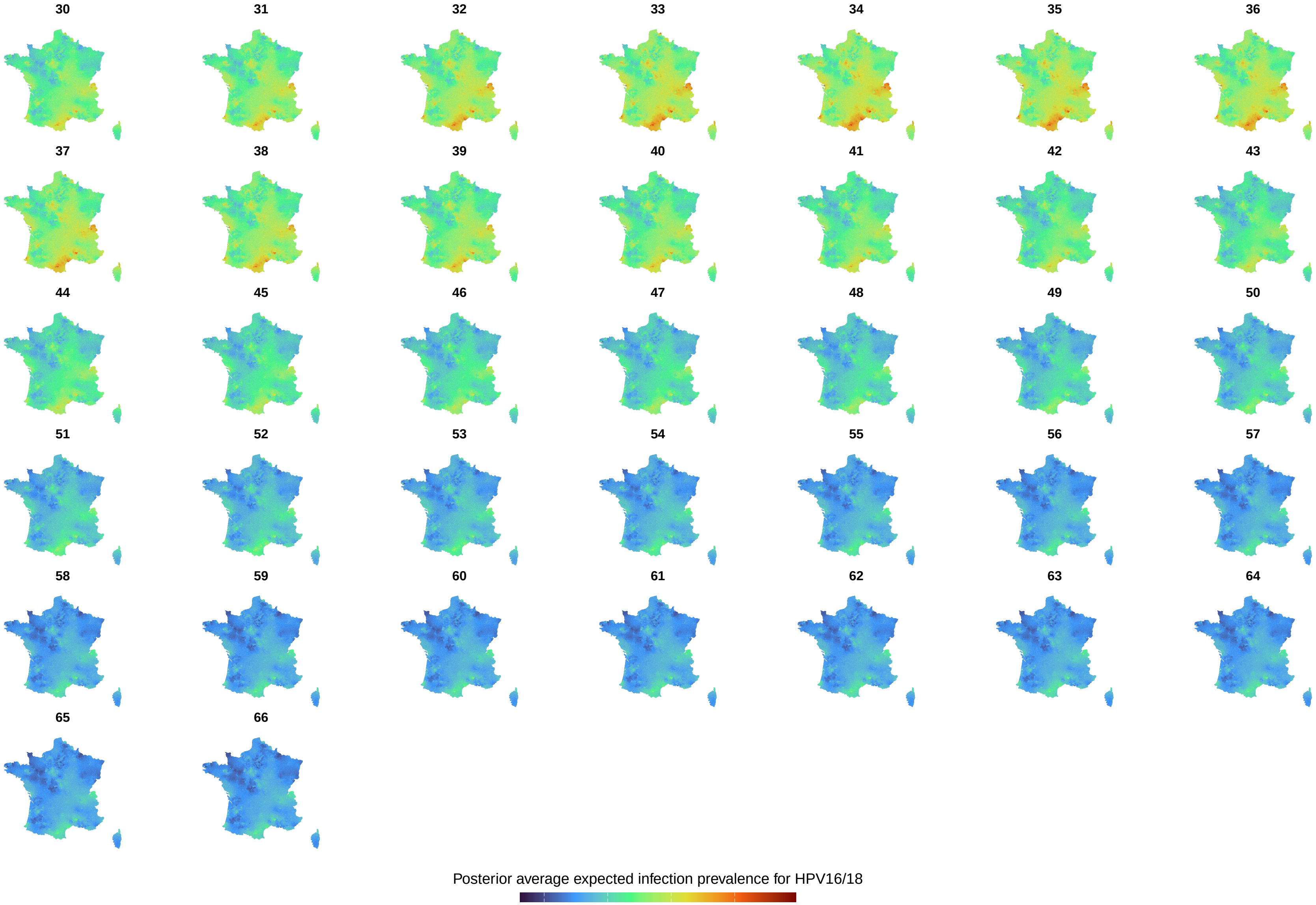

**Figure S32** **Posterior average expected infection prevalence for HPV16/18 under organised screening, across metropolitan France, stratified by age.**

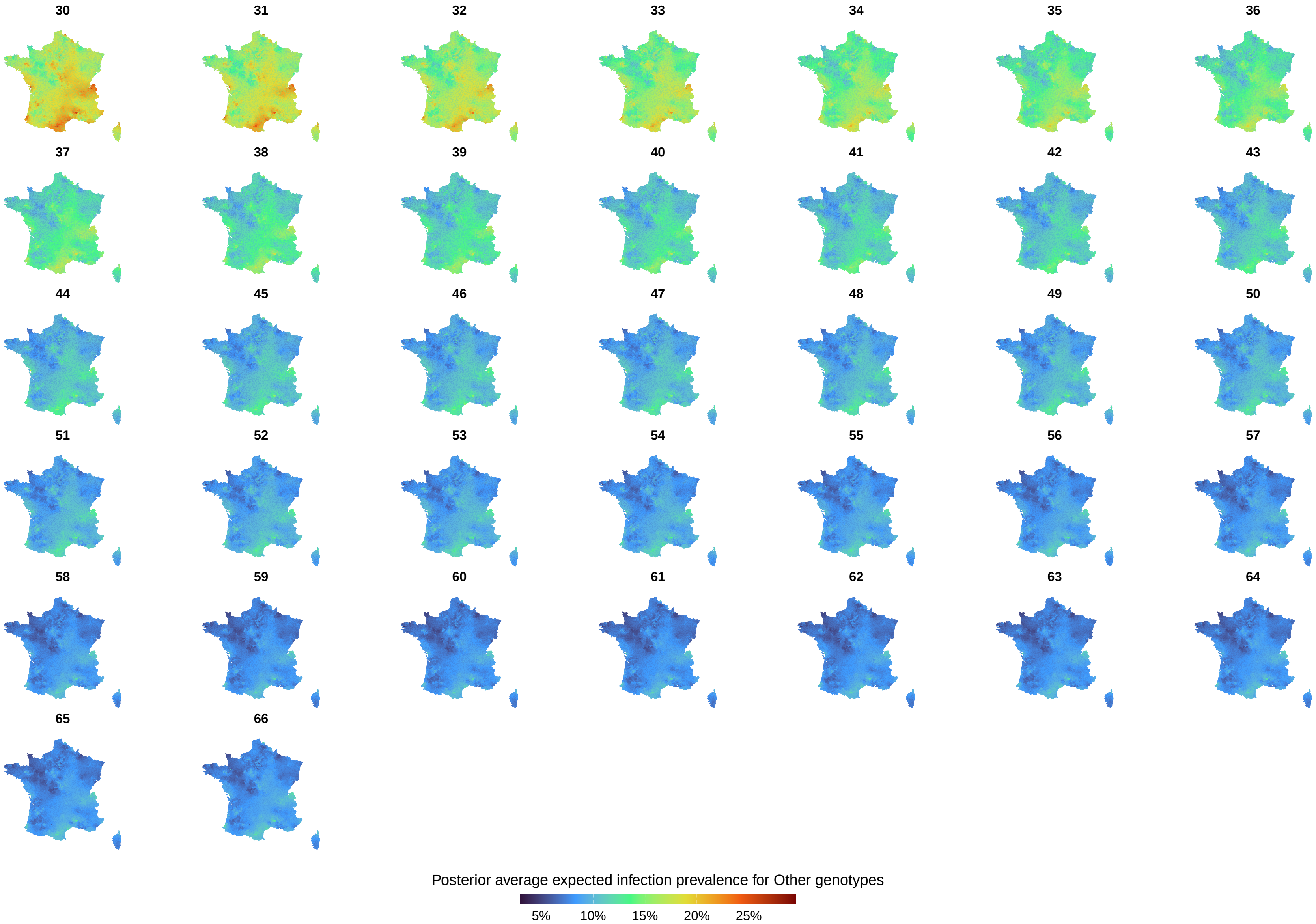

**Figure S33** **Posterior average expected infection prevalence for Other genotypes, under organised screening, across metropolitan France, stratified by age.**

#### S11 Difference in expected HR HPV cervical infection prevalence, between the two groups of genotypes

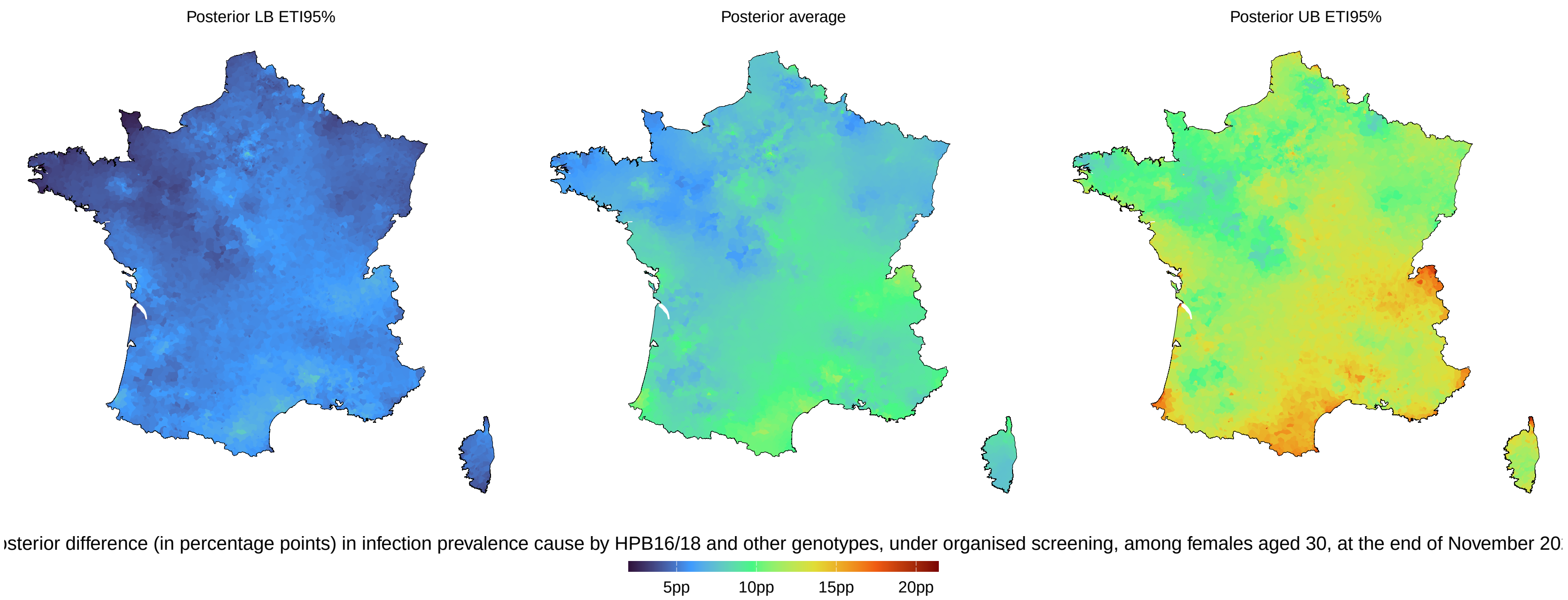

**Figure S34 Summary of the posterior distribution of the difference (in percentage points) in infection prevalence cause by HPB16/18 and other genotypes, under organised screening, among females aged 30, at the end of November 2023.**

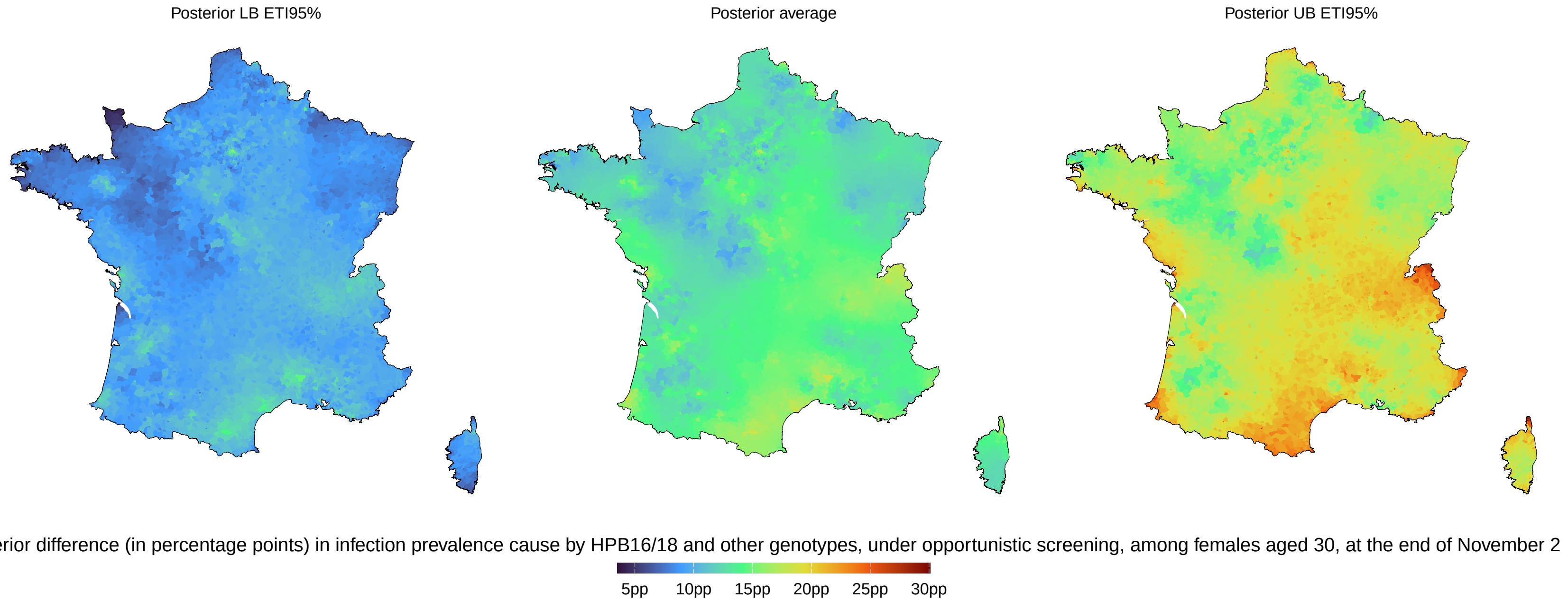

**Figure S35** **Summary of the posterior distribution of the difference (in percentage points) in infection prevalence cause by HPB16/18 and other genotypes, under opportunistic screening, among females aged 30, at the end of November 2023.**

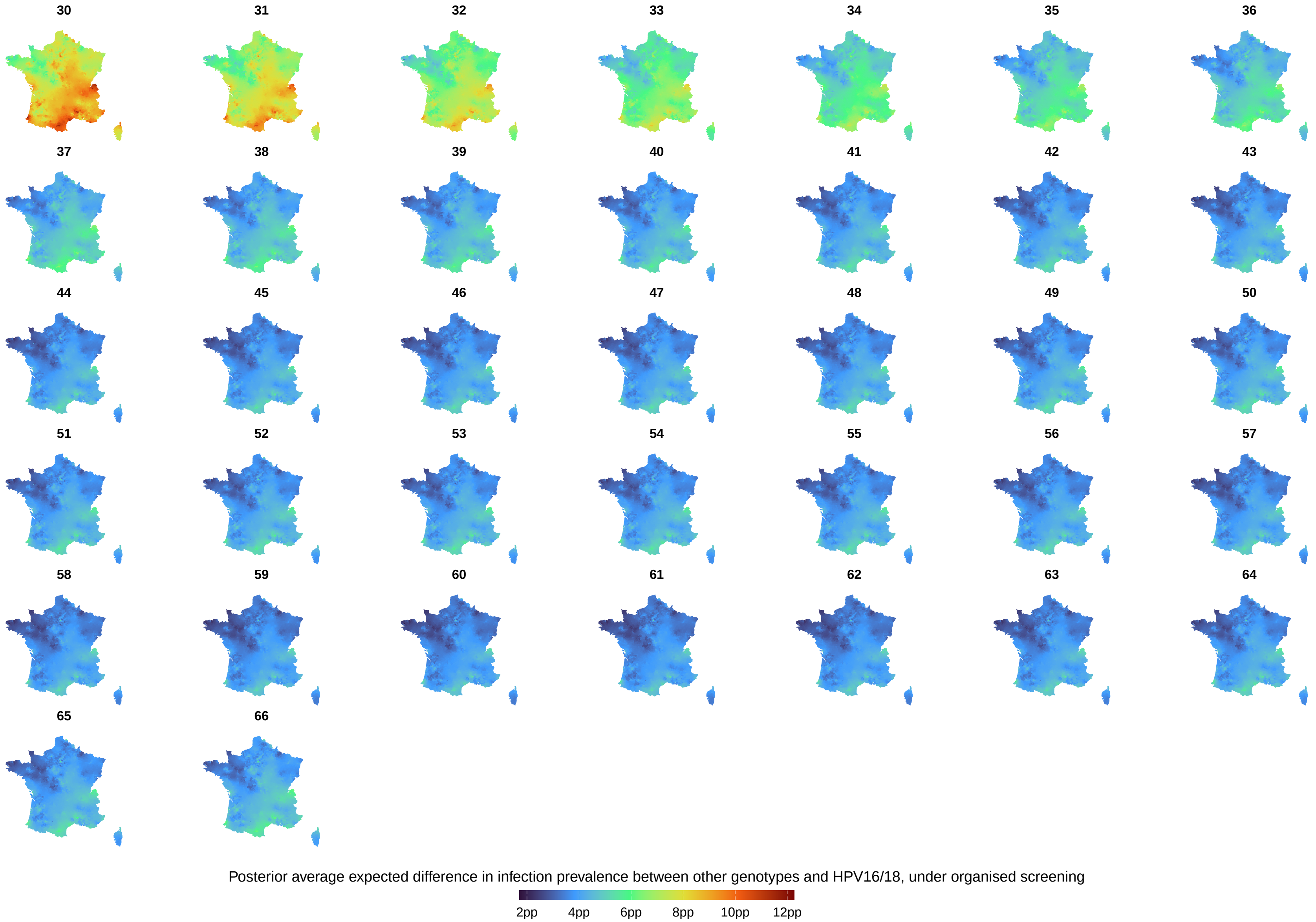

**Figure S36 Posterior average expected difference in infection prevalence between other genotypes and HPV16/18, across metropolitan France, stratified by age, under organised screening.**

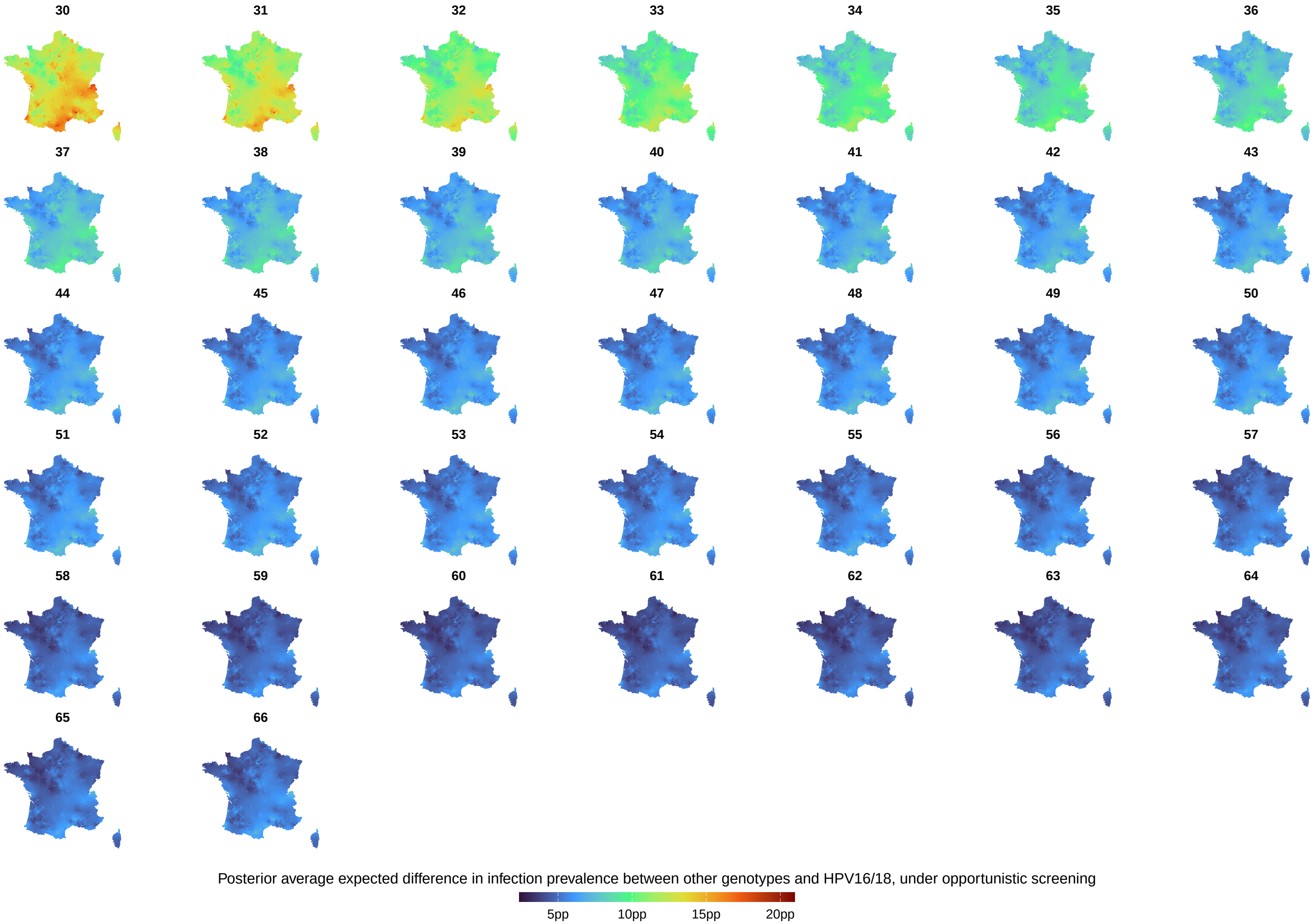

**Figure S37 Posterior average expected difference in infection prevalence between other genotypes and HPV16/18, across metropolitan France, stratified by age, under opportunistic screening.**

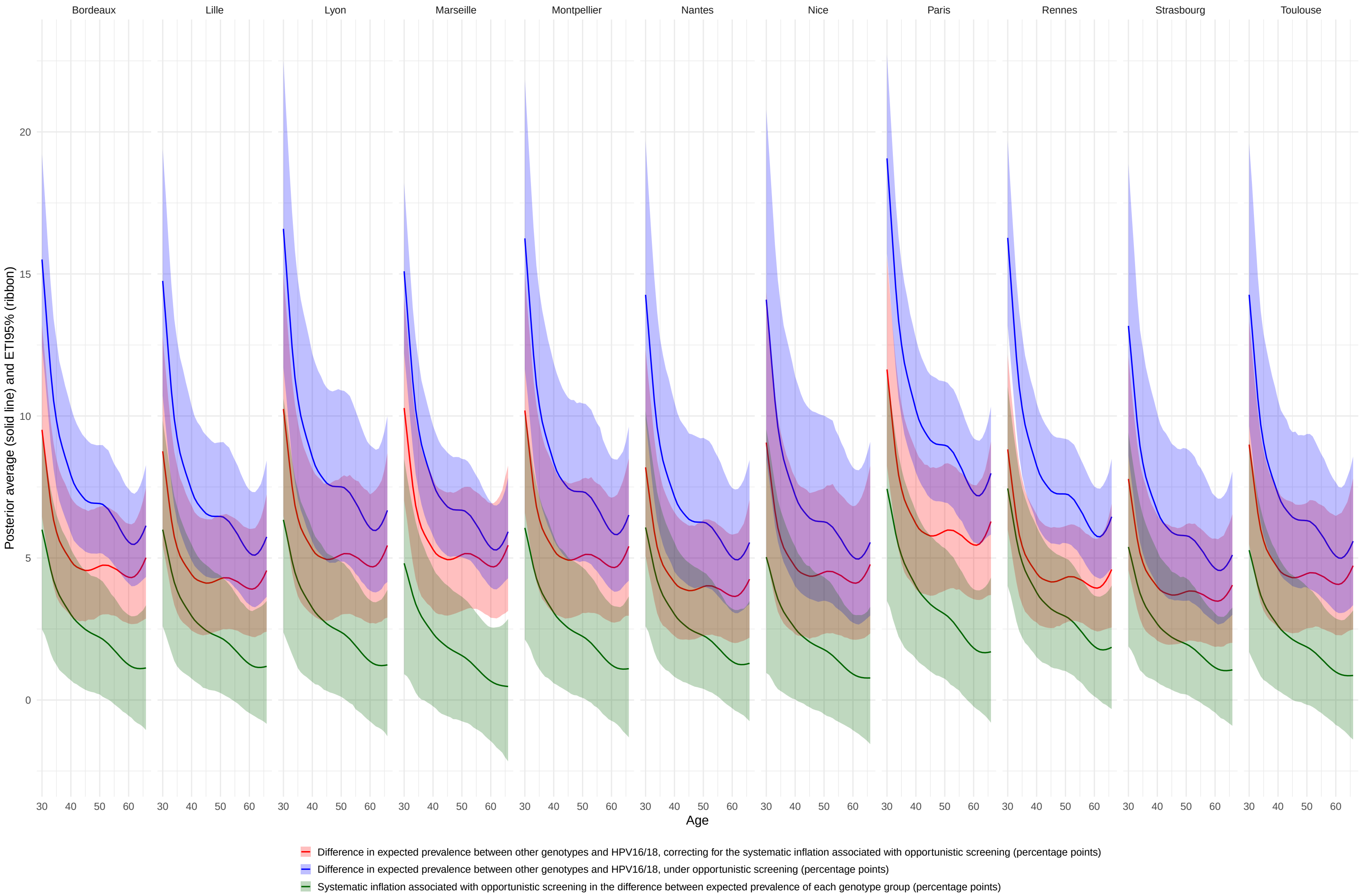

**Figure S38** **Summary of the posterior distribution of the difference (in percentage points), between opportunistic and organised screening, in the expected HR HPV cervical infection prevalence, in major French cities, at the end of November 2023, stratified by age and genotype group.**

#### S12 Figure 3A, B, C, and D in table format

**Table S9 Posterior expected HR HPV cervical infection prevalence (in %) in major French cities, stratified by type of test, city, and age. Table counterpart to Figure 3A**

| | | | | Posterior distribution | | | | | | | |
|---|---|---|---|---|---|---|---|---|---|---|---|
| | | | | | Percentiles | | | | | | |
| Virus | Test | City | Age | Average | 0.01 | 0.025 | 0.1 | 0.5 | 0.9 | 0.975 | 0.99 |
| HPV16/18 | Organised | Nice | 30 | 3.82 | 1.77 | 2.01 | 2.49 | 3.66 | 5.35 | 6.57 | 7.23 |
| HPV16/18 | Organised | Nice | 31 | 4.01 | 1.88 | 2.12 | 2.63 | 3.84 | 5.60 | 6.84 | 7.50 |
| HPV16/18 | Organised | Nice | 32 | 4.18 | 1.99 | 2.23 | 2.75 | 4.01 | 5.80 | 7.05 | 7.82 |
| HPV16/18 | Organised | Nice | 33 | 4.31 | 2.06 | 2.32 | 2.83 | 4.13 | 5.97 | 7.24 | 8.00 |
| HPV16/18 | Organised | Nice | 34 | 4.35 | 2.10 | 2.35 | 2.87 | 4.17 | 6.04 | 7.33 | 8.10 |
| HPV16/18 | Organised | Nice | 35 | 4.31 | 2.10 | 2.33 | 2.86 | 4.13 | 5.98 | 7.24 | 8.04 |
| HPV16/18 | Organised | Nice | 36 | 4.23 | 2.06 | 2.29 | 2.81 | 4.05 | 5.86 | 7.09 | 7.77 |
| HPV16/18 | Organised | Nice | 37 | 4.12 | 2.03 | 2.22 | 2.74 | 3.94 | 5.74 | 6.88 | 7.61 |
| HPV16/18 | Organised | Nice | 38 | 4.01 | 2.00 | 2.15 | 2.66 | 3.84 | 5.59 | 6.71 | 7.38 |
| HPV16/18 | Organised | Nice | 39 | 3.91 | 1.96 | 2.11 | 2.59 | 3.75 | 5.46 | 6.52 | 7.27 |
| HPV16/18 | Organised | Nice | 40 | 3.81 | 1.89 | 2.06 | 2.53 | 3.65 | 5.31 | 6.38 | 7.12 |
| HPV16/18 | Organised | Nice | 41 | 3.72 | 1.84 | 2.00 | 2.47 | 3.56 | 5.19 | 6.20 | 6.93 |
| HPV16/18 | Organised | Nice | 42 | 3.62 | 1.80 | 1.96 | 2.39 | 3.46 | 5.03 | 6.04 | 6.75 |
| HPV16/18 | Organised | Nice | 43 | 3.52 | 1.75 | 1.92 | 2.33 | 3.38 | 4.89 | 5.87 | 6.54 |
| HPV16/18 | Organised | Nice | 44 | 3.45 | 1.72 | 1.88 | 2.27 | 3.30 | 4.79 | 5.77 | 6.39 |
| HPV16/18 | Organised | Nice | 45 | 3.38 | 1.69 | 1.84 | 2.24 | 3.25 | 4.71 | 5.68 | 6.27 |
| HPV16/18 | Organised | Nice | 46 | 3.31 | 1.64 | 1.79 | 2.18 | 3.17 | 4.62 | 5.58 | 6.14 |
| HPV16/18 | Organised | Nice | 47 | 3.24 | 1.59 | 1.75 | 2.14 | 3.12 | 4.50 | 5.45 | 6.06 |
| HPV16/18 | Organised | Nice | 48 | 3.18 | 1.56 | 1.72 | 2.10 | 3.05 | 4.43 | 5.37 | 5.99 |
| HPV16/18 | Organised | Nice | 49 | 3.12 | 1.53 | 1.68 | 2.06 | 2.99 | 4.33 | 5.27 | 5.89 |
| HPV16/18 | Organised | Nice | 50 | 3.06 | 1.49 | 1.65 | 2.02 | 2.93 | 4.26 | 5.18 | 5.74 |
| HPV16/18 | Organised | Nice | 51 | 3.00 | 1.47 | 1.63 | 1.98 | 2.88 | 4.19 | 5.09 | 5.67 |
| HPV16/18 | Organised | Nice | 52 | 2.95 | 1.44 | 1.61 | 1.95 | 2.83 | 4.10 | 5.01 | 5.58 |
| HPV16/18 | Organised | Nice | 53 | 2.89 | 1.39 | 1.58 | 1.91 | 2.77 | 4.03 | 4.91 | 5.47 |
| HPV16/18 | Organised | Nice | 54 | 2.83 | 1.36 | 1.53 | 1.86 | 2.71 | 3.96 | 4.84 | 5.33 |
| HPV16/18 | Organised | Nice | 55 | 2.78 | 1.33 | 1.51 | 1.83 | 2.66 | 3.91 | 4.76 | 5.28 |
| HPV16/18 | Organised | Nice | 56 | 2.75 | 1.31 | 1.49 | 1.79 | 2.63 | 3.87 | 4.65 | 5.23 |

**Table S9 Posterior expected HR HPV cervical infection prevalence (in %) in major French cities, stratified by type of test, city, and age. Table counterpart to Figure 3A *(continued)***

| | | | | Posterior distribution | | | | | | | |
|---|---|---|---|---|---|---|---|---|---|---|---|
| | | | | | Percentiles | | | | | | |
| Virus | Test | City | Age | Average | 0.01 | 0.025 | 0.1 | 0.5 | 0.9 | 0.975 | 0.99 |
| HPV16/18 | Organised | Nice | 57 | 2.73 | 1.29 | 1.47 | 1.78 | 2.61 | 3.84 | 4.62 | 5.20 |
| HPV16/18 | Organised | Nice | 58 | 2.72 | 1.30 | 1.47 | 1.77 | 2.62 | 3.84 | 4.65 | 5.23 |
| HPV16/18 | Organised | Nice | 59 | 2.73 | 1.28 | 1.46 | 1.76 | 2.62 | 3.85 | 4.64 | 5.24 |
| HPV16/18 | Organised | Nice | 60 | 2.74 | 1.30 | 1.46 | 1.77 | 2.63 | 3.87 | 4.68 | 5.27 |
| HPV16/18 | Organised | Nice | 61 | 2.76 | 1.31 | 1.47 | 1.78 | 2.64 | 3.90 | 4.72 | 5.33 |
| HPV16/18 | Organised | Nice | 62 | 2.77 | 1.29 | 1.47 | 1.78 | 2.66 | 3.92 | 4.74 | 5.36 |
| HPV16/18 | Organised | Nice | 63 | 2.78 | 1.30 | 1.47 | 1.78 | 2.66 | 3.95 | 4.77 | 5.35 |
| HPV16/18 | Organised | Nice | 64 | 2.78 | 1.29 | 1.45 | 1.77 | 2.66 | 3.93 | 4.81 | 5.27 |
| HPV16/18 | Organised | Nice | 65 | 2.77 | 1.28 | 1.45 | 1.75 | 2.65 | 3.90 | 4.84 | 5.29 |
| HPV16/18 | Organised | Nice | 66 | 2.75 | 1.27 | 1.43 | 1.73 | 2.63 | 3.91 | 4.89 | 5.32 |
| HPV16/18 | Organised | Marseille | 30 | 4.56 | 2.93 | 3.15 | 3.54 | 4.48 | 5.66 | 6.42 | 6.84 |
| HPV16/18 | Organised | Marseille | 31 | 4.79 | 3.13 | 3.36 | 3.75 | 4.72 | 5.91 | 6.66 | 7.16 |
| HPV16/18 | Organised | Marseille | 32 | 4.99 | 3.27 | 3.51 | 3.94 | 4.93 | 6.12 | 6.85 | 7.29 |
| HPV16/18 | Organised | Marseille | 33 | 5.14 | 3.42 | 3.66 | 4.08 | 5.09 | 6.33 | 7.04 | 7.50 |
| HPV16/18 | Organised | Marseille | 34 | 5.20 | 3.48 | 3.70 | 4.14 | 5.14 | 6.38 | 7.10 | 7.53 |
| HPV16/18 | Organised | Marseille | 35 | 5.15 | 3.49 | 3.69 | 4.10 | 5.09 | 6.31 | 7.04 | 7.43 |
| HPV16/18 | Organised | Marseille | 36 | 5.05 | 3.42 | 3.63 | 4.03 | 5.00 | 6.18 | 6.90 | 7.31 |
| HPV16/18 | Organised | Marseille | 37 | 4.92 | 3.36 | 3.55 | 3.94 | 4.87 | 6.00 | 6.72 | 7.06 |
| HPV16/18 | Organised | Marseille | 38 | 4.80 | 3.27 | 3.45 | 3.83 | 4.74 | 5.85 | 6.54 | 6.91 |
| HPV16/18 | Organised | Marseille | 39 | 4.68 | 3.22 | 3.39 | 3.74 | 4.62 | 5.71 | 6.40 | 6.74 |
| HPV16/18 | Organised | Marseille | 40 | 4.56 | 3.14 | 3.31 | 3.65 | 4.50 | 5.57 | 6.22 | 6.61 |
| HPV16/18 | Organised | Marseille | 41 | 4.45 | 3.05 | 3.22 | 3.55 | 4.39 | 5.42 | 6.07 | 6.40 |
| HPV16/18 | Organised | Marseille | 42 | 4.33 | 2.97 | 3.13 | 3.45 | 4.26 | 5.28 | 5.88 | 6.28 |
| HPV16/18 | Organised | Marseille | 43 | 4.22 | 2.87 | 3.04 | 3.37 | 4.16 | 5.15 | 5.77 | 6.14 |
| HPV16/18 | Organised | Marseille | 44 | 4.12 | 2.81 | 2.97 | 3.29 | 4.06 | 5.02 | 5.68 | 6.02 |
| HPV16/18 | Organised | Marseille | 45 | 4.04 | 2.74 | 2.92 | 3.22 | 3.98 | 4.94 | 5.57 | 5.91 |
| HPV16/18 | Organised | Marseille | 46 | 3.96 | 2.68 | 2.85 | 3.15 | 3.90 | 4.84 | 5.44 | 5.79 |
| HPV16/18 | Organised | Marseille | 47 | 3.88 | 2.62 | 2.79 | 3.08 | 3.82 | 4.74 | 5.34 | 5.66 |
| HPV16/18 | Organised | Marseille | 48 | 3.80 | 2.56 | 2.75 | 3.02 | 3.74 | 4.65 | 5.23 | 5.56 |

**Table S9** **Posterior expected HR HPV cervical infection prevalence (in %) in major French cities, stratified by type of test, city, and age. Table counterpart to Figure 3A *(continued)***

| | | | | Posterior distribution | | | | | | | |
|---|---|---|---|---|---|---|---|---|---|---|---|
| | | | | | Percentiles | | | | | | |
| Virus | Test | City | Age | Average | 0.01 | 0.025 | 0.1 | 0.5 | 0.9 | 0.975 | 0.99 |
| HPV16/18 | Organised | Marseille | 49 | 3.73 | 2.52 | 2.69 | 2.96 | 3.67 | 4.57 | 5.12 | 5.43 |
| HPV16/18 | Organised | Marseille | 50 | 3.66 | 2.47 | 2.63 | 2.91 | 3.61 | 4.47 | 5.05 | 5.36 |
| HPV16/18 | Organised | Marseille | 51 | 3.59 | 2.42 | 2.58 | 2.85 | 3.54 | 4.40 | 4.93 | 5.25 |
| HPV16/18 | Organised | Marseille | 52 | 3.53 | 2.37 | 2.53 | 2.80 | 3.48 | 4.33 | 4.84 | 5.21 |
| HPV16/18 | Organised | Marseille | 53 | 3.46 | 2.32 | 2.47 | 2.74 | 3.41 | 4.25 | 4.75 | 5.08 |
| HPV16/18 | Organised | Marseille | 54 | 3.39 | 2.27 | 2.42 | 2.67 | 3.34 | 4.15 | 4.65 | 4.98 |
| HPV16/18 | Organised | Marseille | 55 | 3.33 | 2.23 | 2.38 | 2.63 | 3.28 | 4.08 | 4.60 | 4.87 |
| HPV16/18 | Organised | Marseille | 56 | 3.29 | 2.18 | 2.35 | 2.60 | 3.24 | 4.03 | 4.53 | 4.77 |
| HPV16/18 | Organised | Marseille | 57 | 3.27 | 2.17 | 2.33 | 2.57 | 3.22 | 4.02 | 4.48 | 4.75 |
| HPV16/18 | Organised | Marseille | 58 | 3.26 | 2.16 | 2.32 | 2.57 | 3.21 | 4.02 | 4.47 | 4.71 |
| HPV16/18 | Organised | Marseille | 59 | 3.27 | 2.15 | 2.32 | 2.57 | 3.22 | 4.02 | 4.48 | 4.75 |
| HPV16/18 | Organised | Marseille | 60 | 3.28 | 2.18 | 2.33 | 2.58 | 3.23 | 4.06 | 4.50 | 4.80 |
| HPV16/18 | Organised | Marseille | 61 | 3.30 | 2.20 | 2.34 | 2.59 | 3.26 | 4.09 | 4.52 | 4.84 |
| HPV16/18 | Organised | Marseille | 62 | 3.32 | 2.19 | 2.32 | 2.59 | 3.27 | 4.13 | 4.56 | 4.92 |
| HPV16/18 | Organised | Marseille | 63 | 3.32 | 2.19 | 2.31 | 2.58 | 3.27 | 4.13 | 4.58 | 4.90 |
| HPV16/18 | Organised | Marseille | 64 | 3.32 | 2.16 | 2.29 | 2.57 | 3.27 | 4.14 | 4.63 | 4.93 |
| HPV16/18 | Organised | Marseille | 65 | 3.31 | 2.13 | 2.25 | 2.53 | 3.26 | 4.16 | 4.66 | 4.96 |
| HPV16/18 | Organised | Marseille | 66 | 3.29 | 2.06 | 2.20 | 2.49 | 3.24 | 4.16 | 4.74 | 5.04 |
| HPV16/18 | Organised | Toulouse | 30 | 3.81 | 2.15 | 2.31 | 2.70 | 3.72 | 5.06 | 5.97 | 6.59 |
| HPV16/18 | Organised | Toulouse | 31 | 4.00 | 2.29 | 2.45 | 2.86 | 3.91 | 5.27 | 6.19 | 6.82 |
| HPV16/18 | Organised | Toulouse | 32 | 4.17 | 2.39 | 2.57 | 3.00 | 4.08 | 5.49 | 6.41 | 7.05 |
| HPV16/18 | Organised | Toulouse | 33 | 4.30 | 2.45 | 2.66 | 3.10 | 4.23 | 5.64 | 6.57 | 7.18 |
| HPV16/18 | Organised | Toulouse | 34 | 4.35 | 2.51 | 2.70 | 3.13 | 4.27 | 5.68 | 6.56 | 7.21 |
| HPV16/18 | Organised | Toulouse | 35 | 4.31 | 2.49 | 2.69 | 3.11 | 4.23 | 5.62 | 6.47 | 7.13 |
| HPV16/18 | Organised | Toulouse | 36 | 4.22 | 2.43 | 2.65 | 3.03 | 4.14 | 5.51 | 6.36 | 7.05 |
| HPV16/18 | Organised | Toulouse | 37 | 4.12 | 2.36 | 2.58 | 2.96 | 4.04 | 5.36 | 6.24 | 6.89 |
| HPV16/18 | Organised | Toulouse | 38 | 4.01 | 2.32 | 2.51 | 2.90 | 3.93 | 5.24 | 6.09 | 6.69 |
| HPV16/18 | Organised | Toulouse | 39 | 3.91 | 2.27 | 2.43 | 2.83 | 3.83 | 5.09 | 5.88 | 6.51 |
| HPV16/18 | Organised | Toulouse | 40 | 3.81 | 2.21 | 2.37 | 2.75 | 3.72 | 4.95 | 5.70 | 6.28 |

**Table S9 Posterior expected HR HPV cervical infection prevalence (in %) in major French cities, stratified by type of test, city, and age. Table counterpart to Figure 3A** ***(continued)***

| | | | | Posterior distribution | | | | | | | |
|---|---|---|---|---|---|---|---|---|---|---|---|
| | | | | | Percentiles | | | | | | |
| Virus | Test | City | Age | Average | 0.01 | 0.025 | 0.1 | 0.5 | 0.9 | 0.975 | 0.99 |
| HPV16/18 | Organised | Toulouse | 41 | 3.71 | 2.15 | 2.30 | 2.68 | 3.63 | 4.84 | 5.59 | 6.09 |
| HPV16/18 | Organised | Toulouse | 42 | 3.61 | 2.09 | 2.23 | 2.61 | 3.53 | 4.71 | 5.42 | 5.91 |
| HPV16/18 | Organised | Toulouse | 43 | 3.52 | 2.03 | 2.18 | 2.53 | 3.44 | 4.59 | 5.29 | 5.76 |
| HPV16/18 | Organised | Toulouse | 44 | 3.44 | 1.98 | 2.14 | 2.47 | 3.37 | 4.49 | 5.19 | 5.61 |
| HPV16/18 | Organised | Toulouse | 45 | 3.38 | 1.96 | 2.10 | 2.43 | 3.30 | 4.40 | 5.09 | 5.47 |
| HPV16/18 | Organised | Toulouse | 46 | 3.30 | 1.91 | 2.04 | 2.38 | 3.23 | 4.32 | 4.99 | 5.35 |
| HPV16/18 | Organised | Toulouse | 47 | 3.23 | 1.85 | 2.00 | 2.33 | 3.17 | 4.23 | 4.90 | 5.24 |
| HPV16/18 | Organised | Toulouse | 48 | 3.17 | 1.82 | 1.96 | 2.27 | 3.10 | 4.14 | 4.80 | 5.15 |
| HPV16/18 | Organised | Toulouse | 49 | 3.11 | 1.79 | 1.93 | 2.23 | 3.04 | 4.06 | 4.71 | 5.03 |
| HPV16/18 | Organised | Toulouse | 50 | 3.05 | 1.75 | 1.89 | 2.18 | 2.99 | 3.99 | 4.63 | 4.96 |
| HPV16/18 | Organised | Toulouse | 51 | 3.00 | 1.72 | 1.85 | 2.13 | 2.94 | 3.93 | 4.52 | 4.89 |
| HPV16/18 | Organised | Toulouse | 52 | 2.94 | 1.70 | 1.82 | 2.09 | 2.89 | 3.86 | 4.44 | 4.85 |
| HPV16/18 | Organised | Toulouse | 53 | 2.88 | 1.66 | 1.78 | 2.05 | 2.82 | 3.79 | 4.38 | 4.76 |
| HPV16/18 | Organised | Toulouse | 54 | 2.83 | 1.62 | 1.74 | 2.01 | 2.77 | 3.70 | 4.31 | 4.68 |
| HPV16/18 | Organised | Toulouse | 55 | 2.78 | 1.59 | 1.72 | 1.98 | 2.72 | 3.63 | 4.24 | 4.59 |
| HPV16/18 | Organised | Toulouse | 56 | 2.74 | 1.57 | 1.70 | 1.95 | 2.68 | 3.59 | 4.20 | 4.56 |
| HPV16/18 | Organised | Toulouse | 57 | 2.72 | 1.55 | 1.69 | 1.93 | 2.67 | 3.57 | 4.16 | 4.52 |
| HPV16/18 | Organised | Toulouse | 58 | 2.72 | 1.54 | 1.66 | 1.93 | 2.65 | 3.58 | 4.12 | 4.50 |
| HPV16/18 | Organised | Toulouse | 59 | 2.72 | 1.53 | 1.66 | 1.93 | 2.65 | 3.58 | 4.17 | 4.48 |
| HPV16/18 | Organised | Toulouse | 60 | 2.74 | 1.54 | 1.67 | 1.93 | 2.67 | 3.61 | 4.19 | 4.51 |
| HPV16/18 | Organised | Toulouse | 61 | 2.75 | 1.57 | 1.66 | 1.95 | 2.69 | 3.63 | 4.22 | 4.56 |
| HPV16/18 | Organised | Toulouse | 62 | 2.77 | 1.57 | 1.67 | 1.95 | 2.70 | 3.64 | 4.25 | 4.62 |
| HPV16/18 | Organised | Toulouse | 63 | 2.77 | 1.57 | 1.66 | 1.96 | 2.69 | 3.66 | 4.29 | 4.70 |
| HPV16/18 | Organised | Toulouse | 64 | 2.77 | 1.54 | 1.65 | 1.94 | 2.69 | 3.65 | 4.30 | 4.74 |
| HPV16/18 | Organised | Toulouse | 65 | 2.76 | 1.52 | 1.63 | 1.92 | 2.67 | 3.65 | 4.32 | 4.81 |
| HPV16/18 | Organised | Toulouse | 66 | 2.75 | 1.46 | 1.60 | 1.89 | 2.65 | 3.66 | 4.39 | 4.86 |
| HPV16/18 | Organised | Bordeaux | 30 | 4.08 | 2.62 | 2.75 | 3.14 | 4.02 | 5.10 | 5.80 | 6.15 |
| HPV16/18 | Organised | Bordeaux | 31 | 4.29 | 2.80 | 2.90 | 3.31 | 4.22 | 5.32 | 6.02 | 6.39 |
| HPV16/18 | Organised | Bordeaux | 32 | 4.47 | 2.91 | 3.06 | 3.47 | 4.41 | 5.55 | 6.22 | 6.58 |

**Table S9** **Posterior expected HR HPV cervical infection prevalence (in %) in major French cities, stratified by type of test, city, and age. Table counterpart to Figure 3A** ***(continued)***

| | | | | Posterior distribution | | | | | | | |
|---|---|---|---|---|---|---|---|---|---|---|---|
| | | | | | Percentiles | | | | | | |
| Virus | Test | City | Age | Average | 0.01 | 0.025 | 0.1 | 0.5 | 0.9 | 0.975 | 0.99 |
| HPV16/18 | Organised | Bordeaux | 33 | 4.61 | 3.02 | 3.18 | 3.60 | 4.55 | 5.71 | 6.37 | 6.74 |
| HPV16/18 | Organised | Bordeaux | 34 | 4.66 | 3.05 | 3.23 | 3.65 | 4.60 | 5.75 | 6.42 | 6.74 |
| HPV16/18 | Organised | Bordeaux | 35 | 4.61 | 3.04 | 3.20 | 3.62 | 4.56 | 5.68 | 6.35 | 6.67 |
| HPV16/18 | Organised | Bordeaux | 36 | 4.52 | 2.97 | 3.17 | 3.56 | 4.47 | 5.55 | 6.22 | 6.55 |
| HPV16/18 | Organised | Bordeaux | 37 | 4.41 | 2.89 | 3.11 | 3.47 | 4.35 | 5.41 | 6.06 | 6.39 |
| HPV16/18 | Organised | Bordeaux | 38 | 4.30 | 2.81 | 3.02 | 3.38 | 4.24 | 5.28 | 5.91 | 6.22 |
| HPV16/18 | Organised | Bordeaux | 39 | 4.19 | 2.74 | 2.96 | 3.30 | 4.13 | 5.15 | 5.77 | 6.06 |
| HPV16/18 | Organised | Bordeaux | 40 | 4.08 | 2.67 | 2.89 | 3.22 | 4.03 | 5.01 | 5.61 | 5.96 |
| HPV16/18 | Organised | Bordeaux | 41 | 3.98 | 2.61 | 2.81 | 3.14 | 3.92 | 4.89 | 5.50 | 5.83 |
| HPV16/18 | Organised | Bordeaux | 42 | 3.87 | 2.55 | 2.74 | 3.05 | 3.82 | 4.77 | 5.35 | 5.65 |
| HPV16/18 | Organised | Bordeaux | 43 | 3.77 | 2.49 | 2.66 | 2.97 | 3.72 | 4.64 | 5.19 | 5.53 |
| HPV16/18 | Organised | Bordeaux | 44 | 3.69 | 2.42 | 2.61 | 2.90 | 3.64 | 4.54 | 5.06 | 5.42 |
| HPV16/18 | Organised | Bordeaux | 45 | 3.62 | 2.37 | 2.54 | 2.84 | 3.57 | 4.45 | 4.95 | 5.32 |
| HPV16/18 | Organised | Bordeaux | 46 | 3.54 | 2.32 | 2.48 | 2.79 | 3.50 | 4.38 | 4.86 | 5.20 |
| HPV16/18 | Organised | Bordeaux | 47 | 3.47 | 2.27 | 2.44 | 2.73 | 3.42 | 4.28 | 4.76 | 5.12 |
| HPV16/18 | Organised | Bordeaux | 48 | 3.40 | 2.22 | 2.40 | 2.68 | 3.35 | 4.17 | 4.67 | 5.01 |
| HPV16/18 | Organised | Bordeaux | 49 | 3.34 | 2.18 | 2.35 | 2.63 | 3.28 | 4.10 | 4.59 | 4.92 |
| HPV16/18 | Organised | Bordeaux | 50 | 3.27 | 2.14 | 2.30 | 2.57 | 3.23 | 4.02 | 4.51 | 4.85 |
| HPV16/18 | Organised | Bordeaux | 51 | 3.21 | 2.10 | 2.25 | 2.53 | 3.17 | 3.95 | 4.45 | 4.77 |
| HPV16/18 | Organised | Bordeaux | 52 | 3.16 | 2.06 | 2.21 | 2.48 | 3.12 | 3.89 | 4.37 | 4.65 |
| HPV16/18 | Organised | Bordeaux | 53 | 3.09 | 2.03 | 2.17 | 2.42 | 3.05 | 3.81 | 4.30 | 4.58 |
| HPV16/18 | Organised | Bordeaux | 54 | 3.03 | 1.98 | 2.11 | 2.38 | 2.99 | 3.74 | 4.22 | 4.49 |
| HPV16/18 | Organised | Bordeaux | 55 | 2.98 | 1.97 | 2.07 | 2.33 | 2.94 | 3.69 | 4.16 | 4.41 |
| HPV16/18 | Organised | Bordeaux | 56 | 2.94 | 1.93 | 2.04 | 2.31 | 2.90 | 3.64 | 4.11 | 4.35 |
| HPV16/18 | Organised | Bordeaux | 57 | 2.92 | 1.89 | 2.01 | 2.29 | 2.88 | 3.60 | 4.10 | 4.35 |
| HPV16/18 | Organised | Bordeaux | 58 | 2.91 | 1.88 | 2.01 | 2.27 | 2.87 | 3.59 | 4.09 | 4.36 |
| HPV16/18 | Organised | Bordeaux | 59 | 2.92 | 1.88 | 2.01 | 2.27 | 2.88 | 3.60 | 4.12 | 4.41 |
| HPV16/18 | Organised | Bordeaux | 60 | 2.94 | 1.88 | 2.02 | 2.29 | 2.89 | 3.63 | 4.16 | 4.45 |
| HPV16/18 | Organised | Bordeaux | 61 | 2.95 | 1.90 | 2.02 | 2.30 | 2.91 | 3.66 | 4.21 | 4.49 |

**Table S9 Posterior expected HR HPV cervical infection prevalence (in %) in major French cities, stratified by type of test, city, and age. Table counterpart to Figure 3A *(continued)***

| | | | | Posterior distribution | | | | | | | |
|---|---|---|---|---|---|---|---|---|---|---|---|
| | | | | | Percentiles | | | | | | |
| Virus | Test | City | Age | Average | 0.01 | 0.025 | 0.1 | 0.5 | 0.9 | 0.975 | 0.99 |
| HPV16/18 | Organised | Bordeaux | 62 | 2.97 | 1.90 | 2.02 | 2.30 | 2.92 | 3.68 | 4.20 | 4.53 |
| HPV16/18 | Organised | Bordeaux | 63 | 2.97 | 1.90 | 2.00 | 2.29 | 2.92 | 3.69 | 4.23 | 4.55 |
| HPV16/18 | Organised | Bordeaux | 64 | 2.97 | 1.89 | 1.98 | 2.27 | 2.91 | 3.73 | 4.27 | 4.55 |
| HPV16/18 | Organised | Bordeaux | 65 | 2.96 | 1.84 | 1.97 | 2.23 | 2.91 | 3.74 | 4.28 | 4.63 |
| HPV16/18 | Organised | Bordeaux | 66 | 2.94 | 1.80 | 1.93 | 2.20 | 2.89 | 3.75 | 4.33 | 4.64 |
| HPV16/18 | Organised | Montpellier | 30 | 4.41 | 2.59 | 2.77 | 3.23 | 4.30 | 5.73 | 6.79 | 7.29 |
| HPV16/18 | Organised | Montpellier | 31 | 4.63 | 2.77 | 2.93 | 3.41 | 4.51 | 5.99 | 7.02 | 7.55 |
| HPV16/18 | Organised | Montpellier | 32 | 4.82 | 2.89 | 3.09 | 3.57 | 4.73 | 6.20 | 7.23 | 7.82 |
| HPV16/18 | Organised | Montpellier | 33 | 4.97 | 2.99 | 3.19 | 3.69 | 4.88 | 6.34 | 7.41 | 8.00 |
| HPV16/18 | Organised | Montpellier | 34 | 5.03 | 3.04 | 3.24 | 3.73 | 4.94 | 6.42 | 7.48 | 8.04 |
| HPV16/18 | Organised | Montpellier | 35 | 4.98 | 3.03 | 3.22 | 3.71 | 4.89 | 6.36 | 7.41 | 7.93 |
| HPV16/18 | Organised | Montpellier | 36 | 4.88 | 2.96 | 3.16 | 3.64 | 4.79 | 6.23 | 7.25 | 7.75 |
| HPV16/18 | Organised | Montpellier | 37 | 4.76 | 2.91 | 3.09 | 3.56 | 4.67 | 6.07 | 7.02 | 7.55 |
| HPV16/18 | Organised | Montpellier | 38 | 4.64 | 2.84 | 3.02 | 3.47 | 4.56 | 5.90 | 6.88 | 7.40 |
| HPV16/18 | Organised | Montpellier | 39 | 4.52 | 2.77 | 2.94 | 3.38 | 4.44 | 5.75 | 6.67 | 7.17 |
| HPV16/18 | Organised | Montpellier | 40 | 4.41 | 2.69 | 2.86 | 3.30 | 4.32 | 5.61 | 6.51 | 7.01 |
| HPV16/18 | Organised | Montpellier | 41 | 4.30 | 2.62 | 2.79 | 3.22 | 4.21 | 5.47 | 6.33 | 6.85 |
| HPV16/18 | Organised | Montpellier | 42 | 4.18 | 2.54 | 2.71 | 3.12 | 4.09 | 5.33 | 6.17 | 6.72 |
| HPV16/18 | Organised | Montpellier | 43 | 4.08 | 2.49 | 2.64 | 3.04 | 3.98 | 5.22 | 6.02 | 6.48 |
| HPV16/18 | Organised | Montpellier | 44 | 3.99 | 2.42 | 2.57 | 2.95 | 3.90 | 5.10 | 5.86 | 6.38 |
| HPV16/18 | Organised | Montpellier | 45 | 3.91 | 2.37 | 2.50 | 2.90 | 3.81 | 5.01 | 5.76 | 6.25 |
| HPV16/18 | Organised | Montpellier | 46 | 3.83 | 2.31 | 2.45 | 2.83 | 3.73 | 4.90 | 5.65 | 6.15 |
| HPV16/18 | Organised | Montpellier | 47 | 3.75 | 2.25 | 2.41 | 2.78 | 3.66 | 4.81 | 5.55 | 6.03 |
| HPV16/18 | Organised | Montpellier | 48 | 3.67 | 2.20 | 2.37 | 2.72 | 3.59 | 4.70 | 5.44 | 6.00 |
| HPV16/18 | Organised | Montpellier | 49 | 3.61 | 2.17 | 2.33 | 2.67 | 3.52 | 4.62 | 5.34 | 5.84 |
| HPV16/18 | Organised | Montpellier | 50 | 3.54 | 2.13 | 2.29 | 2.62 | 3.46 | 4.55 | 5.23 | 5.70 |
| HPV16/18 | Organised | Montpellier | 51 | 3.47 | 2.07 | 2.23 | 2.57 | 3.39 | 4.48 | 5.16 | 5.63 |
| HPV16/18 | Organised | Montpellier | 52 | 3.41 | 2.03 | 2.18 | 2.52 | 3.33 | 4.41 | 5.07 | 5.55 |
| HPV16/18 | Organised | Montpellier | 53 | 3.34 | 2.00 | 2.15 | 2.48 | 3.26 | 4.32 | 4.98 | 5.42 |

**Table S9** **Posterior expected HR HPV cervical infection prevalence (in %) in major French cities, stratified by type of test, city, and age. Table counterpart to Figure 3A** ***(continued)***

| | | | | Posterior distribution | | | | | | | |
|---|---|---|---|---|---|---|---|---|---|---|---|
| | | | | | Percentiles | | | | | | |
| Virus | Test | City | Age | Average | 0.01 | 0.025 | 0.1 | 0.5 | 0.9 | 0.975 | 0.99 |
| HPV16/18 | Organised | Montpellier | 54 | 3.28 | 1.95 | 2.09 | 2.43 | 3.20 | 4.22 | 4.88 | 5.32 |
| HPV16/18 | Organised | Montpellier | 55 | 3.22 | 1.90 | 2.05 | 2.38 | 3.14 | 4.15 | 4.82 | 5.25 |
| HPV16/18 | Organised | Montpellier | 56 | 3.18 | 1.86 | 2.03 | 2.34 | 3.11 | 4.11 | 4.77 | 5.21 |
| HPV16/18 | Organised | Montpellier | 57 | 3.16 | 1.83 | 2.00 | 2.34 | 3.09 | 4.08 | 4.74 | 5.15 |
| HPV16/18 | Organised | Montpellier | 58 | 3.15 | 1.84 | 2.00 | 2.32 | 3.08 | 4.07 | 4.73 | 5.17 |
| HPV16/18 | Organised | Montpellier | 59 | 3.16 | 1.85 | 1.99 | 2.32 | 3.08 | 4.09 | 4.77 | 5.17 |
| HPV16/18 | Organised | Montpellier | 60 | 3.17 | 1.85 | 2.01 | 2.33 | 3.10 | 4.12 | 4.81 | 5.17 |
| HPV16/18 | Organised | Montpellier | 61 | 3.19 | 1.88 | 2.01 | 2.34 | 3.12 | 4.14 | 4.84 | 5.21 |
| HPV16/18 | Organised | Montpellier | 62 | 3.21 | 1.88 | 1.99 | 2.36 | 3.13 | 4.17 | 4.86 | 5.25 |
| HPV16/18 | Organised | Montpellier | 63 | 3.21 | 1.85 | 1.99 | 2.36 | 3.13 | 4.18 | 4.88 | 5.27 |
| HPV16/18 | Organised | Montpellier | 64 | 3.21 | 1.83 | 1.98 | 2.35 | 3.12 | 4.21 | 4.88 | 5.31 |
| HPV16/18 | Organised | Montpellier | 65 | 3.20 | 1.81 | 1.94 | 2.31 | 3.11 | 4.23 | 4.91 | 5.31 |
| HPV16/18 | Organised | Montpellier | 66 | 3.18 | 1.77 | 1.91 | 2.29 | 3.09 | 4.23 | 4.93 | 5.32 |
| HPV16/18 | Organised | Rennes | 30 | 3.93 | 2.55 | 2.71 | 3.04 | 3.88 | 4.89 | 5.56 | 5.83 |
| HPV16/18 | Organised | Rennes | 31 | 4.12 | 2.71 | 2.86 | 3.22 | 4.08 | 5.11 | 5.74 | 6.03 |
| HPV16/18 | Organised | Rennes | 32 | 4.30 | 2.85 | 3.00 | 3.37 | 4.25 | 5.30 | 5.92 | 6.20 |
| HPV16/18 | Organised | Rennes | 33 | 4.43 | 2.94 | 3.11 | 3.51 | 4.39 | 5.48 | 6.04 | 6.38 |
| HPV16/18 | Organised | Rennes | 34 | 4.48 | 3.00 | 3.16 | 3.55 | 4.44 | 5.52 | 6.09 | 6.42 |
| HPV16/18 | Organised | Rennes | 35 | 4.44 | 2.98 | 3.14 | 3.52 | 4.39 | 5.44 | 6.04 | 6.31 |
| HPV16/18 | Organised | Rennes | 36 | 4.35 | 2.94 | 3.09 | 3.46 | 4.30 | 5.32 | 5.90 | 6.17 |
| HPV16/18 | Organised | Rennes | 37 | 4.24 | 2.87 | 3.03 | 3.37 | 4.19 | 5.18 | 5.76 | 6.05 |
| HPV16/18 | Organised | Rennes | 38 | 4.14 | 2.80 | 2.96 | 3.29 | 4.08 | 5.05 | 5.61 | 5.91 |
| HPV16/18 | Organised | Rennes | 39 | 4.03 | 2.75 | 2.89 | 3.20 | 3.98 | 4.92 | 5.48 | 5.78 |
| HPV16/18 | Organised | Rennes | 40 | 3.93 | 2.67 | 2.82 | 3.12 | 3.88 | 4.80 | 5.36 | 5.69 |
| HPV16/18 | Organised | Rennes | 41 | 3.83 | 2.62 | 2.74 | 3.05 | 3.78 | 4.67 | 5.25 | 5.55 |
| HPV16/18 | Organised | Rennes | 42 | 3.72 | 2.53 | 2.67 | 2.97 | 3.68 | 4.56 | 5.13 | 5.41 |
| HPV16/18 | Organised | Rennes | 43 | 3.63 | 2.47 | 2.60 | 2.89 | 3.58 | 4.44 | 5.00 | 5.29 |
| HPV16/18 | Organised | Rennes | 44 | 3.55 | 2.42 | 2.53 | 2.82 | 3.50 | 4.34 | 4.87 | 5.21 |
| HPV16/18 | Organised | Rennes | 45 | 3.48 | 2.36 | 2.48 | 2.76 | 3.43 | 4.25 | 4.78 | 5.11 |

**Table S9 Posterior expected HR HPV cervical infection prevalence (in %) in major French cities, stratified by type of test, city, and age. Table counterpart to Figure 3A** ***(continued)***

| | | | | Posterior distribution | | | | | | | |
|---|---|---|---|---|---|---|---|---|---|---|---|
| | | | | | Percentiles | | | | | | |
| Virus | Test | City | Age | Average | 0.01 | 0.025 | 0.1 | 0.5 | 0.9 | 0.975 | 0.99 |
| HPV16/18 | Organised | Rennes | 46 | 3.41 | 2.31 | 2.44 | 2.71 | 3.36 | 4.16 | 4.68 | 4.99 |
| HPV16/18 | Organised | Rennes | 47 | 3.34 | 2.26 | 2.39 | 2.65 | 3.29 | 4.08 | 4.58 | 4.85 |
| HPV16/18 | Organised | Rennes | 48 | 3.27 | 2.21 | 2.34 | 2.59 | 3.23 | 4.01 | 4.49 | 4.78 |
| HPV16/18 | Organised | Rennes | 49 | 3.21 | 2.16 | 2.30 | 2.54 | 3.17 | 3.93 | 4.42 | 4.70 |
| HPV16/18 | Organised | Rennes | 50 | 3.15 | 2.13 | 2.25 | 2.49 | 3.10 | 3.86 | 4.36 | 4.64 |
| HPV16/18 | Organised | Rennes | 51 | 3.09 | 2.07 | 2.21 | 2.44 | 3.05 | 3.80 | 4.26 | 4.53 |
| HPV16/18 | Organised | Rennes | 52 | 3.04 | 2.01 | 2.16 | 2.39 | 2.99 | 3.73 | 4.20 | 4.43 |
| HPV16/18 | Organised | Rennes | 53 | 2.98 | 1.97 | 2.11 | 2.34 | 2.93 | 3.66 | 4.12 | 4.35 |
| HPV16/18 | Organised | Rennes | 54 | 2.91 | 1.94 | 2.06 | 2.28 | 2.87 | 3.58 | 4.02 | 4.30 |
| HPV16/18 | Organised | Rennes | 55 | 2.86 | 1.91 | 2.03 | 2.25 | 2.82 | 3.52 | 3.95 | 4.26 |
| HPV16/18 | Organised | Rennes | 56 | 2.83 | 1.90 | 2.00 | 2.22 | 2.79 | 3.50 | 3.90 | 4.21 |
| HPV16/18 | Organised | Rennes | 57 | 2.81 | 1.87 | 1.98 | 2.20 | 2.78 | 3.48 | 3.89 | 4.18 |
| HPV16/18 | Organised | Rennes | 58 | 2.80 | 1.85 | 1.96 | 2.19 | 2.77 | 3.46 | 3.89 | 4.17 |
| HPV16/18 | Organised | Rennes | 59 | 2.81 | 1.85 | 1.96 | 2.18 | 2.78 | 3.48 | 3.92 | 4.17 |
| HPV16/18 | Organised | Rennes | 60 | 2.82 | 1.85 | 1.97 | 2.19 | 2.79 | 3.50 | 3.92 | 4.18 |
| HPV16/18 | Organised | Rennes | 61 | 2.84 | 1.86 | 1.97 | 2.21 | 2.81 | 3.52 | 3.96 | 4.21 |
| HPV16/18 | Organised | Rennes | 62 | 2.85 | 1.86 | 1.96 | 2.22 | 2.82 | 3.54 | 3.99 | 4.24 |
| HPV16/18 | Organised | Rennes | 63 | 2.86 | 1.85 | 1.95 | 2.21 | 2.82 | 3.56 | 4.00 | 4.27 |
| HPV16/18 | Organised | Rennes | 64 | 2.86 | 1.82 | 1.95 | 2.20 | 2.82 | 3.57 | 4.03 | 4.27 |
| HPV16/18 | Organised | Rennes | 65 | 2.85 | 1.78 | 1.92 | 2.18 | 2.81 | 3.58 | 4.03 | 4.29 |
| HPV16/18 | Organised | Rennes | 66 | 2.83 | 1.74 | 1.89 | 2.14 | 2.79 | 3.60 | 4.07 | 4.32 |
| HPV16/18 | Organised | Nantes | 30 | 3.62 | 2.01 | 2.16 | 2.56 | 3.52 | 4.79 | 5.60 | 6.19 |
| HPV16/18 | Organised | Nantes | 31 | 3.80 | 2.14 | 2.29 | 2.70 | 3.70 | 5.01 | 5.83 | 6.37 |
| HPV16/18 | Organised | Nantes | 32 | 3.96 | 2.23 | 2.41 | 2.83 | 3.85 | 5.20 | 6.06 | 6.60 |
| HPV16/18 | Organised | Nantes | 33 | 4.08 | 2.32 | 2.50 | 2.94 | 3.97 | 5.34 | 6.25 | 6.77 |
| HPV16/18 | Organised | Nantes | 34 | 4.13 | 2.35 | 2.54 | 2.99 | 4.03 | 5.42 | 6.24 | 6.80 |
| HPV16/18 | Organised | Nantes | 35 | 4.09 | 2.32 | 2.51 | 2.96 | 3.98 | 5.38 | 6.21 | 6.69 |
| HPV16/18 | Organised | Nantes | 36 | 4.01 | 2.29 | 2.49 | 2.91 | 3.90 | 5.29 | 6.11 | 6.55 |
| HPV16/18 | Organised | Nantes | 37 | 3.91 | 2.23 | 2.43 | 2.84 | 3.80 | 5.14 | 5.94 | 6.39 |

**Table S9** **Posterior expected HR HPV cervical infection prevalence (in %) in major French cities, stratified by type of test, city, and age. Table counterpart to Figure 3A** ***(continued)***

| | | | | Posterior distribution | | | | | | | |
|---|---|---|---|---|---|---|---|---|---|---|---|
| | | | | | Percentiles | | | | | | |
| Virus | Test | City | Age | Average | 0.01 | 0.025 | 0.1 | 0.5 | 0.9 | 0.975 | 0.99 |
| HPV16/18 | Organised | Nantes | 38 | 3.81 | 2.18 | 2.38 | 2.77 | 3.71 | 5.02 | 5.80 | 6.17 |
| HPV16/18 | Organised | Nantes | 39 | 3.71 | 2.14 | 2.32 | 2.71 | 3.62 | 4.90 | 5.59 | 6.01 |
| HPV16/18 | Organised | Nantes | 40 | 3.61 | 2.07 | 2.26 | 2.63 | 3.52 | 4.78 | 5.42 | 5.85 |
| HPV16/18 | Organised | Nantes | 41 | 3.52 | 2.01 | 2.19 | 2.57 | 3.43 | 4.66 | 5.26 | 5.65 |
| HPV16/18 | Organised | Nantes | 42 | 3.43 | 1.96 | 2.11 | 2.49 | 3.33 | 4.53 | 5.14 | 5.48 |
| HPV16/18 | Organised | Nantes | 43 | 3.34 | 1.91 | 2.07 | 2.43 | 3.25 | 4.41 | 5.00 | 5.34 |
| HPV16/18 | Organised | Nantes | 44 | 3.26 | 1.86 | 2.03 | 2.37 | 3.18 | 4.30 | 4.90 | 5.26 |
| HPV16/18 | Organised | Nantes | 45 | 3.20 | 1.83 | 1.99 | 2.33 | 3.12 | 4.22 | 4.84 | 5.19 |
| HPV16/18 | Organised | Nantes | 46 | 3.13 | 1.80 | 1.96 | 2.28 | 3.06 | 4.12 | 4.70 | 5.08 |
| HPV16/18 | Organised | Nantes | 47 | 3.07 | 1.74 | 1.92 | 2.24 | 2.99 | 4.04 | 4.62 | 5.01 |
| HPV16/18 | Organised | Nantes | 48 | 3.01 | 1.70 | 1.88 | 2.19 | 2.93 | 3.95 | 4.54 | 4.94 |
| HPV16/18 | Organised | Nantes | 49 | 2.95 | 1.67 | 1.85 | 2.15 | 2.87 | 3.88 | 4.45 | 4.86 |
| HPV16/18 | Organised | Nantes | 50 | 2.89 | 1.64 | 1.81 | 2.11 | 2.81 | 3.81 | 4.36 | 4.81 |
| HPV16/18 | Organised | Nantes | 51 | 2.84 | 1.60 | 1.78 | 2.07 | 2.76 | 3.73 | 4.29 | 4.71 |
| HPV16/18 | Organised | Nantes | 52 | 2.79 | 1.57 | 1.75 | 2.04 | 2.71 | 3.68 | 4.23 | 4.59 |
| HPV16/18 | Organised | Nantes | 53 | 2.73 | 1.54 | 1.71 | 1.99 | 2.66 | 3.58 | 4.14 | 4.50 |
| HPV16/18 | Organised | Nantes | 54 | 2.68 | 1.52 | 1.67 | 1.95 | 2.60 | 3.51 | 4.08 | 4.41 |
| HPV16/18 | Organised | Nantes | 55 | 2.63 | 1.49 | 1.64 | 1.91 | 2.56 | 3.46 | 4.00 | 4.36 |
| HPV16/18 | Organised | Nantes | 56 | 2.60 | 1.47 | 1.61 | 1.89 | 2.53 | 3.42 | 3.97 | 4.33 |
| HPV16/18 | Organised | Nantes | 57 | 2.58 | 1.46 | 1.59 | 1.87 | 2.51 | 3.40 | 3.93 | 4.29 |
| HPV16/18 | Organised | Nantes | 58 | 2.58 | 1.44 | 1.59 | 1.86 | 2.51 | 3.40 | 3.95 | 4.25 |
| HPV16/18 | Organised | Nantes | 59 | 2.58 | 1.45 | 1.58 | 1.87 | 2.51 | 3.42 | 3.97 | 4.24 |
| HPV16/18 | Organised | Nantes | 60 | 2.59 | 1.45 | 1.58 | 1.88 | 2.53 | 3.43 | 3.99 | 4.29 |
| HPV16/18 | Organised | Nantes | 61 | 2.61 | 1.46 | 1.58 | 1.89 | 2.54 | 3.45 | 4.02 | 4.33 |
| HPV16/18 | Organised | Nantes | 62 | 2.62 | 1.46 | 1.59 | 1.89 | 2.55 | 3.47 | 4.05 | 4.35 |
| HPV16/18 | Organised | Nantes | 63 | 2.63 | 1.47 | 1.59 | 1.89 | 2.55 | 3.47 | 4.05 | 4.41 |
| HPV16/18 | Organised | Nantes | 64 | 2.62 | 1.44 | 1.58 | 1.87 | 2.54 | 3.48 | 4.07 | 4.42 |
| HPV16/18 | Organised | Nantes | 65 | 2.62 | 1.42 | 1.55 | 1.86 | 2.53 | 3.48 | 4.12 | 4.53 |
| HPV16/18 | Organised | Nantes | 66 | 2.60 | 1.41 | 1.52 | 1.82 | 2.52 | 3.50 | 4.17 | 4.60 |

**Table S9 Posterior expected HR HPV cervical infection prevalence (in %) in major French cities, stratified by type of test, city, and age. Table counterpart to Figure 3A** ***(continued)***

| | | | | Posterior distribution | | | | | | | |
|---|---|---|---|---|---|---|---|---|---|---|---|
| | | | | | Percentiles | | | | | | |
| Virus | Test | City | Age | Average | 0.01 | 0.025 | 0.1 | 0.5 | 0.9 | 0.975 | 0.99 |
| HPV16/18 | Organised | Lille | 30 | 3.95 | 2.31 | 2.50 | 2.92 | 3.86 | 5.10 | 5.92 | 6.39 |
| HPV16/18 | Organised | Lille | 31 | 4.14 | 2.47 | 2.65 | 3.08 | 4.06 | 5.32 | 6.15 | 6.61 |
| HPV16/18 | Organised | Lille | 32 | 4.32 | 2.57 | 2.80 | 3.22 | 4.23 | 5.55 | 6.38 | 6.80 |
| HPV16/18 | Organised | Lille | 33 | 4.45 | 2.65 | 2.91 | 3.33 | 4.36 | 5.70 | 6.58 | 6.99 |
| HPV16/18 | Organised | Lille | 34 | 4.50 | 2.67 | 2.95 | 3.36 | 4.41 | 5.76 | 6.57 | 7.03 |
| HPV16/18 | Organised | Lille | 35 | 4.46 | 2.68 | 2.92 | 3.34 | 4.37 | 5.72 | 6.50 | 6.91 |
| HPV16/18 | Organised | Lille | 36 | 4.37 | 2.64 | 2.89 | 3.29 | 4.28 | 5.59 | 6.37 | 6.78 |
| HPV16/18 | Organised | Lille | 37 | 4.26 | 2.57 | 2.83 | 3.21 | 4.18 | 5.44 | 6.17 | 6.62 |
| HPV16/18 | Organised | Lille | 38 | 4.15 | 2.53 | 2.74 | 3.13 | 4.07 | 5.30 | 6.01 | 6.48 |
| HPV16/18 | Organised | Lille | 39 | 4.05 | 2.47 | 2.68 | 3.05 | 3.97 | 5.16 | 5.84 | 6.31 |
| HPV16/18 | Organised | Lille | 40 | 3.95 | 2.39 | 2.61 | 2.98 | 3.86 | 5.03 | 5.70 | 6.13 |
| HPV16/18 | Organised | Lille | 41 | 3.85 | 2.34 | 2.54 | 2.90 | 3.77 | 4.90 | 5.60 | 6.06 |
| HPV16/18 | Organised | Lille | 42 | 3.74 | 2.28 | 2.47 | 2.81 | 3.66 | 4.77 | 5.42 | 5.88 |
| HPV16/18 | Organised | Lille | 43 | 3.65 | 2.22 | 2.41 | 2.74 | 3.57 | 4.66 | 5.30 | 5.77 |
| HPV16/18 | Organised | Lille | 44 | 3.57 | 2.18 | 2.35 | 2.67 | 3.49 | 4.55 | 5.17 | 5.65 |
| HPV16/18 | Organised | Lille | 45 | 3.50 | 2.14 | 2.30 | 2.63 | 3.43 | 4.46 | 5.07 | 5.53 |
| HPV16/18 | Organised | Lille | 46 | 3.42 | 2.08 | 2.26 | 2.58 | 3.36 | 4.38 | 4.95 | 5.39 |
| HPV16/18 | Organised | Lille | 47 | 3.35 | 2.02 | 2.19 | 2.52 | 3.29 | 4.29 | 4.84 | 5.32 |
| HPV16/18 | Organised | Lille | 48 | 3.29 | 1.97 | 2.14 | 2.47 | 3.22 | 4.22 | 4.76 | 5.20 |
| HPV16/18 | Organised | Lille | 49 | 3.22 | 1.95 | 2.10 | 2.43 | 3.16 | 4.13 | 4.67 | 5.10 |
| HPV16/18 | Organised | Lille | 50 | 3.16 | 1.91 | 2.05 | 2.38 | 3.09 | 4.06 | 4.59 | 5.02 |
| HPV16/18 | Organised | Lille | 51 | 3.11 | 1.86 | 2.00 | 2.33 | 3.04 | 3.99 | 4.50 | 4.88 |
| HPV16/18 | Organised | Lille | 52 | 3.05 | 1.84 | 1.96 | 2.29 | 2.98 | 3.91 | 4.42 | 4.79 |
| HPV16/18 | Organised | Lille | 53 | 2.99 | 1.80 | 1.93 | 2.23 | 2.92 | 3.84 | 4.34 | 4.67 |
| HPV16/18 | Organised | Lille | 54 | 2.93 | 1.77 | 1.89 | 2.18 | 2.86 | 3.76 | 4.26 | 4.60 |
| HPV16/18 | Organised | Lille | 55 | 2.88 | 1.74 | 1.86 | 2.14 | 2.81 | 3.70 | 4.20 | 4.53 |
| HPV16/18 | Organised | Lille | 56 | 2.84 | 1.69 | 1.85 | 2.11 | 2.78 | 3.65 | 4.17 | 4.49 |
| HPV16/18 | Organised | Lille | 57 | 2.82 | 1.66 | 1.83 | 2.10 | 2.76 | 3.62 | 4.16 | 4.44 |
| HPV16/18 | Organised | Lille | 58 | 2.82 | 1.65 | 1.82 | 2.09 | 2.76 | 3.63 | 4.13 | 4.46 |

**Table S9** **Posterior expected HR HPV cervical infection prevalence (in %) in major French cities, stratified by type of test, city, and age. Table counterpart to Figure 3A** ***(continued)***

| | | | | Posterior distribution | | | | | | | |
|---|---|---|---|---|---|---|---|---|---|---|---|
| | | | | | Percentiles | | | | | | |
| Virus | Test | City | Age | Average | 0.01 | 0.025 | 0.1 | 0.5 | 0.9 | 0.975 | 0.99 |
| HPV16/18 | Organised | Lille | 59 | 2.82 | 1.64 | 1.82 | 2.09 | 2.77 | 3.64 | 4.14 | 4.48 |
| HPV16/18 | Organised | Lille | 60 | 2.84 | 1.64 | 1.81 | 2.10 | 2.78 | 3.66 | 4.14 | 4.51 |
| HPV16/18 | Organised | Lille | 61 | 2.86 | 1.64 | 1.81 | 2.11 | 2.80 | 3.70 | 4.17 | 4.56 |
| HPV16/18 | Organised | Lille | 62 | 2.87 | 1.67 | 1.81 | 2.12 | 2.81 | 3.71 | 4.22 | 4.59 |
| HPV16/18 | Organised | Lille | 63 | 2.87 | 1.66 | 1.81 | 2.12 | 2.81 | 3.73 | 4.26 | 4.65 |
| HPV16/18 | Organised | Lille | 64 | 2.87 | 1.62 | 1.80 | 2.10 | 2.80 | 3.74 | 4.29 | 4.68 |
| HPV16/18 | Organised | Lille | 65 | 2.86 | 1.61 | 1.76 | 2.09 | 2.79 | 3.74 | 4.30 | 4.70 |
| HPV16/18 | Organised | Lille | 66 | 2.85 | 1.60 | 1.72 | 2.05 | 2.77 | 3.76 | 4.36 | 4.74 |
| HPV16/18 | Organised | Strasbourg | 30 | 3.23 | 1.71 | 1.88 | 2.26 | 3.13 | 4.35 | 5.24 | 5.76 |
| HPV16/18 | Organised | Strasbourg | 31 | 3.39 | 1.82 | 1.99 | 2.38 | 3.27 | 4.56 | 5.46 | 5.99 |
| HPV16/18 | Organised | Strasbourg | 32 | 3.54 | 1.90 | 2.08 | 2.49 | 3.42 | 4.72 | 5.71 | 6.24 |
| HPV16/18 | Organised | Strasbourg | 33 | 3.65 | 2.00 | 2.15 | 2.56 | 3.54 | 4.88 | 5.81 | 6.45 |
| HPV16/18 | Organised | Strasbourg | 34 | 3.69 | 2.02 | 2.18 | 2.60 | 3.57 | 4.93 | 5.85 | 6.48 |
| HPV16/18 | Organised | Strasbourg | 35 | 3.66 | 2.02 | 2.17 | 2.58 | 3.53 | 4.86 | 5.79 | 6.46 |
| HPV16/18 | Organised | Strasbourg | 36 | 3.58 | 1.97 | 2.13 | 2.53 | 3.47 | 4.77 | 5.64 | 6.33 |
| HPV16/18 | Organised | Strasbourg | 37 | 3.49 | 1.92 | 2.09 | 2.47 | 3.38 | 4.65 | 5.49 | 6.15 |
| HPV16/18 | Organised | Strasbourg | 38 | 3.40 | 1.87 | 2.04 | 2.41 | 3.29 | 4.53 | 5.35 | 6.01 |
| HPV16/18 | Organised | Strasbourg | 39 | 3.32 | 1.82 | 1.98 | 2.34 | 3.20 | 4.41 | 5.22 | 5.89 |
| HPV16/18 | Organised | Strasbourg | 40 | 3.23 | 1.77 | 1.93 | 2.29 | 3.12 | 4.31 | 5.09 | 5.75 |
| HPV16/18 | Organised | Strasbourg | 41 | 3.15 | 1.72 | 1.87 | 2.23 | 3.04 | 4.21 | 4.97 | 5.63 |
| HPV16/18 | Organised | Strasbourg | 42 | 3.06 | 1.69 | 1.82 | 2.16 | 2.96 | 4.09 | 4.85 | 5.42 |
| HPV16/18 | Organised | Strasbourg | 43 | 2.98 | 1.65 | 1.78 | 2.10 | 2.88 | 3.98 | 4.72 | 5.33 |
| HPV16/18 | Organised | Strasbourg | 44 | 2.92 | 1.61 | 1.74 | 2.06 | 2.82 | 3.89 | 4.62 | 5.20 |
| HPV16/18 | Organised | Strasbourg | 45 | 2.86 | 1.57 | 1.70 | 2.03 | 2.77 | 3.81 | 4.55 | 5.07 |
| HPV16/18 | Organised | Strasbourg | 46 | 2.80 | 1.54 | 1.67 | 1.98 | 2.71 | 3.74 | 4.45 | 4.96 |
| HPV16/18 | Organised | Strasbourg | 47 | 2.74 | 1.50 | 1.64 | 1.94 | 2.65 | 3.67 | 4.34 | 4.86 |
| HPV16/18 | Organised | Strasbourg | 48 | 2.69 | 1.47 | 1.61 | 1.89 | 2.60 | 3.61 | 4.27 | 4.76 |
| HPV16/18 | Organised | Strasbourg | 49 | 2.64 | 1.44 | 1.58 | 1.85 | 2.55 | 3.54 | 4.23 | 4.67 |
| HPV16/18 | Organised | Strasbourg | 50 | 2.59 | 1.40 | 1.55 | 1.82 | 2.50 | 3.46 | 4.15 | 4.60 |

**Table S9** **Posterior expected HR HPV cervical infection prevalence (in %) in major French cities, stratified by type of test, city, and age. Table counterpart to Figure 3A** ***(continued)***

| | | | | Posterior distribution | | | | | | | |
|---|---|---|---|---|---|---|---|---|---|---|---|
| | | | | | Percentiles | | | | | | |
| Virus | Test | City | Age | Average | 0.01 | 0.025 | 0.1 | 0.5 | 0.9 | 0.975 | 0.99 |
| HPV16/18 | Organised | Strasbourg | 51 | 2.54 | 1.38 | 1.52 | 1.78 | 2.46 | 3.41 | 4.11 | 4.53 |
| HPV16/18 | Organised | Strasbourg | 52 | 2.49 | 1.34 | 1.49 | 1.75 | 2.42 | 3.34 | 4.05 | 4.48 |
| HPV16/18 | Organised | Strasbourg | 53 | 2.44 | 1.31 | 1.46 | 1.71 | 2.37 | 3.28 | 3.97 | 4.33 |
| HPV16/18 | Organised | Strasbourg | 54 | 2.39 | 1.29 | 1.43 | 1.68 | 2.32 | 3.21 | 3.89 | 4.21 |
| HPV16/18 | Organised | Strasbourg | 55 | 2.35 | 1.26 | 1.40 | 1.65 | 2.28 | 3.15 | 3.82 | 4.11 |
| HPV16/18 | Organised | Strasbourg | 56 | 2.32 | 1.26 | 1.38 | 1.62 | 2.25 | 3.12 | 3.75 | 4.06 |
| HPV16/18 | Organised | Strasbourg | 57 | 2.31 | 1.25 | 1.36 | 1.61 | 2.23 | 3.11 | 3.72 | 4.06 |
| HPV16/18 | Organised | Strasbourg | 58 | 2.30 | 1.25 | 1.35 | 1.61 | 2.22 | 3.10 | 3.72 | 4.05 |
| HPV16/18 | Organised | Strasbourg | 59 | 2.31 | 1.25 | 1.35 | 1.61 | 2.23 | 3.11 | 3.75 | 4.05 |
| HPV16/18 | Organised | Strasbourg | 60 | 2.32 | 1.26 | 1.36 | 1.62 | 2.25 | 3.14 | 3.75 | 4.07 |
| HPV16/18 | Organised | Strasbourg | 61 | 2.33 | 1.27 | 1.37 | 1.63 | 2.26 | 3.15 | 3.78 | 4.13 |
| HPV16/18 | Organised | Strasbourg | 62 | 2.34 | 1.27 | 1.37 | 1.63 | 2.27 | 3.18 | 3.81 | 4.19 |
| HPV16/18 | Organised | Strasbourg | 63 | 2.35 | 1.26 | 1.37 | 1.62 | 2.27 | 3.20 | 3.83 | 4.19 |
| HPV16/18 | Organised | Strasbourg | 64 | 2.34 | 1.24 | 1.36 | 1.61 | 2.26 | 3.21 | 3.84 | 4.28 |
| HPV16/18 | Organised | Strasbourg | 65 | 2.34 | 1.22 | 1.35 | 1.59 | 2.25 | 3.21 | 3.85 | 4.30 |
| HPV16/18 | Organised | Strasbourg | 66 | 2.33 | 1.18 | 1.31 | 1.57 | 2.23 | 3.20 | 3.86 | 4.30 |
| HPV16/18 | Organised | Lyon | 30 | 4.47 | 2.60 | 2.77 | 3.24 | 4.34 | 5.86 | 6.76 | 7.32 |
| HPV16/18 | Organised | Lyon | 31 | 4.69 | 2.73 | 2.96 | 3.43 | 4.56 | 6.11 | 7.05 | 7.65 |
| HPV16/18 | Organised | Lyon | 32 | 4.89 | 2.87 | 3.10 | 3.58 | 4.76 | 6.36 | 7.28 | 7.90 |
| HPV16/18 | Organised | Lyon | 33 | 5.04 | 2.95 | 3.22 | 3.72 | 4.91 | 6.56 | 7.46 | 8.05 |
| HPV16/18 | Organised | Lyon | 34 | 5.10 | 2.99 | 3.26 | 3.76 | 4.97 | 6.63 | 7.54 | 8.06 |
| HPV16/18 | Organised | Lyon | 35 | 5.05 | 2.96 | 3.24 | 3.73 | 4.91 | 6.56 | 7.47 | 7.97 |
| HPV16/18 | Organised | Lyon | 36 | 4.95 | 2.94 | 3.18 | 3.66 | 4.83 | 6.44 | 7.34 | 7.79 |
| HPV16/18 | Organised | Lyon | 37 | 4.83 | 2.84 | 3.11 | 3.57 | 4.72 | 6.27 | 7.19 | 7.60 |
| HPV16/18 | Organised | Lyon | 38 | 4.71 | 2.76 | 3.04 | 3.48 | 4.59 | 6.10 | 7.01 | 7.40 |
| HPV16/18 | Organised | Lyon | 39 | 4.59 | 2.72 | 2.97 | 3.39 | 4.48 | 5.94 | 6.84 | 7.17 |
| HPV16/18 | Organised | Lyon | 40 | 4.47 | 2.65 | 2.88 | 3.31 | 4.36 | 5.78 | 6.66 | 7.04 |
| HPV16/18 | Organised | Lyon | 41 | 4.36 | 2.58 | 2.80 | 3.24 | 4.25 | 5.63 | 6.47 | 6.86 |
| HPV16/18 | Organised | Lyon | 42 | 4.24 | 2.53 | 2.72 | 3.15 | 4.14 | 5.47 | 6.28 | 6.70 |

**Table S9 Posterior expected HR HPV cervical infection prevalence (in %) in major French cities, stratified by type of test, city, and age. Table counterpart to Figure 3A *(continued)***

| | | | | Posterior distribution | | | | | | | |
|---|---|---|---|---|---|---|---|---|---|---|---|
| | | | | | Percentiles | | | | | | |
| Virus | Test | City | Age | Average | 0.01 | 0.025 | 0.1 | 0.5 | 0.9 | 0.975 | 0.99 |
| HPV16/18 | Organised | Lyon | 43 | 4.13 | 2.45 | 2.66 | 3.06 | 4.04 | 5.33 | 6.11 | 6.55 |
| HPV16/18 | Organised | Lyon | 44 | 4.04 | 2.38 | 2.59 | 2.99 | 3.95 | 5.20 | 5.96 | 6.42 |
| HPV16/18 | Organised | Lyon | 45 | 3.97 | 2.35 | 2.56 | 2.93 | 3.87 | 5.11 | 5.86 | 6.30 |
| HPV16/18 | Organised | Lyon | 46 | 3.88 | 2.30 | 2.50 | 2.87 | 3.79 | 5.01 | 5.77 | 6.15 |
| HPV16/18 | Organised | Lyon | 47 | 3.80 | 2.26 | 2.44 | 2.80 | 3.71 | 4.91 | 5.68 | 6.07 |
| HPV16/18 | Organised | Lyon | 48 | 3.73 | 2.22 | 2.40 | 2.74 | 3.64 | 4.81 | 5.59 | 5.96 |
| HPV16/18 | Organised | Lyon | 49 | 3.66 | 2.20 | 2.35 | 2.69 | 3.57 | 4.73 | 5.50 | 5.85 |
| HPV16/18 | Organised | Lyon | 50 | 3.59 | 2.14 | 2.29 | 2.64 | 3.50 | 4.65 | 5.42 | 5.77 |
| HPV16/18 | Organised | Lyon | 51 | 3.52 | 2.10 | 2.25 | 2.59 | 3.44 | 4.55 | 5.31 | 5.71 |
| HPV16/18 | Organised | Lyon | 52 | 3.46 | 2.05 | 2.22 | 2.54 | 3.38 | 4.49 | 5.23 | 5.59 |
| HPV16/18 | Organised | Lyon | 53 | 3.39 | 2.02 | 2.17 | 2.49 | 3.31 | 4.40 | 5.13 | 5.48 |
| HPV16/18 | Organised | Lyon | 54 | 3.32 | 1.97 | 2.12 | 2.43 | 3.23 | 4.31 | 5.01 | 5.37 |
| HPV16/18 | Organised | Lyon | 55 | 3.27 | 1.94 | 2.08 | 2.40 | 3.18 | 4.23 | 4.93 | 5.31 |
| HPV16/18 | Organised | Lyon | 56 | 3.23 | 1.90 | 2.04 | 2.36 | 3.14 | 4.18 | 4.87 | 5.24 |
| HPV16/18 | Organised | Lyon | 57 | 3.20 | 1.88 | 2.03 | 2.34 | 3.12 | 4.15 | 4.84 | 5.21 |
| HPV16/18 | Organised | Lyon | 58 | 3.20 | 1.87 | 2.03 | 2.32 | 3.11 | 4.15 | 4.81 | 5.20 |
| HPV16/18 | Organised | Lyon | 59 | 3.20 | 1.86 | 2.02 | 2.33 | 3.12 | 4.17 | 4.83 | 5.23 |
| HPV16/18 | Organised | Lyon | 60 | 3.22 | 1.86 | 2.02 | 2.34 | 3.14 | 4.22 | 4.85 | 5.27 |
| HPV16/18 | Organised | Lyon | 61 | 3.24 | 1.87 | 2.03 | 2.34 | 3.15 | 4.26 | 4.90 | 5.31 |
| HPV16/18 | Organised | Lyon | 62 | 3.26 | 1.86 | 2.03 | 2.34 | 3.17 | 4.26 | 4.90 | 5.37 |
| HPV16/18 | Organised | Lyon | 63 | 3.26 | 1.87 | 2.03 | 2.34 | 3.17 | 4.28 | 4.95 | 5.40 |
| HPV16/18 | Organised | Lyon | 64 | 3.26 | 1.88 | 2.01 | 2.32 | 3.15 | 4.30 | 4.98 | 5.44 |
| HPV16/18 | Organised | Lyon | 65 | 3.25 | 1.87 | 1.97 | 2.30 | 3.15 | 4.33 | 5.00 | 5.51 |
| HPV16/18 | Organised | Lyon | 66 | 3.23 | 1.81 | 1.94 | 2.27 | 3.13 | 4.35 | 5.05 | 5.52 |
| HPV16/18 | Organised | Paris | 30 | 4.74 | 3.11 | 3.33 | 3.75 | 4.66 | 5.79 | 6.59 | 6.95 |
| HPV16/18 | Organised | Paris | 31 | 4.97 | 3.31 | 3.55 | 3.96 | 4.91 | 6.04 | 6.81 | 7.22 |
| HPV16/18 | Organised | Paris | 32 | 5.18 | 3.50 | 3.72 | 4.15 | 5.13 | 6.29 | 7.08 | 7.52 |
| HPV16/18 | Organised | Paris | 33 | 5.34 | 3.67 | 3.84 | 4.29 | 5.29 | 6.45 | 7.20 | 7.67 |
| HPV16/18 | Organised | Paris | 34 | 5.40 | 3.73 | 3.89 | 4.34 | 5.35 | 6.51 | 7.21 | 7.70 |

**Table S9 Posterior expected HR HPV cervical infection prevalence (in %) in major French cities, stratified by type of test, city, and age. Table counterpart to Figure 3A *(continued)***

| | | | | Posterior distribution | | | | | | | |
|---|---|---|---|---|---|---|---|---|---|---|---|
| | | | | | Percentiles | | | | | | |
| Virus | Test | City | Age | Average | 0.01 | 0.025 | 0.1 | 0.5 | 0.9 | 0.975 | 0.99 |
| HPV16/18 | Organised | Paris | 35 | 5.35 | 3.71 | 3.88 | 4.29 | 5.30 | 6.43 | 7.11 | 7.62 |
| HPV16/18 | Organised | Paris | 36 | 5.24 | 3.63 | 3.83 | 4.22 | 5.20 | 6.30 | 6.93 | 7.45 |
| HPV16/18 | Organised | Paris | 37 | 5.11 | 3.56 | 3.76 | 4.13 | 5.07 | 6.15 | 6.75 | 7.23 |
| HPV16/18 | Organised | Paris | 38 | 4.98 | 3.51 | 3.68 | 4.03 | 4.94 | 6.00 | 6.59 | 7.05 |
| HPV16/18 | Organised | Paris | 39 | 4.86 | 3.43 | 3.59 | 3.94 | 4.81 | 5.85 | 6.44 | 6.86 |
| HPV16/18 | Organised | Paris | 40 | 4.73 | 3.31 | 3.50 | 3.84 | 4.68 | 5.70 | 6.26 | 6.71 |
| HPV16/18 | Organised | Paris | 41 | 4.62 | 3.22 | 3.40 | 3.75 | 4.56 | 5.57 | 6.13 | 6.55 |
| HPV16/18 | Organised | Paris | 42 | 4.49 | 3.13 | 3.30 | 3.65 | 4.44 | 5.42 | 5.95 | 6.38 |
| HPV16/18 | Organised | Paris | 43 | 4.38 | 3.05 | 3.22 | 3.56 | 4.32 | 5.27 | 5.81 | 6.23 |
| HPV16/18 | Organised | Paris | 44 | 4.28 | 2.98 | 3.16 | 3.47 | 4.22 | 5.15 | 5.72 | 6.07 |
| HPV16/18 | Organised | Paris | 45 | 4.20 | 2.93 | 3.09 | 3.41 | 4.14 | 5.06 | 5.59 | 5.98 |
| HPV16/18 | Organised | Paris | 46 | 4.11 | 2.87 | 3.03 | 3.33 | 4.06 | 4.95 | 5.47 | 5.86 |
| HPV16/18 | Organised | Paris | 47 | 4.03 | 2.80 | 2.96 | 3.26 | 3.98 | 4.85 | 5.36 | 5.73 |
| HPV16/18 | Organised | Paris | 48 | 3.95 | 2.73 | 2.89 | 3.20 | 3.90 | 4.76 | 5.29 | 5.64 |
| HPV16/18 | Organised | Paris | 49 | 3.87 | 2.68 | 2.83 | 3.14 | 3.83 | 4.66 | 5.21 | 5.52 |
| HPV16/18 | Organised | Paris | 50 | 3.80 | 2.63 | 2.76 | 3.07 | 3.75 | 4.58 | 5.11 | 5.43 |
| HPV16/18 | Organised | Paris | 51 | 3.73 | 2.58 | 2.71 | 3.01 | 3.68 | 4.50 | 5.02 | 5.33 |
| HPV16/18 | Organised | Paris | 52 | 3.67 | 2.51 | 2.67 | 2.96 | 3.62 | 4.43 | 4.94 | 5.25 |
| HPV16/18 | Organised | Paris | 53 | 3.59 | 2.46 | 2.63 | 2.90 | 3.54 | 4.34 | 4.85 | 5.16 |
| HPV16/18 | Organised | Paris | 54 | 3.52 | 2.40 | 2.57 | 2.84 | 3.47 | 4.25 | 4.74 | 5.05 |
| HPV16/18 | Organised | Paris | 55 | 3.46 | 2.36 | 2.50 | 2.78 | 3.41 | 4.18 | 4.66 | 4.97 |
| HPV16/18 | Organised | Paris | 56 | 3.42 | 2.31 | 2.47 | 2.75 | 3.38 | 4.13 | 4.63 | 4.90 |
| HPV16/18 | Organised | Paris | 57 | 3.39 | 2.28 | 2.43 | 2.72 | 3.35 | 4.11 | 4.61 | 4.89 |
| HPV16/18 | Organised | Paris | 58 | 3.39 | 2.25 | 2.42 | 2.71 | 3.34 | 4.12 | 4.61 | 4.88 |
| HPV16/18 | Organised | Paris | 59 | 3.39 | 2.25 | 2.42 | 2.71 | 3.35 | 4.14 | 4.61 | 4.88 |
| HPV16/18 | Organised | Paris | 60 | 3.41 | 2.29 | 2.42 | 2.72 | 3.36 | 4.17 | 4.65 | 4.91 |
| HPV16/18 | Organised | Paris | 61 | 3.43 | 2.28 | 2.43 | 2.74 | 3.38 | 4.21 | 4.72 | 4.97 |
| HPV16/18 | Organised | Paris | 62 | 3.45 | 2.31 | 2.43 | 2.74 | 3.39 | 4.23 | 4.72 | 5.01 |
| HPV16/18 | Organised | Paris | 63 | 3.45 | 2.30 | 2.42 | 2.73 | 3.40 | 4.24 | 4.73 | 5.07 |

**Table S9 Posterior expected HR HPV cervical infection prevalence (in %) in major French cities, stratified by type of test, city, and age. Table counterpart to Figure 3A** ***(continued)***

| | | | | Posterior distribution | | | | | | | |
|---|---|---|---|---|---|---|---|---|---|---|---|
| | | | | | Percentiles | | | | | | |
| Virus | Test | City | Age | Average | 0.01 | 0.025 | 0.1 | 0.5 | 0.9 | 0.975 | 0.99 |
| HPV16/18 | Organised | Paris | 64 | 3.45 | 2.29 | 2.42 | 2.71 | 3.40 | 4.26 | 4.77 | 5.13 |
| HPV16/18 | Organised | Paris | 65 | 3.44 | 2.24 | 2.38 | 2.69 | 3.38 | 4.27 | 4.83 | 5.16 |
| HPV16/18 | Organised | Paris | 66 | 3.42 | 2.16 | 2.32 | 2.63 | 3.37 | 4.28 | 4.91 | 5.23 |
| HPV16/18 | Opportunistic | Nice | 30 | 5.00 | 2.46 | 2.75 | 3.37 | 4.83 | 6.88 | 8.38 | 9.11 |
| HPV16/18 | Opportunistic | Nice | 31 | 5.29 | 2.62 | 2.94 | 3.56 | 5.12 | 7.27 | 8.77 | 9.74 |
| HPV16/18 | Opportunistic | Nice | 32 | 5.55 | 2.74 | 3.08 | 3.73 | 5.37 | 7.61 | 9.18 | 10.23 |
| HPV16/18 | Opportunistic | Nice | 33 | 5.75 | 2.87 | 3.18 | 3.85 | 5.55 | 7.88 | 9.57 | 10.61 |
| HPV16/18 | Opportunistic | Nice | 34 | 5.83 | 2.90 | 3.26 | 3.92 | 5.63 | 8.00 | 9.68 | 10.68 |
| HPV16/18 | Opportunistic | Nice | 35 | 5.78 | 2.89 | 3.23 | 3.89 | 5.58 | 7.95 | 9.59 | 10.60 |
| HPV16/18 | Opportunistic | Nice | 36 | 5.67 | 2.81 | 3.15 | 3.81 | 5.48 | 7.76 | 9.40 | 10.38 |
| HPV16/18 | Opportunistic | Nice | 37 | 5.53 | 2.77 | 3.07 | 3.72 | 5.34 | 7.57 | 9.18 | 10.14 |
| HPV16/18 | Opportunistic | Nice | 38 | 5.38 | 2.69 | 2.99 | 3.62 | 5.21 | 7.38 | 8.95 | 9.92 |
| HPV16/18 | Opportunistic | Nice | 39 | 5.24 | 2.62 | 2.90 | 3.54 | 5.06 | 7.21 | 8.70 | 9.61 |
| HPV16/18 | Opportunistic | Nice | 40 | 5.10 | 2.53 | 2.83 | 3.43 | 4.91 | 7.04 | 8.44 | 9.36 |
| HPV16/18 | Opportunistic | Nice | 41 | 4.96 | 2.46 | 2.76 | 3.33 | 4.79 | 6.85 | 8.25 | 9.14 |
| HPV16/18 | Opportunistic | Nice | 42 | 4.81 | 2.38 | 2.68 | 3.23 | 4.65 | 6.63 | 8.03 | 8.91 |
| HPV16/18 | Opportunistic | Nice | 43 | 4.68 | 2.30 | 2.60 | 3.14 | 4.51 | 6.45 | 7.82 | 8.64 |
| HPV16/18 | Opportunistic | Nice | 44 | 4.56 | 2.25 | 2.53 | 3.06 | 4.40 | 6.28 | 7.59 | 8.40 |
| HPV16/18 | Opportunistic | Nice | 45 | 4.47 | 2.20 | 2.45 | 3.00 | 4.30 | 6.14 | 7.42 | 8.23 |
| HPV16/18 | Opportunistic | Nice | 46 | 4.36 | 2.13 | 2.39 | 2.92 | 4.20 | 5.98 | 7.27 | 8.02 |
| HPV16/18 | Opportunistic | Nice | 47 | 4.26 | 2.09 | 2.34 | 2.84 | 4.11 | 5.87 | 7.12 | 7.84 |
| HPV16/18 | Opportunistic | Nice | 48 | 4.16 | 2.06 | 2.29 | 2.78 | 4.01 | 5.74 | 6.95 | 7.71 |
| HPV16/18 | Opportunistic | Nice | 49 | 4.07 | 2.02 | 2.22 | 2.71 | 3.92 | 5.62 | 6.81 | 7.50 |
| HPV16/18 | Opportunistic | Nice | 50 | 3.98 | 1.97 | 2.17 | 2.65 | 3.84 | 5.49 | 6.66 | 7.29 |
| HPV16/18 | Opportunistic | Nice | 51 | 3.89 | 1.94 | 2.12 | 2.60 | 3.76 | 5.35 | 6.52 | 7.13 |
| HPV16/18 | Opportunistic | Nice | 52 | 3.81 | 1.90 | 2.09 | 2.53 | 3.67 | 5.25 | 6.41 | 7.00 |
| HPV16/18 | Opportunistic | Nice | 53 | 3.71 | 1.84 | 2.03 | 2.48 | 3.57 | 5.13 | 6.26 | 6.85 |
| HPV16/18 | Opportunistic | Nice | 54 | 3.62 | 1.77 | 1.97 | 2.40 | 3.49 | 4.99 | 6.12 | 6.69 |
| HPV16/18 | Opportunistic | Nice | 55 | 3.54 | 1.74 | 1.93 | 2.35 | 3.41 | 4.88 | 6.00 | 6.59 |

**Table S9 Posterior expected HR HPV cervical infection prevalence (in %) in major French cities, stratified by type of test, city, and age. Table counterpart to Figure 3A** ***(continued)***

| | | | | Posterior distribution | | | | | | | |
|---|---|---|---|---|---|---|---|---|---|---|---|
| | | | | | Percentiles | | | | | | |
| Virus | Test | City | Age | Average | 0.01 | 0.025 | 0.1 | 0.5 | 0.9 | 0.975 | 0.99 |
| HPV16/18 | Opportunistic | Nice | 56 | 3.48 | 1.71 | 1.91 | 2.32 | 3.36 | 4.80 | 5.89 | 6.48 |
| HPV16/18 | Opportunistic | Nice | 57 | 3.43 | 1.69 | 1.88 | 2.29 | 3.32 | 4.75 | 5.80 | 6.37 |
| HPV16/18 | Opportunistic | Nice | 58 | 3.41 | 1.67 | 1.87 | 2.27 | 3.28 | 4.71 | 5.73 | 6.34 |
| HPV16/18 | Opportunistic | Nice | 59 | 3.40 | 1.67 | 1.85 | 2.25 | 3.27 | 4.71 | 5.68 | 6.32 |
| HPV16/18 | Opportunistic | Nice | 60 | 3.40 | 1.67 | 1.85 | 2.25 | 3.28 | 4.71 | 5.71 | 6.36 |
| HPV16/18 | Opportunistic | Nice | 61 | 3.40 | 1.69 | 1.86 | 2.26 | 3.28 | 4.73 | 5.74 | 6.38 |
| HPV16/18 | Opportunistic | Nice | 62 | 3.40 | 1.69 | 1.85 | 2.25 | 3.27 | 4.71 | 5.69 | 6.33 |
| HPV16/18 | Opportunistic | Nice | 63 | 3.38 | 1.68 | 1.84 | 2.25 | 3.25 | 4.67 | 5.68 | 6.27 |
| HPV16/18 | Opportunistic | Nice | 64 | 3.36 | 1.67 | 1.83 | 2.23 | 3.23 | 4.64 | 5.65 | 6.25 |
| HPV16/18 | Opportunistic | Nice | 65 | 3.33 | 1.64 | 1.82 | 2.20 | 3.20 | 4.58 | 5.62 | 6.18 |
| HPV16/18 | Opportunistic | Nice | 66 | 3.29 | 1.59 | 1.79 | 2.15 | 3.16 | 4.54 | 5.61 | 6.23 |
| HPV16/18 | Opportunistic | Marseille | 30 | 5.67 | 4.23 | 4.45 | 4.85 | 5.64 | 6.52 | 7.08 | 7.41 |
| HPV16/18 | Opportunistic | Marseille | 31 | 5.99 | 4.52 | 4.73 | 5.14 | 5.96 | 6.88 | 7.46 | 7.76 |
| HPV16/18 | Opportunistic | Marseille | 32 | 6.29 | 4.75 | 4.95 | 5.39 | 6.25 | 7.22 | 7.79 | 8.10 |
| HPV16/18 | Opportunistic | Marseille | 33 | 6.51 | 4.91 | 5.13 | 5.58 | 6.48 | 7.46 | 8.03 | 8.34 |
| HPV16/18 | Opportunistic | Marseille | 34 | 6.60 | 4.97 | 5.20 | 5.65 | 6.58 | 7.55 | 8.15 | 8.48 |
| HPV16/18 | Opportunistic | Marseille | 35 | 6.55 | 4.95 | 5.13 | 5.61 | 6.51 | 7.50 | 8.09 | 8.45 |
| HPV16/18 | Opportunistic | Marseille | 36 | 6.42 | 4.88 | 5.03 | 5.51 | 6.38 | 7.39 | 7.96 | 8.26 |
| HPV16/18 | Opportunistic | Marseille | 37 | 6.26 | 4.72 | 4.94 | 5.35 | 6.22 | 7.17 | 7.79 | 8.04 |
| HPV16/18 | Opportunistic | Marseille | 38 | 6.10 | 4.59 | 4.83 | 5.21 | 6.05 | 7.00 | 7.59 | 7.85 |
| HPV16/18 | Opportunistic | Marseille | 39 | 5.93 | 4.47 | 4.69 | 5.08 | 5.90 | 6.82 | 7.38 | 7.66 |
| HPV16/18 | Opportunistic | Marseille | 40 | 5.77 | 4.33 | 4.54 | 4.95 | 5.73 | 6.66 | 7.17 | 7.50 |
| HPV16/18 | Opportunistic | Marseille | 41 | 5.62 | 4.19 | 4.42 | 4.81 | 5.58 | 6.48 | 6.97 | 7.31 |
| HPV16/18 | Opportunistic | Marseille | 42 | 5.45 | 4.08 | 4.29 | 4.68 | 5.41 | 6.30 | 6.75 | 7.11 |
| HPV16/18 | Opportunistic | Marseille | 43 | 5.30 | 3.96 | 4.17 | 4.54 | 5.27 | 6.12 | 6.57 | 6.87 |
| HPV16/18 | Opportunistic | Marseille | 44 | 5.17 | 3.87 | 4.06 | 4.43 | 5.14 | 5.97 | 6.43 | 6.70 |
| HPV16/18 | Opportunistic | Marseille | 45 | 5.06 | 3.78 | 3.98 | 4.32 | 5.03 | 5.85 | 6.28 | 6.54 |
| HPV16/18 | Opportunistic | Marseille | 46 | 4.94 | 3.68 | 3.88 | 4.22 | 4.91 | 5.70 | 6.14 | 6.41 |
| HPV16/18 | Opportunistic | Marseille | 47 | 4.82 | 3.59 | 3.80 | 4.12 | 4.79 | 5.56 | 6.01 | 6.24 |

**Table S9 Posterior expected HR HPV cervical infection prevalence (in %) in major French cities, stratified by type of test, city, and age. Table counterpart to Figure 3A** ***(continued)***

| | | | | Posterior distribution | | | | | | | |
|---|---|---|---|---|---|---|---|---|---|---|---|
| | | | | | Percentiles | | | | | | |
| Virus | Test | City | Age | Average | 0.01 | 0.025 | 0.1 | 0.5 | 0.9 | 0.975 | 0.99 |
| HPV16/18 | Opportunistic | Marseille | 48 | 4.72 | 3.52 | 3.72 | 4.03 | 4.68 | 5.43 | 5.88 | 6.10 |
| HPV16/18 | Opportunistic | Marseille | 49 | 4.61 | 3.45 | 3.64 | 3.94 | 4.58 | 5.32 | 5.75 | 5.96 |
| HPV16/18 | Opportunistic | Marseille | 50 | 4.51 | 3.40 | 3.57 | 3.84 | 4.48 | 5.20 | 5.60 | 5.83 |
| HPV16/18 | Opportunistic | Marseille | 51 | 4.41 | 3.33 | 3.49 | 3.76 | 4.38 | 5.09 | 5.48 | 5.72 |
| HPV16/18 | Opportunistic | Marseille | 52 | 4.32 | 3.24 | 3.41 | 3.68 | 4.29 | 4.98 | 5.38 | 5.60 |
| HPV16/18 | Opportunistic | Marseille | 53 | 4.21 | 3.14 | 3.33 | 3.59 | 4.19 | 4.86 | 5.26 | 5.48 |
| HPV16/18 | Opportunistic | Marseille | 54 | 4.10 | 3.05 | 3.22 | 3.50 | 4.08 | 4.73 | 5.10 | 5.40 |
| HPV16/18 | Opportunistic | Marseille | 55 | 4.01 | 2.98 | 3.15 | 3.42 | 3.98 | 4.63 | 5.01 | 5.31 |
| HPV16/18 | Opportunistic | Marseille | 56 | 3.94 | 2.95 | 3.08 | 3.36 | 3.92 | 4.54 | 4.92 | 5.19 |
| HPV16/18 | Opportunistic | Marseille | 57 | 3.89 | 2.89 | 3.04 | 3.32 | 3.87 | 4.49 | 4.87 | 5.08 |
| HPV16/18 | Opportunistic | Marseille | 58 | 3.86 | 2.86 | 3.02 | 3.30 | 3.84 | 4.45 | 4.84 | 5.00 |
| HPV16/18 | Opportunistic | Marseille | 59 | 3.85 | 2.85 | 3.00 | 3.28 | 3.83 | 4.45 | 4.80 | 5.00 |
| HPV16/18 | Opportunistic | Marseille | 60 | 3.85 | 2.86 | 3.00 | 3.28 | 3.83 | 4.45 | 4.81 | 5.03 |
| HPV16/18 | Opportunistic | Marseille | 61 | 3.86 | 2.86 | 3.01 | 3.29 | 3.84 | 4.47 | 4.84 | 5.05 |
| HPV16/18 | Opportunistic | Marseille | 62 | 3.85 | 2.86 | 3.01 | 3.28 | 3.83 | 4.45 | 4.84 | 5.08 |
| HPV16/18 | Opportunistic | Marseille | 63 | 3.84 | 2.84 | 2.99 | 3.26 | 3.80 | 4.44 | 4.83 | 5.04 |
| HPV16/18 | Opportunistic | Marseille | 64 | 3.81 | 2.82 | 2.95 | 3.23 | 3.78 | 4.42 | 4.81 | 5.02 |
| HPV16/18 | Opportunistic | Marseille | 65 | 3.78 | 2.76 | 2.90 | 3.19 | 3.74 | 4.39 | 4.82 | 5.02 |
| HPV16/18 | Opportunistic | Marseille | 66 | 3.73 | 2.67 | 2.82 | 3.11 | 3.70 | 4.38 | 4.84 | 5.04 |
| HPV16/18 | Opportunistic | Toulouse | 30 | 5.19 | 3.06 | 3.23 | 3.80 | 5.08 | 6.73 | 7.75 | 8.60 |
| HPV16/18 | Opportunistic | Toulouse | 31 | 5.49 | 3.23 | 3.45 | 4.00 | 5.36 | 7.10 | 8.21 | 9.00 |
| HPV16/18 | Opportunistic | Toulouse | 32 | 5.76 | 3.37 | 3.66 | 4.22 | 5.63 | 7.46 | 8.54 | 9.37 |
| HPV16/18 | Opportunistic | Toulouse | 33 | 5.96 | 3.51 | 3.80 | 4.39 | 5.83 | 7.70 | 8.88 | 9.69 |
| HPV16/18 | Opportunistic | Toulouse | 34 | 6.05 | 3.55 | 3.86 | 4.45 | 5.92 | 7.80 | 8.99 | 9.78 |
| HPV16/18 | Opportunistic | Toulouse | 35 | 6.00 | 3.51 | 3.81 | 4.40 | 5.88 | 7.75 | 8.90 | 9.74 |
| HPV16/18 | Opportunistic | Toulouse | 36 | 5.88 | 3.46 | 3.72 | 4.32 | 5.76 | 7.60 | 8.71 | 9.64 |
| HPV16/18 | Opportunistic | Toulouse | 37 | 5.73 | 3.38 | 3.60 | 4.18 | 5.59 | 7.39 | 8.53 | 9.39 |
| HPV16/18 | Opportunistic | Toulouse | 38 | 5.58 | 3.28 | 3.53 | 4.06 | 5.45 | 7.21 | 8.31 | 9.14 |
| HPV16/18 | Opportunistic | Toulouse | 39 | 5.43 | 3.20 | 3.44 | 3.95 | 5.30 | 7.03 | 8.07 | 8.88 |

**Table S9 Posterior expected HR HPV cervical infection prevalence (in %) in major French cities, stratified by type of test, city, and age. Table counterpart to Figure 3A *(continued)***

| | | | | Posterior distribution | | | | | | | |
|---|---|---|---|---|---|---|---|---|---|---|---|
| | | | | | Percentiles | | | | | | |
| Virus | Test | City | Age | Average | 0.01 | 0.025 | 0.1 | 0.5 | 0.9 | 0.975 | 0.99 |
| HPV16/18 | Opportunistic | Toulouse | 40 | 5.28 | 3.11 | 3.35 | 3.84 | 5.15 | 6.84 | 7.82 | 8.54 |
| HPV16/18 | Opportunistic | Toulouse | 41 | 5.14 | 3.00 | 3.25 | 3.73 | 5.02 | 6.65 | 7.66 | 8.34 |
| HPV16/18 | Opportunistic | Toulouse | 42 | 4.99 | 2.91 | 3.15 | 3.63 | 4.87 | 6.46 | 7.47 | 8.09 |
| HPV16/18 | Opportunistic | Toulouse | 43 | 4.85 | 2.83 | 3.06 | 3.54 | 4.73 | 6.28 | 7.26 | 7.92 |
| HPV16/18 | Opportunistic | Toulouse | 44 | 4.73 | 2.74 | 2.97 | 3.45 | 4.63 | 6.13 | 7.08 | 7.74 |
| HPV16/18 | Opportunistic | Toulouse | 45 | 4.63 | 2.67 | 2.91 | 3.37 | 4.54 | 6.01 | 6.93 | 7.58 |
| HPV16/18 | Opportunistic | Toulouse | 46 | 4.52 | 2.61 | 2.85 | 3.29 | 4.43 | 5.87 | 6.75 | 7.36 |
| HPV16/18 | Opportunistic | Toulouse | 47 | 4.41 | 2.54 | 2.78 | 3.22 | 4.33 | 5.74 | 6.60 | 7.17 |
| HPV16/18 | Opportunistic | Toulouse | 48 | 4.31 | 2.48 | 2.72 | 3.15 | 4.22 | 5.62 | 6.48 | 7.05 |
| HPV16/18 | Opportunistic | Toulouse | 49 | 4.22 | 2.42 | 2.66 | 3.07 | 4.12 | 5.47 | 6.34 | 6.93 |
| HPV16/18 | Opportunistic | Toulouse | 50 | 4.12 | 2.35 | 2.60 | 2.99 | 4.02 | 5.33 | 6.21 | 6.78 |
| HPV16/18 | Opportunistic | Toulouse | 51 | 4.04 | 2.33 | 2.54 | 2.93 | 3.94 | 5.22 | 6.07 | 6.63 |
| HPV16/18 | Opportunistic | Toulouse | 52 | 3.95 | 2.25 | 2.48 | 2.86 | 3.85 | 5.11 | 5.95 | 6.47 |
| HPV16/18 | Opportunistic | Toulouse | 53 | 3.85 | 2.21 | 2.41 | 2.79 | 3.76 | 4.99 | 5.82 | 6.34 |
| HPV16/18 | Opportunistic | Toulouse | 54 | 3.75 | 2.16 | 2.35 | 2.72 | 3.66 | 4.90 | 5.65 | 6.20 |
| HPV16/18 | Opportunistic | Toulouse | 55 | 3.67 | 2.12 | 2.28 | 2.66 | 3.58 | 4.78 | 5.51 | 6.04 |
| HPV16/18 | Opportunistic | Toulouse | 56 | 3.61 | 2.08 | 2.24 | 2.61 | 3.52 | 4.69 | 5.44 | 5.95 |
| HPV16/18 | Opportunistic | Toulouse | 57 | 3.56 | 2.06 | 2.23 | 2.60 | 3.48 | 4.65 | 5.36 | 5.88 |
| HPV16/18 | Opportunistic | Toulouse | 58 | 3.53 | 2.03 | 2.21 | 2.57 | 3.45 | 4.62 | 5.32 | 5.79 |
| HPV16/18 | Opportunistic | Toulouse | 59 | 3.52 | 2.02 | 2.20 | 2.56 | 3.43 | 4.58 | 5.33 | 5.77 |
| HPV16/18 | Opportunistic | Toulouse | 60 | 3.52 | 2.03 | 2.19 | 2.56 | 3.44 | 4.58 | 5.35 | 5.75 |
| HPV16/18 | Opportunistic | Toulouse | 61 | 3.53 | 2.03 | 2.20 | 2.56 | 3.44 | 4.59 | 5.34 | 5.75 |
| HPV16/18 | Opportunistic | Toulouse | 62 | 3.52 | 2.04 | 2.19 | 2.55 | 3.45 | 4.58 | 5.31 | 5.75 |
| HPV16/18 | Opportunistic | Toulouse | 63 | 3.51 | 2.02 | 2.18 | 2.53 | 3.43 | 4.55 | 5.28 | 5.76 |
| HPV16/18 | Opportunistic | Toulouse | 64 | 3.48 | 2.01 | 2.18 | 2.50 | 3.40 | 4.53 | 5.23 | 5.74 |
| HPV16/18 | Opportunistic | Toulouse | 65 | 3.45 | 1.97 | 2.15 | 2.47 | 3.37 | 4.48 | 5.25 | 5.70 |
| HPV16/18 | Opportunistic | Toulouse | 66 | 3.41 | 1.93 | 2.09 | 2.44 | 3.33 | 4.47 | 5.21 | 5.77 |
| HPV16/18 | Opportunistic | Bordeaux | 30 | 5.78 | 4.23 | 4.39 | 4.80 | 5.72 | 6.83 | 7.40 | 7.91 |
| HPV16/18 | Opportunistic | Bordeaux | 31 | 6.11 | 4.50 | 4.69 | 5.10 | 6.06 | 7.22 | 7.82 | 8.24 |

**Table S9 Posterior expected HR HPV cervical infection prevalence (in %) in major French cities, stratified by type of test, city, and age. Table counterpart to Figure 3A** ***(continued)***

| | | | | Posterior distribution | | | | | | | |
|---|---|---|---|---|---|---|---|---|---|---|---|
| | | | | | Percentiles | | | | | | |
| Virus | Test | City | Age | Average | 0.01 | 0.025 | 0.1 | 0.5 | 0.9 | 0.975 | 0.99 |
| HPV16/18 | Opportunistic | Bordeaux | 32 | 6.40 | 4.71 | 4.91 | 5.36 | 6.36 | 7.57 | 8.21 | 8.65 |
| HPV16/18 | Opportunistic | Bordeaux | 33 | 6.63 | 4.88 | 5.09 | 5.55 | 6.58 | 7.83 | 8.52 | 8.91 |
| HPV16/18 | Opportunistic | Bordeaux | 34 | 6.73 | 4.98 | 5.17 | 5.62 | 6.68 | 7.93 | 8.65 | 9.06 |
| HPV16/18 | Opportunistic | Bordeaux | 35 | 6.67 | 4.94 | 5.12 | 5.57 | 6.62 | 7.86 | 8.59 | 8.98 |
| HPV16/18 | Opportunistic | Bordeaux | 36 | 6.54 | 4.82 | 5.04 | 5.45 | 6.49 | 7.70 | 8.43 | 8.86 |
| HPV16/18 | Opportunistic | Bordeaux | 37 | 6.38 | 4.69 | 4.90 | 5.31 | 6.32 | 7.52 | 8.20 | 8.62 |
| HPV16/18 | Opportunistic | Bordeaux | 38 | 6.21 | 4.58 | 4.77 | 5.17 | 6.15 | 7.33 | 8.00 | 8.39 |
| HPV16/18 | Opportunistic | Bordeaux | 39 | 6.05 | 4.43 | 4.64 | 5.04 | 6.00 | 7.14 | 7.81 | 8.17 |
| HPV16/18 | Opportunistic | Bordeaux | 40 | 5.88 | 4.29 | 4.51 | 4.89 | 5.83 | 6.96 | 7.60 | 7.97 |
| HPV16/18 | Opportunistic | Bordeaux | 41 | 5.72 | 4.19 | 4.38 | 4.76 | 5.66 | 6.76 | 7.37 | 7.78 |
| HPV16/18 | Opportunistic | Bordeaux | 42 | 5.56 | 4.07 | 4.25 | 4.63 | 5.50 | 6.55 | 7.17 | 7.53 |
| HPV16/18 | Opportunistic | Bordeaux | 43 | 5.40 | 3.97 | 4.13 | 4.50 | 5.35 | 6.37 | 6.97 | 7.30 |
| HPV16/18 | Opportunistic | Bordeaux | 44 | 5.27 | 3.88 | 4.03 | 4.40 | 5.22 | 6.22 | 6.78 | 7.11 |
| HPV16/18 | Opportunistic | Bordeaux | 45 | 5.16 | 3.78 | 3.93 | 4.30 | 5.11 | 6.11 | 6.65 | 6.93 |
| HPV16/18 | Opportunistic | Bordeaux | 46 | 5.04 | 3.70 | 3.84 | 4.19 | 4.98 | 5.97 | 6.51 | 6.74 |
| HPV16/18 | Opportunistic | Bordeaux | 47 | 4.92 | 3.59 | 3.74 | 4.10 | 4.86 | 5.82 | 6.34 | 6.61 |
| HPV16/18 | Opportunistic | Bordeaux | 48 | 4.81 | 3.50 | 3.65 | 4.00 | 4.75 | 5.69 | 6.21 | 6.52 |
| HPV16/18 | Opportunistic | Bordeaux | 49 | 4.70 | 3.42 | 3.58 | 3.91 | 4.65 | 5.56 | 6.07 | 6.36 |
| HPV16/18 | Opportunistic | Bordeaux | 50 | 4.59 | 3.36 | 3.50 | 3.81 | 4.54 | 5.45 | 5.93 | 6.25 |
| HPV16/18 | Opportunistic | Bordeaux | 51 | 4.50 | 3.27 | 3.44 | 3.73 | 4.46 | 5.34 | 5.82 | 6.11 |
| HPV16/18 | Opportunistic | Bordeaux | 52 | 4.40 | 3.20 | 3.36 | 3.65 | 4.36 | 5.22 | 5.70 | 5.96 |
| HPV16/18 | Opportunistic | Bordeaux | 53 | 4.29 | 3.10 | 3.26 | 3.56 | 4.26 | 5.08 | 5.57 | 5.83 |
| HPV16/18 | Opportunistic | Bordeaux | 54 | 4.18 | 3.02 | 3.17 | 3.47 | 4.15 | 4.95 | 5.41 | 5.69 |
| HPV16/18 | Opportunistic | Bordeaux | 55 | 4.09 | 2.96 | 3.09 | 3.40 | 4.05 | 4.85 | 5.31 | 5.58 |
| HPV16/18 | Opportunistic | Bordeaux | 56 | 4.02 | 2.90 | 3.04 | 3.33 | 3.97 | 4.77 | 5.23 | 5.47 |
| HPV16/18 | Opportunistic | Bordeaux | 57 | 3.97 | 2.88 | 3.01 | 3.29 | 3.92 | 4.71 | 5.16 | 5.42 |
| HPV16/18 | Opportunistic | Bordeaux | 58 | 3.94 | 2.84 | 2.98 | 3.27 | 3.89 | 4.68 | 5.12 | 5.39 |
| HPV16/18 | Opportunistic | Bordeaux | 59 | 3.92 | 2.82 | 2.98 | 3.25 | 3.88 | 4.68 | 5.10 | 5.36 |
| HPV16/18 | Opportunistic | Bordeaux | 60 | 3.93 | 2.85 | 2.98 | 3.25 | 3.88 | 4.68 | 5.10 | 5.38 |

**Table S9** **Posterior expected HR HPV cervical infection prevalence (in %) in major French cities, stratified by type of test, city, and age. Table counterpart to Figure 3A** ***(continued)***

| | | | | Posterior distribution | | | | | | | |
|---|---|---|---|---|---|---|---|---|---|---|---|
| | | | | | Percentiles | | | | | | |
| Virus | Test | City | Age | Average | 0.01 | 0.025 | 0.1 | 0.5 | 0.9 | 0.975 | 0.99 |
| HPV16/18 | Opportunistic | Bordeaux | 61 | 3.93 | 2.84 | 2.97 | 3.25 | 3.89 | 4.67 | 5.10 | 5.38 |
| HPV16/18 | Opportunistic | Bordeaux | 62 | 3.93 | 2.85 | 2.97 | 3.25 | 3.88 | 4.66 | 5.10 | 5.35 |
| HPV16/18 | Opportunistic | Bordeaux | 63 | 3.91 | 2.83 | 2.96 | 3.22 | 3.87 | 4.65 | 5.08 | 5.31 |
| HPV16/18 | Opportunistic | Bordeaux | 64 | 3.88 | 2.79 | 2.92 | 3.20 | 3.84 | 4.62 | 5.05 | 5.28 |
| HPV16/18 | Opportunistic | Bordeaux | 65 | 3.85 | 2.75 | 2.88 | 3.16 | 3.81 | 4.60 | 5.05 | 5.25 |
| HPV16/18 | Opportunistic | Bordeaux | 66 | 3.80 | 2.65 | 2.82 | 3.09 | 3.76 | 4.56 | 5.04 | 5.29 |
| HPV16/18 | Opportunistic | Montpellier | 30 | 6.11 | 3.86 | 4.09 | 4.62 | 5.98 | 7.73 | 8.74 | 9.44 |
| HPV16/18 | Opportunistic | Montpellier | 31 | 6.46 | 4.09 | 4.31 | 4.92 | 6.33 | 8.17 | 9.19 | 10.00 |
| HPV16/18 | Opportunistic | Montpellier | 32 | 6.77 | 4.28 | 4.56 | 5.16 | 6.63 | 8.55 | 9.67 | 10.36 |
| HPV16/18 | Opportunistic | Montpellier | 33 | 7.02 | 4.41 | 4.71 | 5.36 | 6.87 | 8.82 | 10.08 | 10.69 |
| HPV16/18 | Opportunistic | Montpellier | 34 | 7.11 | 4.46 | 4.79 | 5.44 | 6.98 | 8.95 | 10.23 | 10.85 |
| HPV16/18 | Opportunistic | Montpellier | 35 | 7.06 | 4.43 | 4.76 | 5.40 | 6.92 | 8.88 | 10.09 | 10.74 |
| HPV16/18 | Opportunistic | Montpellier | 36 | 6.92 | 4.35 | 4.65 | 5.30 | 6.80 | 8.71 | 9.87 | 10.58 |
| HPV16/18 | Opportunistic | Montpellier | 37 | 6.74 | 4.25 | 4.51 | 5.15 | 6.63 | 8.45 | 9.61 | 10.32 |
| HPV16/18 | Opportunistic | Montpellier | 38 | 6.57 | 4.17 | 4.40 | 5.01 | 6.46 | 8.26 | 9.40 | 10.18 |
| HPV16/18 | Opportunistic | Montpellier | 39 | 6.40 | 4.03 | 4.27 | 4.88 | 6.28 | 8.06 | 9.17 | 10.06 |
| HPV16/18 | Opportunistic | Montpellier | 40 | 6.22 | 3.91 | 4.16 | 4.73 | 6.10 | 7.86 | 8.98 | 9.78 |
| HPV16/18 | Opportunistic | Montpellier | 41 | 6.06 | 3.81 | 4.02 | 4.59 | 5.95 | 7.65 | 8.77 | 9.60 |
| HPV16/18 | Opportunistic | Montpellier | 42 | 5.88 | 3.68 | 3.92 | 4.46 | 5.77 | 7.43 | 8.48 | 9.26 |
| HPV16/18 | Opportunistic | Montpellier | 43 | 5.72 | 3.59 | 3.80 | 4.34 | 5.61 | 7.23 | 8.24 | 8.95 |
| HPV16/18 | Opportunistic | Montpellier | 44 | 5.58 | 3.49 | 3.72 | 4.22 | 5.47 | 7.05 | 8.06 | 8.68 |
| HPV16/18 | Opportunistic | Montpellier | 45 | 5.46 | 3.42 | 3.65 | 4.14 | 5.35 | 6.91 | 7.89 | 8.42 |
| HPV16/18 | Opportunistic | Montpellier | 46 | 5.33 | 3.36 | 3.56 | 4.05 | 5.23 | 6.72 | 7.70 | 8.26 |
| HPV16/18 | Opportunistic | Montpellier | 47 | 5.21 | 3.24 | 3.48 | 3.95 | 5.10 | 6.56 | 7.52 | 8.07 |
| HPV16/18 | Opportunistic | Montpellier | 48 | 5.09 | 3.17 | 3.40 | 3.85 | 4.99 | 6.41 | 7.40 | 7.91 |
| HPV16/18 | Opportunistic | Montpellier | 49 | 4.98 | 3.11 | 3.33 | 3.76 | 4.89 | 6.27 | 7.24 | 7.73 |
| HPV16/18 | Opportunistic | Montpellier | 50 | 4.87 | 3.04 | 3.25 | 3.69 | 4.78 | 6.17 | 7.07 | 7.52 |
| HPV16/18 | Opportunistic | Montpellier | 51 | 4.76 | 2.96 | 3.19 | 3.61 | 4.67 | 6.05 | 6.90 | 7.36 |
| HPV16/18 | Opportunistic | Montpellier | 52 | 4.66 | 2.91 | 3.12 | 3.53 | 4.56 | 5.93 | 6.77 | 7.21 |

**Table S9 Posterior expected HR HPV cervical infection prevalence (in %) in major French cities, stratified by type of test, city, and age. Table counterpart to Figure 3A *(continued)***

| | | | | Posterior distribution | | | | | | | |
|---|---|---|---|---|---|---|---|---|---|---|---|
| | | | | | Percentiles | | | | | | |
| Virus | Test | City | Age | Average | 0.01 | 0.025 | 0.1 | 0.5 | 0.9 | 0.975 | 0.99 |
| HPV16/18 | Opportunistic | Montpellier | 53 | 4.55 | 2.83 | 3.04 | 3.45 | 4.44 | 5.78 | 6.59 | 7.01 |
| HPV16/18 | Opportunistic | Montpellier | 54 | 4.43 | 2.75 | 2.95 | 3.36 | 4.33 | 5.64 | 6.42 | 6.86 |
| HPV16/18 | Opportunistic | Montpellier | 55 | 4.34 | 2.67 | 2.88 | 3.28 | 4.24 | 5.52 | 6.26 | 6.74 |
| HPV16/18 | Opportunistic | Montpellier | 56 | 4.26 | 2.64 | 2.83 | 3.22 | 4.17 | 5.42 | 6.15 | 6.67 |
| HPV16/18 | Opportunistic | Montpellier | 57 | 4.21 | 2.59 | 2.80 | 3.18 | 4.12 | 5.36 | 6.08 | 6.59 |
| HPV16/18 | Opportunistic | Montpellier | 58 | 4.17 | 2.60 | 2.77 | 3.14 | 4.09 | 5.30 | 6.07 | 6.56 |
| HPV16/18 | Opportunistic | Montpellier | 59 | 4.16 | 2.59 | 2.76 | 3.13 | 4.08 | 5.27 | 6.04 | 6.55 |
| HPV16/18 | Opportunistic | Montpellier | 60 | 4.16 | 2.61 | 2.77 | 3.15 | 4.08 | 5.27 | 6.06 | 6.52 |
| HPV16/18 | Opportunistic | Montpellier | 61 | 4.17 | 2.61 | 2.77 | 3.14 | 4.07 | 5.28 | 6.07 | 6.58 |
| HPV16/18 | Opportunistic | Montpellier | 62 | 4.16 | 2.60 | 2.77 | 3.15 | 4.08 | 5.27 | 6.05 | 6.59 |
| HPV16/18 | Opportunistic | Montpellier | 63 | 4.15 | 2.59 | 2.75 | 3.13 | 4.06 | 5.25 | 6.01 | 6.61 |
| HPV16/18 | Opportunistic | Montpellier | 64 | 4.12 | 2.54 | 2.71 | 3.11 | 4.04 | 5.23 | 5.99 | 6.58 |
| HPV16/18 | Opportunistic | Montpellier | 65 | 4.08 | 2.55 | 2.67 | 3.06 | 3.99 | 5.20 | 5.93 | 6.49 |
| HPV16/18 | Opportunistic | Montpellier | 66 | 4.03 | 2.45 | 2.61 | 2.99 | 3.94 | 5.15 | 5.92 | 6.44 |
| HPV16/18 | Opportunistic | Rennes | 30 | 6.43 | 4.84 | 5.07 | 5.48 | 6.38 | 7.43 | 8.10 | 8.46 |
| HPV16/18 | Opportunistic | Rennes | 31 | 6.79 | 5.17 | 5.36 | 5.81 | 6.75 | 7.82 | 8.53 | 8.88 |
| HPV16/18 | Opportunistic | Rennes | 32 | 7.12 | 5.44 | 5.62 | 6.10 | 7.08 | 8.20 | 8.93 | 9.29 |
| HPV16/18 | Opportunistic | Rennes | 33 | 7.38 | 5.63 | 5.82 | 6.31 | 7.35 | 8.48 | 9.24 | 9.59 |
| HPV16/18 | Opportunistic | Rennes | 34 | 7.48 | 5.71 | 5.90 | 6.40 | 7.44 | 8.60 | 9.37 | 9.77 |
| HPV16/18 | Opportunistic | Rennes | 35 | 7.42 | 5.67 | 5.85 | 6.36 | 7.38 | 8.52 | 9.28 | 9.67 |
| HPV16/18 | Opportunistic | Rennes | 36 | 7.28 | 5.55 | 5.76 | 6.23 | 7.24 | 8.36 | 9.12 | 9.56 |
| HPV16/18 | Opportunistic | Rennes | 37 | 7.09 | 5.40 | 5.61 | 6.07 | 7.06 | 8.15 | 8.87 | 9.29 |
| HPV16/18 | Opportunistic | Rennes | 38 | 6.91 | 5.28 | 5.46 | 5.91 | 6.86 | 7.96 | 8.66 | 9.04 |
| HPV16/18 | Opportunistic | Rennes | 39 | 6.73 | 5.11 | 5.32 | 5.74 | 6.68 | 7.75 | 8.47 | 8.80 |
| HPV16/18 | Opportunistic | Rennes | 40 | 6.55 | 4.97 | 5.17 | 5.59 | 6.49 | 7.54 | 8.23 | 8.57 |
| HPV16/18 | Opportunistic | Rennes | 41 | 6.37 | 4.83 | 5.03 | 5.43 | 6.33 | 7.35 | 8.01 | 8.35 |
| HPV16/18 | Opportunistic | Rennes | 42 | 6.19 | 4.68 | 4.88 | 5.28 | 6.14 | 7.14 | 7.77 | 8.10 |
| HPV16/18 | Opportunistic | Rennes | 43 | 6.02 | 4.56 | 4.74 | 5.14 | 5.98 | 6.94 | 7.54 | 7.86 |
| HPV16/18 | Opportunistic | Rennes | 44 | 5.87 | 4.43 | 4.61 | 5.02 | 5.83 | 6.77 | 7.37 | 7.69 |

**Table S9** **Posterior expected HR HPV cervical infection prevalence (in %) in major French cities, stratified by type of test, city, and age. Table counterpart to Figure 3A** ***(continued)***

| | | | | Posterior distribution | | | | | | | |
|---|---|---|---|---|---|---|---|---|---|---|---|
| | | | | | Percentiles | | | | | | |
| Virus | Test | City | Age | Average | 0.01 | 0.025 | 0.1 | 0.5 | 0.9 | 0.975 | 0.99 |
| HPV16/18 | Opportunistic | Rennes | 45 | 5.75 | 4.34 | 4.50 | 4.91 | 5.72 | 6.61 | 7.21 | 7.53 |
| HPV16/18 | Opportunistic | Rennes | 46 | 5.61 | 4.23 | 4.40 | 4.80 | 5.58 | 6.47 | 7.05 | 7.40 |
| HPV16/18 | Opportunistic | Rennes | 47 | 5.48 | 4.13 | 4.30 | 4.69 | 5.44 | 6.33 | 6.91 | 7.22 |
| HPV16/18 | Opportunistic | Rennes | 48 | 5.36 | 4.06 | 4.20 | 4.57 | 5.32 | 6.20 | 6.74 | 7.06 |
| HPV16/18 | Opportunistic | Rennes | 49 | 5.24 | 3.99 | 4.11 | 4.48 | 5.20 | 6.05 | 6.62 | 6.91 |
| HPV16/18 | Opportunistic | Rennes | 50 | 5.12 | 3.87 | 4.02 | 4.38 | 5.09 | 5.91 | 6.47 | 6.78 |
| HPV16/18 | Opportunistic | Rennes | 51 | 5.01 | 3.79 | 3.93 | 4.28 | 4.98 | 5.78 | 6.33 | 6.61 |
| HPV16/18 | Opportunistic | Rennes | 52 | 4.91 | 3.70 | 3.85 | 4.18 | 4.88 | 5.67 | 6.19 | 6.45 |
| HPV16/18 | Opportunistic | Rennes | 53 | 4.79 | 3.60 | 3.75 | 4.07 | 4.75 | 5.54 | 6.02 | 6.33 |
| HPV16/18 | Opportunistic | Rennes | 54 | 4.67 | 3.52 | 3.66 | 3.96 | 4.63 | 5.40 | 5.89 | 6.14 |
| HPV16/18 | Opportunistic | Rennes | 55 | 4.56 | 3.43 | 3.58 | 3.89 | 4.53 | 5.29 | 5.76 | 6.03 |
| HPV16/18 | Opportunistic | Rennes | 56 | 4.48 | 3.39 | 3.52 | 3.82 | 4.45 | 5.19 | 5.65 | 5.90 |
| HPV16/18 | Opportunistic | Rennes | 57 | 4.43 | 3.34 | 3.47 | 3.77 | 4.39 | 5.13 | 5.60 | 5.84 |
| HPV16/18 | Opportunistic | Rennes | 58 | 4.39 | 3.30 | 3.43 | 3.74 | 4.36 | 5.10 | 5.56 | 5.76 |
| HPV16/18 | Opportunistic | Rennes | 59 | 4.38 | 3.29 | 3.43 | 3.72 | 4.34 | 5.09 | 5.54 | 5.80 |
| HPV16/18 | Opportunistic | Rennes | 60 | 4.38 | 3.30 | 3.44 | 3.73 | 4.34 | 5.09 | 5.56 | 5.77 |
| HPV16/18 | Opportunistic | Rennes | 61 | 4.39 | 3.30 | 3.46 | 3.73 | 4.35 | 5.09 | 5.57 | 5.75 |
| HPV16/18 | Opportunistic | Rennes | 62 | 4.38 | 3.29 | 3.43 | 3.72 | 4.35 | 5.07 | 5.55 | 5.77 |
| HPV16/18 | Opportunistic | Rennes | 63 | 4.36 | 3.27 | 3.41 | 3.71 | 4.33 | 5.06 | 5.51 | 5.79 |
| HPV16/18 | Opportunistic | Rennes | 64 | 4.33 | 3.23 | 3.36 | 3.68 | 4.30 | 5.03 | 5.48 | 5.79 |
| HPV16/18 | Opportunistic | Rennes | 65 | 4.29 | 3.19 | 3.31 | 3.63 | 4.26 | 5.01 | 5.46 | 5.75 |
| HPV16/18 | Opportunistic | Rennes | 66 | 4.24 | 3.08 | 3.22 | 3.55 | 4.20 | 4.98 | 5.45 | 5.76 |
| HPV16/18 | Opportunistic | Nantes | 30 | 5.47 | 3.31 | 3.52 | 4.04 | 5.34 | 7.10 | 8.08 | 8.70 |
| HPV16/18 | Opportunistic | Nantes | 31 | 5.78 | 3.47 | 3.77 | 4.30 | 5.66 | 7.46 | 8.53 | 9.25 |
| HPV16/18 | Opportunistic | Nantes | 32 | 6.06 | 3.64 | 3.99 | 4.51 | 5.94 | 7.81 | 8.94 | 9.68 |
| HPV16/18 | Opportunistic | Nantes | 33 | 6.28 | 3.82 | 4.13 | 4.67 | 6.13 | 8.06 | 9.24 | 9.95 |
| HPV16/18 | Opportunistic | Nantes | 34 | 6.37 | 3.87 | 4.17 | 4.72 | 6.21 | 8.19 | 9.38 | 10.05 |
| HPV16/18 | Opportunistic | Nantes | 35 | 6.32 | 3.85 | 4.14 | 4.69 | 6.17 | 8.13 | 9.32 | 9.98 |
| HPV16/18 | Opportunistic | Nantes | 36 | 6.19 | 3.79 | 4.04 | 4.60 | 6.05 | 7.97 | 9.13 | 9.86 |

**Table S9** **Posterior expected HR HPV cervical infection prevalence (in %) in major French cities, stratified by type of test, city, and age. Table counterpart to Figure 3A** ***(continued)***

| | | | | Posterior distribution | | | | | | | |
|---|---|---|---|---|---|---|---|---|---|---|---|
| | | | | | Percentiles | | | | | | |
| Virus | Test | City | Age | Average | 0.01 | 0.025 | 0.1 | 0.5 | 0.9 | 0.975 | 0.99 |
| HPV16/18 | Opportunistic | Nantes | 37 | 6.03 | 3.72 | 3.95 | 4.48 | 5.90 | 7.77 | 8.89 | 9.55 |
| HPV16/18 | Opportunistic | Nantes | 38 | 5.88 | 3.60 | 3.85 | 4.36 | 5.74 | 7.59 | 8.69 | 9.36 |
| HPV16/18 | Opportunistic | Nantes | 39 | 5.72 | 3.48 | 3.72 | 4.25 | 5.58 | 7.39 | 8.47 | 9.16 |
| HPV16/18 | Opportunistic | Nantes | 40 | 5.57 | 3.39 | 3.61 | 4.13 | 5.43 | 7.20 | 8.23 | 8.79 |
| HPV16/18 | Opportunistic | Nantes | 41 | 5.42 | 3.28 | 3.52 | 4.01 | 5.28 | 6.99 | 8.02 | 8.60 |
| HPV16/18 | Opportunistic | Nantes | 42 | 5.26 | 3.18 | 3.44 | 3.90 | 5.13 | 6.80 | 7.79 | 8.31 |
| HPV16/18 | Opportunistic | Nantes | 43 | 5.11 | 3.11 | 3.32 | 3.78 | 5.00 | 6.60 | 7.57 | 8.16 |
| HPV16/18 | Opportunistic | Nantes | 44 | 4.99 | 3.02 | 3.25 | 3.69 | 4.87 | 6.45 | 7.40 | 7.93 |
| HPV16/18 | Opportunistic | Nantes | 45 | 4.88 | 2.95 | 3.17 | 3.61 | 4.76 | 6.33 | 7.23 | 7.78 |
| HPV16/18 | Opportunistic | Nantes | 46 | 4.76 | 2.88 | 3.10 | 3.53 | 4.65 | 6.20 | 7.07 | 7.63 |
| HPV16/18 | Opportunistic | Nantes | 47 | 4.65 | 2.80 | 3.02 | 3.45 | 4.54 | 6.06 | 6.91 | 7.50 |
| HPV16/18 | Opportunistic | Nantes | 48 | 4.55 | 2.73 | 2.94 | 3.36 | 4.43 | 5.91 | 6.75 | 7.26 |
| HPV16/18 | Opportunistic | Nantes | 49 | 4.45 | 2.69 | 2.88 | 3.29 | 4.33 | 5.76 | 6.60 | 7.12 |
| HPV16/18 | Opportunistic | Nantes | 50 | 4.35 | 2.62 | 2.81 | 3.22 | 4.24 | 5.64 | 6.46 | 6.97 |
| HPV16/18 | Opportunistic | Nantes | 51 | 4.25 | 2.57 | 2.75 | 3.14 | 4.15 | 5.54 | 6.32 | 6.83 |
| HPV16/18 | Opportunistic | Nantes | 52 | 4.16 | 2.51 | 2.70 | 3.08 | 4.06 | 5.41 | 6.16 | 6.70 |
| HPV16/18 | Opportunistic | Nantes | 53 | 4.06 | 2.44 | 2.62 | 2.99 | 3.96 | 5.27 | 6.03 | 6.56 |
| HPV16/18 | Opportunistic | Nantes | 54 | 3.96 | 2.37 | 2.54 | 2.92 | 3.86 | 5.14 | 5.87 | 6.37 |
| HPV16/18 | Opportunistic | Nantes | 55 | 3.87 | 2.32 | 2.48 | 2.85 | 3.77 | 5.02 | 5.73 | 6.22 |
| HPV16/18 | Opportunistic | Nantes | 56 | 3.80 | 2.26 | 2.44 | 2.80 | 3.71 | 4.93 | 5.64 | 6.13 |
| HPV16/18 | Opportunistic | Nantes | 57 | 3.76 | 2.23 | 2.41 | 2.76 | 3.66 | 4.88 | 5.60 | 6.09 |
| HPV16/18 | Opportunistic | Nantes | 58 | 3.73 | 2.22 | 2.39 | 2.74 | 3.62 | 4.83 | 5.55 | 6.11 |
| HPV16/18 | Opportunistic | Nantes | 59 | 3.71 | 2.20 | 2.38 | 2.73 | 3.62 | 4.82 | 5.56 | 6.08 |
| HPV16/18 | Opportunistic | Nantes | 60 | 3.71 | 2.22 | 2.39 | 2.73 | 3.63 | 4.82 | 5.55 | 6.05 |
| HPV16/18 | Opportunistic | Nantes | 61 | 3.72 | 2.23 | 2.40 | 2.73 | 3.63 | 4.84 | 5.58 | 6.05 |
| HPV16/18 | Opportunistic | Nantes | 62 | 3.72 | 2.23 | 2.39 | 2.73 | 3.63 | 4.81 | 5.55 | 6.01 |
| HPV16/18 | Opportunistic | Nantes | 63 | 3.70 | 2.20 | 2.39 | 2.72 | 3.62 | 4.78 | 5.53 | 5.98 |
| HPV16/18 | Opportunistic | Nantes | 64 | 3.67 | 2.20 | 2.36 | 2.69 | 3.59 | 4.75 | 5.53 | 5.96 |
| HPV16/18 | Opportunistic | Nantes | 65 | 3.64 | 2.18 | 2.33 | 2.66 | 3.55 | 4.74 | 5.52 | 5.94 |

**Table S9** **Posterior expected HR HPV cervical infection prevalence (in %) in major French cities, stratified by type of test, city, and age. Table counterpart to Figure 3A** ***(continued)***

| | | | | Posterior distribution | | | | | | | |
|---|---|---|---|---|---|---|---|---|---|---|---|
| | | | | | Percentiles | | | | | | |
| Virus | Test | City | Age | Average | 0.01 | 0.025 | 0.1 | 0.5 | 0.9 | 0.975 | 0.99 |
| HPV16/18 | Opportunistic | Nantes | 66 | 3.60 | 2.14 | 2.27 | 2.62 | 3.51 | 4.67 | 5.46 | 5.90 |
| HPV16/18 | Opportunistic | Lille | 30 | 5.83 | 3.73 | 4.01 | 4.54 | 5.74 | 7.25 | 8.11 | 8.62 |
| HPV16/18 | Opportunistic | Lille | 31 | 6.17 | 3.94 | 4.25 | 4.82 | 6.08 | 7.62 | 8.57 | 9.04 |
| HPV16/18 | Opportunistic | Lille | 32 | 6.47 | 4.14 | 4.44 | 5.06 | 6.38 | 8.00 | 8.96 | 9.50 |
| HPV16/18 | Opportunistic | Lille | 33 | 6.70 | 4.28 | 4.61 | 5.24 | 6.59 | 8.30 | 9.25 | 9.80 |
| HPV16/18 | Opportunistic | Lille | 34 | 6.79 | 4.35 | 4.68 | 5.32 | 6.68 | 8.41 | 9.42 | 9.97 |
| HPV16/18 | Opportunistic | Lille | 35 | 6.74 | 4.28 | 4.66 | 5.28 | 6.64 | 8.34 | 9.38 | 9.90 |
| HPV16/18 | Opportunistic | Lille | 36 | 6.61 | 4.22 | 4.57 | 5.18 | 6.50 | 8.17 | 9.20 | 9.71 |
| HPV16/18 | Opportunistic | Lille | 37 | 6.44 | 4.11 | 4.42 | 5.03 | 6.34 | 7.94 | 8.97 | 9.52 |
| HPV16/18 | Opportunistic | Lille | 38 | 6.27 | 4.04 | 4.34 | 4.91 | 6.17 | 7.75 | 8.72 | 9.28 |
| HPV16/18 | Opportunistic | Lille | 39 | 6.11 | 3.93 | 4.22 | 4.77 | 6.00 | 7.55 | 8.49 | 9.01 |
| HPV16/18 | Opportunistic | Lille | 40 | 5.94 | 3.83 | 4.12 | 4.64 | 5.85 | 7.36 | 8.28 | 8.77 |
| HPV16/18 | Opportunistic | Lille | 41 | 5.78 | 3.68 | 3.99 | 4.51 | 5.70 | 7.19 | 8.04 | 8.55 |
| HPV16/18 | Opportunistic | Lille | 42 | 5.61 | 3.56 | 3.86 | 4.37 | 5.53 | 6.98 | 7.81 | 8.34 |
| HPV16/18 | Opportunistic | Lille | 43 | 5.46 | 3.44 | 3.74 | 4.25 | 5.37 | 6.78 | 7.64 | 8.15 |
| HPV16/18 | Opportunistic | Lille | 44 | 5.32 | 3.36 | 3.64 | 4.14 | 5.24 | 6.62 | 7.48 | 7.92 |
| HPV16/18 | Opportunistic | Lille | 45 | 5.21 | 3.29 | 3.56 | 4.06 | 5.12 | 6.47 | 7.31 | 7.72 |
| HPV16/18 | Opportunistic | Lille | 46 | 5.09 | 3.22 | 3.49 | 3.97 | 5.00 | 6.32 | 7.15 | 7.54 |
| HPV16/18 | Opportunistic | Lille | 47 | 4.97 | 3.17 | 3.41 | 3.88 | 4.89 | 6.16 | 6.96 | 7.43 |
| HPV16/18 | Opportunistic | Lille | 48 | 4.86 | 3.05 | 3.32 | 3.80 | 4.78 | 6.02 | 6.78 | 7.22 |
| HPV16/18 | Opportunistic | Lille | 49 | 4.75 | 3.00 | 3.26 | 3.72 | 4.68 | 5.91 | 6.64 | 7.05 |
| HPV16/18 | Opportunistic | Lille | 50 | 4.64 | 2.95 | 3.17 | 3.63 | 4.58 | 5.77 | 6.48 | 6.92 |
| HPV16/18 | Opportunistic | Lille | 51 | 4.54 | 2.89 | 3.11 | 3.54 | 4.47 | 5.65 | 6.37 | 6.80 |
| HPV16/18 | Opportunistic | Lille | 52 | 4.45 | 2.82 | 3.05 | 3.46 | 4.37 | 5.51 | 6.21 | 6.63 |
| HPV16/18 | Opportunistic | Lille | 53 | 4.34 | 2.73 | 2.97 | 3.37 | 4.27 | 5.38 | 6.08 | 6.48 |
| HPV16/18 | Opportunistic | Lille | 54 | 4.23 | 2.67 | 2.90 | 3.29 | 4.16 | 5.27 | 5.92 | 6.29 |
| HPV16/18 | Opportunistic | Lille | 55 | 4.13 | 2.59 | 2.84 | 3.21 | 4.06 | 5.14 | 5.81 | 6.16 |
| HPV16/18 | Opportunistic | Lille | 56 | 4.06 | 2.55 | 2.79 | 3.16 | 3.99 | 5.07 | 5.73 | 6.08 |
| HPV16/18 | Opportunistic | Lille | 57 | 4.01 | 2.51 | 2.77 | 3.11 | 3.93 | 5.01 | 5.66 | 5.96 |

**Table S9** **Posterior expected HR HPV cervical infection prevalence (in %) in major French cities, stratified by type of test, city, and age. Table counterpart to Figure 3A *(continued)***

| | | | | Posterior distribution | | | | | | | |
|---|---|---|---|---|---|---|---|---|---|---|---|
| | | | | | Percentiles | | | | | | |
| Virus | Test | City | Age | Average | 0.01 | 0.025 | 0.1 | 0.5 | 0.9 | 0.975 | 0.99 |
| HPV16/18 | Opportunistic | Lille | 58 | 3.98 | 2.50 | 2.75 | 3.09 | 3.90 | 4.97 | 5.61 | 5.89 |
| HPV16/18 | Opportunistic | Lille | 59 | 3.97 | 2.47 | 2.74 | 3.07 | 3.90 | 4.94 | 5.59 | 5.88 |
| HPV16/18 | Opportunistic | Lille | 60 | 3.97 | 2.50 | 2.74 | 3.08 | 3.90 | 4.95 | 5.61 | 5.91 |
| HPV16/18 | Opportunistic | Lille | 61 | 3.97 | 2.52 | 2.73 | 3.07 | 3.91 | 4.96 | 5.60 | 5.93 |
| HPV16/18 | Opportunistic | Lille | 62 | 3.97 | 2.52 | 2.72 | 3.07 | 3.91 | 4.95 | 5.60 | 5.89 |
| HPV16/18 | Opportunistic | Lille | 63 | 3.95 | 2.51 | 2.72 | 3.06 | 3.89 | 4.93 | 5.55 | 5.89 |
| HPV16/18 | Opportunistic | Lille | 64 | 3.92 | 2.49 | 2.69 | 3.03 | 3.86 | 4.91 | 5.52 | 5.94 |
| HPV16/18 | Opportunistic | Lille | 65 | 3.89 | 2.46 | 2.65 | 3.00 | 3.82 | 4.87 | 5.48 | 5.95 |
| HPV16/18 | Opportunistic | Lille | 66 | 3.84 | 2.38 | 2.59 | 2.94 | 3.76 | 4.84 | 5.50 | 5.91 |
| HPV16/18 | Opportunistic | Strasbourg | 30 | 4.66 | 2.62 | 2.85 | 3.35 | 4.53 | 6.14 | 7.25 | 7.86 |
| HPV16/18 | Opportunistic | Strasbourg | 31 | 4.93 | 2.75 | 3.04 | 3.54 | 4.80 | 6.49 | 7.62 | 8.37 |
| HPV16/18 | Opportunistic | Strasbourg | 32 | 5.17 | 2.90 | 3.18 | 3.71 | 5.05 | 6.80 | 7.98 | 8.76 |
| HPV16/18 | Opportunistic | Strasbourg | 33 | 5.36 | 2.98 | 3.29 | 3.84 | 5.22 | 7.05 | 8.28 | 9.01 |
| HPV16/18 | Opportunistic | Strasbourg | 34 | 5.44 | 3.04 | 3.34 | 3.88 | 5.29 | 7.14 | 8.37 | 9.20 |
| HPV16/18 | Opportunistic | Strasbourg | 35 | 5.39 | 3.03 | 3.33 | 3.85 | 5.24 | 7.09 | 8.33 | 9.14 |
| HPV16/18 | Opportunistic | Strasbourg | 36 | 5.29 | 2.97 | 3.26 | 3.78 | 5.14 | 6.93 | 8.18 | 8.96 |
| HPV16/18 | Opportunistic | Strasbourg | 37 | 5.15 | 2.87 | 3.17 | 3.69 | 5.00 | 6.74 | 7.98 | 8.81 |
| HPV16/18 | Opportunistic | Strasbourg | 38 | 5.01 | 2.78 | 3.08 | 3.59 | 4.87 | 6.55 | 7.79 | 8.59 |
| HPV16/18 | Opportunistic | Strasbourg | 39 | 4.88 | 2.70 | 2.99 | 3.50 | 4.74 | 6.40 | 7.58 | 8.26 |
| HPV16/18 | Opportunistic | Strasbourg | 40 | 4.75 | 2.64 | 2.90 | 3.40 | 4.62 | 6.22 | 7.33 | 8.00 |
| HPV16/18 | Opportunistic | Strasbourg | 41 | 4.62 | 2.56 | 2.83 | 3.30 | 4.49 | 6.06 | 7.18 | 7.76 |
| HPV16/18 | Opportunistic | Strasbourg | 42 | 4.48 | 2.49 | 2.74 | 3.20 | 4.36 | 5.90 | 6.98 | 7.52 |
| HPV16/18 | Opportunistic | Strasbourg | 43 | 4.36 | 2.40 | 2.66 | 3.10 | 4.24 | 5.74 | 6.78 | 7.36 |
| HPV16/18 | Opportunistic | Strasbourg | 44 | 4.25 | 2.35 | 2.60 | 3.02 | 4.13 | 5.58 | 6.57 | 7.23 |
| HPV16/18 | Opportunistic | Strasbourg | 45 | 4.16 | 2.29 | 2.55 | 2.97 | 4.03 | 5.47 | 6.50 | 7.05 |
| HPV16/18 | Opportunistic | Strasbourg | 46 | 4.06 | 2.25 | 2.47 | 2.90 | 3.94 | 5.32 | 6.32 | 6.93 |
| HPV16/18 | Opportunistic | Strasbourg | 47 | 3.96 | 2.20 | 2.41 | 2.82 | 3.84 | 5.18 | 6.20 | 6.82 |
| HPV16/18 | Opportunistic | Strasbourg | 48 | 3.87 | 2.13 | 2.36 | 2.75 | 3.75 | 5.06 | 6.06 | 6.63 |
| HPV16/18 | Opportunistic | Strasbourg | 49 | 3.78 | 2.08 | 2.30 | 2.69 | 3.67 | 4.96 | 5.91 | 6.45 |

**Table S9 Posterior expected HR HPV cervical infection prevalence (in %) in major French cities, stratified by type of test, city, and age. Table counterpart to Figure 3A** ***(continued)***

| | | | | Posterior distribution | | | | | | | |
|---|---|---|---|---|---|---|---|---|---|---|---|
| | | | | | Percentiles | | | | | | |
| Virus | Test | City | Age | Average | 0.01 | 0.025 | 0.1 | 0.5 | 0.9 | 0.975 | 0.99 |
| HPV16/18 | Opportunistic | Strasbourg | 50 | 3.70 | 2.05 | 2.25 | 2.63 | 3.58 | 4.86 | 5.79 | 6.30 |
| HPV16/18 | Opportunistic | Strasbourg | 51 | 3.62 | 2.00 | 2.21 | 2.57 | 3.51 | 4.76 | 5.65 | 6.12 |
| HPV16/18 | Opportunistic | Strasbourg | 52 | 3.54 | 1.95 | 2.15 | 2.51 | 3.43 | 4.67 | 5.54 | 6.10 |
| HPV16/18 | Opportunistic | Strasbourg | 53 | 3.46 | 1.90 | 2.10 | 2.45 | 3.35 | 4.54 | 5.43 | 5.97 |
| HPV16/18 | Opportunistic | Strasbourg | 54 | 3.37 | 1.84 | 2.06 | 2.38 | 3.26 | 4.43 | 5.26 | 5.84 |
| HPV16/18 | Opportunistic | Strasbourg | 55 | 3.29 | 1.79 | 2.00 | 2.34 | 3.19 | 4.34 | 5.15 | 5.70 |
| HPV16/18 | Opportunistic | Strasbourg | 56 | 3.23 | 1.75 | 1.97 | 2.30 | 3.14 | 4.25 | 5.04 | 5.54 |
| HPV16/18 | Opportunistic | Strasbourg | 57 | 3.19 | 1.74 | 1.94 | 2.27 | 3.10 | 4.21 | 5.00 | 5.47 |
| HPV16/18 | Opportunistic | Strasbourg | 58 | 3.17 | 1.73 | 1.93 | 2.25 | 3.08 | 4.18 | 4.96 | 5.42 |
| HPV16/18 | Opportunistic | Strasbourg | 59 | 3.16 | 1.71 | 1.92 | 2.24 | 3.06 | 4.17 | 4.97 | 5.42 |
| HPV16/18 | Opportunistic | Strasbourg | 60 | 3.16 | 1.74 | 1.93 | 2.24 | 3.06 | 4.16 | 4.98 | 5.42 |
| HPV16/18 | Opportunistic | Strasbourg | 61 | 3.16 | 1.74 | 1.93 | 2.26 | 3.07 | 4.17 | 4.97 | 5.36 |
| HPV16/18 | Opportunistic | Strasbourg | 62 | 3.16 | 1.75 | 1.92 | 2.25 | 3.06 | 4.17 | 4.98 | 5.46 |
| HPV16/18 | Opportunistic | Strasbourg | 63 | 3.14 | 1.75 | 1.90 | 2.25 | 3.04 | 4.15 | 4.94 | 5.39 |
| HPV16/18 | Opportunistic | Strasbourg | 64 | 3.12 | 1.72 | 1.88 | 2.23 | 3.02 | 4.14 | 4.91 | 5.37 |
| HPV16/18 | Opportunistic | Strasbourg | 65 | 3.09 | 1.70 | 1.85 | 2.20 | 3.00 | 4.12 | 4.86 | 5.33 |
| HPV16/18 | Opportunistic | Strasbourg | 66 | 3.06 | 1.66 | 1.80 | 2.16 | 2.97 | 4.10 | 4.84 | 5.36 |
| HPV16/18 | Opportunistic | Lyon | 30 | 6.32 | 3.82 | 4.13 | 4.75 | 6.20 | 8.07 | 9.39 | 9.89 |
| HPV16/18 | Opportunistic | Lyon | 31 | 6.68 | 4.05 | 4.36 | 5.02 | 6.55 | 8.52 | 9.81 | 10.54 |
| HPV16/18 | Opportunistic | Lyon | 32 | 7.01 | 4.25 | 4.62 | 5.29 | 6.85 | 8.93 | 10.24 | 10.94 |
| HPV16/18 | Opportunistic | Lyon | 33 | 7.25 | 4.40 | 4.77 | 5.47 | 7.08 | 9.24 | 10.56 | 11.42 |
| HPV16/18 | Opportunistic | Lyon | 34 | 7.35 | 4.49 | 4.85 | 5.54 | 7.19 | 9.39 | 10.69 | 11.55 |
| HPV16/18 | Opportunistic | Lyon | 35 | 7.30 | 4.44 | 4.77 | 5.50 | 7.12 | 9.31 | 10.62 | 11.46 |
| HPV16/18 | Opportunistic | Lyon | 36 | 7.16 | 4.33 | 4.68 | 5.39 | 7.00 | 9.13 | 10.47 | 11.23 |
| HPV16/18 | Opportunistic | Lyon | 37 | 6.98 | 4.23 | 4.57 | 5.24 | 6.82 | 8.92 | 10.20 | 10.91 |
| HPV16/18 | Opportunistic | Lyon | 38 | 6.80 | 4.13 | 4.45 | 5.10 | 6.65 | 8.72 | 9.95 | 10.64 |
| HPV16/18 | Opportunistic | Lyon | 39 | 6.62 | 4.03 | 4.33 | 4.96 | 6.47 | 8.48 | 9.71 | 10.38 |
| HPV16/18 | Opportunistic | Lyon | 40 | 6.44 | 3.90 | 4.22 | 4.84 | 6.29 | 8.24 | 9.47 | 10.12 |
| HPV16/18 | Opportunistic | Lyon | 41 | 6.27 | 3.79 | 4.08 | 4.70 | 6.13 | 8.02 | 9.19 | 9.86 |

**Table S9** **Posterior expected HR HPV cervical infection prevalence (in %) in major French cities, stratified by type of test, city, and age. Table counterpart to Figure 3A** ***(continued)***

| | | | | Posterior distribution | | | | | | | |
|---|---|---|---|---|---|---|---|---|---|---|---|
| | | | | | Percentiles | | | | | | |
| Virus | Test | City | Age | Average | 0.01 | 0.025 | 0.1 | 0.5 | 0.9 | 0.975 | 0.99 |
| HPV16/18 | Opportunistic | Lyon | 42 | 6.09 | 3.68 | 3.98 | 4.57 | 5.95 | 7.79 | 8.96 | 9.56 |
| HPV16/18 | Opportunistic | Lyon | 43 | 5.92 | 3.57 | 3.86 | 4.44 | 5.78 | 7.59 | 8.72 | 9.32 |
| HPV16/18 | Opportunistic | Lyon | 44 | 5.77 | 3.48 | 3.78 | 4.34 | 5.63 | 7.37 | 8.48 | 9.15 |
| HPV16/18 | Opportunistic | Lyon | 45 | 5.65 | 3.41 | 3.69 | 4.24 | 5.51 | 7.22 | 8.30 | 9.00 |
| HPV16/18 | Opportunistic | Lyon | 46 | 5.52 | 3.31 | 3.60 | 4.14 | 5.38 | 7.07 | 8.12 | 8.72 |
| HPV16/18 | Opportunistic | Lyon | 47 | 5.39 | 3.24 | 3.51 | 4.04 | 5.27 | 6.93 | 7.94 | 8.48 |
| HPV16/18 | Opportunistic | Lyon | 48 | 5.27 | 3.15 | 3.43 | 3.93 | 5.15 | 6.75 | 7.81 | 8.30 |
| HPV16/18 | Opportunistic | Lyon | 49 | 5.15 | 3.12 | 3.35 | 3.84 | 5.03 | 6.61 | 7.62 | 8.10 |
| HPV16/18 | Opportunistic | Lyon | 50 | 5.04 | 3.04 | 3.29 | 3.76 | 4.92 | 6.47 | 7.43 | 7.91 |
| HPV16/18 | Opportunistic | Lyon | 51 | 4.93 | 2.97 | 3.22 | 3.67 | 4.82 | 6.34 | 7.28 | 7.75 |
| HPV16/18 | Opportunistic | Lyon | 52 | 4.83 | 2.91 | 3.15 | 3.61 | 4.72 | 6.21 | 7.12 | 7.59 |
| HPV16/18 | Opportunistic | Lyon | 53 | 4.71 | 2.83 | 3.06 | 3.53 | 4.60 | 6.05 | 6.94 | 7.43 |
| HPV16/18 | Opportunistic | Lyon | 54 | 4.59 | 2.78 | 2.98 | 3.43 | 4.48 | 5.89 | 6.79 | 7.24 |
| HPV16/18 | Opportunistic | Lyon | 55 | 4.49 | 2.71 | 2.91 | 3.36 | 4.38 | 5.75 | 6.68 | 7.11 |
| HPV16/18 | Opportunistic | Lyon | 56 | 4.41 | 2.67 | 2.86 | 3.29 | 4.31 | 5.66 | 6.57 | 7.02 |
| HPV16/18 | Opportunistic | Lyon | 57 | 4.36 | 2.63 | 2.82 | 3.25 | 4.24 | 5.59 | 6.49 | 6.91 |
| HPV16/18 | Opportunistic | Lyon | 58 | 4.32 | 2.60 | 2.79 | 3.22 | 4.21 | 5.56 | 6.48 | 6.84 |
| HPV16/18 | Opportunistic | Lyon | 59 | 4.31 | 2.57 | 2.78 | 3.22 | 4.21 | 5.56 | 6.41 | 6.86 |
| HPV16/18 | Opportunistic | Lyon | 60 | 4.31 | 2.57 | 2.77 | 3.21 | 4.21 | 5.54 | 6.41 | 6.86 |
| HPV16/18 | Opportunistic | Lyon | 61 | 4.32 | 2.58 | 2.78 | 3.22 | 4.22 | 5.56 | 6.45 | 6.88 |
| HPV16/18 | Opportunistic | Lyon | 62 | 4.31 | 2.58 | 2.79 | 3.21 | 4.22 | 5.54 | 6.46 | 6.87 |
| HPV16/18 | Opportunistic | Lyon | 63 | 4.29 | 2.57 | 2.77 | 3.18 | 4.20 | 5.53 | 6.43 | 6.87 |
| HPV16/18 | Opportunistic | Lyon | 64 | 4.26 | 2.55 | 2.77 | 3.16 | 4.16 | 5.48 | 6.40 | 6.78 |
| HPV16/18 | Opportunistic | Lyon | 65 | 4.22 | 2.50 | 2.73 | 3.13 | 4.11 | 5.47 | 6.38 | 6.74 |
| HPV16/18 | Opportunistic | Lyon | 66 | 4.18 | 2.42 | 2.65 | 3.06 | 4.05 | 5.43 | 6.35 | 6.71 |
| HPV16/18 | Opportunistic | Paris | 30 | 6.70 | 5.01 | 5.23 | 5.69 | 6.65 | 7.79 | 8.41 | 8.71 |
| HPV16/18 | Opportunistic | Paris | 31 | 7.08 | 5.29 | 5.59 | 6.04 | 7.02 | 8.17 | 8.84 | 9.12 |
| HPV16/18 | Opportunistic | Paris | 32 | 7.42 | 5.61 | 5.87 | 6.35 | 7.37 | 8.56 | 9.25 | 9.53 |
| HPV16/18 | Opportunistic | Paris | 33 | 7.69 | 5.77 | 6.07 | 6.58 | 7.63 | 8.85 | 9.58 | 9.87 |

**Table S9** **Posterior expected HR HPV cervical infection prevalence (in %) in major French cities, stratified by type of test, city, and age. Table counterpart to Figure 3A** ***(continued)***

| | | | | Posterior distribution | | | | | | | |
|---|---|---|---|---|---|---|---|---|---|---|---|
| | | | | | Percentiles | | | | | | |
| Virus | Test | City | Age | Average | 0.01 | 0.025 | 0.1 | 0.5 | 0.9 | 0.975 | 0.99 |
| HPV16/18 | Opportunistic | Paris | 34 | 7.79 | 5.91 | 6.17 | 6.67 | 7.73 | 8.99 | 9.71 | 10.01 |
| HPV16/18 | Opportunistic | Paris | 35 | 7.73 | 5.84 | 6.11 | 6.60 | 7.68 | 8.93 | 9.64 | 9.92 |
| HPV16/18 | Opportunistic | Paris | 36 | 7.58 | 5.70 | 6.03 | 6.48 | 7.53 | 8.73 | 9.50 | 9.75 |
| HPV16/18 | Opportunistic | Paris | 37 | 7.39 | 5.53 | 5.86 | 6.31 | 7.34 | 8.52 | 9.24 | 9.56 |
| HPV16/18 | Opportunistic | Paris | 38 | 7.20 | 5.38 | 5.70 | 6.16 | 7.15 | 8.31 | 9.00 | 9.30 |
| HPV16/18 | Opportunistic | Paris | 39 | 7.01 | 5.23 | 5.52 | 6.00 | 6.97 | 8.10 | 8.78 | 9.06 |
| HPV16/18 | Opportunistic | Paris | 40 | 6.82 | 5.12 | 5.38 | 5.83 | 6.78 | 7.87 | 8.54 | 8.86 |
| HPV16/18 | Opportunistic | Paris | 41 | 6.64 | 4.98 | 5.23 | 5.66 | 6.60 | 7.68 | 8.29 | 8.62 |
| HPV16/18 | Opportunistic | Paris | 42 | 6.45 | 4.84 | 5.08 | 5.49 | 6.41 | 7.47 | 8.05 | 8.38 |
| HPV16/18 | Opportunistic | Paris | 43 | 6.27 | 4.71 | 4.92 | 5.35 | 6.24 | 7.26 | 7.83 | 8.14 |
| HPV16/18 | Opportunistic | Paris | 44 | 6.12 | 4.60 | 4.80 | 5.21 | 6.08 | 7.09 | 7.66 | 7.88 |
| HPV16/18 | Opportunistic | Paris | 45 | 5.99 | 4.49 | 4.70 | 5.10 | 5.95 | 6.96 | 7.51 | 7.73 |
| HPV16/18 | Opportunistic | Paris | 46 | 5.85 | 4.38 | 4.58 | 4.98 | 5.81 | 6.78 | 7.34 | 7.61 |
| HPV16/18 | Opportunistic | Paris | 47 | 5.71 | 4.28 | 4.47 | 4.87 | 5.67 | 6.62 | 7.17 | 7.40 |
| HPV16/18 | Opportunistic | Paris | 48 | 5.59 | 4.21 | 4.36 | 4.76 | 5.55 | 6.47 | 7.01 | 7.25 |
| HPV16/18 | Opportunistic | Paris | 49 | 5.46 | 4.09 | 4.26 | 4.65 | 5.42 | 6.34 | 6.85 | 7.09 |
| HPV16/18 | Opportunistic | Paris | 50 | 5.34 | 3.99 | 4.18 | 4.55 | 5.31 | 6.21 | 6.68 | 6.93 |
| HPV16/18 | Opportunistic | Paris | 51 | 5.23 | 3.89 | 4.09 | 4.46 | 5.19 | 6.07 | 6.55 | 6.77 |
| HPV16/18 | Opportunistic | Paris | 52 | 5.12 | 3.82 | 4.00 | 4.36 | 5.08 | 5.94 | 6.43 | 6.66 |
| HPV16/18 | Opportunistic | Paris | 53 | 4.99 | 3.74 | 3.90 | 4.25 | 4.96 | 5.79 | 6.29 | 6.49 |
| HPV16/18 | Opportunistic | Paris | 54 | 4.87 | 3.65 | 3.79 | 4.14 | 4.84 | 5.63 | 6.14 | 6.34 |
| HPV16/18 | Opportunistic | Paris | 55 | 4.76 | 3.57 | 3.71 | 4.04 | 4.73 | 5.51 | 5.99 | 6.21 |
| HPV16/18 | Opportunistic | Paris | 56 | 4.68 | 3.49 | 3.63 | 3.96 | 4.65 | 5.42 | 5.89 | 6.08 |
| HPV16/18 | Opportunistic | Paris | 57 | 4.62 | 3.44 | 3.59 | 3.91 | 4.60 | 5.37 | 5.83 | 6.01 |
| HPV16/18 | Opportunistic | Paris | 58 | 4.58 | 3.43 | 3.56 | 3.88 | 4.56 | 5.33 | 5.79 | 5.96 |
| HPV16/18 | Opportunistic | Paris | 59 | 4.57 | 3.39 | 3.55 | 3.86 | 4.55 | 5.32 | 5.78 | 5.97 |
| HPV16/18 | Opportunistic | Paris | 60 | 4.57 | 3.38 | 3.56 | 3.86 | 4.54 | 5.32 | 5.78 | 5.97 |
| HPV16/18 | Opportunistic | Paris | 61 | 4.58 | 3.41 | 3.57 | 3.88 | 4.55 | 5.33 | 5.80 | 6.01 |
| HPV16/18 | Opportunistic | Paris | 62 | 4.57 | 3.40 | 3.57 | 3.88 | 4.54 | 5.32 | 5.79 | 6.00 |

**Table S9 Posterior expected HR HPV cervical infection prevalence (in %) in major French cities, stratified by type of test, city, and age. Table counterpart to Figure 3A** ***(continued)***

| | | | | Posterior distribution | | | | | | | |
|---|---|---|---|---|---|---|---|---|---|---|---|
| | | | | | Percentiles | | | | | | |
| Virus | Test | City | Age | Average | 0.01 | 0.025 | 0.1 | 0.5 | 0.9 | 0.975 | 0.99 |
| HPV16/18 | Opportunistic | Paris | 63 | 4.55 | 3.38 | 3.55 | 3.86 | 4.52 | 5.30 | 5.77 | 5.96 |
| HPV16/18 | Opportunistic | Paris | 64 | 4.52 | 3.35 | 3.51 | 3.81 | 4.49 | 5.27 | 5.75 | 5.91 |
| HPV16/18 | Opportunistic | Paris | 65 | 4.48 | 3.28 | 3.44 | 3.76 | 4.44 | 5.26 | 5.70 | 5.96 |
| HPV16/18 | Opportunistic | Paris | 66 | 4.43 | 3.18 | 3.35 | 3.69 | 4.39 | 5.23 | 5.70 | 5.94 |
| Other genotypes | Organised | Nice | 30 | 12.88 | 6.80 | 7.47 | 8.98 | 12.49 | 17.12 | 20.26 | 22.18 |
| Other genotypes | Organised | Nice | 31 | 12.26 | 6.51 | 7.16 | 8.52 | 11.90 | 16.33 | 19.41 | 21.17 |
| Other genotypes | Organised | Nice | 32 | 11.61 | 6.23 | 6.78 | 8.06 | 11.28 | 15.51 | 18.40 | 20.01 |
| Other genotypes | Organised | Nice | 33 | 10.96 | 5.84 | 6.36 | 7.61 | 10.69 | 14.65 | 17.26 | 18.88 |
| Other genotypes | Organised | Nice | 34 | 10.42 | 5.53 | 6.01 | 7.20 | 10.16 | 13.91 | 16.36 | 18.04 |
| Other genotypes | Organised | Nice | 35 | 9.98 | 5.28 | 5.79 | 6.93 | 9.70 | 13.36 | 15.81 | 17.30 |
| Other genotypes | Organised | Nice | 36 | 9.60 | 5.07 | 5.56 | 6.68 | 9.33 | 12.86 | 15.23 | 16.72 |
| Other genotypes | Organised | Nice | 37 | 9.29 | 4.91 | 5.36 | 6.42 | 9.01 | 12.46 | 14.81 | 16.18 |
| Other genotypes | Organised | Nice | 38 | 9.01 | 4.74 | 5.19 | 6.23 | 8.73 | 12.06 | 14.40 | 15.72 |
| Other genotypes | Organised | Nice | 39 | 8.76 | 4.63 | 5.05 | 6.06 | 8.48 | 11.70 | 14.02 | 15.29 |
| Other genotypes | Organised | Nice | 40 | 8.50 | 4.47 | 4.91 | 5.90 | 8.24 | 11.40 | 13.68 | 14.89 |
| Other genotypes | Organised | Nice | 41 | 8.28 | 4.37 | 4.74 | 5.73 | 8.03 | 11.10 | 13.36 | 14.53 |
| Other genotypes | Organised | Nice | 42 | 8.10 | 4.25 | 4.63 | 5.58 | 7.84 | 10.88 | 13.08 | 14.20 |
| Other genotypes | Organised | Nice | 43 | 7.95 | 4.18 | 4.57 | 5.48 | 7.69 | 10.68 | 12.91 | 13.86 |
| Other genotypes | Organised | Nice | 44 | 7.84 | 4.11 | 4.51 | 5.39 | 7.58 | 10.53 | 12.72 | 13.69 |
| Other genotypes | Organised | Nice | 45 | 7.74 | 4.08 | 4.42 | 5.33 | 7.49 | 10.40 | 12.53 | 13.47 |
| Other genotypes | Organised | Nice | 46 | 7.67 | 4.04 | 4.38 | 5.29 | 7.41 | 10.30 | 12.41 | 13.39 |
| Other genotypes | Organised | Nice | 47 | 7.62 | 4.00 | 4.32 | 5.25 | 7.36 | 10.20 | 12.30 | 13.27 |
| Other genotypes | Organised | Nice | 48 | 7.59 | 3.98 | 4.31 | 5.21 | 7.35 | 10.19 | 12.25 | 13.28 |
| Other genotypes | Organised | Nice | 49 | 7.57 | 3.97 | 4.34 | 5.20 | 7.31 | 10.18 | 12.31 | 13.36 |
| Other genotypes | Organised | Nice | 50 | 7.55 | 3.99 | 4.33 | 5.18 | 7.29 | 10.15 | 12.25 | 13.12 |
| Other genotypes | Organised | Nice | 51 | 7.53 | 3.95 | 4.30 | 5.17 | 7.26 | 10.12 | 12.27 | 13.13 |
| Other genotypes | Organised | Nice | 52 | 7.48 | 3.92 | 4.27 | 5.14 | 7.21 | 10.03 | 12.15 | 13.07 |
| Other genotypes | Organised | Nice | 53 | 7.41 | 3.89 | 4.23 | 5.10 | 7.16 | 9.95 | 12.03 | 12.87 |
| Other genotypes | Organised | Nice | 54 | 7.30 | 3.80 | 4.15 | 5.02 | 7.05 | 9.83 | 11.79 | 12.73 |

**Table S9 Posterior expected HR HPV cervical infection prevalence (in %) in major French cities, stratified by type of test, city, and age. Table counterpart to Figure 3A *(continued)***

| | | | | Posterior distribution | | | | | | | |
|---|---|---|---|---|---|---|---|---|---|---|---|
| | | | | | Percentiles | | | | | | |
| Virus | Test | City | Age | Average | 0.01 | 0.025 | 0.1 | 0.5 | 0.9 | 0.975 | 0.99 |
| Other genotypes | Organised | Nice | 55 | 7.21 | 3.76 | 4.10 | 4.95 | 6.96 | 9.71 | 11.62 | 12.64 |
| Other genotypes | Organised | Nice | 56 | 7.12 | 3.72 | 4.07 | 4.89 | 6.89 | 9.62 | 11.46 | 12.51 |
| Other genotypes | Organised | Nice | 57 | 7.03 | 3.67 | 4.01 | 4.82 | 6.79 | 9.49 | 11.31 | 12.42 |
| Other genotypes | Organised | Nice | 58 | 6.95 | 3.60 | 3.99 | 4.75 | 6.71 | 9.40 | 11.14 | 12.23 |
| Other genotypes | Organised | Nice | 59 | 6.89 | 3.53 | 3.96 | 4.72 | 6.65 | 9.36 | 11.08 | 12.21 |
| Other genotypes | Organised | Nice | 60 | 6.87 | 3.52 | 3.95 | 4.70 | 6.62 | 9.30 | 11.03 | 12.22 |
| Other genotypes | Organised | Nice | 61 | 6.88 | 3.56 | 3.94 | 4.71 | 6.63 | 9.31 | 11.06 | 12.19 |
| Other genotypes | Organised | Nice | 62 | 6.92 | 3.58 | 3.96 | 4.74 | 6.70 | 9.36 | 11.12 | 12.29 |
| Other genotypes | Organised | Nice | 63 | 7.01 | 3.63 | 4.02 | 4.78 | 6.79 | 9.50 | 11.20 | 12.59 |
| Other genotypes | Organised | Nice | 64 | 7.15 | 3.68 | 4.04 | 4.87 | 6.91 | 9.69 | 11.45 | 12.89 |
| Other genotypes | Organised | Nice | 65 | 7.32 | 3.74 | 4.12 | 4.98 | 7.07 | 9.98 | 11.82 | 13.18 |
| Other genotypes | Organised | Nice | 66 | 7.53 | 3.83 | 4.20 | 5.11 | 7.26 | 10.35 | 12.27 | 13.65 |
| Other genotypes | Organised | Marseille | 30 | 14.84 | 10.58 | 11.17 | 12.20 | 14.72 | 17.67 | 19.42 | 20.45 |
| Other genotypes | Organised | Marseille | 31 | 14.14 | 10.16 | 10.68 | 11.65 | 14.01 | 16.82 | 18.52 | 19.44 |
| Other genotypes | Organised | Marseille | 32 | 13.41 | 9.67 | 10.13 | 11.03 | 13.27 | 15.97 | 17.55 | 18.36 |
| Other genotypes | Organised | Marseille | 33 | 12.67 | 9.10 | 9.54 | 10.40 | 12.52 | 15.10 | 16.58 | 17.33 |
| Other genotypes | Organised | Marseille | 34 | 12.04 | 8.58 | 9.08 | 9.89 | 11.91 | 14.33 | 15.81 | 16.50 |
| Other genotypes | Organised | Marseille | 35 | 11.55 | 8.21 | 8.67 | 9.47 | 11.43 | 13.76 | 15.19 | 15.83 |
| Other genotypes | Organised | Marseille | 36 | 11.11 | 7.90 | 8.32 | 9.12 | 11.00 | 13.31 | 14.65 | 15.23 |
| Other genotypes | Organised | Marseille | 37 | 10.76 | 7.64 | 8.03 | 8.81 | 10.65 | 12.88 | 14.23 | 14.75 |
| Other genotypes | Organised | Marseille | 38 | 10.45 | 7.40 | 7.79 | 8.54 | 10.33 | 12.48 | 13.84 | 14.36 |
| Other genotypes | Organised | Marseille | 39 | 10.15 | 7.22 | 7.56 | 8.30 | 10.06 | 12.16 | 13.42 | 13.93 |
| Other genotypes | Organised | Marseille | 40 | 9.86 | 6.99 | 7.38 | 8.06 | 9.77 | 11.80 | 13.02 | 13.58 |
| Other genotypes | Organised | Marseille | 41 | 9.60 | 6.80 | 7.16 | 7.83 | 9.52 | 11.51 | 12.67 | 13.24 |
| Other genotypes | Organised | Marseille | 42 | 9.40 | 6.62 | 6.96 | 7.66 | 9.30 | 11.28 | 12.43 | 13.05 |
| Other genotypes | Organised | Marseille | 43 | 9.23 | 6.48 | 6.81 | 7.52 | 9.14 | 11.07 | 12.25 | 12.78 |
| Other genotypes | Organised | Marseille | 44 | 9.10 | 6.39 | 6.75 | 7.41 | 9.00 | 10.94 | 12.05 | 12.57 |
| Other genotypes | Organised | Marseille | 45 | 8.99 | 6.31 | 6.65 | 7.31 | 8.89 | 10.83 | 11.90 | 12.50 |
| Other genotypes | Organised | Marseille | 46 | 8.90 | 6.24 | 6.57 | 7.25 | 8.81 | 10.75 | 11.85 | 12.36 |

**Table S9 Posterior expected HR HPV cervical infection prevalence (in %) in major French cities, stratified by type of test, city, and age. Table counterpart to Figure 3A** ***(continued)***

| | | | | Posterior distribution | | | | | | | |
|---|---|---|---|---|---|---|---|---|---|---|---|
| | | | | | Percentiles | | | | | | |
| Virus | Test | City | Age | Average | 0.01 | 0.025 | 0.1 | 0.5 | 0.9 | 0.975 | 0.99 |
| Other genotypes | Organised | Marseille | 47 | 8.85 | 6.20 | 6.53 | 7.21 | 8.77 | 10.68 | 11.80 | 12.28 |
| Other genotypes | Organised | Marseille | 48 | 8.82 | 6.20 | 6.50 | 7.18 | 8.73 | 10.65 | 11.75 | 12.26 |
| Other genotypes | Organised | Marseille | 49 | 8.80 | 6.18 | 6.46 | 7.17 | 8.72 | 10.62 | 11.67 | 12.19 |
| Other genotypes | Organised | Marseille | 50 | 8.78 | 6.16 | 6.45 | 7.14 | 8.71 | 10.60 | 11.71 | 12.19 |
| Other genotypes | Organised | Marseille | 51 | 8.75 | 6.12 | 6.43 | 7.09 | 8.67 | 10.57 | 11.69 | 12.16 |
| Other genotypes | Organised | Marseille | 52 | 8.68 | 6.07 | 6.40 | 7.06 | 8.62 | 10.51 | 11.64 | 12.13 |
| Other genotypes | Organised | Marseille | 53 | 8.61 | 6.06 | 6.36 | 6.98 | 8.53 | 10.43 | 11.54 | 12.02 |
| Other genotypes | Organised | Marseille | 54 | 8.48 | 5.99 | 6.28 | 6.88 | 8.40 | 10.28 | 11.34 | 11.87 |
| Other genotypes | Organised | Marseille | 55 | 8.37 | 5.93 | 6.17 | 6.78 | 8.29 | 10.15 | 11.19 | 11.75 |
| Other genotypes | Organised | Marseille | 56 | 8.28 | 5.86 | 6.11 | 6.70 | 8.19 | 10.01 | 11.10 | 11.57 |
| Other genotypes | Organised | Marseille | 57 | 8.16 | 5.81 | 6.03 | 6.62 | 8.07 | 9.87 | 10.95 | 11.41 |
| Other genotypes | Organised | Marseille | 58 | 8.07 | 5.73 | 5.95 | 6.55 | 7.98 | 9.75 | 10.81 | 11.30 |
| Other genotypes | Organised | Marseille | 59 | 8.01 | 5.67 | 5.88 | 6.49 | 7.92 | 9.65 | 10.73 | 11.22 |
| Other genotypes | Organised | Marseille | 60 | 7.98 | 5.65 | 5.90 | 6.47 | 7.88 | 9.60 | 10.71 | 11.19 |
| Other genotypes | Organised | Marseille | 61 | 7.99 | 5.62 | 5.91 | 6.47 | 7.88 | 9.61 | 10.73 | 11.22 |
| Other genotypes | Organised | Marseille | 62 | 8.04 | 5.61 | 5.92 | 6.51 | 7.94 | 9.70 | 10.80 | 11.33 |
| Other genotypes | Organised | Marseille | 63 | 8.15 | 5.66 | 5.98 | 6.58 | 8.04 | 9.84 | 10.94 | 11.56 |
| Other genotypes | Organised | Marseille | 64 | 8.30 | 5.70 | 6.08 | 6.68 | 8.18 | 10.04 | 11.24 | 11.84 |
| Other genotypes | Organised | Marseille | 65 | 8.50 | 5.79 | 6.15 | 6.81 | 8.40 | 10.36 | 11.58 | 12.21 |
| Other genotypes | Organised | Marseille | 66 | 8.74 | 5.84 | 6.25 | 6.92 | 8.63 | 10.71 | 12.01 | 12.67 |
| Other genotypes | Organised | Toulouse | 30 | 12.80 | 7.80 | 8.35 | 9.55 | 12.57 | 16.27 | 18.42 | 19.55 |
| Other genotypes | Organised | Toulouse | 31 | 12.19 | 7.51 | 7.98 | 9.06 | 11.96 | 15.47 | 17.59 | 18.68 |
| Other genotypes | Organised | Toulouse | 32 | 11.54 | 7.07 | 7.55 | 8.57 | 11.32 | 14.70 | 16.72 | 17.91 |
| Other genotypes | Organised | Toulouse | 33 | 10.89 | 6.70 | 7.11 | 8.08 | 10.69 | 13.90 | 15.68 | 16.98 |
| Other genotypes | Organised | Toulouse | 34 | 10.35 | 6.35 | 6.73 | 7.68 | 10.15 | 13.22 | 14.93 | 16.14 |
| Other genotypes | Organised | Toulouse | 35 | 9.91 | 6.10 | 6.39 | 7.37 | 9.72 | 12.65 | 14.33 | 15.51 |
| Other genotypes | Organised | Toulouse | 36 | 9.53 | 5.88 | 6.14 | 7.08 | 9.34 | 12.18 | 13.84 | 14.82 |
| Other genotypes | Organised | Toulouse | 37 | 9.23 | 5.67 | 5.97 | 6.84 | 9.05 | 11.80 | 13.34 | 14.34 |
| Other genotypes | Organised | Toulouse | 38 | 8.95 | 5.51 | 5.80 | 6.64 | 8.77 | 11.45 | 12.93 | 13.99 |

**Table S9** **Posterior expected HR HPV cervical infection prevalence (in %) in major French cities, stratified by type of test, city, and age. Table counterpart to Figure 3A** ***(continued)***

| | | | | Posterior distribution | | | | | | | |
|---|---|---|---|---|---|---|---|---|---|---|---|
| | | | | | Percentiles | | | | | | |
| Virus | Test | City | Age | Average | 0.01 | 0.025 | 0.1 | 0.5 | 0.9 | 0.975 | 0.99 |
| Other genotypes | Organised | Toulouse | 39 | 8.70 | 5.31 | 5.60 | 6.45 | 8.54 | 11.12 | 12.60 | 13.52 |
| Other genotypes | Organised | Toulouse | 40 | 8.44 | 5.14 | 5.40 | 6.26 | 8.28 | 10.76 | 12.24 | 13.16 |
| Other genotypes | Organised | Toulouse | 41 | 8.22 | 5.01 | 5.26 | 6.10 | 8.05 | 10.47 | 11.97 | 12.84 |
| Other genotypes | Organised | Toulouse | 42 | 8.04 | 4.85 | 5.16 | 5.97 | 7.88 | 10.25 | 11.71 | 12.68 |
| Other genotypes | Organised | Toulouse | 43 | 7.90 | 4.77 | 5.08 | 5.88 | 7.75 | 10.10 | 11.55 | 12.47 |
| Other genotypes | Organised | Toulouse | 44 | 7.78 | 4.67 | 5.01 | 5.82 | 7.64 | 9.94 | 11.41 | 12.31 |
| Other genotypes | Organised | Toulouse | 45 | 7.68 | 4.63 | 4.94 | 5.72 | 7.55 | 9.81 | 11.32 | 12.12 |
| Other genotypes | Organised | Toulouse | 46 | 7.61 | 4.59 | 4.89 | 5.66 | 7.48 | 9.74 | 11.28 | 12.06 |
| Other genotypes | Organised | Toulouse | 47 | 7.56 | 4.55 | 4.87 | 5.61 | 7.41 | 9.69 | 11.21 | 11.97 |
| Other genotypes | Organised | Toulouse | 48 | 7.54 | 4.54 | 4.85 | 5.59 | 7.39 | 9.67 | 11.17 | 11.89 |
| Other genotypes | Organised | Toulouse | 49 | 7.52 | 4.54 | 4.84 | 5.57 | 7.37 | 9.67 | 11.08 | 11.80 |
| Other genotypes | Organised | Toulouse | 50 | 7.50 | 4.53 | 4.83 | 5.55 | 7.38 | 9.66 | 11.06 | 11.80 |
| Other genotypes | Organised | Toulouse | 51 | 7.48 | 4.51 | 4.81 | 5.53 | 7.34 | 9.61 | 11.03 | 11.75 |
| Other genotypes | Organised | Toulouse | 52 | 7.42 | 4.46 | 4.75 | 5.49 | 7.28 | 9.53 | 10.95 | 11.69 |
| Other genotypes | Organised | Toulouse | 53 | 7.35 | 4.42 | 4.71 | 5.44 | 7.22 | 9.45 | 10.83 | 11.61 |
| Other genotypes | Organised | Toulouse | 54 | 7.25 | 4.36 | 4.65 | 5.36 | 7.09 | 9.34 | 10.69 | 11.46 |
| Other genotypes | Organised | Toulouse | 55 | 7.15 | 4.27 | 4.58 | 5.29 | 7.01 | 9.19 | 10.56 | 11.38 |
| Other genotypes | Organised | Toulouse | 56 | 7.07 | 4.22 | 4.53 | 5.22 | 6.93 | 9.07 | 10.49 | 11.24 |
| Other genotypes | Organised | Toulouse | 57 | 6.97 | 4.17 | 4.46 | 5.14 | 6.83 | 8.93 | 10.33 | 11.13 |
| Other genotypes | Organised | Toulouse | 58 | 6.90 | 4.10 | 4.42 | 5.09 | 6.75 | 8.84 | 10.18 | 10.97 |
| Other genotypes | Organised | Toulouse | 59 | 6.84 | 4.08 | 4.38 | 5.03 | 6.70 | 8.78 | 10.08 | 10.86 |
| Other genotypes | Organised | Toulouse | 60 | 6.82 | 4.06 | 4.38 | 5.02 | 6.67 | 8.73 | 10.04 | 10.83 |
| Other genotypes | Organised | Toulouse | 61 | 6.82 | 4.08 | 4.37 | 5.02 | 6.67 | 8.77 | 10.10 | 10.83 |
| Other genotypes | Organised | Toulouse | 62 | 6.87 | 4.08 | 4.39 | 5.04 | 6.71 | 8.88 | 10.22 | 10.93 |
| Other genotypes | Organised | Toulouse | 63 | 6.96 | 4.13 | 4.45 | 5.10 | 6.79 | 9.02 | 10.39 | 11.09 |
| Other genotypes | Organised | Toulouse | 64 | 7.10 | 4.16 | 4.50 | 5.18 | 6.91 | 9.20 | 10.55 | 11.31 |
| Other genotypes | Organised | Toulouse | 65 | 7.27 | 4.22 | 4.57 | 5.28 | 7.09 | 9.41 | 10.83 | 11.63 |
| Other genotypes | Organised | Toulouse | 66 | 7.47 | 4.29 | 4.63 | 5.43 | 7.32 | 9.74 | 11.19 | 12.01 |
| Other genotypes | Organised | Bordeaux | 30 | 13.60 | 9.57 | 10.13 | 11.16 | 13.45 | 16.18 | 17.85 | 18.72 |

**Table S9** **Posterior expected HR HPV cervical infection prevalence (in %) in major French cities, stratified by type of test, city, and age. Table counterpart to Figure 3A *(continued)***

| | | | | Posterior distribution | | | | | | | |
|---|---|---|---|---|---|---|---|---|---|---|---|
| | | | | | Percentiles | | | | | | |
| Virus | Test | City | Age | Average | 0.01 | 0.025 | 0.1 | 0.5 | 0.9 | 0.975 | 0.99 |
| Other genotypes | Organised | Bordeaux | 31 | 12.95 | 9.18 | 9.64 | 10.67 | 12.82 | 15.41 | 16.98 | 17.73 |
| Other genotypes | Organised | Bordeaux | 32 | 12.26 | 8.78 | 9.18 | 10.12 | 12.16 | 14.59 | 16.01 | 16.79 |
| Other genotypes | Organised | Bordeaux | 33 | 11.58 | 8.28 | 8.65 | 9.54 | 11.47 | 13.79 | 15.16 | 15.90 |
| Other genotypes | Organised | Bordeaux | 34 | 11.00 | 7.84 | 8.23 | 9.05 | 10.89 | 13.12 | 14.41 | 15.12 |
| Other genotypes | Organised | Bordeaux | 35 | 10.54 | 7.47 | 7.88 | 8.68 | 10.42 | 12.60 | 13.85 | 14.53 |
| Other genotypes | Organised | Bordeaux | 36 | 10.14 | 7.14 | 7.56 | 8.35 | 10.03 | 12.11 | 13.27 | 13.97 |
| Other genotypes | Organised | Bordeaux | 37 | 9.82 | 6.89 | 7.32 | 8.09 | 9.71 | 11.74 | 12.84 | 13.58 |
| Other genotypes | Organised | Bordeaux | 38 | 9.52 | 6.72 | 7.11 | 7.83 | 9.42 | 11.39 | 12.50 | 13.17 |
| Other genotypes | Organised | Bordeaux | 39 | 9.25 | 6.49 | 6.92 | 7.60 | 9.14 | 11.05 | 12.15 | 12.77 |
| Other genotypes | Organised | Bordeaux | 40 | 8.98 | 6.31 | 6.73 | 7.37 | 8.89 | 10.75 | 11.83 | 12.45 |
| Other genotypes | Organised | Bordeaux | 41 | 8.74 | 6.15 | 6.57 | 7.17 | 8.64 | 10.48 | 11.55 | 12.11 |
| Other genotypes | Organised | Bordeaux | 42 | 8.56 | 6.01 | 6.41 | 7.01 | 8.46 | 10.26 | 11.36 | 11.76 |
| Other genotypes | Organised | Bordeaux | 43 | 8.41 | 5.89 | 6.27 | 6.89 | 8.31 | 10.06 | 11.15 | 11.66 |
| Other genotypes | Organised | Bordeaux | 44 | 8.28 | 5.79 | 6.18 | 6.78 | 8.19 | 9.92 | 11.00 | 11.51 |
| Other genotypes | Organised | Bordeaux | 45 | 8.18 | 5.76 | 6.09 | 6.70 | 8.07 | 9.80 | 10.87 | 11.34 |
| Other genotypes | Organised | Bordeaux | 46 | 8.10 | 5.69 | 6.05 | 6.63 | 8.00 | 9.72 | 10.72 | 11.16 |
| Other genotypes | Organised | Bordeaux | 47 | 8.05 | 5.66 | 5.98 | 6.60 | 7.94 | 9.66 | 10.64 | 11.12 |
| Other genotypes | Organised | Bordeaux | 48 | 8.02 | 5.63 | 5.95 | 6.56 | 7.90 | 9.64 | 10.61 | 11.12 |
| Other genotypes | Organised | Bordeaux | 49 | 8.00 | 5.58 | 5.96 | 6.56 | 7.89 | 9.60 | 10.57 | 11.09 |
| Other genotypes | Organised | Bordeaux | 50 | 7.99 | 5.58 | 5.94 | 6.53 | 7.88 | 9.56 | 10.53 | 10.99 |
| Other genotypes | Organised | Bordeaux | 51 | 7.96 | 5.55 | 5.91 | 6.51 | 7.88 | 9.52 | 10.53 | 11.01 |
| Other genotypes | Organised | Bordeaux | 52 | 7.90 | 5.53 | 5.86 | 6.46 | 7.82 | 9.48 | 10.47 | 10.98 |
| Other genotypes | Organised | Bordeaux | 53 | 7.83 | 5.48 | 5.79 | 6.40 | 7.76 | 9.40 | 10.32 | 10.89 |
| Other genotypes | Organised | Bordeaux | 54 | 7.72 | 5.39 | 5.69 | 6.32 | 7.66 | 9.26 | 10.16 | 10.81 |
| Other genotypes | Organised | Bordeaux | 55 | 7.62 | 5.36 | 5.61 | 6.24 | 7.54 | 9.16 | 10.07 | 10.71 |
| Other genotypes | Organised | Bordeaux | 56 | 7.53 | 5.28 | 5.53 | 6.16 | 7.46 | 9.06 | 10.00 | 10.51 |
| Other genotypes | Organised | Bordeaux | 57 | 7.42 | 5.24 | 5.47 | 6.08 | 7.35 | 8.92 | 9.84 | 10.35 |
| Other genotypes | Organised | Bordeaux | 58 | 7.34 | 5.20 | 5.40 | 6.01 | 7.26 | 8.83 | 9.67 | 10.30 |
| Other genotypes | Organised | Bordeaux | 59 | 7.28 | 5.15 | 5.34 | 5.97 | 7.20 | 8.75 | 9.60 | 10.13 |

**Table S9 Posterior expected HR HPV cervical infection prevalence (in %) in major French cities, stratified by type of test, city, and age. Table counterpart to Figure 3A *(continued)***

| | | | | Posterior distribution | | | | | | | |
|---|---|---|---|---|---|---|---|---|---|---|---|
| | | | | | Percentiles | | | | | | |
| Virus | Test | City | Age | Average | 0.01 | 0.025 | 0.1 | 0.5 | 0.9 | 0.975 | 0.99 |
| Other genotypes | Organised | Bordeaux | 60 | 7.26 | 5.13 | 5.35 | 5.95 | 7.17 | 8.73 | 9.54 | 10.13 |
| Other genotypes | Organised | Bordeaux | 61 | 7.26 | 5.13 | 5.34 | 5.92 | 7.19 | 8.73 | 9.55 | 10.14 |
| Other genotypes | Organised | Bordeaux | 62 | 7.31 | 5.15 | 5.37 | 5.94 | 7.25 | 8.78 | 9.70 | 10.19 |
| Other genotypes | Organised | Bordeaux | 63 | 7.41 | 5.20 | 5.41 | 6.02 | 7.34 | 8.90 | 9.88 | 10.36 |
| Other genotypes | Organised | Bordeaux | 64 | 7.55 | 5.26 | 5.49 | 6.11 | 7.47 | 9.07 | 10.10 | 10.64 |
| Other genotypes | Organised | Bordeaux | 65 | 7.74 | 5.36 | 5.56 | 6.23 | 7.65 | 9.34 | 10.41 | 11.03 |
| Other genotypes | Organised | Bordeaux | 66 | 7.95 | 5.41 | 5.64 | 6.33 | 7.84 | 9.69 | 10.88 | 11.46 |
| Other genotypes | Organised | Montpellier | 30 | 14.60 | 9.46 | 10.05 | 11.35 | 14.41 | 18.13 | 20.60 | 21.91 |
| Other genotypes | Organised | Montpellier | 31 | 13.91 | 9.11 | 9.59 | 10.84 | 13.74 | 17.23 | 19.60 | 20.85 |
| Other genotypes | Organised | Montpellier | 32 | 13.19 | 8.62 | 9.06 | 10.25 | 13.04 | 16.32 | 18.60 | 19.74 |
| Other genotypes | Organised | Montpellier | 33 | 12.46 | 8.13 | 8.59 | 9.69 | 12.32 | 15.49 | 17.54 | 18.78 |
| Other genotypes | Organised | Montpellier | 34 | 11.85 | 7.71 | 8.15 | 9.20 | 11.71 | 14.70 | 16.73 | 18.04 |
| Other genotypes | Organised | Montpellier | 35 | 11.36 | 7.36 | 7.79 | 8.81 | 11.23 | 14.12 | 16.04 | 17.29 |
| Other genotypes | Organised | Montpellier | 36 | 10.93 | 7.10 | 7.48 | 8.45 | 10.79 | 13.57 | 15.47 | 16.57 |
| Other genotypes | Organised | Montpellier | 37 | 10.58 | 6.85 | 7.25 | 8.17 | 10.44 | 13.16 | 14.96 | 16.01 |
| Other genotypes | Organised | Montpellier | 38 | 10.27 | 6.59 | 7.04 | 7.92 | 10.14 | 12.75 | 14.51 | 15.56 |
| Other genotypes | Organised | Montpellier | 39 | 9.99 | 6.39 | 6.83 | 7.69 | 9.84 | 12.43 | 14.09 | 15.22 |
| Other genotypes | Organised | Montpellier | 40 | 9.69 | 6.16 | 6.62 | 7.47 | 9.56 | 12.06 | 13.72 | 14.79 |
| Other genotypes | Organised | Montpellier | 41 | 9.44 | 6.00 | 6.47 | 7.26 | 9.31 | 11.75 | 13.37 | 14.30 |
| Other genotypes | Organised | Montpellier | 42 | 9.24 | 5.86 | 6.32 | 7.10 | 9.12 | 11.50 | 13.08 | 14.01 |
| Other genotypes | Organised | Montpellier | 43 | 9.08 | 5.74 | 6.20 | 6.97 | 8.95 | 11.30 | 12.91 | 13.77 |
| Other genotypes | Organised | Montpellier | 44 | 8.94 | 5.65 | 6.11 | 6.85 | 8.83 | 11.15 | 12.69 | 13.65 |
| Other genotypes | Organised | Montpellier | 45 | 8.83 | 5.59 | 6.01 | 6.78 | 8.72 | 11.01 | 12.61 | 13.57 |
| Other genotypes | Organised | Montpellier | 46 | 8.75 | 5.57 | 5.94 | 6.72 | 8.64 | 10.90 | 12.51 | 13.38 |
| Other genotypes | Organised | Montpellier | 47 | 8.70 | 5.54 | 5.91 | 6.68 | 8.58 | 10.82 | 12.43 | 13.23 |
| Other genotypes | Organised | Montpellier | 48 | 8.67 | 5.47 | 5.90 | 6.65 | 8.55 | 10.79 | 12.39 | 13.34 |
| Other genotypes | Organised | Montpellier | 49 | 8.65 | 5.46 | 5.87 | 6.63 | 8.53 | 10.77 | 12.38 | 13.22 |
| Other genotypes | Organised | Montpellier | 50 | 8.63 | 5.43 | 5.86 | 6.60 | 8.51 | 10.73 | 12.39 | 13.18 |
| Other genotypes | Organised | Montpellier | 51 | 8.60 | 5.41 | 5.83 | 6.58 | 8.48 | 10.73 | 12.32 | 13.06 |

**Table S9** **Posterior expected HR HPV cervical infection prevalence (in %) in major French cities, stratified by type of test, city, and age. Table counterpart to Figure 3A** ***(continued)***

| | | | | Posterior distribution | | | | | | | |
|---|---|---|---|---|---|---|---|---|---|---|---|
| | | | | | Percentiles | | | | | | |
| Virus | Test | City | Age | Average | 0.01 | 0.025 | 0.1 | 0.5 | 0.9 | 0.975 | 0.99 |
| Other genotypes | Organised | Montpellier | 52 | 8.54 | 5.36 | 5.79 | 6.52 | 8.42 | 10.64 | 12.21 | 13.10 |
| Other genotypes | Organised | Montpellier | 53 | 8.46 | 5.37 | 5.77 | 6.45 | 8.35 | 10.55 | 12.06 | 12.87 |
| Other genotypes | Organised | Montpellier | 54 | 8.34 | 5.35 | 5.69 | 6.36 | 8.22 | 10.42 | 11.95 | 12.78 |
| Other genotypes | Organised | Montpellier | 55 | 8.23 | 5.25 | 5.58 | 6.27 | 8.11 | 10.26 | 11.79 | 12.59 |
| Other genotypes | Organised | Montpellier | 56 | 8.14 | 5.19 | 5.53 | 6.20 | 8.02 | 10.13 | 11.61 | 12.34 |
| Other genotypes | Organised | Montpellier | 57 | 8.03 | 5.11 | 5.43 | 6.14 | 7.92 | 10.01 | 11.49 | 12.20 |
| Other genotypes | Organised | Montpellier | 58 | 7.94 | 5.07 | 5.37 | 6.07 | 7.83 | 9.91 | 11.36 | 12.13 |
| Other genotypes | Organised | Montpellier | 59 | 7.88 | 5.03 | 5.33 | 6.02 | 7.77 | 9.83 | 11.25 | 12.04 |
| Other genotypes | Organised | Montpellier | 60 | 7.85 | 4.97 | 5.28 | 5.98 | 7.74 | 9.83 | 11.24 | 11.92 |
| Other genotypes | Organised | Montpellier | 61 | 7.86 | 4.97 | 5.28 | 5.99 | 7.74 | 9.84 | 11.19 | 11.88 |
| Other genotypes | Organised | Montpellier | 62 | 7.91 | 4.99 | 5.30 | 6.01 | 7.78 | 9.92 | 11.24 | 11.93 |
| Other genotypes | Organised | Montpellier | 63 | 8.01 | 5.05 | 5.35 | 6.10 | 7.88 | 10.06 | 11.37 | 12.18 |
| Other genotypes | Organised | Montpellier | 64 | 8.16 | 5.17 | 5.42 | 6.21 | 8.03 | 10.28 | 11.64 | 12.49 |
| Other genotypes | Organised | Montpellier | 65 | 8.36 | 5.23 | 5.52 | 6.32 | 8.22 | 10.56 | 12.02 | 12.99 |
| Other genotypes | Organised | Montpellier | 66 | 8.59 | 5.26 | 5.64 | 6.47 | 8.46 | 10.92 | 12.42 | 13.52 |
| Other genotypes | Organised | Rennes | 30 | 12.75 | 9.04 | 9.47 | 10.57 | 12.66 | 15.11 | 16.70 | 17.41 |
| Other genotypes | Organised | Rennes | 31 | 12.14 | 8.62 | 9.05 | 10.07 | 12.03 | 14.36 | 15.84 | 16.44 |
| Other genotypes | Organised | Rennes | 32 | 11.49 | 8.17 | 8.61 | 9.52 | 11.38 | 13.57 | 14.88 | 15.54 |
| Other genotypes | Organised | Rennes | 33 | 10.84 | 7.73 | 8.12 | 9.01 | 10.75 | 12.84 | 14.06 | 14.68 |
| Other genotypes | Organised | Rennes | 34 | 10.30 | 7.36 | 7.74 | 8.54 | 10.22 | 12.20 | 13.39 | 13.89 |
| Other genotypes | Organised | Rennes | 35 | 9.86 | 7.04 | 7.35 | 8.16 | 9.80 | 11.67 | 12.77 | 13.36 |
| Other genotypes | Organised | Rennes | 36 | 9.48 | 6.81 | 7.06 | 7.83 | 9.42 | 11.24 | 12.28 | 12.92 |
| Other genotypes | Organised | Rennes | 37 | 9.18 | 6.56 | 6.87 | 7.59 | 9.12 | 10.88 | 11.89 | 12.51 |
| Other genotypes | Organised | Rennes | 38 | 8.90 | 6.31 | 6.62 | 7.38 | 8.82 | 10.53 | 11.57 | 12.16 |
| Other genotypes | Organised | Rennes | 39 | 8.65 | 6.16 | 6.46 | 7.16 | 8.58 | 10.27 | 11.26 | 11.74 |
| Other genotypes | Organised | Rennes | 40 | 8.39 | 6.00 | 6.29 | 6.93 | 8.33 | 9.96 | 10.93 | 11.37 |
| Other genotypes | Organised | Rennes | 41 | 8.17 | 5.84 | 6.11 | 6.73 | 8.12 | 9.69 | 10.67 | 11.10 |
| Other genotypes | Organised | Rennes | 42 | 7.99 | 5.71 | 5.99 | 6.58 | 7.94 | 9.48 | 10.44 | 10.88 |
| Other genotypes | Organised | Rennes | 43 | 7.85 | 5.60 | 5.88 | 6.47 | 7.80 | 9.32 | 10.24 | 10.68 |

**Table S9 Posterior expected HR HPV cervical infection prevalence (in %) in major French cities, stratified by type of test, city, and age. Table counterpart to Figure 3A *(continued)***

| | | | | Posterior distribution | | | | | | | |
|---|---|---|---|---|---|---|---|---|---|---|---|
| | | | | | Percentiles | | | | | | |
| Virus | Test | City | Age | Average | 0.01 | 0.025 | 0.1 | 0.5 | 0.9 | 0.975 | 0.99 |
| Other genotypes | Organised | Rennes | 44 | 7.73 | 5.51 | 5.78 | 6.36 | 7.68 | 9.20 | 10.07 | 10.49 |
| Other genotypes | Organised | Rennes | 45 | 7.64 | 5.44 | 5.70 | 6.29 | 7.58 | 9.08 | 9.97 | 10.31 |
| Other genotypes | Organised | Rennes | 46 | 7.57 | 5.40 | 5.66 | 6.22 | 7.51 | 9.01 | 9.87 | 10.27 |
| Other genotypes | Organised | Rennes | 47 | 7.52 | 5.37 | 5.62 | 6.18 | 7.45 | 8.93 | 9.80 | 10.20 |
| Other genotypes | Organised | Rennes | 48 | 7.49 | 5.36 | 5.60 | 6.17 | 7.43 | 8.90 | 9.76 | 10.18 |
| Other genotypes | Organised | Rennes | 49 | 7.47 | 5.34 | 5.61 | 6.14 | 7.41 | 8.88 | 9.74 | 10.17 |
| Other genotypes | Organised | Rennes | 50 | 7.46 | 5.33 | 5.60 | 6.12 | 7.41 | 8.85 | 9.70 | 10.14 |
| Other genotypes | Organised | Rennes | 51 | 7.43 | 5.27 | 5.60 | 6.11 | 7.38 | 8.83 | 9.67 | 10.08 |
| Other genotypes | Organised | Rennes | 52 | 7.38 | 5.25 | 5.55 | 6.04 | 7.34 | 8.78 | 9.60 | 9.99 |
| Other genotypes | Organised | Rennes | 53 | 7.31 | 5.19 | 5.49 | 6.00 | 7.26 | 8.70 | 9.53 | 9.93 |
| Other genotypes | Organised | Rennes | 54 | 7.20 | 5.14 | 5.40 | 5.90 | 7.14 | 8.57 | 9.40 | 9.81 |
| Other genotypes | Organised | Rennes | 55 | 7.11 | 5.07 | 5.34 | 5.82 | 7.06 | 8.47 | 9.27 | 9.68 |
| Other genotypes | Organised | Rennes | 56 | 7.03 | 5.02 | 5.27 | 5.75 | 6.96 | 8.36 | 9.19 | 9.57 |
| Other genotypes | Organised | Rennes | 57 | 6.93 | 4.95 | 5.19 | 5.67 | 6.86 | 8.26 | 9.08 | 9.43 |
| Other genotypes | Organised | Rennes | 58 | 6.85 | 4.89 | 5.13 | 5.62 | 6.79 | 8.17 | 8.97 | 9.31 |
| Other genotypes | Organised | Rennes | 59 | 6.80 | 4.83 | 5.10 | 5.57 | 6.74 | 8.10 | 8.90 | 9.23 |
| Other genotypes | Organised | Rennes | 60 | 6.78 | 4.82 | 5.06 | 5.55 | 6.71 | 8.07 | 8.85 | 9.25 |
| Other genotypes | Organised | Rennes | 61 | 6.78 | 4.82 | 5.07 | 5.56 | 6.72 | 8.08 | 8.84 | 9.29 |
| Other genotypes | Organised | Rennes | 62 | 6.83 | 4.82 | 5.07 | 5.60 | 6.77 | 8.13 | 8.91 | 9.36 |
| Other genotypes | Organised | Rennes | 63 | 6.92 | 4.86 | 5.11 | 5.65 | 6.85 | 8.26 | 9.07 | 9.56 |
| Other genotypes | Organised | Rennes | 64 | 7.05 | 4.94 | 5.17 | 5.75 | 6.97 | 8.44 | 9.28 | 9.83 |
| Other genotypes | Organised | Rennes | 65 | 7.22 | 4.96 | 5.25 | 5.85 | 7.14 | 8.71 | 9.57 | 10.08 |
| Other genotypes | Organised | Rennes | 66 | 7.43 | 5.02 | 5.32 | 5.93 | 7.35 | 9.00 | 9.99 | 10.57 |
| Other genotypes | Organised | Nantes | 30 | 11.81 | 7.26 | 7.76 | 8.85 | 11.59 | 15.07 | 17.09 | 18.07 |
| Other genotypes | Organised | Nantes | 31 | 11.23 | 6.92 | 7.41 | 8.45 | 11.04 | 14.36 | 16.26 | 17.11 |
| Other genotypes | Organised | Nantes | 32 | 10.63 | 6.61 | 7.01 | 7.98 | 10.43 | 13.59 | 15.47 | 16.15 |
| Other genotypes | Organised | Nantes | 33 | 10.03 | 6.21 | 6.61 | 7.53 | 9.82 | 12.82 | 14.64 | 15.21 |
| Other genotypes | Organised | Nantes | 34 | 9.52 | 5.84 | 6.26 | 7.14 | 9.32 | 12.18 | 13.90 | 14.45 |
| Other genotypes | Organised | Nantes | 35 | 9.12 | 5.58 | 6.03 | 6.85 | 8.92 | 11.67 | 13.26 | 13.89 |

**Table S9 Posterior expected HR HPV cervical infection prevalence (in %) in major French cities, stratified by type of test, city, and age. Table counterpart to Figure 3A** ***(continued)***

| | | | | Posterior distribution | | | | | | | |
|---|---|---|---|---|---|---|---|---|---|---|---|
| | | | | | Percentiles | | | | | | |
| Virus | Test | City | Age | Average | 0.01 | 0.025 | 0.1 | 0.5 | 0.9 | 0.975 | 0.99 |
| Other genotypes | Organised | Nantes | 36 | 8.76 | 5.31 | 5.79 | 6.56 | 8.59 | 11.26 | 12.78 | 13.40 |
| Other genotypes | Organised | Nantes | 37 | 8.48 | 5.12 | 5.61 | 6.34 | 8.32 | 10.91 | 12.38 | 13.05 |
| Other genotypes | Organised | Nantes | 38 | 8.23 | 4.95 | 5.44 | 6.15 | 8.08 | 10.58 | 12.01 | 12.71 |
| Other genotypes | Organised | Nantes | 39 | 7.99 | 4.81 | 5.27 | 5.96 | 7.84 | 10.27 | 11.65 | 12.31 |
| Other genotypes | Organised | Nantes | 40 | 7.75 | 4.67 | 5.09 | 5.77 | 7.61 | 9.99 | 11.30 | 11.90 |
| Other genotypes | Organised | Nantes | 41 | 7.54 | 4.53 | 4.94 | 5.61 | 7.40 | 9.74 | 10.97 | 11.74 |
| Other genotypes | Organised | Nantes | 42 | 7.38 | 4.41 | 4.83 | 5.48 | 7.24 | 9.53 | 10.75 | 11.45 |
| Other genotypes | Organised | Nantes | 43 | 7.25 | 4.34 | 4.72 | 5.39 | 7.11 | 9.36 | 10.57 | 11.25 |
| Other genotypes | Organised | Nantes | 44 | 7.14 | 4.28 | 4.64 | 5.29 | 6.99 | 9.24 | 10.41 | 11.15 |
| Other genotypes | Organised | Nantes | 45 | 7.05 | 4.28 | 4.58 | 5.22 | 6.90 | 9.09 | 10.33 | 11.03 |
| Other genotypes | Organised | Nantes | 46 | 6.99 | 4.22 | 4.53 | 5.18 | 6.83 | 9.03 | 10.29 | 10.89 |
| Other genotypes | Organised | Nantes | 47 | 6.94 | 4.16 | 4.49 | 5.13 | 6.79 | 8.94 | 10.19 | 10.83 |
| Other genotypes | Organised | Nantes | 48 | 6.92 | 4.15 | 4.49 | 5.12 | 6.79 | 8.91 | 10.13 | 10.84 |
| Other genotypes | Organised | Nantes | 49 | 6.90 | 4.11 | 4.47 | 5.13 | 6.76 | 8.91 | 10.08 | 10.74 |
| Other genotypes | Organised | Nantes | 50 | 6.88 | 4.08 | 4.47 | 5.11 | 6.73 | 8.90 | 10.07 | 10.70 |
| Other genotypes | Organised | Nantes | 51 | 6.86 | 4.08 | 4.44 | 5.09 | 6.71 | 8.88 | 10.03 | 10.74 |
| Other genotypes | Organised | Nantes | 52 | 6.81 | 4.06 | 4.43 | 5.06 | 6.66 | 8.78 | 9.99 | 10.68 |
| Other genotypes | Organised | Nantes | 53 | 6.75 | 4.03 | 4.39 | 5.01 | 6.60 | 8.71 | 9.92 | 10.58 |
| Other genotypes | Organised | Nantes | 54 | 6.65 | 3.98 | 4.33 | 4.95 | 6.50 | 8.61 | 9.79 | 10.37 |
| Other genotypes | Organised | Nantes | 55 | 6.56 | 3.90 | 4.26 | 4.86 | 6.42 | 8.48 | 9.62 | 10.15 |
| Other genotypes | Organised | Nantes | 56 | 6.48 | 3.84 | 4.21 | 4.80 | 6.33 | 8.39 | 9.53 | 10.05 |
| Other genotypes | Organised | Nantes | 57 | 6.40 | 3.80 | 4.13 | 4.73 | 6.24 | 8.29 | 9.42 | 9.99 |
| Other genotypes | Organised | Nantes | 58 | 6.32 | 3.79 | 4.08 | 4.69 | 6.17 | 8.19 | 9.31 | 9.86 |
| Other genotypes | Organised | Nantes | 59 | 6.27 | 3.73 | 4.06 | 4.66 | 6.12 | 8.11 | 9.21 | 9.73 |
| Other genotypes | Organised | Nantes | 60 | 6.25 | 3.71 | 4.02 | 4.66 | 6.10 | 8.06 | 9.18 | 9.76 |
| Other genotypes | Organised | Nantes | 61 | 6.26 | 3.73 | 4.04 | 4.66 | 6.10 | 8.07 | 9.21 | 9.79 |
| Other genotypes | Organised | Nantes | 62 | 6.30 | 3.74 | 4.06 | 4.70 | 6.15 | 8.13 | 9.28 | 9.88 |
| Other genotypes | Organised | Nantes | 63 | 6.38 | 3.80 | 4.08 | 4.76 | 6.23 | 8.27 | 9.43 | 10.03 |
| Other genotypes | Organised | Nantes | 64 | 6.51 | 3.88 | 4.15 | 4.83 | 6.35 | 8.45 | 9.64 | 10.22 |

**Table S9 Posterior expected HR HPV cervical infection prevalence (in %) in major French cities, stratified by type of test, city, and age. Table counterpart to Figure 3A** ***(continued)***

| | | | | | Posterior distribution | | | | | | |
|---|---|---|---|---|---|---|---|---|---|---|---|
| | | | | | Percentiles | | | | | | |
| Virus | Test | City | Age | Average | 0.01 | 0.025 | 0.1 | 0.5 | 0.9 | 0.975 | 0.99 |
| Other genotypes | Organised | Nantes | 65 | 6.67 | 3.95 | 4.23 | 4.92 | 6.51 | 8.66 | 10.00 | 10.61 |
| Other genotypes | Organised | Nantes | 66 | 6.86 | 4.02 | 4.31 | 5.04 | 6.68 | 8.93 | 10.39 | 11.03 |
| Other genotypes | Organised | Lille | 30 | 12.70 | 8.12 | 8.74 | 9.88 | 12.51 | 15.74 | 17.93 | 18.81 |
| Other genotypes | Organised | Lille | 31 | 12.09 | 7.76 | 8.29 | 9.44 | 11.90 | 14.97 | 16.96 | 17.92 |
| Other genotypes | Organised | Lille | 32 | 11.45 | 7.36 | 7.89 | 8.94 | 11.25 | 14.17 | 16.02 | 17.10 |
| Other genotypes | Organised | Lille | 33 | 10.80 | 6.90 | 7.43 | 8.43 | 10.62 | 13.42 | 15.19 | 16.18 |
| Other genotypes | Organised | Lille | 34 | 10.26 | 6.49 | 7.07 | 8.02 | 10.09 | 12.76 | 14.45 | 15.42 |
| Other genotypes | Organised | Lille | 35 | 9.83 | 6.21 | 6.78 | 7.69 | 9.65 | 12.22 | 13.85 | 14.82 |
| Other genotypes | Organised | Lille | 36 | 9.45 | 5.98 | 6.52 | 7.37 | 9.27 | 11.77 | 13.30 | 14.25 |
| Other genotypes | Organised | Lille | 37 | 9.15 | 5.76 | 6.28 | 7.14 | 8.98 | 11.37 | 12.89 | 13.85 |
| Other genotypes | Organised | Lille | 38 | 8.87 | 5.61 | 6.11 | 6.92 | 8.71 | 11.03 | 12.51 | 13.48 |
| Other genotypes | Organised | Lille | 39 | 8.62 | 5.44 | 5.93 | 6.73 | 8.46 | 10.72 | 12.15 | 13.02 |
| Other genotypes | Organised | Lille | 40 | 8.37 | 5.29 | 5.73 | 6.53 | 8.21 | 10.43 | 11.79 | 12.63 |
| Other genotypes | Organised | Lille | 41 | 8.14 | 5.15 | 5.54 | 6.35 | 8.00 | 10.16 | 11.43 | 12.28 |
| Other genotypes | Organised | Lille | 42 | 7.97 | 5.04 | 5.43 | 6.22 | 7.82 | 9.99 | 11.17 | 11.98 |
| Other genotypes | Organised | Lille | 43 | 7.83 | 4.91 | 5.30 | 6.11 | 7.69 | 9.82 | 11.00 | 11.84 |
| Other genotypes | Organised | Lille | 44 | 7.71 | 4.83 | 5.21 | 6.01 | 7.58 | 9.66 | 10.84 | 11.69 |
| Other genotypes | Organised | Lille | 45 | 7.61 | 4.78 | 5.14 | 5.93 | 7.46 | 9.53 | 10.70 | 11.52 |
| Other genotypes | Organised | Lille | 46 | 7.54 | 4.74 | 5.08 | 5.88 | 7.40 | 9.44 | 10.61 | 11.41 |
| Other genotypes | Organised | Lille | 47 | 7.49 | 4.73 | 5.07 | 5.83 | 7.34 | 9.36 | 10.55 | 11.32 |
| Other genotypes | Organised | Lille | 48 | 7.47 | 4.71 | 5.09 | 5.79 | 7.33 | 9.32 | 10.53 | 11.27 |
| Other genotypes | Organised | Lille | 49 | 7.45 | 4.69 | 5.06 | 5.80 | 7.31 | 9.30 | 10.49 | 11.20 |
| Other genotypes | Organised | Lille | 50 | 7.43 | 4.70 | 5.01 | 5.79 | 7.29 | 9.29 | 10.51 | 11.19 |
| Other genotypes | Organised | Lille | 51 | 7.41 | 4.63 | 5.00 | 5.77 | 7.28 | 9.24 | 10.43 | 11.13 |
| Other genotypes | Organised | Lille | 52 | 7.35 | 4.59 | 4.96 | 5.73 | 7.22 | 9.20 | 10.37 | 11.09 |
| Other genotypes | Organised | Lille | 53 | 7.29 | 4.53 | 4.90 | 5.66 | 7.15 | 9.10 | 10.24 | 10.96 |
| Other genotypes | Organised | Lille | 54 | 7.18 | 4.44 | 4.82 | 5.57 | 7.04 | 9.00 | 10.12 | 10.84 |
| Other genotypes | Organised | Lille | 55 | 7.09 | 4.41 | 4.78 | 5.49 | 6.95 | 8.87 | 9.98 | 10.68 |
| Other genotypes | Organised | Lille | 56 | 7.00 | 4.36 | 4.74 | 5.42 | 6.87 | 8.78 | 9.89 | 10.54 |

**Table S9 Posterior expected HR HPV cervical infection prevalence (in %) in major French cities, stratified by type of test, city, and age. Table counterpart to Figure 3A** ***(continued)***

| | | | | Posterior distribution | | | | | | | |
|---|---|---|---|---|---|---|---|---|---|---|---|
| | | | | | Percentiles | | | | | | |
| Virus | Test | City | Age | Average | 0.01 | 0.025 | 0.1 | 0.5 | 0.9 | 0.975 | 0.99 |
| Other genotypes | Organised | Lille | 57 | 6.91 | 4.32 | 4.67 | 5.33 | 6.78 | 8.67 | 9.73 | 10.44 |
| Other genotypes | Organised | Lille | 58 | 6.83 | 4.25 | 4.61 | 5.28 | 6.70 | 8.57 | 9.61 | 10.22 |
| Other genotypes | Organised | Lille | 59 | 6.78 | 4.23 | 4.60 | 5.23 | 6.65 | 8.49 | 9.59 | 10.19 |
| Other genotypes | Organised | Lille | 60 | 6.75 | 4.22 | 4.57 | 5.22 | 6.62 | 8.46 | 9.57 | 10.15 |
| Other genotypes | Organised | Lille | 61 | 6.76 | 4.24 | 4.59 | 5.23 | 6.63 | 8.48 | 9.59 | 10.20 |
| Other genotypes | Organised | Lille | 62 | 6.80 | 4.26 | 4.59 | 5.25 | 6.66 | 8.52 | 9.63 | 10.32 |
| Other genotypes | Organised | Lille | 63 | 6.90 | 4.30 | 4.66 | 5.29 | 6.77 | 8.66 | 9.78 | 10.42 |
| Other genotypes | Organised | Lille | 64 | 7.03 | 4.37 | 4.75 | 5.40 | 6.92 | 8.86 | 9.98 | 10.69 |
| Other genotypes | Organised | Lille | 65 | 7.20 | 4.45 | 4.82 | 5.52 | 7.08 | 9.12 | 10.27 | 10.99 |
| Other genotypes | Organised | Lille | 66 | 7.40 | 4.56 | 4.88 | 5.60 | 7.25 | 9.45 | 10.68 | 11.44 |
| Other genotypes | Organised | Strasbourg | 30 | 11.01 | 6.37 | 6.89 | 8.09 | 10.76 | 14.28 | 16.54 | 17.85 |
| Other genotypes | Organised | Strasbourg | 31 | 10.48 | 6.04 | 6.56 | 7.68 | 10.23 | 13.54 | 15.75 | 17.04 |
| Other genotypes | Organised | Strasbourg | 32 | 9.91 | 5.74 | 6.17 | 7.27 | 9.67 | 12.85 | 14.93 | 16.15 |
| Other genotypes | Organised | Strasbourg | 33 | 9.34 | 5.42 | 5.80 | 6.86 | 9.12 | 12.12 | 14.12 | 15.26 |
| Other genotypes | Organised | Strasbourg | 34 | 8.87 | 5.04 | 5.48 | 6.49 | 8.64 | 11.50 | 13.42 | 14.54 |
| Other genotypes | Organised | Strasbourg | 35 | 8.49 | 4.80 | 5.26 | 6.20 | 8.27 | 11.00 | 12.86 | 13.98 |
| Other genotypes | Organised | Strasbourg | 36 | 8.16 | 4.61 | 5.01 | 5.96 | 7.94 | 10.59 | 12.42 | 13.34 |
| Other genotypes | Organised | Strasbourg | 37 | 7.89 | 4.47 | 4.88 | 5.75 | 7.70 | 10.24 | 12.04 | 12.88 |
| Other genotypes | Organised | Strasbourg | 38 | 7.66 | 4.36 | 4.74 | 5.56 | 7.46 | 9.96 | 11.70 | 12.61 |
| Other genotypes | Organised | Strasbourg | 39 | 7.43 | 4.21 | 4.60 | 5.39 | 7.22 | 9.69 | 11.38 | 12.24 |
| Other genotypes | Organised | Strasbourg | 40 | 7.21 | 4.09 | 4.45 | 5.22 | 7.01 | 9.40 | 11.02 | 11.88 |
| Other genotypes | Organised | Strasbourg | 41 | 7.02 | 4.01 | 4.34 | 5.08 | 6.81 | 9.14 | 10.70 | 11.59 |
| Other genotypes | Organised | Strasbourg | 42 | 6.86 | 3.92 | 4.23 | 4.97 | 6.67 | 8.96 | 10.53 | 11.38 |
| Other genotypes | Organised | Strasbourg | 43 | 6.74 | 3.87 | 4.16 | 4.88 | 6.55 | 8.81 | 10.33 | 11.28 |
| Other genotypes | Organised | Strasbourg | 44 | 6.64 | 3.80 | 4.10 | 4.80 | 6.45 | 8.69 | 10.13 | 11.06 |
| Other genotypes | Organised | Strasbourg | 45 | 6.56 | 3.75 | 4.05 | 4.74 | 6.38 | 8.60 | 10.03 | 10.89 |
| Other genotypes | Organised | Strasbourg | 46 | 6.50 | 3.71 | 4.00 | 4.72 | 6.31 | 8.53 | 9.94 | 10.76 |
| Other genotypes | Organised | Strasbourg | 47 | 6.45 | 3.69 | 3.97 | 4.69 | 6.28 | 8.45 | 9.88 | 10.79 |
| Other genotypes | Organised | Strasbourg | 48 | 6.43 | 3.69 | 3.97 | 4.66 | 6.25 | 8.43 | 9.87 | 10.79 |

**Table S9 Posterior expected HR HPV cervical infection prevalence (in %) in major French cities, stratified by type of test, city, and age. Table counterpart to Figure 3A** ***(continued)***

| | | | | Posterior distribution | | | | | | | |
|---|---|---|---|---|---|---|---|---|---|---|---|
| | | | | | Percentiles | | | | | | |
| Virus | Test | City | Age | Average | 0.01 | 0.025 | 0.1 | 0.5 | 0.9 | 0.975 | 0.99 |
| Other genotypes | Organised | Strasbourg | 49 | 6.41 | 3.65 | 3.97 | 4.66 | 6.23 | 8.39 | 9.82 | 10.73 |
| Other genotypes | Organised | Strasbourg | 50 | 6.40 | 3.62 | 3.96 | 4.66 | 6.21 | 8.38 | 9.83 | 10.73 |
| Other genotypes | Organised | Strasbourg | 51 | 6.38 | 3.63 | 3.93 | 4.64 | 6.19 | 8.34 | 9.78 | 10.68 |
| Other genotypes | Organised | Strasbourg | 52 | 6.33 | 3.62 | 3.89 | 4.60 | 6.15 | 8.31 | 9.70 | 10.61 |
| Other genotypes | Organised | Strasbourg | 53 | 6.27 | 3.55 | 3.87 | 4.56 | 6.09 | 8.23 | 9.57 | 10.50 |
| Other genotypes | Organised | Strasbourg | 54 | 6.18 | 3.50 | 3.80 | 4.48 | 6.00 | 8.10 | 9.40 | 10.29 |
| Other genotypes | Organised | Strasbourg | 55 | 6.10 | 3.43 | 3.73 | 4.43 | 5.92 | 8.03 | 9.28 | 10.18 |
| Other genotypes | Organised | Strasbourg | 56 | 6.02 | 3.42 | 3.68 | 4.38 | 5.85 | 7.93 | 9.12 | 10.07 |
| Other genotypes | Organised | Strasbourg | 57 | 5.94 | 3.33 | 3.63 | 4.32 | 5.78 | 7.81 | 9.06 | 9.96 |
| Other genotypes | Organised | Strasbourg | 58 | 5.87 | 3.30 | 3.59 | 4.28 | 5.72 | 7.70 | 8.95 | 9.77 |
| Other genotypes | Organised | Strasbourg | 59 | 5.83 | 3.27 | 3.56 | 4.24 | 5.67 | 7.59 | 8.87 | 9.74 |
| Other genotypes | Organised | Strasbourg | 60 | 5.81 | 3.29 | 3.55 | 4.23 | 5.65 | 7.60 | 8.91 | 9.62 |
| Other genotypes | Organised | Strasbourg | 61 | 5.81 | 3.28 | 3.55 | 4.22 | 5.66 | 7.60 | 8.90 | 9.67 |
| Other genotypes | Organised | Strasbourg | 62 | 5.85 | 3.29 | 3.55 | 4.24 | 5.70 | 7.68 | 8.96 | 9.75 |
| Other genotypes | Organised | Strasbourg | 63 | 5.93 | 3.30 | 3.60 | 4.30 | 5.77 | 7.78 | 9.01 | 9.88 |
| Other genotypes | Organised | Strasbourg | 64 | 6.05 | 3.35 | 3.67 | 4.35 | 5.87 | 7.94 | 9.17 | 10.03 |
| Other genotypes | Organised | Strasbourg | 65 | 6.20 | 3.41 | 3.75 | 4.45 | 6.02 | 8.14 | 9.41 | 10.26 |
| Other genotypes | Organised | Strasbourg | 66 | 6.37 | 3.49 | 3.79 | 4.53 | 6.18 | 8.39 | 9.80 | 10.70 |
| Other genotypes | Organised | Lyon | 30 | 14.72 | 9.29 | 10.02 | 11.30 | 14.50 | 18.33 | 20.78 | 21.98 |
| Other genotypes | Organised | Lyon | 31 | 14.03 | 8.86 | 9.58 | 10.77 | 13.82 | 17.44 | 19.80 | 20.91 |
| Other genotypes | Organised | Lyon | 32 | 13.30 | 8.46 | 9.05 | 10.17 | 13.11 | 16.53 | 18.89 | 19.81 |
| Other genotypes | Organised | Lyon | 33 | 12.57 | 7.97 | 8.53 | 9.62 | 12.39 | 15.66 | 17.91 | 18.75 |
| Other genotypes | Organised | Lyon | 34 | 11.95 | 7.60 | 8.08 | 9.12 | 11.78 | 14.92 | 17.06 | 17.98 |
| Other genotypes | Organised | Lyon | 35 | 11.45 | 7.27 | 7.71 | 8.74 | 11.28 | 14.28 | 16.40 | 17.18 |
| Other genotypes | Organised | Lyon | 36 | 11.02 | 6.96 | 7.43 | 8.41 | 10.85 | 13.74 | 15.80 | 16.58 |
| Other genotypes | Organised | Lyon | 37 | 10.68 | 6.72 | 7.22 | 8.15 | 10.53 | 13.28 | 15.29 | 16.18 |
| Other genotypes | Organised | Lyon | 38 | 10.36 | 6.50 | 6.97 | 7.92 | 10.21 | 12.88 | 14.93 | 15.72 |
| Other genotypes | Organised | Lyon | 39 | 10.07 | 6.33 | 6.79 | 7.68 | 9.93 | 12.52 | 14.50 | 15.26 |
| Other genotypes | Organised | Lyon | 40 | 9.78 | 6.14 | 6.56 | 7.44 | 9.63 | 12.19 | 14.13 | 14.75 |

**Table S9** **Posterior expected HR HPV cervical infection prevalence (in %) in major French cities, stratified by type of test, city, and age. Table counterpart to Figure 3A** ***(continued)***

| | | | | | Posterior distribution | | | | | | |
|---|---|---|---|---|---|---|---|---|---|---|---|
| | | | | | Percentiles | | | | | | |
| Virus | Test | City | Age | Average | 0.01 | 0.025 | 0.1 | 0.5 | 0.9 | 0.975 | 0.99 |
| Other genotypes | Organised | Lyon | 41 | 9.52 | 5.93 | 6.37 | 7.23 | 9.39 | 11.89 | 13.73 | 14.38 |
| Other genotypes | Organised | Lyon | 42 | 9.32 | 5.80 | 6.24 | 7.08 | 9.19 | 11.63 | 13.52 | 14.10 |
| Other genotypes | Organised | Lyon | 43 | 9.16 | 5.69 | 6.11 | 6.97 | 9.03 | 11.45 | 13.26 | 13.93 |
| Other genotypes | Organised | Lyon | 44 | 9.02 | 5.61 | 6.00 | 6.86 | 8.90 | 11.27 | 13.06 | 13.81 |
| Other genotypes | Organised | Lyon | 45 | 8.91 | 5.53 | 5.91 | 6.76 | 8.76 | 11.17 | 12.88 | 13.73 |
| Other genotypes | Organised | Lyon | 46 | 8.83 | 5.44 | 5.85 | 6.68 | 8.69 | 11.06 | 12.73 | 13.56 |
| Other genotypes | Organised | Lyon | 47 | 8.77 | 5.45 | 5.81 | 6.65 | 8.65 | 10.99 | 12.70 | 13.47 |
| Other genotypes | Organised | Lyon | 48 | 8.74 | 5.44 | 5.82 | 6.62 | 8.63 | 10.94 | 12.66 | 13.41 |
| Other genotypes | Organised | Lyon | 49 | 8.72 | 5.41 | 5.81 | 6.62 | 8.60 | 10.92 | 12.61 | 13.39 |
| Other genotypes | Organised | Lyon | 50 | 8.70 | 5.40 | 5.81 | 6.61 | 8.58 | 10.90 | 12.60 | 13.36 |
| Other genotypes | Organised | Lyon | 51 | 8.68 | 5.37 | 5.80 | 6.58 | 8.56 | 10.90 | 12.57 | 13.32 |
| Other genotypes | Organised | Lyon | 52 | 8.61 | 5.34 | 5.74 | 6.53 | 8.50 | 10.82 | 12.45 | 13.27 |
| Other genotypes | Organised | Lyon | 53 | 8.54 | 5.31 | 5.66 | 6.46 | 8.42 | 10.69 | 12.33 | 13.19 |
| Other genotypes | Organised | Lyon | 54 | 8.42 | 5.23 | 5.57 | 6.37 | 8.31 | 10.58 | 12.24 | 13.00 |
| Other genotypes | Organised | Lyon | 55 | 8.31 | 5.14 | 5.52 | 6.29 | 8.19 | 10.43 | 12.04 | 13.00 |
| Other genotypes | Organised | Lyon | 56 | 8.21 | 5.07 | 5.44 | 6.21 | 8.10 | 10.31 | 11.97 | 12.93 |
| Other genotypes | Organised | Lyon | 57 | 8.10 | 5.00 | 5.37 | 6.12 | 8.00 | 10.18 | 11.80 | 12.69 |
| Other genotypes | Organised | Lyon | 58 | 8.01 | 4.92 | 5.32 | 6.05 | 7.90 | 10.08 | 11.67 | 12.53 |
| Other genotypes | Organised | Lyon | 59 | 7.95 | 4.88 | 5.28 | 6.00 | 7.83 | 10.03 | 11.58 | 12.45 |
| Other genotypes | Organised | Lyon | 60 | 7.92 | 4.89 | 5.26 | 5.99 | 7.81 | 10.01 | 11.55 | 12.41 |
| Other genotypes | Organised | Lyon | 61 | 7.93 | 4.88 | 5.24 | 5.97 | 7.79 | 10.00 | 11.59 | 12.46 |
| Other genotypes | Organised | Lyon | 62 | 7.98 | 4.90 | 5.26 | 6.00 | 7.86 | 10.09 | 11.66 | 12.58 |
| Other genotypes | Organised | Lyon | 63 | 8.09 | 4.99 | 5.31 | 6.06 | 7.95 | 10.23 | 11.76 | 12.76 |
| Other genotypes | Organised | Lyon | 64 | 8.24 | 5.01 | 5.40 | 6.16 | 8.10 | 10.46 | 12.01 | 12.98 |
| Other genotypes | Organised | Lyon | 65 | 8.44 | 5.06 | 5.51 | 6.32 | 8.29 | 10.76 | 12.36 | 13.29 |
| Other genotypes | Organised | Lyon | 66 | 8.67 | 5.13 | 5.63 | 6.46 | 8.53 | 11.14 | 12.76 | 13.73 |
| Other genotypes | Organised | Paris | 30 | 16.37 | 11.88 | 12.57 | 13.76 | 16.26 | 19.16 | 20.68 | 21.56 |
| Other genotypes | Organised | Paris | 31 | 15.61 | 11.43 | 12.02 | 13.16 | 15.52 | 18.28 | 19.71 | 20.59 |
| Other genotypes | Organised | Paris | 32 | 14.81 | 10.82 | 11.39 | 12.49 | 14.72 | 17.32 | 18.67 | 19.58 |

**Table S9** **Posterior expected HR HPV cervical infection prevalence (in %) in major French cities, stratified by type of test, city, and age. Table counterpart to Figure 3A** ***(continued)***

| | | | | Posterior distribution | | | | | | | |
|---|---|---|---|---|---|---|---|---|---|---|---|
| | | | | | Percentiles | | | | | | |
| Virus | Test | City | Age | Average | 0.01 | 0.025 | 0.1 | 0.5 | 0.9 | 0.975 | 0.99 |
| Other genotypes | Organised | Paris | 33 | 14.01 | 10.19 | 10.76 | 11.78 | 13.91 | 16.35 | 17.69 | 18.39 |
| Other genotypes | Organised | Paris | 34 | 13.33 | 9.74 | 10.26 | 11.23 | 13.24 | 15.58 | 16.86 | 17.45 |
| Other genotypes | Organised | Paris | 35 | 12.79 | 9.30 | 9.87 | 10.75 | 12.71 | 14.94 | 16.24 | 16.73 |
| Other genotypes | Organised | Paris | 36 | 12.31 | 8.98 | 9.48 | 10.34 | 12.24 | 14.38 | 15.67 | 16.16 |
| Other genotypes | Organised | Paris | 37 | 11.93 | 8.73 | 9.18 | 10.01 | 11.87 | 13.91 | 15.16 | 15.69 |
| Other genotypes | Organised | Paris | 38 | 11.58 | 8.49 | 8.92 | 9.71 | 11.52 | 13.54 | 14.73 | 15.15 |
| Other genotypes | Organised | Paris | 39 | 11.26 | 8.27 | 8.68 | 9.43 | 11.20 | 13.16 | 14.40 | 14.81 |
| Other genotypes | Organised | Paris | 40 | 10.94 | 7.96 | 8.38 | 9.16 | 10.87 | 12.78 | 14.02 | 14.46 |
| Other genotypes | Organised | Paris | 41 | 10.65 | 7.78 | 8.14 | 8.89 | 10.58 | 12.48 | 13.61 | 14.16 |
| Other genotypes | Organised | Paris | 42 | 10.43 | 7.61 | 7.94 | 8.72 | 10.36 | 12.24 | 13.35 | 13.88 |
| Other genotypes | Organised | Paris | 43 | 10.25 | 7.49 | 7.82 | 8.57 | 10.19 | 12.06 | 13.13 | 13.69 |
| Other genotypes | Organised | Paris | 44 | 10.10 | 7.40 | 7.68 | 8.44 | 10.03 | 11.89 | 12.99 | 13.52 |
| Other genotypes | Organised | Paris | 45 | 9.98 | 7.30 | 7.56 | 8.30 | 9.91 | 11.73 | 12.86 | 13.39 |
| Other genotypes | Organised | Paris | 46 | 9.89 | 7.23 | 7.52 | 8.22 | 9.83 | 11.60 | 12.74 | 13.28 |
| Other genotypes | Organised | Paris | 47 | 9.82 | 7.18 | 7.47 | 8.18 | 9.77 | 11.52 | 12.61 | 13.21 |
| Other genotypes | Organised | Paris | 48 | 9.79 | 7.14 | 7.44 | 8.14 | 9.74 | 11.48 | 12.58 | 13.15 |
| Other genotypes | Organised | Paris | 49 | 9.77 | 7.12 | 7.43 | 8.14 | 9.71 | 11.45 | 12.57 | 13.21 |
| Other genotypes | Organised | Paris | 50 | 9.75 | 7.13 | 7.40 | 8.12 | 9.68 | 11.45 | 12.52 | 13.06 |
| Other genotypes | Organised | Paris | 51 | 9.72 | 7.08 | 7.41 | 8.09 | 9.65 | 11.40 | 12.48 | 13.00 |
| Other genotypes | Organised | Paris | 52 | 9.65 | 7.03 | 7.36 | 8.01 | 9.58 | 11.33 | 12.35 | 12.97 |
| Other genotypes | Organised | Paris | 53 | 9.56 | 6.98 | 7.27 | 7.93 | 9.49 | 11.21 | 12.26 | 12.86 |
| Other genotypes | Organised | Paris | 54 | 9.43 | 6.86 | 7.15 | 7.82 | 9.36 | 11.05 | 12.09 | 12.61 |
| Other genotypes | Organised | Paris | 55 | 9.30 | 6.74 | 7.04 | 7.73 | 9.23 | 10.92 | 11.89 | 12.42 |
| Other genotypes | Organised | Paris | 56 | 9.20 | 6.67 | 6.98 | 7.64 | 9.13 | 10.76 | 11.79 | 12.23 |
| Other genotypes | Organised | Paris | 57 | 9.07 | 6.58 | 6.88 | 7.55 | 9.01 | 10.63 | 11.64 | 12.06 |
| Other genotypes | Organised | Paris | 58 | 8.97 | 6.50 | 6.85 | 7.47 | 8.91 | 10.54 | 11.53 | 12.00 |
| Other genotypes | Organised | Paris | 59 | 8.90 | 6.47 | 6.77 | 7.41 | 8.85 | 10.44 | 11.43 | 11.98 |
| Other genotypes | Organised | Paris | 60 | 8.87 | 6.46 | 6.75 | 7.37 | 8.83 | 10.39 | 11.36 | 11.94 |
| Other genotypes | Organised | Paris | 61 | 8.88 | 6.45 | 6.76 | 7.37 | 8.85 | 10.42 | 11.35 | 11.91 |

**Table S9** **Posterior expected HR HPV cervical infection prevalence (in %) in major French cities, stratified by type of test, city, and age. Table counterpart to Figure 3A** ***(continued)***

| | | | | Posterior distribution | | | | | | | |
|---|---|---|---|---|---|---|---|---|---|---|---|
| | | | | | Percentiles | | | | | | |
| Virus | Test | City | Age | Average | 0.01 | 0.025 | 0.1 | 0.5 | 0.9 | 0.975 | 0.99 |
| Other genotypes | Organised | Paris | 62 | 8.94 | 6.44 | 6.78 | 7.44 | 8.90 | 10.50 | 11.48 | 12.11 |
| Other genotypes | Organised | Paris | 63 | 9.06 | 6.49 | 6.85 | 7.52 | 9.01 | 10.64 | 11.65 | 12.25 |
| Other genotypes | Organised | Paris | 64 | 9.23 | 6.57 | 6.93 | 7.63 | 9.17 | 10.85 | 11.93 | 12.50 |
| Other genotypes | Organised | Paris | 65 | 9.45 | 6.65 | 7.03 | 7.76 | 9.37 | 11.15 | 12.33 | 12.88 |
| Other genotypes | Organised | Paris | 66 | 9.71 | 6.70 | 7.14 | 7.88 | 9.62 | 11.55 | 12.84 | 13.60 |
| Other genotypes | Opportunistic | Nice | 30 | 19.10 | 10.41 | 11.48 | 13.74 | 18.77 | 24.92 | 28.53 | 30.86 |
| Other genotypes | Opportunistic | Nice | 31 | 18.21 | 9.87 | 10.90 | 13.04 | 17.91 | 23.87 | 27.22 | 29.58 |
| Other genotypes | Opportunistic | Nice | 32 | 17.26 | 9.33 | 10.32 | 12.29 | 16.95 | 22.75 | 25.98 | 28.08 |
| Other genotypes | Opportunistic | Nice | 33 | 16.29 | 8.77 | 9.66 | 11.56 | 16.01 | 21.49 | 24.76 | 26.67 |
| Other genotypes | Opportunistic | Nice | 34 | 15.45 | 8.28 | 9.07 | 10.92 | 15.16 | 20.45 | 23.57 | 25.63 |
| Other genotypes | Opportunistic | Nice | 35 | 14.77 | 7.88 | 8.65 | 10.41 | 14.47 | 19.58 | 22.59 | 24.68 |
| Other genotypes | Opportunistic | Nice | 36 | 14.15 | 7.48 | 8.24 | 9.98 | 13.88 | 18.73 | 21.76 | 23.82 |
| Other genotypes | Opportunistic | Nice | 37 | 13.65 | 7.25 | 7.94 | 9.59 | 13.34 | 18.15 | 21.05 | 23.02 |
| Other genotypes | Opportunistic | Nice | 38 | 13.18 | 7.01 | 7.59 | 9.24 | 12.88 | 17.57 | 20.38 | 22.24 |
| Other genotypes | Opportunistic | Nice | 39 | 12.75 | 6.71 | 7.37 | 8.92 | 12.45 | 17.00 | 19.81 | 21.53 |
| Other genotypes | Opportunistic | Nice | 40 | 12.30 | 6.46 | 7.11 | 8.60 | 12.00 | 16.45 | 19.15 | 20.97 |
| Other genotypes | Opportunistic | Nice | 41 | 11.91 | 6.18 | 6.88 | 8.30 | 11.61 | 15.91 | 18.53 | 20.14 |
| Other genotypes | Opportunistic | Nice | 42 | 11.59 | 5.98 | 6.66 | 8.08 | 11.32 | 15.49 | 18.06 | 19.83 |
| Other genotypes | Opportunistic | Nice | 43 | 11.31 | 5.83 | 6.51 | 7.88 | 11.05 | 15.13 | 17.61 | 19.58 |
| Other genotypes | Opportunistic | Nice | 44 | 11.08 | 5.71 | 6.36 | 7.72 | 10.80 | 14.86 | 17.31 | 19.01 |
| Other genotypes | Opportunistic | Nice | 45 | 10.87 | 5.58 | 6.25 | 7.54 | 10.61 | 14.62 | 17.00 | 18.59 |
| Other genotypes | Opportunistic | Nice | 46 | 10.70 | 5.47 | 6.13 | 7.45 | 10.46 | 14.39 | 16.76 | 18.35 |
| Other genotypes | Opportunistic | Nice | 47 | 10.56 | 5.42 | 6.05 | 7.34 | 10.31 | 14.26 | 16.54 | 18.18 |
| Other genotypes | Opportunistic | Nice | 48 | 10.45 | 5.37 | 5.95 | 7.26 | 10.19 | 14.11 | 16.35 | 17.99 |
| Other genotypes | Opportunistic | Nice | 49 | 10.35 | 5.29 | 5.89 | 7.19 | 10.08 | 14.00 | 16.15 | 18.02 |
| Other genotypes | Opportunistic | Nice | 50 | 10.25 | 5.29 | 5.85 | 7.09 | 9.99 | 13.89 | 16.05 | 17.76 |
| Other genotypes | Opportunistic | Nice | 51 | 10.14 | 5.26 | 5.76 | 7.01 | 9.88 | 13.72 | 15.83 | 17.63 |
| Other genotypes | Opportunistic | Nice | 52 | 9.98 | 5.15 | 5.71 | 6.89 | 9.73 | 13.47 | 15.66 | 17.42 |
| Other genotypes | Opportunistic | Nice | 53 | 9.79 | 5.05 | 5.55 | 6.75 | 9.55 | 13.22 | 15.47 | 17.16 |

**Table S9** **Posterior expected HR HPV cervical infection prevalence (in %) in major French cities, stratified by type of test, city, and age. Table counterpart to Figure 3A** ***(continued)***

| | | | | Posterior distribution | | | | | | | |
|---|---|---|---|---|---|---|---|---|---|---|---|
| | | | | | Percentiles | | | | | | |
| Virus | Test | City | Age | Average | 0.01 | 0.025 | 0.1 | 0.5 | 0.9 | 0.975 | 0.99 |
| Other genotypes | Opportunistic | Nice | 54 | 9.56 | 4.91 | 5.42 | 6.58 | 9.30 | 12.95 | 15.07 | 16.78 |
| Other genotypes | Opportunistic | Nice | 55 | 9.33 | 4.82 | 5.28 | 6.42 | 9.08 | 12.61 | 14.72 | 16.41 |
| Other genotypes | Opportunistic | Nice | 56 | 9.12 | 4.67 | 5.19 | 6.31 | 8.88 | 12.31 | 14.37 | 16.09 |
| Other genotypes | Opportunistic | Nice | 57 | 8.89 | 4.52 | 5.05 | 6.14 | 8.66 | 11.97 | 14.03 | 15.74 |
| Other genotypes | Opportunistic | Nice | 58 | 8.70 | 4.40 | 4.93 | 6.01 | 8.47 | 11.71 | 13.76 | 15.46 |
| Other genotypes | Opportunistic | Nice | 59 | 8.54 | 4.33 | 4.81 | 5.88 | 8.30 | 11.56 | 13.51 | 15.10 |
| Other genotypes | Opportunistic | Nice | 60 | 8.44 | 4.27 | 4.74 | 5.82 | 8.20 | 11.40 | 13.34 | 14.90 |
| Other genotypes | Opportunistic | Nice | 61 | 8.37 | 4.27 | 4.71 | 5.77 | 8.16 | 11.31 | 13.24 | 14.79 |
| Other genotypes | Opportunistic | Nice | 62 | 8.36 | 4.30 | 4.70 | 5.75 | 8.16 | 11.30 | 13.25 | 14.76 |
| Other genotypes | Opportunistic | Nice | 63 | 8.41 | 4.33 | 4.79 | 5.76 | 8.21 | 11.37 | 13.32 | 14.85 |
| Other genotypes | Opportunistic | Nice | 64 | 8.51 | 4.35 | 4.86 | 5.85 | 8.29 | 11.49 | 13.41 | 15.00 |
| Other genotypes | Opportunistic | Nice | 65 | 8.66 | 4.37 | 4.93 | 5.94 | 8.44 | 11.72 | 13.64 | 15.16 |
| Other genotypes | Opportunistic | Nice | 66 | 8.84 | 4.43 | 5.08 | 6.06 | 8.57 | 11.99 | 13.96 | 15.54 |
| Other genotypes | Opportunistic | Marseille | 30 | 20.76 | 16.67 | 17.20 | 18.39 | 20.75 | 23.18 | 24.49 | 25.42 |
| Other genotypes | Opportunistic | Marseille | 31 | 19.81 | 15.84 | 16.40 | 17.56 | 19.79 | 22.11 | 23.37 | 24.25 |
| Other genotypes | Opportunistic | Marseille | 32 | 18.78 | 15.06 | 15.53 | 16.67 | 18.74 | 20.99 | 22.22 | 23.06 |
| Other genotypes | Opportunistic | Marseille | 33 | 17.74 | 14.09 | 14.70 | 15.72 | 17.69 | 19.84 | 21.05 | 21.84 |
| Other genotypes | Opportunistic | Marseille | 34 | 16.83 | 13.33 | 13.94 | 14.89 | 16.79 | 18.86 | 20.05 | 20.70 |
| Other genotypes | Opportunistic | Marseille | 35 | 16.09 | 12.75 | 13.27 | 14.21 | 16.04 | 18.06 | 19.23 | 19.78 |
| Other genotypes | Opportunistic | Marseille | 36 | 15.42 | 12.21 | 12.64 | 13.58 | 15.40 | 17.30 | 18.40 | 19.05 |
| Other genotypes | Opportunistic | Marseille | 37 | 14.88 | 11.78 | 12.14 | 13.09 | 14.85 | 16.73 | 17.74 | 18.41 |
| Other genotypes | Opportunistic | Marseille | 38 | 14.37 | 11.39 | 11.72 | 12.63 | 14.36 | 16.15 | 17.17 | 17.83 |
| Other genotypes | Opportunistic | Marseille | 39 | 13.90 | 11.02 | 11.34 | 12.24 | 13.89 | 15.61 | 16.63 | 17.23 |
| Other genotypes | Opportunistic | Marseille | 40 | 13.42 | 10.55 | 10.93 | 11.78 | 13.40 | 15.11 | 16.09 | 16.61 |
| Other genotypes | Opportunistic | Marseille | 41 | 12.99 | 10.23 | 10.58 | 11.39 | 12.97 | 14.65 | 15.58 | 16.07 |
| Other genotypes | Opportunistic | Marseille | 42 | 12.64 | 9.90 | 10.32 | 11.10 | 12.61 | 14.27 | 15.14 | 15.62 |
| Other genotypes | Opportunistic | Marseille | 43 | 12.35 | 9.65 | 10.02 | 10.83 | 12.31 | 13.92 | 14.84 | 15.33 |
| Other genotypes | Opportunistic | Marseille | 44 | 12.09 | 9.51 | 9.82 | 10.60 | 12.05 | 13.62 | 14.56 | 15.03 |
| Other genotypes | Opportunistic | Marseille | 45 | 11.87 | 9.27 | 9.65 | 10.40 | 11.84 | 13.40 | 14.30 | 14.78 |

**Table S9** **Posterior expected HR HPV cervical infection prevalence (in %) in major French cities, stratified by type of test, city, and age. Table counterpart to Figure 3A** ***(continued)***

| | | | | | Posterior distribution | | | | | | |
|---|---|---|---|---|---|---|---|---|---|---|---|
| | | | | | Percentiles | | | | | | |
| Virus | Test | City | Age | Average | 0.01 | 0.025 | 0.1 | 0.5 | 0.9 | 0.975 | 0.99 |
| Other genotypes | Opportunistic | Marseille | 46 | 11.68 | 9.11 | 9.51 | 10.23 | 11.65 | 13.22 | 14.14 | 14.55 |
| Other genotypes | Opportunistic | Marseille | 47 | 11.53 | 9.00 | 9.41 | 10.09 | 11.49 | 13.02 | 13.91 | 14.35 |
| Other genotypes | Opportunistic | Marseille | 48 | 11.41 | 8.90 | 9.27 | 9.99 | 11.36 | 12.90 | 13.79 | 14.19 |
| Other genotypes | Opportunistic | Marseille | 49 | 11.30 | 8.85 | 9.18 | 9.89 | 11.27 | 12.76 | 13.63 | 14.07 |
| Other genotypes | Opportunistic | Marseille | 50 | 11.19 | 8.78 | 9.05 | 9.80 | 11.17 | 12.64 | 13.48 | 14.00 |
| Other genotypes | Opportunistic | Marseille | 51 | 11.07 | 8.66 | 8.96 | 9.67 | 11.04 | 12.54 | 13.37 | 13.79 |
| Other genotypes | Opportunistic | Marseille | 52 | 10.89 | 8.58 | 8.79 | 9.53 | 10.88 | 12.30 | 13.15 | 13.58 |
| Other genotypes | Opportunistic | Marseille | 53 | 10.69 | 8.39 | 8.67 | 9.35 | 10.66 | 12.07 | 12.95 | 13.44 |
| Other genotypes | Opportunistic | Marseille | 54 | 10.43 | 8.17 | 8.50 | 9.12 | 10.41 | 11.80 | 12.62 | 13.18 |
| Other genotypes | Opportunistic | Marseille | 55 | 10.19 | 7.92 | 8.28 | 8.92 | 10.17 | 11.55 | 12.30 | 12.78 |
| Other genotypes | Opportunistic | Marseille | 56 | 9.96 | 7.74 | 8.07 | 8.70 | 9.94 | 11.29 | 12.10 | 12.53 |
| Other genotypes | Opportunistic | Marseille | 57 | 9.71 | 7.50 | 7.91 | 8.48 | 9.69 | 11.03 | 11.82 | 12.24 |
| Other genotypes | Opportunistic | Marseille | 58 | 9.50 | 7.37 | 7.70 | 8.31 | 9.47 | 10.78 | 11.56 | 11.97 |
| Other genotypes | Opportunistic | Marseille | 59 | 9.34 | 7.28 | 7.56 | 8.13 | 9.30 | 10.59 | 11.38 | 11.75 |
| Other genotypes | Opportunistic | Marseille | 60 | 9.22 | 7.19 | 7.47 | 8.01 | 9.20 | 10.45 | 11.20 | 11.63 |
| Other genotypes | Opportunistic | Marseille | 61 | 9.15 | 7.11 | 7.43 | 7.95 | 9.13 | 10.38 | 11.11 | 11.56 |
| Other genotypes | Opportunistic | Marseille | 62 | 9.14 | 7.09 | 7.43 | 7.96 | 9.11 | 10.37 | 11.05 | 11.58 |
| Other genotypes | Opportunistic | Marseille | 63 | 9.19 | 7.14 | 7.43 | 7.98 | 9.16 | 10.44 | 11.11 | 11.58 |
| Other genotypes | Opportunistic | Marseille | 64 | 9.30 | 7.23 | 7.53 | 8.08 | 9.27 | 10.57 | 11.26 | 11.70 |
| Other genotypes | Opportunistic | Marseille | 65 | 9.46 | 7.30 | 7.62 | 8.18 | 9.42 | 10.79 | 11.51 | 11.97 |
| Other genotypes | Opportunistic | Marseille | 66 | 9.65 | 7.38 | 7.70 | 8.31 | 9.59 | 11.09 | 11.86 | 12.43 |
| Other genotypes | Opportunistic | Toulouse | 30 | 19.46 | 12.21 | 13.05 | 15.14 | 19.27 | 24.23 | 26.94 | 28.53 |
| Other genotypes | Opportunistic | Toulouse | 31 | 18.56 | 11.71 | 12.40 | 14.42 | 18.35 | 23.16 | 25.83 | 27.31 |
| Other genotypes | Opportunistic | Toulouse | 32 | 17.58 | 11.07 | 11.67 | 13.61 | 17.41 | 21.95 | 24.58 | 25.95 |
| Other genotypes | Opportunistic | Toulouse | 33 | 16.59 | 10.36 | 10.93 | 12.85 | 16.40 | 20.78 | 23.22 | 24.60 |
| Other genotypes | Opportunistic | Toulouse | 34 | 15.74 | 9.75 | 10.36 | 12.14 | 15.54 | 19.78 | 22.10 | 23.41 |
| Other genotypes | Opportunistic | Toulouse | 35 | 15.04 | 9.35 | 9.85 | 11.57 | 14.87 | 18.90 | 21.17 | 22.48 |
| Other genotypes | Opportunistic | Toulouse | 36 | 14.41 | 8.85 | 9.41 | 11.04 | 14.22 | 18.16 | 20.40 | 21.72 |
| Other genotypes | Opportunistic | Toulouse | 37 | 13.90 | 8.52 | 9.04 | 10.63 | 13.69 | 17.50 | 19.82 | 21.03 |

**Table S9** **Posterior expected HR HPV cervical infection prevalence (in %) in major French cities, stratified by type of test, city, and age. Table counterpart to Figure 3A** ***(continued)***

| | | | | Posterior distribution | | | | | | | |
|---|---|---|---|---|---|---|---|---|---|---|---|
| | | | | | Percentiles | | | | | | |
| Virus | Test | City | Age | Average | 0.01 | 0.025 | 0.1 | 0.5 | 0.9 | 0.975 | 0.99 |
| Other genotypes | Opportunistic | Toulouse | 38 | 13.43 | 8.16 | 8.73 | 10.28 | 13.25 | 16.97 | 19.09 | 20.35 |
| Other genotypes | Opportunistic | Toulouse | 39 | 12.98 | 7.91 | 8.45 | 9.92 | 12.80 | 16.39 | 18.49 | 19.86 |
| Other genotypes | Opportunistic | Toulouse | 40 | 12.53 | 7.66 | 8.06 | 9.57 | 12.35 | 15.81 | 17.93 | 19.21 |
| Other genotypes | Opportunistic | Toulouse | 41 | 12.13 | 7.41 | 7.81 | 9.26 | 11.94 | 15.35 | 17.32 | 18.55 |
| Other genotypes | Opportunistic | Toulouse | 42 | 11.80 | 7.18 | 7.61 | 8.99 | 11.62 | 14.95 | 16.87 | 18.04 |
| Other genotypes | Opportunistic | Toulouse | 43 | 11.52 | 7.06 | 7.46 | 8.77 | 11.34 | 14.63 | 16.51 | 17.69 |
| Other genotypes | Opportunistic | Toulouse | 44 | 11.28 | 6.84 | 7.27 | 8.62 | 11.10 | 14.31 | 16.13 | 17.30 |
| Other genotypes | Opportunistic | Toulouse | 45 | 11.07 | 6.70 | 7.12 | 8.44 | 10.88 | 14.03 | 15.88 | 17.02 |
| Other genotypes | Opportunistic | Toulouse | 46 | 10.90 | 6.54 | 7.00 | 8.30 | 10.72 | 13.84 | 15.64 | 16.80 |
| Other genotypes | Opportunistic | Toulouse | 47 | 10.75 | 6.45 | 6.93 | 8.20 | 10.55 | 13.67 | 15.40 | 16.59 |
| Other genotypes | Opportunistic | Toulouse | 48 | 10.64 | 6.42 | 6.85 | 8.11 | 10.45 | 13.56 | 15.34 | 16.48 |
| Other genotypes | Opportunistic | Toulouse | 49 | 10.54 | 6.35 | 6.80 | 8.00 | 10.35 | 13.45 | 15.12 | 16.34 |
| Other genotypes | Opportunistic | Toulouse | 50 | 10.44 | 6.25 | 6.72 | 7.93 | 10.25 | 13.29 | 15.09 | 16.25 |
| Other genotypes | Opportunistic | Toulouse | 51 | 10.32 | 6.20 | 6.61 | 7.82 | 10.14 | 13.15 | 14.91 | 15.96 |
| Other genotypes | Opportunistic | Toulouse | 52 | 10.16 | 6.10 | 6.50 | 7.69 | 9.98 | 12.93 | 14.68 | 15.82 |
| Other genotypes | Opportunistic | Toulouse | 53 | 9.97 | 6.01 | 6.41 | 7.55 | 9.79 | 12.65 | 14.40 | 15.48 |
| Other genotypes | Opportunistic | Toulouse | 54 | 9.73 | 5.84 | 6.23 | 7.37 | 9.56 | 12.38 | 14.02 | 15.04 |
| Other genotypes | Opportunistic | Toulouse | 55 | 9.50 | 5.67 | 6.07 | 7.19 | 9.34 | 12.12 | 13.81 | 14.74 |
| Other genotypes | Opportunistic | Toulouse | 56 | 9.28 | 5.55 | 5.96 | 7.01 | 9.11 | 11.86 | 13.52 | 14.43 |
| Other genotypes | Opportunistic | Toulouse | 57 | 9.05 | 5.45 | 5.81 | 6.83 | 8.89 | 11.57 | 13.13 | 14.16 |
| Other genotypes | Opportunistic | Toulouse | 58 | 8.86 | 5.30 | 5.69 | 6.69 | 8.70 | 11.31 | 12.91 | 13.92 |
| Other genotypes | Opportunistic | Toulouse | 59 | 8.70 | 5.20 | 5.55 | 6.57 | 8.57 | 11.12 | 12.71 | 13.63 |
| Other genotypes | Opportunistic | Toulouse | 60 | 8.59 | 5.12 | 5.48 | 6.47 | 8.45 | 10.99 | 12.53 | 13.39 |
| Other genotypes | Opportunistic | Toulouse | 61 | 8.53 | 5.10 | 5.42 | 6.43 | 8.38 | 10.91 | 12.44 | 13.33 |
| Other genotypes | Opportunistic | Toulouse | 62 | 8.52 | 5.06 | 5.40 | 6.43 | 8.37 | 10.89 | 12.46 | 13.28 |
| Other genotypes | Opportunistic | Toulouse | 63 | 8.57 | 5.07 | 5.45 | 6.44 | 8.41 | 10.97 | 12.52 | 13.39 |
| Other genotypes | Opportunistic | Toulouse | 64 | 8.67 | 5.17 | 5.51 | 6.53 | 8.52 | 11.09 | 12.64 | 13.56 |
| Other genotypes | Opportunistic | Toulouse | 65 | 8.82 | 5.26 | 5.56 | 6.63 | 8.65 | 11.30 | 12.85 | 13.77 |
| Other genotypes | Opportunistic | Toulouse | 66 | 9.00 | 5.30 | 5.68 | 6.71 | 8.82 | 11.56 | 13.19 | 14.17 |

**Table S9** **Posterior expected HR HPV cervical infection prevalence (in %) in major French cities, stratified by type of test, city, and age. Table counterpart to Figure 3A** ***(continued)***

| | | | | | Posterior distribution | | | | | | |
|---|---|---|---|---|---|---|---|---|---|---|---|
| | | | | | Percentiles | | | | | | |
| Virus | Test | City | Age | Average | 0.01 | 0.025 | 0.1 | 0.5 | 0.9 | 0.975 | 0.99 |
| Other genotypes | Opportunistic | Bordeaux | 30 | 21.29 | 16.78 | 17.43 | 18.56 | 21.13 | 24.21 | 25.99 | 27.04 |
| Other genotypes | Opportunistic | Bordeaux | 31 | 20.32 | 15.98 | 16.63 | 17.70 | 20.21 | 23.12 | 24.86 | 25.89 |
| Other genotypes | Opportunistic | Bordeaux | 32 | 19.27 | 15.14 | 15.73 | 16.74 | 19.13 | 21.91 | 23.61 | 24.64 |
| Other genotypes | Opportunistic | Bordeaux | 33 | 18.20 | 14.28 | 14.79 | 15.78 | 18.10 | 20.77 | 22.37 | 23.44 |
| Other genotypes | Opportunistic | Bordeaux | 34 | 17.28 | 13.51 | 14.06 | 14.98 | 17.16 | 19.77 | 21.35 | 22.31 |
| Other genotypes | Opportunistic | Bordeaux | 35 | 16.52 | 12.91 | 13.43 | 14.30 | 16.40 | 18.93 | 20.46 | 21.44 |
| Other genotypes | Opportunistic | Bordeaux | 36 | 15.84 | 12.34 | 12.80 | 13.67 | 15.71 | 18.16 | 19.65 | 20.60 |
| Other genotypes | Opportunistic | Bordeaux | 37 | 15.28 | 11.90 | 12.31 | 13.20 | 15.16 | 17.53 | 18.97 | 19.85 |
| Other genotypes | Opportunistic | Bordeaux | 38 | 14.77 | 11.48 | 11.91 | 12.74 | 14.65 | 16.97 | 18.33 | 19.31 |
| Other genotypes | Opportunistic | Bordeaux | 39 | 14.29 | 11.10 | 11.48 | 12.31 | 14.17 | 16.42 | 17.77 | 18.58 |
| Other genotypes | Opportunistic | Bordeaux | 40 | 13.79 | 10.63 | 11.05 | 11.86 | 13.67 | 15.85 | 17.17 | 17.98 |
| Other genotypes | Opportunistic | Bordeaux | 41 | 13.36 | 10.23 | 10.70 | 11.47 | 13.25 | 15.35 | 16.61 | 17.33 |
| Other genotypes | Opportunistic | Bordeaux | 42 | 13.00 | 9.99 | 10.42 | 11.15 | 12.89 | 14.95 | 16.17 | 16.98 |
| Other genotypes | Opportunistic | Bordeaux | 43 | 12.70 | 9.78 | 10.16 | 10.88 | 12.59 | 14.61 | 15.76 | 16.54 |
| Other genotypes | Opportunistic | Bordeaux | 44 | 12.43 | 9.56 | 9.96 | 10.67 | 12.32 | 14.31 | 15.48 | 16.20 |
| Other genotypes | Opportunistic | Bordeaux | 45 | 12.20 | 9.41 | 9.75 | 10.48 | 12.12 | 14.07 | 15.15 | 15.86 |
| Other genotypes | Opportunistic | Bordeaux | 46 | 12.02 | 9.24 | 9.64 | 10.31 | 11.93 | 13.89 | 14.95 | 15.75 |
| Other genotypes | Opportunistic | Bordeaux | 47 | 11.86 | 9.11 | 9.47 | 10.16 | 11.76 | 13.69 | 14.81 | 15.57 |
| Other genotypes | Opportunistic | Bordeaux | 48 | 11.73 | 8.97 | 9.37 | 10.08 | 11.63 | 13.55 | 14.66 | 15.40 |
| Other genotypes | Opportunistic | Bordeaux | 49 | 11.62 | 8.90 | 9.27 | 9.97 | 11.52 | 13.40 | 14.57 | 15.23 |
| Other genotypes | Opportunistic | Bordeaux | 50 | 11.51 | 8.81 | 9.17 | 9.87 | 11.41 | 13.27 | 14.38 | 15.12 |
| Other genotypes | Opportunistic | Bordeaux | 51 | 11.39 | 8.70 | 9.10 | 9.76 | 11.29 | 13.13 | 14.25 | 14.91 |
| Other genotypes | Opportunistic | Bordeaux | 52 | 11.21 | 8.59 | 8.94 | 9.60 | 11.10 | 12.93 | 14.02 | 14.70 |
| Other genotypes | Opportunistic | Bordeaux | 53 | 11.00 | 8.42 | 8.77 | 9.43 | 10.90 | 12.69 | 13.82 | 14.43 |
| Other genotypes | Opportunistic | Bordeaux | 54 | 10.74 | 8.23 | 8.57 | 9.20 | 10.64 | 12.41 | 13.46 | 14.07 |
| Other genotypes | Opportunistic | Bordeaux | 55 | 10.48 | 8.02 | 8.33 | 8.97 | 10.38 | 12.13 | 13.17 | 13.73 |
| Other genotypes | Opportunistic | Bordeaux | 56 | 10.25 | 7.82 | 8.19 | 8.77 | 10.15 | 11.86 | 12.88 | 13.45 |
| Other genotypes | Opportunistic | Bordeaux | 57 | 10.00 | 7.66 | 7.97 | 8.55 | 9.91 | 11.55 | 12.59 | 13.14 |
| Other genotypes | Opportunistic | Bordeaux | 58 | 9.78 | 7.53 | 7.80 | 8.36 | 9.68 | 11.31 | 12.34 | 12.89 |

**Table S9** **Posterior expected HR HPV cervical infection prevalence (in %) in major French cities, stratified by type of test, city, and age. Table counterpart to Figure 3A** ***(continued)***

| | | | | Posterior distribution | | | | | | | |
|---|---|---|---|---|---|---|---|---|---|---|---|
| | | | | | Percentiles | | | | | | |
| Virus | Test | City | Age | Average | 0.01 | 0.025 | 0.1 | 0.5 | 0.9 | 0.975 | 0.99 |
| Other genotypes | Opportunistic | Bordeaux | 59 | 9.61 | 7.38 | 7.65 | 8.21 | 9.53 | 11.10 | 12.15 | 12.59 |
| Other genotypes | Opportunistic | Bordeaux | 60 | 9.49 | 7.25 | 7.57 | 8.09 | 9.41 | 10.97 | 12.00 | 12.52 |
| Other genotypes | Opportunistic | Bordeaux | 61 | 9.42 | 7.22 | 7.51 | 8.02 | 9.33 | 10.89 | 11.87 | 12.46 |
| Other genotypes | Opportunistic | Bordeaux | 62 | 9.41 | 7.18 | 7.48 | 8.01 | 9.32 | 10.91 | 11.91 | 12.45 |
| Other genotypes | Opportunistic | Bordeaux | 63 | 9.46 | 7.22 | 7.52 | 8.04 | 9.37 | 11.00 | 11.93 | 12.52 |
| Other genotypes | Opportunistic | Bordeaux | 64 | 9.57 | 7.29 | 7.58 | 8.14 | 9.49 | 11.14 | 12.00 | 12.67 |
| Other genotypes | Opportunistic | Bordeaux | 65 | 9.73 | 7.34 | 7.68 | 8.25 | 9.65 | 11.34 | 12.29 | 12.90 |
| Other genotypes | Opportunistic | Bordeaux | 66 | 9.94 | 7.34 | 7.74 | 8.38 | 9.84 | 11.64 | 12.65 | 13.22 |
| Other genotypes | Opportunistic | Montpellier | 30 | 22.37 | 15.23 | 16.18 | 18.00 | 22.14 | 26.98 | 30.09 | 31.68 |
| Other genotypes | Opportunistic | Montpellier | 31 | 21.37 | 14.42 | 15.39 | 17.15 | 21.16 | 25.81 | 28.62 | 30.34 |
| Other genotypes | Opportunistic | Montpellier | 32 | 20.28 | 13.69 | 14.53 | 16.24 | 20.10 | 24.50 | 27.37 | 28.84 |
| Other genotypes | Opportunistic | Montpellier | 33 | 19.18 | 12.84 | 13.65 | 15.34 | 18.96 | 23.27 | 26.03 | 27.39 |
| Other genotypes | Opportunistic | Montpellier | 34 | 18.22 | 12.16 | 12.92 | 14.51 | 18.00 | 22.18 | 24.86 | 26.06 |
| Other genotypes | Opportunistic | Montpellier | 35 | 17.43 | 11.69 | 12.34 | 13.84 | 17.22 | 21.24 | 23.81 | 25.23 |
| Other genotypes | Opportunistic | Montpellier | 36 | 16.72 | 11.23 | 11.82 | 13.24 | 16.50 | 20.43 | 22.82 | 24.27 |
| Other genotypes | Opportunistic | Montpellier | 37 | 16.14 | 10.67 | 11.40 | 12.76 | 15.93 | 19.73 | 22.16 | 23.41 |
| Other genotypes | Opportunistic | Montpellier | 38 | 15.61 | 10.30 | 11.03 | 12.32 | 15.40 | 19.11 | 21.43 | 22.85 |
| Other genotypes | Opportunistic | Montpellier | 39 | 15.10 | 9.93 | 10.64 | 11.87 | 14.92 | 18.48 | 20.75 | 22.14 |
| Other genotypes | Opportunistic | Montpellier | 40 | 14.59 | 9.60 | 10.24 | 11.47 | 14.38 | 17.90 | 20.02 | 21.33 |
| Other genotypes | Opportunistic | Montpellier | 41 | 14.13 | 9.25 | 9.94 | 11.12 | 13.95 | 17.32 | 19.49 | 20.84 |
| Other genotypes | Opportunistic | Montpellier | 42 | 13.75 | 9.03 | 9.63 | 10.82 | 13.58 | 16.90 | 19.05 | 20.37 |
| Other genotypes | Opportunistic | Montpellier | 43 | 13.44 | 8.78 | 9.38 | 10.56 | 13.25 | 16.52 | 18.65 | 19.78 |
| Other genotypes | Opportunistic | Montpellier | 44 | 13.16 | 8.56 | 9.20 | 10.33 | 12.98 | 16.18 | 18.24 | 19.44 |
| Other genotypes | Opportunistic | Montpellier | 45 | 12.92 | 8.39 | 9.04 | 10.15 | 12.74 | 15.91 | 17.95 | 19.12 |
| Other genotypes | Opportunistic | Montpellier | 46 | 12.73 | 8.31 | 8.90 | 9.98 | 12.51 | 15.71 | 17.75 | 18.77 |
| Other genotypes | Opportunistic | Montpellier | 47 | 12.56 | 8.22 | 8.75 | 9.87 | 12.36 | 15.47 | 17.48 | 18.55 |
| Other genotypes | Opportunistic | Montpellier | 48 | 12.43 | 8.16 | 8.64 | 9.77 | 12.22 | 15.31 | 17.31 | 18.25 |
| Other genotypes | Opportunistic | Montpellier | 49 | 12.32 | 8.12 | 8.57 | 9.67 | 12.11 | 15.18 | 17.21 | 18.05 |
| Other genotypes | Opportunistic | Montpellier | 50 | 12.20 | 8.02 | 8.48 | 9.56 | 12.00 | 15.07 | 17.02 | 17.98 |

**Table S9** **Posterior expected HR HPV cervical infection prevalence (in %) in major French cities, stratified by type of test, city, and age. Table counterpart to Figure 3A** ***(continued)***

| | | | | Posterior distribution | | | | | | | |
|---|---|---|---|---|---|---|---|---|---|---|---|
| | | | | | Percentiles | | | | | | |
| Virus | Test | City | Age | Average | 0.01 | 0.025 | 0.1 | 0.5 | 0.9 | 0.975 | 0.99 |
| Other genotypes | Opportunistic | Montpellier | 51 | 12.07 | 7.92 | 8.37 | 9.43 | 11.88 | 14.89 | 16.82 | 17.86 |
| Other genotypes | Opportunistic | Montpellier | 52 | 11.88 | 7.69 | 8.24 | 9.27 | 11.68 | 14.67 | 16.54 | 17.64 |
| Other genotypes | Opportunistic | Montpellier | 53 | 11.67 | 7.57 | 8.09 | 9.10 | 11.49 | 14.42 | 16.25 | 17.32 |
| Other genotypes | Opportunistic | Montpellier | 54 | 11.38 | 7.39 | 7.88 | 8.90 | 11.21 | 14.11 | 15.86 | 17.07 |
| Other genotypes | Opportunistic | Montpellier | 55 | 11.12 | 7.22 | 7.70 | 8.70 | 10.96 | 13.79 | 15.57 | 16.61 |
| Other genotypes | Opportunistic | Montpellier | 56 | 10.87 | 7.10 | 7.53 | 8.47 | 10.70 | 13.47 | 15.24 | 16.27 |
| Other genotypes | Opportunistic | Montpellier | 57 | 10.61 | 6.89 | 7.37 | 8.25 | 10.45 | 13.11 | 14.87 | 15.77 |
| Other genotypes | Opportunistic | Montpellier | 58 | 10.38 | 6.75 | 7.22 | 8.07 | 10.23 | 12.85 | 14.55 | 15.52 |
| Other genotypes | Opportunistic | Montpellier | 59 | 10.20 | 6.57 | 7.06 | 7.90 | 10.04 | 12.64 | 14.35 | 15.24 |
| Other genotypes | Opportunistic | Montpellier | 60 | 10.08 | 6.53 | 6.97 | 7.81 | 9.93 | 12.47 | 14.16 | 15.07 |
| Other genotypes | Opportunistic | Montpellier | 61 | 10.00 | 6.49 | 6.90 | 7.75 | 9.87 | 12.35 | 14.10 | 14.90 |
| Other genotypes | Opportunistic | Montpellier | 62 | 9.99 | 6.47 | 6.87 | 7.75 | 9.85 | 12.35 | 14.03 | 14.92 |
| Other genotypes | Opportunistic | Montpellier | 63 | 10.04 | 6.49 | 6.91 | 7.79 | 9.90 | 12.43 | 14.15 | 15.04 |
| Other genotypes | Opportunistic | Montpellier | 64 | 10.16 | 6.53 | 7.05 | 7.88 | 10.03 | 12.61 | 14.31 | 15.24 |
| Other genotypes | Opportunistic | Montpellier | 65 | 10.33 | 6.49 | 7.16 | 7.99 | 10.20 | 12.84 | 14.53 | 15.52 |
| Other genotypes | Opportunistic | Montpellier | 66 | 10.54 | 6.69 | 7.26 | 8.15 | 10.42 | 13.13 | 14.87 | 15.98 |
| Other genotypes | Opportunistic | Rennes | 30 | 22.70 | 18.13 | 18.94 | 20.17 | 22.62 | 25.32 | 26.92 | 27.64 |
| Other genotypes | Opportunistic | Rennes | 31 | 21.68 | 17.40 | 18.02 | 19.27 | 21.59 | 24.19 | 25.69 | 26.39 |
| Other genotypes | Opportunistic | Rennes | 32 | 20.58 | 16.51 | 17.11 | 18.26 | 20.51 | 22.99 | 24.44 | 25.20 |
| Other genotypes | Opportunistic | Rennes | 33 | 19.46 | 15.55 | 16.12 | 17.24 | 19.36 | 21.73 | 23.09 | 23.90 |
| Other genotypes | Opportunistic | Rennes | 34 | 18.49 | 14.69 | 15.24 | 16.38 | 18.42 | 20.68 | 21.96 | 22.73 |
| Other genotypes | Opportunistic | Rennes | 35 | 17.69 | 14.03 | 14.59 | 15.64 | 17.62 | 19.83 | 21.08 | 21.77 |
| Other genotypes | Opportunistic | Rennes | 36 | 16.97 | 13.42 | 13.98 | 14.99 | 16.91 | 19.04 | 20.24 | 20.99 |
| Other genotypes | Opportunistic | Rennes | 37 | 16.38 | 12.90 | 13.51 | 14.47 | 16.29 | 18.38 | 19.65 | 20.33 |
| Other genotypes | Opportunistic | Rennes | 38 | 15.83 | 12.55 | 13.02 | 13.97 | 15.75 | 17.79 | 18.94 | 19.59 |
| Other genotypes | Opportunistic | Rennes | 39 | 15.32 | 12.14 | 12.62 | 13.51 | 15.25 | 17.23 | 18.39 | 18.99 |
| Other genotypes | Opportunistic | Rennes | 40 | 14.80 | 11.67 | 12.19 | 13.03 | 14.73 | 16.66 | 17.78 | 18.30 |
| Other genotypes | Opportunistic | Rennes | 41 | 14.34 | 11.30 | 11.78 | 12.59 | 14.26 | 16.14 | 17.28 | 17.75 |
| Other genotypes | Opportunistic | Rennes | 42 | 13.95 | 11.02 | 11.48 | 12.24 | 13.87 | 15.71 | 16.82 | 17.29 |

**Table S9** **Posterior expected HR HPV cervical infection prevalence (in %) in major French cities, stratified by type of test, city, and age. Table counterpart to Figure 3A** ***(continued)***

| | | | | Posterior distribution | | | | | | | |
|---|---|---|---|---|---|---|---|---|---|---|---|
| | | | | | Percentiles | | | | | | |
| Virus | Test | City | Age | Average | 0.01 | 0.025 | 0.1 | 0.5 | 0.9 | 0.975 | 0.99 |
| Other genotypes | Opportunistic | Rennes | 43 | 13.63 | 10.72 | 11.21 | 11.97 | 13.57 | 15.37 | 16.42 | 16.91 |
| Other genotypes | Opportunistic | Rennes | 44 | 13.35 | 10.51 | 10.93 | 11.71 | 13.29 | 15.08 | 16.08 | 16.60 |
| Other genotypes | Opportunistic | Rennes | 45 | 13.11 | 10.30 | 10.71 | 11.50 | 13.06 | 14.79 | 15.77 | 16.29 |
| Other genotypes | Opportunistic | Rennes | 46 | 12.91 | 10.17 | 10.51 | 11.32 | 12.86 | 14.56 | 15.52 | 16.05 |
| Other genotypes | Opportunistic | Rennes | 47 | 12.74 | 10.04 | 10.33 | 11.17 | 12.68 | 14.38 | 15.38 | 15.90 |
| Other genotypes | Opportunistic | Rennes | 48 | 12.61 | 9.87 | 10.24 | 11.06 | 12.55 | 14.22 | 15.25 | 15.73 |
| Other genotypes | Opportunistic | Rennes | 49 | 12.49 | 9.77 | 10.14 | 10.97 | 12.45 | 14.10 | 15.09 | 15.66 |
| Other genotypes | Opportunistic | Rennes | 50 | 12.37 | 9.72 | 10.03 | 10.86 | 12.32 | 13.96 | 14.94 | 15.47 |
| Other genotypes | Opportunistic | Rennes | 51 | 12.24 | 9.62 | 9.95 | 10.74 | 12.19 | 13.84 | 14.81 | 15.33 |
| Other genotypes | Opportunistic | Rennes | 52 | 12.05 | 9.51 | 9.78 | 10.57 | 12.00 | 13.63 | 14.59 | 15.07 |
| Other genotypes | Opportunistic | Rennes | 53 | 11.83 | 9.28 | 9.63 | 10.36 | 11.79 | 13.39 | 14.33 | 14.79 |
| Other genotypes | Opportunistic | Rennes | 54 | 11.55 | 9.05 | 9.39 | 10.10 | 11.49 | 13.10 | 13.97 | 14.50 |
| Other genotypes | Opportunistic | Rennes | 55 | 11.28 | 8.79 | 9.17 | 9.86 | 11.21 | 12.79 | 13.64 | 14.12 |
| Other genotypes | Opportunistic | Rennes | 56 | 11.03 | 8.63 | 8.93 | 9.63 | 10.98 | 12.50 | 13.39 | 13.79 |
| Other genotypes | Opportunistic | Rennes | 57 | 10.76 | 8.37 | 8.70 | 9.39 | 10.71 | 12.21 | 13.06 | 13.47 |
| Other genotypes | Opportunistic | Rennes | 58 | 10.53 | 8.18 | 8.51 | 9.20 | 10.48 | 11.96 | 12.79 | 13.23 |
| Other genotypes | Opportunistic | Rennes | 59 | 10.35 | 8.04 | 8.37 | 9.03 | 10.30 | 11.72 | 12.57 | 12.95 |
| Other genotypes | Opportunistic | Rennes | 60 | 10.22 | 7.90 | 8.28 | 8.91 | 10.17 | 11.57 | 12.42 | 12.82 |
| Other genotypes | Opportunistic | Rennes | 61 | 10.14 | 7.87 | 8.23 | 8.85 | 10.09 | 11.48 | 12.36 | 12.74 |
| Other genotypes | Opportunistic | Rennes | 62 | 10.13 | 7.86 | 8.17 | 8.81 | 10.07 | 11.49 | 12.31 | 12.74 |
| Other genotypes | Opportunistic | Rennes | 63 | 10.19 | 7.86 | 8.23 | 8.87 | 10.12 | 11.57 | 12.35 | 12.87 |
| Other genotypes | Opportunistic | Rennes | 64 | 10.30 | 8.03 | 8.33 | 8.97 | 10.24 | 11.72 | 12.49 | 13.00 |
| Other genotypes | Opportunistic | Rennes | 65 | 10.48 | 8.15 | 8.46 | 9.11 | 10.42 | 11.92 | 12.74 | 13.23 |
| Other genotypes | Opportunistic | Rennes | 66 | 10.69 | 8.24 | 8.58 | 9.23 | 10.62 | 12.22 | 13.08 | 13.53 |
| Other genotypes | Opportunistic | Nantes | 30 | 19.73 | 12.65 | 13.61 | 15.49 | 19.39 | 24.48 | 27.08 | 28.51 |
| Other genotypes | Opportunistic | Nantes | 31 | 18.82 | 12.09 | 12.94 | 14.72 | 18.49 | 23.44 | 25.94 | 27.46 |
| Other genotypes | Opportunistic | Nantes | 32 | 17.84 | 11.36 | 12.26 | 13.90 | 17.52 | 22.28 | 24.62 | 26.15 |
| Other genotypes | Opportunistic | Nantes | 33 | 16.84 | 10.75 | 11.49 | 13.09 | 16.53 | 21.06 | 23.44 | 25.00 |
| Other genotypes | Opportunistic | Nantes | 34 | 15.97 | 10.17 | 10.86 | 12.37 | 15.68 | 19.96 | 22.34 | 23.81 |

**Table S9 Posterior expected HR HPV cervical infection prevalence (in %) in major French cities, stratified by type of test, city, and age. Table counterpart to Figure 3A *(continued)***

| | | | | Posterior distribution | | | | | | | |
|---|---|---|---|---|---|---|---|---|---|---|---|
| | | | | | Percentiles | | | | | | |
| Virus | Test | City | Age | Average | 0.01 | 0.025 | 0.1 | 0.5 | 0.9 | 0.975 | 0.99 |
| Other genotypes | Opportunistic | Nantes | 35 | 15.27 | 9.69 | 10.37 | 11.80 | 14.98 | 19.10 | 21.39 | 22.90 |
| Other genotypes | Opportunistic | Nantes | 36 | 14.63 | 9.25 | 9.91 | 11.29 | 14.36 | 18.29 | 20.56 | 21.86 |
| Other genotypes | Opportunistic | Nantes | 37 | 14.11 | 8.87 | 9.49 | 10.89 | 13.84 | 17.75 | 19.90 | 21.24 |
| Other genotypes | Opportunistic | Nantes | 38 | 13.63 | 8.53 | 9.17 | 10.51 | 13.37 | 17.14 | 19.33 | 20.55 |
| Other genotypes | Opportunistic | Nantes | 39 | 13.18 | 8.24 | 8.86 | 10.14 | 12.92 | 16.61 | 18.72 | 19.89 |
| Other genotypes | Opportunistic | Nantes | 40 | 12.72 | 7.95 | 8.56 | 9.77 | 12.46 | 16.02 | 18.09 | 19.36 |
| Other genotypes | Opportunistic | Nantes | 41 | 12.32 | 7.69 | 8.23 | 9.44 | 12.07 | 15.53 | 17.53 | 18.75 |
| Other genotypes | Opportunistic | Nantes | 42 | 11.98 | 7.45 | 7.97 | 9.18 | 11.73 | 15.16 | 17.03 | 18.26 |
| Other genotypes | Opportunistic | Nantes | 43 | 11.70 | 7.26 | 7.81 | 8.97 | 11.44 | 14.84 | 16.64 | 17.85 |
| Other genotypes | Opportunistic | Nantes | 44 | 11.46 | 7.14 | 7.60 | 8.76 | 11.22 | 14.54 | 16.39 | 17.46 |
| Other genotypes | Opportunistic | Nantes | 45 | 11.25 | 6.99 | 7.46 | 8.57 | 10.99 | 14.28 | 16.01 | 17.16 |
| Other genotypes | Opportunistic | Nantes | 46 | 11.07 | 6.85 | 7.34 | 8.44 | 10.82 | 14.07 | 15.82 | 16.88 |
| Other genotypes | Opportunistic | Nantes | 47 | 10.92 | 6.77 | 7.24 | 8.31 | 10.68 | 13.90 | 15.61 | 16.74 |
| Other genotypes | Opportunistic | Nantes | 48 | 10.81 | 6.65 | 7.17 | 8.22 | 10.55 | 13.75 | 15.49 | 16.55 |
| Other genotypes | Opportunistic | Nantes | 49 | 10.71 | 6.61 | 7.10 | 8.16 | 10.45 | 13.63 | 15.35 | 16.42 |
| Other genotypes | Opportunistic | Nantes | 50 | 10.60 | 6.52 | 7.02 | 8.07 | 10.38 | 13.48 | 15.24 | 16.28 |
| Other genotypes | Opportunistic | Nantes | 51 | 10.49 | 6.45 | 6.96 | 7.98 | 10.28 | 13.34 | 15.06 | 16.14 |
| Other genotypes | Opportunistic | Nantes | 52 | 10.32 | 6.40 | 6.86 | 7.85 | 10.08 | 13.13 | 14.78 | 15.88 |
| Other genotypes | Opportunistic | Nantes | 53 | 10.13 | 6.26 | 6.74 | 7.70 | 9.91 | 12.90 | 14.52 | 15.47 |
| Other genotypes | Opportunistic | Nantes | 54 | 9.88 | 6.06 | 6.54 | 7.51 | 9.67 | 12.60 | 14.15 | 15.09 |
| Other genotypes | Opportunistic | Nantes | 55 | 9.65 | 5.95 | 6.38 | 7.32 | 9.44 | 12.27 | 13.86 | 14.80 |
| Other genotypes | Opportunistic | Nantes | 56 | 9.43 | 5.82 | 6.24 | 7.14 | 9.22 | 12.01 | 13.57 | 14.50 |
| Other genotypes | Opportunistic | Nantes | 57 | 9.20 | 5.65 | 6.06 | 6.98 | 8.99 | 11.73 | 13.21 | 14.20 |
| Other genotypes | Opportunistic | Nantes | 58 | 9.00 | 5.53 | 5.96 | 6.83 | 8.79 | 11.48 | 12.95 | 13.97 |
| Other genotypes | Opportunistic | Nantes | 59 | 8.84 | 5.44 | 5.83 | 6.69 | 8.63 | 11.28 | 12.75 | 13.66 |
| Other genotypes | Opportunistic | Nantes | 60 | 8.73 | 5.37 | 5.75 | 6.64 | 8.53 | 11.15 | 12.57 | 13.55 |
| Other genotypes | Opportunistic | Nantes | 61 | 8.66 | 5.34 | 5.70 | 6.58 | 8.44 | 11.05 | 12.46 | 13.45 |
| Other genotypes | Opportunistic | Nantes | 62 | 8.65 | 5.33 | 5.67 | 6.57 | 8.44 | 11.04 | 12.48 | 13.38 |
| Other genotypes | Opportunistic | Nantes | 63 | 8.70 | 5.34 | 5.70 | 6.60 | 8.47 | 11.11 | 12.58 | 13.52 |

**Table S9 Posterior expected HR HPV cervical infection prevalence (in %) in major French cities, stratified by type of test, city, and age. Table counterpart to Figure 3A *(continued)***

| | | | | Posterior distribution | | | | | | | |
|---|---|---|---|---|---|---|---|---|---|---|---|
| | | | | | Percentiles | | | | | | |
| Virus | Test | City | Age | Average | 0.01 | 0.025 | 0.1 | 0.5 | 0.9 | 0.975 | 0.99 |
| Other genotypes | Opportunistic | Nantes | 64 | 8.80 | 5.42 | 5.80 | 6.66 | 8.57 | 11.24 | 12.73 | 13.61 |
| Other genotypes | Opportunistic | Nantes | 65 | 8.95 | 5.48 | 5.90 | 6.79 | 8.74 | 11.43 | 13.01 | 13.88 |
| Other genotypes | Opportunistic | Nantes | 66 | 9.14 | 5.57 | 6.03 | 6.91 | 8.91 | 11.67 | 13.35 | 14.20 |
| Other genotypes | Opportunistic | Lille | 30 | 20.59 | 14.09 | 15.02 | 16.77 | 20.40 | 24.67 | 27.11 | 28.52 |
| Other genotypes | Opportunistic | Lille | 31 | 19.65 | 13.34 | 14.32 | 15.99 | 19.43 | 23.61 | 25.99 | 27.21 |
| Other genotypes | Opportunistic | Lille | 32 | 18.63 | 12.61 | 13.51 | 15.09 | 18.45 | 22.46 | 24.66 | 25.96 |
| Other genotypes | Opportunistic | Lille | 33 | 17.59 | 11.80 | 12.74 | 14.20 | 17.37 | 21.28 | 23.33 | 24.70 |
| Other genotypes | Opportunistic | Lille | 34 | 16.69 | 11.28 | 12.06 | 13.46 | 16.49 | 20.24 | 22.20 | 23.62 |
| Other genotypes | Opportunistic | Lille | 35 | 15.96 | 10.77 | 11.52 | 12.82 | 15.77 | 19.40 | 21.21 | 22.53 |
| Other genotypes | Opportunistic | Lille | 36 | 15.30 | 10.20 | 11.01 | 12.28 | 15.11 | 18.62 | 20.42 | 21.79 |
| Other genotypes | Opportunistic | Lille | 37 | 14.76 | 9.79 | 10.62 | 11.85 | 14.60 | 17.96 | 19.79 | 21.02 |
| Other genotypes | Opportunistic | Lille | 38 | 14.26 | 9.52 | 10.23 | 11.41 | 14.09 | 17.39 | 19.09 | 20.33 |
| Other genotypes | Opportunistic | Lille | 39 | 13.79 | 9.10 | 9.87 | 11.05 | 13.64 | 16.81 | 18.53 | 19.76 |
| Other genotypes | Opportunistic | Lille | 40 | 13.32 | 8.82 | 9.51 | 10.66 | 13.13 | 16.26 | 17.97 | 19.15 |
| Other genotypes | Opportunistic | Lille | 41 | 12.89 | 8.60 | 9.22 | 10.28 | 12.72 | 15.78 | 17.30 | 18.60 |
| Other genotypes | Opportunistic | Lille | 42 | 12.54 | 8.31 | 8.95 | 9.98 | 12.38 | 15.37 | 16.83 | 18.14 |
| Other genotypes | Opportunistic | Lille | 43 | 12.25 | 8.11 | 8.73 | 9.73 | 12.08 | 15.02 | 16.46 | 17.66 |
| Other genotypes | Opportunistic | Lille | 44 | 12.00 | 7.94 | 8.53 | 9.55 | 11.83 | 14.74 | 16.16 | 17.30 |
| Other genotypes | Opportunistic | Lille | 45 | 11.78 | 7.76 | 8.33 | 9.37 | 11.60 | 14.50 | 15.85 | 16.94 |
| Other genotypes | Opportunistic | Lille | 46 | 11.59 | 7.65 | 8.19 | 9.23 | 11.42 | 14.21 | 15.62 | 16.75 |
| Other genotypes | Opportunistic | Lille | 47 | 11.44 | 7.54 | 8.09 | 9.11 | 11.27 | 14.03 | 15.46 | 16.50 |
| Other genotypes | Opportunistic | Lille | 48 | 11.32 | 7.46 | 7.98 | 9.00 | 11.16 | 13.87 | 15.30 | 16.38 |
| Other genotypes | Opportunistic | Lille | 49 | 11.21 | 7.37 | 7.92 | 8.93 | 11.05 | 13.77 | 15.12 | 16.28 |
| Other genotypes | Opportunistic | Lille | 50 | 11.11 | 7.33 | 7.88 | 8.84 | 10.96 | 13.65 | 15.02 | 16.17 |
| Other genotypes | Opportunistic | Lille | 51 | 10.98 | 7.29 | 7.74 | 8.74 | 10.84 | 13.50 | 14.89 | 16.04 |
| Other genotypes | Opportunistic | Lille | 52 | 10.81 | 7.14 | 7.62 | 8.60 | 10.65 | 13.29 | 14.65 | 15.83 |
| Other genotypes | Opportunistic | Lille | 53 | 10.61 | 6.98 | 7.49 | 8.42 | 10.47 | 13.03 | 14.41 | 15.63 |
| Other genotypes | Opportunistic | Lille | 54 | 10.36 | 6.80 | 7.28 | 8.20 | 10.21 | 12.73 | 14.11 | 15.23 |
| Other genotypes | Opportunistic | Lille | 55 | 10.11 | 6.62 | 7.15 | 7.98 | 9.97 | 12.39 | 13.75 | 14.84 |

**Table S9 Posterior expected HR HPV cervical infection prevalence (in %) in major French cities, stratified by type of test, city, and age. Table counterpart to Figure 3A** ***(continued)***

| | | | | Posterior distribution | | | | | | | |
|---|---|---|---|---|---|---|---|---|---|---|---|
| | | | | | Percentiles | | | | | | |
| Virus | Test | City | Age | Average | 0.01 | 0.025 | 0.1 | 0.5 | 0.9 | 0.975 | 0.99 |
| Other genotypes | Opportunistic | Lille | 56 | 9.88 | 6.45 | 6.99 | 7.83 | 9.74 | 12.12 | 13.50 | 14.56 |
| Other genotypes | Opportunistic | Lille | 57 | 9.64 | 6.32 | 6.80 | 7.65 | 9.52 | 11.85 | 13.17 | 14.13 |
| Other genotypes | Opportunistic | Lille | 58 | 9.43 | 6.14 | 6.67 | 7.46 | 9.31 | 11.61 | 12.79 | 13.79 |
| Other genotypes | Opportunistic | Lille | 59 | 9.27 | 6.05 | 6.55 | 7.32 | 9.13 | 11.40 | 12.63 | 13.49 |
| Other genotypes | Opportunistic | Lille | 60 | 9.15 | 5.92 | 6.45 | 7.21 | 9.01 | 11.30 | 12.52 | 13.42 |
| Other genotypes | Opportunistic | Lille | 61 | 9.08 | 5.91 | 6.39 | 7.15 | 8.94 | 11.19 | 12.41 | 13.35 |
| Other genotypes | Opportunistic | Lille | 62 | 9.07 | 5.92 | 6.36 | 7.16 | 8.93 | 11.18 | 12.45 | 13.45 |
| Other genotypes | Opportunistic | Lille | 63 | 9.12 | 5.97 | 6.41 | 7.20 | 9.00 | 11.23 | 12.52 | 13.45 |
| Other genotypes | Opportunistic | Lille | 64 | 9.23 | 6.02 | 6.41 | 7.29 | 9.11 | 11.37 | 12.68 | 13.66 |
| Other genotypes | Opportunistic | Lille | 65 | 9.39 | 6.12 | 6.57 | 7.40 | 9.27 | 11.61 | 12.91 | 13.92 |
| Other genotypes | Opportunistic | Lille | 66 | 9.59 | 6.22 | 6.59 | 7.53 | 9.44 | 11.90 | 13.22 | 14.34 |
| Other genotypes | Opportunistic | Strasbourg | 30 | 17.83 | 10.83 | 11.58 | 13.50 | 17.53 | 22.51 | 25.58 | 27.45 |
| Other genotypes | Opportunistic | Strasbourg | 31 | 16.99 | 10.32 | 10.97 | 12.81 | 16.73 | 21.46 | 24.41 | 26.22 |
| Other genotypes | Opportunistic | Strasbourg | 32 | 16.09 | 9.72 | 10.34 | 12.07 | 15.82 | 20.40 | 23.31 | 24.97 |
| Other genotypes | Opportunistic | Strasbourg | 33 | 15.17 | 9.07 | 9.69 | 11.31 | 14.91 | 19.29 | 22.11 | 23.76 |
| Other genotypes | Opportunistic | Strasbourg | 34 | 14.37 | 8.54 | 9.14 | 10.72 | 14.11 | 18.31 | 21.09 | 22.61 |
| Other genotypes | Opportunistic | Strasbourg | 35 | 13.73 | 8.15 | 8.68 | 10.25 | 13.49 | 17.50 | 20.22 | 21.62 |
| Other genotypes | Opportunistic | Strasbourg | 36 | 13.14 | 7.76 | 8.31 | 9.82 | 12.90 | 16.79 | 19.42 | 20.76 |
| Other genotypes | Opportunistic | Strasbourg | 37 | 12.67 | 7.49 | 8.01 | 9.43 | 12.43 | 16.21 | 18.68 | 20.10 |
| Other genotypes | Opportunistic | Strasbourg | 38 | 12.23 | 7.22 | 7.73 | 9.07 | 12.01 | 15.66 | 18.19 | 19.39 |
| Other genotypes | Opportunistic | Strasbourg | 39 | 11.82 | 6.96 | 7.41 | 8.77 | 11.60 | 15.14 | 17.57 | 18.83 |
| Other genotypes | Opportunistic | Strasbourg | 40 | 11.40 | 6.78 | 7.12 | 8.43 | 11.17 | 14.66 | 16.93 | 18.26 |
| Other genotypes | Opportunistic | Strasbourg | 41 | 11.03 | 6.54 | 6.90 | 8.15 | 10.80 | 14.20 | 16.41 | 17.73 |
| Other genotypes | Opportunistic | Strasbourg | 42 | 10.73 | 6.33 | 6.70 | 7.92 | 10.50 | 13.77 | 15.98 | 17.38 |
| Other genotypes | Opportunistic | Strasbourg | 43 | 10.48 | 6.18 | 6.55 | 7.72 | 10.25 | 13.47 | 15.61 | 17.04 |
| Other genotypes | Opportunistic | Strasbourg | 44 | 10.26 | 6.00 | 6.42 | 7.53 | 10.04 | 13.22 | 15.31 | 16.74 |
| Other genotypes | Opportunistic | Strasbourg | 45 | 10.06 | 5.90 | 6.32 | 7.38 | 9.85 | 13.00 | 15.07 | 16.35 |
| Other genotypes | Opportunistic | Strasbourg | 46 | 9.91 | 5.76 | 6.22 | 7.27 | 9.68 | 12.79 | 14.85 | 16.07 |
| Other genotypes | Opportunistic | Strasbourg | 47 | 9.77 | 5.68 | 6.12 | 7.16 | 9.55 | 12.62 | 14.61 | 15.82 |

**Table S9 Posterior expected HR HPV cervical infection prevalence (in %) in major French cities, stratified by type of test, city, and age. Table counterpart to Figure 3A** ***(continued)***

| | | | | Posterior distribution | | | | | | | |
|---|---|---|---|---|---|---|---|---|---|---|---|
| | | | | | Percentiles | | | | | | |
| Virus | Test | City | Age | Average | 0.01 | 0.025 | 0.1 | 0.5 | 0.9 | 0.975 | 0.99 |
| Other genotypes | Opportunistic | Strasbourg | 48 | 9.67 | 5.59 | 6.06 | 7.10 | 9.44 | 12.47 | 14.43 | 15.62 |
| Other genotypes | Opportunistic | Strasbourg | 49 | 9.58 | 5.52 | 5.97 | 7.02 | 9.36 | 12.35 | 14.39 | 15.49 |
| Other genotypes | Opportunistic | Strasbourg | 50 | 9.48 | 5.48 | 5.92 | 6.93 | 9.27 | 12.22 | 14.22 | 15.33 |
| Other genotypes | Opportunistic | Strasbourg | 51 | 9.38 | 5.42 | 5.85 | 6.86 | 9.18 | 12.09 | 14.06 | 15.20 |
| Other genotypes | Opportunistic | Strasbourg | 52 | 9.22 | 5.36 | 5.76 | 6.76 | 9.02 | 11.91 | 13.82 | 14.97 |
| Other genotypes | Opportunistic | Strasbourg | 53 | 9.05 | 5.29 | 5.64 | 6.64 | 8.83 | 11.69 | 13.56 | 14.74 |
| Other genotypes | Opportunistic | Strasbourg | 54 | 8.83 | 5.16 | 5.49 | 6.47 | 8.63 | 11.43 | 13.21 | 14.43 |
| Other genotypes | Opportunistic | Strasbourg | 55 | 8.62 | 4.99 | 5.33 | 6.29 | 8.40 | 11.16 | 12.93 | 14.08 |
| Other genotypes | Opportunistic | Strasbourg | 56 | 8.42 | 4.88 | 5.22 | 6.16 | 8.22 | 10.94 | 12.67 | 13.87 |
| Other genotypes | Opportunistic | Strasbourg | 57 | 8.21 | 4.75 | 5.06 | 5.99 | 8.02 | 10.64 | 12.36 | 13.48 |
| Other genotypes | Opportunistic | Strasbourg | 58 | 8.03 | 4.60 | 4.96 | 5.86 | 7.84 | 10.41 | 12.10 | 13.23 |
| Other genotypes | Opportunistic | Strasbourg | 59 | 7.89 | 4.50 | 4.88 | 5.77 | 7.70 | 10.20 | 11.90 | 12.83 |
| Other genotypes | Opportunistic | Strasbourg | 60 | 7.79 | 4.46 | 4.83 | 5.70 | 7.59 | 10.08 | 11.80 | 12.78 |
| Other genotypes | Opportunistic | Strasbourg | 61 | 7.73 | 4.44 | 4.77 | 5.67 | 7.52 | 10.00 | 11.69 | 12.66 |
| Other genotypes | Opportunistic | Strasbourg | 62 | 7.72 | 4.41 | 4.76 | 5.66 | 7.52 | 10.00 | 11.68 | 12.60 |
| Other genotypes | Opportunistic | Strasbourg | 63 | 7.77 | 4.45 | 4.80 | 5.68 | 7.57 | 10.07 | 11.74 | 12.81 |
| Other genotypes | Opportunistic | Strasbourg | 64 | 7.86 | 4.55 | 4.86 | 5.74 | 7.67 | 10.17 | 11.98 | 13.03 |
| Other genotypes | Opportunistic | Strasbourg | 65 | 7.99 | 4.65 | 4.95 | 5.81 | 7.82 | 10.37 | 12.23 | 13.25 |
| Other genotypes | Opportunistic | Strasbourg | 66 | 8.16 | 4.71 | 5.03 | 5.92 | 7.99 | 10.57 | 12.47 | 13.56 |
| Other genotypes | Opportunistic | Lyon | 30 | 22.91 | 15.05 | 16.25 | 18.11 | 22.61 | 27.79 | 31.20 | 32.62 |
| Other genotypes | Opportunistic | Lyon | 31 | 21.90 | 14.40 | 15.49 | 17.24 | 21.60 | 26.70 | 29.94 | 31.34 |
| Other genotypes | Opportunistic | Lyon | 32 | 20.79 | 13.68 | 14.61 | 16.33 | 20.51 | 25.43 | 28.67 | 29.83 |
| Other genotypes | Opportunistic | Lyon | 33 | 19.67 | 12.79 | 13.78 | 15.39 | 19.40 | 24.14 | 27.19 | 28.43 |
| Other genotypes | Opportunistic | Lyon | 34 | 18.69 | 12.00 | 13.04 | 14.61 | 18.43 | 22.98 | 25.97 | 27.19 |
| Other genotypes | Opportunistic | Lyon | 35 | 17.89 | 11.50 | 12.40 | 13.93 | 17.66 | 22.03 | 25.02 | 26.20 |
| Other genotypes | Opportunistic | Lyon | 36 | 17.16 | 10.96 | 11.86 | 13.34 | 16.90 | 21.18 | 24.03 | 25.25 |
| Other genotypes | Opportunistic | Lyon | 37 | 16.58 | 10.56 | 11.41 | 12.86 | 16.29 | 20.51 | 23.26 | 24.35 |
| Other genotypes | Opportunistic | Lyon | 38 | 16.03 | 10.20 | 11.03 | 12.39 | 15.77 | 19.85 | 22.51 | 23.74 |
| Other genotypes | Opportunistic | Lyon | 39 | 15.51 | 9.83 | 10.66 | 11.97 | 15.26 | 19.23 | 21.77 | 23.01 |

**Table S9 Posterior expected HR HPV cervical infection prevalence (in %) in major French cities, stratified by type of test, city, and age. Table counterpart to Figure 3A** ***(continued)***

| | | | | Posterior distribution | | | | | | | |
|---|---|---|---|---|---|---|---|---|---|---|---|
| | | | | | Percentiles | | | | | | |
| Virus | Test | City | Age | Average | 0.01 | 0.025 | 0.1 | 0.5 | 0.9 | 0.975 | 0.99 |
| Other genotypes | Opportunistic | Lyon | 40 | 14.99 | 9.43 | 10.30 | 11.55 | 14.74 | 18.63 | 21.11 | 22.27 |
| Other genotypes | Opportunistic | Lyon | 41 | 14.52 | 9.16 | 9.97 | 11.18 | 14.26 | 18.07 | 20.53 | 21.55 |
| Other genotypes | Opportunistic | Lyon | 42 | 14.14 | 8.89 | 9.69 | 10.88 | 13.87 | 17.61 | 20.09 | 21.04 |
| Other genotypes | Opportunistic | Lyon | 43 | 13.81 | 8.70 | 9.43 | 10.63 | 13.55 | 17.21 | 19.62 | 20.66 |
| Other genotypes | Opportunistic | Lyon | 44 | 13.53 | 8.49 | 9.21 | 10.41 | 13.27 | 16.82 | 19.25 | 20.21 |
| Other genotypes | Opportunistic | Lyon | 45 | 13.29 | 8.31 | 9.04 | 10.19 | 13.01 | 16.55 | 18.89 | 19.88 |
| Other genotypes | Opportunistic | Lyon | 46 | 13.09 | 8.13 | 8.90 | 10.06 | 12.84 | 16.34 | 18.60 | 19.59 |
| Other genotypes | Opportunistic | Lyon | 47 | 12.91 | 8.06 | 8.77 | 9.92 | 12.66 | 16.12 | 18.36 | 19.39 |
| Other genotypes | Opportunistic | Lyon | 48 | 12.78 | 8.02 | 8.71 | 9.81 | 12.54 | 15.98 | 18.25 | 19.26 |
| Other genotypes | Opportunistic | Lyon | 49 | 12.66 | 7.89 | 8.64 | 9.70 | 12.46 | 15.82 | 18.10 | 19.08 |
| Other genotypes | Opportunistic | Lyon | 50 | 12.54 | 7.84 | 8.51 | 9.63 | 12.33 | 15.63 | 17.93 | 18.91 |
| Other genotypes | Opportunistic | Lyon | 51 | 12.41 | 7.77 | 8.45 | 9.52 | 12.20 | 15.46 | 17.76 | 18.78 |
| Other genotypes | Opportunistic | Lyon | 52 | 12.22 | 7.64 | 8.30 | 9.37 | 11.99 | 15.23 | 17.48 | 18.38 |
| Other genotypes | Opportunistic | Lyon | 53 | 12.00 | 7.45 | 8.15 | 9.18 | 11.77 | 14.99 | 17.26 | 18.11 |
| Other genotypes | Opportunistic | Lyon | 54 | 11.71 | 7.26 | 7.94 | 8.98 | 11.49 | 14.67 | 16.88 | 17.69 |
| Other genotypes | Opportunistic | Lyon | 55 | 11.44 | 7.08 | 7.73 | 8.73 | 11.21 | 14.32 | 16.50 | 17.32 |
| Other genotypes | Opportunistic | Lyon | 56 | 11.19 | 6.87 | 7.56 | 8.54 | 10.97 | 14.05 | 16.05 | 16.96 |
| Other genotypes | Opportunistic | Lyon | 57 | 10.92 | 6.71 | 7.36 | 8.32 | 10.68 | 13.68 | 15.68 | 16.62 |
| Other genotypes | Opportunistic | Lyon | 58 | 10.69 | 6.58 | 7.20 | 8.15 | 10.46 | 13.43 | 15.32 | 16.22 |
| Other genotypes | Opportunistic | Lyon | 59 | 10.50 | 6.48 | 7.08 | 8.01 | 10.28 | 13.20 | 15.08 | 15.92 |
| Other genotypes | Opportunistic | Lyon | 60 | 10.37 | 6.43 | 6.99 | 7.89 | 10.16 | 13.03 | 14.97 | 15.79 |
| Other genotypes | Opportunistic | Lyon | 61 | 10.29 | 6.38 | 6.92 | 7.84 | 10.10 | 12.93 | 14.93 | 15.63 |
| Other genotypes | Opportunistic | Lyon | 62 | 10.28 | 6.39 | 6.92 | 7.83 | 10.07 | 12.95 | 14.92 | 15.61 |
| Other genotypes | Opportunistic | Lyon | 63 | 10.34 | 6.39 | 6.97 | 7.85 | 10.13 | 13.04 | 14.99 | 15.68 |
| Other genotypes | Opportunistic | Lyon | 64 | 10.46 | 6.51 | 7.04 | 7.96 | 10.26 | 13.21 | 15.20 | 15.81 |
| Other genotypes | Opportunistic | Lyon | 65 | 10.63 | 6.61 | 7.16 | 8.09 | 10.42 | 13.44 | 15.32 | 16.28 |
| Other genotypes | Opportunistic | Lyon | 66 | 10.85 | 6.74 | 7.26 | 8.26 | 10.60 | 13.75 | 15.65 | 16.72 |
| Other genotypes | Opportunistic | Paris | 30 | 25.77 | 20.69 | 21.45 | 22.85 | 25.71 | 28.69 | 30.37 | 31.27 |
| Other genotypes | Opportunistic | Paris | 31 | 24.66 | 19.79 | 20.46 | 21.87 | 24.59 | 27.51 | 29.18 | 29.91 |

**Table S9 Posterior expected HR HPV cervical infection prevalence (in %) in major French cities, stratified by type of test, city, and age. Table counterpart to Figure 3A** ***(continued)***

| | | | | Posterior distribution | | | | | | | |
|---|---|---|---|---|---|---|---|---|---|---|---|
| | | | | | Percentiles | | | | | | |
| Virus | Test | City | Age | Average | 0.01 | 0.025 | 0.1 | 0.5 | 0.9 | 0.975 | 0.99 |
| Other genotypes | Opportunistic | Paris | 32 | 23.46 | 18.76 | 19.40 | 20.80 | 23.40 | 26.20 | 27.80 | 28.61 |
| Other genotypes | Opportunistic | Paris | 33 | 22.22 | 17.66 | 18.35 | 19.67 | 22.13 | 24.87 | 26.38 | 27.13 |
| Other genotypes | Opportunistic | Paris | 34 | 21.14 | 16.76 | 17.37 | 18.66 | 21.07 | 23.63 | 25.22 | 25.90 |
| Other genotypes | Opportunistic | Paris | 35 | 20.26 | 16.03 | 16.65 | 17.85 | 20.18 | 22.73 | 24.14 | 24.99 |
| Other genotypes | Opportunistic | Paris | 36 | 19.46 | 15.30 | 15.93 | 17.12 | 19.38 | 21.88 | 23.24 | 24.07 |
| Other genotypes | Opportunistic | Paris | 37 | 18.81 | 14.87 | 15.37 | 16.51 | 18.75 | 21.14 | 22.57 | 23.30 |
| Other genotypes | Opportunistic | Paris | 38 | 18.20 | 14.36 | 14.90 | 15.99 | 18.15 | 20.46 | 21.91 | 22.63 |
| Other genotypes | Opportunistic | Paris | 39 | 17.63 | 13.90 | 14.42 | 15.46 | 17.58 | 19.84 | 21.19 | 21.88 |
| Other genotypes | Opportunistic | Paris | 40 | 17.04 | 13.43 | 13.94 | 14.95 | 17.00 | 19.19 | 20.48 | 21.23 |
| Other genotypes | Opportunistic | Paris | 41 | 16.52 | 13.01 | 13.47 | 14.47 | 16.47 | 18.65 | 19.88 | 20.54 |
| Other genotypes | Opportunistic | Paris | 42 | 16.09 | 12.59 | 13.09 | 14.11 | 16.04 | 18.15 | 19.34 | 20.01 |
| Other genotypes | Opportunistic | Paris | 43 | 15.73 | 12.40 | 12.79 | 13.78 | 15.69 | 17.79 | 18.99 | 19.49 |
| Other genotypes | Opportunistic | Paris | 44 | 15.42 | 12.01 | 12.48 | 13.49 | 15.34 | 17.45 | 18.64 | 19.23 |
| Other genotypes | Opportunistic | Paris | 45 | 15.14 | 11.84 | 12.31 | 13.25 | 15.06 | 17.14 | 18.37 | 18.90 |
| Other genotypes | Opportunistic | Paris | 46 | 14.92 | 11.59 | 12.10 | 13.03 | 14.83 | 16.89 | 18.06 | 18.72 |
| Other genotypes | Opportunistic | Paris | 47 | 14.72 | 11.49 | 11.94 | 12.86 | 14.64 | 16.70 | 17.83 | 18.52 |
| Other genotypes | Opportunistic | Paris | 48 | 14.58 | 11.39 | 11.81 | 12.75 | 14.50 | 16.50 | 17.63 | 18.29 |
| Other genotypes | Opportunistic | Paris | 49 | 14.45 | 11.33 | 11.70 | 12.62 | 14.37 | 16.35 | 17.46 | 18.11 |
| Other genotypes | Opportunistic | Paris | 50 | 14.31 | 11.25 | 11.57 | 12.49 | 14.25 | 16.21 | 17.28 | 17.97 |
| Other genotypes | Opportunistic | Paris | 51 | 14.16 | 11.06 | 11.47 | 12.35 | 14.10 | 16.05 | 17.08 | 17.75 |
| Other genotypes | Opportunistic | Paris | 52 | 13.94 | 10.88 | 11.30 | 12.15 | 13.89 | 15.80 | 16.83 | 17.46 |
| Other genotypes | Opportunistic | Paris | 53 | 13.70 | 10.66 | 11.05 | 11.91 | 13.63 | 15.50 | 16.65 | 17.14 |
| Other genotypes | Opportunistic | Paris | 54 | 13.38 | 10.37 | 10.76 | 11.63 | 13.31 | 15.16 | 16.23 | 16.81 |
| Other genotypes | Opportunistic | Paris | 55 | 13.07 | 10.10 | 10.51 | 11.35 | 13.00 | 14.84 | 15.91 | 16.44 |
| Other genotypes | Opportunistic | Paris | 56 | 12.78 | 9.90 | 10.25 | 11.09 | 12.72 | 14.54 | 15.51 | 16.13 |
| Other genotypes | Opportunistic | Paris | 57 | 12.48 | 9.65 | 10.01 | 10.85 | 12.42 | 14.17 | 15.21 | 15.67 |
| Other genotypes | Opportunistic | Paris | 58 | 12.22 | 9.46 | 9.78 | 10.63 | 12.15 | 13.89 | 14.89 | 15.45 |
| Other genotypes | Opportunistic | Paris | 59 | 12.01 | 9.29 | 9.65 | 10.41 | 11.94 | 13.67 | 14.72 | 15.13 |
| Other genotypes | Opportunistic | Paris | 60 | 11.86 | 9.21 | 9.54 | 10.29 | 11.80 | 13.45 | 14.55 | 14.97 |

**Table S9 Posterior expected HR HPV cervical infection prevalence (in %) in major French cities, stratified by type of test, city, and age. Table counterpart to Figure 3A** ***(continued)***

| | | | | Posterior distribution | | | | | | | |
|---|---|---|---|---|---|---|---|---|---|---|---|
| | | | | | Percentiles | | | | | | |
| Virus | Test | City | Age | Average | 0.01 | 0.025 | 0.1 | 0.5 | 0.9 | 0.975 | 0.99 |
| Other genotypes | Opportunistic | Paris | 61 | 11.78 | 9.16 | 9.51 | 10.21 | 11.71 | 13.37 | 14.43 | 14.90 |
| Other genotypes | Opportunistic | Paris | 62 | 11.76 | 9.12 | 9.45 | 10.20 | 11.70 | 13.38 | 14.41 | 14.89 |
| Other genotypes | Opportunistic | Paris | 63 | 11.83 | 9.15 | 9.50 | 10.25 | 11.77 | 13.47 | 14.44 | 15.07 |
| Other genotypes | Opportunistic | Paris | 64 | 11.96 | 9.25 | 9.58 | 10.36 | 11.90 | 13.61 | 14.56 | 15.27 |
| Other genotypes | Opportunistic | Paris | 65 | 12.16 | 9.32 | 9.70 | 10.52 | 12.11 | 13.86 | 14.81 | 15.58 |
| Other genotypes | Opportunistic | Paris | 66 | 12.41 | 9.41 | 9.84 | 10.66 | 12.35 | 14.22 | 15.26 | 15.95 |

**Table S10 Posterior difference in expected HR HPV cervical infection prevalence (in percentage points) in major French cities, between opportunistic and organised screening, stratified by type of test, city, and age. Table counterpart to Figure 3B.**

| | | | Posterior distribution | | | | | | | |
|---|---|---|---|---|---|---|---|---|---|---|
| | | | | Percentiles | | | | | | |
| Virus | City | Age | Average | 0.01 | 0.025 | 0.1 | 0.5 | 0.9 | 0.975 | 0.99 |
| HPV16/18 | Bordeaux | 30 | 1.69 | -0.16 | 0.17 | 0.70 | 1.69 | 2.65 | 3.22 | 3.55 |
| HPV16/18 | Bordeaux | 31 | 1.82 | -0.09 | 0.25 | 0.83 | 1.81 | 2.81 | 3.41 | 3.77 |
| HPV16/18 | Bordeaux | 32 | 1.94 | -0.04 | 0.35 | 0.92 | 1.92 | 2.95 | 3.56 | 3.93 |
| HPV16/18 | Bordeaux | 33 | 2.02 | 0.00 | 0.39 | 1.00 | 2.02 | 3.07 | 3.71 | 4.06 |
| HPV16/18 | Bordeaux | 34 | 2.07 | 0.04 | 0.44 | 1.04 | 2.06 | 3.11 | 3.74 | 4.11 |
| HPV16/18 | Bordeaux | 35 | 2.06 | 0.09 | 0.48 | 1.04 | 2.04 | 3.08 | 3.69 | 4.08 |
| HPV16/18 | Bordeaux | 36 | 2.02 | 0.12 | 0.49 | 1.03 | 2.01 | 3.02 | 3.60 | 4.05 |
| HPV16/18 | Bordeaux | 37 | 1.96 | 0.13 | 0.51 | 1.00 | 1.96 | 2.95 | 3.52 | 3.93 |
| HPV16/18 | Bordeaux | 38 | 1.91 | 0.07 | 0.49 | 0.97 | 1.91 | 2.85 | 3.44 | 3.81 |
| HPV16/18 | Bordeaux | 39 | 1.86 | 0.07 | 0.45 | 0.93 | 1.85 | 2.78 | 3.32 | 3.70 |
| HPV16/18 | Bordeaux | 40 | 1.80 | 0.08 | 0.42 | 0.91 | 1.79 | 2.71 | 3.24 | 3.58 |
| HPV16/18 | Bordeaux | 41 | 1.74 | 0.09 | 0.39 | 0.88 | 1.74 | 2.62 | 3.18 | 3.44 |
| HPV16/18 | Bordeaux | 42 | 1.68 | 0.08 | 0.37 | 0.84 | 1.68 | 2.54 | 3.07 | 3.35 |
| HPV16/18 | Bordeaux | 43 | 1.63 | 0.08 | 0.35 | 0.82 | 1.63 | 2.45 | 2.96 | 3.26 |

**Table S10 Posterior difference in expected HR HPV cervical infection prevalence (in percentage points) in major French cities, between opportunistic and organised screening, stratified by type of test, city, and age. Table counterpart to Figure 3B.** ***(continued)***

| | | | Posterior distribution | | | | | | | |
|---|---|---|---|---|---|---|---|---|---|---|
| | | | | Percentiles | | | | | | |
| Virus | City | Age | Average | 0.01 | 0.025 | 0.1 | 0.5 | 0.9 | 0.975 | 0.99 |
| HPV16/18 | Bordeaux | 44 | 1.58 | 0.05 | 0.32 | 0.78 | 1.58 | 2.38 | 2.88 | 3.15 |
| HPV16/18 | Bordeaux | 45 | 1.54 | 0.06 | 0.30 | 0.75 | 1.54 | 2.33 | 2.81 | 3.09 |
| HPV16/18 | Bordeaux | 46 | 1.49 | 0.04 | 0.27 | 0.72 | 1.49 | 2.27 | 2.74 | 3.02 |
| HPV16/18 | Bordeaux | 47 | 1.45 | 0.02 | 0.25 | 0.69 | 1.44 | 2.21 | 2.68 | 2.94 |
| HPV16/18 | Bordeaux | 48 | 1.41 | -0.01 | 0.22 | 0.67 | 1.40 | 2.15 | 2.62 | 2.85 |
| HPV16/18 | Bordeaux | 49 | 1.36 | -0.03 | 0.19 | 0.65 | 1.36 | 2.10 | 2.56 | 2.79 |
| HPV16/18 | Bordeaux | 50 | 1.32 | -0.03 | 0.18 | 0.62 | 1.33 | 2.05 | 2.49 | 2.75 |
| HPV16/18 | Bordeaux | 51 | 1.28 | -0.05 | 0.14 | 0.59 | 1.28 | 1.99 | 2.44 | 2.68 |
| HPV16/18 | Bordeaux | 52 | 1.24 | -0.09 | 0.11 | 0.56 | 1.24 | 1.94 | 2.36 | 2.59 |
| HPV16/18 | Bordeaux | 53 | 1.20 | -0.12 | 0.10 | 0.53 | 1.20 | 1.88 | 2.29 | 2.51 |
| HPV16/18 | Bordeaux | 54 | 1.15 | -0.12 | 0.09 | 0.50 | 1.15 | 1.82 | 2.22 | 2.44 |
| HPV16/18 | Bordeaux | 55 | 1.11 | -0.14 | 0.08 | 0.47 | 1.11 | 1.76 | 2.15 | 2.37 |
| HPV16/18 | Bordeaux | 56 | 1.08 | -0.16 | 0.06 | 0.44 | 1.08 | 1.73 | 2.11 | 2.33 |
| HPV16/18 | Bordeaux | 57 | 1.05 | -0.18 | 0.03 | 0.39 | 1.05 | 1.70 | 2.09 | 2.32 |
| HPV16/18 | Bordeaux | 58 | 1.02 | -0.20 | 0.00 | 0.36 | 1.03 | 1.67 | 2.07 | 2.30 |
| HPV16/18 | Bordeaux | 59 | 1.01 | -0.24 | -0.02 | 0.34 | 1.01 | 1.66 | 2.06 | 2.29 |
| HPV16/18 | Bordeaux | 60 | 0.99 | -0.26 | -0.07 | 0.32 | 1.00 | 1.65 | 2.05 | 2.28 |
| HPV16/18 | Bordeaux | 61 | 0.98 | -0.32 | -0.10 | 0.29 | 0.98 | 1.65 | 2.06 | 2.25 |
| HPV16/18 | Bordeaux | 62 | 0.96 | -0.37 | -0.12 | 0.27 | 0.97 | 1.64 | 2.04 | 2.22 |
| HPV16/18 | Bordeaux | 63 | 0.94 | -0.44 | -0.18 | 0.24 | 0.95 | 1.63 | 2.01 | 2.21 |
| HPV16/18 | Bordeaux | 64 | 0.91 | -0.47 | -0.22 | 0.22 | 0.92 | 1.62 | 2.01 | 2.22 |
| HPV16/18 | Bordeaux | 65 | 0.89 | -0.54 | -0.28 | 0.18 | 0.90 | 1.60 | 2.02 | 2.22 |
| HPV16/18 | Bordeaux | 66 | 0.86 | -0.62 | -0.34 | 0.13 | 0.86 | 1.58 | 2.00 | 2.24 |
| HPV16/18 | Lille | 30 | 1.89 | -0.04 | 0.29 | 0.82 | 1.87 | 3.05 | 3.75 | 4.04 |
| HPV16/18 | Lille | 31 | 2.02 | 0.07 | 0.39 | 0.93 | 1.99 | 3.20 | 3.95 | 4.26 |
| HPV16/18 | Lille | 32 | 2.15 | 0.15 | 0.47 | 1.02 | 2.10 | 3.36 | 4.15 | 4.43 |
| HPV16/18 | Lille | 33 | 2.24 | 0.19 | 0.55 | 1.08 | 2.18 | 3.49 | 4.33 | 4.61 |
| HPV16/18 | Lille | 34 | 2.29 | 0.21 | 0.63 | 1.12 | 2.23 | 3.54 | 4.39 | 4.73 |
| HPV16/18 | Lille | 35 | 2.28 | 0.23 | 0.64 | 1.14 | 2.22 | 3.52 | 4.34 | 4.66 |

**Table S10** **Posterior difference in expected HR HPV cervical infection prevalence (in percentage points) in major French cities, between opportunistic and organised screening, stratified by type of test, city, and age. Table counterpart to Figure 3B.** ***(continued)***

| | | | Posterior distribution | | | | | | | |
|---|---|---|---|---|---|---|---|---|---|---|
| | | | | Percentiles | | | | | | |
| Virus | City | Age | Average | 0.01 | 0.025 | 0.1 | 0.5 | 0.9 | 0.975 | 0.99 |
| HPV16/18 | Lille | 36 | 2.23 | 0.23 | 0.60 | 1.12 | 2.18 | 3.46 | 4.23 | 4.55 |
| HPV16/18 | Lille | 37 | 2.17 | 0.24 | 0.59 | 1.08 | 2.12 | 3.36 | 4.12 | 4.46 |
| HPV16/18 | Lille | 38 | 2.12 | 0.22 | 0.59 | 1.05 | 2.06 | 3.27 | 4.02 | 4.34 |
| HPV16/18 | Lille | 39 | 2.06 | 0.20 | 0.57 | 1.02 | 2.00 | 3.17 | 3.92 | 4.25 |
| HPV16/18 | Lille | 40 | 1.99 | 0.20 | 0.53 | 0.98 | 1.95 | 3.09 | 3.82 | 4.13 |
| HPV16/18 | Lille | 41 | 1.93 | 0.21 | 0.49 | 0.95 | 1.89 | 3.00 | 3.72 | 4.03 |
| HPV16/18 | Lille | 42 | 1.87 | 0.20 | 0.46 | 0.91 | 1.82 | 2.91 | 3.61 | 3.89 |
| HPV16/18 | Lille | 43 | 1.81 | 0.19 | 0.43 | 0.88 | 1.76 | 2.81 | 3.52 | 3.75 |
| HPV16/18 | Lille | 44 | 1.76 | 0.17 | 0.42 | 0.85 | 1.72 | 2.74 | 3.43 | 3.67 |
| HPV16/18 | Lille | 45 | 1.71 | 0.12 | 0.40 | 0.83 | 1.68 | 2.68 | 3.35 | 3.59 |
| HPV16/18 | Lille | 46 | 1.66 | 0.10 | 0.38 | 0.80 | 1.63 | 2.59 | 3.25 | 3.50 |
| HPV16/18 | Lille | 47 | 1.61 | 0.08 | 0.34 | 0.78 | 1.58 | 2.52 | 3.15 | 3.42 |
| HPV16/18 | Lille | 48 | 1.57 | 0.07 | 0.33 | 0.74 | 1.53 | 2.45 | 3.05 | 3.35 |
| HPV16/18 | Lille | 49 | 1.52 | 0.05 | 0.30 | 0.71 | 1.50 | 2.39 | 2.97 | 3.28 |
| HPV16/18 | Lille | 50 | 1.48 | 0.03 | 0.29 | 0.68 | 1.45 | 2.33 | 2.91 | 3.20 |
| HPV16/18 | Lille | 51 | 1.44 | 0.01 | 0.26 | 0.65 | 1.41 | 2.27 | 2.84 | 3.11 |
| HPV16/18 | Lille | 52 | 1.40 | 0.01 | 0.23 | 0.62 | 1.37 | 2.22 | 2.76 | 3.05 |
| HPV16/18 | Lille | 53 | 1.35 | -0.02 | 0.21 | 0.59 | 1.32 | 2.15 | 2.67 | 2.97 |
| HPV16/18 | Lille | 54 | 1.30 | -0.04 | 0.18 | 0.55 | 1.27 | 2.07 | 2.59 | 2.88 |
| HPV16/18 | Lille | 55 | 1.26 | -0.05 | 0.16 | 0.52 | 1.22 | 2.02 | 2.53 | 2.80 |
| HPV16/18 | Lille | 56 | 1.22 | -0.06 | 0.14 | 0.49 | 1.19 | 1.97 | 2.49 | 2.75 |
| HPV16/18 | Lille | 57 | 1.19 | -0.09 | 0.12 | 0.46 | 1.16 | 1.93 | 2.45 | 2.72 |
| HPV16/18 | Lille | 58 | 1.16 | -0.12 | 0.10 | 0.44 | 1.14 | 1.90 | 2.45 | 2.67 |
| HPV16/18 | Lille | 59 | 1.14 | -0.15 | 0.05 | 0.41 | 1.12 | 1.89 | 2.42 | 2.65 |
| HPV16/18 | Lille | 60 | 1.13 | -0.19 | 0.01 | 0.39 | 1.11 | 1.89 | 2.42 | 2.67 |
| HPV16/18 | Lille | 61 | 1.12 | -0.23 | -0.03 | 0.36 | 1.10 | 1.90 | 2.42 | 2.64 |
| HPV16/18 | Lille | 62 | 1.10 | -0.29 | -0.05 | 0.33 | 1.09 | 1.89 | 2.40 | 2.64 |
| HPV16/18 | Lille | 63 | 1.08 | -0.36 | -0.10 | 0.31 | 1.07 | 1.87 | 2.39 | 2.63 |
| HPV16/18 | Lille | 64 | 1.05 | -0.42 | -0.11 | 0.28 | 1.04 | 1.85 | 2.38 | 2.66 |

**Table S10** **Posterior difference in expected HR HPV cervical infection prevalence (in percentage points) in major French cities, between opportunistic and organised screening, stratified by type of test, city, and age. Table counterpart to Figure 3B.** ***(continued)***

| | | | Posterior distribution | | | | | | | |
|---|---|---|---|---|---|---|---|---|---|---|
| | | | | Percentiles | | | | | | |
| Virus | City | Age | Average | 0.01 | 0.025 | 0.1 | 0.5 | 0.9 | 0.975 | 0.99 |
| HPV16/18 | Lille | 65 | 1.03 | -0.49 | -0.18 | 0.23 | 1.02 | 1.83 | 2.35 | 2.65 |
| HPV16/18 | Lille | 66 | 1.00 | -0.56 | -0.23 | 0.19 | 0.99 | 1.81 | 2.31 | 2.67 |
| HPV16/18 | Lyon | 30 | 1.85 | -0.30 | 0.04 | 0.67 | 1.82 | 3.12 | 3.92 | 4.49 |
| HPV16/18 | Lyon | 31 | 1.99 | -0.22 | 0.15 | 0.78 | 1.94 | 3.30 | 4.16 | 4.72 |
| HPV16/18 | Lyon | 32 | 2.11 | -0.14 | 0.24 | 0.87 | 2.05 | 3.45 | 4.34 | 4.93 |
| HPV16/18 | Lyon | 33 | 2.21 | -0.05 | 0.32 | 0.94 | 2.14 | 3.58 | 4.51 | 5.11 |
| HPV16/18 | Lyon | 34 | 2.26 | -0.02 | 0.37 | 0.97 | 2.18 | 3.63 | 4.57 | 5.13 |
| HPV16/18 | Lyon | 35 | 2.24 | 0.02 | 0.37 | 0.97 | 2.16 | 3.62 | 4.52 | 5.09 |
| HPV16/18 | Lyon | 36 | 2.20 | 0.06 | 0.37 | 0.96 | 2.12 | 3.55 | 4.44 | 5.00 |
| HPV16/18 | Lyon | 37 | 2.15 | 0.07 | 0.33 | 0.94 | 2.07 | 3.45 | 4.35 | 4.81 |
| HPV16/18 | Lyon | 38 | 2.09 | 0.06 | 0.32 | 0.93 | 2.01 | 3.37 | 4.24 | 4.70 |
| HPV16/18 | Lyon | 39 | 2.03 | 0.05 | 0.33 | 0.90 | 1.95 | 3.28 | 4.11 | 4.58 |
| HPV16/18 | Lyon | 40 | 1.97 | 0.03 | 0.33 | 0.87 | 1.90 | 3.17 | 3.99 | 4.48 |
| HPV16/18 | Lyon | 41 | 1.91 | 0.00 | 0.29 | 0.84 | 1.84 | 3.10 | 3.86 | 4.34 |
| HPV16/18 | Lyon | 42 | 1.84 | -0.02 | 0.27 | 0.80 | 1.79 | 3.01 | 3.74 | 4.21 |
| HPV16/18 | Lyon | 43 | 1.78 | -0.06 | 0.24 | 0.75 | 1.73 | 2.92 | 3.60 | 4.10 |
| HPV16/18 | Lyon | 44 | 1.73 | -0.06 | 0.22 | 0.72 | 1.67 | 2.85 | 3.51 | 4.05 |
| HPV16/18 | Lyon | 45 | 1.69 | -0.08 | 0.19 | 0.69 | 1.62 | 2.77 | 3.43 | 3.94 |
| HPV16/18 | Lyon | 46 | 1.64 | -0.10 | 0.17 | 0.66 | 1.58 | 2.70 | 3.34 | 3.87 |
| HPV16/18 | Lyon | 47 | 1.59 | -0.11 | 0.17 | 0.63 | 1.53 | 2.62 | 3.27 | 3.74 |
| HPV16/18 | Lyon | 48 | 1.54 | -0.11 | 0.16 | 0.62 | 1.49 | 2.56 | 3.21 | 3.64 |
| HPV16/18 | Lyon | 49 | 1.50 | -0.13 | 0.14 | 0.59 | 1.43 | 2.50 | 3.13 | 3.57 |
| HPV16/18 | Lyon | 50 | 1.45 | -0.14 | 0.12 | 0.55 | 1.39 | 2.43 | 3.06 | 3.47 |
| HPV16/18 | Lyon | 51 | 1.41 | -0.16 | 0.11 | 0.54 | 1.35 | 2.37 | 2.96 | 3.38 |
| HPV16/18 | Lyon | 52 | 1.37 | -0.19 | 0.10 | 0.52 | 1.32 | 2.30 | 2.89 | 3.33 |
| HPV16/18 | Lyon | 53 | 1.32 | -0.20 | 0.07 | 0.49 | 1.27 | 2.23 | 2.82 | 3.21 |
| HPV16/18 | Lyon | 54 | 1.27 | -0.22 | 0.03 | 0.45 | 1.22 | 2.15 | 2.73 | 3.08 |
| HPV16/18 | Lyon | 55 | 1.22 | -0.24 | 0.00 | 0.43 | 1.18 | 2.10 | 2.66 | 3.01 |
| HPV16/18 | Lyon | 56 | 1.19 | -0.28 | -0.04 | 0.39 | 1.14 | 2.05 | 2.58 | 2.97 |

**Table S10 Posterior difference in expected HR HPV cervical infection prevalence (in percentage points) in major French cities, between opportunistic and organised screening, stratified by type of test, city, and age. Table counterpart to Figure 3B.** ***(continued)***

| | | | Posterior distribution | | | | | | | |
|---|---|---|---|---|---|---|---|---|---|---|
| | | | | Percentiles | | | | | | |
| Virus | City | Age | Average | 0.01 | 0.025 | 0.1 | 0.5 | 0.9 | 0.975 | 0.99 |
| HPV16/18 | Lyon | 57 | 1.15 | -0.31 | -0.08 | 0.36 | 1.11 | 2.00 | 2.56 | 2.87 |
| HPV16/18 | Lyon | 58 | 1.13 | -0.33 | -0.10 | 0.34 | 1.09 | 1.97 | 2.51 | 2.87 |
| HPV16/18 | Lyon | 59 | 1.11 | -0.35 | -0.12 | 0.33 | 1.06 | 1.94 | 2.52 | 2.89 |
| HPV16/18 | Lyon | 60 | 1.09 | -0.37 | -0.17 | 0.30 | 1.06 | 1.92 | 2.51 | 2.87 |
| HPV16/18 | Lyon | 61 | 1.08 | -0.42 | -0.21 | 0.28 | 1.04 | 1.92 | 2.51 | 2.85 |
| HPV16/18 | Lyon | 62 | 1.06 | -0.48 | -0.26 | 0.25 | 1.02 | 1.90 | 2.49 | 2.82 |
| HPV16/18 | Lyon | 63 | 1.03 | -0.53 | -0.29 | 0.21 | 1.00 | 1.89 | 2.48 | 2.76 |
| HPV16/18 | Lyon | 64 | 1.01 | -0.58 | -0.33 | 0.18 | 0.98 | 1.88 | 2.46 | 2.73 |
| HPV16/18 | Lyon | 65 | 0.98 | -0.66 | -0.38 | 0.14 | 0.96 | 1.86 | 2.43 | 2.72 |
| HPV16/18 | Lyon | 66 | 0.94 | -0.70 | -0.44 | 0.09 | 0.93 | 1.84 | 2.43 | 2.71 |
| HPV16/18 | Marseille | 30 | 1.11 | -1.02 | -0.65 | 0.04 | 1.16 | 2.13 | 2.57 | 2.91 |
| HPV16/18 | Marseille | 31 | 1.21 | -0.92 | -0.58 | 0.12 | 1.26 | 2.22 | 2.72 | 3.02 |
| HPV16/18 | Marseille | 32 | 1.30 | -0.91 | -0.53 | 0.18 | 1.34 | 2.34 | 2.87 | 3.13 |
| HPV16/18 | Marseille | 33 | 1.37 | -0.85 | -0.47 | 0.22 | 1.42 | 2.43 | 2.97 | 3.25 |
| HPV16/18 | Marseille | 34 | 1.40 | -0.81 | -0.42 | 0.26 | 1.45 | 2.47 | 2.98 | 3.33 |
| HPV16/18 | Marseille | 35 | 1.40 | -0.77 | -0.40 | 0.27 | 1.45 | 2.45 | 2.96 | 3.31 |
| HPV16/18 | Marseille | 36 | 1.37 | -0.72 | -0.39 | 0.27 | 1.42 | 2.41 | 2.92 | 3.26 |
| HPV16/18 | Marseille | 37 | 1.33 | -0.69 | -0.39 | 0.26 | 1.38 | 2.33 | 2.86 | 3.17 |
| HPV16/18 | Marseille | 38 | 1.30 | -0.67 | -0.38 | 0.24 | 1.34 | 2.27 | 2.78 | 3.09 |
| HPV16/18 | Marseille | 39 | 1.26 | -0.67 | -0.37 | 0.25 | 1.29 | 2.20 | 2.70 | 2.98 |
| HPV16/18 | Marseille | 40 | 1.21 | -0.64 | -0.37 | 0.22 | 1.26 | 2.14 | 2.63 | 2.89 |
| HPV16/18 | Marseille | 41 | 1.17 | -0.65 | -0.38 | 0.21 | 1.21 | 2.09 | 2.55 | 2.79 |
| HPV16/18 | Marseille | 42 | 1.13 | -0.66 | -0.38 | 0.18 | 1.16 | 2.03 | 2.48 | 2.72 |
| HPV16/18 | Marseille | 43 | 1.09 | -0.64 | -0.38 | 0.18 | 1.12 | 1.96 | 2.41 | 2.63 |
| HPV16/18 | Marseille | 44 | 1.05 | -0.68 | -0.39 | 0.16 | 1.08 | 1.91 | 2.34 | 2.57 |
| HPV16/18 | Marseille | 45 | 1.02 | -0.68 | -0.40 | 0.14 | 1.05 | 1.87 | 2.29 | 2.48 |
| HPV16/18 | Marseille | 46 | 0.98 | -0.65 | -0.40 | 0.13 | 1.01 | 1.82 | 2.23 | 2.40 |
| HPV16/18 | Marseille | 47 | 0.95 | -0.66 | -0.39 | 0.11 | 0.97 | 1.76 | 2.17 | 2.34 |
| HPV16/18 | Marseille | 48 | 0.91 | -0.69 | -0.42 | 0.09 | 0.94 | 1.71 | 2.11 | 2.30 |

**Table S10** **Posterior difference in expected HR HPV cervical infection prevalence (in percentage points) in major French cities, between opportunistic and organised screening, stratified by type of test, city, and age. Table counterpart to Figure 3B.** ***(continued)***

| | | | Posterior distribution | | | | | | | |
|---|---|---|---|---|---|---|---|---|---|---|
| | | | | Percentiles | | | | | | |
| Virus | City | Age | Average | 0.01 | 0.025 | 0.1 | 0.5 | 0.9 | 0.975 | 0.99 |
| HPV16/18 | Marseille | 49 | 0.88 | -0.69 | -0.44 | 0.07 | 0.91 | 1.67 | 2.04 | 2.22 |
| HPV16/18 | Marseille | 50 | 0.85 | -0.71 | -0.45 | 0.05 | 0.87 | 1.63 | 1.99 | 2.17 |
| HPV16/18 | Marseille | 51 | 0.82 | -0.73 | -0.45 | 0.03 | 0.85 | 1.58 | 1.94 | 2.13 |
| HPV16/18 | Marseille | 52 | 0.79 | -0.75 | -0.46 | 0.01 | 0.81 | 1.53 | 1.89 | 2.09 |
| HPV16/18 | Marseille | 53 | 0.75 | -0.78 | -0.48 | 0.00 | 0.78 | 1.49 | 1.84 | 2.00 |
| HPV16/18 | Marseille | 54 | 0.72 | -0.77 | -0.50 | -0.03 | 0.74 | 1.44 | 1.77 | 1.96 |
| HPV16/18 | Marseille | 55 | 0.68 | -0.77 | -0.50 | -0.05 | 0.71 | 1.40 | 1.73 | 1.89 |
| HPV16/18 | Marseille | 56 | 0.65 | -0.79 | -0.52 | -0.08 | 0.68 | 1.37 | 1.70 | 1.85 |
| HPV16/18 | Marseille | 57 | 0.63 | -0.83 | -0.53 | -0.11 | 0.65 | 1.34 | 1.66 | 1.82 |
| HPV16/18 | Marseille | 58 | 0.60 | -0.86 | -0.58 | -0.13 | 0.63 | 1.31 | 1.64 | 1.80 |
| HPV16/18 | Marseille | 59 | 0.59 | -0.90 | -0.61 | -0.16 | 0.61 | 1.29 | 1.65 | 1.79 |
| HPV16/18 | Marseille | 60 | 0.57 | -0.93 | -0.63 | -0.17 | 0.60 | 1.29 | 1.64 | 1.79 |
| HPV16/18 | Marseille | 61 | 0.55 | -0.97 | -0.68 | -0.21 | 0.59 | 1.28 | 1.64 | 1.77 |
| HPV16/18 | Marseille | 62 | 0.54 | -1.01 | -0.70 | -0.25 | 0.57 | 1.27 | 1.61 | 1.79 |
| HPV16/18 | Marseille | 63 | 0.51 | -1.05 | -0.77 | -0.29 | 0.56 | 1.26 | 1.61 | 1.80 |
| HPV16/18 | Marseille | 64 | 0.49 | -1.10 | -0.83 | -0.32 | 0.54 | 1.25 | 1.60 | 1.79 |
| HPV16/18 | Marseille | 65 | 0.47 | -1.15 | -0.88 | -0.38 | 0.51 | 1.23 | 1.60 | 1.78 |
| HPV16/18 | Marseille | 66 | 0.44 | -1.21 | -0.95 | -0.42 | 0.48 | 1.22 | 1.59 | 1.80 |
| HPV16/18 | Montpellier | 30 | 1.70 | -0.28 | 0.02 | 0.58 | 1.68 | 2.87 | 3.53 | 3.89 |
| HPV16/18 | Montpellier | 31 | 1.83 | -0.18 | 0.12 | 0.69 | 1.80 | 3.02 | 3.70 | 4.10 |
| HPV16/18 | Montpellier | 32 | 1.95 | -0.10 | 0.19 | 0.79 | 1.91 | 3.17 | 3.89 | 4.33 |
| HPV16/18 | Montpellier | 33 | 2.04 | 0.00 | 0.23 | 0.85 | 1.99 | 3.29 | 4.01 | 4.47 |
| HPV16/18 | Montpellier | 34 | 2.09 | 0.02 | 0.29 | 0.89 | 2.03 | 3.32 | 4.07 | 4.56 |
| HPV16/18 | Montpellier | 35 | 2.08 | 0.00 | 0.30 | 0.90 | 2.02 | 3.29 | 4.02 | 4.55 |
| HPV16/18 | Montpellier | 36 | 2.04 | 0.04 | 0.29 | 0.90 | 1.99 | 3.24 | 3.93 | 4.47 |
| HPV16/18 | Montpellier | 37 | 1.98 | 0.04 | 0.30 | 0.87 | 1.94 | 3.15 | 3.84 | 4.33 |
| HPV16/18 | Montpellier | 38 | 1.93 | 0.01 | 0.29 | 0.85 | 1.89 | 3.07 | 3.75 | 4.22 |
| HPV16/18 | Montpellier | 39 | 1.88 | -0.02 | 0.26 | 0.82 | 1.83 | 3.00 | 3.64 | 4.11 |
| HPV16/18 | Montpellier | 40 | 1.82 | -0.03 | 0.22 | 0.78 | 1.77 | 2.92 | 3.58 | 4.00 |

**Table S10** **Posterior difference in expected HR HPV cervical infection prevalence (in percentage points) in major French cities, between opportunistic and organised screening, stratified by type of test, city, and age. Table counterpart to Figure 3B.** ***(continued)***

| | | | Posterior distribution | | | | | | | |
|---|---|---|---|---|---|---|---|---|---|---|
| | | | | Percentiles | | | | | | |
| Virus | City | Age | Average | 0.01 | 0.025 | 0.1 | 0.5 | 0.9 | 0.975 | 0.99 |
| HPV16/18 | Montpellier | 41 | 1.76 | -0.07 | 0.20 | 0.75 | 1.71 | 2.84 | 3.50 | 3.88 |
| HPV16/18 | Montpellier | 42 | 1.70 | -0.09 | 0.19 | 0.72 | 1.66 | 2.73 | 3.40 | 3.77 |
| HPV16/18 | Montpellier | 43 | 1.64 | -0.12 | 0.18 | 0.69 | 1.61 | 2.64 | 3.29 | 3.64 |
| HPV16/18 | Montpellier | 44 | 1.60 | -0.12 | 0.16 | 0.66 | 1.56 | 2.59 | 3.21 | 3.55 |
| HPV16/18 | Montpellier | 45 | 1.55 | -0.14 | 0.14 | 0.63 | 1.52 | 2.52 | 3.12 | 3.48 |
| HPV16/18 | Montpellier | 46 | 1.51 | -0.13 | 0.12 | 0.60 | 1.47 | 2.44 | 3.05 | 3.39 |
| HPV16/18 | Montpellier | 47 | 1.46 | -0.14 | 0.11 | 0.57 | 1.42 | 2.38 | 2.97 | 3.33 |
| HPV16/18 | Montpellier | 48 | 1.42 | -0.17 | 0.09 | 0.54 | 1.39 | 2.33 | 2.89 | 3.21 |
| HPV16/18 | Montpellier | 49 | 1.37 | -0.17 | 0.08 | 0.52 | 1.35 | 2.27 | 2.83 | 3.14 |
| HPV16/18 | Montpellier | 50 | 1.33 | -0.17 | 0.07 | 0.49 | 1.30 | 2.20 | 2.76 | 3.08 |
| HPV16/18 | Montpellier | 51 | 1.29 | -0.20 | 0.06 | 0.47 | 1.27 | 2.15 | 2.71 | 3.04 |
| HPV16/18 | Montpellier | 52 | 1.25 | -0.22 | 0.03 | 0.44 | 1.23 | 2.10 | 2.62 | 2.96 |
| HPV16/18 | Montpellier | 53 | 1.21 | -0.23 | -0.01 | 0.41 | 1.20 | 2.03 | 2.54 | 2.87 |
| HPV16/18 | Montpellier | 54 | 1.16 | -0.24 | -0.04 | 0.38 | 1.14 | 1.96 | 2.47 | 2.77 |
| HPV16/18 | Montpellier | 55 | 1.12 | -0.26 | -0.06 | 0.35 | 1.10 | 1.90 | 2.40 | 2.71 |
| HPV16/18 | Montpellier | 56 | 1.08 | -0.28 | -0.10 | 0.32 | 1.06 | 1.86 | 2.35 | 2.63 |
| HPV16/18 | Montpellier | 57 | 1.05 | -0.31 | -0.12 | 0.30 | 1.03 | 1.83 | 2.31 | 2.59 |
| HPV16/18 | Montpellier | 58 | 1.02 | -0.32 | -0.13 | 0.28 | 1.01 | 1.81 | 2.28 | 2.56 |
| HPV16/18 | Montpellier | 59 | 1.00 | -0.36 | -0.18 | 0.25 | 0.99 | 1.78 | 2.26 | 2.54 |
| HPV16/18 | Montpellier | 60 | 0.99 | -0.40 | -0.21 | 0.23 | 0.98 | 1.76 | 2.27 | 2.54 |
| HPV16/18 | Montpellier | 61 | 0.98 | -0.43 | -0.24 | 0.20 | 0.96 | 1.76 | 2.25 | 2.55 |
| HPV16/18 | Montpellier | 62 | 0.96 | -0.49 | -0.28 | 0.18 | 0.94 | 1.75 | 2.27 | 2.52 |
| HPV16/18 | Montpellier | 63 | 0.93 | -0.58 | -0.30 | 0.15 | 0.92 | 1.73 | 2.24 | 2.53 |
| HPV16/18 | Montpellier | 64 | 0.91 | -0.64 | -0.36 | 0.11 | 0.89 | 1.71 | 2.22 | 2.49 |
| HPV16/18 | Montpellier | 65 | 0.88 | -0.71 | -0.40 | 0.07 | 0.87 | 1.69 | 2.21 | 2.48 |
| HPV16/18 | Montpellier | 66 | 0.85 | -0.76 | -0.46 | 0.02 | 0.85 | 1.67 | 2.20 | 2.47 |
| HPV16/18 | Nantes | 30 | 1.85 | 0.09 | 0.37 | 0.82 | 1.80 | 2.98 | 3.72 | 4.08 |
| HPV16/18 | Nantes | 31 | 1.98 | 0.19 | 0.46 | 0.91 | 1.92 | 3.15 | 3.93 | 4.34 |
| HPV16/18 | Nantes | 32 | 2.10 | 0.27 | 0.54 | 1.01 | 2.03 | 3.32 | 4.09 | 4.55 |

**Table S10 Posterior difference in expected HR HPV cervical infection prevalence (in percentage points) in major French cities, between opportunistic and organised screening, stratified by type of test, city, and age. Table counterpart to Figure 3B.** *(continued)*

| | | | Posterior distribution | | | | | | | |
|---|---|---|---|---|---|---|---|---|---|---|
| | | | | Percentiles | | | | | | |
| Virus | City | Age | Average | 0.01 | 0.025 | 0.1 | 0.5 | 0.9 | 0.975 | 0.99 |
| HPV16/18 | Nantes | 33 | 2.20 | 0.32 | 0.60 | 1.08 | 2.12 | 3.45 | 4.23 | 4.66 |
| HPV16/18 | Nantes | 34 | 2.24 | 0.37 | 0.64 | 1.11 | 2.16 | 3.49 | 4.30 | 4.74 |
| HPV16/18 | Nantes | 35 | 2.23 | 0.37 | 0.63 | 1.13 | 2.14 | 3.45 | 4.28 | 4.75 |
| HPV16/18 | Nantes | 36 | 2.19 | 0.39 | 0.61 | 1.11 | 2.11 | 3.39 | 4.19 | 4.66 |
| HPV16/18 | Nantes | 37 | 2.13 | 0.38 | 0.62 | 1.08 | 2.06 | 3.31 | 4.06 | 4.55 |
| HPV16/18 | Nantes | 38 | 2.07 | 0.39 | 0.61 | 1.04 | 2.00 | 3.22 | 3.97 | 4.43 |
| HPV16/18 | Nantes | 39 | 2.01 | 0.38 | 0.60 | 1.01 | 1.94 | 3.13 | 3.85 | 4.29 |
| HPV16/18 | Nantes | 40 | 1.95 | 0.35 | 0.57 | 0.98 | 1.88 | 3.05 | 3.73 | 4.21 |
| HPV16/18 | Nantes | 41 | 1.89 | 0.32 | 0.56 | 0.95 | 1.82 | 2.96 | 3.63 | 4.06 |
| HPV16/18 | Nantes | 42 | 1.83 | 0.30 | 0.53 | 0.92 | 1.76 | 2.87 | 3.55 | 3.97 |
| HPV16/18 | Nantes | 43 | 1.77 | 0.28 | 0.49 | 0.89 | 1.70 | 2.79 | 3.43 | 3.82 |
| HPV16/18 | Nantes | 44 | 1.72 | 0.26 | 0.47 | 0.85 | 1.65 | 2.71 | 3.35 | 3.72 |
| HPV16/18 | Nantes | 45 | 1.68 | 0.23 | 0.46 | 0.82 | 1.61 | 2.66 | 3.27 | 3.64 |
| HPV16/18 | Nantes | 46 | 1.63 | 0.22 | 0.44 | 0.80 | 1.57 | 2.59 | 3.17 | 3.55 |
| HPV16/18 | Nantes | 47 | 1.58 | 0.20 | 0.42 | 0.77 | 1.53 | 2.52 | 3.10 | 3.48 |
| HPV16/18 | Nantes | 48 | 1.54 | 0.18 | 0.40 | 0.75 | 1.49 | 2.46 | 3.00 | 3.38 |
| HPV16/18 | Nantes | 49 | 1.50 | 0.18 | 0.36 | 0.71 | 1.44 | 2.40 | 2.92 | 3.28 |
| HPV16/18 | Nantes | 50 | 1.45 | 0.17 | 0.35 | 0.68 | 1.40 | 2.33 | 2.86 | 3.18 |
| HPV16/18 | Nantes | 51 | 1.41 | 0.14 | 0.33 | 0.65 | 1.37 | 2.27 | 2.80 | 3.08 |
| HPV16/18 | Nantes | 52 | 1.37 | 0.14 | 0.30 | 0.63 | 1.32 | 2.21 | 2.72 | 3.00 |
| HPV16/18 | Nantes | 53 | 1.33 | 0.13 | 0.28 | 0.60 | 1.28 | 2.14 | 2.64 | 2.96 |
| HPV16/18 | Nantes | 54 | 1.28 | 0.10 | 0.26 | 0.57 | 1.24 | 2.08 | 2.55 | 2.86 |
| HPV16/18 | Nantes | 55 | 1.24 | 0.09 | 0.24 | 0.54 | 1.20 | 2.02 | 2.47 | 2.78 |
| HPV16/18 | Nantes | 56 | 1.20 | 0.06 | 0.22 | 0.52 | 1.16 | 1.97 | 2.42 | 2.75 |
| HPV16/18 | Nantes | 57 | 1.17 | 0.02 | 0.20 | 0.49 | 1.14 | 1.93 | 2.38 | 2.66 |
| HPV16/18 | Nantes | 58 | 1.15 | -0.02 | 0.16 | 0.47 | 1.11 | 1.90 | 2.34 | 2.63 |
| HPV16/18 | Nantes | 59 | 1.13 | -0.05 | 0.13 | 0.46 | 1.09 | 1.88 | 2.34 | 2.63 |
| HPV16/18 | Nantes | 60 | 1.12 | -0.07 | 0.10 | 0.44 | 1.08 | 1.87 | 2.34 | 2.61 |
| HPV16/18 | Nantes | 61 | 1.11 | -0.09 | 0.07 | 0.42 | 1.07 | 1.87 | 2.32 | 2.60 |

**Table S10** **Posterior difference in expected HR HPV cervical infection prevalence (in percentage points) in major French cities, between opportunistic and organised screening, stratified by type of test, city, and age. Table counterpart to Figure 3B.** ***(continued)***

| | | | Posterior distribution | | | | | | | |
|---|---|---|---|---|---|---|---|---|---|---|
| | | | | Percentiles | | | | | | |
| Virus | City | Age | Average | 0.01 | 0.025 | 0.1 | 0.5 | 0.9 | 0.975 | 0.99 |
| HPV16/18 | Nantes | 62 | 1.09 | -0.13 | 0.04 | 0.41 | 1.06 | 1.86 | 2.30 | 2.58 |
| HPV16/18 | Nantes | 63 | 1.07 | -0.20 | 0.02 | 0.38 | 1.04 | 1.85 | 2.29 | 2.57 |
| HPV16/18 | Nantes | 64 | 1.05 | -0.25 | -0.02 | 0.35 | 1.02 | 1.83 | 2.27 | 2.57 |
| HPV16/18 | Nantes | 65 | 1.02 | -0.31 | -0.07 | 0.31 | 0.99 | 1.80 | 2.27 | 2.52 |
| HPV16/18 | Nantes | 66 | 0.99 | -0.41 | -0.12 | 0.28 | 0.96 | 1.77 | 2.28 | 2.52 |
| HPV16/18 | Nice | 30 | 1.19 | -1.05 | -0.56 | 0.09 | 1.13 | 2.35 | 3.24 | 3.72 |
| HPV16/18 | Nice | 31 | 1.29 | -0.97 | -0.49 | 0.16 | 1.23 | 2.48 | 3.40 | 3.92 |
| HPV16/18 | Nice | 32 | 1.38 | -0.87 | -0.47 | 0.22 | 1.31 | 2.61 | 3.58 | 4.10 |
| HPV16/18 | Nice | 33 | 1.45 | -0.84 | -0.41 | 0.26 | 1.38 | 2.71 | 3.73 | 4.26 |
| HPV16/18 | Nice | 34 | 1.48 | -0.81 | -0.39 | 0.27 | 1.41 | 2.77 | 3.76 | 4.30 |
| HPV16/18 | Nice | 35 | 1.47 | -0.76 | -0.38 | 0.29 | 1.40 | 2.75 | 3.76 | 4.24 |
| HPV16/18 | Nice | 36 | 1.45 | -0.74 | -0.37 | 0.28 | 1.38 | 2.69 | 3.67 | 4.20 |
| HPV16/18 | Nice | 37 | 1.41 | -0.74 | -0.36 | 0.29 | 1.34 | 2.62 | 3.57 | 4.08 |
| HPV16/18 | Nice | 38 | 1.37 | -0.75 | -0.35 | 0.29 | 1.30 | 2.54 | 3.49 | 3.99 |
| HPV16/18 | Nice | 39 | 1.33 | -0.71 | -0.35 | 0.27 | 1.26 | 2.49 | 3.38 | 3.90 |
| HPV16/18 | Nice | 40 | 1.28 | -0.75 | -0.34 | 0.26 | 1.22 | 2.41 | 3.29 | 3.79 |
| HPV16/18 | Nice | 41 | 1.24 | -0.74 | -0.36 | 0.25 | 1.17 | 2.34 | 3.21 | 3.73 |
| HPV16/18 | Nice | 42 | 1.20 | -0.71 | -0.36 | 0.22 | 1.12 | 2.29 | 3.11 | 3.59 |
| HPV16/18 | Nice | 43 | 1.16 | -0.72 | -0.37 | 0.20 | 1.08 | 2.21 | 2.98 | 3.51 |
| HPV16/18 | Nice | 44 | 1.12 | -0.71 | -0.37 | 0.18 | 1.05 | 2.15 | 2.91 | 3.44 |
| HPV16/18 | Nice | 45 | 1.09 | -0.71 | -0.38 | 0.17 | 1.03 | 2.09 | 2.86 | 3.37 |
| HPV16/18 | Nice | 46 | 1.05 | -0.69 | -0.39 | 0.15 | 0.99 | 2.03 | 2.77 | 3.29 |
| HPV16/18 | Nice | 47 | 1.02 | -0.69 | -0.40 | 0.14 | 0.96 | 1.99 | 2.69 | 3.22 |
| HPV16/18 | Nice | 48 | 0.98 | -0.70 | -0.40 | 0.13 | 0.93 | 1.94 | 2.64 | 3.13 |
| HPV16/18 | Nice | 49 | 0.95 | -0.70 | -0.41 | 0.11 | 0.91 | 1.89 | 2.56 | 3.02 |
| HPV16/18 | Nice | 50 | 0.92 | -0.69 | -0.42 | 0.10 | 0.88 | 1.84 | 2.49 | 2.97 |
| HPV16/18 | Nice | 51 | 0.89 | -0.70 | -0.43 | 0.07 | 0.85 | 1.79 | 2.44 | 2.88 |
| HPV16/18 | Nice | 52 | 0.86 | -0.71 | -0.43 | 0.05 | 0.82 | 1.72 | 2.39 | 2.80 |
| HPV16/18 | Nice | 53 | 0.82 | -0.71 | -0.44 | 0.03 | 0.79 | 1.67 | 2.31 | 2.73 |

**Table S10** **Posterior difference in expected HR HPV cervical infection prevalence (in percentage points) in major French cities, between opportunistic and organised screening, stratified by type of test, city, and age. Table counterpart to Figure 3B.** ***(continued)***

| | | | Posterior distribution | | | | | | | |
|---|---|---|---|---|---|---|---|---|---|---|
| | | | | Percentiles | | | | | | |
| Virus | City | Age | Average | 0.01 | 0.025 | 0.1 | 0.5 | 0.9 | 0.975 | 0.99 |
| HPV16/18 | Nice | 54 | 0.79 | -0.71 | -0.46 | 0.00 | 0.75 | 1.61 | 2.26 | 2.61 |
| HPV16/18 | Nice | 55 | 0.76 | -0.71 | -0.48 | -0.01 | 0.72 | 1.57 | 2.20 | 2.56 |
| HPV16/18 | Nice | 56 | 0.73 | -0.73 | -0.49 | -0.03 | 0.70 | 1.53 | 2.15 | 2.50 |
| HPV16/18 | Nice | 57 | 0.70 | -0.76 | -0.51 | -0.05 | 0.67 | 1.51 | 2.11 | 2.44 |
| HPV16/18 | Nice | 58 | 0.68 | -0.76 | -0.52 | -0.07 | 0.65 | 1.49 | 2.06 | 2.43 |
| HPV16/18 | Nice | 59 | 0.67 | -0.78 | -0.53 | -0.09 | 0.64 | 1.46 | 2.02 | 2.40 |
| HPV16/18 | Nice | 60 | 0.65 | -0.80 | -0.55 | -0.11 | 0.63 | 1.45 | 2.02 | 2.36 |
| HPV16/18 | Nice | 61 | 0.64 | -0.85 | -0.58 | -0.13 | 0.62 | 1.45 | 2.02 | 2.36 |
| HPV16/18 | Nice | 62 | 0.62 | -0.88 | -0.61 | -0.17 | 0.60 | 1.44 | 2.00 | 2.38 |
| HPV16/18 | Nice | 63 | 0.60 | -0.92 | -0.64 | -0.19 | 0.59 | 1.42 | 1.97 | 2.35 |
| HPV16/18 | Nice | 64 | 0.58 | -0.95 | -0.68 | -0.22 | 0.56 | 1.40 | 1.99 | 2.35 |
| HPV16/18 | Nice | 65 | 0.56 | -1.00 | -0.71 | -0.26 | 0.54 | 1.39 | 1.95 | 2.36 |
| HPV16/18 | Nice | 66 | 0.54 | -1.07 | -0.75 | -0.29 | 0.52 | 1.36 | 1.94 | 2.31 |
| HPV16/18 | Paris | 30 | 1.97 | -0.10 | 0.23 | 0.86 | 1.97 | 3.05 | 3.67 | 3.99 |
| HPV16/18 | Paris | 31 | 2.11 | 0.03 | 0.38 | 1.00 | 2.11 | 3.22 | 3.84 | 4.21 |
| HPV16/18 | Paris | 32 | 2.24 | 0.14 | 0.48 | 1.11 | 2.24 | 3.39 | 4.01 | 4.34 |
| HPV16/18 | Paris | 33 | 2.35 | 0.21 | 0.56 | 1.21 | 2.34 | 3.51 | 4.17 | 4.46 |
| HPV16/18 | Paris | 34 | 2.39 | 0.30 | 0.59 | 1.26 | 2.38 | 3.56 | 4.22 | 4.50 |
| HPV16/18 | Paris | 35 | 2.38 | 0.30 | 0.60 | 1.27 | 2.37 | 3.53 | 4.15 | 4.47 |
| HPV16/18 | Paris | 36 | 2.34 | 0.30 | 0.59 | 1.26 | 2.32 | 3.46 | 4.10 | 4.38 |
| HPV16/18 | Paris | 37 | 2.28 | 0.30 | 0.60 | 1.23 | 2.26 | 3.37 | 3.98 | 4.27 |
| HPV16/18 | Paris | 38 | 2.22 | 0.31 | 0.59 | 1.20 | 2.20 | 3.28 | 3.85 | 4.17 |
| HPV16/18 | Paris | 39 | 2.15 | 0.29 | 0.58 | 1.16 | 2.14 | 3.19 | 3.75 | 4.05 |
| HPV16/18 | Paris | 40 | 2.09 | 0.29 | 0.54 | 1.12 | 2.09 | 3.08 | 3.66 | 3.96 |
| HPV16/18 | Paris | 41 | 2.03 | 0.26 | 0.54 | 1.07 | 2.03 | 3.00 | 3.54 | 3.82 |
| HPV16/18 | Paris | 42 | 1.96 | 0.21 | 0.49 | 1.02 | 1.96 | 2.91 | 3.46 | 3.71 |
| HPV16/18 | Paris | 43 | 1.89 | 0.21 | 0.46 | 1.00 | 1.90 | 2.82 | 3.35 | 3.59 |
| HPV16/18 | Paris | 44 | 1.84 | 0.16 | 0.46 | 0.96 | 1.85 | 2.75 | 3.26 | 3.51 |
| HPV16/18 | Paris | 45 | 1.79 | 0.17 | 0.44 | 0.93 | 1.80 | 2.68 | 3.18 | 3.41 |

**Table S10** **Posterior difference in expected HR HPV cervical infection prevalence (in percentage points) in major French cities, between opportunistic and organised screening, stratified by type of test, city, and age. Table counterpart to Figure 3B.** ***(continued)***

| Virus | City | Age | Posterior distribution | | | | | | | |
|---|---|---|---|---|---|---|---|---|---|---|
| | | | | Percentiles | | | | | | |
| | | | Average | 0.01 | 0.025 | 0.1 | 0.5 | 0.9 | 0.975 | 0.99 |
| HPV16/18 | Paris | 46 | 1.74 | 0.15 | 0.41 | 0.90 | 1.75 | 2.61 | 3.10 | 3.32 |
| HPV16/18 | Paris | 47 | 1.69 | 0.13 | 0.37 | 0.86 | 1.69 | 2.54 | 3.02 | 3.25 |
| HPV16/18 | Paris | 48 | 1.64 | 0.11 | 0.34 | 0.83 | 1.64 | 2.47 | 2.94 | 3.17 |
| HPV16/18 | Paris | 49 | 1.59 | 0.09 | 0.32 | 0.78 | 1.59 | 2.41 | 2.86 | 3.12 |
| HPV16/18 | Paris | 50 | 1.54 | 0.07 | 0.29 | 0.74 | 1.55 | 2.35 | 2.78 | 3.03 |
| HPV16/18 | Paris | 51 | 1.50 | 0.05 | 0.26 | 0.70 | 1.50 | 2.30 | 2.73 | 2.94 |
| HPV16/18 | Paris | 52 | 1.45 | 0.05 | 0.24 | 0.67 | 1.45 | 2.25 | 2.65 | 2.90 |
| HPV16/18 | Paris | 53 | 1.40 | 0.02 | 0.22 | 0.64 | 1.41 | 2.17 | 2.56 | 2.79 |
| HPV16/18 | Paris | 54 | 1.35 | 0.00 | 0.18 | 0.60 | 1.35 | 2.09 | 2.49 | 2.73 |
| HPV16/18 | Paris | 55 | 1.30 | -0.03 | 0.16 | 0.56 | 1.31 | 2.03 | 2.44 | 2.67 |
| HPV16/18 | Paris | 56 | 1.26 | -0.07 | 0.14 | 0.52 | 1.27 | 1.99 | 2.38 | 2.62 |
| HPV16/18 | Paris | 57 | 1.22 | -0.11 | 0.11 | 0.49 | 1.24 | 1.95 | 2.34 | 2.60 |
| HPV16/18 | Paris | 58 | 1.20 | -0.12 | 0.08 | 0.46 | 1.20 | 1.91 | 2.33 | 2.58 |
| HPV16/18 | Paris | 59 | 1.18 | -0.15 | 0.06 | 0.42 | 1.18 | 1.89 | 2.32 | 2.58 |
| HPV16/18 | Paris | 60 | 1.16 | -0.21 | 0.02 | 0.40 | 1.17 | 1.89 | 2.34 | 2.59 |
| HPV16/18 | Paris | 61 | 1.14 | -0.24 | -0.01 | 0.37 | 1.15 | 1.90 | 2.34 | 2.58 |
| HPV16/18 | Paris | 62 | 1.12 | -0.28 | -0.04 | 0.34 | 1.13 | 1.89 | 2.33 | 2.56 |
| HPV16/18 | Paris | 63 | 1.10 | -0.34 | -0.10 | 0.29 | 1.11 | 1.87 | 2.31 | 2.59 |
| HPV16/18 | Paris | 64 | 1.07 | -0.41 | -0.14 | 0.26 | 1.08 | 1.86 | 2.29 | 2.56 |
| HPV16/18 | Paris | 65 | 1.04 | -0.50 | -0.20 | 0.21 | 1.05 | 1.84 | 2.28 | 2.54 |
| HPV16/18 | Paris | 66 | 1.00 | -0.56 | -0.28 | 0.16 | 1.02 | 1.83 | 2.30 | 2.49 |
| HPV16/18 | Rennes | 30 | 2.50 | 0.65 | 0.99 | 1.47 | 2.49 | 3.54 | 4.07 | 4.48 |
| HPV16/18 | Rennes | 31 | 2.67 | 0.77 | 1.10 | 1.63 | 2.67 | 3.74 | 4.29 | 4.71 |
| HPV16/18 | Rennes | 32 | 2.82 | 0.88 | 1.22 | 1.75 | 2.83 | 3.92 | 4.50 | 4.93 |
| HPV16/18 | Rennes | 33 | 2.94 | 1.00 | 1.31 | 1.85 | 2.95 | 4.06 | 4.66 | 5.13 |
| HPV16/18 | Rennes | 34 | 3.00 | 1.08 | 1.33 | 1.90 | 2.99 | 4.12 | 4.73 | 5.19 |
| HPV16/18 | Rennes | 35 | 2.98 | 1.05 | 1.34 | 1.89 | 2.97 | 4.09 | 4.70 | 5.14 |
| HPV16/18 | Rennes | 36 | 2.92 | 1.04 | 1.34 | 1.86 | 2.91 | 4.02 | 4.61 | 5.01 |
| HPV16/18 | Rennes | 37 | 2.85 | 1.02 | 1.31 | 1.81 | 2.83 | 3.92 | 4.47 | 4.88 |

**Table S10** **Posterior difference in expected HR HPV cervical infection prevalence (in percentage points) in major French cities, between opportunistic and organised screening, stratified by type of test, city, and age. Table counterpart to Figure 3B.** ***(continued)***

| | | | Posterior distribution | | | | | | | |
|---|---|---|---|---|---|---|---|---|---|---|
| | | | | Percentiles | | | | | | |
| Virus | City | Age | Average | 0.01 | 0.025 | 0.1 | 0.5 | 0.9 | 0.975 | 0.99 |
| HPV16/18 | Rennes | 38 | 2.77 | 1.02 | 1.29 | 1.76 | 2.75 | 3.82 | 4.36 | 4.80 |
| HPV16/18 | Rennes | 39 | 2.70 | 0.99 | 1.24 | 1.71 | 2.68 | 3.72 | 4.24 | 4.65 |
| HPV16/18 | Rennes | 40 | 2.62 | 0.97 | 1.20 | 1.66 | 2.60 | 3.61 | 4.12 | 4.50 |
| HPV16/18 | Rennes | 41 | 2.54 | 0.94 | 1.14 | 1.61 | 2.53 | 3.51 | 4.00 | 4.39 |
| HPV16/18 | Rennes | 42 | 2.46 | 0.89 | 1.12 | 1.55 | 2.44 | 3.39 | 3.90 | 4.26 |
| HPV16/18 | Rennes | 43 | 2.39 | 0.85 | 1.07 | 1.50 | 2.37 | 3.29 | 3.79 | 4.15 |
| HPV16/18 | Rennes | 44 | 2.32 | 0.83 | 1.03 | 1.45 | 2.30 | 3.21 | 3.70 | 4.03 |
| HPV16/18 | Rennes | 45 | 2.27 | 0.80 | 0.99 | 1.42 | 2.25 | 3.14 | 3.60 | 3.93 |
| HPV16/18 | Rennes | 46 | 2.20 | 0.76 | 0.95 | 1.37 | 2.19 | 3.06 | 3.53 | 3.84 |
| HPV16/18 | Rennes | 47 | 2.14 | 0.73 | 0.90 | 1.31 | 2.13 | 2.99 | 3.42 | 3.74 |
| HPV16/18 | Rennes | 48 | 2.08 | 0.70 | 0.87 | 1.27 | 2.07 | 2.91 | 3.34 | 3.68 |
| HPV16/18 | Rennes | 49 | 2.03 | 0.67 | 0.83 | 1.24 | 2.01 | 2.85 | 3.27 | 3.58 |
| HPV16/18 | Rennes | 50 | 1.97 | 0.64 | 0.78 | 1.20 | 1.96 | 2.77 | 3.18 | 3.47 |
| HPV16/18 | Rennes | 51 | 1.92 | 0.60 | 0.75 | 1.16 | 1.91 | 2.71 | 3.12 | 3.40 |
| HPV16/18 | Rennes | 52 | 1.87 | 0.59 | 0.74 | 1.11 | 1.86 | 2.64 | 3.02 | 3.35 |
| HPV16/18 | Rennes | 53 | 1.81 | 0.55 | 0.70 | 1.07 | 1.80 | 2.57 | 2.96 | 3.22 |
| HPV16/18 | Rennes | 54 | 1.75 | 0.50 | 0.67 | 1.02 | 1.73 | 2.49 | 2.88 | 3.13 |
| HPV16/18 | Rennes | 55 | 1.70 | 0.46 | 0.65 | 0.99 | 1.69 | 2.43 | 2.79 | 3.05 |
| HPV16/18 | Rennes | 56 | 1.65 | 0.43 | 0.62 | 0.95 | 1.64 | 2.37 | 2.74 | 2.96 |
| HPV16/18 | Rennes | 57 | 1.62 | 0.38 | 0.60 | 0.92 | 1.60 | 2.33 | 2.68 | 2.96 |
| HPV16/18 | Rennes | 58 | 1.59 | 0.35 | 0.55 | 0.89 | 1.58 | 2.31 | 2.68 | 2.91 |
| HPV16/18 | Rennes | 59 | 1.57 | 0.31 | 0.52 | 0.87 | 1.56 | 2.29 | 2.67 | 2.92 |
| HPV16/18 | Rennes | 60 | 1.56 | 0.28 | 0.49 | 0.85 | 1.55 | 2.30 | 2.67 | 2.90 |
| HPV16/18 | Rennes | 61 | 1.55 | 0.24 | 0.46 | 0.84 | 1.54 | 2.30 | 2.67 | 2.88 |
| HPV16/18 | Rennes | 62 | 1.53 | 0.19 | 0.43 | 0.80 | 1.52 | 2.28 | 2.65 | 2.87 |
| HPV16/18 | Rennes | 63 | 1.50 | 0.16 | 0.40 | 0.77 | 1.49 | 2.26 | 2.63 | 2.89 |
| HPV16/18 | Rennes | 64 | 1.48 | 0.10 | 0.34 | 0.73 | 1.46 | 2.24 | 2.65 | 2.86 |
| HPV16/18 | Rennes | 65 | 1.44 | 0.05 | 0.26 | 0.69 | 1.44 | 2.21 | 2.61 | 2.84 |
| HPV16/18 | Rennes | 66 | 1.41 | -0.01 | 0.20 | 0.63 | 1.40 | 2.19 | 2.61 | 2.85 |

**Table S10** **Posterior difference in expected HR HPV cervical infection prevalence (in percentage points) in major French cities, between opportunistic and organised screening, stratified by type of test, city, and age. Table counterpart to Figure 3B.** ***(continued)***

| | | | Posterior distribution | | | | | | | |
|---|---|---|---|---|---|---|---|---|---|---|
| | | | | Percentiles | | | | | | |
| Virus | City | Age | Average | 0.01 | 0.025 | 0.1 | 0.5 | 0.9 | 0.975 | 0.99 |
| HPV16/18 | Strasbourg | 30 | 1.43 | -0.31 | 0.00 | 0.51 | 1.37 | 2.45 | 3.16 | 3.63 |
| HPV16/18 | Strasbourg | 31 | 1.54 | -0.25 | 0.07 | 0.58 | 1.48 | 2.59 | 3.35 | 3.79 |
| HPV16/18 | Strasbourg | 32 | 1.63 | -0.20 | 0.14 | 0.64 | 1.57 | 2.72 | 3.53 | 4.03 |
| HPV16/18 | Strasbourg | 33 | 1.71 | -0.15 | 0.19 | 0.69 | 1.64 | 2.83 | 3.67 | 4.22 |
| HPV16/18 | Strasbourg | 34 | 1.75 | -0.13 | 0.19 | 0.72 | 1.67 | 2.87 | 3.69 | 4.25 |
| HPV16/18 | Strasbourg | 35 | 1.74 | -0.08 | 0.19 | 0.73 | 1.66 | 2.85 | 3.67 | 4.26 |
| HPV16/18 | Strasbourg | 36 | 1.70 | -0.07 | 0.20 | 0.72 | 1.62 | 2.80 | 3.60 | 4.17 |
| HPV16/18 | Strasbourg | 37 | 1.66 | -0.06 | 0.20 | 0.70 | 1.58 | 2.72 | 3.49 | 4.09 |
| HPV16/18 | Strasbourg | 38 | 1.61 | -0.06 | 0.19 | 0.68 | 1.54 | 2.64 | 3.38 | 3.92 |
| HPV16/18 | Strasbourg | 39 | 1.56 | -0.07 | 0.20 | 0.65 | 1.49 | 2.57 | 3.29 | 3.80 |
| HPV16/18 | Strasbourg | 40 | 1.52 | -0.08 | 0.19 | 0.63 | 1.45 | 2.50 | 3.19 | 3.68 |
| HPV16/18 | Strasbourg | 41 | 1.47 | -0.08 | 0.17 | 0.61 | 1.41 | 2.42 | 3.10 | 3.59 |
| HPV16/18 | Strasbourg | 42 | 1.42 | -0.08 | 0.16 | 0.58 | 1.36 | 2.35 | 3.00 | 3.47 |
| HPV16/18 | Strasbourg | 43 | 1.37 | -0.08 | 0.15 | 0.57 | 1.32 | 2.28 | 2.90 | 3.37 |
| HPV16/18 | Strasbourg | 44 | 1.33 | -0.09 | 0.14 | 0.54 | 1.27 | 2.23 | 2.83 | 3.29 |
| HPV16/18 | Strasbourg | 45 | 1.30 | -0.10 | 0.12 | 0.51 | 1.24 | 2.17 | 2.78 | 3.22 |
| HPV16/18 | Strasbourg | 46 | 1.26 | -0.12 | 0.11 | 0.48 | 1.20 | 2.10 | 2.71 | 3.14 |
| HPV16/18 | Strasbourg | 47 | 1.22 | -0.13 | 0.11 | 0.46 | 1.17 | 2.04 | 2.63 | 3.11 |
| HPV16/18 | Strasbourg | 48 | 1.18 | -0.16 | 0.10 | 0.44 | 1.14 | 1.99 | 2.57 | 3.06 |
| HPV16/18 | Strasbourg | 49 | 1.15 | -0.17 | 0.08 | 0.42 | 1.10 | 1.93 | 2.50 | 2.98 |
| HPV16/18 | Strasbourg | 50 | 1.11 | -0.18 | 0.08 | 0.41 | 1.07 | 1.89 | 2.43 | 2.92 |
| HPV16/18 | Strasbourg | 51 | 1.08 | -0.16 | 0.06 | 0.39 | 1.04 | 1.85 | 2.38 | 2.76 |
| HPV16/18 | Strasbourg | 52 | 1.05 | -0.18 | 0.04 | 0.37 | 1.01 | 1.80 | 2.30 | 2.72 |
| HPV16/18 | Strasbourg | 53 | 1.01 | -0.18 | 0.02 | 0.34 | 0.97 | 1.74 | 2.23 | 2.65 |
| HPV16/18 | Strasbourg | 54 | 0.97 | -0.19 | 0.00 | 0.32 | 0.93 | 1.68 | 2.16 | 2.57 |
| HPV16/18 | Strasbourg | 55 | 0.94 | -0.21 | -0.01 | 0.30 | 0.90 | 1.63 | 2.12 | 2.52 |
| HPV16/18 | Strasbourg | 56 | 0.91 | -0.23 | -0.02 | 0.28 | 0.87 | 1.59 | 2.09 | 2.47 |
| HPV16/18 | Strasbourg | 57 | 0.89 | -0.25 | -0.04 | 0.26 | 0.85 | 1.55 | 2.03 | 2.41 |
| HPV16/18 | Strasbourg | 58 | 0.87 | -0.26 | -0.07 | 0.25 | 0.83 | 1.54 | 2.00 | 2.43 |

**Table S10 Posterior difference in expected HR HPV cervical infection prevalence (in percentage points) in major French cities, between opportunistic and organised screening, stratified by type of test, city, and age. Table counterpart to Figure 3B.** ***(continued)***

| | | | Posterior distribution | | | | | | | |
|---|---|---|---|---|---|---|---|---|---|---|
| | | | | Percentiles | | | | | | |
| Virus | City | Age | Average | 0.01 | 0.025 | 0.1 | 0.5 | 0.9 | 0.975 | 0.99 |
| HPV16/18 | Strasbourg | 59 | 0.85 | -0.27 | -0.09 | 0.23 | 0.81 | 1.53 | 1.99 | 2.41 |
| HPV16/18 | Strasbourg | 60 | 0.84 | -0.30 | -0.11 | 0.21 | 0.80 | 1.52 | 2.00 | 2.43 |
| HPV16/18 | Strasbourg | 61 | 0.83 | -0.34 | -0.13 | 0.20 | 0.79 | 1.52 | 2.00 | 2.39 |
| HPV16/18 | Strasbourg | 62 | 0.82 | -0.38 | -0.16 | 0.18 | 0.77 | 1.51 | 1.99 | 2.39 |
| HPV16/18 | Strasbourg | 63 | 0.80 | -0.45 | -0.18 | 0.16 | 0.76 | 1.50 | 1.97 | 2.40 |
| HPV16/18 | Strasbourg | 64 | 0.78 | -0.47 | -0.22 | 0.13 | 0.74 | 1.49 | 1.95 | 2.41 |
| HPV16/18 | Strasbourg | 65 | 0.76 | -0.50 | -0.26 | 0.11 | 0.72 | 1.48 | 1.95 | 2.37 |
| HPV16/18 | Strasbourg | 66 | 0.73 | -0.54 | -0.30 | 0.07 | 0.69 | 1.46 | 1.98 | 2.39 |
| HPV16/18 | Toulouse | 30 | 1.38 | -0.49 | -0.18 | 0.35 | 1.36 | 2.41 | 3.04 | 3.45 |
| HPV16/18 | Toulouse | 31 | 1.49 | -0.44 | -0.12 | 0.44 | 1.48 | 2.55 | 3.19 | 3.66 |
| HPV16/18 | Toulouse | 32 | 1.59 | -0.41 | -0.05 | 0.50 | 1.57 | 2.68 | 3.36 | 3.84 |
| HPV16/18 | Toulouse | 33 | 1.66 | -0.35 | 0.01 | 0.55 | 1.64 | 2.79 | 3.54 | 3.96 |
| HPV16/18 | Toulouse | 34 | 1.70 | -0.31 | 0.04 | 0.60 | 1.67 | 2.84 | 3.59 | 4.11 |
| HPV16/18 | Toulouse | 35 | 1.69 | -0.27 | 0.05 | 0.61 | 1.65 | 2.83 | 3.55 | 4.09 |
| HPV16/18 | Toulouse | 36 | 1.66 | -0.28 | 0.02 | 0.62 | 1.61 | 2.77 | 3.50 | 4.01 |
| HPV16/18 | Toulouse | 37 | 1.62 | -0.27 | 0.05 | 0.61 | 1.57 | 2.70 | 3.37 | 3.87 |
| HPV16/18 | Toulouse | 38 | 1.57 | -0.26 | 0.07 | 0.59 | 1.53 | 2.63 | 3.28 | 3.75 |
| HPV16/18 | Toulouse | 39 | 1.53 | -0.25 | 0.07 | 0.56 | 1.48 | 2.57 | 3.21 | 3.67 |
| HPV16/18 | Toulouse | 40 | 1.48 | -0.24 | 0.05 | 0.54 | 1.43 | 2.50 | 3.13 | 3.59 |
| HPV16/18 | Toulouse | 41 | 1.43 | -0.25 | 0.02 | 0.53 | 1.38 | 2.43 | 3.04 | 3.46 |
| HPV16/18 | Toulouse | 42 | 1.38 | -0.25 | 0.02 | 0.50 | 1.33 | 2.35 | 2.94 | 3.34 |
| HPV16/18 | Toulouse | 43 | 1.33 | -0.25 | 0.01 | 0.48 | 1.28 | 2.27 | 2.86 | 3.21 |
| HPV16/18 | Toulouse | 44 | 1.29 | -0.26 | 0.00 | 0.47 | 1.25 | 2.20 | 2.79 | 3.12 |
| HPV16/18 | Toulouse | 45 | 1.26 | -0.26 | -0.01 | 0.44 | 1.21 | 2.15 | 2.73 | 3.04 |
| HPV16/18 | Toulouse | 46 | 1.22 | -0.27 | -0.03 | 0.42 | 1.17 | 2.09 | 2.64 | 2.97 |
| HPV16/18 | Toulouse | 47 | 1.18 | -0.27 | -0.02 | 0.40 | 1.14 | 2.03 | 2.57 | 2.88 |
| HPV16/18 | Toulouse | 48 | 1.14 | -0.26 | -0.04 | 0.38 | 1.10 | 1.98 | 2.52 | 2.80 |
| HPV16/18 | Toulouse | 49 | 1.11 | -0.29 | -0.07 | 0.36 | 1.07 | 1.93 | 2.47 | 2.71 |
| HPV16/18 | Toulouse | 50 | 1.07 | -0.28 | -0.08 | 0.33 | 1.03 | 1.87 | 2.41 | 2.64 |

**Table S10 Posterior difference in expected HR HPV cervical infection prevalence (in percentage points) in major French cities, between opportunistic and organised screening, stratified by type of test, city, and age. Table counterpart to Figure 3B.** ***(continued)***

| | | | Posterior distribution | | | | | | | |
|---|---|---|---|---|---|---|---|---|---|---|
| | | | | Percentiles | | | | | | |
| Virus | City | Age | Average | 0.01 | 0.025 | 0.1 | 0.5 | 0.9 | 0.975 | 0.99 |
| HPV16/18 | Toulouse | 51 | 1.04 | -0.30 | -0.09 | 0.31 | 1.00 | 1.81 | 2.35 | 2.59 |
| HPV16/18 | Toulouse | 52 | 1.01 | -0.31 | -0.10 | 0.29 | 0.97 | 1.77 | 2.30 | 2.54 |
| HPV16/18 | Toulouse | 53 | 0.97 | -0.33 | -0.11 | 0.27 | 0.93 | 1.71 | 2.24 | 2.47 |
| HPV16/18 | Toulouse | 54 | 0.93 | -0.35 | -0.13 | 0.25 | 0.90 | 1.65 | 2.17 | 2.39 |
| HPV16/18 | Toulouse | 55 | 0.89 | -0.37 | -0.15 | 0.23 | 0.86 | 1.60 | 2.11 | 2.35 |
| HPV16/18 | Toulouse | 56 | 0.86 | -0.38 | -0.16 | 0.21 | 0.84 | 1.56 | 2.05 | 2.29 |
| HPV16/18 | Toulouse | 57 | 0.84 | -0.41 | -0.18 | 0.19 | 0.81 | 1.52 | 2.02 | 2.29 |
| HPV16/18 | Toulouse | 58 | 0.82 | -0.43 | -0.20 | 0.16 | 0.79 | 1.50 | 1.99 | 2.27 |
| HPV16/18 | Toulouse | 59 | 0.80 | -0.45 | -0.22 | 0.14 | 0.78 | 1.50 | 1.97 | 2.26 |
| HPV16/18 | Toulouse | 60 | 0.79 | -0.46 | -0.24 | 0.12 | 0.77 | 1.49 | 1.95 | 2.26 |
| HPV16/18 | Toulouse | 61 | 0.77 | -0.49 | -0.26 | 0.10 | 0.76 | 1.50 | 1.96 | 2.25 |
| HPV16/18 | Toulouse | 62 | 0.76 | -0.52 | -0.29 | 0.08 | 0.74 | 1.48 | 1.95 | 2.24 |
| HPV16/18 | Toulouse | 63 | 0.74 | -0.56 | -0.32 | 0.05 | 0.72 | 1.46 | 1.92 | 2.21 |
| HPV16/18 | Toulouse | 64 | 0.72 | -0.59 | -0.36 | 0.02 | 0.70 | 1.45 | 1.94 | 2.19 |
| HPV16/18 | Toulouse | 65 | 0.69 | -0.65 | -0.41 | -0.02 | 0.68 | 1.42 | 1.91 | 2.18 |
| HPV16/18 | Toulouse | 66 | 0.67 | -0.74 | -0.48 | -0.06 | 0.66 | 1.41 | 1.90 | 2.17 |
| Other genotypes | Bordeaux | 30 | 7.69 | 3.00 | 3.81 | 5.20 | 7.68 | 10.20 | 11.47 | 12.23 |
| Other genotypes | Bordeaux | 31 | 7.37 | 2.93 | 3.72 | 5.05 | 7.37 | 9.76 | 10.94 | 11.66 |
| Other genotypes | Bordeaux | 32 | 7.01 | 2.86 | 3.47 | 4.79 | 7.02 | 9.30 | 10.41 | 11.06 |
| Other genotypes | Bordeaux | 33 | 6.62 | 2.65 | 3.24 | 4.52 | 6.63 | 8.81 | 9.86 | 10.48 |
| Other genotypes | Bordeaux | 34 | 6.28 | 2.53 | 3.05 | 4.26 | 6.28 | 8.37 | 9.37 | 9.97 |
| Other genotypes | Bordeaux | 35 | 5.98 | 2.35 | 2.94 | 4.02 | 5.97 | 7.97 | 8.99 | 9.58 |
| Other genotypes | Bordeaux | 36 | 5.70 | 2.21 | 2.79 | 3.81 | 5.69 | 7.62 | 8.67 | 9.19 |
| Other genotypes | Bordeaux | 37 | 5.47 | 2.06 | 2.64 | 3.64 | 5.44 | 7.33 | 8.34 | 8.91 |
| Other genotypes | Bordeaux | 38 | 5.25 | 1.92 | 2.50 | 3.46 | 5.22 | 7.04 | 8.02 | 8.64 |
| Other genotypes | Bordeaux | 39 | 5.03 | 1.80 | 2.34 | 3.29 | 5.02 | 6.79 | 7.76 | 8.37 |
| Other genotypes | Bordeaux | 40 | 4.81 | 1.61 | 2.21 | 3.12 | 4.80 | 6.51 | 7.49 | 8.01 |
| Other genotypes | Bordeaux | 41 | 4.61 | 1.53 | 2.07 | 2.97 | 4.62 | 6.26 | 7.21 | 7.76 |
| Other genotypes | Bordeaux | 42 | 4.44 | 1.43 | 1.95 | 2.81 | 4.45 | 6.05 | 6.99 | 7.54 |

**Table S10** **Posterior difference in expected HR HPV cervical infection prevalence (in percentage points) in major French cities, between opportunistic and organised screening, stratified by type of test, city, and age. Table counterpart to Figure 3B.** ***(continued)***

| | | | Posterior distribution | | | | | | | |
|---|---|---|---|---|---|---|---|---|---|---|
| | | | | Percentiles | | | | | | |
| Virus | City | Age | Average | 0.01 | 0.025 | 0.1 | 0.5 | 0.9 | 0.975 | 0.99 |
| Other genotypes | Bordeaux | 43 | 4.29 | 1.31 | 1.85 | 2.71 | 4.31 | 5.88 | 6.79 | 7.35 |
| Other genotypes | Bordeaux | 44 | 4.15 | 1.23 | 1.74 | 2.59 | 4.15 | 5.71 | 6.58 | 7.16 |
| Other genotypes | Bordeaux | 45 | 4.02 | 1.13 | 1.65 | 2.49 | 4.03 | 5.57 | 6.40 | 6.99 |
| Other genotypes | Bordeaux | 46 | 3.91 | 1.03 | 1.54 | 2.38 | 3.91 | 5.44 | 6.24 | 6.87 |
| Other genotypes | Bordeaux | 47 | 3.81 | 0.96 | 1.45 | 2.31 | 3.80 | 5.31 | 6.14 | 6.74 |
| Other genotypes | Bordeaux | 48 | 3.71 | 0.89 | 1.37 | 2.21 | 3.70 | 5.20 | 6.09 | 6.61 |
| Other genotypes | Bordeaux | 49 | 3.62 | 0.81 | 1.28 | 2.13 | 3.62 | 5.10 | 5.96 | 6.58 |
| Other genotypes | Bordeaux | 50 | 3.53 | 0.72 | 1.19 | 2.05 | 3.53 | 4.99 | 5.86 | 6.43 |
| Other genotypes | Bordeaux | 51 | 3.43 | 0.66 | 1.11 | 1.97 | 3.44 | 4.89 | 5.69 | 6.27 |
| Other genotypes | Bordeaux | 52 | 3.31 | 0.57 | 1.02 | 1.86 | 3.32 | 4.75 | 5.61 | 6.09 |
| Other genotypes | Bordeaux | 53 | 3.17 | 0.49 | 0.93 | 1.73 | 3.19 | 4.57 | 5.39 | 5.90 |
| Other genotypes | Bordeaux | 54 | 3.02 | 0.43 | 0.80 | 1.60 | 3.04 | 4.38 | 5.21 | 5.70 |
| Other genotypes | Bordeaux | 55 | 2.87 | 0.34 | 0.68 | 1.48 | 2.89 | 4.21 | 4.99 | 5.48 |
| Other genotypes | Bordeaux | 56 | 2.72 | 0.23 | 0.58 | 1.36 | 2.73 | 4.03 | 4.81 | 5.28 |
| Other genotypes | Bordeaux | 57 | 2.57 | 0.08 | 0.48 | 1.25 | 2.60 | 3.87 | 4.61 | 5.10 |
| Other genotypes | Bordeaux | 58 | 2.44 | 0.04 | 0.38 | 1.13 | 2.46 | 3.74 | 4.42 | 4.92 |
| Other genotypes | Bordeaux | 59 | 2.33 | -0.05 | 0.26 | 1.02 | 2.34 | 3.62 | 4.27 | 4.73 |
| Other genotypes | Bordeaux | 60 | 2.23 | -0.18 | 0.17 | 0.92 | 2.25 | 3.52 | 4.18 | 4.58 |
| Other genotypes | Bordeaux | 61 | 2.16 | -0.29 | 0.06 | 0.83 | 2.18 | 3.45 | 4.13 | 4.47 |
| Other genotypes | Bordeaux | 62 | 2.09 | -0.37 | 0.00 | 0.75 | 2.11 | 3.41 | 4.10 | 4.45 |
| Other genotypes | Bordeaux | 63 | 2.05 | -0.48 | -0.06 | 0.68 | 2.06 | 3.39 | 4.08 | 4.45 |
| Other genotypes | Bordeaux | 64 | 2.02 | -0.58 | -0.15 | 0.62 | 2.02 | 3.40 | 4.10 | 4.51 |
| Other genotypes | Bordeaux | 65 | 2.00 | -0.70 | -0.28 | 0.55 | 2.00 | 3.41 | 4.16 | 4.55 |
| Other genotypes | Bordeaux | 66 | 1.98 | -0.87 | -0.39 | 0.47 | 1.99 | 3.47 | 4.25 | 4.68 |
| Other genotypes | Lille | 30 | 7.89 | 3.01 | 3.86 | 5.07 | 7.80 | 10.81 | 12.58 | 13.59 |
| Other genotypes | Lille | 31 | 7.56 | 2.96 | 3.75 | 4.88 | 7.46 | 10.35 | 12.08 | 13.10 |
| Other genotypes | Lille | 32 | 7.18 | 2.88 | 3.57 | 4.64 | 7.07 | 9.90 | 11.52 | 12.51 |
| Other genotypes | Lille | 33 | 6.79 | 2.80 | 3.35 | 4.35 | 6.69 | 9.35 | 10.98 | 11.96 |
| Other genotypes | Lille | 34 | 6.43 | 2.64 | 3.13 | 4.09 | 6.34 | 8.91 | 10.47 | 11.40 |

**Table S10 Posterior difference in expected HR HPV cervical infection prevalence (in percentage points) in major French cities, between opportunistic and organised screening, stratified by type of test, city, and age. Table counterpart to Figure 3B.** ***(continued)***

| | | | Posterior distribution | | | | | | | |
|---|---|---|---|---|---|---|---|---|---|---|
| | | | | Percentiles | | | | | | |
| Virus | City | Age | Average | 0.01 | 0.025 | 0.1 | 0.5 | 0.9 | 0.975 | 0.99 |
| Other genotypes | Lille | 35 | 6.13 | 2.48 | 2.95 | 3.90 | 6.03 | 8.52 | 10.06 | 11.01 |
| Other genotypes | Lille | 36 | 5.85 | 2.34 | 2.77 | 3.70 | 5.76 | 8.13 | 9.61 | 10.50 |
| Other genotypes | Lille | 37 | 5.61 | 2.20 | 2.64 | 3.54 | 5.52 | 7.83 | 9.27 | 10.11 |
| Other genotypes | Lille | 38 | 5.39 | 2.07 | 2.50 | 3.36 | 5.29 | 7.53 | 8.97 | 9.70 |
| Other genotypes | Lille | 39 | 5.17 | 1.95 | 2.39 | 3.22 | 5.08 | 7.29 | 8.65 | 9.43 |
| Other genotypes | Lille | 40 | 4.95 | 1.78 | 2.21 | 3.07 | 4.86 | 7.02 | 8.28 | 9.09 |
| Other genotypes | Lille | 41 | 4.75 | 1.68 | 2.08 | 2.92 | 4.67 | 6.76 | 8.01 | 8.84 |
| Other genotypes | Lille | 42 | 4.58 | 1.60 | 1.96 | 2.79 | 4.50 | 6.53 | 7.74 | 8.52 |
| Other genotypes | Lille | 43 | 4.43 | 1.50 | 1.88 | 2.68 | 4.34 | 6.33 | 7.54 | 8.32 |
| Other genotypes | Lille | 44 | 4.29 | 1.42 | 1.80 | 2.56 | 4.20 | 6.15 | 7.36 | 8.08 |
| Other genotypes | Lille | 45 | 4.16 | 1.32 | 1.71 | 2.47 | 4.08 | 6.01 | 7.20 | 7.89 |
| Other genotypes | Lille | 46 | 4.05 | 1.24 | 1.62 | 2.38 | 3.97 | 5.84 | 7.00 | 7.81 |
| Other genotypes | Lille | 47 | 3.95 | 1.19 | 1.52 | 2.29 | 3.87 | 5.71 | 6.85 | 7.68 |
| Other genotypes | Lille | 48 | 3.85 | 1.12 | 1.44 | 2.22 | 3.78 | 5.61 | 6.76 | 7.52 |
| Other genotypes | Lille | 49 | 3.76 | 1.05 | 1.35 | 2.15 | 3.69 | 5.51 | 6.70 | 7.33 |
| Other genotypes | Lille | 50 | 3.67 | 0.93 | 1.30 | 2.07 | 3.59 | 5.41 | 6.55 | 7.21 |
| Other genotypes | Lille | 51 | 3.58 | 0.85 | 1.21 | 1.97 | 3.51 | 5.29 | 6.44 | 7.09 |
| Other genotypes | Lille | 52 | 3.46 | 0.75 | 1.10 | 1.87 | 3.39 | 5.17 | 6.32 | 6.88 |
| Other genotypes | Lille | 53 | 3.33 | 0.65 | 1.02 | 1.78 | 3.26 | 5.00 | 6.16 | 6.69 |
| Other genotypes | Lille | 54 | 3.17 | 0.52 | 0.90 | 1.65 | 3.11 | 4.80 | 5.89 | 6.42 |
| Other genotypes | Lille | 55 | 3.02 | 0.37 | 0.81 | 1.53 | 2.96 | 4.61 | 5.69 | 6.26 |
| Other genotypes | Lille | 56 | 2.88 | 0.25 | 0.70 | 1.40 | 2.83 | 4.40 | 5.48 | 5.95 |
| Other genotypes | Lille | 57 | 2.73 | 0.15 | 0.58 | 1.29 | 2.69 | 4.23 | 5.25 | 5.70 |
| Other genotypes | Lille | 58 | 2.60 | 0.07 | 0.49 | 1.17 | 2.56 | 4.08 | 5.11 | 5.55 |
| Other genotypes | Lille | 59 | 2.49 | -0.01 | 0.39 | 1.07 | 2.45 | 3.95 | 4.97 | 5.41 |
| Other genotypes | Lille | 60 | 2.40 | -0.07 | 0.31 | 0.99 | 2.36 | 3.85 | 4.82 | 5.33 |
| Other genotypes | Lille | 61 | 2.32 | -0.13 | 0.25 | 0.92 | 2.29 | 3.79 | 4.75 | 5.23 |
| Other genotypes | Lille | 62 | 2.27 | -0.21 | 0.16 | 0.86 | 2.23 | 3.75 | 4.72 | 5.17 |
| Other genotypes | Lille | 63 | 2.23 | -0.29 | 0.09 | 0.81 | 2.18 | 3.73 | 4.69 | 5.16 |

**Table S10 Posterior difference in expected HR HPV cervical infection prevalence (in percentage points) in major French cities, between opportunistic and organised screening, stratified by type of test, city, and age. Table counterpart to Figure 3B.** ***(continued)***

| | | | Posterior distribution | | | | | | | |
|---|---|---|---|---|---|---|---|---|---|---|
| | | | | Percentiles | | | | | | |
| Virus | City | Age | Average | 0.01 | 0.025 | 0.1 | 0.5 | 0.9 | 0.975 | 0.99 |
| Other genotypes | Lille | 64 | 2.20 | -0.40 | 0.01 | 0.73 | 2.15 | 3.75 | 4.71 | 5.19 |
| Other genotypes | Lille | 65 | 2.19 | -0.58 | -0.08 | 0.66 | 2.13 | 3.78 | 4.79 | 5.31 |
| Other genotypes | Lille | 66 | 2.18 | -0.70 | -0.17 | 0.60 | 2.11 | 3.84 | 4.87 | 5.49 |
| Other genotypes | Lyon | 30 | 8.19 | 2.46 | 3.49 | 5.07 | 8.08 | 11.43 | 13.41 | 14.54 |
| Other genotypes | Lyon | 31 | 7.87 | 2.49 | 3.44 | 4.86 | 7.74 | 10.98 | 12.94 | 13.95 |
| Other genotypes | Lyon | 32 | 7.50 | 2.35 | 3.22 | 4.62 | 7.37 | 10.50 | 12.36 | 13.43 |
| Other genotypes | Lyon | 33 | 7.10 | 2.19 | 2.98 | 4.38 | 6.95 | 9.99 | 11.71 | 12.79 |
| Other genotypes | Lyon | 34 | 6.74 | 2.05 | 2.79 | 4.11 | 6.62 | 9.49 | 11.19 | 12.21 |
| Other genotypes | Lyon | 35 | 6.44 | 1.89 | 2.65 | 3.89 | 6.31 | 9.08 | 10.71 | 11.76 |
| Other genotypes | Lyon | 36 | 6.15 | 1.75 | 2.49 | 3.70 | 6.01 | 8.74 | 10.37 | 11.36 |
| Other genotypes | Lyon | 37 | 5.90 | 1.68 | 2.37 | 3.53 | 5.77 | 8.42 | 10.08 | 10.97 |
| Other genotypes | Lyon | 38 | 5.67 | 1.60 | 2.25 | 3.37 | 5.53 | 8.10 | 9.67 | 10.64 |
| Other genotypes | Lyon | 39 | 5.44 | 1.47 | 2.10 | 3.21 | 5.32 | 7.81 | 9.37 | 10.28 |
| Other genotypes | Lyon | 40 | 5.21 | 1.36 | 1.98 | 3.03 | 5.09 | 7.54 | 9.02 | 9.90 |
| Other genotypes | Lyon | 41 | 5.00 | 1.32 | 1.80 | 2.89 | 4.88 | 7.26 | 8.70 | 9.67 |
| Other genotypes | Lyon | 42 | 4.82 | 1.12 | 1.69 | 2.74 | 4.69 | 7.02 | 8.45 | 9.38 |
| Other genotypes | Lyon | 43 | 4.66 | 1.02 | 1.59 | 2.64 | 4.54 | 6.82 | 8.24 | 9.22 |
| Other genotypes | Lyon | 44 | 4.51 | 0.97 | 1.51 | 2.53 | 4.39 | 6.63 | 8.02 | 9.00 |
| Other genotypes | Lyon | 45 | 4.37 | 0.84 | 1.40 | 2.43 | 4.26 | 6.46 | 7.84 | 8.78 |
| Other genotypes | Lyon | 46 | 4.25 | 0.79 | 1.32 | 2.34 | 4.14 | 6.30 | 7.68 | 8.63 |
| Other genotypes | Lyon | 47 | 4.14 | 0.72 | 1.23 | 2.23 | 4.03 | 6.17 | 7.53 | 8.46 |
| Other genotypes | Lyon | 48 | 4.04 | 0.63 | 1.18 | 2.17 | 3.93 | 6.06 | 7.39 | 8.33 |
| Other genotypes | Lyon | 49 | 3.94 | 0.51 | 1.10 | 2.07 | 3.84 | 5.90 | 7.28 | 8.17 |
| Other genotypes | Lyon | 50 | 3.84 | 0.39 | 1.04 | 1.98 | 3.75 | 5.78 | 7.11 | 8.03 |
| Other genotypes | Lyon | 51 | 3.73 | 0.34 | 0.95 | 1.90 | 3.63 | 5.66 | 6.95 | 7.91 |
| Other genotypes | Lyon | 52 | 3.60 | 0.25 | 0.86 | 1.80 | 3.50 | 5.49 | 6.77 | 7.69 |
| Other genotypes | Lyon | 53 | 3.46 | 0.16 | 0.71 | 1.69 | 3.37 | 5.34 | 6.58 | 7.39 |
| Other genotypes | Lyon | 54 | 3.29 | 0.08 | 0.59 | 1.54 | 3.19 | 5.15 | 6.39 | 7.18 |
| Other genotypes | Lyon | 55 | 3.13 | 0.01 | 0.49 | 1.41 | 3.04 | 4.93 | 6.17 | 6.86 |

**Table S10 Posterior difference in expected HR HPV cervical infection prevalence (in percentage points) in major French cities, between opportunistic and organised screening, stratified by type of test, city, and age. Table counterpart to Figure 3B. *(continued)***

| Virus | City | Age | Average | 0.01 | 0.025 | 0.1 | 0.5 | 0.9 | 0.975 | 0.99 |
|---|---|---|---|---|---|---|---|---|---|---|
| | | | Posterior distribution | | | | | | | |
| | | | | Percentiles | | | | | | |
| Other genotypes | Lyon | 56 | 2.98 | -0.09 | 0.37 | 1.28 | 2.90 | 4.74 | 5.95 | 6.55 |
| Other genotypes | Lyon | 57 | 2.82 | -0.18 | 0.24 | 1.17 | 2.74 | 4.57 | 5.69 | 6.28 |
| Other genotypes | Lyon | 58 | 2.67 | -0.28 | 0.12 | 1.05 | 2.61 | 4.37 | 5.52 | 6.09 |
| Other genotypes | Lyon | 59 | 2.55 | -0.37 | 0.03 | 0.93 | 2.49 | 4.21 | 5.31 | 6.06 |
| Other genotypes | Lyon | 60 | 2.45 | -0.45 | -0.03 | 0.84 | 2.39 | 4.10 | 5.15 | 5.89 |
| Other genotypes | Lyon | 61 | 2.36 | -0.54 | -0.09 | 0.78 | 2.31 | 4.02 | 5.03 | 5.77 |
| Other genotypes | Lyon | 62 | 2.30 | -0.65 | -0.17 | 0.69 | 2.26 | 3.97 | 4.99 | 5.67 |
| Other genotypes | Lyon | 63 | 2.25 | -0.73 | -0.28 | 0.60 | 2.20 | 3.94 | 5.03 | 5.65 |
| Other genotypes | Lyon | 64 | 2.22 | -0.85 | -0.37 | 0.54 | 2.16 | 3.93 | 5.03 | 5.65 |
| Other genotypes | Lyon | 65 | 2.19 | -1.01 | -0.48 | 0.47 | 2.15 | 3.92 | 5.08 | 5.79 |
| Other genotypes | Lyon | 66 | 2.18 | -1.21 | -0.62 | 0.40 | 2.15 | 3.98 | 5.16 | 5.90 |
| Other genotypes | Marseille | 30 | 5.92 | 0.49 | 1.51 | 3.14 | 6.01 | 8.61 | 9.88 | 10.50 |
| Other genotypes | Marseille | 31 | 5.67 | 0.51 | 1.47 | 3.01 | 5.75 | 8.21 | 9.33 | 10.02 |
| Other genotypes | Marseille | 32 | 5.38 | 0.54 | 1.35 | 2.84 | 5.44 | 7.79 | 8.88 | 9.54 |
| Other genotypes | Marseille | 33 | 5.07 | 0.53 | 1.18 | 2.66 | 5.12 | 7.35 | 8.36 | 9.03 |
| Other genotypes | Marseille | 34 | 4.79 | 0.46 | 1.08 | 2.47 | 4.86 | 6.96 | 7.86 | 8.56 |
| Other genotypes | Marseille | 35 | 4.54 | 0.34 | 0.98 | 2.30 | 4.63 | 6.63 | 7.56 | 8.17 |
| Other genotypes | Marseille | 36 | 4.31 | 0.21 | 0.87 | 2.13 | 4.41 | 6.35 | 7.23 | 7.83 |
| Other genotypes | Marseille | 37 | 4.11 | 0.14 | 0.78 | 2.02 | 4.19 | 6.09 | 6.93 | 7.53 |
| Other genotypes | Marseille | 38 | 3.93 | -0.01 | 0.65 | 1.88 | 4.02 | 5.85 | 6.66 | 7.26 |
| Other genotypes | Marseille | 39 | 3.75 | -0.08 | 0.54 | 1.75 | 3.82 | 5.62 | 6.44 | 6.92 |
| Other genotypes | Marseille | 40 | 3.56 | -0.13 | 0.45 | 1.63 | 3.63 | 5.38 | 6.18 | 6.63 |
| Other genotypes | Marseille | 41 | 3.39 | -0.22 | 0.36 | 1.52 | 3.47 | 5.17 | 5.95 | 6.39 |
| Other genotypes | Marseille | 42 | 3.25 | -0.32 | 0.25 | 1.42 | 3.31 | 5.00 | 5.77 | 6.22 |
| Other genotypes | Marseille | 43 | 3.11 | -0.41 | 0.16 | 1.33 | 3.17 | 4.83 | 5.59 | 6.03 |
| Other genotypes | Marseille | 44 | 2.99 | -0.49 | 0.04 | 1.21 | 3.07 | 4.68 | 5.42 | 5.90 |
| Other genotypes | Marseille | 45 | 2.88 | -0.58 | -0.03 | 1.11 | 2.96 | 4.56 | 5.30 | 5.69 |
| Other genotypes | Marseille | 46 | 2.78 | -0.60 | -0.11 | 1.00 | 2.84 | 4.44 | 5.20 | 5.58 |
| Other genotypes | Marseille | 47 | 2.68 | -0.69 | -0.21 | 0.92 | 2.75 | 4.34 | 5.10 | 5.48 |

**Table S10 Posterior difference in expected HR HPV cervical infection prevalence (in percentage points) in major French cities, between opportunistic and organised screening, stratified by type of test, city, and age. Table counterpart to Figure 3B.** ***(continued)***

| | | | Posterior distribution | | | | | | | |
|---|---|---|---|---|---|---|---|---|---|---|
| | | | | Percentiles | | | | | | |
| Virus | City | Age | Average | 0.01 | 0.025 | 0.1 | 0.5 | 0.9 | 0.975 | 0.99 |
| Other genotypes | Marseille | 48 | 2.59 | -0.79 | -0.31 | 0.85 | 2.65 | 4.23 | 5.00 | 5.34 |
| Other genotypes | Marseille | 49 | 2.50 | -0.86 | -0.39 | 0.76 | 2.58 | 4.13 | 4.90 | 5.27 |
| Other genotypes | Marseille | 50 | 2.41 | -0.97 | -0.43 | 0.68 | 2.49 | 4.03 | 4.80 | 5.17 |
| Other genotypes | Marseille | 51 | 2.32 | -1.07 | -0.49 | 0.61 | 2.39 | 3.92 | 4.72 | 5.06 |
| Other genotypes | Marseille | 52 | 2.21 | -1.14 | -0.56 | 0.52 | 2.29 | 3.80 | 4.55 | 4.90 |
| Other genotypes | Marseille | 53 | 2.09 | -1.22 | -0.61 | 0.40 | 2.17 | 3.66 | 4.40 | 4.77 |
| Other genotypes | Marseille | 54 | 1.95 | -1.30 | -0.69 | 0.30 | 2.02 | 3.49 | 4.25 | 4.63 |
| Other genotypes | Marseille | 55 | 1.81 | -1.41 | -0.76 | 0.18 | 1.88 | 3.31 | 4.05 | 4.46 |
| Other genotypes | Marseille | 56 | 1.68 | -1.45 | -0.88 | 0.08 | 1.75 | 3.16 | 3.90 | 4.28 |
| Other genotypes | Marseille | 57 | 1.55 | -1.50 | -1.00 | -0.04 | 1.62 | 3.01 | 3.73 | 4.10 |
| Other genotypes | Marseille | 58 | 1.43 | -1.59 | -1.08 | -0.15 | 1.50 | 2.88 | 3.56 | 3.94 |
| Other genotypes | Marseille | 59 | 1.32 | -1.66 | -1.18 | -0.24 | 1.40 | 2.76 | 3.41 | 3.76 |
| Other genotypes | Marseille | 60 | 1.23 | -1.78 | -1.26 | -0.31 | 1.31 | 2.68 | 3.30 | 3.65 |
| Other genotypes | Marseille | 61 | 1.16 | -1.86 | -1.33 | -0.40 | 1.23 | 2.61 | 3.22 | 3.65 |
| Other genotypes | Marseille | 62 | 1.09 | -1.96 | -1.45 | -0.49 | 1.18 | 2.56 | 3.20 | 3.59 |
| Other genotypes | Marseille | 63 | 1.04 | -2.12 | -1.57 | -0.56 | 1.12 | 2.54 | 3.21 | 3.59 |
| Other genotypes | Marseille | 64 | 0.99 | -2.24 | -1.70 | -0.64 | 1.07 | 2.53 | 3.22 | 3.61 |
| Other genotypes | Marseille | 65 | 0.95 | -2.41 | -1.86 | -0.74 | 1.04 | 2.55 | 3.29 | 3.66 |
| Other genotypes | Marseille | 66 | 0.91 | -2.68 | -2.05 | -0.85 | 1.02 | 2.57 | 3.36 | 3.83 |
| Other genotypes | Montpellier | 30 | 7.76 | 2.45 | 3.25 | 4.88 | 7.72 | 10.70 | 12.56 | 13.97 |
| Other genotypes | Montpellier | 31 | 7.45 | 2.45 | 3.16 | 4.68 | 7.42 | 10.28 | 12.02 | 13.44 |
| Other genotypes | Montpellier | 32 | 7.10 | 2.35 | 3.00 | 4.45 | 7.02 | 9.84 | 11.44 | 12.81 |
| Other genotypes | Montpellier | 33 | 6.71 | 2.22 | 2.81 | 4.17 | 6.64 | 9.35 | 10.86 | 12.21 |
| Other genotypes | Montpellier | 34 | 6.37 | 1.95 | 2.64 | 3.91 | 6.29 | 8.91 | 10.39 | 11.65 |
| Other genotypes | Montpellier | 35 | 6.07 | 1.81 | 2.52 | 3.71 | 6.00 | 8.49 | 10.03 | 11.02 |
| Other genotypes | Montpellier | 36 | 5.80 | 1.66 | 2.34 | 3.53 | 5.72 | 8.15 | 9.59 | 10.59 |
| Other genotypes | Montpellier | 37 | 5.56 | 1.52 | 2.18 | 3.36 | 5.48 | 7.84 | 9.26 | 10.22 |
| Other genotypes | Montpellier | 38 | 5.33 | 1.39 | 2.06 | 3.18 | 5.26 | 7.57 | 8.87 | 9.92 |
| Other genotypes | Montpellier | 39 | 5.12 | 1.30 | 1.96 | 3.03 | 5.05 | 7.32 | 8.54 | 9.52 |

**Table S10 Posterior difference in expected HR HPV cervical infection prevalence (in percentage points) in major French cities, between opportunistic and organised screening, stratified by type of test, city, and age. Table counterpart to Figure 3B.** ***(continued)***

| | | | Posterior distribution | | | | | | | |
|---|---|---|---|---|---|---|---|---|---|---|
| | | | | Percentiles | | | | | | |
| Virus | City | Age | Average | 0.01 | 0.025 | 0.1 | 0.5 | 0.9 | 0.975 | 0.99 |
| Other genotypes | Montpellier | 40 | 4.89 | 1.16 | 1.80 | 2.86 | 4.84 | 7.02 | 8.33 | 9.16 |
| Other genotypes | Montpellier | 41 | 4.69 | 1.05 | 1.68 | 2.72 | 4.65 | 6.77 | 8.04 | 8.85 |
| Other genotypes | Montpellier | 42 | 4.52 | 0.96 | 1.54 | 2.59 | 4.48 | 6.57 | 7.80 | 8.56 |
| Other genotypes | Montpellier | 43 | 4.36 | 0.81 | 1.46 | 2.50 | 4.31 | 6.38 | 7.58 | 8.30 |
| Other genotypes | Montpellier | 44 | 4.22 | 0.75 | 1.35 | 2.38 | 4.18 | 6.19 | 7.41 | 8.11 |
| Other genotypes | Montpellier | 45 | 4.09 | 0.65 | 1.26 | 2.28 | 4.05 | 6.02 | 7.22 | 7.94 |
| Other genotypes | Montpellier | 46 | 3.97 | 0.53 | 1.17 | 2.19 | 3.94 | 5.90 | 7.03 | 7.78 |
| Other genotypes | Montpellier | 47 | 3.86 | 0.46 | 1.09 | 2.12 | 3.82 | 5.74 | 6.90 | 7.72 |
| Other genotypes | Montpellier | 48 | 3.76 | 0.38 | 1.01 | 2.02 | 3.72 | 5.62 | 6.78 | 7.54 |
| Other genotypes | Montpellier | 49 | 3.67 | 0.34 | 0.94 | 1.94 | 3.62 | 5.52 | 6.65 | 7.33 |
| Other genotypes | Montpellier | 50 | 3.57 | 0.29 | 0.86 | 1.84 | 3.53 | 5.40 | 6.50 | 7.20 |
| Other genotypes | Montpellier | 51 | 3.47 | 0.17 | 0.77 | 1.73 | 3.41 | 5.30 | 6.32 | 7.06 |
| Other genotypes | Montpellier | 52 | 3.34 | 0.09 | 0.69 | 1.63 | 3.29 | 5.14 | 6.16 | 6.95 |
| Other genotypes | Montpellier | 53 | 3.20 | 0.02 | 0.58 | 1.52 | 3.16 | 4.99 | 6.03 | 6.74 |
| Other genotypes | Montpellier | 54 | 3.04 | -0.05 | 0.46 | 1.37 | 3.00 | 4.79 | 5.84 | 6.53 |
| Other genotypes | Montpellier | 55 | 2.89 | -0.17 | 0.34 | 1.26 | 2.85 | 4.59 | 5.64 | 6.30 |
| Other genotypes | Montpellier | 56 | 2.74 | -0.28 | 0.24 | 1.14 | 2.70 | 4.40 | 5.43 | 6.11 |
| Other genotypes | Montpellier | 57 | 2.58 | -0.35 | 0.14 | 1.02 | 2.54 | 4.21 | 5.19 | 5.83 |
| Other genotypes | Montpellier | 58 | 2.45 | -0.43 | 0.02 | 0.92 | 2.41 | 4.04 | 5.00 | 5.65 |
| Other genotypes | Montpellier | 59 | 2.32 | -0.50 | -0.04 | 0.81 | 2.29 | 3.91 | 4.90 | 5.48 |
| Other genotypes | Montpellier | 60 | 2.23 | -0.60 | -0.10 | 0.72 | 2.19 | 3.79 | 4.77 | 5.30 |
| Other genotypes | Montpellier | 61 | 2.14 | -0.67 | -0.20 | 0.63 | 2.11 | 3.69 | 4.74 | 5.19 |
| Other genotypes | Montpellier | 62 | 2.08 | -0.75 | -0.29 | 0.57 | 2.06 | 3.62 | 4.65 | 5.20 |
| Other genotypes | Montpellier | 63 | 2.03 | -0.85 | -0.41 | 0.50 | 2.02 | 3.59 | 4.59 | 5.16 |
| Other genotypes | Montpellier | 64 | 1.99 | -0.96 | -0.50 | 0.41 | 1.98 | 3.59 | 4.57 | 5.18 |
| Other genotypes | Montpellier | 65 | 1.97 | -1.12 | -0.57 | 0.32 | 1.96 | 3.61 | 4.64 | 5.31 |
| Other genotypes | Montpellier | 66 | 1.95 | -1.35 | -0.71 | 0.22 | 1.93 | 3.68 | 4.73 | 5.39 |
| Other genotypes | Nantes | 30 | 7.93 | 3.25 | 3.86 | 5.21 | 7.71 | 11.02 | 13.12 | 14.11 |
| Other genotypes | Nantes | 31 | 7.59 | 3.09 | 3.78 | 5.00 | 7.38 | 10.53 | 12.56 | 13.54 |

**Table S10** **Posterior difference in expected HR HPV cervical infection prevalence (in percentage points) in major French cities, between opportunistic and organised screening, stratified by type of test, city, and age. Table counterpart to Figure 3B.** ***(continued)***

| | | | Posterior distribution | | | | | | | |
|---|---|---|---|---|---|---|---|---|---|---|
| | | | | Percentiles | | | | | | |
| Virus | City | Age | Average | 0.01 | 0.025 | 0.1 | 0.5 | 0.9 | 0.975 | 0.99 |
| Other genotypes | Nantes | 32 | 7.21 | 3.02 | 3.61 | 4.75 | 7.00 | 10.02 | 12.02 | 13.01 |
| Other genotypes | Nantes | 33 | 6.81 | 2.76 | 3.40 | 4.47 | 6.59 | 9.49 | 11.36 | 12.37 |
| Other genotypes | Nantes | 34 | 6.45 | 2.55 | 3.23 | 4.21 | 6.25 | 9.02 | 10.74 | 11.74 |
| Other genotypes | Nantes | 35 | 6.15 | 2.41 | 3.06 | 3.98 | 5.97 | 8.62 | 10.29 | 11.19 |
| Other genotypes | Nantes | 36 | 5.87 | 2.20 | 2.91 | 3.76 | 5.70 | 8.21 | 9.87 | 10.73 |
| Other genotypes | Nantes | 37 | 5.63 | 2.02 | 2.79 | 3.60 | 5.46 | 7.92 | 9.55 | 10.36 |
| Other genotypes | Nantes | 38 | 5.41 | 1.89 | 2.61 | 3.42 | 5.24 | 7.62 | 9.22 | 10.02 |
| Other genotypes | Nantes | 39 | 5.19 | 1.73 | 2.46 | 3.28 | 5.03 | 7.31 | 8.87 | 9.62 |
| Other genotypes | Nantes | 40 | 4.97 | 1.63 | 2.36 | 3.11 | 4.81 | 7.03 | 8.60 | 9.27 |
| Other genotypes | Nantes | 41 | 4.77 | 1.54 | 2.21 | 2.96 | 4.61 | 6.75 | 8.28 | 8.96 |
| Other genotypes | Nantes | 42 | 4.60 | 1.46 | 2.10 | 2.85 | 4.45 | 6.54 | 8.04 | 8.66 |
| Other genotypes | Nantes | 43 | 4.45 | 1.38 | 1.97 | 2.73 | 4.29 | 6.34 | 7.79 | 8.50 |
| Other genotypes | Nantes | 44 | 4.32 | 1.29 | 1.90 | 2.64 | 4.15 | 6.18 | 7.55 | 8.30 |
| Other genotypes | Nantes | 45 | 4.19 | 1.27 | 1.79 | 2.53 | 4.03 | 6.03 | 7.37 | 8.06 |
| Other genotypes | Nantes | 46 | 4.08 | 1.23 | 1.69 | 2.45 | 3.94 | 5.91 | 7.22 | 7.97 |
| Other genotypes | Nantes | 47 | 3.98 | 1.16 | 1.63 | 2.35 | 3.85 | 5.80 | 7.12 | 7.79 |
| Other genotypes | Nantes | 48 | 3.89 | 1.09 | 1.57 | 2.27 | 3.76 | 5.67 | 6.96 | 7.63 |
| Other genotypes | Nantes | 49 | 3.81 | 1.09 | 1.49 | 2.19 | 3.68 | 5.56 | 6.82 | 7.57 |
| Other genotypes | Nantes | 50 | 3.72 | 1.05 | 1.42 | 2.11 | 3.59 | 5.47 | 6.69 | 7.38 |
| Other genotypes | Nantes | 51 | 3.62 | 0.93 | 1.35 | 2.04 | 3.50 | 5.36 | 6.59 | 7.24 |
| Other genotypes | Nantes | 52 | 3.51 | 0.89 | 1.29 | 1.96 | 3.38 | 5.21 | 6.43 | 7.06 |
| Other genotypes | Nantes | 53 | 3.38 | 0.83 | 1.19 | 1.87 | 3.25 | 5.06 | 6.28 | 6.88 |
| Other genotypes | Nantes | 54 | 3.23 | 0.72 | 1.10 | 1.75 | 3.11 | 4.87 | 6.09 | 6.67 |
| Other genotypes | Nantes | 55 | 3.09 | 0.63 | 1.02 | 1.64 | 2.97 | 4.68 | 5.85 | 6.41 |
| Other genotypes | Nantes | 56 | 2.95 | 0.52 | 0.92 | 1.53 | 2.82 | 4.51 | 5.64 | 6.17 |
| Other genotypes | Nantes | 57 | 2.80 | 0.41 | 0.81 | 1.44 | 2.68 | 4.32 | 5.44 | 5.96 |
| Other genotypes | Nantes | 58 | 2.68 | 0.32 | 0.74 | 1.34 | 2.56 | 4.17 | 5.27 | 5.75 |
| Other genotypes | Nantes | 59 | 2.57 | 0.24 | 0.64 | 1.24 | 2.45 | 4.03 | 5.12 | 5.57 |
| Other genotypes | Nantes | 60 | 2.48 | 0.18 | 0.57 | 1.15 | 2.37 | 3.93 | 4.96 | 5.48 |

**Table S10 Posterior difference in expected HR HPV cervical infection prevalence (in percentage points) in major French cities, between opportunistic and organised screening, stratified by type of test, city, and age. Table counterpart to Figure 3B.** ***(continued)***

| | | | Posterior distribution | | | | | | | |
|---|---|---|---|---|---|---|---|---|---|---|
| | | | | Percentiles | | | | | | |
| Virus | City | Age | Average | 0.01 | 0.025 | 0.1 | 0.5 | 0.9 | 0.975 | 0.99 |
| Other genotypes | Nantes | 61 | 2.41 | 0.04 | 0.52 | 1.08 | 2.30 | 3.86 | 4.88 | 5.37 |
| Other genotypes | Nantes | 62 | 2.35 | -0.02 | 0.44 | 1.02 | 2.25 | 3.80 | 4.81 | 5.34 |
| Other genotypes | Nantes | 63 | 2.32 | -0.17 | 0.37 | 0.96 | 2.22 | 3.79 | 4.81 | 5.35 |
| Other genotypes | Nantes | 64 | 2.30 | -0.29 | 0.28 | 0.89 | 2.20 | 3.80 | 4.87 | 5.35 |
| Other genotypes | Nantes | 65 | 2.29 | -0.37 | 0.17 | 0.84 | 2.19 | 3.85 | 4.93 | 5.46 |
| Other genotypes | Nantes | 66 | 2.29 | -0.49 | 0.06 | 0.78 | 2.19 | 3.93 | 4.99 | 5.54 |
| Other genotypes | Nice | 30 | 6.22 | 0.26 | 1.39 | 3.00 | 6.11 | 9.61 | 11.72 | 13.06 |
| Other genotypes | Nice | 31 | 5.95 | 0.34 | 1.30 | 2.86 | 5.85 | 9.20 | 11.28 | 12.54 |
| Other genotypes | Nice | 32 | 5.65 | 0.30 | 1.18 | 2.72 | 5.55 | 8.77 | 10.74 | 11.89 |
| Other genotypes | Nice | 33 | 5.32 | 0.28 | 1.07 | 2.49 | 5.23 | 8.29 | 10.20 | 11.32 |
| Other genotypes | Nice | 34 | 5.04 | 0.24 | 0.91 | 2.35 | 4.95 | 7.87 | 9.66 | 10.80 |
| Other genotypes | Nice | 35 | 4.79 | 0.16 | 0.79 | 2.22 | 4.70 | 7.50 | 9.21 | 10.46 |
| Other genotypes | Nice | 36 | 4.55 | 0.08 | 0.73 | 2.07 | 4.45 | 7.19 | 8.89 | 10.04 |
| Other genotypes | Nice | 37 | 4.36 | -0.01 | 0.66 | 1.94 | 4.26 | 6.91 | 8.59 | 9.81 |
| Other genotypes | Nice | 38 | 4.17 | -0.09 | 0.59 | 1.83 | 4.06 | 6.67 | 8.29 | 9.48 |
| Other genotypes | Nice | 39 | 3.99 | -0.14 | 0.48 | 1.71 | 3.89 | 6.42 | 8.02 | 9.15 |
| Other genotypes | Nice | 40 | 3.80 | -0.20 | 0.35 | 1.60 | 3.70 | 6.16 | 7.69 | 8.79 |
| Other genotypes | Nice | 41 | 3.63 | -0.28 | 0.27 | 1.48 | 3.53 | 5.92 | 7.46 | 8.48 |
| Other genotypes | Nice | 42 | 3.49 | -0.34 | 0.24 | 1.40 | 3.37 | 5.74 | 7.21 | 8.22 |
| Other genotypes | Nice | 43 | 3.36 | -0.45 | 0.20 | 1.30 | 3.26 | 5.57 | 7.01 | 7.93 |
| Other genotypes | Nice | 44 | 3.24 | -0.53 | 0.15 | 1.21 | 3.15 | 5.40 | 6.83 | 7.70 |
| Other genotypes | Nice | 45 | 3.13 | -0.60 | 0.09 | 1.13 | 3.03 | 5.27 | 6.66 | 7.51 |
| Other genotypes | Nice | 46 | 3.04 | -0.66 | 0.02 | 1.05 | 2.95 | 5.14 | 6.55 | 7.35 |
| Other genotypes | Nice | 47 | 2.94 | -0.74 | -0.03 | 1.00 | 2.86 | 5.02 | 6.43 | 7.24 |
| Other genotypes | Nice | 48 | 2.86 | -0.80 | -0.09 | 0.93 | 2.78 | 4.91 | 6.30 | 7.09 |
| Other genotypes | Nice | 49 | 2.78 | -0.86 | -0.15 | 0.86 | 2.70 | 4.78 | 6.16 | 6.95 |
| Other genotypes | Nice | 50 | 2.70 | -0.94 | -0.21 | 0.79 | 2.63 | 4.67 | 6.08 | 6.81 |
| Other genotypes | Nice | 51 | 2.61 | -0.97 | -0.28 | 0.71 | 2.53 | 4.54 | 5.92 | 6.65 |
| Other genotypes | Nice | 52 | 2.50 | -1.08 | -0.35 | 0.63 | 2.43 | 4.43 | 5.73 | 6.48 |

**Table S10 Posterior difference in expected HR HPV cervical infection prevalence (in percentage points) in major French cities, between opportunistic and organised screening, stratified by type of test, city, and age. Table counterpart to Figure 3B.** ***(continued)***

| | | | Posterior distribution | | | | | | | |
|---|---|---|---|---|---|---|---|---|---|---|
| | | | | Percentiles | | | | | | |
| Virus | City | Age | Average | 0.01 | 0.025 | 0.1 | 0.5 | 0.9 | 0.975 | 0.99 |
| Other genotypes | Nice | 53 | 2.39 | -1.13 | -0.43 | 0.54 | 2.32 | 4.28 | 5.56 | 6.33 |
| Other genotypes | Nice | 54 | 2.25 | -1.13 | -0.51 | 0.46 | 2.19 | 4.12 | 5.33 | 6.14 |
| Other genotypes | Nice | 55 | 2.12 | -1.18 | -0.58 | 0.39 | 2.05 | 3.94 | 5.13 | 5.98 |
| Other genotypes | Nice | 56 | 2.00 | -1.27 | -0.65 | 0.29 | 1.92 | 3.78 | 4.91 | 5.75 |
| Other genotypes | Nice | 57 | 1.87 | -1.34 | -0.72 | 0.19 | 1.80 | 3.61 | 4.76 | 5.54 |
| Other genotypes | Nice | 58 | 1.75 | -1.40 | -0.81 | 0.12 | 1.69 | 3.46 | 4.54 | 5.38 |
| Other genotypes | Nice | 59 | 1.65 | -1.45 | -0.85 | 0.04 | 1.60 | 3.33 | 4.42 | 5.20 |
| Other genotypes | Nice | 60 | 1.57 | -1.49 | -0.94 | -0.02 | 1.52 | 3.22 | 4.30 | 5.09 |
| Other genotypes | Nice | 61 | 1.50 | -1.61 | -1.02 | -0.09 | 1.47 | 3.16 | 4.20 | 4.97 |
| Other genotypes | Nice | 62 | 1.44 | -1.70 | -1.09 | -0.15 | 1.41 | 3.10 | 4.13 | 4.95 |
| Other genotypes | Nice | 63 | 1.40 | -1.78 | -1.19 | -0.24 | 1.37 | 3.09 | 4.12 | 4.92 |
| Other genotypes | Nice | 64 | 1.36 | -1.83 | -1.28 | -0.31 | 1.34 | 3.08 | 4.13 | 5.02 |
| Other genotypes | Nice | 65 | 1.34 | -1.95 | -1.43 | -0.38 | 1.33 | 3.10 | 4.12 | 5.02 |
| Other genotypes | Nice | 66 | 1.31 | -2.11 | -1.56 | -0.46 | 1.30 | 3.12 | 4.18 | 5.06 |
| Other genotypes | Paris | 30 | 9.40 | 4.31 | 5.10 | 6.50 | 9.45 | 12.23 | 13.75 | 14.50 |
| Other genotypes | Paris | 31 | 9.05 | 4.16 | 5.01 | 6.30 | 9.08 | 11.72 | 13.20 | 13.80 |
| Other genotypes | Paris | 32 | 8.64 | 4.06 | 4.79 | 6.06 | 8.67 | 11.16 | 12.61 | 13.18 |
| Other genotypes | Paris | 33 | 8.21 | 3.92 | 4.49 | 5.76 | 8.19 | 10.61 | 11.97 | 12.51 |
| Other genotypes | Paris | 34 | 7.82 | 3.68 | 4.21 | 5.46 | 7.78 | 10.13 | 11.42 | 12.02 |
| Other genotypes | Paris | 35 | 7.48 | 3.53 | 4.00 | 5.22 | 7.46 | 9.72 | 10.94 | 11.52 |
| Other genotypes | Paris | 36 | 7.15 | 3.32 | 3.79 | 5.00 | 7.13 | 9.32 | 10.52 | 11.16 |
| Other genotypes | Paris | 37 | 6.88 | 3.21 | 3.62 | 4.78 | 6.86 | 9.00 | 10.12 | 10.76 |
| Other genotypes | Paris | 38 | 6.62 | 3.04 | 3.50 | 4.57 | 6.60 | 8.65 | 9.75 | 10.31 |
| Other genotypes | Paris | 39 | 6.36 | 2.89 | 3.35 | 4.38 | 6.36 | 8.34 | 9.39 | 10.01 |
| Other genotypes | Paris | 40 | 6.10 | 2.69 | 3.17 | 4.15 | 6.09 | 8.03 | 9.06 | 9.70 |
| Other genotypes | Paris | 41 | 5.87 | 2.51 | 3.01 | 3.97 | 5.85 | 7.75 | 8.77 | 9.39 |
| Other genotypes | Paris | 42 | 5.66 | 2.38 | 2.87 | 3.80 | 5.64 | 7.50 | 8.47 | 9.15 |
| Other genotypes | Paris | 43 | 5.48 | 2.27 | 2.75 | 3.66 | 5.46 | 7.29 | 8.16 | 8.95 |
| Other genotypes | Paris | 44 | 5.32 | 2.13 | 2.59 | 3.52 | 5.31 | 7.10 | 8.00 | 8.70 |

**Table S10** **Posterior difference in expected HR HPV cervical infection prevalence (in percentage points) in major French cities, between opportunistic and organised screening, stratified by type of test, city, and age. Table counterpart to Figure 3B.** ***(continued)***

| | | | Posterior distribution | | | | | | | |
|---|---|---|---|---|---|---|---|---|---|---|
| | | | | Percentiles | | | | | | |
| Virus | City | Age | Average | 0.01 | 0.025 | 0.1 | 0.5 | 0.9 | 0.975 | 0.99 |
| Other genotypes | Paris | 45 | 5.16 | 2.02 | 2.46 | 3.37 | 5.16 | 6.92 | 7.81 | 8.50 |
| Other genotypes | Paris | 46 | 5.03 | 1.89 | 2.34 | 3.25 | 5.02 | 6.74 | 7.66 | 8.41 |
| Other genotypes | Paris | 47 | 4.90 | 1.81 | 2.23 | 3.14 | 4.89 | 6.62 | 7.56 | 8.30 |
| Other genotypes | Paris | 48 | 4.79 | 1.72 | 2.13 | 3.04 | 4.77 | 6.49 | 7.45 | 8.07 |
| Other genotypes | Paris | 49 | 4.68 | 1.62 | 2.03 | 2.93 | 4.66 | 6.36 | 7.31 | 7.97 |
| Other genotypes | Paris | 50 | 4.56 | 1.49 | 1.92 | 2.81 | 4.55 | 6.23 | 7.17 | 7.82 |
| Other genotypes | Paris | 51 | 4.44 | 1.42 | 1.78 | 2.72 | 4.44 | 6.12 | 7.03 | 7.66 |
| Other genotypes | Paris | 52 | 4.30 | 1.26 | 1.65 | 2.58 | 4.29 | 5.95 | 6.86 | 7.37 |
| Other genotypes | Paris | 53 | 4.14 | 1.16 | 1.50 | 2.45 | 4.13 | 5.77 | 6.64 | 7.17 |
| Other genotypes | Paris | 54 | 3.95 | 1.01 | 1.36 | 2.31 | 3.96 | 5.57 | 6.41 | 7.00 |
| Other genotypes | Paris | 55 | 3.77 | 0.88 | 1.21 | 2.16 | 3.77 | 5.34 | 6.18 | 6.78 |
| Other genotypes | Paris | 56 | 3.59 | 0.71 | 1.11 | 2.01 | 3.58 | 5.13 | 5.97 | 6.48 |
| Other genotypes | Paris | 57 | 3.41 | 0.56 | 0.97 | 1.88 | 3.41 | 4.93 | 5.72 | 6.24 |
| Other genotypes | Paris | 58 | 3.25 | 0.40 | 0.86 | 1.74 | 3.25 | 4.73 | 5.54 | 6.05 |
| Other genotypes | Paris | 59 | 3.10 | 0.32 | 0.75 | 1.63 | 3.11 | 4.58 | 5.38 | 5.82 |
| Other genotypes | Paris | 60 | 2.99 | 0.22 | 0.64 | 1.51 | 3.00 | 4.46 | 5.26 | 5.66 |
| Other genotypes | Paris | 61 | 2.90 | 0.07 | 0.56 | 1.40 | 2.90 | 4.40 | 5.18 | 5.60 |
| Other genotypes | Paris | 62 | 2.82 | -0.08 | 0.52 | 1.32 | 2.83 | 4.34 | 5.11 | 5.51 |
| Other genotypes | Paris | 63 | 2.77 | -0.18 | 0.44 | 1.26 | 2.78 | 4.32 | 5.13 | 5.56 |
| Other genotypes | Paris | 64 | 2.74 | -0.31 | 0.31 | 1.17 | 2.73 | 4.32 | 5.13 | 5.60 |
| Other genotypes | Paris | 65 | 2.71 | -0.41 | 0.20 | 1.05 | 2.71 | 4.36 | 5.24 | 5.66 |
| Other genotypes | Paris | 66 | 2.70 | -0.56 | 0.07 | 0.94 | 2.71 | 4.44 | 5.40 | 5.78 |
| Other genotypes | Rennes | 30 | 9.95 | 5.50 | 6.07 | 7.36 | 9.90 | 12.64 | 14.04 | 14.98 |
| Other genotypes | Rennes | 31 | 9.55 | 5.36 | 5.91 | 7.07 | 9.51 | 12.15 | 13.40 | 14.29 |
| Other genotypes | Rennes | 32 | 9.10 | 5.15 | 5.58 | 6.73 | 9.08 | 11.57 | 12.74 | 13.56 |
| Other genotypes | Rennes | 33 | 8.62 | 4.85 | 5.30 | 6.36 | 8.59 | 10.95 | 12.06 | 12.83 |
| Other genotypes | Rennes | 34 | 8.19 | 4.55 | 5.04 | 6.04 | 8.14 | 10.39 | 11.51 | 12.28 |
| Other genotypes | Rennes | 35 | 7.83 | 4.36 | 4.81 | 5.74 | 7.77 | 9.95 | 11.04 | 11.77 |
| Other genotypes | Rennes | 36 | 7.49 | 4.12 | 4.61 | 5.47 | 7.43 | 9.53 | 10.60 | 11.27 |

**Table S10** **Posterior difference in expected HR HPV cervical infection prevalence (in percentage points) in major French cities, between opportunistic and organised screening, stratified by type of test, city, and age. Table counterpart to Figure 3B.** ***(continued)***

| | | | Posterior distribution | | | | | | | |
|---|---|---|---|---|---|---|---|---|---|---|
| | | | | Percentiles | | | | | | |
| Virus | City | Age | Average | 0.01 | 0.025 | 0.1 | 0.5 | 0.9 | 0.975 | 0.99 |
| Other genotypes | Rennes | 37 | 7.20 | 3.91 | 4.43 | 5.26 | 7.13 | 9.20 | 10.24 | 10.87 |
| Other genotypes | Rennes | 38 | 6.93 | 3.72 | 4.24 | 5.07 | 6.86 | 8.87 | 9.90 | 10.48 |
| Other genotypes | Rennes | 39 | 6.67 | 3.54 | 4.06 | 4.85 | 6.61 | 8.55 | 9.56 | 10.11 |
| Other genotypes | Rennes | 40 | 6.41 | 3.37 | 3.87 | 4.65 | 6.35 | 8.26 | 9.22 | 9.75 |
| Other genotypes | Rennes | 41 | 6.17 | 3.22 | 3.68 | 4.46 | 6.12 | 7.94 | 8.90 | 9.38 |
| Other genotypes | Rennes | 42 | 5.96 | 3.12 | 3.52 | 4.29 | 5.92 | 7.73 | 8.63 | 9.13 |
| Other genotypes | Rennes | 43 | 5.78 | 2.97 | 3.39 | 4.12 | 5.75 | 7.51 | 8.39 | 8.90 |
| Other genotypes | Rennes | 44 | 5.62 | 2.81 | 3.24 | 4.00 | 5.59 | 7.27 | 8.23 | 8.71 |
| Other genotypes | Rennes | 45 | 5.47 | 2.72 | 3.11 | 3.89 | 5.45 | 7.11 | 8.05 | 8.49 |
| Other genotypes | Rennes | 46 | 5.34 | 2.61 | 3.01 | 3.78 | 5.31 | 6.98 | 7.90 | 8.39 |
| Other genotypes | Rennes | 47 | 5.22 | 2.54 | 2.91 | 3.68 | 5.18 | 6.83 | 7.75 | 8.29 |
| Other genotypes | Rennes | 48 | 5.12 | 2.41 | 2.82 | 3.57 | 5.08 | 6.72 | 7.62 | 8.19 |
| Other genotypes | Rennes | 49 | 5.02 | 2.30 | 2.74 | 3.47 | 4.98 | 6.62 | 7.51 | 8.11 |
| Other genotypes | Rennes | 50 | 4.92 | 2.24 | 2.64 | 3.37 | 4.87 | 6.50 | 7.42 | 8.00 |
| Other genotypes | Rennes | 51 | 4.81 | 2.19 | 2.58 | 3.28 | 4.76 | 6.40 | 7.25 | 7.85 |
| Other genotypes | Rennes | 52 | 4.67 | 2.05 | 2.46 | 3.15 | 4.63 | 6.25 | 7.10 | 7.65 |
| Other genotypes | Rennes | 53 | 4.52 | 1.93 | 2.30 | 3.02 | 4.48 | 6.09 | 6.95 | 7.45 |
| Other genotypes | Rennes | 54 | 4.34 | 1.78 | 2.17 | 2.88 | 4.32 | 5.88 | 6.70 | 7.19 |
| Other genotypes | Rennes | 55 | 4.17 | 1.61 | 2.01 | 2.73 | 4.15 | 5.69 | 6.48 | 6.94 |
| Other genotypes | Rennes | 56 | 4.00 | 1.54 | 1.85 | 2.59 | 3.98 | 5.50 | 6.27 | 6.73 |
| Other genotypes | Rennes | 57 | 3.83 | 1.35 | 1.71 | 2.46 | 3.82 | 5.30 | 6.04 | 6.47 |
| Other genotypes | Rennes | 58 | 3.68 | 1.26 | 1.58 | 2.32 | 3.67 | 5.13 | 5.84 | 6.32 |
| Other genotypes | Rennes | 59 | 3.55 | 1.16 | 1.48 | 2.20 | 3.53 | 4.98 | 5.68 | 6.15 |
| Other genotypes | Rennes | 60 | 3.44 | 1.05 | 1.36 | 2.10 | 3.42 | 4.85 | 5.55 | 6.04 |
| Other genotypes | Rennes | 61 | 3.36 | 0.94 | 1.25 | 2.02 | 3.34 | 4.78 | 5.47 | 5.90 |
| Other genotypes | Rennes | 62 | 3.30 | 0.83 | 1.15 | 1.95 | 3.28 | 4.72 | 5.45 | 5.82 |
| Other genotypes | Rennes | 63 | 3.27 | 0.73 | 1.05 | 1.88 | 3.25 | 4.71 | 5.44 | 5.83 |
| Other genotypes | Rennes | 64 | 3.25 | 0.58 | 0.98 | 1.83 | 3.24 | 4.73 | 5.49 | 5.90 |
| Other genotypes | Rennes | 65 | 3.25 | 0.49 | 0.90 | 1.80 | 3.23 | 4.78 | 5.56 | 6.03 |

**Table S10 Posterior difference in expected HR HPV cervical infection prevalence (in percentage points) in major French cities, between opportunistic and organised screening, stratified by type of test, city, and age. Table counterpart to Figure 3B. *(continued)***

| | | | Posterior distribution | | | | | | | |
|---|---|---|---|---|---|---|---|---|---|---|
| | | | | Percentiles | | | | | | |
| Virus | City | Age | Average | 0.01 | 0.025 | 0.1 | 0.5 | 0.9 | 0.975 | 0.99 |
| Other genotypes | Rennes | 66 | 3.27 | 0.30 | 0.80 | 1.72 | 3.24 | 4.86 | 5.68 | 6.25 |
| Other genotypes | Strasbourg | 30 | 6.82 | 1.96 | 2.70 | 4.02 | 6.66 | 9.82 | 11.78 | 13.13 |
| Other genotypes | Strasbourg | 31 | 6.52 | 1.89 | 2.62 | 3.87 | 6.31 | 9.45 | 11.30 | 12.60 |
| Other genotypes | Strasbourg | 32 | 6.18 | 1.76 | 2.49 | 3.68 | 5.99 | 8.99 | 10.74 | 11.93 |
| Other genotypes | Strasbourg | 33 | 5.82 | 1.62 | 2.32 | 3.46 | 5.64 | 8.50 | 10.19 | 11.33 |
| Other genotypes | Strasbourg | 34 | 5.50 | 1.48 | 2.15 | 3.28 | 5.33 | 8.06 | 9.74 | 10.90 |
| Other genotypes | Strasbourg | 35 | 5.24 | 1.42 | 2.02 | 3.10 | 5.05 | 7.69 | 9.29 | 10.44 |
| Other genotypes | Strasbourg | 36 | 4.99 | 1.33 | 1.88 | 2.91 | 4.81 | 7.35 | 8.92 | 9.96 |
| Other genotypes | Strasbourg | 37 | 4.77 | 1.26 | 1.74 | 2.77 | 4.62 | 7.04 | 8.61 | 9.57 |
| Other genotypes | Strasbourg | 38 | 4.58 | 1.19 | 1.64 | 2.62 | 4.42 | 6.79 | 8.24 | 9.27 |
| Other genotypes | Strasbourg | 39 | 4.39 | 1.10 | 1.55 | 2.48 | 4.23 | 6.54 | 8.00 | 8.94 |
| Other genotypes | Strasbourg | 40 | 4.19 | 1.00 | 1.45 | 2.34 | 4.04 | 6.28 | 7.62 | 8.62 |
| Other genotypes | Strasbourg | 41 | 4.02 | 0.92 | 1.36 | 2.23 | 3.88 | 6.03 | 7.37 | 8.27 |
| Other genotypes | Strasbourg | 42 | 3.87 | 0.89 | 1.27 | 2.11 | 3.73 | 5.84 | 7.16 | 7.97 |
| Other genotypes | Strasbourg | 43 | 3.74 | 0.82 | 1.20 | 2.01 | 3.60 | 5.68 | 6.96 | 7.80 |
| Other genotypes | Strasbourg | 44 | 3.62 | 0.73 | 1.12 | 1.94 | 3.48 | 5.50 | 6.82 | 7.63 |
| Other genotypes | Strasbourg | 45 | 3.51 | 0.66 | 1.06 | 1.85 | 3.38 | 5.35 | 6.63 | 7.44 |
| Other genotypes | Strasbourg | 46 | 3.41 | 0.62 | 1.02 | 1.79 | 3.29 | 5.26 | 6.53 | 7.33 |
| Other genotypes | Strasbourg | 47 | 3.32 | 0.50 | 0.97 | 1.72 | 3.20 | 5.13 | 6.44 | 7.16 |
| Other genotypes | Strasbourg | 48 | 3.24 | 0.48 | 0.93 | 1.66 | 3.13 | 5.02 | 6.32 | 7.04 |
| Other genotypes | Strasbourg | 49 | 3.16 | 0.47 | 0.89 | 1.59 | 3.05 | 4.94 | 6.19 | 6.96 |
| Other genotypes | Strasbourg | 50 | 3.08 | 0.38 | 0.83 | 1.52 | 2.98 | 4.85 | 6.11 | 6.89 |
| Other genotypes | Strasbourg | 51 | 3.00 | 0.34 | 0.77 | 1.46 | 2.90 | 4.73 | 5.98 | 6.74 |
| Other genotypes | Strasbourg | 52 | 2.89 | 0.29 | 0.71 | 1.39 | 2.81 | 4.60 | 5.83 | 6.58 |
| Other genotypes | Strasbourg | 53 | 2.78 | 0.24 | 0.63 | 1.29 | 2.69 | 4.47 | 5.66 | 6.42 |
| Other genotypes | Strasbourg | 54 | 2.65 | 0.17 | 0.54 | 1.20 | 2.55 | 4.29 | 5.44 | 6.18 |
| Other genotypes | Strasbourg | 55 | 2.52 | 0.05 | 0.46 | 1.10 | 2.41 | 4.09 | 5.28 | 5.93 |
| Other genotypes | Strasbourg | 56 | 2.40 | -0.03 | 0.37 | 0.99 | 2.30 | 3.93 | 5.09 | 5.75 |
| Other genotypes | Strasbourg | 57 | 2.27 | -0.10 | 0.27 | 0.91 | 2.17 | 3.79 | 4.90 | 5.59 |

**Table S10 Posterior difference in expected HR HPV cervical infection prevalence (in percentage points) in major French cities, between opportunistic and organised screening, stratified by type of test, city, and age. Table counterpart to Figure 3B.** ***(continued)***

| | | | Posterior distribution | | | | | | | |
|---|---|---|---|---|---|---|---|---|---|---|
| | | | | Percentiles | | | | | | |
| Virus | City | Age | Average | 0.01 | 0.025 | 0.1 | 0.5 | 0.9 | 0.975 | 0.99 |
| Other genotypes | Strasbourg | 58 | 2.16 | -0.20 | 0.18 | 0.82 | 2.05 | 3.63 | 4.72 | 5.38 |
| Other genotypes | Strasbourg | 59 | 2.06 | -0.24 | 0.11 | 0.76 | 1.96 | 3.49 | 4.60 | 5.19 |
| Other genotypes | Strasbourg | 60 | 1.98 | -0.32 | 0.04 | 0.71 | 1.87 | 3.41 | 4.50 | 5.09 |
| Other genotypes | Strasbourg | 61 | 1.92 | -0.42 | -0.03 | 0.64 | 1.81 | 3.32 | 4.43 | 4.97 |
| Other genotypes | Strasbourg | 62 | 1.87 | -0.47 | -0.10 | 0.57 | 1.77 | 3.27 | 4.37 | 4.97 |
| Other genotypes | Strasbourg | 63 | 1.83 | -0.53 | -0.14 | 0.53 | 1.73 | 3.25 | 4.35 | 4.98 |
| Other genotypes | Strasbourg | 64 | 1.81 | -0.65 | -0.24 | 0.47 | 1.71 | 3.28 | 4.36 | 4.97 |
| Other genotypes | Strasbourg | 65 | 1.80 | -0.75 | -0.33 | 0.41 | 1.70 | 3.32 | 4.44 | 5.00 |
| Other genotypes | Strasbourg | 66 | 1.79 | -0.89 | -0.41 | 0.33 | 1.69 | 3.36 | 4.49 | 5.06 |
| Other genotypes | Toulouse | 30 | 6.65 | 1.36 | 2.35 | 3.88 | 6.54 | 9.61 | 11.27 | 12.11 |
| Other genotypes | Toulouse | 31 | 6.37 | 1.48 | 2.33 | 3.75 | 6.24 | 9.19 | 10.79 | 11.53 |
| Other genotypes | Toulouse | 32 | 6.04 | 1.48 | 2.18 | 3.57 | 5.94 | 8.75 | 10.28 | 11.10 |
| Other genotypes | Toulouse | 33 | 5.70 | 1.32 | 2.04 | 3.33 | 5.58 | 8.24 | 9.71 | 10.48 |
| Other genotypes | Toulouse | 34 | 5.39 | 1.30 | 1.95 | 3.11 | 5.28 | 7.84 | 9.25 | 9.95 |
| Other genotypes | Toulouse | 35 | 5.13 | 1.25 | 1.78 | 2.93 | 5.01 | 7.46 | 8.90 | 9.59 |
| Other genotypes | Toulouse | 36 | 4.88 | 1.01 | 1.63 | 2.79 | 4.77 | 7.14 | 8.49 | 9.26 |
| Other genotypes | Toulouse | 37 | 4.67 | 0.99 | 1.49 | 2.63 | 4.58 | 6.88 | 8.20 | 8.91 |
| Other genotypes | Toulouse | 38 | 4.48 | 0.88 | 1.40 | 2.48 | 4.40 | 6.61 | 7.91 | 8.62 |
| Other genotypes | Toulouse | 39 | 4.29 | 0.79 | 1.30 | 2.36 | 4.22 | 6.37 | 7.62 | 8.26 |
| Other genotypes | Toulouse | 40 | 4.09 | 0.73 | 1.17 | 2.21 | 4.02 | 6.12 | 7.34 | 7.97 |
| Other genotypes | Toulouse | 41 | 3.91 | 0.58 | 1.07 | 2.07 | 3.83 | 5.87 | 7.07 | 7.63 |
| Other genotypes | Toulouse | 42 | 3.76 | 0.48 | 0.98 | 1.97 | 3.68 | 5.69 | 6.87 | 7.37 |
| Other genotypes | Toulouse | 43 | 3.63 | 0.42 | 0.90 | 1.85 | 3.55 | 5.50 | 6.65 | 7.14 |
| Other genotypes | Toulouse | 44 | 3.50 | 0.34 | 0.85 | 1.77 | 3.43 | 5.35 | 6.51 | 6.99 |
| Other genotypes | Toulouse | 45 | 3.39 | 0.32 | 0.80 | 1.67 | 3.32 | 5.20 | 6.33 | 6.82 |
| Other genotypes | Toulouse | 46 | 3.29 | 0.27 | 0.73 | 1.58 | 3.23 | 5.08 | 6.16 | 6.69 |
| Other genotypes | Toulouse | 47 | 3.19 | 0.20 | 0.67 | 1.51 | 3.13 | 4.96 | 6.02 | 6.55 |
| Other genotypes | Toulouse | 48 | 3.11 | 0.15 | 0.62 | 1.46 | 3.04 | 4.85 | 5.91 | 6.38 |
| Other genotypes | Toulouse | 49 | 3.02 | 0.10 | 0.57 | 1.38 | 2.96 | 4.76 | 5.78 | 6.29 |

**Table S10 Posterior difference in expected HR HPV cervical infection prevalence (in percentage points) in major French cities, between opportunistic and organised screening, stratified by type of test, city, and age. Table counterpart to Figure 3B.** ***(continued)***

| | | | Posterior distribution | | | | | | | |
|---|---|---|---|---|---|---|---|---|---|---|
| | | | | Percentiles | | | | | | |
| Virus | City | Age | Average | 0.01 | 0.025 | 0.1 | 0.5 | 0.9 | 0.975 | 0.99 |
| Other genotypes | Toulouse | 50 | 2.94 | 0.05 | 0.50 | 1.31 | 2.88 | 4.63 | 5.66 | 6.20 |
| Other genotypes | Toulouse | 51 | 2.85 | -0.07 | 0.40 | 1.24 | 2.79 | 4.52 | 5.56 | 6.08 |
| Other genotypes | Toulouse | 52 | 2.74 | -0.16 | 0.32 | 1.15 | 2.68 | 4.39 | 5.39 | 5.92 |
| Other genotypes | Toulouse | 53 | 2.62 | -0.25 | 0.24 | 1.06 | 2.57 | 4.26 | 5.21 | 5.75 |
| Other genotypes | Toulouse | 54 | 2.48 | -0.39 | 0.14 | 0.95 | 2.44 | 4.06 | 5.00 | 5.56 |
| Other genotypes | Toulouse | 55 | 2.34 | -0.49 | 0.06 | 0.85 | 2.30 | 3.89 | 4.83 | 5.36 |
| Other genotypes | Toulouse | 56 | 2.21 | -0.56 | -0.01 | 0.74 | 2.18 | 3.72 | 4.66 | 5.14 |
| Other genotypes | Toulouse | 57 | 2.08 | -0.67 | -0.11 | 0.64 | 2.05 | 3.57 | 4.45 | 4.97 |
| Other genotypes | Toulouse | 58 | 1.96 | -0.81 | -0.20 | 0.54 | 1.93 | 3.43 | 4.29 | 4.81 |
| Other genotypes | Toulouse | 59 | 1.86 | -0.90 | -0.28 | 0.45 | 1.82 | 3.32 | 4.15 | 4.64 |
| Other genotypes | Toulouse | 60 | 1.77 | -0.98 | -0.37 | 0.38 | 1.74 | 3.21 | 4.06 | 4.56 |
| Other genotypes | Toulouse | 61 | 1.70 | -0.96 | -0.43 | 0.32 | 1.66 | 3.13 | 4.00 | 4.46 |
| Other genotypes | Toulouse | 62 | 1.65 | -1.07 | -0.53 | 0.27 | 1.59 | 3.10 | 3.98 | 4.44 |
| Other genotypes | Toulouse | 63 | 1.60 | -1.19 | -0.59 | 0.20 | 1.55 | 3.07 | 3.95 | 4.44 |
| Other genotypes | Toulouse | 64 | 1.57 | -1.27 | -0.70 | 0.14 | 1.52 | 3.09 | 4.00 | 4.48 |
| Other genotypes | Toulouse | 65 | 1.55 | -1.34 | -0.77 | 0.04 | 1.50 | 3.10 | 4.06 | 4.53 |
| Other genotypes | Toulouse | 66 | 1.53 | -1.49 | -0.91 | -0.04 | 1.48 | 3.16 | 4.10 | 4.62 |

**Table S11 Posterior percentage of postcodes with a greater expected infection prevalence under opportunistic screening, than under organised screening, stratified by age and genotype group, as of November. Table counterpart to Figure 3C.**

| | | Posterior distribution | | | | | | |
|---|---|---|---|---|---|---|---|---|
| | | | Percentiles | | | | | |
| Virus | Age | Average | 0.01 | 0.025 | 0.1 | 0.5 | 0.9 | 0.975 | 0.99 |
| HPV16/18 | 30 | 97.18 | 57.72 | 72.90 | 92.79 | 99.85 | 100.00 | 100.00 | 100.00 |
| HPV16/18 | 31 | 97.75 | 64.86 | 78.97 | 94.58 | 99.88 | 100.00 | 100.00 | 100.00 |
| HPV16/18 | 32 | 98.11 | 70.76 | 82.61 | 95.54 | 99.90 | 100.00 | 100.00 | 100.00 |

**Table S11 Posterior percentage of postcodes with a greater expected infection prevalence under opportunistic screening, than under organised screening, stratified by age and genotype group, as of November. Table counterpart to Figure 3C.** ***(continued)***

| | | Posterior distribution | | | | | | | |
|---|---|---|---|---|---|---|---|---|---|
| | | | Percentiles | | | | | | |
| Virus | Age | Average | 0.01 | 0.025 | 0.1 | 0.5 | 0.9 | 0.975 | 0.99 |
| HPV16/18 | 33 | 98.36 | 74.42 | 85.45 | 96.10 | 99.92 | 100.00 | 100.00 | 100.00 |
| HPV16/18 | 34 | 98.51 | 76.23 | 86.59 | 96.44 | 99.92 | 100.00 | 100.00 | 100.00 |
| HPV16/18 | 35 | 98.59 | 76.76 | 87.55 | 96.67 | 99.92 | 100.00 | 100.00 | 100.00 |
| HPV16/18 | 36 | 98.64 | 78.88 | 87.87 | 96.80 | 99.92 | 100.00 | 100.00 | 100.00 |
| HPV16/18 | 37 | 98.64 | 78.97 | 87.77 | 96.87 | 99.92 | 100.00 | 100.00 | 100.00 |
| HPV16/18 | 38 | 98.64 | 78.73 | 87.67 | 96.93 | 99.92 | 100.00 | 100.00 | 100.00 |
| HPV16/18 | 39 | 98.62 | 77.87 | 87.42 | 96.85 | 99.90 | 100.00 | 100.00 | 100.00 |
| HPV16/18 | 40 | 98.57 | 78.00 | 87.12 | 96.77 | 99.90 | 100.00 | 100.00 | 100.00 |
| HPV16/18 | 41 | 98.51 | 77.14 | 86.46 | 96.50 | 99.90 | 100.00 | 100.00 | 100.00 |
| HPV16/18 | 42 | 98.44 | 75.87 | 85.75 | 96.30 | 99.88 | 100.00 | 100.00 | 100.00 |
| HPV16/18 | 43 | 98.38 | 74.61 | 84.75 | 96.14 | 99.88 | 100.00 | 100.00 | 100.00 |
| HPV16/18 | 44 | 98.31 | 73.11 | 84.55 | 95.92 | 99.88 | 100.00 | 100.00 | 100.00 |
| HPV16/18 | 45 | 98.23 | 72.04 | 83.61 | 95.74 | 99.88 | 100.00 | 100.00 | 100.00 |
| HPV16/18 | 46 | 98.14 | 70.64 | 83.11 | 95.51 | 99.87 | 100.00 | 100.00 | 100.00 |
| HPV16/18 | 47 | 98.06 | 69.52 | 83.13 | 95.19 | 99.85 | 100.00 | 100.00 | 100.00 |
| HPV16/18 | 48 | 97.94 | 68.92 | 82.12 | 94.83 | 99.85 | 100.00 | 100.00 | 100.00 |
| HPV16/18 | 49 | 97.85 | 67.68 | 81.73 | 94.40 | 99.83 | 100.00 | 100.00 | 100.00 |
| HPV16/18 | 50 | 97.71 | 65.90 | 80.71 | 94.13 | 99.82 | 100.00 | 100.00 | 100.00 |
| HPV16/18 | 51 | 97.58 | 64.45 | 79.79 | 93.67 | 99.80 | 100.00 | 100.00 | 100.00 |
| HPV16/18 | 52 | 97.43 | 64.15 | 78.92 | 93.05 | 99.78 | 100.00 | 100.00 | 100.00 |
| HPV16/18 | 53 | 97.23 | 63.50 | 77.21 | 92.46 | 99.75 | 100.00 | 100.00 | 100.00 |
| HPV16/18 | 54 | 97.00 | 60.57 | 74.69 | 91.76 | 99.72 | 100.00 | 100.00 | 100.00 |
| HPV16/18 | 55 | 96.75 | 55.99 | 73.63 | 91.02 | 99.69 | 100.00 | 100.00 | 100.00 |
| HPV16/18 | 56 | 96.49 | 55.41 | 71.14 | 90.00 | 99.64 | 100.00 | 100.00 | 100.00 |
| HPV16/18 | 57 | 96.15 | 52.64 | 69.07 | 88.91 | 99.59 | 100.00 | 100.00 | 100.00 |
| HPV16/18 | 58 | 95.81 | 50.06 | 67.08 | 87.75 | 99.54 | 100.00 | 100.00 | 100.00 |
| HPV16/18 | 59 | 95.43 | 48.62 | 64.48 | 86.37 | 99.47 | 100.00 | 100.00 | 100.00 |
| HPV16/18 | 60 | 95.01 | 44.11 | 61.30 | 84.87 | 99.42 | 100.00 | 100.00 | 100.00 |
| HPV16/18 | 61 | 94.53 | 41.95 | 57.65 | 83.72 | 99.35 | 100.00 | 100.00 | 100.00 |

**Table S11 Posterior percentage of postcodes with a greater expected infection prevalence under opportunistic screening, than under organised screening, stratified by age and genotype group, as of November. Table counterpart to Figure 3C.** ***(continued)***

| | | Posterior distribution | | | | | | | |
|---|---|---|---|---|---|---|---|---|---|
| | | | Percentiles | | | | | | |
| Virus | Age | Average | 0.01 | 0.025 | 0.1 | 0.5 | 0.9 | 0.975 | 0.99 |
| HPV16/18 | 62 | 93.98 | 37.41 | 53.77 | 81.45 | 99.32 | 100.00 | 100.00 | 100.00 |
| HPV16/18 | 63 | 93.30 | 32.11 | 48.47 | 78.85 | 99.20 | 100.00 | 100.00 | 100.00 |
| HPV16/18 | 64 | 92.51 | 26.80 | 43.74 | 74.77 | 99.09 | 100.00 | 100.00 | 100.00 |
| HPV16/18 | 65 | 91.58 | 22.10 | 37.81 | 71.11 | 98.99 | 100.00 | 100.00 | 100.00 |
| HPV16/18 | 66 | 90.50 | 17.11 | 31.43 | 67.41 | 98.82 | 100.00 | 100.00 | 100.00 |
| Other genotypes | 30 | 99.92 | 98.77 | 99.34 | 99.85 | 100.00 | 100.00 | 100.00 | 100.00 |
| Other genotypes | 31 | 99.92 | 98.94 | 99.35 | 99.85 | 100.00 | 100.00 | 100.00 | 100.00 |
| Other genotypes | 32 | 99.93 | 98.94 | 99.35 | 99.85 | 100.00 | 100.00 | 100.00 | 100.00 |
| Other genotypes | 33 | 99.92 | 98.84 | 99.32 | 99.85 | 100.00 | 100.00 | 100.00 | 100.00 |
| Other genotypes | 34 | 99.92 | 98.76 | 99.27 | 99.83 | 100.00 | 100.00 | 100.00 | 100.00 |
| Other genotypes | 35 | 99.92 | 98.62 | 99.17 | 99.80 | 100.00 | 100.00 | 100.00 | 100.00 |
| Other genotypes | 36 | 99.90 | 98.49 | 99.17 | 99.78 | 100.00 | 100.00 | 100.00 | 100.00 |
| Other genotypes | 37 | 99.88 | 98.26 | 99.07 | 99.75 | 100.00 | 100.00 | 100.00 | 100.00 |
| Other genotypes | 38 | 99.88 | 98.06 | 98.96 | 99.72 | 100.00 | 100.00 | 100.00 | 100.00 |
| Other genotypes | 39 | 99.87 | 97.56 | 98.87 | 99.69 | 99.98 | 100.00 | 100.00 | 100.00 |
| Other genotypes | 40 | 99.83 | 97.23 | 98.71 | 99.62 | 99.98 | 100.00 | 100.00 | 100.00 |
| Other genotypes | 41 | 99.82 | 96.87 | 98.44 | 99.57 | 99.98 | 100.00 | 100.00 | 100.00 |
| Other genotypes | 42 | 99.78 | 96.42 | 98.16 | 99.50 | 99.98 | 100.00 | 100.00 | 100.00 |
| Other genotypes | 43 | 99.77 | 95.91 | 97.96 | 99.44 | 99.98 | 100.00 | 100.00 | 100.00 |
| Other genotypes | 44 | 99.72 | 95.91 | 97.56 | 99.34 | 99.98 | 100.00 | 100.00 | 100.00 |
| Other genotypes | 45 | 99.69 | 95.24 | 97.31 | 99.27 | 99.97 | 100.00 | 100.00 | 100.00 |
| Other genotypes | 46 | 99.64 | 94.60 | 96.85 | 99.17 | 99.97 | 100.00 | 100.00 | 100.00 |
| Other genotypes | 47 | 99.59 | 93.90 | 96.42 | 99.02 | 99.97 | 100.00 | 100.00 | 100.00 |
| Other genotypes | 48 | 99.54 | 93.32 | 95.84 | 98.87 | 99.95 | 100.00 | 100.00 | 100.00 |
| Other genotypes | 49 | 99.47 | 92.34 | 95.47 | 98.72 | 99.95 | 100.00 | 100.00 | 100.00 |
| Other genotypes | 50 | 99.40 | 91.84 | 94.93 | 98.48 | 99.93 | 100.00 | 100.00 | 100.00 |
| Other genotypes | 51 | 99.30 | 91.18 | 94.36 | 98.24 | 99.92 | 100.00 | 100.00 | 100.00 |
| Other genotypes | 52 | 99.19 | 89.56 | 93.49 | 97.93 | 99.90 | 100.00 | 100.00 | 100.00 |
| Other genotypes | 53 | 99.04 | 87.85 | 92.29 | 97.58 | 99.88 | 100.00 | 100.00 | 100.00 |

**Table S11** **Posterior percentage of postcodes with a greater expected infection prevalence under opportunistic screening, than under organised screening, stratified by age and genotype group, as of November. Table counterpart to Figure 3C.** ***(continued)***

| | | Posterior distribution | | | | | | | |
|---|---|---|---|---|---|---|---|---|---|
| | | | Percentiles | | | | | | |
| Virus | Age | Average | 0.01 | 0.025 | 0.1 | 0.5 | 0.9 | 0.975 | 0.99 |
| Other genotypes | 54 | 98.84 | 85.76 | 91.18 | 97.03 | 99.83 | 100.00 | 100.00 | 100.00 |
| Other genotypes | 55 | 98.61 | 82.89 | 89.41 | 96.22 | 99.78 | 100.00 | 100.00 | 100.00 |
| Other genotypes | 56 | 98.33 | 80.37 | 87.45 | 95.28 | 99.70 | 100.00 | 100.00 | 100.00 |
| Other genotypes | 57 | 97.98 | 77.41 | 85.36 | 94.33 | 99.62 | 100.00 | 100.00 | 100.00 |
| Other genotypes | 58 | 97.60 | 74.69 | 83.09 | 93.32 | 99.50 | 100.00 | 100.00 | 100.00 |
| Other genotypes | 59 | 97.15 | 72.40 | 80.66 | 91.96 | 99.35 | 100.00 | 100.00 | 100.00 |
| Other genotypes | 60 | 96.68 | 71.01 | 77.71 | 90.52 | 99.19 | 100.00 | 100.00 | 100.00 |
| Other genotypes | 61 | 96.19 | 67.06 | 75.39 | 88.84 | 99.04 | 100.00 | 100.00 | 100.00 |
| Other genotypes | 62 | 95.64 | 61.40 | 72.75 | 87.29 | 98.84 | 100.00 | 100.00 | 100.00 |
| Other genotypes | 63 | 95.01 | 56.41 | 68.84 | 85.25 | 98.61 | 100.00 | 100.00 | 100.00 |
| Other genotypes | 64 | 94.30 | 51.33 | 64.25 | 82.84 | 98.41 | 100.00 | 100.00 | 100.00 |
| Other genotypes | 65 | 93.49 | 43.54 | 58.86 | 80.14 | 98.24 | 100.00 | 100.00 | 100.00 |
| Other genotypes | 66 | 92.49 | 41.07 | 52.69 | 76.79 | 97.94 | 100.00 | 100.00 | 100.00 |

**Table S12** **Posterior distribution of the difference in the number of postcodes with a higher expected prevalence under opportunistic screening versus organised screening at each age, relative to females aged 30, as of end of November 2023. Table counterpart to Figure 3D.**

| | | Posterior distribution | | | | | | | | |
|---|---|---|---|---|---|---|---|---|---|---|
| | | | Percentiles | | | | | | | |
| Virus | Age | Average | 0.01 | 0.025 | 0.1 | 0.5 | 0.9 | 0.975 | 0.99 | Proba < 0 |
| HPV16/18 | 31 | 34 | -21 | -8 | -1 | 0 | 83 | 363 | 595 | 0.12 |
| HPV16/18 | 32 | 56 | -47 | -16 | -2 | 0 | 140 | 606 | 1,010 | 0.15 |
| HPV16/18 | 33 | 71 | -72 | -30 | -3 | 0 | 174 | 784 | 1,272 | 0.17 |
| HPV16/18 | 34 | 80 | -100 | -44 | -5 | 0 | 196 | 915 | 1,466 | 0.19 |
| HPV16/18 | 35 | 85 | -149 | -58 | -7 | 0 | 210 | 1,016 | 1,559 | 0.21 |
| HPV16/18 | 36 | 88 | -185 | -84 | -10 | 0 | 219 | 1,060 | 1,623 | 0.23 |
| HPV16/18 | 37 | 88 | -246 | -115 | -14 | 0 | 223 | 1,087 | 1,689 | 0.26 |

**Table S12 Posterior distribution of the difference in the number of postcodes with a higher expected prevalence under opportunistic screening versus organised screening at each age, relative to females aged 30, as of end of November 2023. Table counterpart to Figure 3D.** ***(continued)***

| | | Posterior distribution | | | | | | | | |
|---|---|---|---|---|---|---|---|---|---|---|
| | | | Percentiles | | | | | | | |
| Virus | Age | Average | 0.01 | 0.025 | 0.1 | 0.5 | 0.9 | 0.975 | 0.99 | Proba < 0 |
| HPV16/18 | 38 | 88 | -306 | -158 | -18 | 0 | 227 | 1,103 | 1,732 | 0.27 |
| HPV16/18 | 39 | 87 | -384 | -196 | -26 | 0 | 236 | 1,143 | 1,854 | 0.28 |
| HPV16/18 | 40 | 84 | -462 | -219 | -34 | 0 | 232 | 1,155 | 1,857 | 0.30 |
| HPV16/18 | 41 | 80 | -542 | -254 | -43 | 0 | 233 | 1,130 | 1,878 | 0.31 |
| HPV16/18 | 42 | 76 | -607 | -304 | -50 | 0 | 236 | 1,126 | 1,908 | 0.33 |
| HPV16/18 | 43 | 72 | -700 | -341 | -55 | 0 | 232 | 1,118 | 1,921 | 0.34 |
| HPV16/18 | 44 | 68 | -762 | -398 | -65 | 0 | 235 | 1,121 | 1,903 | 0.35 |
| HPV16/18 | 45 | 63 | -869 | -443 | -81 | 0 | 231 | 1,136 | 1,907 | 0.36 |
| HPV16/18 | 46 | 58 | -914 | -491 | -89 | 0 | 229 | 1,133 | 1,907 | 0.37 |
| HPV16/18 | 47 | 52 | -908 | -525 | -99 | 0 | 227 | 1,117 | 1,905 | 0.39 |
| HPV16/18 | 48 | 46 | -1,030 | -571 | -112 | 0 | 220 | 1,124 | 1,902 | 0.40 |
| HPV16/18 | 49 | 40 | -1,104 | -652 | -133 | 0 | 220 | 1,128 | 1,900 | 0.41 |
| HPV16/18 | 50 | 32 | -1,154 | -713 | -146 | 0 | 209 | 1,115 | 1,908 | 0.43 |
| HPV16/18 | 51 | 24 | -1,297 | -791 | -162 | 0 | 207 | 1,113 | 1,886 | 0.44 |
| HPV16/18 | 52 | 14 | -1,376 | -839 | -182 | 0 | 191 | 1,076 | 1,886 | 0.46 |
| HPV16/18 | 53 | 3 | -1,479 | -892 | -208 | 0 | 187 | 1,023 | 1,895 | 0.47 |
| HPV16/18 | 54 | -11 | -1,569 | -1,001 | -245 | 0 | 174 | 979 | 1,898 | 0.49 |
| HPV16/18 | 55 | -26 | -1,645 | -1,133 | -304 | -1 | 163 | 951 | 1,842 | 0.51 |
| HPV16/18 | 56 | -43 | -1,850 | -1,275 | -352 | -1 | 150 | 951 | 1,878 | 0.52 |
| HPV16/18 | 57 | -62 | -2,086 | -1,353 | -406 | -2 | 139 | 911 | 1,854 | 0.54 |
| HPV16/18 | 58 | -83 | -2,271 | -1,536 | -450 | -3 | 129 | 876 | 1,804 | 0.56 |
| HPV16/18 | 59 | -106 | -2,325 | -1,623 | -515 | -4 | 108 | 848 | 1,774 | 0.58 |
| HPV16/18 | 60 | -131 | -2,510 | -1,807 | -611 | -5 | 101 | 780 | 1,676 | 0.59 |
| HPV16/18 | 61 | -160 | -2,712 | -2,001 | -719 | -7 | 93 | 762 | 1,588 | 0.60 |
| HPV16/18 | 62 | -193 | -3,054 | -2,179 | -818 | -9 | 85 | 731 | 1,565 | 0.61 |
| HPV16/18 | 63 | -234 | -3,556 | -2,458 | -971 | -12 | 76 | 701 | 1,516 | 0.63 |
| HPV16/18 | 64 | -282 | -3,899 | -2,726 | -1,195 | -15 | 73 | 697 | 1,485 | 0.64 |
| HPV16/18 | 65 | -338 | -4,244 | -3,146 | -1,420 | -18 | 65 | 657 | 1,432 | 0.65 |

**Table S12 Posterior distribution of the difference in the number of postcodes with a higher expected prevalence under opportunistic screening versus organised screening at each age, relative to females aged 30, as of end of November 2023. Table counterpart to Figure 3D.** ***(continued)***

| Virus | Age | Posterior distribution | | | | | | | | |
|---|---|---|---|---|---|---|---|---|---|---|
| | | | Percentiles | | | | | | | |
| | | Average | 0.01 | 0.025 | 0.1 | 0.5 | 0.9 | 0.975 | 0.99 | Proba < 0 |
| HPV16/18 | 66 | -403 | -4,592 | -3,597 | -1,653 | -23 | 61 | 636 | 1,467 | 0.66 |
| Other genotypes | 31 | 1 | -6 | -3 | 0 | 0 | 0 | 6 | 15 | 0.10 |
| Other genotypes | 32 | 1 | -13 | -6 | -1 | 0 | 1 | 8 | 22 | 0.14 |
| Other genotypes | 33 | 1 | -19 | -10 | -2 | 0 | 1 | 10 | 25 | 0.19 |
| Other genotypes | 34 | 0 | -28 | -15 | -3 | 0 | 1 | 11 | 28 | 0.22 |
| Other genotypes | 35 | 0 | -41 | -19 | -4 | 0 | 1 | 11 | 32 | 0.25 |
| Other genotypes | 36 | -1 | -50 | -24 | -5 | 0 | 1 | 11 | 35 | 0.28 |
| Other genotypes | 37 | -1 | -64 | -30 | -7 | 0 | 0 | 10 | 34 | 0.31 |
| Other genotypes | 38 | -2 | -77 | -37 | -8 | 0 | 0 | 10 | 33 | 0.34 |
| Other genotypes | 39 | -3 | -91 | -45 | -10 | 0 | 0 | 10 | 31 | 0.37 |
| Other genotypes | 40 | -4 | -110 | -57 | -13 | 0 | 0 | 8 | 30 | 0.40 |
| Other genotypes | 41 | -6 | -129 | -71 | -17 | 0 | 0 | 8 | 31 | 0.43 |
| Other genotypes | 42 | -8 | -150 | -85 | -20 | 0 | 0 | 8 | 32 | 0.45 |
| Other genotypes | 43 | -9 | -178 | -103 | -24 | 0 | 0 | 7 | 29 | 0.47 |
| Other genotypes | 44 | -11 | -210 | -120 | -29 | 0 | 0 | 6 | 27 | 0.49 |
| Other genotypes | 45 | -14 | -239 | -143 | -34 | -1 | 0 | 6 | 29 | 0.52 |
| Other genotypes | 46 | -16 | -277 | -163 | -42 | -1 | 0 | 5 | 22 | 0.55 |
| Other genotypes | 47 | -20 | -326 | -191 | -49 | -1 | 0 | 4 | 18 | 0.57 |
| Other genotypes | 48 | -23 | -357 | -216 | -58 | -2 | 0 | 3 | 13 | 0.59 |
| Other genotypes | 49 | -27 | -432 | -244 | -66 | -2 | 0 | 2 | 11 | 0.61 |
| Other genotypes | 50 | -31 | -446 | -276 | -79 | -3 | 0 | 1 | 11 | 0.64 |
| Other genotypes | 51 | -37 | -514 | -315 | -94 | -4 | 0 | 0 | 6 | 0.66 |
| Other genotypes | 52 | -44 | -591 | -362 | -113 | -5 | 0 | 0 | 6 | 0.68 |
| Other genotypes | 53 | -53 | -713 | -426 | -138 | -6 | 0 | 0 | 3 | 0.70 |
| Other genotypes | 54 | -65 | -811 | -513 | -169 | -9 | 0 | 0 | 1 | 0.73 |
| Other genotypes | 55 | -79 | -995 | -598 | -217 | -12 | 0 | 0 | 0 | 0.75 |
| Other genotypes | 56 | -96 | -1,136 | -716 | -269 | -17 | 0 | 0 | 0 | 0.78 |
| Other genotypes | 57 | -116 | -1,324 | -837 | -323 | -22 | 0 | 0 | 0 | 0.80 |

**Table S12 Posterior distribution of the difference in the number of postcodes with a higher expected prevalence under opportunistic screening versus organised screening at each age, relative to females aged 30, as of end of November 2023. Table counterpart to Figure 3D. *(continued)***

| | | Posterior distribution | | | | | | | | |
|---|---|---|---|---|---|---|---|---|---|---|
| | | | Percentiles | | | | | | | |
| Virus | Age | Average | 0.01 | 0.025 | 0.1 | 0.5 | 0.9 | 0.975 | 0.99 | Proba < 0 |
| Other genotypes | 58 | -140 | -1,484 | -981 | -392 | -29 | 0 | 0 | 0 | 0.83 |
| Other genotypes | 59 | -166 | -1,622 | -1,122 | -476 | -37 | 0 | 0 | 0 | 0.84 |
| Other genotypes | 60 | -194 | -1,730 | -1,295 | -564 | -47 | 0 | 0 | 0 | 0.86 |
| Other genotypes | 61 | -224 | -1,948 | -1,442 | -663 | -56 | 0 | 0 | 0 | 0.87 |
| Other genotypes | 62 | -258 | -2,235 | -1,626 | -753 | -69 | 0 | 0 | 0 | 0.88 |
| Other genotypes | 63 | -296 | -2,579 | -1,854 | -872 | -81 | 0 | 0 | 0 | 0.89 |
| Other genotypes | 64 | -339 | -2,914 | -2,134 | -1,017 | -94 | 0 | 0 | 0 | 0.89 |
| Other genotypes | 65 | -388 | -3,335 | -2,459 | -1,185 | -104 | 0 | 0 | 0 | 0.89 |
| Other genotypes | 66 | -448 | -3,514 | -2,821 | -1,380 | -122 | 0 | 0 | 0 | 0.89 |

### S13 Difference in expected HR HPV cervical prevalence between the last and first week of the study period, stratified by age

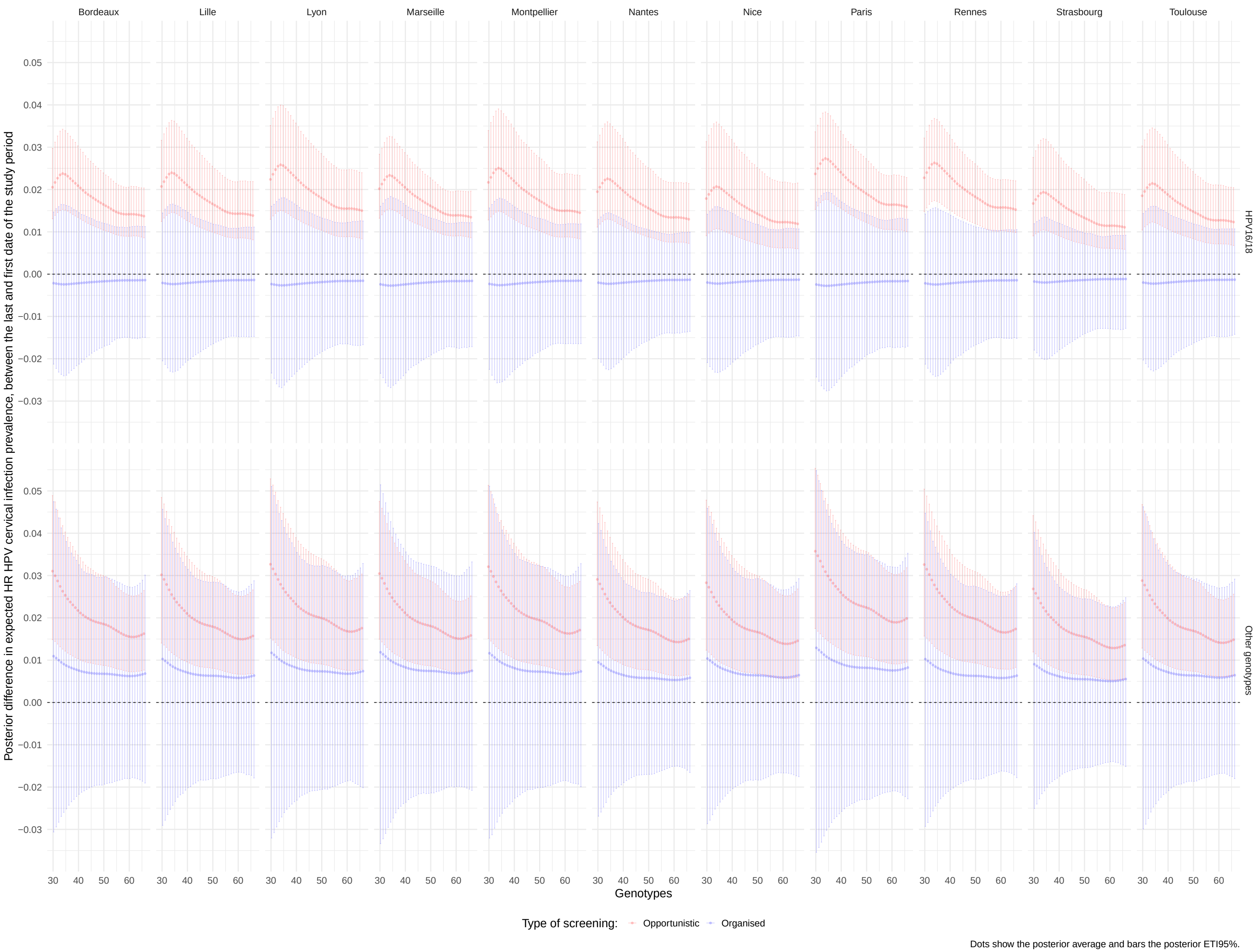

**Figure S39** Difference in expected HR HPV cervical infection prevalence, between the last and first week of the study period in major French cities, stratified by type of screening and age, for HPV16/18.

#### S14 Figure 4B and C in table format

**Table S13 Posterior expected HR HPV infection prevalence (in %) assuming all data would have been collected with opportunistic or organised screening, not stratified or stratified by age, year, or age and year. Provide values from Figure 4B in table format.**

| | | | Posterior distribution | | | | | | | |
|---|---|---|---|---|---|---|---|---|---|---|
| | | | | Percentiles | | | | | | |
| Genotype | Year | Age | Average | 0.01 | 0.025 | 0.1 | 0.5 | 0.9 | 0.975 | 0.99 |
| HPV16/18 | 2020 | | 3.19 | 3.05 | 3.08 | 3.12 | 3.19 | 3.27 | 3.32 | 3.34 |
| HPV16/18 | 2021 | | 3.65 | 3.54 | 3.56 | 3.59 | 3.65 | 3.70 | 3.74 | 3.75 |
| HPV16/18 | 2022 | | 4.03 | 3.86 | 3.89 | 3.93 | 4.02 | 4.12 | 4.17 | 4.19 |
| HPV16/18 | 2023 | | 4.50 | 4.31 | 4.34 | 4.40 | 4.50 | 4.61 | 4.67 | 4.70 |
| HPV16/18 | 2020 | | 3.44 | 2.38 | 2.50 | 2.80 | 3.39 | 4.15 | 4.61 | 4.81 |
| HPV16/18 | 2021 | | 3.31 | 2.66 | 2.77 | 2.94 | 3.30 | 3.70 | 3.95 | 4.10 |
| HPV16/18 | 2022 | | 3.08 | 2.72 | 2.76 | 2.87 | 3.07 | 3.29 | 3.41 | 3.47 |
| HPV16/18 | 2023 | | 3.13 | 2.69 | 2.76 | 2.88 | 3.13 | 3.38 | 3.51 | 3.60 |
| Other genotypes | 2020 | | 9.45 | 9.23 | 9.26 | 9.32 | 9.45 | 9.58 | 9.65 | 9.69 |
| Other genotypes | 2021 | | 8.80 | 8.64 | 8.67 | 8.70 | 8.80 | 8.89 | 8.95 | 8.97 |
| Other genotypes | 2022 | | 9.30 | 9.07 | 9.11 | 9.17 | 9.30 | 9.44 | 9.51 | 9.54 |
| Other genotypes | 2023 | | 10.95 | 10.66 | 10.70 | 10.78 | 10.95 | 11.11 | 11.21 | 11.25 |
| Other genotypes | 2020 | | 7.17 | 5.49 | 5.72 | 6.14 | 7.13 | 8.27 | 8.89 | 9.26 |
| Other genotypes | 2021 | | 7.02 | 6.09 | 6.20 | 6.49 | 7.01 | 7.57 | 7.87 | 8.04 |
| Other genotypes | 2022 | | 7.47 | 6.88 | 6.94 | 7.13 | 7.47 | 7.81 | 7.97 | 8.07 |
| Other genotypes | 2023 | | 7.75 | 7.07 | 7.16 | 7.34 | 7.74 | 8.15 | 8.40 | 8.50 |
| HPV16/18 | | [30,39] | 4.24 | 3.24 | 3.37 | 3.59 | 4.50 | 4.65 | 4.70 | 4.73 |
| HPV16/18 | | (39,49] | 3.49 | 2.71 | 2.80 | 3.00 | 3.67 | 3.81 | 3.86 | 3.89 |
| HPV16/18 | | (49,59] | 2.83 | 2.28 | 2.34 | 2.49 | 2.93 | 3.05 | 3.11 | 3.14 |
| HPV16/18 | | (59,66] | 2.69 | 2.16 | 2.24 | 2.40 | 2.74 | 2.89 | 2.97 | 3.06 |
| Other genotypes | | [30,39] | 10.64 | 8.03 | 8.20 | 8.53 | 11.33 | 12.38 | 12.45 | 12.48 |
| Other genotypes | | (39,49] | 7.60 | 5.89 | 6.00 | 6.27 | 8.19 | 8.66 | 8.73 | 8.76 |

**Table S13** **Posterior expected HR HPV infection prevalence (in %) assuming all data would have been collected with opportunistic or organised screening, not stratified or stratified by age, year, or age and year. Provide values from Figure 4B in table format.** ***(continued)***

| | | | Posterior distribution | | | | | | | |
|---|---|---|---|---|---|---|---|---|---|---|
| | | | | Percentiles | | | | | | |
| Genotype | Year | Age | Average | 0.01 | 0.025 | 0.1 | 0.5 | 0.9 | 0.975 | 0.99 |
| Other genotypes | | (49,59] | 6.67 | 5.46 | 5.55 | 5.80 | 7.03 | 7.34 | 7.42 | 7.46 |
| Other genotypes | | (59,66] | 6.17 | 5.23 | 5.33 | 5.60 | 6.31 | 6.57 | 6.68 | 6.73 |
| HPV16/18 | 2020 | [30,39] | 3.87 | 3.67 | 3.71 | 3.76 | 3.86 | 3.97 | 4.04 | 4.07 |
| HPV16/18 | 2020 | (39,49] | 3.18 | 3.01 | 3.05 | 3.09 | 3.18 | 3.27 | 3.32 | 3.35 |
| HPV16/18 | 2020 | (49,59] | 2.53 | 2.39 | 2.41 | 2.45 | 2.53 | 2.62 | 2.65 | 2.68 |
| HPV16/18 | 2020 | (59,66] | 2.35 | 2.17 | 2.20 | 2.25 | 2.35 | 2.45 | 2.50 | 2.53 |
| HPV16/18 | 2021 | [30,39] | 4.46 | 4.30 | 4.33 | 4.37 | 4.46 | 4.54 | 4.58 | 4.60 |
| HPV16/18 | 2021 | (39,49] | 3.65 | 3.50 | 3.53 | 3.57 | 3.65 | 3.73 | 3.78 | 3.81 |
| HPV16/18 | 2021 | (49,59] | 2.91 | 2.77 | 2.79 | 2.84 | 2.91 | 2.99 | 3.02 | 3.06 |
| HPV16/18 | 2021 | (59,66] | 2.70 | 2.53 | 2.55 | 2.60 | 2.70 | 2.80 | 2.86 | 2.89 |
| HPV16/18 | 2022 | [30,39] | 4.89 | 4.67 | 4.70 | 4.77 | 4.89 | 5.01 | 5.08 | 5.11 |
| HPV16/18 | 2022 | (39,49] | 4.01 | 3.82 | 3.84 | 3.90 | 4.01 | 4.11 | 4.18 | 4.21 |
| HPV16/18 | 2022 | (49,59] | 3.19 | 3.03 | 3.05 | 3.09 | 3.19 | 3.29 | 3.34 | 3.37 |
| HPV16/18 | 2022 | (59,66] | 2.95 | 2.73 | 2.77 | 2.83 | 2.95 | 3.08 | 3.14 | 3.17 |
| HPV16/18 | 2023 | [30,39] | 5.45 | 5.20 | 5.23 | 5.31 | 5.45 | 5.59 | 5.66 | 5.71 |
| HPV16/18 | 2023 | (39,49] | 4.50 | 4.27 | 4.31 | 4.36 | 4.50 | 4.63 | 4.70 | 4.74 |
| HPV16/18 | 2023 | (49,59] | 3.58 | 3.38 | 3.41 | 3.47 | 3.58 | 3.70 | 3.76 | 3.81 |
| HPV16/18 | 2023 | (59,66] | 3.31 | 3.05 | 3.10 | 3.17 | 3.31 | 3.45 | 3.52 | 3.56 |
| HPV16/18 | 2020 | [30,39] | 4.09 | 2.77 | 2.94 | 3.27 | 4.03 | 5.00 | 5.56 | 5.90 |
| HPV16/18 | 2020 | (39,49] | 3.39 | 2.33 | 2.43 | 2.73 | 3.34 | 4.11 | 4.55 | 4.79 |
| HPV16/18 | 2020 | (49,59] | 2.80 | 1.92 | 2.04 | 2.28 | 2.76 | 3.36 | 3.73 | 3.89 |
| HPV16/18 | 2020 | (59,66] | 2.71 | 1.91 | 1.99 | 2.21 | 2.68 | 3.25 | 3.62 | 3.82 |
| HPV16/18 | 2021 | [30,39] | 3.97 | 3.11 | 3.24 | 3.44 | 3.95 | 4.51 | 4.86 | 5.08 |
| HPV16/18 | 2021 | (39,49] | 3.28 | 2.60 | 2.67 | 2.87 | 3.27 | 3.70 | 3.94 | 4.09 |
| HPV16/18 | 2021 | (49,59] | 2.71 | 2.17 | 2.25 | 2.40 | 2.71 | 3.05 | 3.22 | 3.33 |

**Table S13** **Posterior expected HR HPV infection prevalence (in %) assuming all data would have been collected with opportunistic or organised screening, not stratified or stratified by age, year, or age and year. Provide values from Figure 4B in table format.** ***(continued)***

| Genotype | Year | Age | Posterior distribution | | | | | | | |
|---|---|---|---|---|---|---|---|---|---|---|
| | | | | Percentiles | | | | | | |
| | | | Average | 0.01 | 0.025 | 0.1 | 0.5 | 0.9 | 0.975 | 0.99 |
| HPV16/18 | 2021 | (59,66] | 2.64 | 2.10 | 2.16 | 2.30 | 2.63 | 2.97 | 3.20 | 3.28 |
| HPV16/18 | 2022 | [30,39] | 3.67 | 3.09 | 3.19 | 3.35 | 3.66 | 4.01 | 4.18 | 4.27 |
| HPV16/18 | 2022 | (39,49] | 3.03 | 2.60 | 2.67 | 2.79 | 3.02 | 3.28 | 3.41 | 3.49 |
| HPV16/18 | 2022 | (49,59] | 2.50 | 2.17 | 2.20 | 2.30 | 2.50 | 2.71 | 2.82 | 2.90 |
| HPV16/18 | 2022 | (59,66] | 2.42 | 1.98 | 2.05 | 2.17 | 2.42 | 2.70 | 2.84 | 2.93 |
| HPV16/18 | 2023 | [30,39] | 3.73 | 3.12 | 3.19 | 3.36 | 3.73 | 4.12 | 4.27 | 4.40 |
| HPV16/18 | 2023 | (39,49] | 3.09 | 2.62 | 2.69 | 2.82 | 3.08 | 3.36 | 3.51 | 3.61 |
| HPV16/18 | 2023 | (49,59] | 2.55 | 2.13 | 2.19 | 2.32 | 2.54 | 2.80 | 2.95 | 3.04 |
| HPV16/18 | 2023 | (59,66] | 2.46 | 1.92 | 2.02 | 2.18 | 2.45 | 2.76 | 2.92 | 3.06 |
| Other genotypes | 2020 | [30,39] | 12.31 | 11.99 | 12.03 | 12.12 | 12.30 | 12.49 | 12.59 | 12.63 |
| Other genotypes | 2020 | (39,49] | 8.65 | 8.39 | 8.42 | 8.50 | 8.64 | 8.79 | 8.87 | 8.91 |
| Other genotypes | 2020 | (49,59] | 7.31 | 7.07 | 7.10 | 7.16 | 7.30 | 7.45 | 7.53 | 7.58 |
| Other genotypes | 2020 | (59,66] | 6.48 | 6.16 | 6.22 | 6.30 | 6.47 | 6.66 | 6.75 | 6.81 |
| Other genotypes | 2021 | [30,39] | 11.58 | 11.33 | 11.36 | 11.44 | 11.58 | 11.72 | 11.80 | 11.84 |
| Other genotypes | 2021 | (39,49] | 8.12 | 7.91 | 7.94 | 8.00 | 8.12 | 8.23 | 8.30 | 8.33 |
| Other genotypes | 2021 | (49,59] | 6.85 | 6.64 | 6.68 | 6.74 | 6.85 | 6.97 | 7.04 | 7.08 |
| Other genotypes | 2021 | (59,66] | 6.12 | 5.84 | 5.88 | 5.96 | 6.11 | 6.27 | 6.37 | 6.41 |
| Other genotypes | 2022 | [30,39] | 12.15 | 11.82 | 11.87 | 11.97 | 12.15 | 12.34 | 12.43 | 12.46 |
| Other genotypes | 2022 | (39,49] | 8.47 | 8.23 | 8.25 | 8.33 | 8.47 | 8.62 | 8.70 | 8.75 |
| Other genotypes | 2022 | (49,59] | 7.17 | 6.89 | 6.95 | 7.02 | 7.16 | 7.31 | 7.39 | 7.45 |
| Other genotypes | 2022 | (59,66] | 6.36 | 6.06 | 6.10 | 6.19 | 6.36 | 6.54 | 6.64 | 6.69 |
| Other genotypes | 2023 | [30,39] | 14.28 | 13.88 | 13.94 | 14.05 | 14.28 | 14.51 | 14.63 | 14.67 |
| Other genotypes | 2023 | (39,49] | 9.99 | 9.67 | 9.71 | 9.80 | 9.99 | 10.18 | 10.28 | 10.32 |
| Other genotypes | 2023 | (49,59] | 8.45 | 8.15 | 8.20 | 8.28 | 8.45 | 8.64 | 8.73 | 8.80 |
| Other genotypes | 2023 | (59,66] | 7.49 | 7.12 | 7.19 | 7.28 | 7.49 | 7.70 | 7.82 | 7.88 |

**Table S13 Posterior expected HR HPV infection prevalence (in %) assuming all data would have been collected with opportunistic or organised screening, not stratified or stratified by age, year, or age and year. Provide values from Figure 4B in table format.** ***(continued)***

| | | | Posterior distribution | | | | | | | |
|---|---|---|---|---|---|---|---|---|---|---|
| | | | | Percentiles | | | | | | |
| Genotype | Year | Age | Average | 0.01 | 0.025 | 0.1 | 0.5 | 0.9 | 0.975 | 0.99 |
| Other genotypes | 2020 | [30,39] | 8.83 | 6.68 | 7.04 | 7.56 | 8.78 | 10.24 | 11.05 | 11.49 |
| Other genotypes | 2020 | (39,49] | 6.53 | 4.93 | 5.15 | 5.54 | 6.50 | 7.56 | 8.21 | 8.55 |
| Other genotypes | 2020 | (49,59] | 6.00 | 4.59 | 4.77 | 5.09 | 5.95 | 6.94 | 7.51 | 7.87 |
| Other genotypes | 2020 | (59,66] | 5.78 | 4.45 | 4.58 | 4.94 | 5.75 | 6.63 | 7.17 | 7.59 |
| Other genotypes | 2021 | [30,39] | 8.69 | 7.45 | 7.62 | 7.98 | 8.67 | 9.47 | 9.92 | 10.11 |
| Other genotypes | 2021 | (39,49] | 6.42 | 5.48 | 5.60 | 5.85 | 6.41 | 7.02 | 7.36 | 7.53 |
| Other genotypes | 2021 | (49,59] | 5.90 | 5.08 | 5.17 | 5.42 | 5.90 | 6.41 | 6.64 | 6.79 |
| Other genotypes | 2021 | (59,66] | 5.74 | 4.91 | 5.00 | 5.24 | 5.75 | 6.23 | 6.49 | 6.62 |
| Other genotypes | 2022 | [30,39] | 9.20 | 8.28 | 8.39 | 8.67 | 9.19 | 9.77 | 10.04 | 10.22 |
| Other genotypes | 2022 | (39,49] | 6.77 | 6.11 | 6.19 | 6.38 | 6.76 | 7.15 | 7.39 | 7.51 |
| Other genotypes | 2022 | (49,59] | 6.24 | 5.62 | 5.70 | 5.87 | 6.23 | 6.61 | 6.80 | 6.87 |
| Other genotypes | 2022 | (59,66] | 6.03 | 5.30 | 5.40 | 5.58 | 6.02 | 6.49 | 6.75 | 6.88 |
| Other genotypes | 2023 | [30,39] | 9.57 | 8.59 | 8.70 | 8.95 | 9.56 | 10.20 | 10.54 | 10.77 |
| Other genotypes | 2023 | (39,49] | 7.01 | 6.27 | 6.38 | 6.59 | 7.00 | 7.47 | 7.71 | 7.88 |
| Other genotypes | 2023 | (49,59] | 6.45 | 5.75 | 5.85 | 6.04 | 6.44 | 6.86 | 7.08 | 7.19 |
| Other genotypes | 2023 | (59,66] | 6.21 | 5.38 | 5.50 | 5.72 | 6.19 | 6.73 | 6.99 | 7.13 |

**Table S14 Marginal Difference in Expected Prevalence (in percentage points), not stratified or stratified by age, year, or age and year. Provide values from Figure 4C in table format.**

| | | | Posterior distribution | | | | | | | | |
|---|---|---|---|---|---|---|---|---|---|---|---|
| | | | | Percentiles | | | | | | | |
| Genotype | Year | Age | Average | 0.01 | 0.025 | 0.1 | 0.5 | 0.9 | 0.975 | 0.99 | Pr(MDEP>0) |
| HPV16/18 | 2020 | | -0.25 | -1.60 | -1.38 | -0.96 | -0.21 | 0.40 | 0.71 | 0.85 | 0.33 |
| HPV16/18 | 2021 | | 0.34 | -0.47 | -0.29 | -0.06 | 0.35 | 0.72 | 0.89 | 1.00 | 0.86 |
| HPV16/18 | 2022 | | 0.95 | 0.52 | 0.61 | 0.73 | 0.95 | 1.17 | 1.28 | 1.35 | 1.00 |
| HPV16/18 | 2023 | | 1.37 | 0.89 | 0.96 | 1.11 | 1.37 | 1.64 | 1.79 | 1.85 | 1.00 |
| Other genotypes | 2020 | | 2.28 | 0.19 | 0.52 | 1.21 | 2.33 | 3.31 | 3.73 | 3.95 | 0.99 |
| Other genotypes | 2021 | | 1.78 | 0.75 | 0.90 | 1.21 | 1.80 | 2.32 | 2.62 | 2.73 | 1.00 |
| Other genotypes | 2022 | | 1.83 | 1.20 | 1.29 | 1.46 | 1.83 | 2.20 | 2.38 | 2.47 | 1.00 |
| Other genotypes | 2023 | | 3.20 | 2.40 | 2.51 | 2.77 | 3.21 | 3.63 | 3.84 | 3.96 | 1.00 |
| HPV16/18 | | [30,39] | 0.71 | -0.16 | -0.02 | 0.25 | 0.73 | 1.14 | 1.34 | 1.45 | 0.97 |
| HPV16/18 | | (39,49] | 0.53 | -0.11 | 0.00 | 0.18 | 0.54 | 0.87 | 1.03 | 1.09 | 0.97 |
| HPV16/18 | | (49,59] | 0.34 | -0.18 | -0.08 | 0.07 | 0.35 | 0.60 | 0.72 | 0.78 | 0.94 |
| HPV16/18 | | (59,66] | 0.22 | -0.34 | -0.27 | -0.08 | 0.23 | 0.51 | 0.62 | 0.69 | 0.83 |
| Other genotypes | | [30,39] | 3.33 | 2.14 | 2.29 | 2.64 | 3.35 | 3.96 | 4.23 | 4.39 | 1.00 |
| Other genotypes | | (39,49] | 1.99 | 0.98 | 1.16 | 1.48 | 2.01 | 2.47 | 2.70 | 2.82 | 1.00 |
| Other genotypes | | (49,59] | 1.19 | 0.42 | 0.55 | 0.77 | 1.21 | 1.60 | 1.78 | 1.88 | 1.00 |
| Other genotypes | | (59,66] | 0.58 | -0.23 | -0.12 | 0.13 | 0.57 | 1.03 | 1.26 | 1.35 | 0.95 |
| HPV16/18 | 2020 | [30,39] | -0.23 | -2.05 | -1.64 | -1.15 | -0.17 | 0.60 | 0.96 | 1.12 | 0.40 |
| HPV16/18 | 2021 | [30,39] | 0.49 | -0.63 | -0.44 | -0.06 | 0.51 | 1.02 | 1.24 | 1.40 | 0.87 |
| HPV16/18 | 2022 | [30,39] | 1.22 | 0.59 | 0.67 | 0.86 | 1.23 | 1.55 | 1.71 | 1.82 | 1.00 |
| HPV16/18 | 2023 | [30,39] | 1.72 | 1.02 | 1.14 | 1.35 | 1.72 | 2.11 | 2.26 | 2.38 | 1.00 |
| HPV16/18 | 2020 | (39,49] | -0.21 | -1.59 | -1.35 | -0.93 | -0.18 | 0.44 | 0.76 | 0.88 | 0.36 |
| HPV16/18 | 2021 | (39,49] | 0.37 | -0.42 | -0.29 | -0.07 | 0.38 | 0.79 | 0.97 | 1.06 | 0.87 |
| HPV16/18 | 2022 | (39,49] | 0.98 | 0.51 | 0.59 | 0.72 | 0.99 | 1.23 | 1.35 | 1.42 | 1.00 |
| HPV16/18 | 2023 | (39,49] | 1.41 | 0.87 | 0.95 | 1.13 | 1.42 | 1.69 | 1.83 | 1.90 | 1.00 |
| HPV16/18 | 2020 | (49,59] | -0.27 | -1.34 | -1.21 | -0.84 | -0.24 | 0.25 | 0.49 | 0.61 | 0.27 |
| HPV16/18 | 2021 | (49,59] | 0.20 | -0.43 | -0.35 | -0.14 | 0.21 | 0.52 | 0.67 | 0.75 | 0.79 |
| HPV16/18 | 2022 | (49,59] | 0.69 | 0.27 | 0.34 | 0.46 | 0.69 | 0.90 | 1.01 | 1.06 | 1.00 |

**Table S14 Marginal Difference in Expected Prevalence (in percentage points), not stratified or stratified by age, year, or age and year. Provide values from Figure 4C in table format.** ***(continued)***

| | | | Posterior distribution | | | | | | | | |
|---|---|---|---|---|---|---|---|---|---|---|---|
| | | | | Percentiles | | | | | | | |
| Genotype | Year | Age | Average | 0.01 | 0.025 | 0.1 | 0.5 | 0.9 | 0.975 | 0.99 | Pr(MDEP>0) |
| HPV16/18 | 2023 | (49,59] | 1.03 | 0.58 | 0.64 | 0.78 | 1.04 | 1.28 | 1.43 | 1.51 | 1.00 |
| HPV16/18 | 2020 | (59,66] | -0.37 | -1.48 | -1.27 | -0.91 | -0.33 | 0.14 | 0.38 | 0.44 | 0.19 |
| HPV16/18 | 2021 | (59,66] | 0.07 | -0.59 | -0.51 | -0.29 | 0.08 | 0.41 | 0.54 | 0.61 | 0.62 |
| HPV16/18 | 2022 | (59,66] | 0.53 | 0.00 | 0.11 | 0.24 | 0.54 | 0.81 | 0.93 | 1.01 | 0.99 |
| HPV16/18 | 2023 | (59,66] | 0.85 | 0.26 | 0.33 | 0.52 | 0.86 | 1.17 | 1.33 | 1.41 | 1.00 |
| Other genotypes | 2020 | [30,39] | 3.47 | 0.76 | 1.29 | 2.09 | 3.51 | 4.79 | 5.28 | 5.54 | 1.00 |
| Other genotypes | 2021 | [30,39] | 2.89 | 1.44 | 1.65 | 2.12 | 2.91 | 3.62 | 3.99 | 4.14 | 1.00 |
| Other genotypes | 2022 | [30,39] | 2.95 | 1.84 | 2.06 | 2.35 | 2.96 | 3.51 | 3.79 | 3.98 | 1.00 |
| Other genotypes | 2023 | [30,39] | 4.71 | 3.51 | 3.70 | 4.06 | 4.71 | 5.33 | 5.65 | 5.78 | 1.00 |
| Other genotypes | 2020 | (39,49] | 2.12 | 0.04 | 0.44 | 1.07 | 2.15 | 3.10 | 3.51 | 3.71 | 0.99 |
| Other genotypes | 2021 | (39,49] | 1.70 | 0.60 | 0.75 | 1.10 | 1.71 | 2.26 | 2.53 | 2.65 | 1.00 |
| Other genotypes | 2022 | (39,49] | 1.71 | 0.98 | 1.05 | 1.30 | 1.71 | 2.11 | 2.30 | 2.41 | 1.00 |
| Other genotypes | 2023 | (39,49] | 2.98 | 2.06 | 2.22 | 2.50 | 2.99 | 3.43 | 3.66 | 3.76 | 1.00 |
| Other genotypes | 2020 | (49,59] | 1.31 | -0.51 | -0.18 | 0.36 | 1.36 | 2.20 | 2.55 | 2.71 | 0.96 |
| Other genotypes | 2021 | (49,59] | 0.95 | 0.01 | 0.21 | 0.46 | 0.96 | 1.44 | 1.70 | 1.79 | 0.99 |
| Other genotypes | 2022 | (49,59] | 0.93 | 0.21 | 0.35 | 0.56 | 0.94 | 1.30 | 1.49 | 1.61 | 1.00 |
| Other genotypes | 2023 | (49,59] | 2.01 | 1.23 | 1.35 | 1.55 | 2.02 | 2.44 | 2.65 | 2.76 | 1.00 |
| Other genotypes | 2020 | (59,66] | 0.70 | -1.12 | -0.70 | -0.17 | 0.73 | 1.54 | 1.92 | 2.08 | 0.84 |
| Other genotypes | 2021 | (59,66] | 0.38 | -0.55 | -0.40 | -0.11 | 0.37 | 0.90 | 1.15 | 1.26 | 0.84 |
| Other genotypes | 2022 | (59,66] | 0.33 | -0.55 | -0.44 | -0.17 | 0.34 | 0.81 | 1.03 | 1.12 | 0.80 |
| Other genotypes | 2023 | (59,66] | 1.28 | 0.26 | 0.46 | 0.75 | 1.29 | 1.81 | 2.08 | 2.23 | 1.00 |

### S15 Sensitivity analyses

#### S15.1 Change in the variance of priors assigned to the (latent) correlation parameters

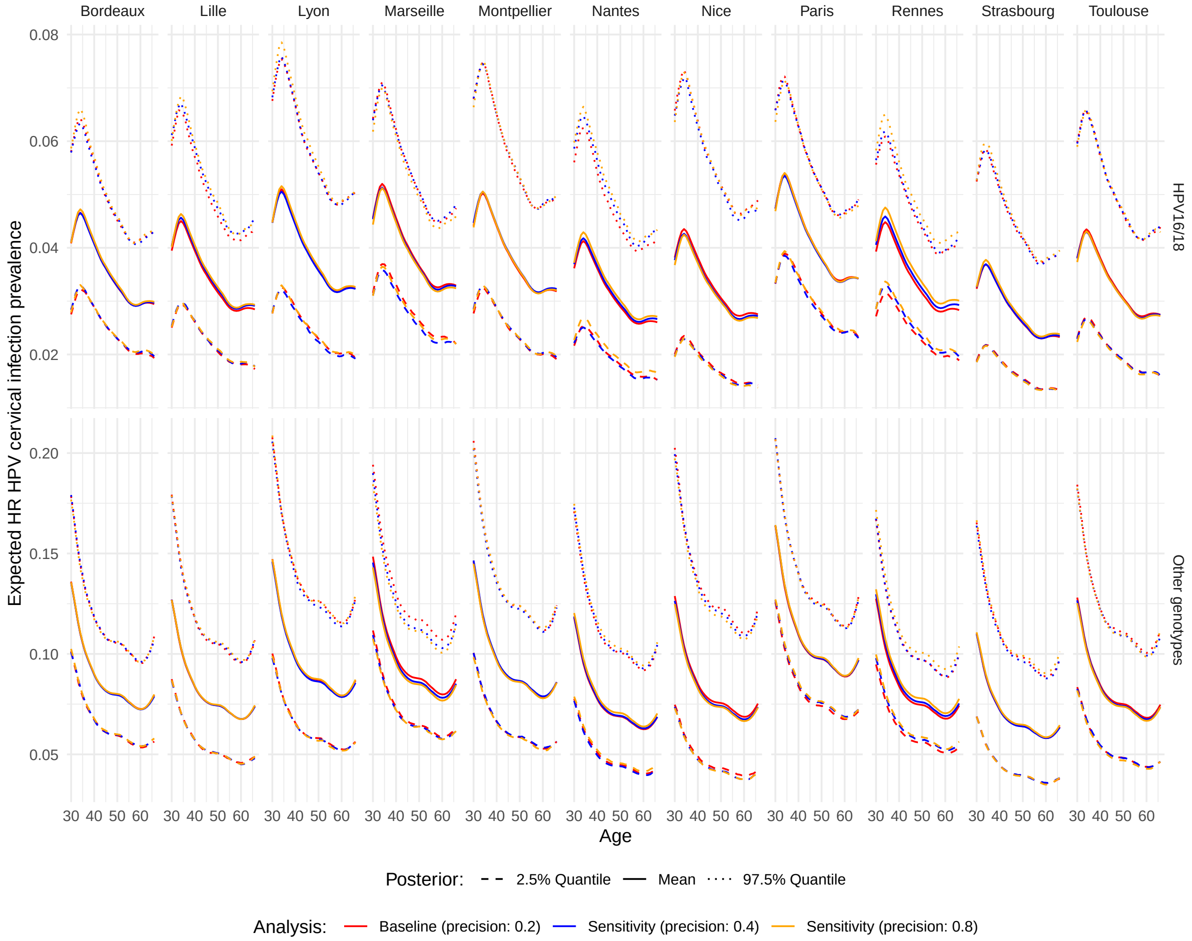

**Figure S40** **Posterior average (ETI95%) for the expected HR HPV cervical infection prevalence in 11 major cities in France. The precision is defined as 1/variance: the greater the precision, the lower the variance. The baseline case, precision = 0.2, has a U-shape. A precision of 0.4 looks like a uniform distribution on [-1,1]. At a precision of 0.8, extreme values for the correlation are very unlikely.**

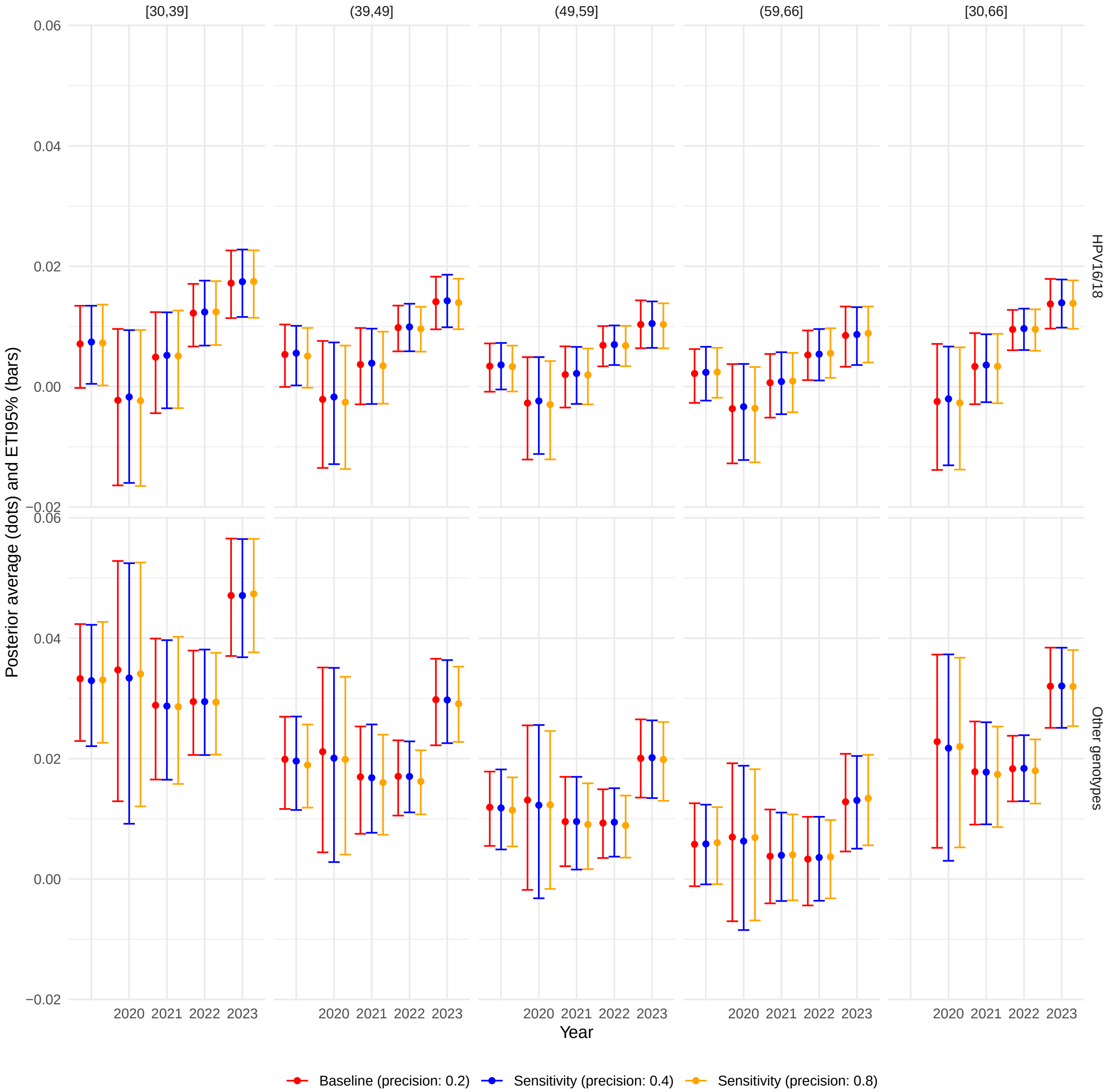

**Figure S41** **Posterior average (ETI95%) for the Marginal Difference in Expected Prevalence, stratified by precision for the priors of the latent correlation parameters, age, and year. The precision is defined as 1/variance: the greater the precision, the lower the variance. The baseline case, precision = 0.2, has a U-shape. A precision of 0.4 looks like a uniform distribution on [-1,1]. At a precision of 0.8, extreme values for the correlation are very unlikely.**

#### S15.2 Non-selected competing models

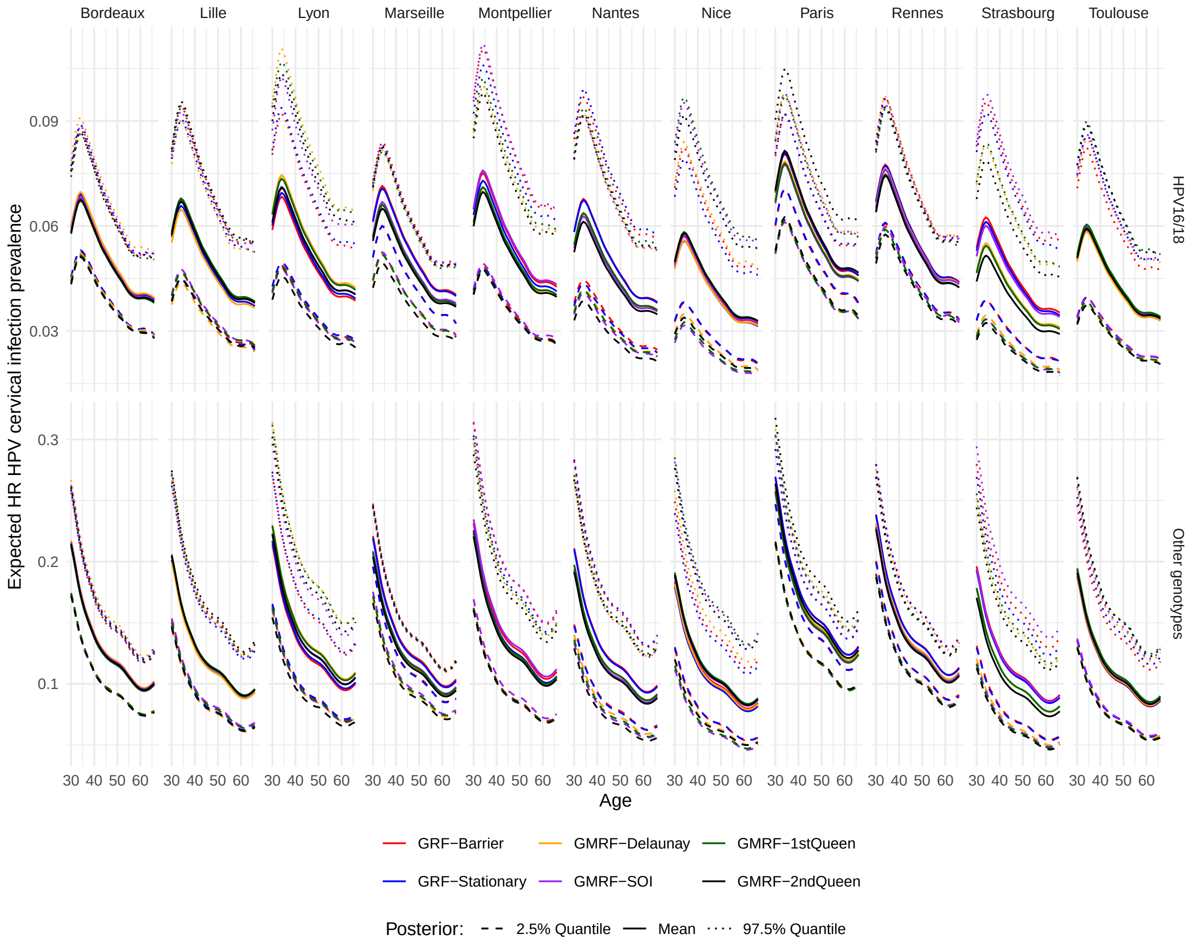

**Figure S42 Posterior average (ETI95%) for the conditional prevalence in major French cities, at the end of November 2023, stratified by genotype group and type of Matérn covariance function (stationary or non-stationary model).**

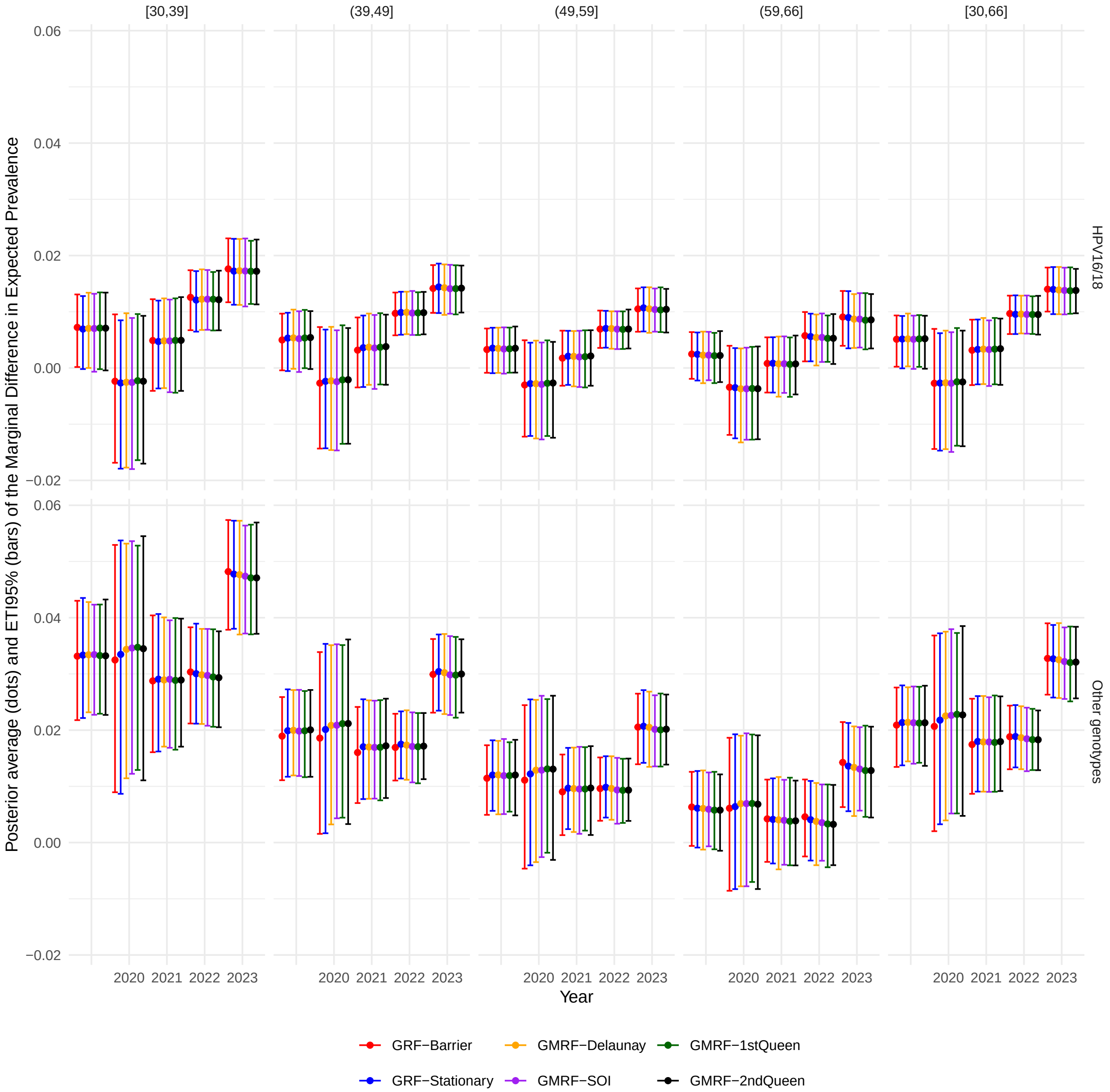

**Figure S43** **Posterior average (ETI95%) for the Marginal Difference in Expected Prevalence, stratified by the type of spatial component, age, and year.**
